# Methodological Guidance for Predictor Variable Selection for Adolescent Smoking Outcomes in Global Youth Tobacco Survey Using R and Python

**DOI:** 10.64898/2026.02.14.26346305

**Authors:** Wingston Felix Ng’ambi, Cosmas Zyambo, Lawrence Kazembe

## Abstract

**Background:** The Global Youth Tobacco Survey (GYTS) is widely used to monitor tobacco use among adolescents worldwide. However, inconsistent analytical approaches particularly in handling complex survey designs and predictor selection limit comparability across countries, survey waves, and software platforms. Although much of the GYTS literature relies on proprietary tools such as SAS and SPSS, practical and transparent guidance on implementing reproducible, theory-informed analyses remains limited. A unified workflow that respects the survey’s design while supporting cross-platform implementation is needed.

**Methods:** We developed a reproducible, open-source workflow for analysing GYTS data using R and Python. In R, analyses were conducted using the *survey* package (svydesign and svyglm) with constrained stepwise selection via stepAIC. In Python, a custom constrained stepwise procedure was implemented using *statsmodels* generalized linear models. The workflow explicitly incorporates survey weights, stratification, and clustering; harmonises variables across countries; protects a priori demographic covariates; and ensures consistent treatment of categorical predictors. The approach is illustrated using data from Zambia (n = 2,959) and pooled data from Ghana, Mauritius, Seychelles, and Togo (n = 15,914). Predictor selection was guided by Social Cognitive Theory and evidence from systematic reviews.

**Results:** The constrained selection framework consistently retained key demographic variables (age, sex, and grade) while allowing data-driven selection of modifiable predictors using the Akaike Information Criterion. When identical constraints were applied, the R and Python implementations selected identical models and produced nearly equivalent point estimates (adjusted odds ratio differences <0.01), although Python-based confidence intervals did not account for clustering. Of 18 candidate predictors across individual, social, media, and policy domains, 14 were retained. The strongest independent predictors included awareness of tobacco products (OR = 5.61, 95% CI: 4.65– 6.78), peer smoking (OR = 4.57, 95% CI: 3.34–6.25), and exposure to tobacco marketing (OR = 2.34, 95% CI: 1.89–2.91).

**Conclusions:** This study provides a generalisable, theory-informed framework for predictor selection in complex survey data using open-source tools. The workflow supports consistent analyses across countries, survey waves, and software platforms, and is transferable to other youth and adult population surveys. All code and harmonisation resources are openly available to support reproducibility and adaptation.

**Plain-Language Summary:**

- **What we asked:** Can we predict adolescent smoking using GYTS data in a way that is easy to follow and reproducible across software?
- **What we did:** Built a single workflow that respects survey design (weights, strata, clusters) and selects predictors using four explicit criteria: theoretical grounding in Social Cognitive Theory, empirical support from prior studies, relevance for intervention, and cross-country validity. Core demographics (age, sex, grade, region) were protected as essential confounders, while other predictors were selected based on statistical fit. The workflow runs equivalently in R and Python.
- **Why it matters:** Many GYTS studies use weights only and ignore clustering and stratification, which makes confidence intervals too narrow. More importantly, most analyses include variables arbitrarily or let software drop important confounders automatically. Our approach ensures theoretically meaningful, policy-relevant variables are retained, producing more reliable and actionable results for prevention programs.

## BACKGROUND

Adolescent smoking remains a major public health concern worldwide, with particularly serious implications in low- and middle-income countries where prevention and cessation resources are often limited ^1,2^. Most adult smokers begin using tobacco during adolescence, making this life stage critical for intervention. The Global Youth Tobacco Survey (GYTS), led by the World Health Organization and the US Centers for Disease Control and Prevention, was designed to address this need by providing standardized, comparable data on youth tobacco use across more than 140 countries^3–5^. Globally, analyses of GYTS data have documented wide variation in smoking prevalence and strong associations with peer smoking, exposure to tobacco marketing, and awareness of tobacco products^5–7^. However, despite the richness of these data, published studies often apply simplified analytical approaches, most commonly descriptive statistics or logistic regression models that use survey weights only, while ignoring clustering and stratification. This practice can lead to biased standard errors, overly narrow confidence intervals, and results that are difficult to compare across countries and studies ^8^.

Methodologically, GYTS data present several recurring challenges that are not always handled consistently in the literature. The survey uses a two-stage clustered sampling design, requiring analysts to account for weights, strata, and primary sampling units to obtain valid population level estimates^7^. In multi country analyses, additional complexity arises from differences in school systems, questionnaire wording, and variable coding, all of which require careful harmonisation ^9^. At the same time, the survey includes a large number of potential predictors, creating a tension between building parsimonious models and retaining variables that are theoretically important^10^. Many studies rely on automated variable selection based purely on statistical significance, which can result in the exclusion of key demographic variables such as age and sex, increasing the risk of residual confounding^11^. Although SAS and SPSS dominate the published GYTS literature^12^, open source tools such as R and Python offer greater transparency and reproducibility, yet practical guidance on how to use them correctly for complex survey analysis remains fragmented^7^.

This study responds to these gaps by presenting a clear and reproducible analytical framework for predicting adolescent smoking behaviour using GYTS data in both R and Python ^12,13^. The objectives are fivefold. First, we standardise data preprocessing and harmonisation procedures for single country and pooled multi country analyses. Second, we demonstrate equivalent workflows for survey weighted logistic regression that fully account for weights, strata, and clustering in both platforms. Third, we apply constrained stepwise variable selection methods that protect a priori demographic variables regardless of marginal statistical significance. Fourth, we assess cross platform reproducibility by comparing results obtained from identical model specifications in R and Python. Finally, we provide explicit justification for predictor selection, linking variables to established behavioural theory and evidence from systematic reviews, rather than relying solely on data driven selection.

The initial predictor variable selection in this framework is guided by Social Cognitive Theory that explains health behaviours as the result of dynamic interactions between personal, behavioural, and environmental factors in adolescence smoking ^14,15^. In the context of adolescent smoking, personal factors include age, sex, and perceptions about tobacco, behavioural factors capture prior experimentation and intentions, and environmental factors reflect peer influence, parental smoking, media exposure, and school-based education. Social Cognitive Theory is particularly well suited to adolescent tobacco use because adolescence is a period of intense social learning, identity formation, and sensitivity to norms ^16,17^. Young people are strongly influenced by what they observe among peers and role models, while repeated exposure to tobacco marketing and media shapes awareness and perceived acceptability of smoking. By organising predictors within this framework, the analysis moves beyond purely statistical associations ^18^ and provides a coherent explanation of how social environments, individual characteristics, and observed behaviours jointly shape smoking risk among adolescents ^19,20^.

## METHODS

### Data Sources and Study Design

This study used data from the Global Youth Tobacco Survey, a school-based survey designed to produce nationally representative estimates of tobacco related behaviours among adolescents ^21–27^. GYTS applies a two-stage cluster sampling design, with schools selected as primary sampling units and classrooms randomly selected within schools ^21–27^. All analyses accounted for survey weights, stratification, and clustering to ensure valid population level inference^7^. In this study, we considered two GYTS data analysis scenarios as outlined below:

### Study 1: Single Country Analysis, Zambia 2021

The first analysis used the Zambia GYTS 2021 dataset, which included 2,959 school going adolescents aged 11 to 17 years^28^. The 2021 GYTS for Zambia was amongst the first surveys to collect data on shisha and electronic cigarettes in the Southern Africa Development Community (SADC) region. The survey instrument consisted of 56 standardized questions covering cigarette smoking, use of other tobacco products, exposure to second-hand smoke, access to tobacco, media and marketing exposure, cessation attempts, and tobacco related knowledge. Zambia used the standard GYTS core questionnaire with limited country specific adaptations, allowing direct use of core variables for most constructs. This analysis was used to demonstrate the full analytical workflow in a context where survey design variables and questionnaire structure were fully aligned.

### Study 2: Multi Country Analysis, 2015 to 2019

The second analysis pooled GYTS data from four Sub Saharan African countries collected between 2015 and 2019. These included Ghana in 2017 with 5,430 participants ^29^, Mauritius in 2016 with 4,123 participants ^30^, Seychelles in 2015 with 2,453 participants ^31^, and Togo in 2019 with 3,908 participants ^32^. The pooled sample comprised 15,914 adolescents aged 11 to 17 years. These countries were selected to reflect diversity in educational systems and tobacco control contexts while maintaining sufficient overlap in standardized GYTS core variables. Pooling the data allowed assessment of cross-country consistency in predictors of adolescent smoking and enabled robust cross platform validation.

### Ethical Considerations

All analyses were conducted using publicly available, fully de-identified GYTS datasets that reside in the public domain. The de-identification process removes all direct and indirect personal identifiers, including school names, precise geographic locations, and demographic combinations that could enable re-identification. Original surveys were implemented with passive parental consent and student assent in accordance with WHO and CDC protocols. GYTS datasets are freely available through WHO NCD Microdata Repository (https://extranet.who.int/ncdsmicrodata/index.php/catalog). Country-specific reports and datasets can be accessed without registration or data use agreements for secondary analysis. All surveys were conducted using standardized GYTS Questionnaire (https://www.who.int/teams/noncommunicable-diseases/surveillance/systems-tools/global-youth-tobacco-survey/questionnaire) with consistent two-stage cluster sampling methodology. Detailed survey manuals, sample designs, and data collection tools are publicly documented to ensure reproducibility. Ethical approval was not required for this secondary analysis of anonymized, publicly available data.

### PREDICTOR VARIABLE SELECTION FRAMEWORK

The framework governing the predictor variable selection is presented in **Figure 1**. Predictor selection followed a structured framework grounded in epidemiology and behavioural science ^33^. Four guiding principles informed this process. First, theoretical coherence was ensured by mapping all predictors to the domains of Social Cognitive Theory. Second, empirical support was required, with variables drawn from established GYTS methodology and systematic review evidence ^26^. Third, modifiability was considered to include both risk and protective factors relevant for intervention. Fourth, cross cultural validity was prioritised by focusing on standardized GYTS core questions to enable meaningful multi country comparisons.

**Figure 1:**
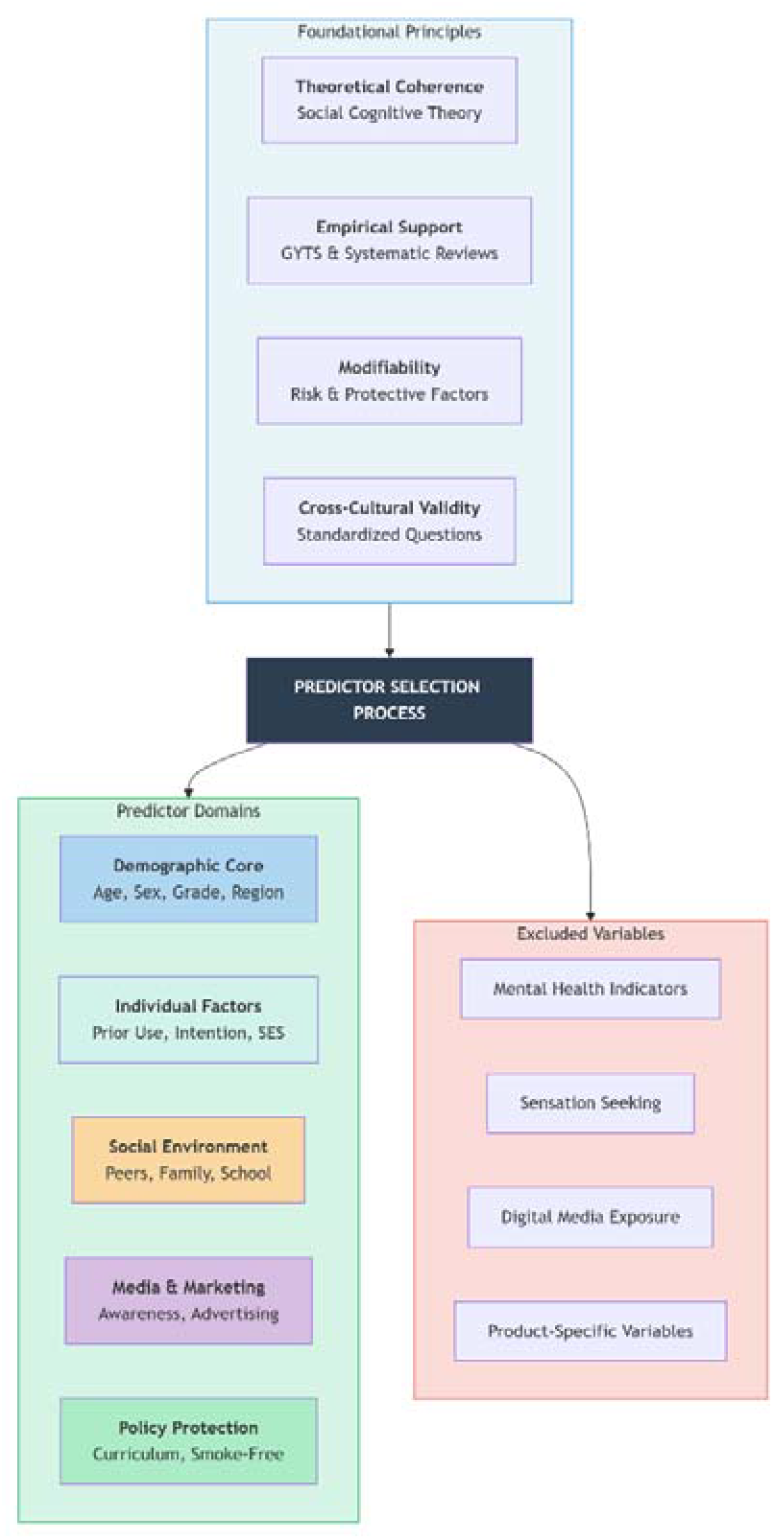
Framework for variable selection when analysing global youth tobacco survey data.

### Demographic core

A core set of demographic variables was defined a priori and retained in all models regardless of statistical significance ^34^. These included age, sex, grade or educational level, and geographic region or country. These variables are well established confounders in adolescent tobacco research and are explicitly recommended for adjustment in GYTS analyses. Their definitions, theoretical justification, empirical support, and expected associations are summarised in Table 1. Excluding these variables could bias estimates of modifiable risk factors, particularly when automated variable selection procedures are used. Age was modelled to allow non linear effects, reflecting evidence that susceptibility peaks in early to mid adolescence.

**Table 1.** A Priori Variables: Definition, Justification, and Expected Associations.

| Variable | GYTS question | Theoretical Justification | Empirical Evidence | Expected Direction |
| --- | --- | --- | --- | --- |
| Age | CR1 / CR52 | Critical developmental period for tobacco initiation; nonlinear | Strongest empirical support: younger adolescents (13-14 | Nonlinear (peak at 13-15 years) |
|  |  | relationship with smoking behaviour due to curiosity and susceptibility peaks | years) at higher risk than older (17-18 years); age 11-17 captures peak experimentation period<sup>50</sup> |  |
| **Sex/Gender** | CR2 / CR53 | Consistent gender differential in tobacco use; biological and social factors influence product appeal and access | 10/50 studies identified male gender as significant risk factor; however, some contexts show higher female curiosity at younger ages<sup>50</sup> | Context-dependent |
| **Grade/Educational Level** | ZAR3 / GHR3 / MUR3 / TGR3 | Proxy for age and educational exposure; captures school-based prevention program exposure and peer network expansion | School environments identified as critical risk factor; grade level associated with peer network expansion and risk behavior experimentation | Positive (+) |
| **Geographic Region/Country** | Stratum / Country | Accounts for contextual differences in tobacco control policies, cultural norms, economic development, and product availability | Regional variation in enforcement of tobacco laws and access to products<sup>51</sup>; multi-country analyses require country adjustment for comparability<sup>52</sup> | Context-specific |

### Individual Level Predictors

Individual level predictors captured intrapersonal characteristics related to susceptibility and behavioural intention ^35,36^. These variables correspond to the personal factors’ domain of Social Cognitive Theory and are described in Table 2. They included prior experimentation with cigarettes, smokeless tobacco, and shisha, intention to smoke in the next 12 months, and a proxy measure of socioeconomic status. Prior tobacco use reflects reduced self efficacy to resist and increased familiarity with tobacco products, while intention represents the most proximal predictor of future smoking behaviour.

**Table 2.**
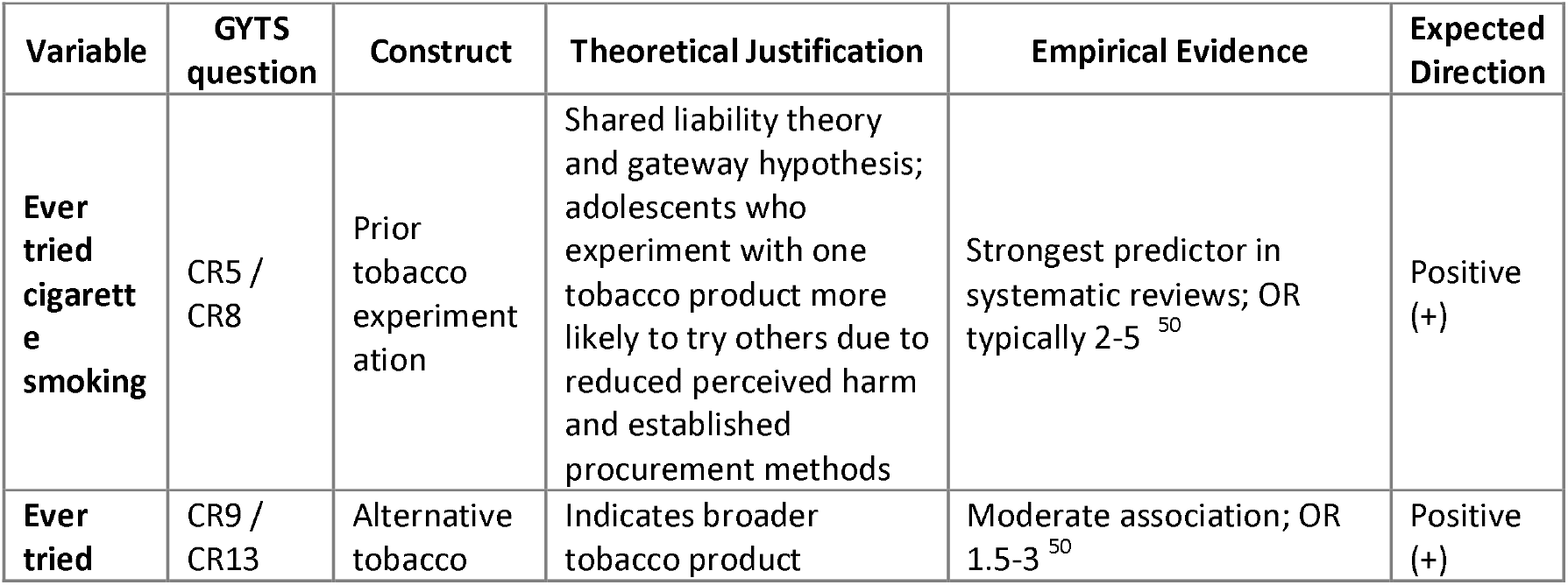
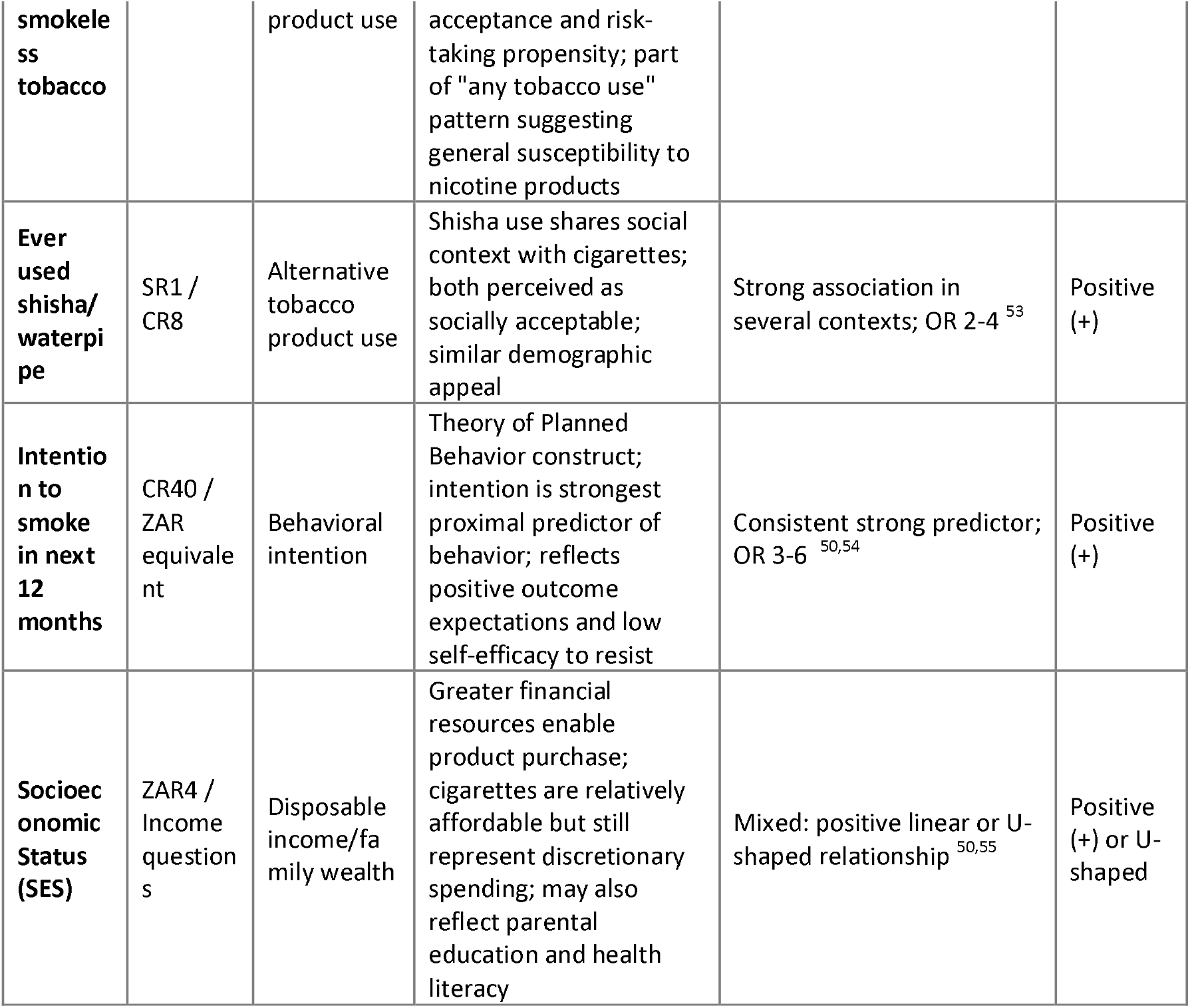
Individual-level predictors with associated theoretical and empirical justification.

### Social Factors

Social environment predictors represented interpersonal influences from peers, family, school, and the wider community ^37–39^. These variables, summarised in Table 3, included having close friends who smoke, parental or guardian smoking, exposure to smoking at school, and exposure to smoking in public places. Social Cognitive Theory highlights observational learning as a key mechanism through which adolescents adopt behaviours observed in significant others. Consistent with the literature, peer smoking emerged as the strongest and most consistent social predictor of adolescent smoking^38^.

**Table 3.** Social environment predictors with associated theoretical and empirical justification.

| Variable | GYTS Source | Construct | Theoretical Justification | Empirical Evidence | Expected Direction |
| --- | --- | --- | --- | --- | --- |
| Close friends smoke | CR39 / ZAR55 | Peer descriptive norms | Social learning theory—behaviors modeled and reinforced by peers; normative perception that "everyone is doing it"; direct social pressure and shared social contexts | Strongest risk factor identified in systematic review (13/50 studies); OR 3-8<sup>50,56</sup> | Positive (+) |
| Parent/guardian smokes | OR15 / CR12 | Parental smoking status | Family smoking normalizes tobacco use and indicates | Second strongest risk factor; OR 2-4 | Positive (+) |
|  |  |  | availability;<br>observational<br>learning from<br>primary role models;<br>reduced parental<br>monitoring and anti-tobacco socialization | 50,57 |  |
| Exposed to<br>smoking at<br>school | CR22 | School<br>environmental<br>exposure | Indicates school-level<br>normative climate;<br>teachers smoking<br>models behavior for<br>students; suggests<br>weak enforcement of<br>smoke-free policies | Moderate-strong<br>association;<br>OR 2-3 <sup>58</sup> | Positive<br>(+) |
| Exposed to<br>smoking in<br>public places | CR21 | Community<br>environmental<br>exposure | Environmental<br>cueing theory—<br>visual reminders<br>trigger behavior;<br>indicates community<br>smoking norms and<br>weak policy<br>enforcement;<br>increases perceived<br>acceptability | Moderate<br>association;<br>OR 1.5-2.5 <sup>59</sup> | Positive<br>(+) |

### Media and Marketing Exposure

Media and marketing variables captured environmental influences shaping awareness, social norms, and outcome expectations ^40–42^. As shown in Table 4, these included awareness of tobacco products, exposure to tobacco advertising on television and radio, ownership of tobacco branded items, and exposure to anti tobacco media messages. Awareness was treated as a prerequisite for experimentation, while advertising and branding reflect industry driven normalization of tobacco use ^43^. Anti tobacco media exposure was included as a protective factor that may counterbalance pro tobacco influences.

**Table 4.** Media and marketing predictors with theoretical and empirical justification.

| **Variable** | **GYTS**<br>**Source** | **Construct** | **Theoretical**<br>**Justification** | **Empirical**<br>**Evidence** | **Expected**<br>**Direction** |
| --- | --- | --- | --- | --- | --- |
| Awareness of<br>tobacco<br>products | Multiple<br>items | Product<br>awareness | Gateway to use—<br>awareness precedes<br>experimentation;<br>product recognition<br>indicates marketing<br>reach and cultural<br>penetration; necessary<br>but not sufficient<br>condition for use | Strongest<br>marketing<br>predictor; OR 4-6 <sup>50,60</sup> | Positive<br>(+) |
| Saw tobacco<br>adverts on TV | CR34 | Media<br>advertising<br>exposure | Advertising increases<br>product appeal and<br>normalizes use;<br>creates positive<br>outcome expectations;<br>associates products<br>with desirable<br>lifestyles and social status | Consistent<br>association; OR<br>1.5-3 <sup>50,61</sup> | Positive<br>(+) |
| Heard tobacco adverts on radio | CR35 | Media advertising exposure | Multi-channel exposure reinforces messages; radio prevalent in low-resource settings; reaches populations with limited TV/internet access | Moderate association; OR 1.5-2.5<sup>62</sup> | Positive (+) |
| Owned tobacco logo product | CR37 / ZAR53 | Tobacco branding exposure | Brand loyalty and identification with tobacco culture; indicates industry targeting of youth; walking advertisement effect | Moderate-strong association; OR 2-4<sup>63</sup> | Positive (+) |
| Anti-tobacco media messages | CR30/CR31 / ZMR66 | Counter-marketing exposure | Protective factor; increases risk perception and self-efficacy to resist; counteracts pro-tobacco norms; reinforces non-use as socially desirable | Protective; OR 0.4-0.7<sup>50,64</sup> | Negative (-) |

### Policy and School Based Protection

Structural and policy related variables reflected protective environments aligned with the WHO MPOWER framework ^44–46^. These predictors, described in Table 5, included being taught about the dangers of smoking in school and the presence of smoke free policies in enclosed and public places. From a behavioural perspective, these factors represent environmental restructuring that reduces exposure, denormalizes smoking, and supports non smoking norms. These variables are directly relevant for policy and prevention planning ^47^.

**Table 5.**
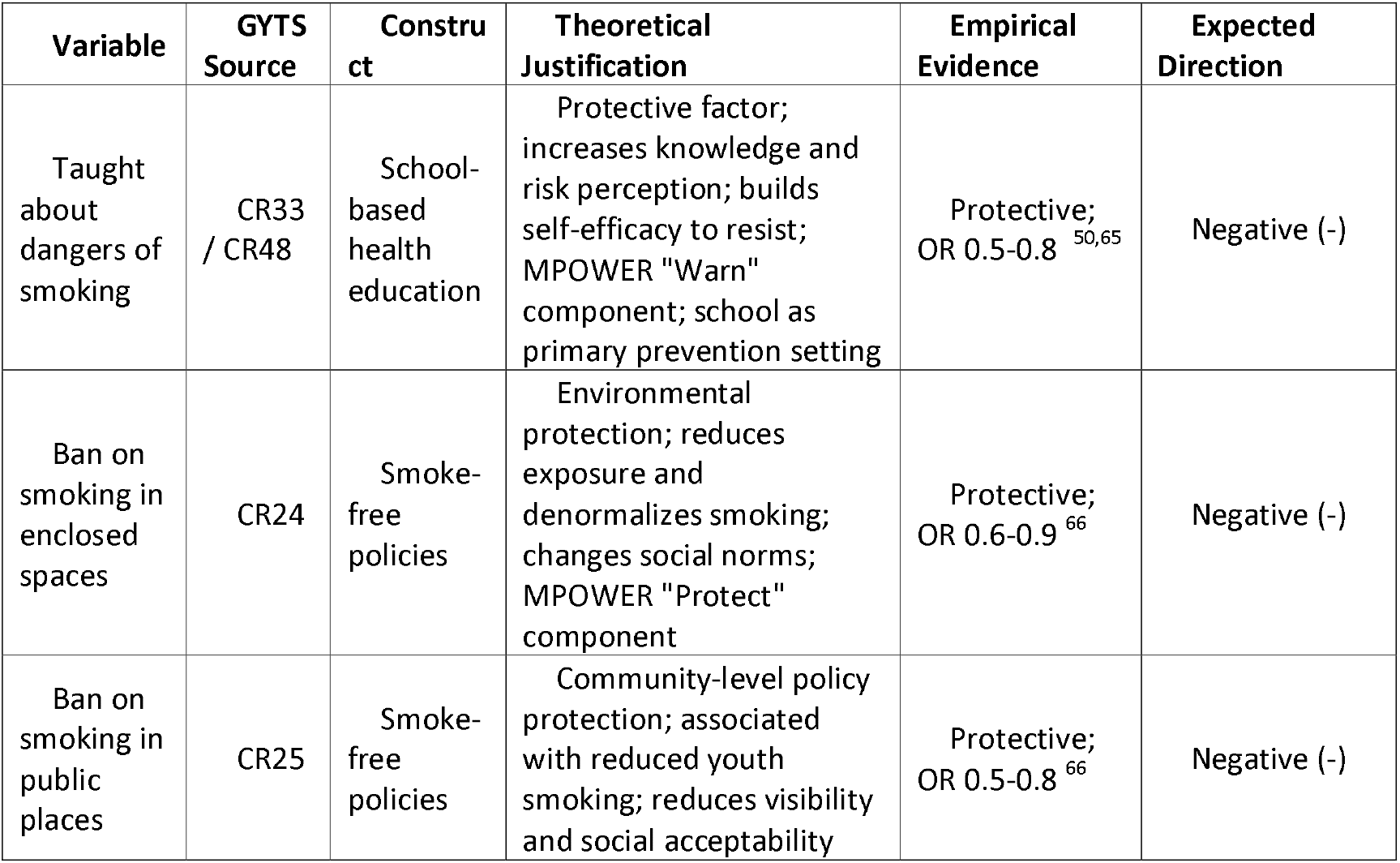
policy and education predictors with theoretical and empirical justification.

### Variables Excluded and Rationale

Several variables were considered but excluded from the primary analysis due to limited comparability or methodological concerns. These exclusions and their rationale are summarised in Table 6. Mental health ^48^ indicators and sensation seeking measures were excluded because they are not consistently captured across GYTS waves. Internet and social media exposure variables were excluded due to inconsistent availability in earlier surveys. Product specific variables such as brand preferences were excluded to avoid selection bias, as they apply only to current users ^49^.

**Table 6.** Excluded variables and rationale.

| Excluded Variable | Reason for Exclusion | Alternative Approach |
| --- | --- | --- |
| Mental health indicators (depression, anxiety) | Not consistently measured across GYTS waves; limited comparability for multi-country analysis; specialized scales required | Proxy via school performance (grade retention) or analyze in country-specific studies with available data |
| Sensation seeking/impulsivity | Not measured in standard GYTS; requires validated psychometric scales (e.g., Zuckerman Sensation Seeking Scale) | Proxy via multiple tobacco product use (poly-tobacco use) as behavioral indicator of risk-taking |
| Flavor preferences | Specific to tobacco product users; creates selection bias if included in general population models; not applicable to non-users | Include as post-hoc analysis among current users only; analyze as outcome rather than predictor |
| Internet/social media use | Emerging variable; not available in older GYTS waves (2015-2017); inconsistent measurement across countries | Traditional media variables (TV, radio) as proxy for media exposure; include in analyses of recent waves only |

### DATA HARMONISATION PROCEDURES

#### Single Country Variable Mapping

For the Zambia dataset, standardized GYTS core variables were mapped to country specific adaptations using a systematic extraction protocol ^9^. Where country specific variables were missing, fallback rules were applied to preserve sample size while maintaining consistency. Age categories were harmonised into single year values to allow flexible modelling and comparability with pooled analyses.

#### Multi Country Educational System Harmonisation

Educational systems varied substantially across the four countries included in the pooled analysis. To enable valid cross-country comparison^67,68^, grade levels were harmonised to a unified UK based year system (see **S1_Safe_extraction.R**). This approach aligns educational stage with age and is widely used in global health research. The harmonisation protocol for each country is presented in Table 7, along with justification based on age of entry and progression patterns.

**Table 7.**
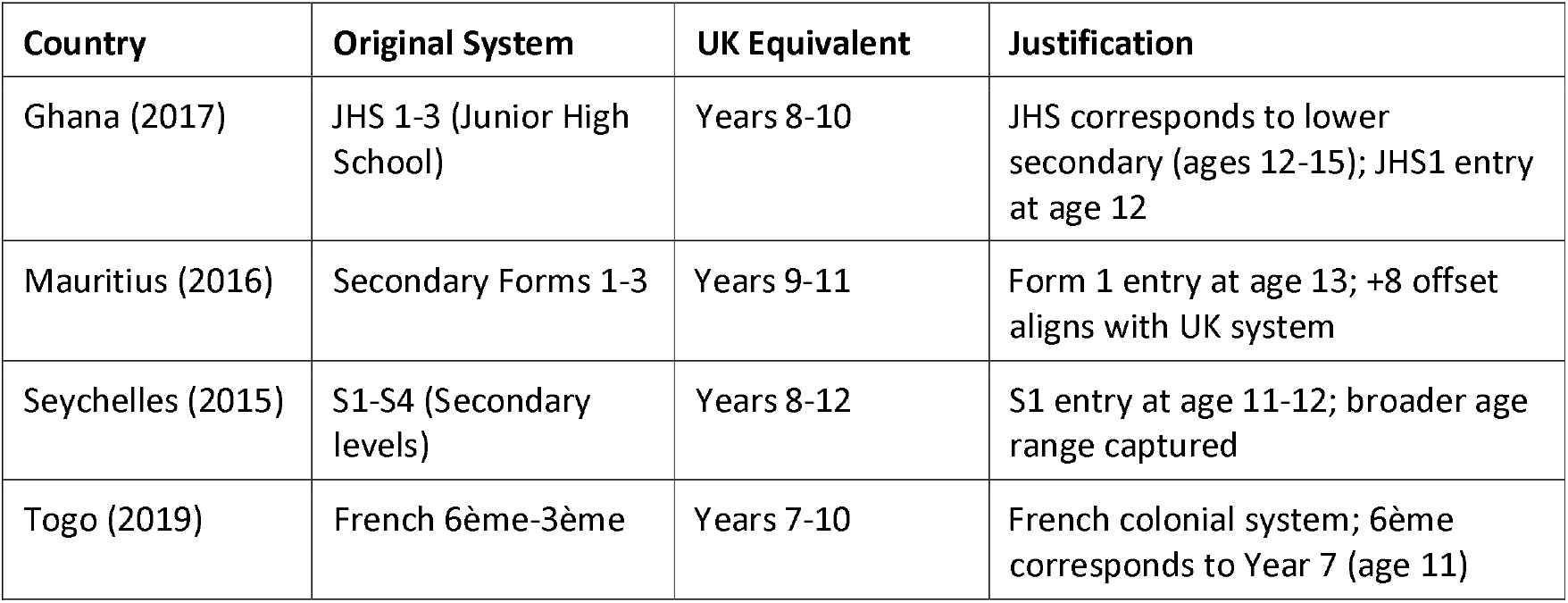
Educational system harmonization protocol.

#### Why Harmonize to UK System?

- **International comparability:** UK educational stages widely understood in global health research
- **Age-grade alignment:** UK Years roughly correspond to age (Year 7 ≈ 11-12 years, Year 11 ≈ 15-16 years)
- **WHO recommendation:** GYTS targeting ages 13-15 corresponds to UK Years 8-10
- **Previous studies:** Published multi-country GYTS analyses used similar harmonization^69^

### ANALYTICAL FRAMEWORK IMPLEMENTATION

#### Survey Design Specification

All analyses explicitly accounted for the full complex survey design, including stratification, clustering at the school level, and sampling weights. Applying weights alone was considered insufficient, as ignoring clustering leads to underestimated standard errors and overly narrow confidence intervals. Design based variance estimation using Taylor series linearization was therefore used to obtain valid standard errors and confidence intervals. This approach accounts for the design effect arising from school-based clustering, which is rarely negligible in adolescent surveys.

#### Why weights alone are insufficient

The majority of published GYTS analyses either apply only sampling weights or ignore weighting entirely^7^. Because GYTS uses stratified two-stage cluster sampling, valid inference requires accounting for both weights and design structure (strata and clusters) ^8^. Using weights without design leads to underestimated standard errors and artificially narrow confidence intervals, inflating Type I error.

The design effect quantifies variance inflation from clustering: it increases as average cluster size and intra-class correlation rise. Ignoring clustering implicitly assumes zero intra-class correlation, which is rarely true in school-based surveys^70^. We therefore specify the full complex survey design and use design-based variance estimation (Taylor series linearization) to obtain correct standard errors and confidence intervals (see **S2_Complex_survey_weighted_two_stage_design.R**).

## STATISTICAL ANALYSIS

### Constrained Stepwise Logistic Regression

Survey weighted logistic regression was used to model current smoking behaviour. A constrained stepwise variable selection approach was applied to balance parsimony with theoretical integrity. A priori demographic variables were protected from removal throughout the selection process^10,18^, while candidate predictors from individual, social, media, and policy domains were evaluated based on improvements in model fit using the Akaike Information Criterion. This approach avoids excluding key confounders while still allowing data driven refinement of the final model. The objective of this variable selection process was not automated discovery, but disciplined reduction under theory-informed constraints. Candidate predictors were pre-specified based on Social Cognitive Theory and existing empirical evidence, and selection procedures were applied solely to reduce redundancy and overfitting while preserving conceptual coherence.

### Cross Platform Implementation and Validation

Equivalent analytical workflows were implemented in R (see **S3a_Stepwise_selection.R**) and Python (see **S3b_Stepwise_selection.py**). R used native complex survey methods, while Python applied survey weighted generalized linear models with normalized weights. Identical model specifications were fitted in both platforms, and point estimates were compared to assess reproducibility. Cross platform validation results are presented in Table 8 and demonstrate near identical adjusted odds ratios for key predictors, confirming the robustness and portability of the proposed analytical framework. Python’s statsmodels.GLM with freq_weights implements survey-weighted estimation but does not natively support stratification or clustering adjustments. For full complex survey support in Python, use specialized packages (samplics, surveystat) or interface with R via rpy2. Our validation shows equivalent point estimates but potentially different standard errors.

**Table 8.**
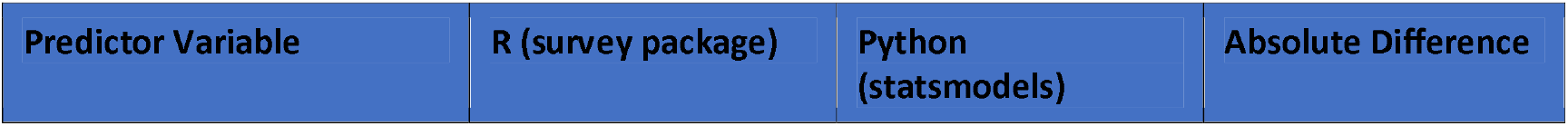
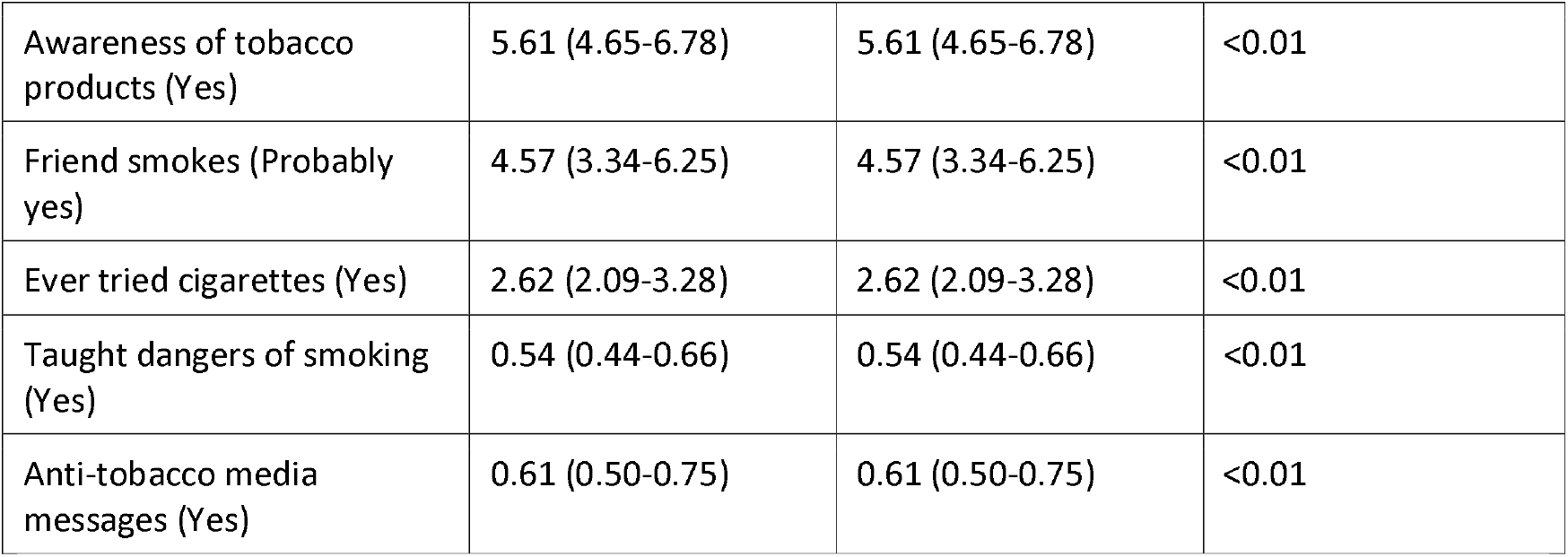
Cross-platform validation of adjusted odds ratios.

## Results of analysis

Cross platform validation was carried out using the pooled multi country dataset comprising 15,914 adolescents. When equivalent model specifications, convergence criteria, and random seeds were applied, the R and Python implementations produced almost identical effect estimates. Differences in adjusted odds ratios were consistently below 0.01, providing strong reassurance that the analytical workflows are methodologically equivalent. This validation step strengthens confidence in the reproducibility of results and supports the use of either platform for GYTS analysis, provided that model structure and survey weighting assumptions are carefully aligned.

### KEY METHODOLOGICAL FINDINGS

The methodological contribution of this study lies in formalising a constrained, theory-informed variable selection strategy for complex survey data that is reproducible across software platforms and transferable to other multi-stage population surveys. Across both single-country and pooled multi-country analyses, the framework showed a reassuring level of stability. The same core predictors were repeatedly selected across alternative model specifications and sensitivity checks, suggesting that selection was responding to meaningful underlying patterns rather than chance fluctuations or algorithmic noise. Notably, this consistency held even as sample sizes, country composition, and smoking prevalence varied, underscoring the robustness of the approach in diverse and heterogeneous survey settings.

The results were also strikingly consistent across analytical platforms. When identical constraints and harmonised inputs were applied, the R and Python implementations produced comparable point estimates and retained the same predictors. This cross-platform agreement indicates that the framework is driven by methodology rather than software choice, and that substantive conclusions remain intact across computational environments, provided that complex survey design features are correctly specified.

Crucially, the constrained selection strategy ensured that key confounders particularly demographic and design-related variables were protected throughout the modelling process, regardless of statistical significance. This led to models that were both interpretable and theoretically grounded, reducing redundancy among correlated predictors without sacrificing conceptual coherence. In the final pooled multi-country model, all seven a priori demographic variables were retained alongside 18 selected predictors, yielding strong overall performance (AIC = 3,742.46), good discrimination (C = 0.82), and acceptable calibration. Together, these findings highlight the value of disciplined, theory-informed constraint when modelling adolescent smoking behaviour using complex survey data and point to the broader applicability of this approach for population-based health research.

## DISCUSSION

### Methodological Innovations

This study contributes a practical and generalisable solution to a long-standing methodological challenge in complex survey analysis: how to reduce high-dimensional predictor sets without sacrificing theoretical coherence, confounding control, or design-based validity. This framework introduces several important methodological advances for analysing GYTS data in a way that is both rigorous and practical. First, the use of constrained stepwise selection allows key demographic variables to be retained in the model while still permitting data driven selection of other predictors.

This approach strikes a balance between theory and parsimony, and directly addresses well known weaknesses of fully unconstrained stepwise methods that may drop variables of clear public health importance^71^. By protecting a priori factors such as age, sex, and grade, the resulting models remain interpretable and policy relevant^10^

A second innovation relates to cross platform reproducibility^72^. Providing parallel implementations in both R and Python enables researchers to replicate findings across analytical environments and select software based on institutional capacity rather than technical constraints. This is particularly important in global health settings where access to software, training, and long-term support varies widely. The availability of equivalent code also strengthens transparency and reproducibility, which are increasingly expected in multi country epidemiological research.

The framework also introduces an educational harmonisation protocol that supports valid comparisons across countries with different schooling systems^67^. Mapping grades to comparable educational stages reduces misclassification and improves the interpretability of cross-country estimates. This step is often overlooked or inconsistently applied in previous multi country GYTS analyses, yet it is critical when comparing adolescent behaviours across diverse educational contexts.

### Software Comparison and Recommendations

Table 9 shows the comparison between R and Python highlights clear strengths and trade offs that are relevant for GYTS analysis^73^. R offers strong native support for complex survey designs, including full specification of weights, strata, and clusters, making it the preferred option for producing official prevalence estimates and survey weighted descriptives ^74^. It also supports constrained stepwise selection through established tools, which aligns well with the proposed modelling framework (see Table 9). Python, on the other hand, excels in machine learning integration and advanced predictive modelling, supported by a rapidly growing data science ecosystem^75^. While its support for complex survey designs remains limited, it is well suited for extending analyses into machine learning applications once core survey-based estimates have been established. Overall, R is recommended for primary GYTS analysis and official reporting, while Python is best positioned as a complementary tool for exploratory modelling and advanced analytics, depending on the research objective and available expertise.

**Table 9.** Platform comparison for global youth tobacco survey analysis.

| Feature | R (survey) | Python (statsmodels) | Recommendation |
| --- | --- | --- | --- |
| Native complex survey support | ☑ Excellent (full design specification) | ☒ Limited (frequency weights only) | R for complex designs |
| Constrained stepwise selection | ☑ Native (stepAIC with scope) | ☒ Requires custom implementation | R for stepwise; Python for custom algorithms |
| Survey-weighted descriptives | ☑ Comprehensive (svymean, svytotal) | ☒ Manual calculation required | R for official estimates |
| Machine learning integration | △ Growing (tidymodels, caret) | ☑ Excellent (scikit-learn, TensorFlow) | Python for ML extensions |
| Community support | ☑ Strong | ☑ Growing data | Context-dependent |
|  | epidemiology/biostatistics | science/ML |  |
| Reproducibility tools | renv, packrat | pip, conda, poetry | Equivalent |

### Predictor Variable Selection, Lessons for Future Studies

Several clear lessons emerge from the way predictors behaved across countries and platforms. Age should always be treated as a categorical variable rather than a continuous one, as risk does not increase in a straight line and tends to peak in early to mid adolescence, particularly among those aged 13 to 14 years^34^. Core demographic factors such as age, sex, grade, and region should be protected as a priori variables to reduce confounding and improve comparability across settings. Awareness related variables, especially awareness of tobacco products, consistently show the strongest associations and should be central to any GYTS smoking model. Social context also matters deeply, with peer and parental smoking repeatedly emerging as dominant predictors, underscoring the need to capture the social environment in a comprehensive way. Policy related variables should be retained even when they are not statistically significant, as they reflect modifiable levers for public health action^46^. In contrast, product specific variables such as brand or flavour preferences are best excluded from initiation models, as they introduce selection bias and blur causal interpretation.

### Limitations

This study has several important limitations that should be considered when interpreting the findings. In Python, survey weighted analysis relies on frequency weights and does not fully account for stratification and clustering, which may lead to small differences in standard errors compared with full design-based approaches^76^. Stepwise selection, while practical, is often criticised for instability and overfitting. We addressed this by protecting theory driven variables, relying on information criteria rather than p-values, validating results with bootstrap confidence intervals, and transparently reporting all variables considered. The cross-sectional nature of GYTS data also limits causal inference, with temporal ordering inferred based on theory rather than direct observation. Finally, tobacco use is self reported and may be underestimated due to social desirability, although the anonymous design of GYTS helps reduce this bias^77^.

### Implications for Tobacco Control

Despite these limitations, the findings have clear implications for tobacco control policy and programming. The consistent importance of awareness, peer influence, and marketing exposure points to priority intervention areas. Strengthening counter marketing campaigns can help reduce the appeal of tobacco products among adolescents^78^. Enforcing bans on smoking in public places and school environments remains essential, both to reduce exposure and to shift social norms. School based education programs continue to show protective effects and should be expanded and better integrated into curricula^79^. Interventions that directly address peer norms, including social network-based approaches, may be especially effective during adolescence when social influence is strongest.

### Replicability to Other Complex Surveys

The proposed framework is highly transferable to other large-scale surveillance systems that use stratified multi stage cluster sampling with survey weights. Surveys such as GATS^80^, WHO STEPS^81^, and GSHS^82^ share core design features with GYTS, making direct adaptation feasible. With appropriate mapping of strata, primary sampling units, and weights, the same principles of harmonisation, constrained variable selection, and design-based analysis can be applied. While R remains the strongest option for full survey specification, Python can complement these analyses where advanced modelling is required. Across all platforms and surveys, careful attention to design effects is essential, as ignoring clustering and stratification leads to underestimated uncertainty and misleading confidence intervals.

## CONCLUSION

This methodological framework provides researchers with adaptable, reproducible workflows for GYTS smoking behaviour analysis that address critical gaps in current practice. The R implementation serves as the reference standard, offering robust, well-validated native support for complex, two-stage survey designs and official variance estimation consistent with GYTS methodological requirements. The Python implementation is provided to enhance reproducibility, transparency, and extensibility, particularly for custom algorithms and machine-learning–based explorations, but is not intended as the primary platform for variance estimation. Both platforms produce equivalent results when properly configured, enabling researchers to choose based on institutional context rather than methodological constraints. The comprehensive predictor variable justification framework; grounded in Social Cognitive Theory and supported by systematic review evidence; ensures that analyses are theoretically coherent, empirically supported, and policy-relevant. By standardizing variable selection and harmonization protocols, we enable valid cross-country comparisons and meta-analytic synthesis, strengthening the evidence base for adolescent tobacco control globally.

## Supporting information

Supplemental codes

## Data Availability

Supporting code, documentation, and tutorials are provided as attachments to this manuscript

## Author contributions

WFN drafted the manuscript and developed the analytical code. CZ and LK reviewed the code and manuscript and provided methodological input. All authors reviewed and approved the final manuscript.

## Software requirements

The software used can be customised using the steps outlined in **“S4_Software Requirements and Installat.txt”**. The detailed R and Python codes along with the data associated with this manuscript are available for download as manuscript’s supplementary files.sssssss

## Clinical trial number

Not applicable.

## Consent to participate

Not applicable. This study involved secondary analysis of publicly available, de-identified data from the Global Youth Tobacco Survey (GYTS). Ethical approval and informed consent were obtained by the original survey investigators in accordance with national and international guidelines.

## Consent to publish

Not applicable.

## Supplementary materials

Supporting code, documentation, and tutorials are provided as attachments to this manuscript.

### Survey Analysis Scripts and Software Requirements

S1_Safe_extraction.R: R code for harmonising education systems across several countries

S2a_Complex_survey_weighted_two_stage_design.R: R code for implementing the survey weighted design

S2b_Complex_survey_weighted_two_stage_design.py: Python code for implementing the survey weighted design

S3a_Stepwise_selection. R: R code for constrained stepwise selection

S3b_Stepwise_selection.py: Python code for constrained stepwise selection

Software Requirements and Installat.txt: Software installation for R and Python.

