## Supplemental codes for "Methodological Guidance for Predictor Variable Selection for Adolescent Smoking Outcomes in Global Youth Tobacco Survey Using R and Python": data_cleaning_ecig_afric_2026.do

```
cd "C:\Users\LENOVO\Documents\GitHub\Global_Tobacco_Survey_Analysis_2025\data\gtys\ecig"

*Ghana
use "C:\Users\LENOVO\Documents\GitHub\Global_Tobacco_Survey_Analysis_2025\data\gtys\ecig\GHANA_17.dta", clear
keep FinalWgt CR1 CR2 GHR3 GHR4 CR5 CR13 CR21 CR22 CR24 CR25 CR33 CR34 CR35 CR37 CR39 CR40 ELR1 ELR2 SR1 Stratum PSU country year


ren CR1 Age
ren CR2 Sex
ren GHR3 Grade
ren GHR4 income
egen windex = xtile(income), nq(3)
drop income
ren CR5 smk_try
ta smk_try
ren CR13 smkless_try
ren CR21 smk_public
ta smk_public
recode smk_public 0=0 1/max=1
la def smk_public 0 No 1 Yes
la val smk_public smk_public
ren CR22 smk_school
ren CR24 ban_closed_smoking
ren CR25 ban_public_smoking
ren CR33 taught_smking_dangers
ren CR34 advert_tv_smk
ren CR35 advert_radio_smk
ren CR37 tobaco_logo_product
ren CR39 friend_smk
ren CR40 smk_intend_12mo
ren ELR1 heard_ecig
ren ELR2 smk_ecig
ren SR1 shisha


recode smk_ecig 1=0 2/max=1
la def smk_ecig 0 No 1 Yes
la val smk_ecig smk_ecig

* Convert Ghana JHS grades to British school years
gen uk_year = .
replace uk_year = 8 if Grade == 1  // JHS1 -> Year 8
replace uk_year = 9 if Grade == 2  // JHS2 -> Year 9  
replace uk_year = 10 if Grade == 3 // JHS3 -> Year 10

* Add value labels for UK years
label define uk_year_lbl 8 "Year 8" 9 "Year 9" 10 "Year 10"
label values uk_year uk_year_lbl

* Verify the conversion
move uk_year Grade 

drop Grade
ren uk_year Grade

compress

save ghana_ecig_17, replace
 
*Mauritania
use "C:\Users\LENOVO\Documents\GitHub\Global_Tobacco_Survey_Analysis_2025\data\gtys\ecig\MAURITANIA_DATA_18.dta", clear

keep FinalWgt CR1 CR2 QAR3 QAR4 CR30 CR31 CR5 CR13 CR21 CR22 CR24 CR25 CR33 CR34 CR35 CR37 CR39 CR40 ELR1 ELR2 SR1 Stratum PSU country year OR45 OR59 OR60 OR46

ren CR1 Age
ren CR2 Sex
ren QAR3 Grade
ren QAR4 income
egen windex = xtile(income), nq(3)
drop income
ren CR5 smk_try
ta smk_try
ren CR13 smkless_try
ren CR21 smk_public
ta smk_public
recode smk_public 0=0 1/max=1
la def smk_public 0 No 1 Yes
la val smk_public smk_public
ren CR22 smk_school
ren CR24 ban_closed_smoking
ren CR25 ban_public_smoking
ren CR33 taught_smking_dangers
ren CR34 advert_tv_smk
ren CR35 advert_radio_smk
ren CR37 tobaco_logo_product
ren CR39 friend_smk
ren CR40 smk_intend_12mo
ren ELR1 heard_ecig
ren ELR2 smk_ecig
ren SR1 shisha


recode smk_ecig 1=0 2/max=1
la def smk_ecig 0 No 1 Yes
la val smk_ecig smk_ecig

* Convert Mauritania secondary grades to UK years
gen uk_year = Grade + 6  // 1→7, 2→8, 3→9, 4→10

* Add value labels
label define uk_year_lbl 7 "Year 7" 8 "Year 8" 9 "Year 9" 10 "Year 10"
label values uk_year uk_year_lbl

* Verify the conversion
move uk_year Grade 

drop Grade
ren uk_year Grade

drop OR45- OR60 CR30 CR31

compress

save mauritania_ecig_18, replace


*Mauritius
use "MUS2016.dta", clear
 
keep FinalWgt CR1 CR2 MUR3 MUR4 CR5 CR13 CR21 CR22 CR24 CR25 CR33 CR34 CR35 CR37 CR39 CR40 ELR1 ELR2 Stratum PSU country year 

ren CR1 Age
ren CR2 Sex
ren MUR3 Grade
ren MUR4 income
egen windex = xtile(income), nq(3)
drop income
ren CR5 smk_try
ta smk_try
ren CR13 smkless_try
ren CR21 smk_public
ta smk_public
recode smk_public 0=0 1/max=1
la def smk_public 0 No 1 Yes
la val smk_public smk_public
ren CR22 smk_school
ren CR24 ban_closed_smoking
ren CR25 ban_public_smoking
ren CR33 taught_smking_dangers
ren CR34 advert_tv_smk
ren CR35 advert_radio_smk
ren CR37 tobaco_logo_product
ren CR39 friend_smk
ren CR40 smk_intend_12mo
ren ELR1 heard_ecig
ren ELR2 smk_ecig
gen shisha=smkless_try


recode smk_ecig 1=0 2/max=1
la def smk_ecig 0 No 1 Yes
la val smk_ecig smk_ecig

* Keep original variable and create UK equivalent
clonevar Grade_mauritius = Grade
gen uk_year = Grade + 8
label define uk_year_lbl 9 "Year 9" 10 "Year 10" 11 "Year 11"
label values uk_year uk_year_lbl

* Verify the conversion
move uk_year Grade 

drop Grade
ren uk_year Grade

drop Grade_mauritius

compress

save mauritius_ecig_16, replace


*Sychelles
use SYC2015.dta, clear
 
keep FinalWgt CR1 CR2 SCR3 SCR4 CR5 CR13 CR21 CR22 CR24 CR25 CR33 CR34 CR35 CR37 CR39 CR40 ELR1 ELR2 SR1 Stratum PSU country year

ren CR1 Age
ren CR2 Sex
ren SCR3 Grade
ren SCR4 income
egen windex = xtile(income), nq(3)
drop income
ren CR5 smk_try
ta smk_try
ren CR13 smkless_try
ren CR21 smk_public
ta smk_public
recode smk_public 0=0 1/max=1
la def smk_public 0 No 1 Yes
la val smk_public smk_public
ren CR22 smk_school
ren CR24 ban_closed_smoking
ren CR25 ban_public_smoking
ren CR33 taught_smking_dangers
ren CR34 advert_tv_smk
ren CR35 advert_radio_smk
ren CR37 tobaco_logo_product
ren CR39 friend_smk
ren CR40 smk_intend_12mo
ren ELR1 heard_ecig
ren ELR2 smk_ecig
ren SR1 shisha


recode smk_ecig 1=0 2/max=1
la def smk_ecig 0 No 1 Yes
la val smk_ecig smk_ecig

* Create a combined variable with both systems
clonevar grade_seychelles = Grade
gen uk_year = Grade + 7
label define uk_year_lbl 8 "Year 8" 9 "Year 9" 10 "Year 10" 11 "Year 11" 12 "Year 12"
label values uk_year uk_year_lbl

* Verify the conversion
move uk_year Grade 

drop Grade
ren uk_year Grade

drop grade_seychelles

compress

save sychelles_ecig_15, replace


*Togo
use TOGO_DATA_19.dta, clear
 
keep FinalWgt CR1 CR2 TGR3 TGR4 CR5 CR13 CR21 CR22 CR24 CR25 CR33 CR34 CR35 CR37 CR39 CR40 ELR1 ELR2 SR1 Stratum PSU country year

ren CR1 Age
ren CR2 Sex
ren TGR3 Grade
ren TGR4 income
egen windex = xtile(income), nq(3)
drop income
ren CR5 smk_try
ta smk_try
ren CR13 smkless_try
ren CR21 smk_public
ta smk_public
recode smk_public 0=0 1/max=1
la def smk_public 0 No 1 Yes
la val smk_public smk_public
ren CR22 smk_school
ren CR24 ban_closed_smoking
ren CR25 ban_public_smoking
ren CR33 taught_smking_dangers
ren CR34 advert_tv_smk
ren CR35 advert_radio_smk
ren CR37 tobaco_logo_product
ren CR39 friend_smk
ren CR40 smk_intend_12mo
ren ELR1 heard_ecig
ren ELR2 smk_ecig
ren SR1 shisha


recode smk_ecig 1=0 2/max=1
la def smk_ecig 0 No 1 Yes
la val smk_ecig smk_ecig

* Convert Togo French-system grades to UK years
gen uk_year = .
replace uk_year = 7 if Grade == 1   // 6ème -> Year 7 (age 11-12)
replace uk_year = 8 if Grade == 2   // 5ème -> Year 8 (age 12-13)
replace uk_year = 9 if Grade == 3   // 4ème -> Year 9 (age 13-14)
replace uk_year = 10 if Grade == 4  // 3ème -> Year 10 (age 14-15)

* Add value labels
label define uk_year_lbl 7 "Year 7" 8 "Year 8" 9 "Year 9" 10 "Year 10"
label values uk_year uk_year_lbl

* Verify the conversion
move uk_year Grade 

drop Grade
ren uk_year Grade

compress

save togo_ecig_19, replace

use using ghana_ecig_17, clear
append using sychelles_ecig_15
append using mauritius_ecig_16
append using togo_ecig_19
ren country Country
la var Country Country
la var year Year
ren year Year
ren windex SES
la var SES "Social economic status"
la var Grade Grade
la var PSU PSU

recode smk_intend_12mo 1/2=0 3/4=1
la def smk_intend_12mo 0 No 1 Yes
la val smk_intend_12mo smk_intend_12mo

recode heard_ecig 2=0
recode shisha 2=0
drop if missing(smk_ecig)
recode smk_school 2=0
recode smk_try 2=0
recode ban_closed_smoking 2=0
recode ban_public_smoking 2=0
recode tobaco_logo_product 2=0
recode advert_radio_smk 1=0 3=0 2=1
recode advert_tv_smk 1=0 3=0 2=1
recode taught_smking_dangers 2/3=0
 

 la val advert_tv_smk smk_ecig
 la val smk_intend_12mo smk_ecig
 la val heard_ecig smk_ecig
 la val shisha smk_ecig
 la val smk_school smk_ecig
 la val smk_try smk_ecig
 la val ban_closed_smoking smk_ecig
 la val ban_public_smoking smk_ecig
 la val tobaco_logo_product smk_ecig
 la val advert_radio_smk smk_ecig
 la val taught_smking_dangers smk_ecig
 
 
 replace Age= Age+10
 
 
 * Add UK-style value labels
label define grade_uk 7 "Year 7" 8 "Year 8" 9 "Year 9" 10 "Year 10" 11 "Year 11" 12 "Year 12"
label values Grade grade_uk


recode friend_smk 1/2=0 3/4=1
la def friend_smk 0 No 1 Yes
la val friend_smk friend_smk

compress

save gyts_ecig_analysis_clean, replace
```
