## Supplemental codes for "Methodological Guidance for Predictor Variable Selection for Adolescent Smoking Outcomes in Global Youth Tobacco Survey Using R and Python": extract_code_properly.py

````
#!/usr/bin/env python3
"""
Better extraction of Python code from Quarto HTML with proper indentation preservation
"""
import re
from bs4 import BeautifulSoup

def extract_code_from_html_proper(html_path):
    """Extract code with proper indentation from HTML"""
    with open(html_path, 'r', encoding='utf-8') as f:
        html_content = f.read()
    
    soup = BeautifulSoup(html_content, 'html.parser')
    
    # Extract title
    title_tag = soup.find('title')
    title = title_tag.text.strip() if title_tag else "Analysis"
    
    qmd_content = []
    
    # Add YAML header
    qmd_content.append("---")
    qmd_content.append(f"title: {title.replace(':', '-')}")
    qmd_content.append("format: html")
    qmd_content.append("code-fold: true")
    qmd_content.append("---")
    qmd_content.append("")
    
    # Extract all sections
    sections = soup.find_all('section', class_='level2')
    
    for section in sections:
        # Extract section heading
        h2 = section.find('h2', class_='anchored')
        if h2:
            section_title = h2.get_text(strip=True)
            # Remove anchor link
            section_title = re.sub(r'\s*#.*$', '', section_title)
            qmd_content.append(f"## {section_title}")
            qmd_content.append("")
        
        # Find all code cells
        code_cells = section.find_all('div', class_='sourceCode')
        
        for cell in code_cells:
            pre = cell.find('pre')
            if pre:
                code = pre.find('code', class_='sourceCode python')
                if code:
                    # Extract lines with proper indentation
                    # Each line is in a span with id="cbX-Y"
                    line_spans = code.find_all('span', id=True)
                    
                    # Sort by line number
                    def get_line_num(span):
                        span_id = span.get('id', '')
                        match = re.search(r'cb\d+-(\d+)', span_id)
                        return int(match.group(1)) if match else 9999
                    
                    line_spans.sort(key=get_line_num)
                    
                    # Extract text and detect indentation from HTML structure
                    code_lines = []
                    for span in line_spans:
                        # Get text content, preserving structure
                        line_text = span.get_text()
                        
                        # Try to detect indentation from parent elements or HTML structure
                        # Count leading whitespace in the raw HTML or use the text structure
                        span_html = str(span)
                        
                        # Find the text content and detect indentation
                        # Simple approach: look at the actual text and infer indentation
                        # from control flow structures
                        cleaned_line = line_text.strip()
                        if cleaned_line:
                            code_lines.append(cleaned_line)
                    
                    if code_lines:
                        # Basic indentation fix based on control flow
                        fixed_lines = []
                        indent_level = 0
                        for i, line in enumerate(code_lines):
                            stripped = line.strip()
                            if not stripped or stripped.startswith('#'):
                                fixed_lines.append(stripped)
                                continue
                            
                            # Adjust indent based on previous line
                            if i > 0:
                                prev_line = code_lines[i-1].strip()
                                # If previous line ends with ':', increase indent
                                if prev_line.endswith(':') and not prev_line.startswith('#'):
                                    indent_level += 1
                                # If current line starts with elif/else/except, decrease indent
                                elif stripped.startswith(('elif ', 'else:', 'except ', 'finally:')):
                                    indent_level = max(0, indent_level - 1)
                            
                            # Apply indentation
                            indent = ' ' * (indent_level * 4)
                            fixed_lines.append(indent + stripped)
                            
                            # Adjust for next iteration
                            if stripped.endswith(':') and not any(k in stripped for k in ['#', '"""', "'''"]):
                                pass  # Will increase indent for next line
                            else:
                                # Check if we should decrease indent
                                if i < len(code_lines) - 1:
                                    next_line = code_lines[i+1].strip()
                                    if not next_line.startswith(('elif ', 'else:', 'except ', 'finally')):
                                        # Don't decrease if next line continues block
                                        if not prev_line.endswith(':'):
                                            indent_level = max(0, indent_level - 1)
                        
                        code_text = '\n'.join(fixed_lines)
                        qmd_content.append("```{python}")
                        qmd_content.append(code_text)
                        qmd_content.append("```")
                        qmd_content.append("")
    
    return '\n'.join(qmd_content)

if __name__ == "__main__":
    html_path = r"Predicting the uptake of e-cigarette use in Ghana, Mauritius, Seychelles, and Togo_ Findings from Global Youth Tobacco Survey (2023-2025).html"
    output_path = r"GTYS_ecig_analysis_2025.qmd"
    
    try:
        qmd_content = extract_code_from_html_proper(html_path)
        
        with open(output_path, 'w', encoding='utf-8') as f:
            f.write(qmd_content)
        
        print(f"Successfully created {output_path}")
        print(f"Total lines: {len(qmd_content.splitlines())}")
    except Exception as e:
        print(f"Error: {e}")
        import traceback
        traceback.print_exc()
````
