## Supplemental codes for "Methodological Guidance for Predictor Variable Selection for Adolescent Smoking Outcomes in Global Youth Tobacco Survey Using R and Python": extract_code_to_qmd.py

````
#!/usr/bin/env python3
"""
Extract Python code chunks from Quarto HTML output and convert to .qmd format
"""
import re
from bs4 import BeautifulSoup
from html import unescape

def extract_code_from_html(html_path):
    """Extract code chunks and sections from HTML file"""
    with open(html_path, 'r', encoding='utf-8') as f:
        html_content = f.read()
    
    soup = BeautifulSoup(html_content, 'html.parser')
    
    # Extract title
    title_tag = soup.find('title')
    title = title_tag.text.strip() if title_tag else "Analysis"
    
    # Extract all sections with code
    sections = soup.find_all('section', class_='level2')
    
    qmd_content = []
    
    # Add YAML header
    qmd_content.append("---")
    qmd_content.append(f"title: {title.replace(':', '-')}")
    qmd_content.append("format: html")
    qmd_content.append("code-fold: true")
    qmd_content.append("---")
    qmd_content.append("")
    
    for section in sections:
        # Extract section heading
        h2 = section.find('h2', class_='anchored')
        if h2:
            section_title = h2.get_text(strip=True)
            # Remove anchor link text
            section_title = re.sub(r'\s*#.*$', '', section_title)
            qmd_content.append(f"## {section_title}")
            qmd_content.append("")
        
        # Find all code cells in this section
        code_cells = section.find_all('div', class_='sourceCode')
        
        for cell in code_cells:
            pre = cell.find('pre')
            if pre:
                code = pre.find('code', class_='sourceCode python')
                if code:
                    # Extract lines - each line is in a span with id="cbX-Y"
                    # Sort spans by their id to maintain order
                    line_spans = code.find_all('span', id=True)
                    
                    # Sort by span id (e.g., "cb2-1", "cb2-2", etc.)
                    def get_line_number(span):
                        span_id = span.get('id', '')
                        match = re.search(r'cb\d+-(\d+)', span_id)
                        return int(match.group(1)) if match else 9999
                    
                    line_spans.sort(key=get_line_number)
                    
                    # Extract text from each line span
                    code_lines = []
                    for span in line_spans:
                        # Get all text from the span (including nested spans)
                        line_text = span.get_text()
                        # Clean up: remove extra whitespace but preserve structure
                        line_text = line_text.strip()
                        if line_text:  # Only add non-empty lines
                            code_lines.append(line_text)
                    
                    if code_lines:
                        # Join code lines
                        code_text = '\n'.join(code_lines)
                        # Clean up HTML entities
                        code_text = unescape(code_text)
                        
                        qmd_content.append("```{python}")
                        qmd_content.append(code_text)
                        qmd_content.append("```")
                        qmd_content.append("")
    
    return '\n'.join(qmd_content)

if __name__ == "__main__":
    html_path = r"Predicting the uptake of e-cigarette use in Ghana, Mauritius, Seychelles, and Togo_ Findings from Global Youth Tobacco Survey (2023-2025).html"
    output_path = r"GTYS_ecig_analysis_2025.qmd"
    
    try:
        qmd_content = extract_code_from_html(html_path)
        
        with open(output_path, 'w', encoding='utf-8') as f:
            f.write(qmd_content)
        
        print(f"Successfully created {output_path}")
        print(f"Total lines: {len(qmd_content.splitlines())}")
    except Exception as e:
        print(f"Error: {e}")
        import traceback
        traceback.print_exc()
````
