## Supplemental codes for "Methodological Guidance for Predictor Variable Selection for Adolescent Smoking Outcomes in Global Youth Tobacco Survey Using R and Python": fix_indentation.py

````
#!/usr/bin/env python3
"""
Fix indentation in Python code blocks within a Quarto markdown file.
This script attempts to fix indentation by detecting Python code blocks
and ensuring proper indentation based on control flow structures.
"""
import re

def fix_python_indentation(code):
    """Fix indentation in a Python code block"""
    lines = code.split('\n')
    fixed_lines = []
    indent_level = 0
    indent_size = 4
    
    for i, line in enumerate(lines):
        stripped = line.lstrip()
        if not stripped or stripped.startswith('#'):
            # Preserve empty lines and comments as-is
            fixed_lines.append(line)
            continue
        
        # Calculate expected indent based on previous line
        if i > 0:
            prev_line = lines[i-1].rstrip()
            prev_stripped = prev_line.lstrip()
            
            # If previous line ends with colon, next line should be indented
            if prev_line.endswith(':') and prev_stripped and not prev_stripped.startswith('#'):
                indent_level += 1
            # If current line starts a block that should close previous indent
            elif stripped and not any(keyword in prev_stripped for keyword in ['def ', 'class ', 'if ', 'elif ', 'else:', 'for ', 'while ', 'try:', 'except', 'with ']):
                # Check if we should dedent
                if stripped.startswith(('elif ', 'else:', 'except', 'finally')):
                    indent_level = max(0, indent_level - 1)
        
        # Apply indentation
        indent = ' ' * (indent_level * indent_size)
        fixed_lines.append(indent + stripped)
        
        # Adjust indent for next iteration if line ends with colon
        if stripped.endswith(':') and not any(keyword in stripped for keyword in ['#', '"""', "'''"]):
            pass  # Indent will be added at start of next iteration
        elif i < len(lines) - 1:
            next_stripped = lines[i+1].lstrip()
            if next_stripped and not next_stripped.startswith(('elif ', 'else:', 'except', 'finally')):
                # If next line doesn't continue the block, reduce indent
                if not (line.rstrip().endswith(':') or line.rstrip().endswith('\\')):
                    if not any(keyword in stripped for keyword in ['def ', 'class ', 'if ', 'for ', 'while ', 'try:', 'with ']):
                        indent_level = max(0, indent_level - 1)
    
    return '\n'.join(fixed_lines)

def fix_qmd_file(filepath):
    """Fix indentation in all Python code blocks in a .qmd file"""
    with open(filepath, 'r', encoding='utf-8') as f:
        content = f.read()
    
    # Pattern to match Python code blocks
    pattern = r'```\{python\}(.*?)```'
    
    def fix_code_block(match):
        code = match.group(1)
        # Remove leading/trailing newlines
        code = code.strip()
        # Fix indentation
        fixed_code = fix_python_indentation(code)
        return f'```{{python}}\n{fixed_code}\n```'
    
    # Replace all Python code blocks
    fixed_content = re.sub(pattern, fix_code_block, content, flags=re.DOTALL)
    
    # Write back
    with open(filepath, 'w', encoding='utf-8') as f:
        f.write(fixed_content)
    
    print(f"Fixed indentation in {filepath}")

if __name__ == "__main__":
    import sys
    qmd_file = r"C:\Users\LENOVO\Documents\GitHub\Global_Tobacco_Survey_Analysis_2025\data\gtys\ecig\GTYS_ecig_analysis_2025.qmd"
    fix_qmd_file(qmd_file)
````
