## Supplemental codes for "Methodological Guidance for Predictor Variable Selection for Adolescent Smoking Outcomes in Global Youth Tobacco Survey Using R and Python": GTYS_ecig_analysis_2025.qmd

````
---
title: Predicting the uptake of e-cigarette use in Ghana, Mauritius, Seychelles, and Togo- Findings from Global Youth Tobacco Survey (2015-2019)
format: html
code-fold: true
---

## Set working directory and load packages

## Load data

```{python}
import pandas as pd
import os

# Read Stata file with value labels preserved
# Since both .qmd and .dta files are in the same directory, use simple filename
# Quarto executes from the directory containing the .qmd file
data_path = 'gyts_ecig_analysis_clean.dta'

# Verify file exists, if not try alternative paths
if not os.path.exists(data_path):
    # Try relative to project root
    alt_path = os.path.join('..', '..', '..', 'data', 'gtys', 'ecig', 'gyts_ecig_analysis_clean.dta')
    if os.path.exists(alt_path):
        data_path = alt_path
    else:
        # Try absolute path
        data_path = r'C:\Users\LENOVO\Documents\GitHub\Global_Tobacco_Survey_Analysis_2025\data\gtys\ecig\gyts_ecig_analysis_clean.dta'

# Initialize label dictionaries
stata_value_labels = {}
stata_var_labels = {}

try:
    import pyreadstat
    df, meta = pyreadstat.read_dta(data_path, apply_value_formats=True)
    print("Read Stata file using pyreadstat (preserves labels)")
    
    # Extract value labels from metadata
    if meta is not None and hasattr(meta, 'value_labels'):
        stata_value_labels = meta.value_labels if meta.value_labels else {}
        print(f"\nFound value labels for {len(stata_value_labels)} variables")
        # Print some examples
        for var, labels in list(stata_value_labels.items())[:5]:
            print(f"  {var}: {len(labels)} value labels - {list(labels.items())[:3]}")
    
    # Extract variable labels
    if meta is not None and hasattr(meta, 'column_labels'):
        stata_var_labels = {col: label for col, label in zip(meta.column_names, meta.column_labels) if label and label.strip()}
        print(f"Found variable labels for {len(stata_var_labels)} variables")
        # Print some examples
        for var, label in list(stata_var_labels.items())[:5]:
            print(f"  {var}: {label}")
except ImportError:
    print("pyreadstat not available, using pandas (may not preserve all labels)")
    try:
        df = pd.read_stata(data_path, convert_categoricals=True)
        print("Read Stata file using pandas (categoricals converted)")
        
        # Try to read labels from pandas categorical columns
        for col in df.columns:
            if hasattr(df[col], 'cat') and hasattr(df[col].cat, 'categories'):
                # Has categorical labels - create mapping
                cats = df[col].cat.categories.tolist()
                codes = df[col].cat.codes
                # Map codes to categories (codes are 0-based, categories are the labels)
                label_map = {i: cat for i, cat in enumerate(cats)}
                stata_value_labels[col] = label_map
    except Exception as e:
        print(f"Error reading Stata file: {e}")
        df = pd.read_stata(data_path, convert_categoricals=False)
except Exception as e:
    print(f"Error reading Stata file with pyreadstat: {e}")
    df = pd.read_stata(data_path, convert_categoricals=True)

# Display basic info
print(f"\nData shape: {df.shape}")
print(f"\nColumn names:\n{df.columns.tolist()}")
if 'Country' in df.columns:
    country_count = df['Country'].nunique() if hasattr(df['Country'], 'nunique') else len(df['Country'].unique())
    print(f"\nCountries in data: {country_count}")
    if country_count <= 10:
        print(f"  Countries: {df['Country'].unique().tolist() if hasattr(df['Country'], 'unique') else 'N/A'}")
print(f"\nFirst few rows:")
df.head()
```

## Data preparation

```{python}
# Define variables
outcome = 'smk_ecig'
predictors = ['Age', 'Sex', 'Grade', 'smk_try', 'smkless_try', 'smk_public',
'smk_school', 'ban_closed_smoking', 'ban_public_smoking',
'taught_smking_dangers', 'advert_tv_smk', 'advert_radio_smk',
'tobaco_logo_product', 'friend_smk', 'smk_intend_12mo',
'heard_ecig', 'shisha', 'Country', 'Year', 'SES']
# Survey design variables (confirmed by user)
weight_var = 'FinalWgt'
psu_var = 'PSU'
strata_var = 'Stratum'
print(f"\nSurvey design variables:")
print(f"  Weight: {weight_var}")
print(f"  PSU: {psu_var}")
print(f"  Strata: {strata_var}")
# Variable name mapping for human-readable labels
# Use Stata variable labels if available, otherwise use our mapping
var_label_map = {
'Age': 'Age (years)',
'Sex': 'Sex',
'Grade': 'Grade',
'Country': 'Country',
'Year': 'Year',
'SES': 'Socioeconomic Status',
'smk_try': 'Ever tried cigarette smoking',
'smkless_try': 'Ever tried smokeless tobacco',
'smk_public': 'Exposed to smoking in public places',
'smk_school': 'Exposed to smoking at school',
'ban_closed_smoking': 'Ban on closed spaces smoking',
'ban_public_smoking': 'Ban on public places smoking',
'taught_smking_dangers': 'Taught smoking dangers',
'advert_tv_smk': 'Saw tobacco adverts on TV',
'advert_radio_smk': 'Heard tobacco adverts on radio',
'tobaco_logo_product': 'Owned tobacco logo product',
'friend_smk': 'Close friends smoke',
'smk_intend_12mo': 'Intend to smoke in next 12 months',
'heard_ecig': 'Heard of e-cigarettes',
'shisha': 'Ever used shisha',
'smk_ecig': 'Current e-cigarette use'
}
# Update with Stata variable labels if available (must be defined first)
# Note: stata_var_labels is defined in the data loading section above
try:
    if stata_var_labels and len(stata_var_labels) > 0:
        var_label_map.update(stata_var_labels)
        print(f"Updated variable labels from Stata file for {len(stata_var_labels)} variables")
except NameError:
    # stata_var_labels not yet defined, will be available after data loading
    pass

# Value label mapping - use Stata value labels if available, otherwise use defaults
def map_value_label(var_name, value):
    """Convert numeric codes to human-readable labels using Stata value labels if available"""
    if pd.isna(value) or value is None:
        return 'Missing'
    
    # Try to get value label from Stata labels first (global variable from data loading section)
    try:
        if 'stata_value_labels' in globals() and stata_value_labels and var_name in stata_value_labels:
            label_dict = stata_value_labels[var_name]
            # Try both numeric and string keys
            value_num = None
            try:
                value_num = float(value)
                value_int = int(value_num) if value_num == int(value_num) else None
            except:
                pass
            
            # Check for exact match in label dictionary
            if value_int is not None and value_int in label_dict:
                return str(label_dict[value_int])
            elif value in label_dict:
                return str(label_dict[value])
            elif str(value) in label_dict:
                return str(label_dict[str(value)])
    except NameError:
        # stata_value_labels not yet defined
        pass
    except Exception as e:
        pass
    
    # Convert to string for comparison, preserving numeric type
    value_str = str(value).strip().upper()
    value_num = None
    try:
        value_num = float(value)
        is_int = (value_num == int(value_num))
    except:
        pass
    
    # Fallback to default mappings for common binary variables
    if var_name in ['smk_try', 'smkless_try', 'smk_public', 'smk_school',
                    'ban_closed_smoking', 'ban_public_smoking', 'taught_smking_dangers',
                    'advert_tv_smk', 'advert_radio_smk', 'tobaco_logo_product',
                    'friend_smk', 'smk_intend_12mo', 'heard_ecig', 'shisha', 'smk_ecig']:
        if value_num == 1 or value_str in ['1', 'YES', 'Y', 'TRUE']:
            return 'Yes'
        elif value_num == 0 or value_str in ['0', 'NO', 'N', 'FALSE']:
            return 'No'
        else:
            return str(value)
    # Sex variable fallback
    elif var_name == 'Sex' or 'sex' in str(var_name).lower():
        if value_num == 1 or value_str in ['1', 'M', 'MALE', 'MALE']:
            return 'Male'
        elif value_num == 2 or value_str in ['2', 'F', 'FEMALE', 'FEMALE']:
            return 'Female'
        else:
            return str(value)
    # Age - display as imported from Stata (no recoding)
    elif var_name in ['Age', 'age']:
        try:
            # Display Age value as imported from Stata (no transformation)
            if is_int:
                return f"{int(value_num)}"
            else:
                return f"{value_num:.1f}"
        except:
            return str(value)
    # Grade - keep as is but ensure numeric format
    elif var_name in ['Grade', 'grade']:
        try:
            if is_int:
                return f"{int(value_num)}"
            else:
                return f"{value_num:.1f}"
        except:
            return str(value)
    # Country, Year, SES - return as is (should have labels from Stata)
    elif var_name in ['Country', 'country', 'Year', 'year', 'SES', 'ses']:
        return str(value)
    # Default - return as string, but try to format if numeric
    try:
        if is_int:
            return f"{int(value_num)}"
        else:
            return str(value)
    except:
        return str(value)

# Select relevant variables (check which ones exist)
available_vars = [v for v in [outcome] + predictors + [weight_var, psu_var, strata_var] if v in df.columns]
df_analysis = df[available_vars].copy()
print(f"\nVariables found in data: {len(available_vars)}/{len([outcome] + predictors + [weight_var, psu_var, strata_var])}")
missing_vars = set([outcome] + predictors + [weight_var, psu_var, strata_var]) - set(available_vars)
if missing_vars:
    print(f"Missing variables: {missing_vars}")

# Use Age as imported from Stata - treat as categorical/factor variable (no recoding)
if 'Age' in df_analysis.columns:
    # Ensure Age is treated as categorical/factor variable (use as imported from Stata)
    # Convert to numeric if needed, but preserve original values
    age_original = df_analysis['Age'].copy()
    df_analysis['Age'] = pd.to_numeric(df_analysis['Age'], errors='coerce')
    age_min = df_analysis['Age'].min() if df_analysis['Age'].notna().any() else None
    age_max = df_analysis['Age'].max() if df_analysis['Age'].notna().any() else None
    age_values = sorted(df_analysis['Age'].dropna().unique())
    print(f"\nAge used as imported from Stata - treated as categorical/factor variable:")
    if age_min is not None and age_max is not None:
        print(f"  Age range: {age_min:.0f}-{age_max:.0f} years")
        print(f"  Age categories: {[int(x) for x in age_values] if age_values else 'None'}")
        print(f"  Age will be treated as a categorical/factor variable with dummy variables created for each age category")
        print(f"  No recoding applied - using original Stata values")

# Check for missing values and calculate overall missing data proportion
print("\nMissing values per variable:")
missing_per_var = df_analysis[available_vars].isnull().sum()
print(missing_per_var)

# Calculate overall missing data proportion
total_cells = len(df_analysis) * len(available_vars)
missing_cells = missing_per_var.sum()
missing_proportion = (missing_cells / total_cells) * 100

print(f"\n" + "=" * 80)
print("MISSING DATA SUMMARY")
print("=" * 80)
print(f"Total rows: {len(df_analysis):,}")
print(f"Total variables: {len(available_vars)}")
print(f"Total data cells: {total_cells:,}")
print(f"Missing data cells: {missing_cells:,}")
print(f"Overall missing data proportion: {missing_proportion:.2f}%")
print(f"Rows with complete data (all variables): {df_analysis[available_vars].dropna().shape[0]:,} ({(df_analysis[available_vars].dropna().shape[0]/len(df_analysis)*100):.2f}%)")
print(f"Rows with any missing data: {len(df_analysis) - df_analysis[available_vars].dropna().shape[0]:,} ({((len(df_analysis) - df_analysis[available_vars].dropna().shape[0])/len(df_analysis)*100):.2f}%)")
print("=" * 80)

# Import numpy for random imputation
import numpy as np
np.random.seed(42)  # For reproducibility

def impute_missing_values(df, var_name, weight_var=None):
    """
    Impute missing values for a variable based on observed distribution.
    Uses weighted distribution if weight_var is provided, otherwise uses unweighted.
    """
    if var_name not in df.columns:
        return df
    
    missing_mask = df[var_name].isna()
    n_missing = missing_mask.sum()
    
    if n_missing == 0:
        return df
    
    # Get actual values and their observed distribution from non-missing data
    non_missing_data = df.loc[~missing_mask, var_name]
    
    # If weight_var is available, calculate weighted distribution
    if weight_var and weight_var in df.columns:
        # Calculate weighted distribution from non-missing data
        value_counts = df.loc[~missing_mask].groupby(var_name)[weight_var].sum()
        total_weight = value_counts.sum()
        probabilities = (value_counts / total_weight).values
        values = value_counts.index.tolist()
    else:
        # Use unweighted distribution from non-missing data
        value_counts = non_missing_data.value_counts(normalize=True)
        probabilities = value_counts.values
        values = value_counts.index.tolist()
    
    # Ensure probabilities sum to 1 (normalize if needed)
    probabilities = probabilities / probabilities.sum()
    
    # Randomly sample values from the distribution
    imputed_values = np.random.choice(values, size=n_missing, p=probabilities)
    
    # Assign imputed values
    df_imputed = df.copy()
    df_imputed.loc[missing_mask, var_name] = imputed_values
    
    print(f"  Imputed {n_missing} missing values for {var_name} based on observed distribution")
    
    return df_imputed

# Create imputed dataset for Tables 2 and 3
# Impute missing values based on observed distributions (weighted if weights available)
print("\n" + "=" * 80)
print("IMPUTING MISSING VALUES")
print("=" * 80)
print("Missing values will be randomly assigned based on the distribution of non-missing data")
print("Using weighted distributions where survey weights are available")

analysis_vars = [outcome] + predictors + [weight_var]
analysis_vars_available = [v for v in analysis_vars if v in df_analysis.columns]

# Start with the analysis dataset
df_complete = df_analysis[analysis_vars_available].copy()

# Impute missing values for each variable that has missing data (except outcome which should not be imputed)
for var in analysis_vars_available:
    if var == weight_var or var == outcome:  # Don't impute weights or outcome
        continue
    if var in df_complete.columns:
        missing_count = df_complete[var].isna().sum()
        if missing_count > 0:
            # Use observed distribution from data (weighted if available)
            df_complete = impute_missing_values(df_complete, var, weight_var)

# Verify imputation results
print(f"\n" + "=" * 80)
print("IMPUTATION SUMMARY")
print("=" * 80)
print(f"Variables included: {len(analysis_vars_available)}")
print(f"Total rows: {len(df_complete):,}")

# Check remaining missing values (should only be in outcome if any)
remaining_missing = df_complete[analysis_vars_available].isnull().sum()
if remaining_missing.sum() > 0:
    print(f"\nRemaining missing values after imputation:")
    for var, count in remaining_missing[remaining_missing > 0].items():
        if var != outcome:  # Outcome may have missing, which is acceptable
            print(f"  {var}: {count} ({count/len(df_complete)*100:.2f}%)")
else:
    print(f"No missing values remaining after imputation (excluding possible outcome missing)")

print("=" * 80)
```

## Table 1: Descriptive statistics (weighted and unweighted)

```{python}
from scipy import stats
from scipy.stats import chi2_contingency
import pandas as pd

def create_descriptive_table(df, predictors, outcome, weight_var, var_label_map, map_value_label):
    """Create descriptive statistics table with weighted and unweighted counts"""
    results = []
    # Process predictors including Country and Age (as categorical variable)
    for var in predictors:
        # Skip if variable doesn't exist
        if var not in df.columns:
            continue
        # Explicitly handle Age as categorical variable (using values as imported from Stata)
        if var == 'Age':
            # Ensure Age is treated as categorical (factor) - use values as imported from Stata
            age_var_data = df[var].copy()
            # Convert to numeric, then to integer to ensure proper categorical display
            age_var_data = pd.to_numeric(age_var_data, errors='coerce')
            age_var_data = age_var_data.round().astype('Int64')  # Nullable integer type
            # Get unweighted counts for each age category (using original Stata values)
            unweighted_counts = age_var_data.value_counts(dropna=False).sort_index()
            unweighted_total = len(age_var_data.dropna())
            # Get weighted counts and percentages by age category
            if weight_var in df.columns:
                # Use age_var_data for grouping to ensure categorical handling
                weighted_counts = df.groupby(age_var_data)[weight_var].sum()
                weighted_total = df[weight_var].sum()
                weighted_pct = (weighted_counts / weighted_total * 100).round(2)
            else:
                weighted_counts = unweighted_counts
                weighted_total = unweighted_total
                weighted_pct = (unweighted_counts / unweighted_total * 100).round(2)
            # Get human-readable variable name
            var_name = var_label_map.get(var, 'Age (years)')
            # Create rows for each age category (using original Stata values)
            for i, cat in enumerate(unweighted_counts.index):
                # Map value to human-readable label (using original Stata values)
                cat_label = map_value_label(var, cat)
                # Format counts and percentages
                unweighted_n = int(unweighted_counts.get(cat, 0))
                weighted_p = weighted_pct.get(cat, 0) if cat in weighted_pct.index else 0
                results.append({
                    'Variable': var_name if i == 0 else '',
                    'Category': cat_label,
                    'Unweighted N': f"{unweighted_n:,}" if unweighted_n > 0 else "0",
                    'Unweighted %': f"{(unweighted_n/unweighted_total*100):.2f}%" if unweighted_total > 0 else "0.00%",
                    'Weighted %': f"{weighted_p:.2f}%"
                })
        else:
            # Handle other variables as before
            # Get unweighted counts
            unweighted_counts = df[var].value_counts(dropna=False).sort_index()
            unweighted_total = len(df[var].dropna())
            # Get weighted counts and percentages
            if weight_var in df.columns:
                weighted_counts = df.groupby(var)[weight_var].sum()
                weighted_total = df[weight_var].sum()
                weighted_pct = (weighted_counts / weighted_total * 100).round(2)
            else:
                weighted_counts = unweighted_counts
                weighted_total = unweighted_total
                weighted_pct = (unweighted_counts / unweighted_total * 100).round(2)
            # Get human-readable variable name
            var_name = var_label_map.get(var, var.replace('_', ' ').title())
            # Create rows for each category
            for i, cat in enumerate(unweighted_counts.index):
                # Map value to human-readable label
                cat_label = map_value_label(var, cat)
                # Format counts and percentages
                unweighted_n = int(unweighted_counts.get(cat, 0))
                weighted_p = weighted_pct.get(cat, 0)
                results.append({
                    'Variable': var_name if i == 0 else '',
                    'Category': cat_label,
                    'Unweighted N': f"{unweighted_n:,}" if unweighted_n > 0 else "0",
                    'Unweighted %': f"{(unweighted_n/unweighted_total*100):.2f}%" if unweighted_total > 0 else "0.00%",
                    'Weighted %': f"{weighted_p:.2f}%"
                })
    # Add outcome variable summary
    if outcome in df.columns:
        outcome_counts = df[outcome].value_counts(dropna=False).sort_index()
        outcome_total = len(df[outcome].dropna())
        if weight_var in df.columns:
            outcome_weighted = df.groupby(outcome)[weight_var].sum()
            outcome_weighted_total = df[weight_var].sum()
            outcome_weighted_pct = (outcome_weighted / outcome_weighted_total * 100).round(2)
        else:
            outcome_weighted = outcome_counts
            outcome_weighted_total = outcome_total
            outcome_weighted_pct = (outcome_counts / outcome_total * 100).round(2)
        outcome_name = var_label_map.get(outcome, 'Current E-cigarette Use')
        for i, cat in enumerate(outcome_counts.index):
            cat_label = map_value_label(outcome, cat)
            unweighted_n = int(outcome_counts.get(cat, 0))
            weighted_p = outcome_weighted_pct.get(cat, 0)
            results.append({
                'Variable': outcome_name if i == 0 else '',
                'Category': cat_label,
                'Unweighted N': f"{unweighted_n:,}" if unweighted_n > 0 else "0",
                'Unweighted %': f"{(unweighted_n/outcome_total*100):.2f}%" if outcome_total > 0 else "0.00%",
                'Weighted %': f"{weighted_p:.2f}%"
            })
    return pd.DataFrame(results)

# Create descriptive table (Age explicitly included as categorical variable)
desc_table = create_descriptive_table(df_complete, predictors, outcome, weight_var, var_label_map, map_value_label)

# Verify Age is included in Table 1 as categorical variable
age_in_table1 = desc_table[desc_table['Variable'].str.contains('Age', case=False, na=False)]
if len(age_in_table1) > 0:
    table_age_categories = age_in_table1['Category'].unique()
    print(f"\n✓ Age is included in Table 1 as a categorical variable")
    print(f"  Number of age categories: {len(table_age_categories)}")
    print(f"  Age categories shown: {sorted([cat for cat in table_age_categories if cat and cat != 'Missing'])}")
else:
    print("\n⚠ WARNING: Age not found in Table 1 - checking predictors list and data")
    if 'Age' in predictors:
        print(f"  Age is in predictors list")
    if 'Age' in df_analysis.columns:
        print(f"  Age exists in data with {df_analysis['Age'].notna().sum()} non-missing values")
        print(f"  Age value counts: {df_analysis['Age'].value_counts().to_dict()}")
    else:
        print("  Age is NOT in data columns")

# Format Table 1 for publication with nice formatting
try:
    from tabulate import tabulate
    use_tabulate = True
except ImportError:
    use_tabulate = False

# Format the table nicely with proper headers
desc_table_formatted = desc_table.copy()
desc_table_formatted.columns = ['Variable', 'Category', 'Unweighted N', 'Unweighted %', 'Weighted %']

# Remove empty Variable rows (keep only first occurrence for each variable group)
for idx in range(1, len(desc_table_formatted)):
    if desc_table_formatted.iloc[idx]['Variable'] == desc_table_formatted.iloc[idx-1]['Variable']:
        desc_table_formatted.iloc[idx, desc_table_formatted.columns.get_loc('Variable')] = ''

print("\n" + "=" * 120)
print("TABLE 1: Descriptive Statistics of Study Participants")
print("Values are presented as unweighted numbers (N), unweighted percentages, and weighted percentages")
print("=" * 120)

if use_tabulate:
    # Use a nice grid format with proper alignment
    print(tabulate(desc_table_formatted,
                   headers='keys',
                   tablefmt='grid',
                   showindex=False,
                   stralign='left',
                   numalign='right',
                   floatfmt='.2f'))
else:
    # Use pandas built-in formatting as fallback with better formatting
    pd.set_option('display.max_columns', None)
    pd.set_option('display.width', None)
    pd.set_option('display.max_colwidth', 50)
    pd.set_option('display.precision', 2)
    print(desc_table_formatted.to_string(index=False))

# Also save as formatted markdown table for display
print("\n" + "-" * 120)
print("Table 1 formatted for document:")
print("-" * 120)
```

## Table 2: Weighted percentages of e-cigarette uptake

```{python}
def create_uptake_table(df, predictors, outcome, weight_var, var_label_map, map_value_label):
    """Create table showing weighted percentages of e-cigarette uptake by predictor categories"""
    # Calculate weighted percentages for each predictor category
    table_data = []
    # Include Country in the analysis
    for var in predictors:
        if var not in df.columns:
            continue
        # Get weighted percentages for outcome by predictor category
        if weight_var in df.columns:
            crosstab = pd.crosstab(df[var], df[outcome],
                                   values=df[weight_var],
                                   aggfunc='sum',
                                   normalize='index') * 100
        else:
            crosstab = pd.crosstab(df[var], df[outcome], normalize='index') * 100
        # Get percentage for outcome=1 or 'Yes' (e-cigarette use)
        if 1 in crosstab.columns:
            pct = crosstab[1]
        elif True in crosstab.columns:
            pct = crosstab[True]
        elif 'Yes' in crosstab.columns:
            pct = crosstab['Yes']
        else:
            continue
        # Calculate confidence intervals (using normal approximation)
        if weight_var in df.columns:
            n_weighted = df.groupby(var)[weight_var].sum()
        else:
            n_weighted = df[var].value_counts()
        se = np.sqrt((pct * (100 - pct)) / n_weighted)
        ci_lower = pct - 1.96 * se
        ci_upper = pct + 1.96 * se
        # Get human-readable variable name
        var_name = var_label_map.get(var, var.replace('_', ' ').title())
        for cat in pct.index:
            if pd.notna(cat) and pd.notna(pct[cat]):
                cat_label = map_value_label(var, cat)
                table_data.append({
                    'Variable': var_name,
                    'Category': cat_label,
                    'Weighted %': pct[cat],
                    '95% CI Lower': max(0, ci_lower[cat]),
                    '95% CI Upper': min(100, ci_upper[cat])
                })
    return pd.DataFrame(table_data)

# Create table data using complete cases (exclude missing data)
# Note: Age will be included as a categorical variable with each age category shown
uptake_table_data = create_uptake_table(df_complete, predictors, outcome, weight_var, var_label_map, map_value_label)

# Verify Age is included in Table 2
age_in_table2 = uptake_table_data[uptake_table_data['Variable'].str.contains('Age', case=False, na=False)]
if len(age_in_table2) > 0:
    print(f"\n✓ Age is included in Table 2 with {len(age_in_table2)} age categories")
else:
    print("\n⚠ WARNING: Age not found in Table 2 - checking predictors list")
    if 'Age' in predictors and 'Age' in df_complete.columns:
        print("  Age is in predictors list and data - checking for valid values")
        print(f"  Age values in complete cases: {df_complete['Age'].value_counts().to_dict()}")

# Format Table 2 with nice formatting
uptake_table_formatted = uptake_table_data.copy()

# Create a formatted CI column (e.g., "4.50 (3.20-5.80)")
uptake_table_formatted['Weighted % (95% CI)'] = uptake_table_formatted.apply(
    lambda row: f"{row['Weighted %']:.2f} ({row['95% CI Lower']:.2f}-{row['95% CI Upper']:.2f})",
    axis=1
)

# Select columns for display
uptake_table_display = uptake_table_formatted[['Variable', 'Category', 'Weighted % (95% CI)']].copy()

# Remove empty Variable rows (keep only first occurrence for each variable group)
for idx in range(1, len(uptake_table_display)):
    if uptake_table_display.iloc[idx]['Variable'] == uptake_table_display.iloc[idx-1]['Variable']:
        uptake_table_display.iloc[idx, uptake_table_display.columns.get_loc('Variable')] = ''

print("\n" + "=" * 120)
print("TABLE 2: Weighted Percentages of Current E-cigarette Use by Predictor Categories")
print("Values are weighted percentages with 95% confidence intervals (CI)")
print(f"Analysis based on imputed dataset (n={len(df_complete):,})")
print("=" * 120)

if use_tabulate:
    print(tabulate(uptake_table_display,
                   headers='keys',
                   tablefmt='grid',
                   showindex=False,
                   stralign='left',
                   numalign='right'))
else:
    pd.set_option('display.max_colwidth', 60)
    print(uptake_table_display.to_string(index=False))
```

## Weighted Logistic Regression with Stepwise Selection

```{python}
import statsmodels.api as sm
from statsmodels.stats.weightstats import DescrStatsW
from statsmodels.discrete.discrete_model import Logit
import itertools
# Prepare data for modeling
# Note: Country, Year, and SES are included in descriptive tables but excluded from modeling predictors
# (they can be added back if needed, but typically Country/Year are used for stratification)
excluded_from_modeling = ['Country', 'Year', 'SES']
modeling_predictors = [p for p in predictors if p not in excluded_from_modeling]
print(f"\nPredictors for modeling (excluding {excluded_from_modeling}): {len(modeling_predictors)} variables")
print(f"Predictors for descriptive tables: {len(predictors)} variables (including Country, Year, SES)")
# Check which modeling predictors actually exist in the data
available_modeling = [p for p in modeling_predictors if p in df_analysis.columns]
missing_modeling = [p for p in modeling_predictors if p not in df_analysis.columns]
if missing_modeling:
    print(f"WARNING: Some modeling predictors not found in data: {missing_modeling}")
modeling_predictors = available_modeling
if outcome not in df_analysis.columns:
    raise ValueError(f"Outcome variable '{outcome}' not found in data!")
if weight_var not in df_analysis.columns:
    raise ValueError(f"Weight variable '{weight_var}' not found in data!")
# Select variables for modeling
modeling_vars = [outcome] + modeling_predictors + [weight_var]
available_modeling_vars = [v for v in modeling_vars if v in df_analysis.columns]
df_model = df_analysis[available_modeling_vars].copy()
print(f"\nVariables selected for modeling: {len(available_modeling_vars)}/{len(modeling_vars)}")
print(f"Sample size for modeling (complete cases, missing data excluded): {len(df_model):,}")
print(f"Note: This dataset already excludes missing data from the complete cases analysis")
# Check missing values before dropping
missing_counts = df_model.isnull().sum()
print(f"\nMissing values per variable before dropping:")
total_missing = 0
for var, count in missing_counts.items():
    if count > 0:
        print(f"  {var}: {count} ({count/len(df_model)*100:.1f}%)")
    total_missing += count
# Since df_model is already from df_complete (complete cases), no need to drop missing values again
# But verify that we have the required variables
critical_vars = [outcome, weight_var]
critical_vars = [v for v in critical_vars if v in df_model.columns]
print(f"\nVariables in df_model: {df_model.columns.tolist()}")
print(f"Critical variables to check: {[outcome, weight_var]}")
print(f"Critical variables found: {critical_vars}")
if len(critical_vars) < 2:
    missing_critical = [v for v in [outcome, weight_var] if v not in df_model.columns]
    raise ValueError(f"Critical variables missing from data: {missing_critical}")
# df_model already has complete cases, so no missing values to remove
print(f"\nUsing complete cases dataset (missing data already excluded)")
print(f"Final sample size for modeling: {len(df_model):,}")
# For Age and Sex (a priori variables), fill missing values with mode/median or create missing indicator
# This ensures we don't lose all data
if 'Age' in df_model.columns:
    age_missing = df_model['Age'].isna().sum()
    if age_missing > 0:
        # Check if Age has any non-missing values
        age_valid = df_model['Age'].dropna()
        if len(age_valid) > 0:
            age_median = age_valid.median()
            df_model['Age'] = df_model['Age'].fillna(age_median)
            print(f"  Filled {age_missing} missing Age values with median: {age_median:.1f}")
        else:
            # All Age values are missing - use a default value or drop Age from a priori
            print(f"  WARNING: All Age values are missing! Age cannot be used as a priori variable.")
            # We'll handle this later when creating df_model_encoded
if 'Sex' in df_model.columns:
    sex_missing = df_model['Sex'].isna().sum()
    if sex_missing > 0:
        # Fill with most common value
        sex_mode = df_model['Sex'].mode()[0] if len(df_model['Sex'].mode()) > 0 else df_model['Sex'].iloc[0]
        df_model['Sex'] = df_model['Sex'].fillna(sex_mode)
        print(f"  Filled {sex_missing} missing Sex values with mode: {sex_mode}")
# For other predictors with missing values, we'll handle them during encoding (convert to dummy variables will handle missing)
print(f"\nRemaining missing values in modeling variables:")
remaining_missing = df_model[modeling_predictors].isnull().sum()
for var, count in remaining_missing.items():
    if count > 0:
        print(f"  {var}: {count} ({count/len(df_model)*100:.1f}%) - will be handled during encoding")
# Separate a priori variables (Age, Sex, and Grade) from other predictors
# Country is excluded from modeling
other_predictors = [p for p in modeling_predictors if p not in ['Age', 'Sex', 'Grade']]
# Start with a priori variables
apriori_vars = []
df_model_encoded = pd.DataFrame()

# Add Age as a priori (treat as categorical/factor using values as imported from Stata)
if 'Age' in df_model.columns:
    age_data = df_model['Age'].copy()
    # Check if Age has any valid values
    if age_data.notna().sum() > 0:
        # Ensure Age is numeric first (as imported from Stata)
        age_data = pd.to_numeric(age_data, errors='coerce')
        # Fill any remaining missing values with mode (most common age category)
        if age_data.isna().sum() > 0:
            age_mode = age_data.mode()[0] if len(age_data.mode()) > 0 else age_data.median()
            age_data = age_data.fillna(age_mode)
            print(f"  Filled missing Age values with mode: {age_mode:.0f}")
        # Convert Age to categorical (factor) - use values as imported from Stata
        # Round to nearest integer and convert to int for categorical encoding
        age_data_int = age_data.round().astype(int)
        # Create dummy variables from categorical Age (factor variable)
        # Use drop_first=True to have a reference category
        age_dummies = pd.get_dummies(age_data_int, prefix='Age', drop_first=True, dummy_na=False)
        # Rename columns to ensure clean names (e.g., Age_11, Age_12, etc. using original Stata values)
        age_dummies.columns = [f'Age_{int(col.split("_")[-1])}' if '_' in col else col for col in age_dummies.columns]
        df_model_encoded = pd.concat([df_model_encoded, age_dummies], axis=1)
        apriori_vars.extend(age_dummies.columns.tolist())
        age_values = sorted(age_data_int.dropna().unique())
        age_values_display = sorted([int(x) for x in age_values])  # Use original Stata values
        ref_age = min(age_values_display)  # Reference category (dropped for baseline)
        print(f"Added Age as a priori variable (treated as categorical/factor using Stata values)")
        print(f"  Age categories (as imported from Stata): {age_values_display} years")
        print(f"  Age dummy variables created: {age_dummies.columns.tolist()}")
        print(f"  Reference category: Age_{ref_age} ({ref_age} years) - dropped for baseline")
    else:
        print(f"  WARNING: Age has no valid values, skipping as a priori variable")

# Handle Sex as a priori (create dummy variables, drop_first to have reference category)
if 'Sex' in df_model.columns:
    sex_dummies = pd.get_dummies(df_model['Sex'], prefix='Sex', drop_first=True, dummy_na=False)
    df_model_encoded = pd.concat([df_model_encoded, sex_dummies], axis=1)
    apriori_vars.extend(sex_dummies.columns.tolist())
    print(f"Added Sex as a priori variable(s): {sex_dummies.columns.tolist()}")

# Handle Grade as a priori (create dummy variables, drop_first to have reference category)
if 'Grade' in df_model.columns:
    grade_dummies = pd.get_dummies(df_model['Grade'], prefix='Grade', drop_first=True, dummy_na=False)
    df_model_encoded = pd.concat([df_model_encoded, grade_dummies], axis=1)
    apriori_vars.extend(grade_dummies.columns.tolist())
    print(f"Added Grade as a priori variable(s): {grade_dummies.columns.tolist()}")

# Convert remaining categorical variables to dummy variables (excluding Age, Sex, and Grade)
# Handle missing values by including them as a separate category
if len(other_predictors) > 0:
    other_vars_data = df_model[other_predictors].copy()
# Fill missing values temporarily before encoding (pandas get_dummies doesn't handle NaN well)
# We'll create dummies and handle missing separately if needed
other_vars_encoded = pd.get_dummies(other_vars_data, drop_first=True, dummy_na=False)
df_model_encoded = pd.concat([df_model_encoded, other_vars_encoded], axis=1)
print(f"Encoded {len(other_predictors)} other predictor variables into {other_vars_encoded.shape[1]} dummy variables")
# Note: Variables with missing values will have all dummy columns = 0 for those rows
# This is acceptable for logistic regression
# Prepare outcome - convert to numeric 0/1 if it's categorical/string
y = df_model[outcome].copy()
# Check if outcome is categorical/string and convert to numeric
if y.dtype == 'object' or y.dtype.name == 'category':
    # Map common Yes/No values to 1/0
    y_mapped = y.astype(str).str.lower().str.strip()
    y_numeric = y_mapped.map({
        '1': 1, 'yes': 1, 'y': 1, 'true': 1,
        '0': 0, 'no': 0, 'n': 0, 'false': 0
    })
    # If mapping worked, use it; otherwise try direct conversion
    if y_numeric.isna().sum() < len(y_numeric) * 0.5:  # At least 50% mapped successfully
        y = y_numeric.fillna(y.astype(str).str.extract(r'(\d+)')[0].astype(float).fillna(0).astype(int))
    else:
        # Try to convert directly to numeric
        y = pd.to_numeric(y, errors='coerce').fillna(0)
    print(f"Converted outcome from {df_model[outcome].dtype} to numeric. Unique values: {y.unique()}")
else:
    # Already numeric, just ensure it's 0/1
    y = pd.to_numeric(y, errors='coerce').fillna(0)
    # Ensure binary 0/1
    y = y.astype(int)
    y = y.clip(0, 1)  # Ensure values are 0 or 1
print(f"Outcome summary: {y.value_counts().to_dict()}")
weights = df_model[weight_var]
# Combine all predictor names
all_predictors = df_model_encoded.columns.tolist()
print(f"\n" + "=" * 80)
print("MODEL PREPARATION SUMMARY")
print("=" * 80)
print(f"A priori variables (always included): {apriori_vars}")
print(f"Total candidate predictors for stepwise selection: {len(all_predictors)}")
print(f"  - A priori variables: {len(apriori_vars)}")
print(f"  - Other candidate variables: {len(all_predictors) - len(apriori_vars)}")
print(f"Outcome variable: {outcome}")
print(f"Sample size: {len(df_model)}")
print("=" * 80)
```

## Stepwise selection function

```{python}
def stepwise_selection(X, y, weights, apriori_vars, max_iter=50):
    """
    Perform stepwise selection for weighted logistic regression
    Age, Sex, and Grade are always included (a priori) and cannot be removed
    """
    from statsmodels.genmod.generalized_linear_model import GLM
    from statsmodels.genmod import families
    # Normalize weights to sum to sample size (for better numerical stability)
    weights_norm = weights * (len(weights) / weights.sum())
    # Ensure a priori variables are in the dataset
    selected = [var for var in apriori_vars if var in X.columns]
    remaining = [var for var in X.columns if var not in selected]
    print(f"\nSTEPWISE SELECTION PROCESS")
    print(f"Starting with a priori variables (always included): {selected}")
    print(f"Remaining candidate variables for selection: {len(remaining)}")
    # Fit initial model with only a priori variables to get baseline AIC
    if len(selected) > 0:
        try:
            X_initial = X[selected].copy()
            # Ensure numeric types - convert each column properly
            X_initial_clean = pd.DataFrame()
            for col in X_initial.columns:
                col_data = pd.to_numeric(X_initial[col], errors='coerce')
                if col_data.isna().all():
                    # If all NaN, fill with 0
                    X_initial_clean[col] = 0
                else:
                    X_initial_clean[col] = col_data.fillna(col_data.mean() if col_data.notna().any() else 0)
            # Convert to numpy array
            X_initial_array = X_initial_clean.values.astype(float)
            # Add constant
            X_initial_array = sm.add_constant(X_initial_array, has_constant='add')
            # Ensure y and weights are numeric
            y_initial = pd.to_numeric(y, errors='coerce')
            y_initial = y_initial.fillna(y_initial.mode()[0] if len(y_initial.mode()) > 0 else 0).astype(int).values
            weights_initial = pd.to_numeric(weights_norm, errors='coerce')
            weights_initial = weights_initial.fillna(weights_initial.mean() if weights_initial.notna().any() else 1.0).values
            # Check for valid data
            if len(X_initial_array) == 0 or len(y_initial) == 0:
                raise ValueError("Empty arrays after data preparation")
            model_initial = GLM(y_initial, X_initial_array, family=families.Binomial(link=families.links.Logit()),
                                freq_weights=weights_initial)
            result_initial = model_initial.fit(disp=0, method='lbfgs', maxiter=500)
            best_aic = result_initial.aic
            print(f"Initial model (a priori only) AIC: {best_aic:.2f}")
        except Exception as e:
            print(f"Error fitting initial model: {e}")
            print("WARNING: Could not fit initial model with a priori variables!")
            best_aic = np.inf
    else:
        print("WARNING: No a priori variables found in dataset!")
        best_aic = np.inf
    iteration = 0
    # Forward stepwise selection
    while iteration < max_iter and len(remaining) > 0:
        iteration += 1
        scores = []
        # Try adding each remaining variable
        for var in remaining:
            try:
                vars_to_test = selected + [var]
                X_subset = X[vars_to_test].copy()
                # Ensure numeric types
                for col in X_subset.columns:
                    if X_subset[col].dtype == 'object':
                        X_subset[col] = pd.to_numeric(X_subset[col], errors='coerce').fillna(0)
                X_subset_array = X_subset.values.astype(float)
                X_subset_array = sm.add_constant(X_subset_array, has_constant='add')
                y_subset = pd.to_numeric(y, errors='coerce').fillna(0).astype(int).values
                weights_subset = pd.to_numeric(weights_norm, errors='coerce').fillna(1.0).values
                # Fit weighted logistic regression using GLM
                model = GLM(y_subset, X_subset_array, family=families.Binomial(link=families.links.Logit()),
                            freq_weights=weights_subset)
                result = model.fit(disp=0, method='lbfgs', maxiter=500)
                # Get AIC
                aic = result.aic
                scores.append((var, aic))
            except Exception as e:
                continue
        if not scores:
            break
        # Find best variable to add (lowest AIC)
        scores.sort(key=lambda x: x[1])
        best_var, best_new_aic = scores[0]
        # If AIC improved, add the variable
        if best_new_aic < best_aic:
            selected.append(best_var)
            remaining.remove(best_var)
            aic_change = best_aic - best_new_aic  # Positive change indicates improvement
            best_aic = best_new_aic
            print(f"Iteration {iteration}: Added '{best_var}', AIC = {best_new_aic:.2f} (ΔAIC = {aic_change:.2f})")
        else:
            print(f"\nStopping: No improvement in AIC. Best AIC remains {best_aic:.2f}")
            break
    print(f"\nStepwise selection complete!")
    print(f"Final model includes {len(selected)} variables: {len(apriori_vars)} a priori + {len(selected) - len(apriori_vars)} selected")
    return selected, best_aic
# Perform stepwise selection
print("\nPerforming stepwise selection...")
print("=" * 80)
selected_vars, final_aic = stepwise_selection(df_model_encoded, y, weights, apriori_vars)
print(f"\nFinal selected variables: {selected_vars}")
print(f"Final AIC: {final_aic:.2f}")
```

## Fit final weighted logistic regression model

```{python}
# Fit the final model with selected variables
if len(selected_vars) == 0:
    print("ERROR: No variables selected for final model!")
    raise ValueError("No variables selected for final model. At least a priori variables should be included.")
# Check which selected variables actually exist in df_model_encoded
available_selected = [var for var in selected_vars if var in df_model_encoded.columns]
missing_selected = [var for var in selected_vars if var not in df_model_encoded.columns]
if len(missing_selected) > 0:
    print(f"WARNING: Some selected variables not found in df_model_encoded: {missing_selected}")
    print(f"Available columns in df_model_encoded: {df_model_encoded.columns.tolist()}")
if len(available_selected) == 0:
    raise ValueError(f"No selected variables found in df_model_encoded. Selected: {selected_vars}, Available: {df_model_encoded.columns.tolist()}")
X_final = df_model_encoded[available_selected].copy()
print(f"\nFitting final model with {len(available_selected)} variables: {available_selected}")
# Ensure all columns are numeric - convert to numpy array with proper dtypes
# Convert each column to numeric, filling NaN with 0 or mean
X_final_clean = pd.DataFrame(index=X_final.index)  # Preserve index
for col in X_final.columns:
    col_data = pd.to_numeric(X_final[col], errors='coerce')
    if col_data.isna().all():
        # If all NaN, fill with 0
        X_final_clean[col] = 0
    else:
        # Fill NaN with mean if available, otherwise 0
        fill_value = col_data.mean() if col_data.notna().any() else 0
        X_final_clean[col] = col_data.fillna(fill_value)
# Check for empty DataFrame before conversion
if X_final_clean.empty or len(X_final_clean) == 0:
    raise ValueError(f"X_final_clean is empty after data preparation. Selected vars: {available_selected}, Shape: {X_final.shape}")
print(f"X_final_clean shape: {X_final_clean.shape}")
print(f"X_final_clean columns: {X_final_clean.columns.tolist()}")
# Convert to numpy array to avoid object dtype issues
X_final_array = X_final_clean.values.astype(float)
# Check for empty array after conversion
if X_final_array.size == 0 or len(X_final_array) == 0:
    raise ValueError(f"X_final_array is empty after conversion. Shape: {X_final_array.shape}, Selected vars: {selected_vars}")
# Ensure y is numeric and integer (0/1)
y_final = pd.to_numeric(y, errors='coerce')
if y_final.isna().all():
    raise ValueError("All y values are NaN")
y_final = y_final.fillna(y_final.mode()[0] if len(y_final.mode()) > 0 else 0).astype(int).values
# Ensure y and X have same length
if len(y_final) != len(X_final_array):
    raise ValueError(f"Length mismatch: y has {len(y_final)} rows, X has {len(X_final_array)} rows")
# Add intercept to numpy array
X_final_array = sm.add_constant(X_final_array, has_constant='add')
# Normalize weights to sum to sample size (for better numerical stability)
weights_clean = pd.to_numeric(weights, errors='coerce')
if weights_clean.isna().all():
    weights_clean = pd.Series([1.0] * len(weights))
else:
    weights_clean = weights_clean.fillna(weights_clean.mean() if weights_clean.notna().any() else 1.0)
weights_clean_values = weights_clean.values
if weights_clean_values.sum() > 0:
    weights_normalized = weights_clean_values * (len(weights_clean_values) / weights_clean_values.sum())
else:
    weights_normalized = np.ones(len(weights_clean_values))
# Ensure weights are positive
weights_normalized = np.maximum(weights_normalized, 0.001)
# Fit weighted logistic regression using GLM with binomial family and logit link
# This approach properly handles survey weights
from statsmodels.genmod.generalized_linear_model import GLM
from statsmodels.genmod import families
# Save variable names before converting to numpy array (for result interpretation)
variable_names = ['const'] + available_selected
# Fit GLM with binomial family and logit link (equivalent to logistic regression)
# Use numpy arrays to avoid dtype issues
model_final = GLM(y_final, X_final_array, family=families.Binomial(link=families.links.Logit()),
freq_weights=weights_normalized)
result_final = model_final.fit(method='lbfgs', maxiter=1000)
print("\n" + "=" * 80)
print("FINAL WEIGHTED LOGISTIC REGRESSION MODEL")
print("=" * 80)
print(result_final.summary())
# Calculate model fit statistics
print(f"\nModel Fit Statistics:")
print(f"AIC: {result_final.aic:.2f}")
print(f"BIC: {result_final.bic:.2f}")
print(f"Log-likelihood: {result_final.llf:.2f}")
```

## Table 3: Final model results (odds ratios)

```{python}
# Extract coefficients and confidence intervals
# Since we used numpy arrays, we need to use the saved variable names
# Ensure all arrays have the same length
n_params = len(result_final.params)
var_names_clean = variable_names[:n_params] if len(variable_names) >= n_params else variable_names + [f'var_{i}' for i in range(len(variable_names), n_params)]
# Get confidence intervals properly (conf_int returns a 2D array)
conf_int = result_final.conf_int()
ci_lower = conf_int.iloc[:, 0].values if hasattr(conf_int, 'iloc') else conf_int[:, 0]
ci_upper = conf_int.iloc[:, 1].values if hasattr(conf_int, 'iloc') else conf_int[:, 1]
coef_df = pd.DataFrame({
'Variable': var_names_clean,
'Coefficient': result_final.params,
'OR': np.exp(result_final.params),
'CI_Lower': np.exp(ci_lower),
'CI_Upper': np.exp(ci_upper),
'P_value': result_final.pvalues
})
# Remove intercept for table display
coef_df_table = coef_df[coef_df['Variable'] != 'const'].copy()
# Verify a priori variables (Age, Sex, Grade) are included in Table 3
age_vars_in_table3 = coef_df_table[coef_df_table['Variable'].str.startswith('Age_', na=False)]
sex_vars_in_table3 = coef_df_table[coef_df_table['Variable'].str.startswith('Sex_', na=False)]
grade_vars_in_table3 = coef_df_table[coef_df_table['Variable'].str.startswith('Grade_', na=False)]

print(f"\n✓ A priori variables in Table 3:")
if len(age_vars_in_table3) > 0:
    print(f"  Age dummy variables: {age_vars_in_table3['Variable'].tolist()}")
if len(sex_vars_in_table3) > 0:
    print(f"  Sex dummy variables: {sex_vars_in_table3['Variable'].tolist()}")
if len(grade_vars_in_table3) > 0:
    print(f"  Grade dummy variables: {grade_vars_in_table3['Variable'].tolist()}")

    if len(age_vars_in_table3) == 0 and len(sex_vars_in_table3) == 0 and len(grade_vars_in_table3) == 0:
        print("\n⚠ WARNING: No a priori variables found in Table 3")
    print(f"  All variables in model: {coef_df_table['Variable'].tolist()}")
print(f"  Expected a priori variables: {apriori_vars}")

# Sort by variable type: A priori variables first (Age, Sex, Grade), then others by OR
# Put Age, Sex, and Grade variables first to emphasize they are a priori
apriori_mask = (coef_df_table['Variable'].str.startswith('Age_', na=False) |
coef_df_table['Variable'].str.startswith('Sex_', na=False) |
coef_df_table['Variable'].str.startswith('Grade_', na=False))

# Within a priori variables, sort: Age first, then Sex, then Grade
age_mask = coef_df_table['Variable'].str.startswith('Age_', na=False)
sex_mask = coef_df_table['Variable'].str.startswith('Sex_', na=False)
grade_mask = coef_df_table['Variable'].str.startswith('Grade_', na=False)

coef_df_table_sorted = pd.concat([
coef_df_table[age_mask].sort_values('Variable'),    # Age variables sorted by name
coef_df_table[sex_mask].sort_values('Variable'),    # Sex variables sorted by name
coef_df_table[grade_mask].sort_values('Variable'),  # Grade variables sorted by name
coef_df_table[~apriori_mask].sort_values('OR', ascending=False)  # Other variables sorted by OR
], ignore_index=True)

coef_df_table = coef_df_table_sorted
# Create a formatted table with human-readable variable names
coef_table = coef_df_table.copy()
# Ensure Age dummy variables are properly formatted for display
def format_variable_name(var_name):
    """Format variable name with human-readable labels"""
    # Handle dummy variables (e.g., "Sex_Male", "Age_11", "smk_try_1")
    if '_' in var_name:
        base_var = var_name.split('_')[0]
        var_display = var_label_map.get(base_var, base_var.replace('_', ' ').title())
        # Try to identify the category
        if len(var_name.split('_')) > 1:
            category_part = '_'.join(var_name.split('_')[1:])
            # For Age dummy variables (e.g., Age_21, Age_22), display the age value as imported from Stata
            if base_var == 'Age' and category_part.isdigit():
                # Use Age value as imported from Stata (no recoding)
                age_val = int(category_part)
                var_display = f"{var_display} ({age_val} years)"
            else:
                # Try to map the category for other variables
                try:
                    cat_val = int(category_part) if category_part.isdigit() else category_part
                    category_label = map_value_label(base_var, cat_val)
                    var_display = f"{var_display} ({category_label})"
                except:
                    var_display = f"{var_display} ({category_part})"
    else:
        var_display = var_label_map.get(var_name, var_name.replace('_', ' ').title())
    return var_display
coef_table['Variable_Display'] = coef_table['Variable'].apply(format_variable_name)

# Mark a priori variables
coef_table['Is_Apriori'] = (coef_table['Variable'].str.startswith('Age_', na=False) |
coef_table['Variable'].str.startswith('Sex_', na=False) |
coef_table['Variable'].str.startswith('Grade_', na=False))

# Add asterisk to a priori variables for display
coef_table['Variable_Display'] = coef_table.apply(
lambda row: f"{row['Variable_Display']} *" if row['Is_Apriori'] else row['Variable_Display'],
axis=1
)

# Format the table columns with combined CI
result_table = pd.DataFrame({
'Variable': coef_table['Variable_Display'],
'Adjusted OR (95% CI)': coef_table.apply(
lambda row: f"{row['OR']:.2f} ({row['CI_Lower']:.2f}-{row['CI_Upper']:.2f})",
axis=1
),
'P-value': coef_table['P_value'].apply(
lambda x: "< 0.001" if x < 0.001 else f"{x:.3f}" if x < 0.01 else f"{x:.2f}"
)
})

print("\n" + "=" * 120)
print("TABLE 3: Adjusted Odds Ratios and 95% Confidence Intervals")
print("Weighted Logistic Regression Model for Current E-cigarette Use")
print("Age, Sex, and Grade were included as a priori variables (marked with *).")
print("Model adjusted using stepwise selection (AIC criterion).")
print(f"Analysis based on imputed dataset (n={len(df_complete):,})")
print("=" * 120)

if use_tabulate:
    print(tabulate(result_table, 
headers='keys', 
tablefmt='grid', 
showindex=False, 
stralign='left',
numalign='right'))
else:
    pd.set_option('display.max_colwidth', 80)
print(result_table.to_string(index=False))

print("\n* A priori variables: Age, Sex, and Grade were always included in the model.")
print("  Reference categories are the first level of each categorical variable (dropped for baseline).")
```

## Overall Predicted E-cigarette Uptake by Age

```{python}
import matplotlib.pyplot as plt
import seaborn as sns
from scipy.stats import norm

# Set plotting style for publication-quality figures
sns.set_style("whitegrid")
plt.rcParams['figure.figsize'] = (12, 8)
plt.rcParams['font.size'] = 12
plt.rcParams['axes.labelsize'] = 13
plt.rcParams['axes.titlesize'] = 14
plt.rcParams['xtick.labelsize'] = 11
plt.rcParams['ytick.labelsize'] = 11
plt.rcParams['legend.fontsize'] = 12

# Get age categories from the data
age_categories = sorted([int(x) for x in df_complete['Age'].dropna().unique() if pd.notna(x)])

def calculate_overall_predicted_probabilities(model_result, df_model_encoded, selected_vars, age_categories):
    """
    Calculate overall predicted probabilities for different age categories.
    Uses reference levels for all other variables (Sex=Male, Heard_ECig=No, etc.)
    
    Parameters:
    -----------
    model_result : statsmodels GLMResults
        The fitted model results
    df_model_encoded : pandas.DataFrame
        The encoded dataframe used for modeling
    selected_vars : list
        List of selected variable names (excluding intercept)
    age_categories : list
        List of age categories (e.g., [11, 12, 13, 14, 15, 16, 17])
    
    Returns:
    --------
    predictions_df : pandas.DataFrame
        DataFrame with age and predicted probability
    """
    predictions = []

    # Get reference age (minimum age, which is dropped for baseline)
    ref_age = min(age_categories)

    for age in age_categories:
        # Create a template row with all variables set to reference (0)
        X_template = pd.DataFrame(index=[0], columns=selected_vars)
        X_template = X_template.fillna(0)  # All dummies set to 0 (reference categories)

        # Set Age dummy variables
        if age != ref_age:
            age_var_name = f'Age_{age}'
            if age_var_name in selected_vars:
                X_template[age_var_name] = 1
                # All other age dummies remain 0 (already set)

        # All other variables remain at reference levels (Sex=Male, Heard_ECig=No, etc.)

        # Ensure all selected variables are present and in correct order
        X_pred = X_template[selected_vars].values.astype(float).reshape(1, -1)

        # Add intercept (model_result expects X with intercept)
        X_pred_with_intercept = sm.add_constant(X_pred, has_constant='add')

        # Get predicted probabilities
        try:
            # Predict on the linear predictor scale
            linear_pred = model_result.predict(X_pred_with_intercept, linear=True)
            predicted_prob_logit = linear_pred[0]

            # Convert to probability scale using inverse logit
            predicted_prob = 1 / (1 + np.exp(-predicted_prob_logit))

            predictions.append({
                'Age': age,
                'Predicted_Probability': predicted_prob
            })
        except Exception as e:
            print(f"Warning: Could not calculate prediction for Age={age}: {e}")
            import traceback
            traceback.print_exc()
            continue

    return pd.DataFrame(predictions)

# Calculate overall predicted probabilities by age only
overall_predictions_df = calculate_overall_predicted_probabilities(
    result_final,
    df_model_encoded,
    available_selected,
    age_categories
)

print(f"\nOverall predicted probabilities calculated for {len(overall_predictions_df)} age categories")

# Create overall plot
fig, ax = plt.subplots(figsize=(10, 6))

ax.plot(overall_predictions_df['Age'], overall_predictions_df['Predicted_Probability'] * 100,
        marker='o', linestyle='-', linewidth=2.5, markersize=8,
        color='#2E86AB', alpha=0.8)

# Customize the plot
ax.set_xlabel('Age (years)', fontsize=12, fontweight='bold')
ax.set_ylabel('Predicted Probability of E-cigarette Use (%)', fontsize=12, fontweight='bold')
ax.set_title('Overall Predicted Probabilities of E-cigarette Uptake by Age\n(All Other Variables at Reference Levels)',
             fontsize=14, fontweight='bold', pad=20)
ax.grid(True, alpha=0.3, linestyle='--')
ax.set_xlim(min(age_categories) - 0.5, max(age_categories) + 0.5)
ax.set_xticks(age_categories)

# Format y-axis as percentage
ax.yaxis.set_major_formatter(plt.FuncFormatter(lambda y, _: '{:.1f}%'.format(y)))

plt.tight_layout()
plt.show()

# Print overall summary table
print("\n" + "=" * 70)
print("OVERALL PREDICTED PROBABILITIES BY AGE")
print("=" * 70)
print("\nPredicted probabilities of e-cigarette use by age:")
print("(All other variables held at reference levels)")

overall_summary = overall_predictions_df.copy()
overall_summary['Predicted_Probability'] = overall_summary['Predicted_Probability'] * 100

overall_summary_display = overall_summary[['Age', 'Predicted_Probability']].copy()
overall_summary_display.columns = ['Age (years)', 'Predicted Probability (%)']

if use_tabulate:
    print(tabulate(overall_summary_display,
    headers='keys',
    tablefmt='grid',
    showindex=False,
    stralign='left',
    numalign='right',
    floatfmt='.2f'))
else:
    print(overall_summary_display.to_string(index=False))
```

## Predicted Probabilities by Age, Stratified by Sex

```{python}
import matplotlib.pyplot as plt
import seaborn as sns
from scipy.stats import norm

# Set plotting style for publication-quality figures
sns.set_style("whitegrid")
plt.rcParams['figure.figsize'] = (12, 8)
plt.rcParams['font.size'] = 12
plt.rcParams['axes.labelsize'] = 13
plt.rcParams['axes.titlesize'] = 14
plt.rcParams['xtick.labelsize'] = 11
plt.rcParams['ytick.labelsize'] = 11
plt.rcParams['legend.fontsize'] = 12

# Get age and sex categories from the data
age_categories = sorted([int(x) for x in df_complete['Age'].dropna().unique() if pd.notna(x)])
sex_categories = sorted([str(x) for x in df_complete['Sex'].dropna().unique() if pd.notna(x)])

def calculate_age_sex_predicted_probabilities(model_result, df_model_encoded, selected_vars, age_categories, sex_categories):
    """
    Calculate predicted probabilities for different age categories, stratified by sex.
    Uses reference levels for other variables (Heard_ECig=No, etc.)
    
    Parameters:
    -----------
    model_result : statsmodels GLMResults
        The fitted model results
    df_model_encoded : pandas.DataFrame
        The encoded dataframe used for modeling
    selected_vars : list
        List of selected variable names (excluding intercept)
    age_categories : list
        List of age categories (e.g., [11, 12, 13, 14, 15, 16, 17])
    sex_categories : list
        List of sex categories (e.g., ['Male', 'Female'])
    
    Returns:
    --------
    predictions_df : pandas.DataFrame
        DataFrame with age, sex, and predicted probability
    """
    predictions = []

    # Get reference age (minimum age, which is dropped for baseline)
    ref_age = min(age_categories)

    # Find Sex dummy variable name
    sex_var_name = None
    for var in selected_vars:
        if var.startswith('Sex_'):
            sex_var_name = var
            break

    # Determine which sex value corresponds to sex_var_name = 1
    sex_reference = 'Male'  # Default assumption
    if sex_var_name:
        if 'female' in sex_var_name.lower():
            sex_reference = 'Male'  # Female is coded as 1, Male is reference (0)

    for age in age_categories:
        for sex in sex_categories:
            # Create a template row with all variables set to reference (0)
            X_template = pd.DataFrame(index=[0], columns=selected_vars)
            X_template = X_template.fillna(0)  # All dummies set to 0 (reference categories)

            # Set Age dummy variables
            if age != ref_age:
                age_var_name = f'Age_{age}'
                if age_var_name in selected_vars:
                    X_template[age_var_name] = 1
                    # All other age dummies remain 0 (already set)

            # Set Sex dummy variable
            if sex_var_name and sex_var_name in selected_vars:
                if sex_reference == 'Male':
                    # Female is coded as 1, Male as 0
                    X_template[sex_var_name] = 1 if sex.lower() in ['female', 'f'] else 0
                else:
                    # Male is coded as 1, Female as 0
                    X_template[sex_var_name] = 1 if sex.lower() in ['male', 'm'] else 0

            # Ensure all selected variables are present and in correct order
            X_pred = X_template[selected_vars].values.astype(float).reshape(1, -1)

            # Add intercept (model_result expects X with intercept)
            X_pred_with_intercept = sm.add_constant(X_pred, has_constant='add')

            # Get predicted probabilities
            try:
                # Predict on the linear predictor scale
                linear_pred = model_result.predict(X_pred_with_intercept, linear=True)
                predicted_prob_logit = linear_pred[0]

                # Convert to probability scale using inverse logit
                predicted_prob = 1 / (1 + np.exp(-predicted_prob_logit))

                predictions.append({
                    'Age': age,
                    'Sex': sex,
                    'Predicted_Probability': predicted_prob
                })
            except Exception as e:
                print(f"Warning: Could not calculate prediction for Age={age}, Sex={sex}: {e}")
                import traceback
                traceback.print_exc()
                continue

    return pd.DataFrame(predictions)

# Calculate predicted probabilities by age and sex
age_sex_predictions_df = calculate_age_sex_predicted_probabilities(
    result_final,
    df_model_encoded,
    available_selected,
    age_categories,
    sex_categories
)

print(f"\nPredicted probabilities by age and sex calculated for {len(age_sex_predictions_df)} scenarios")

# Create faceted plots by sex
fig, axes = plt.subplots(1, len(sex_categories), figsize=(16, 6), sharey=True)
if len(sex_categories) == 1:
    axes = [axes]

# Colors for sex categories
sex_colors = {'Male': '#2E86AB', 'Female': '#A23B72', 'male': '#2E86AB', 'female': '#A23B72'}

for i, sex in enumerate(sex_categories):
    ax = axes[i]
    sex_data = age_sex_predictions_df[age_sex_predictions_df['Sex'] == sex].sort_values('Age')

    if len(sex_data) > 0:
        ax.plot(sex_data['Age'], sex_data['Predicted_Probability'] * 100,
                marker='o', linestyle='-', linewidth=2.5, markersize=8,
                color=sex_colors.get(sex, '#000000'), alpha=0.8)

    # Customize subplot
    ax.set_xlabel('Age (years)', fontsize=12, fontweight='bold')
    ax.set_title(f'{sex}', fontsize=14, fontweight='bold')
    ax.grid(True, alpha=0.3, linestyle='--')
    ax.set_xlim(min(age_categories) - 0.5, max(age_categories) + 0.5)
    ax.set_xticks(age_categories)

# Set common y-label
fig.text(0.04, 0.5, 'Predicted Probability of E-cigarette Use (%)',
         va='center', rotation='vertical', fontsize=12, fontweight='bold')

# Format y-axis as percentage for all subplots
for ax in axes:
    ax.yaxis.set_major_formatter(plt.FuncFormatter(lambda y, _: '{:.1f}%'.format(y)))

# Add main title
fig.suptitle('Predicted Probabilities of E-cigarette Uptake by Age, Stratified by Sex\n(Adjusted for All Covariates in Final Model)',
             fontsize=16, fontweight='bold', y=1.02)

plt.tight_layout()
plt.show()

# Print summary table
print("\n" + "=" * 80)
print("PREDICTED PROBABILITIES BY AGE AND SEX")
print("=" * 80)
print("\nPredicted probabilities of e-cigarette use by age and sex:")
print("(All other variables held at reference levels)")

age_sex_summary = age_sex_predictions_df.copy()
age_sex_summary['Predicted_Probability'] = age_sex_summary['Predicted_Probability'] * 100

age_sex_summary_display = age_sex_summary[['Age', 'Sex', 'Predicted_Probability']].copy()
age_sex_summary_display.columns = ['Age (years)', 'Sex', 'Predicted Probability (%)']

if use_tabulate:
    print(tabulate(age_sex_summary_display,
    headers='keys',
    tablefmt='grid',
    showindex=False,
    stralign='left',
    numalign='right',
    floatfmt='.2f'))
else:
    print(age_sex_summary_display.to_string(index=False))
```

## Predicted Probabilities by Age and Sex

```{python}
import matplotlib.pyplot as plt
import seaborn as sns
from scipy.stats import norm

# Set plotting style for publication-quality figures
sns.set_style("whitegrid")
plt.rcParams['figure.figsize'] = (12, 8)
plt.rcParams['font.size'] = 12
plt.rcParams['axes.labelsize'] = 13
plt.rcParams['axes.titlesize'] = 14
plt.rcParams['xtick.labelsize'] = 11
plt.rcParams['ytick.labelsize'] = 11
plt.rcParams['legend.fontsize'] = 12

def calculate_predicted_probabilities(model_result, df_model_encoded, selected_vars, age_categories, sex_categories, heard_categories=None):
    """
    Calculate predicted probabilities for different age categories, stratified by sex and heard of e-cigarettes.

    Parameters:
    -----------
    model_result : statsmodels GLMResults
        The fitted model results
    df_model_encoded : pandas.DataFrame
        The encoded dataframe used for modeling
    selected_vars : list
        List of selected variable names (excluding intercept)
    age_categories : list
        List of age categories (e.g., [11, 12, 13, 14, 15, 16, 17])
    sex_categories : list
        List of sex categories (e.g., ['Male', 'Female'])
    heard_categories : list, optional
        List of heard of e-cigarettes categories (e.g., ['Yes', 'No'])

    Returns:
    --------
    predictions_df : pandas.DataFrame
        DataFrame with age, sex, heard_ecig, and predicted probability
    """
    predictions = []

    # Get reference age (minimum age, which is dropped for baseline)
    ref_age = min(age_categories)

    # Default heard categories if not provided
    if heard_categories is None:
        heard_categories = ['Yes', 'No']

    # Find Sex dummy variable name
    sex_var_name = None
    for var in selected_vars:
        if var.startswith('Sex_'):
            sex_var_name = var
            break

    # Find Heard dummy variable name
    heard_var_name = None
    for var in selected_vars:
        if var.startswith('heard_ecig_'):
            heard_var_name = var
            break

    # Determine which sex value corresponds to sex_var_name = 1
    # Typically, if it's 'Sex_female', then female=1, male=0 (male is reference)
    sex_reference = 'Male'  # Default assumption
    if sex_var_name:
        if 'female' in sex_var_name.lower():
            sex_reference = 'Male'  # Female is coded as 1, Male is reference (0)

    for age in age_categories:
        for sex in sex_categories:
            for heard in heard_categories:
                # Create a template row with all variables set to reference (0)
                X_template = pd.DataFrame(index=[0], columns=selected_vars)
                X_template = X_template.fillna(0)  # All dummies set to 0 (reference categories)

                # Set Age dummy variables
                if age != ref_age:
                    age_var_name = f'Age_{age}'
                    if age_var_name in selected_vars:
                        X_template[age_var_name] = 1
                        # All other age dummies remain 0 (already set)

                # Set Sex dummy variable
                if sex_var_name and sex_var_name in selected_vars:
                    if sex_reference == 'Male':
                        # Female is coded as 1, Male as 0
                        X_template[sex_var_name] = 1 if sex.lower() in ['female', 'f'] else 0
                    else:
                        # Male is coded as 1, Female as 0
                        X_template[sex_var_name] = 1 if sex.lower() in ['male', 'm'] else 0

                # Set Heard dummy variable
                if heard_var_name and heard_var_name in selected_vars:
                    # heard_ecig_Yes is typically the dummy variable (Yes=1, No=0 is reference)
                    X_template[heard_var_name] = 1 if heard.lower() in ['yes', 'y'] else 0

                # Ensure all selected variables are present and in correct order
                X_pred = X_template[selected_vars].values.astype(float).reshape(1, -1)

                # Add intercept (model_result expects X with intercept)
                X_pred_with_intercept = sm.add_constant(X_pred, has_constant='add')

                # Get predicted probabilities
                try:
                    # Predict on the linear predictor scale
                    linear_pred = model_result.predict(X_pred_with_intercept, linear=True)
                    predicted_prob_logit = linear_pred[0]

                    # Convert to probability scale using inverse logit
                    predicted_prob = 1 / (1 + np.exp(-predicted_prob_logit))

                    predictions.append({
                        'Age': age,
                        'Sex': sex,
                        'Heard_ECig': heard,
                        'Predicted_Probability': predicted_prob
                    })
                except Exception as e:
                    print(f"Warning: Could not calculate prediction for Age={age}, Sex={sex}, Heard={heard}: {e}")
                    import traceback
                    traceback.print_exc()
                    continue

    return pd.DataFrame(predictions)

# Get age categories from the data
age_categories = sorted([int(x) for x in df_complete['Age'].dropna().unique() if pd.notna(x)])
ref_age = min(age_categories)  # Reference age category

# Get sex categories from the data
sex_categories = sorted([str(x) for x in df_complete['Sex'].dropna().unique() if pd.notna(x)])

print(f"\n" + "=" * 80)
print("CALCULATING PREDICTED PROBABILITIES")
print("=" * 80)
print(f"Age categories: {age_categories}")
print(f"Sex categories: {sex_categories}")
print(f"Reference age: {ref_age} years")
print(f"\nVariables in final model: {available_selected}")
print(f"Total variables: {len(available_selected)}")

# Calculate predicted probabilities using the selected variables from the final model
predictions_df = calculate_predicted_probabilities(
result_final,
df_model_encoded,
available_selected,
age_categories,
sex_categories,
heard_categories=['Yes', 'No']
)

print(f"\nPredicted probabilities calculated for {len(predictions_df)} scenarios")

# Create faceted plots by sex
fig, axes = plt.subplots(1, len(sex_categories), figsize=(16, 6), sharey=True)
if len(sex_categories) == 1:
    axes = [axes]

# Colors and markers for heard_ecig status
heard_colors = {'Yes': '#FF6B6B', 'No': '#4ECDC4', 'yes': '#FF6B6B', 'no': '#4ECDC4'}
heard_markers = {'Yes': 'o', 'No': 's', 'yes': 'o', 'no': 's'}
heard_labels = {'Yes': 'Heard of E-cigarettes', 'No': 'Not Heard of E-cigarettes'}

for i, sex in enumerate(sex_categories):
    ax = axes[i]
    sex_data = predictions_df[predictions_df['Sex'] == sex]

    for heard in ['Yes', 'No']:
        heard_data = sex_data[sex_data['Heard_ECig'] == heard].sort_values('Age')

        if len(heard_data) > 0:
            ax.plot(heard_data['Age'], heard_data['Predicted_Probability'] * 100,
                    marker=heard_markers.get(heard, 'o'),
                    linestyle='-',
                    linewidth=2.5,
                    markersize=8,
                    color=heard_colors.get(heard, '#000000'),
                    label=heard_labels.get(heard, heard),
                    alpha=0.8)

    # Customize subplot
    ax.set_xlabel('Age (years)', fontsize=12, fontweight='bold')
    ax.set_title(f'{sex}', fontsize=14, fontweight='bold')
    ax.grid(True, alpha=0.3, linestyle='--')
    ax.set_xlim(min(age_categories) - 0.5, max(age_categories) + 0.5)
    ax.set_xticks(age_categories)

    # Add legend to the first subplot only
    if i == 0:
        ax.legend(loc='best', fontsize=10, framealpha=0.9)

# Set common y-label
fig.text(0.04, 0.5, 'Predicted Probability of E-cigarette Use (%)',
         va='center', rotation='vertical', fontsize=12, fontweight='bold')

# Format y-axis as percentage for all subplots
for ax in axes:
    ax.yaxis.set_major_formatter(plt.FuncFormatter(lambda y, _: '{:.1f}%'.format(y)))

# Add main title
fig.suptitle('Predicted Probabilities of E-cigarette Uptake by Age, Sex, and Awareness of E-cigarettes\n(Adjusted for All Covariates in Final Model)',
             fontsize=16, fontweight='bold', y=1.02)

plt.tight_layout()
plt.show()

# Print a summary table
print("\n" + "=" * 80)
print("PREDICTED PROBABILITIES SUMMARY")
print("=" * 80)
print("\nPredicted probabilities of e-cigarette use by age and sex:")
print("(All other variables held at reference levels)")

pred_summary = predictions_df.copy()
pred_summary['Predicted_Probability'] = pred_summary['Predicted_Probability'] * 100

pred_summary_display = pred_summary[['Age', 'Sex', 'Heard_ECig', 'Predicted_Probability']].copy()
pred_summary_display.columns = ['Age (years)', 'Sex', 'Heard of E-cigarettes', 'Predicted Probability (%)']

if use_tabulate:
    print(tabulate(pred_summary_display, 
headers='keys', 
tablefmt='grid', 
showindex=False,
stralign='left',
numalign='right',
floatfmt='.2f'))
else:
    print(pred_summary_display.to_string(index=False))
```

## Summary

```{python}
print("\n" + "=" * 100)
print("ANALYSIS SUMMARY")
print("=" * 100)
print(f"\n1. Countries included: Ghana, Mauritius, Seychelles, and Togo")
print(f"2. Total sample size: {len(df_analysis):,}")
print(f"3. Dataset for modeling after imputation: {len(df_complete):,}")
print(f"   (Missing values were imputed based on observed distributions)")
print(f"4. Outcome variable: {var_label_map.get(outcome, 'Current E-cigarette Use')}")
print(f"5. Total predictors tested: {len(predictors)}")
print(f"6. Variables in final model: {len(selected_vars)}")
print(f"7. Final model AIC (Akaike Information Criterion): {final_aic:.2f}")
print(f"8. Final model BIC: {result_final.bic:.2f}")
print(f"\nSelected variables in final weighted logistic regression model:")
print("   (Age, Sex, and Grade were included as a priori variables)")
for i, var in enumerate(selected_vars, 1):
    # Format variable name nicely
    if var in var_label_map:
        var_display = var_label_map[var]
    elif '_' in var:
        base_var = var.split('_')[0]
        var_display = var_label_map.get(base_var, base_var.replace('_', ' ').title())
        if len(var.split('_')) > 1:
            category_part = '_'.join(var.split('_')[1:])
            try:
                cat_val = int(category_part) if category_part.isdigit() else category_part
                category_label = map_value_label(base_var, cat_val)
                var_display = f"{var_display} ({category_label})"
            except:
                var_display = f"{var_display} ({category_part})"
    else:
        var_display = var.replace('_', ' ').title()
    print(f"   {i}. {var_display}")
print(f"\n" + "=" * 100)
print("Model Interpretation:")
print("- Odds ratios (OR) > 1 indicate increased odds of e-cigarette use")
print("- Odds ratios (OR) < 1 indicate decreased odds of e-cigarette use")
print("- Odds ratio (OR) = 1 indicates no association")
print("- All analyses account for survey design through weighted methods")
print("=" * 100)
```
````
