## Supplemental codes for "Methodological Guidance for Predictor Variable Selection for Adolescent Smoking Outcomes in Global Youth Tobacco Survey Using R and Python": S2b_Complex_survey_weighted_two_stage_design.py

```
# Python Implementation (statsmodels):
# Python: Survey-Weighted GLM

import pandas as pd
import statsmodels.api as sm
from statsmodels.genmod.generalized_linear_model import GLM
from statsmodels.genmod import families

# Python requires manual weight normalization for numerical stability
weights_norm = df["FinalWgt"] * (len(df) / df["FinalWgt"].sum())

# Design specification is implicit in GLM through freq_weights
# Note: No explicit strata/PSU specification in statsmodels GLM
# For complex designs with clustering, consider samplics or survey package via rpy2

model = GLM(
    y,
    X,
    family=families.Binomial(link=families.links.Logit()),
    freq_weights=weights_norm,
)
```
