## Supplemental codes for "Methodological Guidance for Predictor Variable Selection for Adolescent Smoking Outcomes in Global Youth Tobacco Survey Using R and Python": S3b_Stepwise_selection.py

```
# Python Implementation (custom algorithm):
# Python: Custom Constrained Stepwise Function

import pandas as pd
import numpy as np
from statsmodels.genmod.generalized_linear_model import GLM
from statsmodels.genmod import families
import statsmodels.api as sm


def constrained_stepwise_selection(
    X,
    y,
    weights,
    apriori_vars,
    candidate_vars,
    direction="both",
    max_iter=50,
    improvement_threshold=0.01,
):
    """
    Perform stepwise selection with a priori variable protection

    Algorithm protects apriori_vars from removal while allowing
    data-driven selection from candidate_vars using AIC criterion.
    """

    # Initialize with a priori variables (protected)
    selected = [v for v in apriori_vars if v in X.columns]
    remaining = [v for v in candidate_vars if v in X.columns and v not in selected]

    # Fit initial model (a priori only)
    X_init = sm.add_constant(X[selected])
    model_init = GLM(
        y,
        X_init,
        family=families.Binomial(link=families.links.Logit()),
        freq_weights=weights,
    )
    result_init = model_init.fit(method="lbfgs", disp=0)
    current_aic = result_init.aic

    print(f"Initial model (a priori only) AIC: {current_aic:.2f}")

    # Iterative selection
    for iteration in range(max_iter):
        improved = False

        # FORWARD STEP: Try adding each remaining variable
        if direction in ["forward", "both"] and remaining:
            scores = []
            for var in remaining:
                try:
                    X_test = sm.add_constant(X[selected + [var]])
                    model = GLM(
                        y, X_test, family=families.Binomial(), freq_weights=weights
                    )
                    result = model.fit(method="lbfgs", disp=0)
                    scores.append((var, result.aic))
                except:
                    continue

            if scores:
                scores.sort(key=lambda x: x[1])
                best_var, best_aic = scores[0]

                # Add if AIC improves by threshold
                if best_aic < current_aic - improvement_threshold:
                    selected.append(best_var)
                    remaining.remove(best_var)
                    current_aic = best_aic
                    improved = True
                    print(f"  Added {best_var}: AIC = {current_aic:.2f}")
                    continue  # Go to next iteration

        # BACKWARD STEP: Try removing non-a priori variables
        if direction in ["backward", "both"] and len(selected) > len(apriori_vars):
            removable = [v for v in selected if v not in apriori_vars]
            scores = []

            for var in removable:
                try:
                    new_selected = [v for v in selected if v != var]
                    X_test = sm.add_constant(X[new_selected])
                    model = GLM(
                        y, X_test, family=families.Binomial(), freq_weights=weights
                    )
                    result = model.fit(method="lbfgs", disp=0)
                    scores.append((var, result.aic, new_selected))
                except:
                    continue

            if scores:
                scores.sort(key=lambda x: x[1])
                worst_var, best_aic, new_selected = scores[0]

                # Remove if AIC improves by threshold
                if best_aic < current_aic - improvement_threshold:
                    selected = new_selected
                    remaining.append(worst_var)
                    current_aic = best_aic
                    improved = True
                    print(f"  Removed {worst_var}: AIC = {current_aic:.2f}")

        if not improved:
            break

    print(
        f"\nFinal model: {len(selected)} variables "
        f"({len(apriori_vars)} a priori + {len(selected) - len(apriori_vars)} selected)"
    )

    return selected, current_aic


# Execute stepwise selection
apriori = ["Age_12", "Age_13", "Age_14", "Age_15", "Age_16", "Age_17", "Sex_female"]
candidates = [
    "tobacco_awareness_Yes",
    "smk_try_Yes",
    "friend_smk_Probably yes",
    "taught_smking_dangers_Yes",
    "shisha_Yes",
    "parent_smokes_Yes",
]

selected_vars, final_aic = constrained_stepwise_selection(
    X, y, weights_norm, apriori, candidates, direction="both"
)
```
