## Supplemental codes for "Methodological Guidance for Predictor Variable Selection for Adolescent Smoking Outcomes in Global Youth Tobacco Survey Using R and Python": Software Requirements and Installat.txt

Software Requirements and Installation
Python Requirements
•	Python 3.10+
•	Packages: pandas, numpy, statsmodels, scipy
Install Python
1.	Download from python.org and run the installer.
2.	Select “Add Python to PATH” during installation.
3.	Verify: python --version in a terminal.
Set Up a Virtual Environment
1.	Create: python -m venv .venv
2.	Activate (Windows): .venv\\Scripts\\activate
3.	Install: pip install pandas numpy statsmodels scipy
VSCode and Quarto
1.	Install VSCode from code.visualstudio.com.
2.	Add extensions: Python, R, and Quarto.
3.	Install Quarto from quarto.org and verify with quarto --version.

R Requirements
•	R 4.2+
•	Packages: survey, MASS, broom, dplyr, tidyr
Install R and RStudio
1.	Install R from cran.r-project.org.
2.	Install RStudio from posit.co.
3.	Verify R: run R --version or open RStudio.
Install R Packages
install.packages(c(\"survey\", \"MASS\", \"broom\", \"dplyr\", \"tidyr\"))
Reproducible Documents
•	Use Quarto for cross-platform reports; author in .qmd and render to HTML/PDF/DOCX.
•	Configure project environments with R’s renv or Python’s venv/conda.
