## Supplemental codes for "Methodological Guidance for Predictor Variable Selection for Adolescent Smoking Outcomes in Global Youth Tobacco Survey Using R and Python": FIXES_NEEDED.md

# Fixes Needed for QMD Files

## Syntax Error Found

All files have an invalid syntax error in the region assignment:
- **Error**: `grepl("Lusaka", .$`Region` | .$`Province`, ignore.case = TRUE)`
- **Fix**: `grepl("Lusaka", Region, ignore.case = TRUE) | grepl("Lusaka", Province, ignore.case = TRUE)`

The `.$` syntax doesn't work in dplyr `mutate()`. Use column names directly.

## Files Fixed
- ✅ paper01_tobacco_growing_association.qmd
- ✅ paper02_factors_shisha.qmd

## Files Status
- ✅ ALL 20 FILES - Syntax error fixed

## Missing Components from Prompts

### Paper 1 (Tobacco Growing Association)
- ✅ Added: Sensitivity analyses structure (excluding occasional users, different definitions)
- ✅ Added: Supplementary Table 2 placeholder (Sensitivity analysis results)
- ✅ Added: Prevalence ratios (PR) with Tobacco_Growing as reference
- ✅ Added: Pairwise comparisons with Bonferroni correction structure
- ✅ Added: Model 3 structure (Full model - add socioeconomic factors when available)
- ⚠️ Note: Figure 3 (Map) requires geographic data - placeholder structure provided

### Paper 2 (Shisha)
- ⚠️ Missing: Table 2 (Distribution of predictor variables by shisha smoking status within each region)
- ⚠️ Missing: Region-specific bivariate analysis tables
- ⚠️ Missing: Meta-regression to explain regional differences
- ⚠️ Missing: I² statistic for between-region heterogeneity
- ⚠️ Missing: Figure 3 (Pathway diagram)

### Paper 3 (Trends)
- ℹ️ Note: Requires access to previous GYTS waves - structure is in place
- ⚠️ Missing: Joinpoint regression analysis
- ⚠️ Missing: Annual percentage change (APC) calculations
- ⚠️ Missing: Projections to 2025 and 2030

### Paper 4 (Teaching)
- ⚠️ Missing: Mediation analysis by region
- ⚠️ Missing: Figure 3 (Mediation pathway diagrams)
- ⚠️ Missing: Dose-response analysis by quartiles

### Paper 5 (Reasons)
- ℹ️ Note: Requires identification of reason variables in dataset
- Structure is in place as placeholder

### Paper 6 (Cigarette Smoking)
- ⚠️ Missing: Table 2 (Heterogeneity tests)
- ⚠️ Missing: Table 3 (Machine learning variable importance)
- ⚠️ Missing: Table 4 (Integrated model with interaction terms)
- ⚠️ Missing: Machine learning approach (Random Forest)

## General Notes

1. **Variable Names**: All files use placeholder variable names (e.g., Region, Province, WEIGHT, SCHOOL_ID). These need to be verified against the actual dataset.

2. **Survey Weights**: All files check for "WEIGHT" variable. Update to match actual weight variable name.

3. **School Clustering**: Multilevel models reference "SCHOOL_ID". Update if different.

4. **Region Classification**: The region logic assumes certain province names. Verify actual region codes in your dataset.
