## Supplemental codes for "Methodological Guidance for Predictor Variable Selection for Adolescent Smoking Outcomes in Global Youth Tobacco Survey Using R and Python": HOW_TO_RENDER.md

# How to Render All Papers

## Quick Start

```powershell
# From project root directory
cd draft_papers
quarto render paper*.qmd
```

Or render individually:
```powershell
quarto render paper01_tobacco_growing_association.qmd
quarto render paper02_factors_shisha.qmd
# etc.
```

## Before First Render

1. **Install R packages** (if not already installed):
```r
install.packages(c("readxl", "tidyverse", "survey", "gtsummary", 
                   "flextable", "patchwork", "scales", "viridis", 
                   "RColorBrewer", "knitr", "kableExtra", "broom", 
                   "lme4", "emmeans", "ggplot2", "ggthemes", 
                   "stringr", "purrr"))
```

2. **Verify variable names** in your dataset and update in the .qmd files:
   - Region/Province column names
   - Weight variable name
   - School ID variable name
   - Questionnaire variable codes (CR1, CR3, etc.)

## Expected Output

Each paper will generate:
- **HTML file**: `paper##_filename.html` 
- **Figures folder**: With high-resolution PNG/PDF figures
- **Publication-ready tables**: Formatted with flextable
- **Professional visualizations**: With error bars, proper labels

## Troubleshooting

- **Variable not found errors**: Update variable names to match your dataset
- **Package missing errors**: Install the required R packages
- **File path errors**: Ensure you're running from the correct directory
