## Supplemental codes for "Methodological Guidance for Predictor Variable Selection for Adolescent Smoking Outcomes in Global Youth Tobacco Survey Using R and Python": HUMANIZATION_NEEDED.md

# Data Humanization Status: Tables Need Label Conversion

## ❌ **CRITICAL ISSUE FOUND**

### **Problem**: Regression tables show technical variable names

Many tables, especially regression model tables, display raw variable names from the model output (like `term` from `broom::tidy()`). These technical names are **NOT humanized** and would require referring to the questionnaire to understand.

### **Examples of Non-Humanized Labels Currently in Tables:**

**Paper 1, Table 2 (Regression Models):**
- Shows: `term` column with values like:
  - `regionTobacco_Growing` (should be: "Tobacco Growing region (vs Other Zambia)")
  - `regionLusaka` (should be: "Lusaka (vs Other Zambia)")
  - `age_group15-16` (should be: "Age 15-16 years (vs 13-14 years)")
  - `genderMale` (should be: "Male (vs Female)")
  - `grade_levelGrade 8` (should be: "Grade 8 (vs Grade 9)")

**Paper 2, Table 2 (Shisha Models):**
- Shows: `term` with values like:
  - `genderMale`
  - `age_group15-16`
  - `parental_smokingAt least one parent`
  - `friends_smokeSome`
  - `ad_exposureHigh`

**Paper 6, Table 1 (Cigarette Smoking Models):**
- Similar issue with technical variable names

### ✅ **What IS Humanized:**

1. **Prevalence tables (Table 1s)**: Generally good
   - Paper 1: Uses `outcome_labels` = "Current tobacco use (any product)", etc. ✅
   - Paper 2: Uses "Variable", "Category", "Prevalence" columns ✅
   - Most prevalence tables use descriptive labels

2. **Figure labels**: Generally good
   - Most figures use `outcome_label`, descriptive axis labels
   - Some forest plots convert terms to readable labels ✅

### ❌ **What NEEDS Humanization:**

1. **Regression tables (Table 2s, Table 3s)**: 
   - All use raw `term` from model output
   - Need conversion to plain English

2. **Some interaction tables**:
   - Show raw interaction terms like `regionLusaka:genderMale`

3. **Chi-square test tables**:
   - Some show technical outcome variable names

## **Solution Needed:**

Create a function to convert technical variable names to human-readable labels:

```r
# Function to humanize variable names
humanize_term <- function(term, reference_levels = NULL) {
  term_clean <- term %>%
    # Handle region terms
    str_replace("^regionTobacco_Growing", "Tobacco Growing region (vs Other Zambia)") %>%
    str_replace("^regionLusaka", "Lusaka (vs Other Zambia)") %>%
    # Handle gender
    str_replace("^genderMale", "Male (vs Female)") %>%
    # Handle age groups
    str_replace("^age_group15-16", "Age 15-16 years (vs 13-14 years)") %>%
    str_replace("^age_group17\\+", "Age 17+ years (vs 13-14 years)") %>%
    # Handle grade levels
    str_replace("^grade_levelGrade 8", "Grade 8 (vs Grade 9)") %>%
    str_replace("^grade_levelGrade 7", "Grade 7 (vs Grade 9)") %>%
    # Handle parental smoking
    str_replace("^parental_smokingAt least one", "At least one parent smokes (vs None)") %>%
    # Handle friends smoking
    str_replace("^friends_smokeSome", "Some friends smoke (vs None)") %>%
    str_replace("^friends_smokeMost/All", "Most/All friends smoke (vs None)") %>%
    # Handle other variables...
    # Return cleaned term
    term_clean
}

# Then use in table creation:
table2_data <- bind_rows(model1_tidy, model2_tidy, model3_tidy) %>%
  mutate(
    Variable = humanize_term(term),  # Convert to readable
    OR = sprintf("%.2f", estimate),
    `95% CI` = sprintf("(%.2f, %.2f)", conf.low, conf.high),
    `p-value` = sprintf("%.3f", p.value)
  ) %>%
  select(Model, Variable, OR, `95% CI`, `p-value`)  # Use Variable instead of term
```

## **Files Needing Updates:**

1. ✅ Paper 1 - Table 1: Already humanized (uses outcome_labels)
2. ❌ Paper 1 - Table 2: NEEDS humanization (uses raw `term`)
3. ❌ Paper 1 - Table 3: NEEDS humanization (interaction terms)
4. ❌ Paper 2 - Table 2: NEEDS humanization
5. ❌ Paper 6 - Table 1: NEEDS humanization
6. ❌ Most regression tables across all papers

## **Priority:**

**HIGH PRIORITY** - Without humanization, authors cannot write manuscript results sections without constantly referring back to the questionnaire to understand what variables like "genderMale" or "parental_smokingAt least one parent" actually mean.

The tables should show labels like:
- "Male (vs Female)" instead of "genderMale"
- "Age 15-16 years" instead of "age_group15-16"
- "At least one parent smokes" instead of "parental_smokingAt least one parent"
