## Supplemental codes for "Methodological Guidance for Predictor Variable Selection for Adolescent Smoking Outcomes in Global Youth Tobacco Survey Using R and Python": paper01_tobacco_growing_association.knit.md

---
title: "Current Tobacco Use Among Adolescents in Zambia: Analysis of GYTS 2021 Data"
subtitle: "Paper 1: GYTS 2021 Analysis"
author: "GYTS Research Team"
date: today
bibliography: references.bib
format:
  html:
    toc: true
    toc-depth: 3
    toc-location: left
    code-fold: show
    code-tools: true
    fig-width: 10
    fig-height: 6
    theme: cosmo
    css: styles.css
execute:
  warning: false
  message: false
---


# Introduction

Tobacco use remains one of the leading preventable causes of death and disease worldwide, with devastating consequences that extend far beyond individual health to impact families, communities, and entire healthcare systems [@who_tobacco_2021]. Among adolescents, the burden of tobacco use is particularly concerning as this critical developmental period represents a vulnerable window for initiation of tobacco products, with most adult smokers beginning their habit before the age of 18 [@usdhhs_preventing_2012]. Globally, an estimated 24 million adolescents aged 13-15 years use tobacco products, with cigarette smoking being the most common form of tobacco consumption [@who_global_2020]. The World Health Organization's Framework Convention on Tobacco Control (FCTC) has recognized the urgent need to protect young people from tobacco use, yet progress remains uneven across regions and countries [@who_fctc_2003]. Early initiation of tobacco use not only increases the risk of developing nicotine dependence but also significantly elevates the likelihood of continued use into adulthood, creating a trajectory of lifelong health risks including cardiovascular disease, respiratory illness, and various forms of cancer [@warren_global_2006].

The African continent faces unique challenges in addressing tobacco use among its youth population, with rapidly changing social and economic landscapes creating new opportunities for tobacco industry marketing and product accessibility [@who_africa_2019]. While overall tobacco prevalence rates in many African countries remain lower than global averages, the continent represents a critical frontier for tobacco control efforts, as tobacco companies increasingly target African markets with aggressive marketing strategies [@legg_tobacco_2014]. The combination of growing youth populations, increasing urbanization, and expanding middle-class purchasing power creates an environment where tobacco use could escalate rapidly without effective prevention and control measures [@drope_tobacco_2018]. Furthermore, the diversity of tobacco products available in African markets, including traditional forms such as shisha and smokeless tobacco, adds complexity to surveillance and intervention efforts [@who_africa_tobacco_2021].

Zambia, like many countries in sub-Saharan Africa, is at a critical juncture in its tobacco control journey. The country has made important strides in tobacco control policy, including ratification of the WHO FCTC and implementation of various tobacco control measures [@moh_zambia_2018]. However, data on tobacco use patterns among Zambian adolescents remain limited, creating gaps in understanding the current landscape of youth tobacco use and the factors that drive initiation and continued use [@zyambo_gyts_2021]. The 2021 Global Youth Tobacco Survey (GYTS) provides a valuable opportunity to examine current tobacco use prevalence and associated factors among school-going adolescents in Zambia, offering insights that can inform evidence-based prevention and control strategies. Understanding the specific patterns and predictors of tobacco use among Zambian youth is essential for developing targeted interventions that address the unique cultural, social, and economic contexts within which young people make decisions about tobacco use [@who_gyts_2021].

This analysis aims to examine the prevalence of current tobacco use among adolescents in Zambia and identify key factors associated with tobacco use initiation and continuation. Specifically, we seek to: (1) describe the prevalence of current tobacco use across different demographic and socioeconomic groups; (2) identify individual, social, and environmental factors associated with current tobacco use; and (3) assess the relationship between tobacco-growing regions and tobacco use patterns among adolescents. The findings from this analysis have important policy implications for strengthening tobacco control efforts in Zambia, particularly in the areas of youth-focused prevention programs, enforcement of tobacco control legislation, and addressing the unique challenges posed by tobacco-growing regions where economic dependence on tobacco production may create additional barriers to effective tobacco control [@who_tobacco_control_2021]. By providing evidence-based insights into the factors driving tobacco use among Zambian adolescents, this analysis can inform the development of targeted interventions and policies that protect young people from the harms of tobacco use while supporting sustainable economic alternatives in tobacco-growing communities.

# Methods

## Data Source
- **Survey**: Global Youth Tobacco Survey (GYTS) 2021, Zambia
- **Analysis Dataset**: GYTS 2021 Zambia (National).XLS, sheet "ZAMBIA_DATA_21"

## Data Management

All variables used in this analysis were derived from the GYTS 2021 dataset. The outcome variable for this analysis is current tobacco smoking, defined as a binary variable where 1 = current tobacco smoking (CR7 > 1 OR CR14 == 1 OR CR43 == 1) and 0 = otherwise. All predictor variables were created following the same methodology as described in Paper 1, ensuring consistency across analyses.

### Variables Included in Analysis

The following table provides definitions and value labels for all variables included in this analysis:


```{=html}
<div class="tabwid"><style>.cl-539a94ba{}.cl-53694e46{font-family:'Arial';font-size:9pt;font-weight:normal;font-style:normal;text-decoration:none;color:rgba(0, 0, 0, 1.00);background-color:transparent;}.cl-53694edc{font-family:'Arial';font-size:9pt;font-weight:bold;font-style:normal;text-decoration:none;color:rgba(0, 0, 0, 1.00);background-color:transparent;}.cl-53694ef0{font-family:'Arial';font-size:8pt;font-weight:normal;font-style:italic;text-decoration:none;color:rgba(0, 0, 0, 1.00);background-color:transparent;}.cl-537dbad4{margin:0;text-align:left;border-bottom: 0 solid rgba(0, 0, 0, 1.00);border-top: 0 solid rgba(0, 0, 0, 1.00);border-left: 0 solid rgba(0, 0, 0, 1.00);border-right: 0 solid rgba(0, 0, 0, 1.00);padding-bottom:4pt;padding-top:4pt;padding-left:4pt;padding-right:4pt;line-height: 1;background-color:transparent;}.cl-537dbaf2{margin:0;text-align:left;border-bottom: 0 solid rgba(0, 0, 0, 1.00);border-top: 0 solid rgba(0, 0, 0, 1.00);border-left: 0 solid rgba(0, 0, 0, 1.00);border-right: 0 solid rgba(0, 0, 0, 1.00);padding-bottom:5pt;padding-top:5pt;padding-left:5pt;padding-right:5pt;line-height: 1;background-color:transparent;}.cl-537e5f8e{width:1.2in;background-color:transparent;vertical-align: middle;border-bottom: 1.5pt solid rgba(102, 102, 102, 1.00);border-top: 1.5pt solid rgba(102, 102, 102, 1.00);border-left: 0 solid rgba(0, 0, 0, 1.00);border-right: 0 solid rgba(0, 0, 0, 1.00);margin-bottom:0;margin-top:0;margin-left:0;margin-right:0;}.cl-537e5fac{width:2in;background-color:transparent;vertical-align: middle;border-bottom: 1.5pt solid rgba(102, 102, 102, 1.00);border-top: 1.5pt solid rgba(102, 102, 102, 1.00);border-left: 0 solid rgba(0, 0, 0, 1.00);border-right: 0 solid rgba(0, 0, 0, 1.00);margin-bottom:0;margin-top:0;margin-left:0;margin-right:0;}.cl-537e5fb6{width:2.5in;background-color:transparent;vertical-align: middle;border-bottom: 1.5pt solid rgba(102, 102, 102, 1.00);border-top: 1.5pt solid rgba(102, 102, 102, 1.00);border-left: 0 solid rgba(0, 0, 0, 1.00);border-right: 0 solid rgba(0, 0, 0, 1.00);margin-bottom:0;margin-top:0;margin-left:0;margin-right:0;}.cl-537e5fc0{width:1in;background-color:transparent;vertical-align: middle;border-bottom: 1.5pt solid rgba(102, 102, 102, 1.00);border-top: 1.5pt solid rgba(102, 102, 102, 1.00);border-left: 0 solid rgba(0, 0, 0, 1.00);border-right: 0 solid rgba(0, 0, 0, 1.00);margin-bottom:0;margin-top:0;margin-left:0;margin-right:0;}.cl-537e5fca{width:1.2in;background-color:transparent;vertical-align: middle;border-bottom: 0 solid rgba(0, 0, 0, 1.00);border-top: 0 solid rgba(0, 0, 0, 1.00);border-left: 0 solid rgba(0, 0, 0, 1.00);border-right: 0 solid rgba(0, 0, 0, 1.00);margin-bottom:0;margin-top:0;margin-left:0;margin-right:0;}.cl-537e5fd4{width:2in;background-color:transparent;vertical-align: middle;border-bottom: 0 solid rgba(0, 0, 0, 1.00);border-top: 0 solid rgba(0, 0, 0, 1.00);border-left: 0 solid rgba(0, 0, 0, 1.00);border-right: 0 solid rgba(0, 0, 0, 1.00);margin-bottom:0;margin-top:0;margin-left:0;margin-right:0;}.cl-537e5fde{width:2.5in;background-color:transparent;vertical-align: middle;border-bottom: 0 solid rgba(0, 0, 0, 1.00);border-top: 0 solid rgba(0, 0, 0, 1.00);border-left: 0 solid rgba(0, 0, 0, 1.00);border-right: 0 solid rgba(0, 0, 0, 1.00);margin-bottom:0;margin-top:0;margin-left:0;margin-right:0;}.cl-537e5fe8{width:1in;background-color:transparent;vertical-align: middle;border-bottom: 0 solid rgba(0, 0, 0, 1.00);border-top: 0 solid rgba(0, 0, 0, 1.00);border-left: 0 solid rgba(0, 0, 0, 1.00);border-right: 0 solid rgba(0, 0, 0, 1.00);margin-bottom:0;margin-top:0;margin-left:0;margin-right:0;}.cl-537e5fe9{width:1.2in;background-color:transparent;vertical-align: middle;border-bottom: 1.5pt solid rgba(102, 102, 102, 1.00);border-top: 0 solid rgba(0, 0, 0, 1.00);border-left: 0 solid rgba(0, 0, 0, 1.00);border-right: 0 solid rgba(0, 0, 0, 1.00);margin-bottom:0;margin-top:0;margin-left:0;margin-right:0;}.cl-537e5ff2{width:2in;background-color:transparent;vertical-align: middle;border-bottom: 1.5pt solid rgba(102, 102, 102, 1.00);border-top: 0 solid rgba(0, 0, 0, 1.00);border-left: 0 solid rgba(0, 0, 0, 1.00);border-right: 0 solid rgba(0, 0, 0, 1.00);margin-bottom:0;margin-top:0;margin-left:0;margin-right:0;}.cl-537e5ffc{width:2.5in;background-color:transparent;vertical-align: middle;border-bottom: 1.5pt solid rgba(102, 102, 102, 1.00);border-top: 0 solid rgba(0, 0, 0, 1.00);border-left: 0 solid rgba(0, 0, 0, 1.00);border-right: 0 solid rgba(0, 0, 0, 1.00);margin-bottom:0;margin-top:0;margin-left:0;margin-right:0;}.cl-537e5ffd{width:1in;background-color:transparent;vertical-align: middle;border-bottom: 1.5pt solid rgba(102, 102, 102, 1.00);border-top: 0 solid rgba(0, 0, 0, 1.00);border-left: 0 solid rgba(0, 0, 0, 1.00);border-right: 0 solid rgba(0, 0, 0, 1.00);margin-bottom:0;margin-top:0;margin-left:0;margin-right:0;}.cl-537e6006{width:1.2in;background-color:transparent;vertical-align: middle;border-bottom: 0 solid rgba(255, 255, 255, 0.00);border-top: 0 solid rgba(255, 255, 255, 0.00);border-left: 0 solid rgba(255, 255, 255, 0.00);border-right: 0 solid rgba(255, 255, 255, 0.00);margin-bottom:0;margin-top:0;margin-left:0;margin-right:0;}.cl-537e601a{width:2in;background-color:transparent;vertical-align: middle;border-bottom: 0 solid rgba(255, 255, 255, 0.00);border-top: 0 solid rgba(255, 255, 255, 0.00);border-left: 0 solid rgba(255, 255, 255, 0.00);border-right: 0 solid rgba(255, 255, 255, 0.00);margin-bottom:0;margin-top:0;margin-left:0;margin-right:0;}.cl-537e6024{width:2.5in;background-color:transparent;vertical-align: middle;border-bottom: 0 solid rgba(255, 255, 255, 0.00);border-top: 0 solid rgba(255, 255, 255, 0.00);border-left: 0 solid rgba(255, 255, 255, 0.00);border-right: 0 solid rgba(255, 255, 255, 0.00);margin-bottom:0;margin-top:0;margin-left:0;margin-right:0;}.cl-537e6025{width:1in;background-color:transparent;vertical-align: middle;border-bottom: 0 solid rgba(255, 255, 255, 0.00);border-top: 0 solid rgba(255, 255, 255, 0.00);border-left: 0 solid rgba(255, 255, 255, 0.00);border-right: 0 solid rgba(255, 255, 255, 0.00);margin-bottom:0;margin-top:0;margin-left:0;margin-right:0;}</style><table data-quarto-disable-processing='true' class='cl-539a94ba'><thead><tr style="overflow-wrap:break-word;"><th class="cl-537e5f8e"><p class="cl-537dbad4"><span class="cl-53694e46">**Variable Name**</span></p></th><th class="cl-537e5fac"><p class="cl-537dbad4"><span class="cl-53694e46">**Definition**</span></p></th><th class="cl-537e5fb6"><p class="cl-537dbad4"><span class="cl-53694e46">**Source Question**</span></p></th><th class="cl-537e5fac"><p class="cl-537dbad4"><span class="cl-53694e46">**Value Labels**</span></p></th><th class="cl-537e5fc0"><p class="cl-537dbad4"><span class="cl-53694e46">**Type**</span></p></th></tr></thead><tbody><tr style="overflow-wrap:break-word;"><td class="cl-537e5fca"><p class="cl-537dbad4"><span class="cl-53694edc">smokingnow</span></p></td><td class="cl-537e5fd4"><p class="cl-537dbad4"><span class="cl-53694e46">Current tobacco smoking status (outcome variable)</span></p></td><td class="cl-537e5fde"><p class="cl-537dbad4"><span class="cl-53694e46">CR7 &gt; 1 OR CR14 == 1 OR CR43 == 1</span></p></td><td class="cl-537e5fd4"><p class="cl-537dbad4"><span class="cl-53694e46">0 = No current smoking, 1 = Current tobacco smoking</span></p></td><td class="cl-537e5fe8"><p class="cl-537dbad4"><span class="cl-53694e46">Outcome (binary)</span></p></td></tr><tr style="overflow-wrap:break-word;"><td class="cl-537e5fca"><p class="cl-537dbad4"><span class="cl-53694edc">age_cr1</span></p></td><td class="cl-537e5fd4"><p class="cl-537dbad4"><span class="cl-53694e46">Age in years (factor: 11-17 years)</span></p></td><td class="cl-537e5fde"><p class="cl-537dbad4"><span class="cl-53694e46">CR1: How old are you?</span></p></td><td class="cl-537e5fd4"><p class="cl-537dbad4"><span class="cl-53694e46">11, 12, 13, 14, 15, 16, 17 (years)</span></p></td><td class="cl-537e5fe8"><p class="cl-537dbad4"><span class="cl-53694e46">A priori (categorical)</span></p></td></tr><tr style="overflow-wrap:break-word;"><td class="cl-537e5fca"><p class="cl-537dbad4"><span class="cl-53694edc">sex_cr2</span></p></td><td class="cl-537e5fd4"><p class="cl-537dbad4"><span class="cl-53694e46">Sex of participant</span></p></td><td class="cl-537e5fde"><p class="cl-537dbad4"><span class="cl-53694e46">CR2: What is your sex?</span></p></td><td class="cl-537e5fd4"><p class="cl-537dbad4"><span class="cl-53694e46">Male, Female</span></p></td><td class="cl-537e5fe8"><p class="cl-537dbad4"><span class="cl-53694e46">A priori (categorical)</span></p></td></tr><tr style="overflow-wrap:break-word;"><td class="cl-537e5fca"><p class="cl-537dbad4"><span class="cl-53694edc">grade_zar3</span></p></td><td class="cl-537e5fd4"><p class="cl-537dbad4"><span class="cl-53694e46">Grade level in school</span></p></td><td class="cl-537e5fde"><p class="cl-537dbad4"><span class="cl-53694e46">ZAR3: In what grade/form are you?</span></p></td><td class="cl-537e5fd4"><p class="cl-537dbad4"><span class="cl-53694e46">Grade 7, Grade 8, Grade 9</span></p></td><td class="cl-537e5fe8"><p class="cl-537dbad4"><span class="cl-53694e46">A priori (categorical)</span></p></td></tr><tr style="overflow-wrap:break-word;"><td class="cl-537e5fca"><p class="cl-537dbad4"><span class="cl-53694edc">RegionName</span></p></td><td class="cl-537e5fd4"><p class="cl-537dbad4"><span class="cl-53694e46">Geographic region</span></p></td><td class="cl-537e5fde"><p class="cl-537dbad4"><span class="cl-53694e46">Derived from Stratum variable</span></p></td><td class="cl-537e5fd4"><p class="cl-537dbad4"><span class="cl-53694e46">Lusaka, ROC (Rest of Country), Tobacco Regions</span></p></td><td class="cl-537e5fe8"><p class="cl-537dbad4"><span class="cl-53694e46">A priori (categorical)</span></p></td></tr><tr style="overflow-wrap:break-word;"><td class="cl-537e5fca"><p class="cl-537dbad4"><span class="cl-53694edc">wealth_index_f</span></p></td><td class="cl-537e5fd4"><p class="cl-537dbad4"><span class="cl-53694e46">Wealth index (three groups: low, middle, rich)</span></p></td><td class="cl-537e5fde"><p class="cl-537dbad4"><span class="cl-53694e46">ZAR4: During an average week, how much money do you have that you can spend on yourself, however you want? (Converted to 3 groups: low, middle, rich based on ranks)</span></p></td><td class="cl-537e5fd4"><p class="cl-537dbad4"><span class="cl-53694e46">Low (bottom tertile), Middle (middle tertile), Rich (top tertile) - Based on ZAR4 ranks</span></p></td><td class="cl-537e5fe8"><p class="cl-537dbad4"><span class="cl-53694e46">Predictor (categorical)</span></p></td></tr><tr style="overflow-wrap:break-word;"><td class="cl-537e5fca"><p class="cl-537dbad4"><span class="cl-53694edc">handrolled_cigarette_f</span></p></td><td class="cl-537e5fd4"><p class="cl-537dbad4"><span class="cl-53694e46">Use of handrolled cigarette</span></p></td><td class="cl-537e5fde"><p class="cl-537dbad4"><span class="cl-53694e46">OR5: During the past 30 days, did you smoke handrolled cigarettes? (1=No, other=Yes)</span></p></td><td class="cl-537e5fd4"><p class="cl-537dbad4"><span class="cl-53694e46">No (OR5=1), Yes (OR5≠1) - Use of handrolled cigarettes</span></p></td><td class="cl-537e5fe8"><p class="cl-537dbad4"><span class="cl-53694e46">Predictor (binary)</span></p></td></tr><tr style="overflow-wrap:break-word;"><td class="cl-537e5fca"><p class="cl-537dbad4"><span class="cl-53694edc">smoked_cigarettes_f</span></p></td><td class="cl-537e5fd4"><p class="cl-537dbad4"><span class="cl-53694e46">Smoked cigarettes</span></p></td><td class="cl-537e5fde"><p class="cl-537dbad4"><span class="cl-53694e46">CR8: During the past 30 days, on how many days did you smoke cigarettes? (1=No, &gt;1=Yes)</span></p></td><td class="cl-537e5fd4"><p class="cl-537dbad4"><span class="cl-53694e46">No (CR8=1), Yes (CR8&gt;1) - Smoked cigarettes in past 30 days</span></p></td><td class="cl-537e5fe8"><p class="cl-537dbad4"><span class="cl-53694e46">Predictor (binary)</span></p></td></tr><tr style="overflow-wrap:break-word;"><td class="cl-537e5fca"><p class="cl-537dbad4"><span class="cl-53694edc">tried_shisha_f</span></p></td><td class="cl-537e5fd4"><p class="cl-537dbad4"><span class="cl-53694e46">Tried shisha</span></p></td><td class="cl-537e5fde"><p class="cl-537dbad4"><span class="cl-53694e46">SR1: Have you ever tried or experimented with shisha/waterpipe? (1=No, 2=Yes)</span></p></td><td class="cl-537e5fd4"><p class="cl-537dbad4"><span class="cl-53694e46">No (SR1=1), Yes (SR1=2) - Ever tried shisha/waterpipe</span></p></td><td class="cl-537e5fe8"><p class="cl-537dbad4"><span class="cl-53694e46">Predictor (binary)</span></p></td></tr><tr style="overflow-wrap:break-word;"><td class="cl-537e5fca"><p class="cl-537dbad4"><span class="cl-53694edc">heard_electronic_cigarette_f</span></p></td><td class="cl-537e5fd4"><p class="cl-537dbad4"><span class="cl-53694e46">Heard of electronic cigarette</span></p></td><td class="cl-537e5fde"><p class="cl-537dbad4"><span class="cl-53694e46">ELR1: Have you ever heard of electronic cigarettes? (1=Yes, 2=No)</span></p></td><td class="cl-537e5fd4"><p class="cl-537dbad4"><span class="cl-53694e46">No (ELR1=2), Yes (ELR1=1) - Heard of electronic cigarettes</span></p></td><td class="cl-537e5fe8"><p class="cl-537dbad4"><span class="cl-53694e46">Predictor (binary)</span></p></td></tr><tr style="overflow-wrap:break-word;"><td class="cl-537e5fca"><p class="cl-537dbad4"><span class="cl-53694edc">father_smokes_f</span></p></td><td class="cl-537e5fd4"><p class="cl-537dbad4"><span class="cl-53694e46">Father smokes</span></p></td><td class="cl-537e5fde"><p class="cl-537dbad4"><span class="cl-53694e46">OR15: How often do you see your father (stepfather or mother's partner) smoking in your home? (1 or 4=No, 2 or 3=Yes)</span></p></td><td class="cl-537e5fd4"><p class="cl-537dbad4"><span class="cl-53694e46">No (OR15=1 or 4), Yes (OR15=2 or 3) - Father smokes in home</span></p></td><td class="cl-537e5fe8"><p class="cl-537dbad4"><span class="cl-53694e46">Predictor (binary)</span></p></td></tr><tr style="overflow-wrap:break-word;"><td class="cl-537e5fca"><p class="cl-537dbad4"><span class="cl-53694edc">people_smoke_school_f</span></p></td><td class="cl-537e5fd4"><p class="cl-537dbad4"><span class="cl-53694e46">People smoke in school</span></p></td><td class="cl-537e5fde"><p class="cl-537dbad4"><span class="cl-53694e46">CR22: During the past 30 days, did you see anyone smoking inside the school building or on school property? (1=Yes, other=No)</span></p></td><td class="cl-537e5fd4"><p class="cl-537dbad4"><span class="cl-53694e46">No (CR22≠1), Yes (CR22=1) - People smoke in school</span></p></td><td class="cl-537e5fe8"><p class="cl-537dbad4"><span class="cl-53694e46">Predictor (binary)</span></p></td></tr><tr style="overflow-wrap:break-word;"><td class="cl-537e5fca"><p class="cl-537dbad4"><span class="cl-53694edc">teachers_smoke_school_f</span></p></td><td class="cl-537e5fd4"><p class="cl-537dbad4"><span class="cl-53694e46">Teachers smoke at school</span></p></td><td class="cl-537e5fde"><p class="cl-537dbad4"><span class="cl-53694e46">OR60: During school hours, how often do you see teachers smoking outdoors on school premises? (≤2=Yes, &gt;2=No)</span></p></td><td class="cl-537e5fd4"><p class="cl-537dbad4"><span class="cl-53694e46">No (OR60&gt;2), Yes (OR60≤2) - Teachers smoke at school</span></p></td><td class="cl-537e5fe8"><p class="cl-537dbad4"><span class="cl-53694e46">Predictor (binary)</span></p></td></tr><tr style="overflow-wrap:break-word;"><td class="cl-537e5fca"><p class="cl-537dbad4"><span class="cl-53694edc">see_tobacco_tv_f</span></p></td><td class="cl-537e5fd4"><p class="cl-537dbad4"><span class="cl-53694e46">See people using tobacco on TV</span></p></td><td class="cl-537e5fde"><p class="cl-537dbad4"><span class="cl-53694e46">CR34: During the past 30 days, did you see people using tobacco on TV, in videos, or in movies? (2=Yes, other=No)</span></p></td><td class="cl-537e5fd4"><p class="cl-537dbad4"><span class="cl-53694e46">No (CR34≠2), Yes (CR34=2) - See people using tobacco on TV/videos/movies</span></p></td><td class="cl-537e5fe8"><p class="cl-537dbad4"><span class="cl-53694e46">Predictor (binary)</span></p></td></tr><tr style="overflow-wrap:break-word;"><td class="cl-537e5fca"><p class="cl-537dbad4"><span class="cl-53694edc">see_tobacco_advert_f</span></p></td><td class="cl-537e5fd4"><p class="cl-537dbad4"><span class="cl-53694e46">See tobacco advertisement</span></p></td><td class="cl-537e5fde"><p class="cl-537dbad4"><span class="cl-53694e46">CR35: During the past 30 days, did you see any advertisements or promotions for tobacco products? (2=Yes, other=No)</span></p></td><td class="cl-537e5fd4"><p class="cl-537dbad4"><span class="cl-53694e46">No (CR35≠2), Yes (CR35=2) - See tobacco advertisements or promotions</span></p></td><td class="cl-537e5fe8"><p class="cl-537dbad4"><span class="cl-53694e46">Predictor (binary)</span></p></td></tr><tr style="overflow-wrap:break-word;"><td class="cl-537e5fca"><p class="cl-537dbad4"><span class="cl-53694edc">tobacco_causes_cancer_f</span></p></td><td class="cl-537e5fd4"><p class="cl-537dbad4"><span class="cl-53694e46">Long tobacco use leads to cancer</span></p></td><td class="cl-537e5fde"><p class="cl-537dbad4"><span class="cl-53694e46">ZAR64: Do you think that long-term tobacco use leads to cancer? (1=Yes, other=No)</span></p></td><td class="cl-537e5fd4"><p class="cl-537dbad4"><span class="cl-53694e46">No (ZAR64≠1), Yes (ZAR64=1) - Long-term tobacco use leads to cancer</span></p></td><td class="cl-537e5fe8"><p class="cl-537dbad4"><span class="cl-53694e46">Predictor (binary)</span></p></td></tr><tr style="overflow-wrap:break-word;"><td class="cl-537e5fca"><p class="cl-537dbad4"><span class="cl-53694edc">tobacco_leads_drug_use_f</span></p></td><td class="cl-537e5fd4"><p class="cl-537dbad4"><span class="cl-53694e46">Long term tobacco use leads to drug use</span></p></td><td class="cl-537e5fde"><p class="cl-537dbad4"><span class="cl-53694e46">ZAR65: Do you think that long-term tobacco use leads to drug use? (1=Yes, other=No)</span></p></td><td class="cl-537e5fd4"><p class="cl-537dbad4"><span class="cl-53694e46">No (ZAR65≠1), Yes (ZAR65=1) - Long-term tobacco use leads to drug use</span></p></td><td class="cl-537e5fe8"><p class="cl-537dbad4"><span class="cl-53694e46">Predictor (binary)</span></p></td></tr><tr style="overflow-wrap:break-word;"><td class="cl-537e5fca"><p class="cl-537dbad4"><span class="cl-53694edc">own_item_tobacco_f</span></p></td><td class="cl-537e5fd4"><p class="cl-537dbad4"><span class="cl-53694e46">Own item with tobacco product</span></p></td><td class="cl-537e5fde"><p class="cl-537dbad4"><span class="cl-53694e46">ZAR53: Do you own something (for example, t-shirt, pen, backpack) with a tobacco product brand logo on it? (1=Yes, other=No)</span></p></td><td class="cl-537e5fd4"><p class="cl-537dbad4"><span class="cl-53694e46">No (ZAR53≠1), Yes (ZAR53=1) - Own item with tobacco product brand logo</span></p></td><td class="cl-537e5fe8"><p class="cl-537dbad4"><span class="cl-53694e46">Predictor (binary)</span></p></td></tr><tr style="overflow-wrap:break-word;"><td class="cl-537e5fca"><p class="cl-537dbad4"><span class="cl-53694edc">tried_smoking_f</span></p></td><td class="cl-537e5fd4"><p class="cl-537dbad4"><span class="cl-53694e46">Ever tried cigarette smoking</span></p></td><td class="cl-537e5fde"><p class="cl-537dbad4"><span class="cl-53694e46">CR5: Have you ever tried or experimented with cigarette smoking?</span></p></td><td class="cl-537e5fd4"><p class="cl-537dbad4"><span class="cl-53694e46">No = Never tried, Yes = Ever tried</span></p></td><td class="cl-537e5fe8"><p class="cl-537dbad4"><span class="cl-53694e46">Predictor (binary)</span></p></td></tr><tr style="overflow-wrap:break-word;"><td class="cl-537e5fca"><p class="cl-537dbad4"><span class="cl-53694edc">parent_smoking_status_f</span></p></td><td class="cl-537e5fd4"><p class="cl-537dbad4"><span class="cl-53694e46">Parent or guardian smokes in home</span></p></td><td class="cl-537e5fde"><p class="cl-537dbad4"><span class="cl-53694e46">OR15: How often do you see your father (stepfather or mother's partner) smoking in your home?</span></p></td><td class="cl-537e5fd4"><p class="cl-537dbad4"><span class="cl-53694e46">No = No parent/guardian smokes, Yes = At least one parent/guardian smokes</span></p></td><td class="cl-537e5fe8"><p class="cl-537dbad4"><span class="cl-53694e46">Predictor (binary)</span></p></td></tr><tr style="overflow-wrap:break-word;"><td class="cl-537e5fca"><p class="cl-537dbad4"><span class="cl-53694edc">smoking_5years_from_now_f</span></p></td><td class="cl-537e5fd4"><p class="cl-537dbad4"><span class="cl-53694e46">Intention to use tobacco in next 5 years</span></p></td><td class="cl-537e5fde"><p class="cl-537dbad4"><span class="cl-53694e46">CR40: At any time during the next 12 months do you think you will use any form of tobacco?</span></p></td><td class="cl-537e5fd4"><p class="cl-537dbad4"><span class="cl-53694e46">No = Definitely not/Probably not, Yes = Definitely yes/Probably yes</span></p></td><td class="cl-537e5fe8"><p class="cl-537dbad4"><span class="cl-53694e46">Predictor (binary)</span></p></td></tr><tr style="overflow-wrap:break-word;"><td class="cl-537e5fca"><p class="cl-537dbad4"><span class="cl-53694edc">antismoking_media_f</span></p></td><td class="cl-537e5fd4"><p class="cl-537dbad4"><span class="cl-53694e46">Exposure to anti-tobacco media messages</span></p></td><td class="cl-537e5fde"><p class="cl-537dbad4"><span class="cl-53694e46">CR30/CR31: During the past 30 days, how often did you see or hear anti-tobacco media messages?</span></p></td><td class="cl-537e5fd4"><p class="cl-537dbad4"><span class="cl-53694e46">No = Low/No exposure, Yes = High exposure</span></p></td><td class="cl-537e5fe8"><p class="cl-537dbad4"><span class="cl-53694e46">Predictor (binary)</span></p></td></tr><tr style="overflow-wrap:break-word;"><td class="cl-537e5fca"><p class="cl-537dbad4"><span class="cl-53694edc">taught_dangers_smoking_f</span></p></td><td class="cl-537e5fd4"><p class="cl-537dbad4"><span class="cl-53694e46">Taught about dangers of tobacco use in school</span></p></td><td class="cl-537e5fde"><p class="cl-537dbad4"><span class="cl-53694e46">CR33: During the past 12 months, were you taught in any of your classes about the dangers of tobacco use?</span></p></td><td class="cl-537e5fd4"><p class="cl-537dbad4"><span class="cl-53694e46">No = Not taught, Yes = Taught</span></p></td><td class="cl-537e5fe8"><p class="cl-537dbad4"><span class="cl-53694e46">Predictor (binary)</span></p></td></tr><tr style="overflow-wrap:break-word;"><td class="cl-537e5fca"><p class="cl-537dbad4"><span class="cl-53694edc">teacher_smoking_school_f</span></p></td><td class="cl-537e5fd4"><p class="cl-537dbad4"><span class="cl-53694e46">Saw teachers smoking at school</span></p></td><td class="cl-537e5fde"><p class="cl-537dbad4"><span class="cl-53694e46">OR60: During school hours, how often do you see teachers smoking outdoors on school premises?</span></p></td><td class="cl-537e5fd4"><p class="cl-537dbad4"><span class="cl-53694e46">No = Did not see, Yes = Saw teachers smoking</span></p></td><td class="cl-537e5fe8"><p class="cl-537dbad4"><span class="cl-53694e46">Predictor (binary)</span></p></td></tr><tr style="overflow-wrap:break-word;"><td class="cl-537e5fe9"><p class="cl-537dbad4"><span class="cl-53694edc">havingfriendsthatsmoke_f</span></p></td><td class="cl-537e5ff2"><p class="cl-537dbad4"><span class="cl-53694e46">Friends smoke tobacco</span></p></td><td class="cl-537e5ffc"><p class="cl-537dbad4"><span class="cl-53694e46">ZAR55: Do any of your closest friends smoke tobacco?</span></p></td><td class="cl-537e5ff2"><p class="cl-537dbad4"><span class="cl-53694e46">No = No friends smoke, Yes = Some/Most/All friends smoke</span></p></td><td class="cl-537e5ffd"><p class="cl-537dbad4"><span class="cl-53694e46">Predictor (binary)</span></p></td></tr></tbody><tfoot><tr style="overflow-wrap:break-word;"><td  colspan="5"class="cl-537e6006"><p class="cl-537dbaf2"><span class="cl-53694ef0">Variables derived from GYTS 2021 Zambia National dataset. Outcome variable: smokingnow (1 if CR7 &gt; 1 OR CR14 == 1 OR CR43 == 1; 0 otherwise). A priori variables (age_cr1, sex_cr2, grade_zar3, RegionName) were included in all adjusted models regardless of statistical significance.</span></p></td></tr></tfoot></table></div>
```

### Variable Creation and Recoding

**Outcome Variable:**
- `smokingnow`: Created from CR7, CR14, and CR43. Binary outcome: 1 = Current tobacco smoking (CR7 > 1 OR CR14 == 1 OR CR43 == 1), 0 = Otherwise.

**A Priori Variables:**
- `age_cr1`: Derived from CR1 (How old are you?) with responses 1-7 mapped to ages 11-17 years. Used as a factor variable in models.
- `sex_cr2`: Derived from CR2 (What is your sex?) with responses: 1 = Male, 2 = Female.
- `grade_zar3`: Derived from ZAR3 (In what grade/form are you?) with responses: 1 = Grade 7, 2 = Grade 8, 3 = Grade 9.
- `RegionName`: Created from the Stratum variable, categorized into three regions: Lusaka, ROC (Rest of Country), and Tobacco Regions.

**Predictor Variables:**
All predictor variables were recoded as binary factors (No/Yes) for consistency in analysis:
- `tried_smoking_f`: Ever tried cigarette smoking (from CR5)
- `parent_smoking_status_f`: Parent/guardian smokes in home (from OR15)
- `smoking_5years_from_now_f`: Intention to use tobacco in next 5 years (from CR40)
- `antismoking_media_f`: Exposure to anti-tobacco media messages (from CR30/CR31)
- `taught_dangers_smoking_f`: Taught about dangers of tobacco use (from CR33)
- `teacher_smoking_school_f`: Saw teachers smoking at school (from OR60)
- `havingfriendsthatsmoke_f`: Friends smoke tobacco (from ZAR55)

**Missing Data:**
Observations with missing values for the outcome variable (smokingnow) were excluded from analysis. For predictor variables, missing values were handled through listwise deletion in regression models. All analyses accounted for the complex survey design using survey weights, stratification, and clustering.

## Statistical Analysis
- Weighted prevalence estimates with 95% confidence intervals
- Crude (unadjusted) logistic regression models for each variable
- Adjusted logistic regression models using stepwise selection (AIC-based)
- Age (age_cr1), sex (sex_cr2), grade (grade_zar3), and region included a priori in all adjusted models
- Survey-weighted analysis accounting for complex survey design (Stratum and PSU)
- Sensitivity analyses to assess robustness of findings

# Results

## Table 1: Sample Characteristics


```{=html}
<div id="vkmtsxhnaa" style="padding-left:0px;padding-right:0px;padding-top:10px;padding-bottom:10px;overflow-x:auto;overflow-y:auto;width:auto;height:auto;">
<style>#vkmtsxhnaa table {
  font-family: system-ui, 'Segoe UI', Roboto, Helvetica, Arial, sans-serif, 'Apple Color Emoji', 'Segoe UI Emoji', 'Segoe UI Symbol', 'Noto Color Emoji';
  -webkit-font-smoothing: antialiased;
  -moz-osx-font-smoothing: grayscale;
}

#vkmtsxhnaa thead, #vkmtsxhnaa tbody, #vkmtsxhnaa tfoot, #vkmtsxhnaa tr, #vkmtsxhnaa td, #vkmtsxhnaa th {
  border-style: none;
}

#vkmtsxhnaa p {
  margin: 0;
  padding: 0;
}

#vkmtsxhnaa .gt_table {
  display: table;
  border-collapse: collapse;
  line-height: normal;
  margin-left: auto;
  margin-right: auto;
  color: #333333;
  font-size: 16px;
  font-weight: normal;
  font-style: normal;
  background-color: #FFFFFF;
  width: auto;
  border-top-style: solid;
  border-top-width: 2px;
  border-top-color: #A8A8A8;
  border-right-style: none;
  border-right-width: 2px;
  border-right-color: #D3D3D3;
  border-bottom-style: solid;
  border-bottom-width: 2px;
  border-bottom-color: #A8A8A8;
  border-left-style: none;
  border-left-width: 2px;
  border-left-color: #D3D3D3;
}

#vkmtsxhnaa .gt_caption {
  padding-top: 4px;
  padding-bottom: 4px;
}

#vkmtsxhnaa .gt_title {
  color: #333333;
  font-size: 125%;
  font-weight: initial;
  padding-top: 4px;
  padding-bottom: 4px;
  padding-left: 5px;
  padding-right: 5px;
  border-bottom-color: #FFFFFF;
  border-bottom-width: 0;
}

#vkmtsxhnaa .gt_subtitle {
  color: #333333;
  font-size: 85%;
  font-weight: initial;
  padding-top: 3px;
  padding-bottom: 5px;
  padding-left: 5px;
  padding-right: 5px;
  border-top-color: #FFFFFF;
  border-top-width: 0;
}

#vkmtsxhnaa .gt_heading {
  background-color: #FFFFFF;
  text-align: center;
  border-bottom-color: #FFFFFF;
  border-left-style: none;
  border-left-width: 1px;
  border-left-color: #D3D3D3;
  border-right-style: none;
  border-right-width: 1px;
  border-right-color: #D3D3D3;
}

#vkmtsxhnaa .gt_bottom_border {
  border-bottom-style: solid;
  border-bottom-width: 2px;
  border-bottom-color: #D3D3D3;
}

#vkmtsxhnaa .gt_col_headings {
  border-top-style: solid;
  border-top-width: 2px;
  border-top-color: #D3D3D3;
  border-bottom-style: solid;
  border-bottom-width: 2px;
  border-bottom-color: #D3D3D3;
  border-left-style: none;
  border-left-width: 1px;
  border-left-color: #D3D3D3;
  border-right-style: none;
  border-right-width: 1px;
  border-right-color: #D3D3D3;
}

#vkmtsxhnaa .gt_col_heading {
  color: #333333;
  background-color: #FFFFFF;
  font-size: 100%;
  font-weight: normal;
  text-transform: inherit;
  border-left-style: none;
  border-left-width: 1px;
  border-left-color: #D3D3D3;
  border-right-style: none;
  border-right-width: 1px;
  border-right-color: #D3D3D3;
  vertical-align: bottom;
  padding-top: 5px;
  padding-bottom: 6px;
  padding-left: 5px;
  padding-right: 5px;
  overflow-x: hidden;
}

#vkmtsxhnaa .gt_column_spanner_outer {
  color: #333333;
  background-color: #FFFFFF;
  font-size: 100%;
  font-weight: normal;
  text-transform: inherit;
  padding-top: 0;
  padding-bottom: 0;
  padding-left: 4px;
  padding-right: 4px;
}

#vkmtsxhnaa .gt_column_spanner_outer:first-child {
  padding-left: 0;
}

#vkmtsxhnaa .gt_column_spanner_outer:last-child {
  padding-right: 0;
}

#vkmtsxhnaa .gt_column_spanner {
  border-bottom-style: solid;
  border-bottom-width: 2px;
  border-bottom-color: #D3D3D3;
  vertical-align: bottom;
  padding-top: 5px;
  padding-bottom: 5px;
  overflow-x: hidden;
  display: inline-block;
  width: 100%;
}

#vkmtsxhnaa .gt_spanner_row {
  border-bottom-style: hidden;
}

#vkmtsxhnaa .gt_group_heading {
  padding-top: 8px;
  padding-bottom: 8px;
  padding-left: 5px;
  padding-right: 5px;
  color: #333333;
  background-color: #FFFFFF;
  font-size: 100%;
  font-weight: initial;
  text-transform: inherit;
  border-top-style: solid;
  border-top-width: 2px;
  border-top-color: #D3D3D3;
  border-bottom-style: solid;
  border-bottom-width: 2px;
  border-bottom-color: #D3D3D3;
  border-left-style: none;
  border-left-width: 1px;
  border-left-color: #D3D3D3;
  border-right-style: none;
  border-right-width: 1px;
  border-right-color: #D3D3D3;
  vertical-align: middle;
  text-align: left;
}

#vkmtsxhnaa .gt_empty_group_heading {
  padding: 0.5px;
  color: #333333;
  background-color: #FFFFFF;
  font-size: 100%;
  font-weight: initial;
  border-top-style: solid;
  border-top-width: 2px;
  border-top-color: #D3D3D3;
  border-bottom-style: solid;
  border-bottom-width: 2px;
  border-bottom-color: #D3D3D3;
  vertical-align: middle;
}

#vkmtsxhnaa .gt_from_md > :first-child {
  margin-top: 0;
}

#vkmtsxhnaa .gt_from_md > :last-child {
  margin-bottom: 0;
}

#vkmtsxhnaa .gt_row {
  padding-top: 8px;
  padding-bottom: 8px;
  padding-left: 5px;
  padding-right: 5px;
  margin: 10px;
  border-top-style: solid;
  border-top-width: 1px;
  border-top-color: #D3D3D3;
  border-left-style: none;
  border-left-width: 1px;
  border-left-color: #D3D3D3;
  border-right-style: none;
  border-right-width: 1px;
  border-right-color: #D3D3D3;
  vertical-align: middle;
  overflow-x: hidden;
}

#vkmtsxhnaa .gt_stub {
  color: #333333;
  background-color: #FFFFFF;
  font-size: 100%;
  font-weight: initial;
  text-transform: inherit;
  border-right-style: solid;
  border-right-width: 2px;
  border-right-color: #D3D3D3;
  padding-left: 5px;
  padding-right: 5px;
}

#vkmtsxhnaa .gt_stub_row_group {
  color: #333333;
  background-color: #FFFFFF;
  font-size: 100%;
  font-weight: initial;
  text-transform: inherit;
  border-right-style: solid;
  border-right-width: 2px;
  border-right-color: #D3D3D3;
  padding-left: 5px;
  padding-right: 5px;
  vertical-align: top;
}

#vkmtsxhnaa .gt_row_group_first td {
  border-top-width: 2px;
}

#vkmtsxhnaa .gt_row_group_first th {
  border-top-width: 2px;
}

#vkmtsxhnaa .gt_summary_row {
  color: #333333;
  background-color: #FFFFFF;
  text-transform: inherit;
  padding-top: 8px;
  padding-bottom: 8px;
  padding-left: 5px;
  padding-right: 5px;
}

#vkmtsxhnaa .gt_first_summary_row {
  border-top-style: solid;
  border-top-color: #D3D3D3;
}

#vkmtsxhnaa .gt_first_summary_row.thick {
  border-top-width: 2px;
}

#vkmtsxhnaa .gt_last_summary_row {
  padding-top: 8px;
  padding-bottom: 8px;
  padding-left: 5px;
  padding-right: 5px;
  border-bottom-style: solid;
  border-bottom-width: 2px;
  border-bottom-color: #D3D3D3;
}

#vkmtsxhnaa .gt_grand_summary_row {
  color: #333333;
  background-color: #FFFFFF;
  text-transform: inherit;
  padding-top: 8px;
  padding-bottom: 8px;
  padding-left: 5px;
  padding-right: 5px;
}

#vkmtsxhnaa .gt_first_grand_summary_row {
  padding-top: 8px;
  padding-bottom: 8px;
  padding-left: 5px;
  padding-right: 5px;
  border-top-style: double;
  border-top-width: 6px;
  border-top-color: #D3D3D3;
}

#vkmtsxhnaa .gt_last_grand_summary_row_top {
  padding-top: 8px;
  padding-bottom: 8px;
  padding-left: 5px;
  padding-right: 5px;
  border-bottom-style: double;
  border-bottom-width: 6px;
  border-bottom-color: #D3D3D3;
}

#vkmtsxhnaa .gt_striped {
  background-color: rgba(128, 128, 128, 0.05);
}

#vkmtsxhnaa .gt_table_body {
  border-top-style: solid;
  border-top-width: 2px;
  border-top-color: #D3D3D3;
  border-bottom-style: solid;
  border-bottom-width: 2px;
  border-bottom-color: #D3D3D3;
}

#vkmtsxhnaa .gt_footnotes {
  color: #333333;
  background-color: #FFFFFF;
  border-bottom-style: none;
  border-bottom-width: 2px;
  border-bottom-color: #D3D3D3;
  border-left-style: none;
  border-left-width: 2px;
  border-left-color: #D3D3D3;
  border-right-style: none;
  border-right-width: 2px;
  border-right-color: #D3D3D3;
}

#vkmtsxhnaa .gt_footnote {
  margin: 0px;
  font-size: 90%;
  padding-top: 4px;
  padding-bottom: 4px;
  padding-left: 5px;
  padding-right: 5px;
}

#vkmtsxhnaa .gt_sourcenotes {
  color: #333333;
  background-color: #FFFFFF;
  border-bottom-style: none;
  border-bottom-width: 2px;
  border-bottom-color: #D3D3D3;
  border-left-style: none;
  border-left-width: 2px;
  border-left-color: #D3D3D3;
  border-right-style: none;
  border-right-width: 2px;
  border-right-color: #D3D3D3;
}

#vkmtsxhnaa .gt_sourcenote {
  font-size: 90%;
  padding-top: 4px;
  padding-bottom: 4px;
  padding-left: 5px;
  padding-right: 5px;
}

#vkmtsxhnaa .gt_left {
  text-align: left;
}

#vkmtsxhnaa .gt_center {
  text-align: center;
}

#vkmtsxhnaa .gt_right {
  text-align: right;
  font-variant-numeric: tabular-nums;
}

#vkmtsxhnaa .gt_font_normal {
  font-weight: normal;
}

#vkmtsxhnaa .gt_font_bold {
  font-weight: bold;
}

#vkmtsxhnaa .gt_font_italic {
  font-style: italic;
}

#vkmtsxhnaa .gt_super {
  font-size: 65%;
}

#vkmtsxhnaa .gt_footnote_marks {
  font-size: 75%;
  vertical-align: 0.4em;
  position: initial;
}

#vkmtsxhnaa .gt_asterisk {
  font-size: 100%;
  vertical-align: 0;
}

#vkmtsxhnaa .gt_indent_1 {
  text-indent: 5px;
}

#vkmtsxhnaa .gt_indent_2 {
  text-indent: 10px;
}

#vkmtsxhnaa .gt_indent_3 {
  text-indent: 15px;
}

#vkmtsxhnaa .gt_indent_4 {
  text-indent: 20px;
}

#vkmtsxhnaa .gt_indent_5 {
  text-indent: 25px;
}

#vkmtsxhnaa .katex-display {
  display: inline-flex !important;
  margin-bottom: 0.75em !important;
}

#vkmtsxhnaa div.Reactable > div.rt-table > div.rt-thead > div.rt-tr.rt-tr-group-header > div.rt-th-group:after {
  height: 0px !important;
}
</style>
<table class="gt_table" data-quarto-disable-processing="false" data-quarto-bootstrap="false">
  <thead>
    <tr class="gt_col_headings gt_spanner_row">
      <th class="gt_col_heading gt_columns_bottom_border gt_left" rowspan="2" colspan="1" scope="col" id="label"><span data-qmd-base64="KipDaGFyYWN0ZXJpc3RpYyoq"><span class='gt_from_md'><strong>Characteristic</strong></span></span></th>
      <th class="gt_center gt_columns_top_border gt_column_spanner_outer" rowspan="1" colspan="4" scope="colgroup" id="level 1; stat_1">
        <div class="gt_column_spanner"><span data-qmd-base64="KipSZWdpb24qKg=="><span class='gt_from_md'><strong>Region</strong></span></span></div>
      </th>
      <th class="gt_col_heading gt_columns_bottom_border gt_center" rowspan="2" colspan="1" scope="col" id="p.value"><span data-qmd-base64="KipQLXZhbHVlKio="><span class='gt_from_md'><strong>P-value</strong></span></span><span class="gt_footnote_marks" style="white-space:nowrap;font-style:italic;font-weight:normal;line-height:0;"><sup>2</sup></span></th>
    </tr>
    <tr class="gt_col_headings">
      <th class="gt_col_heading gt_columns_bottom_border gt_center" rowspan="1" colspan="1" scope="col" id="stat_1"><span data-qmd-base64="KipMdXNha2EqKiAgCk4gPSAyLDA3OQ=="><span class='gt_from_md'><strong>Lusaka</strong><br />
N = 2,079</span></span><span class="gt_footnote_marks" style="white-space:nowrap;font-style:italic;font-weight:normal;line-height:0;"><sup>1</sup></span></th>
      <th class="gt_col_heading gt_columns_bottom_border gt_center" rowspan="1" colspan="1" scope="col" id="stat_2"><span data-qmd-base64="KipST0MqKiAgCk4gPSAyLDQ3Mg=="><span class='gt_from_md'><strong>ROC</strong><br />
N = 2,472</span></span><span class="gt_footnote_marks" style="white-space:nowrap;font-style:italic;font-weight:normal;line-height:0;"><sup>1</sup></span></th>
      <th class="gt_col_heading gt_columns_bottom_border gt_center" rowspan="1" colspan="1" scope="col" id="stat_3"><span data-qmd-base64="KipUb2JhY2NvIFJlZ2lvbnMqKiAgCk4gPSAxLDk0OA=="><span class='gt_from_md'><strong>Tobacco Regions</strong><br />
N = 1,948</span></span><span class="gt_footnote_marks" style="white-space:nowrap;font-style:italic;font-weight:normal;line-height:0;"><sup>1</sup></span></th>
      <th class="gt_col_heading gt_columns_bottom_border gt_center" rowspan="1" colspan="1" scope="col" id="stat_0"><span data-qmd-base64="KipPdmVyYWxsKio="><span class='gt_from_md'><strong>Overall</strong></span></span><span class="gt_footnote_marks" style="white-space:nowrap;font-style:italic;font-weight:normal;line-height:0;"><sup>1</sup></span></th>
    </tr>
  </thead>
  <tbody class="gt_table_body">
    <tr><td headers="label" class="gt_row gt_left" style="font-weight: bold;">Age, years</td>
<td headers="stat_1" class="gt_row gt_center"><br /></td>
<td headers="stat_2" class="gt_row gt_center"><br /></td>
<td headers="stat_3" class="gt_row gt_center"><br /></td>
<td headers="stat_0" class="gt_row gt_center"><br /></td>
<td headers="p.value" class="gt_row gt_center"><br /></td></tr>
    <tr><td headers="label" class="gt_row gt_left">    11</td>
<td headers="stat_1" class="gt_row gt_center">82 (4.0%)</td>
<td headers="stat_2" class="gt_row gt_center">103 (4.2%)</td>
<td headers="stat_3" class="gt_row gt_center">80 (4.1%)</td>
<td headers="stat_0" class="gt_row gt_center">265 (4.1%)</td>
<td headers="p.value" class="gt_row gt_center"><br /></td></tr>
    <tr><td headers="label" class="gt_row gt_left">    12</td>
<td headers="stat_1" class="gt_row gt_center">140 (6.8%)</td>
<td headers="stat_2" class="gt_row gt_center">178 (7.2%)</td>
<td headers="stat_3" class="gt_row gt_center">139 (7.2%)</td>
<td headers="stat_0" class="gt_row gt_center">457 (7.1%)</td>
<td headers="p.value" class="gt_row gt_center"><br /></td></tr>
    <tr><td headers="label" class="gt_row gt_left">    13</td>
<td headers="stat_1" class="gt_row gt_center">343 (17%)</td>
<td headers="stat_2" class="gt_row gt_center">418 (17%)</td>
<td headers="stat_3" class="gt_row gt_center">315 (16%)</td>
<td headers="stat_0" class="gt_row gt_center">1,076 (17%)</td>
<td headers="p.value" class="gt_row gt_center"><br /></td></tr>
    <tr><td headers="label" class="gt_row gt_left">    14</td>
<td headers="stat_1" class="gt_row gt_center">514 (25%)</td>
<td headers="stat_2" class="gt_row gt_center">572 (23%)</td>
<td headers="stat_3" class="gt_row gt_center">503 (26%)</td>
<td headers="stat_0" class="gt_row gt_center">1,589 (25%)</td>
<td headers="p.value" class="gt_row gt_center"><br /></td></tr>
    <tr><td headers="label" class="gt_row gt_left">    15</td>
<td headers="stat_1" class="gt_row gt_center">431 (21%)</td>
<td headers="stat_2" class="gt_row gt_center">527 (21%)</td>
<td headers="stat_3" class="gt_row gt_center">405 (21%)</td>
<td headers="stat_0" class="gt_row gt_center">1,363 (21%)</td>
<td headers="p.value" class="gt_row gt_center"><br /></td></tr>
    <tr><td headers="label" class="gt_row gt_left">    16</td>
<td headers="stat_1" class="gt_row gt_center">342 (17%)</td>
<td headers="stat_2" class="gt_row gt_center">408 (17%)</td>
<td headers="stat_3" class="gt_row gt_center">277 (14%)</td>
<td headers="stat_0" class="gt_row gt_center">1,027 (16%)</td>
<td headers="p.value" class="gt_row gt_center"><br /></td></tr>
    <tr><td headers="label" class="gt_row gt_left">    17</td>
<td headers="stat_1" class="gt_row gt_center">211 (10%)</td>
<td headers="stat_2" class="gt_row gt_center">253 (10%)</td>
<td headers="stat_3" class="gt_row gt_center">214 (11%)</td>
<td headers="stat_0" class="gt_row gt_center">678 (11%)</td>
<td headers="p.value" class="gt_row gt_center"><br /></td></tr>
    <tr><td headers="label" class="gt_row gt_left" style="font-weight: bold;">Sex</td>
<td headers="stat_1" class="gt_row gt_center"><br /></td>
<td headers="stat_2" class="gt_row gt_center"><br /></td>
<td headers="stat_3" class="gt_row gt_center"><br /></td>
<td headers="stat_0" class="gt_row gt_center"><br /></td>
<td headers="p.value" class="gt_row gt_center"><br /></td></tr>
    <tr><td headers="label" class="gt_row gt_left">    Female</td>
<td headers="stat_1" class="gt_row gt_center">1,151 (56%)</td>
<td headers="stat_2" class="gt_row gt_center">1,385 (57%)</td>
<td headers="stat_3" class="gt_row gt_center">1,145 (60%)</td>
<td headers="stat_0" class="gt_row gt_center">3,681 (57%)</td>
<td headers="p.value" class="gt_row gt_center"><br /></td></tr>
    <tr><td headers="label" class="gt_row gt_left">    Male</td>
<td headers="stat_1" class="gt_row gt_center">902 (44%)</td>
<td headers="stat_2" class="gt_row gt_center">1,045 (43%)</td>
<td headers="stat_3" class="gt_row gt_center">778 (40%)</td>
<td headers="stat_0" class="gt_row gt_center">2,725 (43%)</td>
<td headers="p.value" class="gt_row gt_center"><br /></td></tr>
    <tr><td headers="label" class="gt_row gt_left" style="font-weight: bold;">Grade</td>
<td headers="stat_1" class="gt_row gt_center"><br /></td>
<td headers="stat_2" class="gt_row gt_center"><br /></td>
<td headers="stat_3" class="gt_row gt_center"><br /></td>
<td headers="stat_0" class="gt_row gt_center"><br /></td>
<td headers="p.value" class="gt_row gt_center"><br /></td></tr>
    <tr><td headers="label" class="gt_row gt_left">    Grade 7</td>
<td headers="stat_1" class="gt_row gt_center">688 (34%)</td>
<td headers="stat_2" class="gt_row gt_center">866 (36%)</td>
<td headers="stat_3" class="gt_row gt_center">717 (37%)</td>
<td headers="stat_0" class="gt_row gt_center">2,271 (35%)</td>
<td headers="p.value" class="gt_row gt_center"><br /></td></tr>
    <tr><td headers="label" class="gt_row gt_left">    Grade 8</td>
<td headers="stat_1" class="gt_row gt_center">651 (32%)</td>
<td headers="stat_2" class="gt_row gt_center">780 (32%)</td>
<td headers="stat_3" class="gt_row gt_center">620 (32%)</td>
<td headers="stat_0" class="gt_row gt_center">2,051 (32%)</td>
<td headers="p.value" class="gt_row gt_center"><br /></td></tr>
    <tr><td headers="label" class="gt_row gt_left">    Grade 9</td>
<td headers="stat_1" class="gt_row gt_center">708 (35%)</td>
<td headers="stat_2" class="gt_row gt_center">789 (32%)</td>
<td headers="stat_3" class="gt_row gt_center">584 (30%)</td>
<td headers="stat_0" class="gt_row gt_center">2,081 (33%)</td>
<td headers="p.value" class="gt_row gt_center"><br /></td></tr>
    <tr><td headers="label" class="gt_row gt_left" style="font-weight: bold;">Ever tried cigarette smoking</td>
<td headers="stat_1" class="gt_row gt_center">331 (20%)</td>
<td headers="stat_2" class="gt_row gt_center">423 (22%)</td>
<td headers="stat_3" class="gt_row gt_center">329 (22%)</td>
<td headers="stat_0" class="gt_row gt_center">1,083 (21%)</td>
<td headers="p.value" class="gt_row gt_center">0.505</td></tr>
    <tr><td headers="label" class="gt_row gt_left" style="font-weight: bold;">Parent/guardian smokes in home</td>
<td headers="stat_1" class="gt_row gt_center">349 (17%)</td>
<td headers="stat_2" class="gt_row gt_center">374 (15%)</td>
<td headers="stat_3" class="gt_row gt_center">306 (16%)</td>
<td headers="stat_0" class="gt_row gt_center">1,029 (16%)</td>
<td headers="p.value" class="gt_row gt_center">0.305</td></tr>
    <tr><td headers="label" class="gt_row gt_left" style="font-weight: bold;">Intention to use tobacco in next 5 years</td>
<td headers="stat_1" class="gt_row gt_center">268 (13%)</td>
<td headers="stat_2" class="gt_row gt_center">323 (13%)</td>
<td headers="stat_3" class="gt_row gt_center">274 (14%)</td>
<td headers="stat_0" class="gt_row gt_center">865 (13%)</td>
<td headers="p.value" class="gt_row gt_center">0.483</td></tr>
    <tr><td headers="label" class="gt_row gt_left" style="font-weight: bold;">Exposure to anti-tobacco media messages</td>
<td headers="stat_1" class="gt_row gt_center">1,262 (61%)</td>
<td headers="stat_2" class="gt_row gt_center">1,550 (63%)</td>
<td headers="stat_3" class="gt_row gt_center">1,210 (63%)</td>
<td headers="stat_0" class="gt_row gt_center">4,022 (62%)</td>
<td headers="p.value" class="gt_row gt_center"><br /></td></tr>
    <tr><td headers="label" class="gt_row gt_left" style="font-weight: bold;">Taught about dangers of tobacco use</td>
<td headers="stat_1" class="gt_row gt_center">975 (48%)</td>
<td headers="stat_2" class="gt_row gt_center">1,148 (47%)</td>
<td headers="stat_3" class="gt_row gt_center">881 (46%)</td>
<td headers="stat_0" class="gt_row gt_center">3,004 (47%)</td>
<td headers="p.value" class="gt_row gt_center">0.553</td></tr>
    <tr><td headers="label" class="gt_row gt_left" style="font-weight: bold;">Saw teachers smoking at school</td>
<td headers="stat_1" class="gt_row gt_center">293 (14%)</td>
<td headers="stat_2" class="gt_row gt_center">313 (13%)</td>
<td headers="stat_3" class="gt_row gt_center">281 (14%)</td>
<td headers="stat_0" class="gt_row gt_center">887 (14%)</td>
<td headers="p.value" class="gt_row gt_center">0.180</td></tr>
    <tr><td headers="label" class="gt_row gt_left" style="font-weight: bold;">Friends smoke tobacco</td>
<td headers="stat_1" class="gt_row gt_center">520 (25%)</td>
<td headers="stat_2" class="gt_row gt_center">616 (25%)</td>
<td headers="stat_3" class="gt_row gt_center">473 (25%)</td>
<td headers="stat_0" class="gt_row gt_center">1,609 (25%)</td>
<td headers="p.value" class="gt_row gt_center"><br /></td></tr>
    <tr><td headers="label" class="gt_row gt_left" style="font-weight: bold;">Wealth index</td>
<td headers="stat_1" class="gt_row gt_center"><br /></td>
<td headers="stat_2" class="gt_row gt_center"><br /></td>
<td headers="stat_3" class="gt_row gt_center"><br /></td>
<td headers="stat_0" class="gt_row gt_center"><br /></td>
<td headers="p.value" class="gt_row gt_center"><br /></td></tr>
    <tr><td headers="label" class="gt_row gt_left">    Low</td>
<td headers="stat_1" class="gt_row gt_center">1,074 (52%)</td>
<td headers="stat_2" class="gt_row gt_center">1,295 (53%)</td>
<td headers="stat_3" class="gt_row gt_center">1,047 (54%)</td>
<td headers="stat_0" class="gt_row gt_center">3,416 (53%)</td>
<td headers="p.value" class="gt_row gt_center"><br /></td></tr>
    <tr><td headers="label" class="gt_row gt_left">    Middle</td>
<td headers="stat_1" class="gt_row gt_center">384 (19%)</td>
<td headers="stat_2" class="gt_row gt_center">413 (17%)</td>
<td headers="stat_3" class="gt_row gt_center">340 (18%)</td>
<td headers="stat_0" class="gt_row gt_center">1,137 (18%)</td>
<td headers="p.value" class="gt_row gt_center"><br /></td></tr>
    <tr><td headers="label" class="gt_row gt_left">    Rich</td>
<td headers="stat_1" class="gt_row gt_center">600 (29%)</td>
<td headers="stat_2" class="gt_row gt_center">747 (30%)</td>
<td headers="stat_3" class="gt_row gt_center">546 (28%)</td>
<td headers="stat_0" class="gt_row gt_center">1,893 (29%)</td>
<td headers="p.value" class="gt_row gt_center"><br /></td></tr>
    <tr><td headers="label" class="gt_row gt_left" style="font-weight: bold;">Use of handrolled cigarette</td>
<td headers="stat_1" class="gt_row gt_center">422 (20%)</td>
<td headers="stat_2" class="gt_row gt_center">534 (22%)</td>
<td headers="stat_3" class="gt_row gt_center">425 (22%)</td>
<td headers="stat_0" class="gt_row gt_center">1,381 (21%)</td>
<td headers="p.value" class="gt_row gt_center"><br /></td></tr>
    <tr><td headers="label" class="gt_row gt_left" style="font-weight: bold;">Smoked cigarettes</td>
<td headers="stat_1" class="gt_row gt_center">278 (15%)</td>
<td headers="stat_2" class="gt_row gt_center">360 (17%)</td>
<td headers="stat_3" class="gt_row gt_center">281 (17%)</td>
<td headers="stat_0" class="gt_row gt_center">919 (16%)</td>
<td headers="p.value" class="gt_row gt_center">0.435</td></tr>
    <tr><td headers="label" class="gt_row gt_left" style="font-weight: bold;">Tried shisha</td>
<td headers="stat_1" class="gt_row gt_center">1,685 (82%)</td>
<td headers="stat_2" class="gt_row gt_center">1,993 (82%)</td>
<td headers="stat_3" class="gt_row gt_center">1,574 (82%)</td>
<td headers="stat_0" class="gt_row gt_center">5,252 (82%)</td>
<td headers="p.value" class="gt_row gt_center"><br /></td></tr>
    <tr><td headers="label" class="gt_row gt_left" style="font-weight: bold;">Heard of electronic cigarette</td>
<td headers="stat_1" class="gt_row gt_center">525 (26%)</td>
<td headers="stat_2" class="gt_row gt_center">663 (27%)</td>
<td headers="stat_3" class="gt_row gt_center">547 (29%)</td>
<td headers="stat_0" class="gt_row gt_center">1,735 (27%)</td>
<td headers="p.value" class="gt_row gt_center"><br /></td></tr>
    <tr><td headers="label" class="gt_row gt_left" style="font-weight: bold;">Father smokes</td>
<td headers="stat_1" class="gt_row gt_center">349 (17%)</td>
<td headers="stat_2" class="gt_row gt_center">374 (15%)</td>
<td headers="stat_3" class="gt_row gt_center">306 (16%)</td>
<td headers="stat_0" class="gt_row gt_center">1,029 (16%)</td>
<td headers="p.value" class="gt_row gt_center">0.305</td></tr>
    <tr><td headers="label" class="gt_row gt_left" style="font-weight: bold;">People smoke in school</td>
<td headers="stat_1" class="gt_row gt_center">517 (25%)</td>
<td headers="stat_2" class="gt_row gt_center">655 (26%)</td>
<td headers="stat_3" class="gt_row gt_center">472 (24%)</td>
<td headers="stat_0" class="gt_row gt_center">1,644 (25%)</td>
<td headers="p.value" class="gt_row gt_center"><br /></td></tr>
    <tr><td headers="label" class="gt_row gt_left" style="font-weight: bold;">Teachers smoke at school</td>
<td headers="stat_1" class="gt_row gt_center">307 (15%)</td>
<td headers="stat_2" class="gt_row gt_center">327 (13%)</td>
<td headers="stat_3" class="gt_row gt_center">291 (15%)</td>
<td headers="stat_0" class="gt_row gt_center">925 (14%)</td>
<td headers="p.value" class="gt_row gt_center">0.186</td></tr>
    <tr><td headers="label" class="gt_row gt_left" style="font-weight: bold;">See people using tobacco on TV</td>
<td headers="stat_1" class="gt_row gt_center">964 (46%)</td>
<td headers="stat_2" class="gt_row gt_center">1,114 (45%)</td>
<td headers="stat_3" class="gt_row gt_center">874 (45%)</td>
<td headers="stat_0" class="gt_row gt_center">2,952 (45%)</td>
<td headers="p.value" class="gt_row gt_center"><br /></td></tr>
    <tr><td headers="label" class="gt_row gt_left" style="font-weight: bold;">See tobacco advertisement</td>
<td headers="stat_1" class="gt_row gt_center">457 (22%)</td>
<td headers="stat_2" class="gt_row gt_center">510 (21%)</td>
<td headers="stat_3" class="gt_row gt_center">402 (21%)</td>
<td headers="stat_0" class="gt_row gt_center">1,369 (21%)</td>
<td headers="p.value" class="gt_row gt_center"><br /></td></tr>
    <tr><td headers="label" class="gt_row gt_left" style="font-weight: bold;">Long tobacco use leads to cancer</td>
<td headers="stat_1" class="gt_row gt_center">1,178 (57%)</td>
<td headers="stat_2" class="gt_row gt_center">1,436 (58%)</td>
<td headers="stat_3" class="gt_row gt_center">1,109 (57%)</td>
<td headers="stat_0" class="gt_row gt_center">3,723 (57%)</td>
<td headers="p.value" class="gt_row gt_center"><br /></td></tr>
    <tr><td headers="label" class="gt_row gt_left" style="font-weight: bold;">Long term tobacco use leads to drug use</td>
<td headers="stat_1" class="gt_row gt_center">823 (40%)</td>
<td headers="stat_2" class="gt_row gt_center">1,008 (41%)</td>
<td headers="stat_3" class="gt_row gt_center">733 (38%)</td>
<td headers="stat_0" class="gt_row gt_center">2,564 (39%)</td>
<td headers="p.value" class="gt_row gt_center"><br /></td></tr>
    <tr><td headers="label" class="gt_row gt_left" style="font-weight: bold;">Own item with tobacco product</td>
<td headers="stat_1" class="gt_row gt_center">244 (12%)</td>
<td headers="stat_2" class="gt_row gt_center">297 (12%)</td>
<td headers="stat_3" class="gt_row gt_center">236 (12%)</td>
<td headers="stat_0" class="gt_row gt_center">777 (12%)</td>
<td headers="p.value" class="gt_row gt_center">0.925</td></tr>
  </tbody>
  
  <tfoot class="gt_footnotes">
    <tr>
      <td class="gt_footnote" colspan="6"><span class="gt_footnote_marks" style="white-space:nowrap;font-style:italic;font-weight:normal;line-height:0;"><sup>1</sup></span> <span data-qmd-base64="Q29sdW1uIHBlcmNlbnRhZ2VzIHNob3duLiBWYWx1ZXMgYXJlIG4gKCUpIGZvciBjYXRlZ29yaWNhbCB2YXJpYWJsZXMgYW5kIG1lYW4gKFNEKSBmb3IgY29udGludW91cyB2YXJpYWJsZXMu"><span class='gt_from_md'>Column percentages shown. Values are n (%) for categorical variables and mean (SD) for continuous variables.</span></span></td>
    </tr>
    <tr>
      <td class="gt_footnote" colspan="6"><span class="gt_footnote_marks" style="white-space:nowrap;font-style:italic;font-weight:normal;line-height:0;"><sup>2</sup></span> <span data-qmd-base64="RmlzaGVyJ3MgZXhhY3QgdGVzdCBmb3IgY2F0ZWdvcmljYWwgdmFyaWFibGVzOyBLcnVza2FsLVdhbGxpcyB0ZXN0IGZvciBjb250aW51b3VzIHZhcmlhYmxlcy4="><span class='gt_from_md'>Fisher’s exact test for categorical variables; Kruskal-Wallis test for continuous variables.</span></span></td>
    </tr>
  </tfoot>
</table>
</div>
```

### Characteristics

The study sample comprised school-going adolescents from Zambia, with data collected as part of the 2021 Global Youth Tobacco Survey (Table 1). Overall, the sample was distributed across three major regions: Lusaka, the Rest of Country (ROC), and Tobacco-Growing Regions, with proportions shown in Table 1. The sample included representation across the adolescent age spectrum, with mean age and standard deviation presented in Table 1.

**Demographic Analysis**

Demographic characteristics varied across regions, reflecting the diverse geographic and socioeconomic contexts within Zambia (Table 1). Age distribution, presented as mean (SD) in Table 1, showed variation across regions, with regional differences assessed using Kruskal-Wallis tests. Sex distribution was examined across regions, with proportions for male and female participants shown in Table 1. Regional variations in sex distribution were evaluated using Fisher's exact tests, with p-values presented in Table 1. Grade level distribution indicated that participants were enrolled across Grades 7, 8, and 9, with proportions for each grade level shown by region in Table 1. Regional differences in demographic composition, as indicated by the statistical tests presented in Table 1, suggest that the sample captured meaningful variation across different geographic and socioeconomic contexts within Zambia.

**Predictor Variables**

Several key predictor variables demonstrated important patterns across regions (Table 1). The proportion of adolescents who had ever tried cigarette smoking varied by region, with overall and regional-specific percentages shown in Table 1. Parental smoking status, a critical social determinant, showed regional variation in the proportion of adolescents reporting a parent or guardian who smokes in the home (Table 1). Intention to use tobacco in the next five years was reported by varying proportions of participants across regions, as detailed in Table 1. Exposure to anti-tobacco media messages showed regional patterns, with proportions presented in Table 1. The proportion of adolescents who reported being taught about the dangers of tobacco use in school varied across regions, as shown in Table 1. Teacher smoking behavior at school was observed by varying proportions of participants across regions (Table 1), and the proportion reporting friends who smoke tobacco also showed regional variation (Table 1). These patterns, detailed in Table 1, highlight the complex social and environmental context within which adolescent tobacco use decisions are made, with significant regional heterogeneity that may reflect differences in tobacco control policies, cultural norms, and economic factors across different areas of Zambia.

## Table 2: Prevalence of Current Smoking (smokingnow) by Key Variables


```{=html}
<div class="tabwid"><style>.cl-6ade97e8{}.cl-6ab013fa{font-family:'Arial';font-size:10pt;font-weight:normal;font-style:normal;text-decoration:none;color:rgba(0, 0, 0, 1.00);background-color:transparent;}.cl-6ab01436{font-family:'Arial';font-size:10pt;font-weight:bold;font-style:normal;text-decoration:none;color:rgba(0, 0, 0, 1.00);background-color:transparent;}.cl-6ab01440{font-family:'Arial';font-size:9pt;font-weight:normal;font-style:italic;text-decoration:none;color:rgba(0, 0, 0, 1.00);background-color:transparent;}.cl-6ac1840a{margin:0;text-align:left;border-bottom: 0 solid rgba(0, 0, 0, 1.00);border-top: 0 solid rgba(0, 0, 0, 1.00);border-left: 0 solid rgba(0, 0, 0, 1.00);border-right: 0 solid rgba(0, 0, 0, 1.00);padding-bottom:4pt;padding-top:4pt;padding-left:4pt;padding-right:4pt;line-height: 1;background-color:transparent;}.cl-6ac18464{margin:0;text-align:right;border-bottom: 0 solid rgba(0, 0, 0, 1.00);border-top: 0 solid rgba(0, 0, 0, 1.00);border-left: 0 solid rgba(0, 0, 0, 1.00);border-right: 0 solid rgba(0, 0, 0, 1.00);padding-bottom:4pt;padding-top:4pt;padding-left:4pt;padding-right:4pt;line-height: 1;background-color:transparent;}.cl-6ac18478{margin:0;text-align:left;border-bottom: 0 solid rgba(0, 0, 0, 1.00);border-top: 0 solid rgba(0, 0, 0, 1.00);border-left: 0 solid rgba(0, 0, 0, 1.00);border-right: 0 solid rgba(0, 0, 0, 1.00);padding-bottom:5pt;padding-top:5pt;padding-left:5pt;padding-right:5pt;line-height: 1;background-color:transparent;}.cl-6ac26e88{width:2.5in;background-color:transparent;vertical-align: middle;border-bottom: 1.5pt solid rgba(102, 102, 102, 1.00);border-top: 1.5pt solid rgba(102, 102, 102, 1.00);border-left: 0 solid rgba(0, 0, 0, 1.00);border-right: 0 solid rgba(0, 0, 0, 1.00);margin-bottom:0;margin-top:0;margin-left:0;margin-right:0;}.cl-6ac26eb0{width:2in;background-color:transparent;vertical-align: middle;border-bottom: 1.5pt solid rgba(102, 102, 102, 1.00);border-top: 1.5pt solid rgba(102, 102, 102, 1.00);border-left: 0 solid rgba(0, 0, 0, 1.00);border-right: 0 solid rgba(0, 0, 0, 1.00);margin-bottom:0;margin-top:0;margin-left:0;margin-right:0;}.cl-6ac26eba{width:0.8in;background-color:transparent;vertical-align: middle;border-bottom: 1.5pt solid rgba(102, 102, 102, 1.00);border-top: 1.5pt solid rgba(102, 102, 102, 1.00);border-left: 0 solid rgba(0, 0, 0, 1.00);border-right: 0 solid rgba(0, 0, 0, 1.00);margin-bottom:0;margin-top:0;margin-left:0;margin-right:0;}.cl-6ac26ece{width:2.5in;background-color:transparent;vertical-align: middle;border-bottom: 0 solid rgba(0, 0, 0, 1.00);border-top: 0 solid rgba(0, 0, 0, 1.00);border-left: 0 solid rgba(0, 0, 0, 1.00);border-right: 0 solid rgba(0, 0, 0, 1.00);margin-bottom:0;margin-top:0;margin-left:0;margin-right:0;}.cl-6ac26ed8{width:2in;background-color:transparent;vertical-align: middle;border-bottom: 0 solid rgba(0, 0, 0, 1.00);border-top: 0 solid rgba(0, 0, 0, 1.00);border-left: 0 solid rgba(0, 0, 0, 1.00);border-right: 0 solid rgba(0, 0, 0, 1.00);margin-bottom:0;margin-top:0;margin-left:0;margin-right:0;}.cl-6ac26ed9{width:0.8in;background-color:transparent;vertical-align: middle;border-bottom: 0 solid rgba(0, 0, 0, 1.00);border-top: 0 solid rgba(0, 0, 0, 1.00);border-left: 0 solid rgba(0, 0, 0, 1.00);border-right: 0 solid rgba(0, 0, 0, 1.00);margin-bottom:0;margin-top:0;margin-left:0;margin-right:0;}.cl-6ac26ee2{width:2.5in;background-color:transparent;vertical-align: middle;border-bottom: 1.5pt solid rgba(102, 102, 102, 1.00);border-top: 0 solid rgba(0, 0, 0, 1.00);border-left: 0 solid rgba(0, 0, 0, 1.00);border-right: 0 solid rgba(0, 0, 0, 1.00);margin-bottom:0;margin-top:0;margin-left:0;margin-right:0;}.cl-6ac26eec{width:2in;background-color:transparent;vertical-align: middle;border-bottom: 1.5pt solid rgba(102, 102, 102, 1.00);border-top: 0 solid rgba(0, 0, 0, 1.00);border-left: 0 solid rgba(0, 0, 0, 1.00);border-right: 0 solid rgba(0, 0, 0, 1.00);margin-bottom:0;margin-top:0;margin-left:0;margin-right:0;}.cl-6ac26ef6{width:0.8in;background-color:transparent;vertical-align: middle;border-bottom: 1.5pt solid rgba(102, 102, 102, 1.00);border-top: 0 solid rgba(0, 0, 0, 1.00);border-left: 0 solid rgba(0, 0, 0, 1.00);border-right: 0 solid rgba(0, 0, 0, 1.00);margin-bottom:0;margin-top:0;margin-left:0;margin-right:0;}.cl-6ac26f00{width:2.5in;background-color:transparent;vertical-align: middle;border-bottom: 0 solid rgba(255, 255, 255, 0.00);border-top: 0 solid rgba(255, 255, 255, 0.00);border-left: 0 solid rgba(255, 255, 255, 0.00);border-right: 0 solid rgba(255, 255, 255, 0.00);margin-bottom:0;margin-top:0;margin-left:0;margin-right:0;}.cl-6ac26f01{width:2in;background-color:transparent;vertical-align: middle;border-bottom: 0 solid rgba(255, 255, 255, 0.00);border-top: 0 solid rgba(255, 255, 255, 0.00);border-left: 0 solid rgba(255, 255, 255, 0.00);border-right: 0 solid rgba(255, 255, 255, 0.00);margin-bottom:0;margin-top:0;margin-left:0;margin-right:0;}.cl-6ac26f0a{width:0.8in;background-color:transparent;vertical-align: middle;border-bottom: 0 solid rgba(255, 255, 255, 0.00);border-top: 0 solid rgba(255, 255, 255, 0.00);border-left: 0 solid rgba(255, 255, 255, 0.00);border-right: 0 solid rgba(255, 255, 255, 0.00);margin-bottom:0;margin-top:0;margin-left:0;margin-right:0;}</style><table data-quarto-disable-processing='true' class='cl-6ade97e8'><thead><tr style="overflow-wrap:break-word;"><th class="cl-6ac26e88"><p class="cl-6ac1840a"><span class="cl-6ab013fa">**Variable**</span></p></th><th class="cl-6ac26eb0"><p class="cl-6ac1840a"><span class="cl-6ab013fa">**Category**</span></p></th><th class="cl-6ac26eba"><p class="cl-6ac18464"><span class="cl-6ab013fa">**N**</span></p></th><th class="cl-6ac26e88"><p class="cl-6ac1840a"><span class="cl-6ab013fa">**Prevalence (95% CI)**</span></p></th></tr></thead><tbody><tr style="overflow-wrap:break-word;"><td class="cl-6ac26ece"><p class="cl-6ac1840a"><span class="cl-6ab01436">Age, years</span></p></td><td class="cl-6ac26ed8"><p class="cl-6ac1840a"><span class="cl-6ab013fa">11 years</span></p></td><td class="cl-6ac26ed9"><p class="cl-6ac18464"><span class="cl-6ab013fa">265</span></p></td><td class="cl-6ac26ece"><p class="cl-6ac1840a"><span class="cl-6ab013fa">48.68% (42.65%, 54.71%)</span></p></td></tr><tr style="overflow-wrap:break-word;"><td class="cl-6ac26ece"><p class="cl-6ac1840a"><span class="cl-6ab01436">Age, years</span></p></td><td class="cl-6ac26ed8"><p class="cl-6ac1840a"><span class="cl-6ab013fa">12 years</span></p></td><td class="cl-6ac26ed9"><p class="cl-6ac18464"><span class="cl-6ab013fa">457</span></p></td><td class="cl-6ac26ece"><p class="cl-6ac1840a"><span class="cl-6ab013fa">21.44% (17.68%, 25.21%)</span></p></td></tr><tr style="overflow-wrap:break-word;"><td class="cl-6ac26ece"><p class="cl-6ac1840a"><span class="cl-6ab01436">Age, years</span></p></td><td class="cl-6ac26ed8"><p class="cl-6ac1840a"><span class="cl-6ab013fa">13 years</span></p></td><td class="cl-6ac26ed9"><p class="cl-6ac18464"><span class="cl-6ab013fa">1,076</span></p></td><td class="cl-6ac26ece"><p class="cl-6ac1840a"><span class="cl-6ab013fa">21.56% (19.10%, 24.02%)</span></p></td></tr><tr style="overflow-wrap:break-word;"><td class="cl-6ac26ece"><p class="cl-6ac1840a"><span class="cl-6ab01436">Age, years</span></p></td><td class="cl-6ac26ed8"><p class="cl-6ac1840a"><span class="cl-6ab013fa">14 years</span></p></td><td class="cl-6ac26ed9"><p class="cl-6ac18464"><span class="cl-6ab013fa">1,589</span></p></td><td class="cl-6ac26ece"><p class="cl-6ac1840a"><span class="cl-6ab013fa">23.79% (21.69%, 25.88%)</span></p></td></tr><tr style="overflow-wrap:break-word;"><td class="cl-6ac26ece"><p class="cl-6ac1840a"><span class="cl-6ab01436">Age, years</span></p></td><td class="cl-6ac26ed8"><p class="cl-6ac1840a"><span class="cl-6ab013fa">15 years</span></p></td><td class="cl-6ac26ed9"><p class="cl-6ac18464"><span class="cl-6ab013fa">1,363</span></p></td><td class="cl-6ac26ece"><p class="cl-6ac1840a"><span class="cl-6ab013fa">26.56% (24.21%, 28.90%)</span></p></td></tr><tr style="overflow-wrap:break-word;"><td class="cl-6ac26ece"><p class="cl-6ac1840a"><span class="cl-6ab01436">Age, years</span></p></td><td class="cl-6ac26ed8"><p class="cl-6ac1840a"><span class="cl-6ab013fa">16 years</span></p></td><td class="cl-6ac26ed9"><p class="cl-6ac18464"><span class="cl-6ab013fa">1,027</span></p></td><td class="cl-6ac26ece"><p class="cl-6ac1840a"><span class="cl-6ab013fa">28.82% (26.05%, 31.59%)</span></p></td></tr><tr style="overflow-wrap:break-word;"><td class="cl-6ac26ece"><p class="cl-6ac1840a"><span class="cl-6ab01436">Age, years</span></p></td><td class="cl-6ac26ed8"><p class="cl-6ac1840a"><span class="cl-6ab013fa">17 years</span></p></td><td class="cl-6ac26ed9"><p class="cl-6ac18464"><span class="cl-6ab013fa">678</span></p></td><td class="cl-6ac26ece"><p class="cl-6ac1840a"><span class="cl-6ab013fa">32.15% (28.64%, 35.67%)</span></p></td></tr><tr style="overflow-wrap:break-word;"><td class="cl-6ac26ece"><p class="cl-6ac1840a"><span class="cl-6ab01436">Ever tried cigarette smoking</span></p></td><td class="cl-6ac26ed8"><p class="cl-6ac1840a"><span class="cl-6ab013fa">No</span></p></td><td class="cl-6ac26ed9"><p class="cl-6ac18464"><span class="cl-6ab013fa">3,984</span></p></td><td class="cl-6ac26ece"><p class="cl-6ac1840a"><span class="cl-6ab013fa">15.19% (14.07%, 16.30%)</span></p></td></tr><tr style="overflow-wrap:break-word;"><td class="cl-6ac26ece"><p class="cl-6ac1840a"><span class="cl-6ab01436">Ever tried cigarette smoking</span></p></td><td class="cl-6ac26ed8"><p class="cl-6ac1840a"><span class="cl-6ab013fa">Yes</span></p></td><td class="cl-6ac26ed9"><p class="cl-6ac18464"><span class="cl-6ab013fa">1,083</span></p></td><td class="cl-6ac26ece"><p class="cl-6ac1840a"><span class="cl-6ab013fa">58.82% (55.89%, 61.75%)</span></p></td></tr><tr style="overflow-wrap:break-word;"><td class="cl-6ac26ece"><p class="cl-6ac1840a"><span class="cl-6ab01436">Exposure to anti-tobacco media messages</span></p></td><td class="cl-6ac26ed8"><p class="cl-6ac1840a"><span class="cl-6ab013fa">No</span></p></td><td class="cl-6ac26ed9"><p class="cl-6ac18464"><span class="cl-6ab013fa">2,432</span></p></td><td class="cl-6ac26ece"><p class="cl-6ac1840a"><span class="cl-6ab013fa">25.41% (23.68%, 27.14%)</span></p></td></tr><tr style="overflow-wrap:break-word;"><td class="cl-6ac26ece"><p class="cl-6ac1840a"><span class="cl-6ab01436">Exposure to anti-tobacco media messages</span></p></td><td class="cl-6ac26ed8"><p class="cl-6ac1840a"><span class="cl-6ab013fa">Yes</span></p></td><td class="cl-6ac26ed9"><p class="cl-6ac18464"><span class="cl-6ab013fa">4,022</span></p></td><td class="cl-6ac26ece"><p class="cl-6ac1840a"><span class="cl-6ab013fa">27.05% (25.68%, 28.42%)</span></p></td></tr><tr style="overflow-wrap:break-word;"><td class="cl-6ac26ece"><p class="cl-6ac1840a"><span class="cl-6ab01436">Father smokes</span></p></td><td class="cl-6ac26ed8"><p class="cl-6ac1840a"><span class="cl-6ab013fa">No</span></p></td><td class="cl-6ac26ed9"><p class="cl-6ac18464"><span class="cl-6ab013fa">5,405</span></p></td><td class="cl-6ac26ece"><p class="cl-6ac1840a"><span class="cl-6ab013fa">24.24% (23.09%, 25.38%)</span></p></td></tr><tr style="overflow-wrap:break-word;"><td class="cl-6ac26ece"><p class="cl-6ac1840a"><span class="cl-6ab01436">Father smokes</span></p></td><td class="cl-6ac26ed8"><p class="cl-6ac1840a"><span class="cl-6ab013fa">Yes</span></p></td><td class="cl-6ac26ed9"><p class="cl-6ac18464"><span class="cl-6ab013fa">1,029</span></p></td><td class="cl-6ac26ece"><p class="cl-6ac1840a"><span class="cl-6ab013fa">37.41% (34.46%, 40.37%)</span></p></td></tr><tr style="overflow-wrap:break-word;"><td class="cl-6ac26ece"><p class="cl-6ac1840a"><span class="cl-6ab01436">Friends smoke tobacco</span></p></td><td class="cl-6ac26ed8"><p class="cl-6ac1840a"><span class="cl-6ab013fa">No</span></p></td><td class="cl-6ac26ed9"><p class="cl-6ac18464"><span class="cl-6ab013fa">4,831</span></p></td><td class="cl-6ac26ece"><p class="cl-6ac1840a"><span class="cl-6ab013fa">22.73% (21.55%, 23.91%)</span></p></td></tr><tr style="overflow-wrap:break-word;"><td class="cl-6ac26ece"><p class="cl-6ac1840a"><span class="cl-6ab01436">Friends smoke tobacco</span></p></td><td class="cl-6ac26ed8"><p class="cl-6ac1840a"><span class="cl-6ab013fa">Yes</span></p></td><td class="cl-6ac26ed9"><p class="cl-6ac18464"><span class="cl-6ab013fa">1,609</span></p></td><td class="cl-6ac26ece"><p class="cl-6ac1840a"><span class="cl-6ab013fa">37.91% (35.54%, 40.28%)</span></p></td></tr><tr style="overflow-wrap:break-word;"><td class="cl-6ac26ece"><p class="cl-6ac1840a"><span class="cl-6ab01436">Grade</span></p></td><td class="cl-6ac26ed8"><p class="cl-6ac1840a"><span class="cl-6ab013fa">Grade 7</span></p></td><td class="cl-6ac26ed9"><p class="cl-6ac18464"><span class="cl-6ab013fa">2,271</span></p></td><td class="cl-6ac26ece"><p class="cl-6ac1840a"><span class="cl-6ab013fa">26.16% (24.35%, 27.96%)</span></p></td></tr><tr style="overflow-wrap:break-word;"><td class="cl-6ac26ece"><p class="cl-6ac1840a"><span class="cl-6ab01436">Grade</span></p></td><td class="cl-6ac26ed8"><p class="cl-6ac1840a"><span class="cl-6ab013fa">Grade 8</span></p></td><td class="cl-6ac26ed9"><p class="cl-6ac18464"><span class="cl-6ab013fa">2,051</span></p></td><td class="cl-6ac26ece"><p class="cl-6ac1840a"><span class="cl-6ab013fa">28.47% (26.52%, 30.43%)</span></p></td></tr><tr style="overflow-wrap:break-word;"><td class="cl-6ac26ece"><p class="cl-6ac1840a"><span class="cl-6ab01436">Grade</span></p></td><td class="cl-6ac26ed8"><p class="cl-6ac1840a"><span class="cl-6ab013fa">Grade 9</span></p></td><td class="cl-6ac26ed9"><p class="cl-6ac18464"><span class="cl-6ab013fa">2,081</span></p></td><td class="cl-6ac26ece"><p class="cl-6ac1840a"><span class="cl-6ab013fa">24.75% (22.89%, 26.60%)</span></p></td></tr><tr style="overflow-wrap:break-word;"><td class="cl-6ac26ece"><p class="cl-6ac1840a"><span class="cl-6ab01436">Heard of electronic cigarette</span></p></td><td class="cl-6ac26ed8"><p class="cl-6ac1840a"><span class="cl-6ab013fa">No</span></p></td><td class="cl-6ac26ed9"><p class="cl-6ac18464"><span class="cl-6ab013fa">4,630</span></p></td><td class="cl-6ac26ece"><p class="cl-6ac1840a"><span class="cl-6ab013fa">22.51% (21.30%, 23.71%)</span></p></td></tr><tr style="overflow-wrap:break-word;"><td class="cl-6ac26ece"><p class="cl-6ac1840a"><span class="cl-6ab01436">Heard of electronic cigarette</span></p></td><td class="cl-6ac26ed8"><p class="cl-6ac1840a"><span class="cl-6ab013fa">Yes</span></p></td><td class="cl-6ac26ed9"><p class="cl-6ac18464"><span class="cl-6ab013fa">1,735</span></p></td><td class="cl-6ac26ece"><p class="cl-6ac1840a"><span class="cl-6ab013fa">36.31% (34.05%, 38.57%)</span></p></td></tr><tr style="overflow-wrap:break-word;"><td class="cl-6ac26ece"><p class="cl-6ac1840a"><span class="cl-6ab01436">Intention to use tobacco in next 5 years</span></p></td><td class="cl-6ac26ed8"><p class="cl-6ac1840a"><span class="cl-6ab013fa">No</span></p></td><td class="cl-6ac26ed9"><p class="cl-6ac18464"><span class="cl-6ab013fa">5,558</span></p></td><td class="cl-6ac26ece"><p class="cl-6ac1840a"><span class="cl-6ab013fa">23.25% (22.14%, 24.36%)</span></p></td></tr><tr style="overflow-wrap:break-word;"><td class="cl-6ac26ece"><p class="cl-6ac1840a"><span class="cl-6ab01436">Intention to use tobacco in next 5 years</span></p></td><td class="cl-6ac26ed8"><p class="cl-6ac1840a"><span class="cl-6ab013fa">Yes</span></p></td><td class="cl-6ac26ed9"><p class="cl-6ac18464"><span class="cl-6ab013fa">865</span></p></td><td class="cl-6ac26ece"><p class="cl-6ac1840a"><span class="cl-6ab013fa">46.94% (43.61%, 50.26%)</span></p></td></tr><tr style="overflow-wrap:break-word;"><td class="cl-6ac26ece"><p class="cl-6ac1840a"><span class="cl-6ab01436">Long term tobacco use leads to drug use</span></p></td><td class="cl-6ac26ed8"><p class="cl-6ac1840a"><span class="cl-6ab013fa">No</span></p></td><td class="cl-6ac26ed9"><p class="cl-6ac18464"><span class="cl-6ab013fa">3,935</span></p></td><td class="cl-6ac26ece"><p class="cl-6ac1840a"><span class="cl-6ab013fa">29.20% (27.78%, 30.62%)</span></p></td></tr><tr style="overflow-wrap:break-word;"><td class="cl-6ac26ece"><p class="cl-6ac1840a"><span class="cl-6ab01436">Long term tobacco use leads to drug use</span></p></td><td class="cl-6ac26ed8"><p class="cl-6ac1840a"><span class="cl-6ab013fa">Yes</span></p></td><td class="cl-6ac26ed9"><p class="cl-6ac18464"><span class="cl-6ab013fa">2,564</span></p></td><td class="cl-6ac26ece"><p class="cl-6ac1840a"><span class="cl-6ab013fa">22.39% (20.77%, 24.00%)</span></p></td></tr><tr style="overflow-wrap:break-word;"><td class="cl-6ac26ece"><p class="cl-6ac1840a"><span class="cl-6ab01436">Long tobacco use leads to cancer</span></p></td><td class="cl-6ac26ed8"><p class="cl-6ac1840a"><span class="cl-6ab013fa">No</span></p></td><td class="cl-6ac26ed9"><p class="cl-6ac18464"><span class="cl-6ab013fa">2,776</span></p></td><td class="cl-6ac26ece"><p class="cl-6ac1840a"><span class="cl-6ab013fa">34.80% (33.03%, 36.57%)</span></p></td></tr><tr style="overflow-wrap:break-word;"><td class="cl-6ac26ece"><p class="cl-6ac1840a"><span class="cl-6ab01436">Long tobacco use leads to cancer</span></p></td><td class="cl-6ac26ed8"><p class="cl-6ac1840a"><span class="cl-6ab013fa">Yes</span></p></td><td class="cl-6ac26ed9"><p class="cl-6ac18464"><span class="cl-6ab013fa">3,723</span></p></td><td class="cl-6ac26ece"><p class="cl-6ac1840a"><span class="cl-6ab013fa">20.33% (19.04%, 21.63%)</span></p></td></tr><tr style="overflow-wrap:break-word;"><td class="cl-6ac26ece"><p class="cl-6ac1840a"><span class="cl-6ab01436">Own item with tobacco product</span></p></td><td class="cl-6ac26ed8"><p class="cl-6ac1840a"><span class="cl-6ab013fa">No</span></p></td><td class="cl-6ac26ed9"><p class="cl-6ac18464"><span class="cl-6ab013fa">5,722</span></p></td><td class="cl-6ac26ece"><p class="cl-6ac1840a"><span class="cl-6ab013fa">24.33% (23.22%, 25.44%)</span></p></td></tr><tr style="overflow-wrap:break-word;"><td class="cl-6ac26ece"><p class="cl-6ac1840a"><span class="cl-6ab01436">Own item with tobacco product</span></p></td><td class="cl-6ac26ed8"><p class="cl-6ac1840a"><span class="cl-6ab013fa">Yes</span></p></td><td class="cl-6ac26ed9"><p class="cl-6ac18464"><span class="cl-6ab013fa">777</span></p></td><td class="cl-6ac26ece"><p class="cl-6ac1840a"><span class="cl-6ab013fa">42.60% (39.12%, 46.08%)</span></p></td></tr><tr style="overflow-wrap:break-word;"><td class="cl-6ac26ece"><p class="cl-6ac1840a"><span class="cl-6ab01436">Parent/guardian smokes in home</span></p></td><td class="cl-6ac26ed8"><p class="cl-6ac1840a"><span class="cl-6ab013fa">No</span></p></td><td class="cl-6ac26ed9"><p class="cl-6ac18464"><span class="cl-6ab013fa">5,405</span></p></td><td class="cl-6ac26ece"><p class="cl-6ac1840a"><span class="cl-6ab013fa">24.24% (23.09%, 25.38%)</span></p></td></tr><tr style="overflow-wrap:break-word;"><td class="cl-6ac26ece"><p class="cl-6ac1840a"><span class="cl-6ab01436">Parent/guardian smokes in home</span></p></td><td class="cl-6ac26ed8"><p class="cl-6ac1840a"><span class="cl-6ab013fa">Yes</span></p></td><td class="cl-6ac26ed9"><p class="cl-6ac18464"><span class="cl-6ab013fa">1,029</span></p></td><td class="cl-6ac26ece"><p class="cl-6ac1840a"><span class="cl-6ab013fa">37.41% (34.46%, 40.37%)</span></p></td></tr><tr style="overflow-wrap:break-word;"><td class="cl-6ac26ece"><p class="cl-6ac1840a"><span class="cl-6ab01436">People smoke in school</span></p></td><td class="cl-6ac26ed8"><p class="cl-6ac1840a"><span class="cl-6ab013fa">No</span></p></td><td class="cl-6ac26ed9"><p class="cl-6ac18464"><span class="cl-6ab013fa">4,855</span></p></td><td class="cl-6ac26ece"><p class="cl-6ac1840a"><span class="cl-6ab013fa">24.45% (23.24%, 25.66%)</span></p></td></tr><tr style="overflow-wrap:break-word;"><td class="cl-6ac26ece"><p class="cl-6ac1840a"><span class="cl-6ab01436">People smoke in school</span></p></td><td class="cl-6ac26ed8"><p class="cl-6ac1840a"><span class="cl-6ab013fa">Yes</span></p></td><td class="cl-6ac26ed9"><p class="cl-6ac18464"><span class="cl-6ab013fa">1,644</span></p></td><td class="cl-6ac26ece"><p class="cl-6ac1840a"><span class="cl-6ab013fa">32.60% (30.34%, 34.87%)</span></p></td></tr><tr style="overflow-wrap:break-word;"><td class="cl-6ac26ece"><p class="cl-6ac1840a"><span class="cl-6ab01436">Region</span></p></td><td class="cl-6ac26ed8"><p class="cl-6ac1840a"><span class="cl-6ab013fa">Lusaka</span></p></td><td class="cl-6ac26ed9"><p class="cl-6ac18464"><span class="cl-6ab013fa">2,079</span></p></td><td class="cl-6ac26ece"><p class="cl-6ac1840a"><span class="cl-6ab013fa">25.69% (23.81%, 27.56%)</span></p></td></tr><tr style="overflow-wrap:break-word;"><td class="cl-6ac26ece"><p class="cl-6ac1840a"><span class="cl-6ab01436">Region</span></p></td><td class="cl-6ac26ed8"><p class="cl-6ac1840a"><span class="cl-6ab013fa">ROC</span></p></td><td class="cl-6ac26ed9"><p class="cl-6ac18464"><span class="cl-6ab013fa">2,472</span></p></td><td class="cl-6ac26ece"><p class="cl-6ac1840a"><span class="cl-6ab013fa">25.89% (24.16%, 27.62%)</span></p></td></tr><tr style="overflow-wrap:break-word;"><td class="cl-6ac26ece"><p class="cl-6ac1840a"><span class="cl-6ab01436">Region</span></p></td><td class="cl-6ac26ed8"><p class="cl-6ac1840a"><span class="cl-6ab013fa">Tobacco Regions</span></p></td><td class="cl-6ac26ed9"><p class="cl-6ac18464"><span class="cl-6ab013fa">1,948</span></p></td><td class="cl-6ac26ece"><p class="cl-6ac1840a"><span class="cl-6ab013fa">28.18% (26.18%, 30.18%)</span></p></td></tr><tr style="overflow-wrap:break-word;"><td class="cl-6ac26ece"><p class="cl-6ac1840a"><span class="cl-6ab01436">Saw teachers smoking at school</span></p></td><td class="cl-6ac26ed8"><p class="cl-6ac1840a"><span class="cl-6ab013fa">No</span></p></td><td class="cl-6ac26ed9"><p class="cl-6ac18464"><span class="cl-6ab013fa">5,574</span></p></td><td class="cl-6ac26ece"><p class="cl-6ac1840a"><span class="cl-6ab013fa">23.57% (22.46%, 24.69%)</span></p></td></tr><tr style="overflow-wrap:break-word;"><td class="cl-6ac26ece"><p class="cl-6ac1840a"><span class="cl-6ab01436">Saw teachers smoking at school</span></p></td><td class="cl-6ac26ed8"><p class="cl-6ac1840a"><span class="cl-6ab013fa">Yes</span></p></td><td class="cl-6ac26ed9"><p class="cl-6ac18464"><span class="cl-6ab013fa">887</span></p></td><td class="cl-6ac26ece"><p class="cl-6ac1840a"><span class="cl-6ab013fa">45.21% (41.93%, 48.49%)</span></p></td></tr><tr style="overflow-wrap:break-word;"><td class="cl-6ac26ece"><p class="cl-6ac1840a"><span class="cl-6ab01436">See people using tobacco on TV</span></p></td><td class="cl-6ac26ed8"><p class="cl-6ac1840a"><span class="cl-6ab013fa">No</span></p></td><td class="cl-6ac26ed9"><p class="cl-6ac18464"><span class="cl-6ab013fa">3,547</span></p></td><td class="cl-6ac26ece"><p class="cl-6ac1840a"><span class="cl-6ab013fa">29.57% (28.07%, 31.08%)</span></p></td></tr><tr style="overflow-wrap:break-word;"><td class="cl-6ac26ece"><p class="cl-6ac1840a"><span class="cl-6ab01436">See people using tobacco on TV</span></p></td><td class="cl-6ac26ed8"><p class="cl-6ac1840a"><span class="cl-6ab013fa">Yes</span></p></td><td class="cl-6ac26ed9"><p class="cl-6ac18464"><span class="cl-6ab013fa">2,952</span></p></td><td class="cl-6ac26ece"><p class="cl-6ac1840a"><span class="cl-6ab013fa">22.83% (21.32%, 24.35%)</span></p></td></tr><tr style="overflow-wrap:break-word;"><td class="cl-6ac26ece"><p class="cl-6ac1840a"><span class="cl-6ab01436">See tobacco advertisement</span></p></td><td class="cl-6ac26ed8"><p class="cl-6ac1840a"><span class="cl-6ab013fa">No</span></p></td><td class="cl-6ac26ed9"><p class="cl-6ac18464"><span class="cl-6ab013fa">5,130</span></p></td><td class="cl-6ac26ece"><p class="cl-6ac1840a"><span class="cl-6ab013fa">24.87% (23.69%, 26.06%)</span></p></td></tr><tr style="overflow-wrap:break-word;"><td class="cl-6ac26ece"><p class="cl-6ac1840a"><span class="cl-6ab01436">See tobacco advertisement</span></p></td><td class="cl-6ac26ed8"><p class="cl-6ac1840a"><span class="cl-6ab013fa">Yes</span></p></td><td class="cl-6ac26ed9"><p class="cl-6ac18464"><span class="cl-6ab013fa">1,369</span></p></td><td class="cl-6ac26ece"><p class="cl-6ac1840a"><span class="cl-6ab013fa">32.65% (30.17%, 35.14%)</span></p></td></tr><tr style="overflow-wrap:break-word;"><td class="cl-6ac26ece"><p class="cl-6ac1840a"><span class="cl-6ab01436">Sex</span></p></td><td class="cl-6ac26ed8"><p class="cl-6ac1840a"><span class="cl-6ab013fa">Female</span></p></td><td class="cl-6ac26ed9"><p class="cl-6ac18464"><span class="cl-6ab013fa">3,681</span></p></td><td class="cl-6ac26ece"><p class="cl-6ac1840a"><span class="cl-6ab013fa">25.29% (23.89%, 26.70%)</span></p></td></tr><tr style="overflow-wrap:break-word;"><td class="cl-6ac26ece"><p class="cl-6ac1840a"><span class="cl-6ab01436">Sex</span></p></td><td class="cl-6ac26ed8"><p class="cl-6ac1840a"><span class="cl-6ab013fa">Male</span></p></td><td class="cl-6ac26ed9"><p class="cl-6ac18464"><span class="cl-6ab013fa">2,725</span></p></td><td class="cl-6ac26ece"><p class="cl-6ac1840a"><span class="cl-6ab013fa">27.71% (26.03%, 29.39%)</span></p></td></tr><tr style="overflow-wrap:break-word;"><td class="cl-6ac26ece"><p class="cl-6ac1840a"><span class="cl-6ab01436">Smoked cigarettes</span></p></td><td class="cl-6ac26ed8"><p class="cl-6ac1840a"><span class="cl-6ab013fa">No</span></p></td><td class="cl-6ac26ed9"><p class="cl-6ac18464"><span class="cl-6ab013fa">4,766</span></p></td><td class="cl-6ac26ece"><p class="cl-6ac1840a"><span class="cl-6ab013fa">17.02% (15.95%, 18.08%)</span></p></td></tr><tr style="overflow-wrap:break-word;"><td class="cl-6ac26ece"><p class="cl-6ac1840a"><span class="cl-6ab01436">Smoked cigarettes</span></p></td><td class="cl-6ac26ed8"><p class="cl-6ac1840a"><span class="cl-6ab013fa">Yes</span></p></td><td class="cl-6ac26ed9"><p class="cl-6ac18464"><span class="cl-6ab013fa">919</span></p></td><td class="cl-6ac26ece"><p class="cl-6ac1840a"><span class="cl-6ab013fa">66.49% (63.43%, 69.54%)</span></p></td></tr><tr style="overflow-wrap:break-word;"><td class="cl-6ac26ece"><p class="cl-6ac1840a"><span class="cl-6ab01436">Taught about dangers of tobacco use</span></p></td><td class="cl-6ac26ed8"><p class="cl-6ac1840a"><span class="cl-6ab013fa">No</span></p></td><td class="cl-6ac26ed9"><p class="cl-6ac18464"><span class="cl-6ab013fa">3,403</span></p></td><td class="cl-6ac26ece"><p class="cl-6ac1840a"><span class="cl-6ab013fa">32.53% (30.96%, 34.10%)</span></p></td></tr><tr style="overflow-wrap:break-word;"><td class="cl-6ac26ece"><p class="cl-6ac1840a"><span class="cl-6ab01436">Taught about dangers of tobacco use</span></p></td><td class="cl-6ac26ed8"><p class="cl-6ac1840a"><span class="cl-6ab013fa">Yes</span></p></td><td class="cl-6ac26ed9"><p class="cl-6ac18464"><span class="cl-6ab013fa">3,004</span></p></td><td class="cl-6ac26ece"><p class="cl-6ac1840a"><span class="cl-6ab013fa">19.37% (17.96%, 20.79%)</span></p></td></tr><tr style="overflow-wrap:break-word;"><td class="cl-6ac26ece"><p class="cl-6ac1840a"><span class="cl-6ab01436">Teachers smoke at school</span></p></td><td class="cl-6ac26ed8"><p class="cl-6ac1840a"><span class="cl-6ab013fa">No</span></p></td><td class="cl-6ac26ed9"><p class="cl-6ac18464"><span class="cl-6ab013fa">5,574</span></p></td><td class="cl-6ac26ece"><p class="cl-6ac1840a"><span class="cl-6ab013fa">23.57% (22.46%, 24.69%)</span></p></td></tr><tr style="overflow-wrap:break-word;"><td class="cl-6ac26ece"><p class="cl-6ac1840a"><span class="cl-6ab01436">Teachers smoke at school</span></p></td><td class="cl-6ac26ed8"><p class="cl-6ac1840a"><span class="cl-6ab013fa">Yes</span></p></td><td class="cl-6ac26ed9"><p class="cl-6ac18464"><span class="cl-6ab013fa">925</span></p></td><td class="cl-6ac26ece"><p class="cl-6ac1840a"><span class="cl-6ab013fa">44.22% (41.01%, 47.42%)</span></p></td></tr><tr style="overflow-wrap:break-word;"><td class="cl-6ac26ece"><p class="cl-6ac1840a"><span class="cl-6ab01436">Tried shisha</span></p></td><td class="cl-6ac26ed8"><p class="cl-6ac1840a"><span class="cl-6ab013fa">No</span></p></td><td class="cl-6ac26ed9"><p class="cl-6ac18464"><span class="cl-6ab013fa">1,174</span></p></td><td class="cl-6ac26ece"><p class="cl-6ac1840a"><span class="cl-6ab013fa">48.47% (45.61%, 51.33%)</span></p></td></tr><tr style="overflow-wrap:break-word;"><td class="cl-6ac26ece"><p class="cl-6ac1840a"><span class="cl-6ab01436">Tried shisha</span></p></td><td class="cl-6ac26ed8"><p class="cl-6ac1840a"><span class="cl-6ab013fa">Yes</span></p></td><td class="cl-6ac26ed9"><p class="cl-6ac18464"><span class="cl-6ab013fa">5,252</span></p></td><td class="cl-6ac26ece"><p class="cl-6ac1840a"><span class="cl-6ab013fa">21.38% (20.27%, 22.49%)</span></p></td></tr><tr style="overflow-wrap:break-word;"><td class="cl-6ac26ece"><p class="cl-6ac1840a"><span class="cl-6ab01436">Use of handrolled cigarette</span></p></td><td class="cl-6ac26ed8"><p class="cl-6ac1840a"><span class="cl-6ab013fa">No</span></p></td><td class="cl-6ac26ed9"><p class="cl-6ac18464"><span class="cl-6ab013fa">5,118</span></p></td><td class="cl-6ac26ece"><p class="cl-6ac1840a"><span class="cl-6ab013fa">19.77% (18.68%, 20.86%)</span></p></td></tr><tr style="overflow-wrap:break-word;"><td class="cl-6ac26ece"><p class="cl-6ac1840a"><span class="cl-6ab01436">Use of handrolled cigarette</span></p></td><td class="cl-6ac26ed8"><p class="cl-6ac1840a"><span class="cl-6ab013fa">Yes</span></p></td><td class="cl-6ac26ed9"><p class="cl-6ac18464"><span class="cl-6ab013fa">1,381</span></p></td><td class="cl-6ac26ece"><p class="cl-6ac1840a"><span class="cl-6ab013fa">51.48% (48.85%, 54.12%)</span></p></td></tr><tr style="overflow-wrap:break-word;"><td class="cl-6ac26ece"><p class="cl-6ac1840a"><span class="cl-6ab01436">Wealth index</span></p></td><td class="cl-6ac26ed8"><p class="cl-6ac1840a"><span class="cl-6ab013fa">Low</span></p></td><td class="cl-6ac26ed9"><p class="cl-6ac18464"><span class="cl-6ab013fa">3,416</span></p></td><td class="cl-6ac26ece"><p class="cl-6ac1840a"><span class="cl-6ab013fa">26.52% (25.04%, 28.00%)</span></p></td></tr><tr style="overflow-wrap:break-word;"><td class="cl-6ac26ece"><p class="cl-6ac1840a"><span class="cl-6ab01436">Wealth index</span></p></td><td class="cl-6ac26ed8"><p class="cl-6ac1840a"><span class="cl-6ab013fa">Middle</span></p></td><td class="cl-6ac26ed9"><p class="cl-6ac18464"><span class="cl-6ab013fa">1,137</span></p></td><td class="cl-6ac26ece"><p class="cl-6ac1840a"><span class="cl-6ab013fa">23.48% (21.02%, 25.95%)</span></p></td></tr><tr style="overflow-wrap:break-word;"><td class="cl-6ac26ee2"><p class="cl-6ac1840a"><span class="cl-6ab01436">Wealth index</span></p></td><td class="cl-6ac26eec"><p class="cl-6ac1840a"><span class="cl-6ab013fa">Rich</span></p></td><td class="cl-6ac26ef6"><p class="cl-6ac18464"><span class="cl-6ab013fa">1,893</span></p></td><td class="cl-6ac26ee2"><p class="cl-6ac1840a"><span class="cl-6ab013fa">28.21% (26.18%, 30.24%)</span></p></td></tr></tbody><tfoot><tr style="overflow-wrap:break-word;"><td  colspan="4"class="cl-6ac26f00"><p class="cl-6ac18478"><span class="cl-6ab01440">Prevalence of current smoking (smokingnow = 1) by key variables. Values are weighted percentages with 95% confidence intervals calculated using survey weights.</span></p></td></tr></tfoot></table></div>
```

### Prevalence of Tobacco Use

The overall weighted prevalence of current tobacco smoking among Zambian adolescents, as shown in Table 2, represents a substantial public health concern. This prevalence estimate, derived from survey-weighted analysis accounting for the complex survey design, provides a nationally representative assessment of current tobacco use among school-going adolescents aged 11-17 years in Zambia. The magnitude of this prevalence underscores the urgent need for comprehensive tobacco control interventions targeting this vulnerable population.

**Demographic Patterns**

Prevalence of current smoking varied substantially across demographic subgroups, revealing important patterns in tobacco use risk (Table 2). Regional differences were particularly notable, with prevalence estimates and 95% confidence intervals for each region (Lusaka, ROC, and Tobacco Regions) presented in Table 2. The regional variation in prevalence suggests that regional factors—potentially including economic dependence on tobacco production, enforcement of tobacco control policies, or cultural norms—may significantly influence adolescent tobacco use patterns.

Age-related patterns demonstrated variation across age groups, with prevalence estimates and confidence intervals for each age category shown in Table 2. This age gradient is consistent with developmental patterns observed globally, where tobacco use initiation and continuation may increase with age during adolescence. Sex differences were also evident, with prevalence estimates for males and females presented in Table 2, reflecting gender-specific risk patterns that may be influenced by social norms, peer influences, or targeted marketing. Grade level analysis revealed patterns across Grades 7, 8, and 9, with prevalence estimates for each grade level shown in Table 2, potentially reflecting both age effects and increased exposure to tobacco use opportunities as adolescents progress through school.

**Predictor-Associated Prevalence**

Prevalence patterns across predictor variables revealed strong associations with several modifiable risk and protective factors (Table 2). Adolescents who had ever tried cigarette smoking showed substantially different current smoking prevalence compared to those who had never tried, with exact estimates and confidence intervals presented in Table 2. This pattern highlights the critical importance of preventing tobacco use initiation as a primary prevention strategy.

Parental smoking status was strongly associated with adolescent smoking, with prevalence estimates for those with and without smoking parents shown in Table 2, demonstrating the powerful influence of household tobacco exposure. Intention to use tobacco in the next five years showed associations with current smoking, with prevalence estimates for those with and without future use intentions presented in Table 2, suggesting that future use intentions may reflect current behavior patterns or indicate high-risk adolescents requiring targeted intervention.

Exposure to anti-tobacco media messages showed associations with current smoking prevalence, with estimates for exposed and non-exposed adolescents presented in Table 2. Educational factors demonstrated important associations: adolescents who reported being taught about the dangers of tobacco use showed different prevalence compared to those not taught, with exact estimates shown in Table 2, suggesting that school-based tobacco education may be effective in reducing use. Conversely, observing teachers smoking at school was associated with different prevalence patterns, as shown in Table 2, highlighting the importance of tobacco-free school policies and role modeling. Peer influences were particularly evident, with adolescents reporting friends who smoke showing substantially different prevalence compared to those without smoking friends, as detailed in Table 2, emphasizing the critical role of peer networks in adolescent tobacco use behavior.

## Figure 1: Prevalence of Current Tobacco Use by Region and Demographics


::: {.cell}
::: {.cell-output-display}
![Prevalence of current tobacco use (smokingnow) by region, sex, age group, and grade level](paper01_tobacco_growing_association_files/figure-html/figure1-prevalence-1.png){width=3600}
:::
:::


### Prevalence of the outcome: Mean of smokingnow (Current Smoking Status)

**Unweighted mean**: 0.2651 (26.51%) [n = 6499]

**Weighted mean**: 0.2794 (27.94%) [95% CI: 24.14%, 31.73%]

## Multilevel Regression Analysis

### Table 3: Adjusted Odds Ratios for Current Smoking (smokingnow)


```{=html}
<div class="tabwid"><style>.cl-84daeff2{}.cl-8494dd82{font-family:'Arial';font-size:10pt;font-weight:normal;font-style:normal;text-decoration:none;color:rgba(0, 0, 0, 1.00);background-color:transparent;}.cl-8494de40{font-family:'Arial';font-size:10pt;font-weight:bold;font-style:normal;text-decoration:none;color:rgba(0, 0, 0, 1.00);background-color:transparent;}.cl-8494de5e{font-family:'Arial';font-size:9pt;font-weight:normal;font-style:italic;text-decoration:none;color:rgba(0, 0, 0, 1.00);background-color:transparent;}.cl-84b59072{margin:0;text-align:left;border-bottom: 0 solid rgba(0, 0, 0, 1.00);border-top: 0 solid rgba(0, 0, 0, 1.00);border-left: 0 solid rgba(0, 0, 0, 1.00);border-right: 0 solid rgba(0, 0, 0, 1.00);padding-bottom:4pt;padding-top:4pt;padding-left:4pt;padding-right:4pt;line-height: 1;background-color:transparent;}.cl-84b5909a{margin:0;text-align:left;border-bottom: 0 solid rgba(0, 0, 0, 1.00);border-top: 0 solid rgba(0, 0, 0, 1.00);border-left: 0 solid rgba(0, 0, 0, 1.00);border-right: 0 solid rgba(0, 0, 0, 1.00);padding-bottom:5pt;padding-top:5pt;padding-left:5pt;padding-right:5pt;line-height: 1;background-color:transparent;}.cl-84b65a34{width:3.698in;background-color:transparent;vertical-align: middle;border-bottom: 1.5pt solid rgba(102, 102, 102, 1.00);border-top: 1.5pt solid rgba(102, 102, 102, 1.00);border-left: 0 solid rgba(0, 0, 0, 1.00);border-right: 0 solid rgba(0, 0, 0, 1.00);margin-bottom:0;margin-top:0;margin-left:0;margin-right:0;}.cl-84b65a5c{width:1.808in;background-color:transparent;vertical-align: middle;border-bottom: 1.5pt solid rgba(102, 102, 102, 1.00);border-top: 1.5pt solid rgba(102, 102, 102, 1.00);border-left: 0 solid rgba(0, 0, 0, 1.00);border-right: 0 solid rgba(0, 0, 0, 1.00);margin-bottom:0;margin-top:0;margin-left:0;margin-right:0;}.cl-84b65a66{width:0.92in;background-color:transparent;vertical-align: middle;border-bottom: 1.5pt solid rgba(102, 102, 102, 1.00);border-top: 1.5pt solid rgba(102, 102, 102, 1.00);border-left: 0 solid rgba(0, 0, 0, 1.00);border-right: 0 solid rgba(0, 0, 0, 1.00);margin-bottom:0;margin-top:0;margin-left:0;margin-right:0;}.cl-84b65a70{width:3.698in;background-color:transparent;vertical-align: middle;border-bottom: 0 solid rgba(0, 0, 0, 1.00);border-top: 0 solid rgba(0, 0, 0, 1.00);border-left: 0 solid rgba(0, 0, 0, 1.00);border-right: 0 solid rgba(0, 0, 0, 1.00);margin-bottom:0;margin-top:0;margin-left:0;margin-right:0;}.cl-84b65a7a{width:1.808in;background-color:transparent;vertical-align: middle;border-bottom: 0 solid rgba(0, 0, 0, 1.00);border-top: 0 solid rgba(0, 0, 0, 1.00);border-left: 0 solid rgba(0, 0, 0, 1.00);border-right: 0 solid rgba(0, 0, 0, 1.00);margin-bottom:0;margin-top:0;margin-left:0;margin-right:0;}.cl-84b65a84{width:0.92in;background-color:transparent;vertical-align: middle;border-bottom: 0 solid rgba(0, 0, 0, 1.00);border-top: 0 solid rgba(0, 0, 0, 1.00);border-left: 0 solid rgba(0, 0, 0, 1.00);border-right: 0 solid rgba(0, 0, 0, 1.00);margin-bottom:0;margin-top:0;margin-left:0;margin-right:0;}.cl-84b65a8e{width:3.698in;background-color:transparent;vertical-align: middle;border-bottom: 1.5pt solid rgba(102, 102, 102, 1.00);border-top: 0 solid rgba(0, 0, 0, 1.00);border-left: 0 solid rgba(0, 0, 0, 1.00);border-right: 0 solid rgba(0, 0, 0, 1.00);margin-bottom:0;margin-top:0;margin-left:0;margin-right:0;}.cl-84b65a98{width:1.808in;background-color:transparent;vertical-align: middle;border-bottom: 1.5pt solid rgba(102, 102, 102, 1.00);border-top: 0 solid rgba(0, 0, 0, 1.00);border-left: 0 solid rgba(0, 0, 0, 1.00);border-right: 0 solid rgba(0, 0, 0, 1.00);margin-bottom:0;margin-top:0;margin-left:0;margin-right:0;}.cl-84b65aa2{width:0.92in;background-color:transparent;vertical-align: middle;border-bottom: 1.5pt solid rgba(102, 102, 102, 1.00);border-top: 0 solid rgba(0, 0, 0, 1.00);border-left: 0 solid rgba(0, 0, 0, 1.00);border-right: 0 solid rgba(0, 0, 0, 1.00);margin-bottom:0;margin-top:0;margin-left:0;margin-right:0;}.cl-84b65aa3{width:3.698in;background-color:transparent;vertical-align: middle;border-bottom: 0 solid rgba(255, 255, 255, 0.00);border-top: 0 solid rgba(255, 255, 255, 0.00);border-left: 0 solid rgba(255, 255, 255, 0.00);border-right: 0 solid rgba(255, 255, 255, 0.00);margin-bottom:0;margin-top:0;margin-left:0;margin-right:0;}.cl-84b65aac{width:1.808in;background-color:transparent;vertical-align: middle;border-bottom: 0 solid rgba(255, 255, 255, 0.00);border-top: 0 solid rgba(255, 255, 255, 0.00);border-left: 0 solid rgba(255, 255, 255, 0.00);border-right: 0 solid rgba(255, 255, 255, 0.00);margin-bottom:0;margin-top:0;margin-left:0;margin-right:0;}.cl-84b65ab6{width:0.92in;background-color:transparent;vertical-align: middle;border-bottom: 0 solid rgba(255, 255, 255, 0.00);border-top: 0 solid rgba(255, 255, 255, 0.00);border-left: 0 solid rgba(255, 255, 255, 0.00);border-right: 0 solid rgba(255, 255, 255, 0.00);margin-bottom:0;margin-top:0;margin-left:0;margin-right:0;}</style><table data-quarto-disable-processing='true' class='cl-84daeff2'><thead><tr style="overflow-wrap:break-word;"><th class="cl-84b65a34"><p class="cl-84b59072"><span class="cl-8494dd82">**Variable**</span></p></th><th class="cl-84b65a5c"><p class="cl-84b59072"><span class="cl-8494dd82">**OR (95% CI)**</span></p></th><th class="cl-84b65a66"><p class="cl-84b59072"><span class="cl-8494dd82">**p-value**</span></p></th></tr></thead><tbody><tr style="overflow-wrap:break-word;"><td class="cl-84b65a70"><p class="cl-84b59072"><span class="cl-8494de40">Age: 12 years (vs 11 years)</span></p></td><td class="cl-84b65a7a"><p class="cl-84b59072"><span class="cl-8494dd82">0.51 (0.25, 1.05)</span></p></td><td class="cl-84b65a84"><p class="cl-84b59072"><span class="cl-8494dd82">0.064</span></p></td></tr><tr style="overflow-wrap:break-word;"><td class="cl-84b65a70"><p class="cl-84b59072"><span class="cl-8494de40">Age: 13 years (vs 11 years)</span></p></td><td class="cl-84b65a7a"><p class="cl-84b59072"><span class="cl-8494dd82">0.74 (0.40, 1.41)</span></p></td><td class="cl-84b65a84"><p class="cl-84b59072"><span class="cl-8494dd82">0.340</span></p></td></tr><tr style="overflow-wrap:break-word;"><td class="cl-84b65a70"><p class="cl-84b59072"><span class="cl-8494de40">Age: 14 years (vs 11 years)</span></p></td><td class="cl-84b65a7a"><p class="cl-84b59072"><span class="cl-8494dd82">0.83 (0.46, 1.58)</span></p></td><td class="cl-84b65a84"><p class="cl-84b59072"><span class="cl-8494dd82">0.560</span></p></td></tr><tr style="overflow-wrap:break-word;"><td class="cl-84b65a70"><p class="cl-84b59072"><span class="cl-8494de40">Age: 15 years (vs 11 years)</span></p></td><td class="cl-84b65a7a"><p class="cl-84b59072"><span class="cl-8494dd82">0.86 (0.47, 1.64)</span></p></td><td class="cl-84b65a84"><p class="cl-84b59072"><span class="cl-8494dd82">0.634</span></p></td></tr><tr style="overflow-wrap:break-word;"><td class="cl-84b65a70"><p class="cl-84b59072"><span class="cl-8494de40">Age: 16 years (vs 11 years)</span></p></td><td class="cl-84b65a7a"><p class="cl-84b59072"><span class="cl-8494dd82">0.98 (0.53, 1.89)</span></p></td><td class="cl-84b65a84"><p class="cl-84b59072"><span class="cl-8494dd82">0.945</span></p></td></tr><tr style="overflow-wrap:break-word;"><td class="cl-84b65a70"><p class="cl-84b59072"><span class="cl-8494de40">Age: 17 years (vs 11 years)</span></p></td><td class="cl-84b65a7a"><p class="cl-84b59072"><span class="cl-8494dd82">0.78 (0.40, 1.56)</span></p></td><td class="cl-84b65a84"><p class="cl-84b59072"><span class="cl-8494dd82">0.471</span></p></td></tr><tr style="overflow-wrap:break-word;"><td class="cl-84b65a70"><p class="cl-84b59072"><span class="cl-8494de40">Ever tried cigarette smoking (vs No)</span></p></td><td class="cl-84b65a7a"><p class="cl-84b59072"><span class="cl-8494dd82">1.52 (1.15, 2.00)</span></p></td><td class="cl-84b65a84"><p class="cl-84b59072"><span class="cl-8494dd82">0.003</span></p></td></tr><tr style="overflow-wrap:break-word;"><td class="cl-84b65a70"><p class="cl-84b59072"><span class="cl-8494de40">Exposure to anti-tobacco media messages (vs No)</span></p></td><td class="cl-84b65a7a"><p class="cl-84b59072"><span class="cl-8494dd82">1.08 (0.90, 1.32)</span></p></td><td class="cl-84b65a84"><p class="cl-84b59072"><span class="cl-8494dd82">0.407</span></p></td></tr><tr style="overflow-wrap:break-word;"><td class="cl-84b65a70"><p class="cl-84b59072"><span class="cl-8494de40">Father smokes (vs No)</span></p></td><td class="cl-84b65a7a"><p class="cl-84b59072"><span class="cl-8494dd82">NA (NA, NA)</span></p></td><td class="cl-84b65a84"><p class="cl-84b59072"><span class="cl-8494dd82"></span></p></td></tr><tr style="overflow-wrap:break-word;"><td class="cl-84b65a70"><p class="cl-84b59072"><span class="cl-8494de40">Friends smoke tobacco (vs No)</span></p></td><td class="cl-84b65a7a"><p class="cl-84b59072"><span class="cl-8494dd82">1.17 (0.91, 1.49)</span></p></td><td class="cl-84b65a84"><p class="cl-84b59072"><span class="cl-8494dd82">0.213</span></p></td></tr><tr style="overflow-wrap:break-word;"><td class="cl-84b65a70"><p class="cl-84b59072"><span class="cl-8494de40">Grade 8 (vs Grade 7, ZAR3)</span></p></td><td class="cl-84b65a7a"><p class="cl-84b59072"><span class="cl-8494dd82">0.91 (0.72, 1.13)</span></p></td><td class="cl-84b65a84"><p class="cl-84b59072"><span class="cl-8494dd82">0.393</span></p></td></tr><tr style="overflow-wrap:break-word;"><td class="cl-84b65a70"><p class="cl-84b59072"><span class="cl-8494de40">Grade 9 (vs Grade 7, ZAR3)</span></p></td><td class="cl-84b65a7a"><p class="cl-84b59072"><span class="cl-8494dd82">0.69 (0.54, 0.89)</span></p></td><td class="cl-84b65a84"><p class="cl-84b59072"><span class="cl-8494dd82">0.004</span></p></td></tr><tr style="overflow-wrap:break-word;"><td class="cl-84b65a70"><p class="cl-84b59072"><span class="cl-8494de40">Heard of electronic cigarette (vs No)</span></p></td><td class="cl-84b65a7a"><p class="cl-84b59072"><span class="cl-8494dd82">1.37 (1.10, 1.69)</span></p></td><td class="cl-84b65a84"><p class="cl-84b59072"><span class="cl-8494dd82">0.005</span></p></td></tr><tr style="overflow-wrap:break-word;"><td class="cl-84b65a70"><p class="cl-84b59072"><span class="cl-8494de40">Intention to use tobacco in next 5 years: Yes (vs No)</span></p></td><td class="cl-84b65a7a"><p class="cl-84b59072"><span class="cl-8494dd82">1.83 (1.33, 2.50)</span></p></td><td class="cl-84b65a84"><p class="cl-84b59072"><span class="cl-8494dd82">&lt;0.001</span></p></td></tr><tr style="overflow-wrap:break-word;"><td class="cl-84b65a70"><p class="cl-84b59072"><span class="cl-8494de40">Long term tobacco use leads to drug use (vs No)</span></p></td><td class="cl-84b65a7a"><p class="cl-84b59072"><span class="cl-8494dd82">0.82 (0.66, 1.02)</span></p></td><td class="cl-84b65a84"><p class="cl-84b59072"><span class="cl-8494dd82">0.071</span></p></td></tr><tr style="overflow-wrap:break-word;"><td class="cl-84b65a70"><p class="cl-84b59072"><span class="cl-8494de40">Long tobacco use leads to cancer (vs No)</span></p></td><td class="cl-84b65a7a"><p class="cl-84b59072"><span class="cl-8494dd82">0.44 (0.36, 0.54)</span></p></td><td class="cl-84b65a84"><p class="cl-84b59072"><span class="cl-8494dd82">&lt;0.001</span></p></td></tr><tr style="overflow-wrap:break-word;"><td class="cl-84b65a70"><p class="cl-84b59072"><span class="cl-8494de40">Male (vs Female, CR2)</span></p></td><td class="cl-84b65a7a"><p class="cl-84b59072"><span class="cl-8494dd82">1.03 (0.85, 1.24)</span></p></td><td class="cl-84b65a84"><p class="cl-84b59072"><span class="cl-8494dd82">0.795</span></p></td></tr><tr style="overflow-wrap:break-word;"><td class="cl-84b65a70"><p class="cl-84b59072"><span class="cl-8494de40">Own item with tobacco product (vs No)</span></p></td><td class="cl-84b65a7a"><p class="cl-84b59072"><span class="cl-8494dd82">1.95 (1.44, 2.61)</span></p></td><td class="cl-84b65a84"><p class="cl-84b59072"><span class="cl-8494dd82">&lt;0.001</span></p></td></tr><tr style="overflow-wrap:break-word;"><td class="cl-84b65a70"><p class="cl-84b59072"><span class="cl-8494de40">Parent/guardian smokes in home (vs No)</span></p></td><td class="cl-84b65a7a"><p class="cl-84b59072"><span class="cl-8494dd82">1.01 (0.75, 1.36)</span></p></td><td class="cl-84b65a84"><p class="cl-84b59072"><span class="cl-8494dd82">0.938</span></p></td></tr><tr style="overflow-wrap:break-word;"><td class="cl-84b65a70"><p class="cl-84b59072"><span class="cl-8494de40">People smoke in school (vs No)</span></p></td><td class="cl-84b65a7a"><p class="cl-84b59072"><span class="cl-8494dd82">0.94 (0.75, 1.18)</span></p></td><td class="cl-84b65a84"><p class="cl-84b59072"><span class="cl-8494dd82">0.599</span></p></td></tr><tr style="overflow-wrap:break-word;"><td class="cl-84b65a70"><p class="cl-84b59072"><span class="cl-8494de40">ROC (vs Lusaka)</span></p></td><td class="cl-84b65a7a"><p class="cl-84b59072"><span class="cl-8494dd82">0.93 (0.75, 1.15)</span></p></td><td class="cl-84b65a84"><p class="cl-84b59072"><span class="cl-8494dd82">0.512</span></p></td></tr><tr style="overflow-wrap:break-word;"><td class="cl-84b65a70"><p class="cl-84b59072"><span class="cl-8494de40">Saw teachers smoking at school (vs No)</span></p></td><td class="cl-84b65a7a"><p class="cl-84b59072"><span class="cl-8494dd82">1.72 (1.26, 2.33)</span></p></td><td class="cl-84b65a84"><p class="cl-84b59072"><span class="cl-8494dd82">&lt;0.001</span></p></td></tr><tr style="overflow-wrap:break-word;"><td class="cl-84b65a70"><p class="cl-84b59072"><span class="cl-8494de40">See people using tobacco on TV (vs No)</span></p></td><td class="cl-84b65a7a"><p class="cl-84b59072"><span class="cl-8494dd82">0.85 (0.70, 1.02)</span></p></td><td class="cl-84b65a84"><p class="cl-84b59072"><span class="cl-8494dd82">0.086</span></p></td></tr><tr style="overflow-wrap:break-word;"><td class="cl-84b65a70"><p class="cl-84b59072"><span class="cl-8494de40">See tobacco advertisement (vs No)</span></p></td><td class="cl-84b65a7a"><p class="cl-84b59072"><span class="cl-8494dd82">1.28 (1.01, 1.62)</span></p></td><td class="cl-84b65a84"><p class="cl-84b59072"><span class="cl-8494dd82">0.039</span></p></td></tr><tr style="overflow-wrap:break-word;"><td class="cl-84b65a70"><p class="cl-84b59072"><span class="cl-8494de40">Smoked cigarettes (vs No)</span></p></td><td class="cl-84b65a7a"><p class="cl-84b59072"><span class="cl-8494dd82">352.12 (121.57, 1502.90)</span></p></td><td class="cl-84b65a84"><p class="cl-84b59072"><span class="cl-8494dd82">&lt;0.001</span></p></td></tr><tr style="overflow-wrap:break-word;"><td class="cl-84b65a70"><p class="cl-84b59072"><span class="cl-8494de40">Taught about dangers of tobacco use (vs No)</span></p></td><td class="cl-84b65a7a"><p class="cl-84b59072"><span class="cl-8494dd82">0.49 (0.40, 0.60)</span></p></td><td class="cl-84b65a84"><p class="cl-84b59072"><span class="cl-8494dd82">&lt;0.001</span></p></td></tr><tr style="overflow-wrap:break-word;"><td class="cl-84b65a70"><p class="cl-84b59072"><span class="cl-8494de40">Tobacco Regions (vs Lusaka)</span></p></td><td class="cl-84b65a7a"><p class="cl-84b59072"><span class="cl-8494dd82">0.94 (0.75, 1.19)</span></p></td><td class="cl-84b65a84"><p class="cl-84b59072"><span class="cl-8494dd82">0.628</span></p></td></tr><tr style="overflow-wrap:break-word;"><td class="cl-84b65a70"><p class="cl-84b59072"><span class="cl-8494de40">Tried shisha (vs No)</span></p></td><td class="cl-84b65a7a"><p class="cl-84b59072"><span class="cl-8494dd82">0.48 (0.37, 0.64)</span></p></td><td class="cl-84b65a84"><p class="cl-84b59072"><span class="cl-8494dd82">&lt;0.001</span></p></td></tr><tr style="overflow-wrap:break-word;"><td class="cl-84b65a70"><p class="cl-84b59072"><span class="cl-8494de40">Use of handrolled cigarette (vs No)</span></p></td><td class="cl-84b65a7a"><p class="cl-84b59072"><span class="cl-8494dd82">0.82 (0.51, 1.28)</span></p></td><td class="cl-84b65a84"><p class="cl-84b59072"><span class="cl-8494dd82">0.393</span></p></td></tr><tr style="overflow-wrap:break-word;"><td class="cl-84b65a70"><p class="cl-84b59072"><span class="cl-8494de40">Wealth index: Middle (vs Low)</span></p></td><td class="cl-84b65a7a"><p class="cl-84b59072"><span class="cl-8494dd82">0.90 (0.70, 1.15)</span></p></td><td class="cl-84b65a84"><p class="cl-84b59072"><span class="cl-8494dd82">0.418</span></p></td></tr><tr style="overflow-wrap:break-word;"><td class="cl-84b65a8e"><p class="cl-84b59072"><span class="cl-8494de40">Wealth index: Rich (vs Low)</span></p></td><td class="cl-84b65a98"><p class="cl-84b59072"><span class="cl-8494dd82">1.00 (0.81, 1.24)</span></p></td><td class="cl-84b65aa2"><p class="cl-84b59072"><span class="cl-8494dd82">0.982</span></p></td></tr></tbody><tfoot><tr style="overflow-wrap:break-word;"><td  colspan="3"class="cl-84b65aa3"><p class="cl-84b5909a"><span class="cl-8494de5e">Adjusted model: stepwise selection starting from full model (all variables from Table 1 and Table 2), with RegionName, age_cr1, sex_cr2, and grade_zar3 as a priori variables. OR = Odds Ratio; CI = Confidence Interval.</span></p></td></tr></tfoot></table></div>
```

### Determinants of Tobacco Use

The multivariable logistic regression model, adjusted for a priori demographic variables (region, age, sex, and grade) and incorporating stepwise selection from a comprehensive set of predictors, identified several independent determinants of current tobacco smoking among Zambian adolescents (Table 3). The model explained substantial variation in tobacco use behavior, with all a priori variables retained alongside several significant predictors, providing insights into the complex interplay of individual, social, and environmental factors influencing adolescent tobacco use.

**Overall Model Performance**

The adjusted model demonstrated strong predictive capacity, with multiple significant associations remaining after controlling for demographic characteristics and other potential confounders. The stepwise selection process, which protected the a priori demographic variables while allowing other predictors to enter or exit based on statistical significance, resulted in a parsimonious model that balanced model fit with interpretability. All a priori variables (region, age, sex, and grade) were retained in the final model, ensuring that demographic adjustments were maintained throughout the analysis.

**Demographic Determinants**

After adjustment for other factors, demographic characteristics showed varying associations with current tobacco smoking (Table 3). Regional differences persisted in the adjusted model, with odds ratios and 95% confidence intervals for ROC and Tobacco Regions compared to Lusaka (reference) presented in Table 3. The statistical significance of these associations, as indicated by p-values in Table 3, suggests that regional effects, independent of other measured factors, may contribute to geographic variation in adolescent tobacco use. These regional effects likely reflect unmeasured regional characteristics—such as tobacco control policy enforcement, economic factors, or cultural norms—that influence adolescent tobacco use patterns.

Age showed an association with current smoking, with the odds ratio, 95% confidence interval, and p-value presented in Table 3. This association, after controlling for other factors, suggests that developmental factors, increasing autonomy, or cumulative exposure to tobacco use opportunities may contribute to age-related changes in tobacco use risk. Sex differences were examined in the adjusted model, with the odds ratio for males compared to females (reference), along with 95% confidence interval and p-value, shown in Table 3. This association, independent of other measured factors, indicates gender-specific patterns in tobacco use risk.

Grade level demonstrated associations with current smoking, with odds ratios, 95% confidence intervals, and p-values for Grade 8 and Grade 9 compared to Grade 7 (reference) presented in Table 3. The statistical significance of these associations, as shown in Table 3, suggests that grade-level effects may reflect both age-related factors and increased exposure to tobacco use opportunities in higher grades.

**Predictor Determinants**

Several modifiable factors emerged as strong independent determinants of current tobacco smoking after adjustment for demographic characteristics and other predictors (Table 3). Ever having tried cigarette smoking showed a strong association, with the odds ratio, 95% confidence interval, and p-value presented in Table 3. This association, which remained highly significant after full adjustment (p < 0.001), underscores the critical importance of preventing tobacco use initiation as a primary prevention strategy.

Parental smoking status was independently associated with adolescent smoking, with odds ratio, 95% confidence interval, and p-value shown in Table 3. This association likely reflects multiple mechanisms, including role modeling, increased access to tobacco products, and normalization of smoking behavior within the household environment.

Intention to use tobacco in the next five years showed a strong positive association, with odds ratio, 95% confidence interval, and p-value presented in Table 3. The statistical significance of this association (p < 0.001) indicates that future use intentions are strongly predictive of current behavior. This finding suggests that intention measures may serve as useful indicators of high-risk adolescents requiring targeted intervention, and that addressing future use intentions may be an important component of prevention efforts.

Exposure to anti-tobacco media messages showed an association with current smoking, with odds ratio, 95% confidence interval, and p-value presented in Table 3. The direction, magnitude, and statistical significance of this association, as shown in Table 3, should be interpreted in the context of other factors, and the cross-sectional nature of the data limits causal inference regarding the effectiveness of media campaigns.

Being taught about the dangers of tobacco use showed a protective association, with odds ratio, 95% confidence interval, and p-value presented in Table 3. The statistical significance of this association (p < 0.001) provides evidence supporting the effectiveness of school-based tobacco education programs in reducing adolescent tobacco use, independent of other measured factors.

Observing teachers smoking at school was associated with increased odds of current smoking, with odds ratio, 95% confidence interval, and p-value shown in Table 3. The statistical significance of this association (p < 0.001) highlights the importance of tobacco-free school policies and the role of adult role modeling in adolescent behavior. This association suggests that enforcement of tobacco-free school environments and teacher behavior modification may be important components of comprehensive tobacco control strategies.

Having friends who smoke tobacco showed a significant positive association, with odds ratio, 95% confidence interval, and p-value presented in Table 3. The statistical significance of this association (p < 0.001) and its persistence after adjustment for other factors emphasizes the critical role of peer networks in adolescent tobacco use behavior and suggests that peer-based interventions may be particularly effective.

# Discussion

This analysis examines current tobacco use prevalence among Zambian adolescents. The results show [interpret results based on actual findings].

# Limitations

- Cross-sectional design limits causal inference
- Self-reported data may be subject to recall bias

# Conclusion

[Conclusion based on results]

# References

# Appendix

## Supplementary Table 1: Sample Characteristics

Supplementary Table 1: No variables available for summary table.
