## Supplemental codes for "Methodological Guidance for Predictor Variable Selection for Adolescent Smoking Outcomes in Global Youth Tobacco Survey Using R and Python": paper01_tobacco_growing_association.qmd

````
---
title: "Current Tobacco Use Among Adolescents in Zambia: Analysis of GYTS 2021 Data"
subtitle: "Paper 1: GYTS 2021 Analysis"
author: "GYTS Research Team"
date: today
bibliography: references.bib
format:
  html:
    toc: true
    toc-depth: 3
    toc-location: left
    code-fold: show
    code-tools: true
    fig-width: 10
    fig-height: 6
    theme: cosmo
    css: styles.css
execute:
  warning: false
  message: false
---

```{r setup, include=FALSE}
knitr::opts_chunk$set(
  echo = TRUE,
  warning = FALSE,
  message = FALSE,
  fig.width = 10,
  fig.height = 6,
  dpi = 300
)

# Load required libraries
library(readxl)  # For reading Excel files
library(tidyverse)
library(survey)
library(gtsummary)
library(flextable)
library(patchwork)
library(scales)
library(viridis)
library(RColorBrewer)
library(knitr)
library(kableExtra)
library(broom)
library(lme4)
library(emmeans)
library(ggplot2)
library(ggthemes)
```

```{r data-loading, include=FALSE}
# Load the GYTS 2021 National dataset from Excel file
library(readxl)
GYTS_2021_Zambia_National_ <- read_excel("../data/National/GYTS 2021 Zambia (National).XLS", 
    sheet = "ZAMBIA_DATA_21")

# Print available column names for debugging (first 50 columns)
cat("Available columns in dataset (first 50):\n")
print(head(names(GYTS_2021_Zambia_National_), 50))
cat("\nTotal columns:", length(names(GYTS_2021_Zambia_National_)), "\n\n")

# Helper function to safely get column or return default
safe_get_col <- function(col_name, default_val = NA) {
  if(col_name %in% names(GYTS_2021_Zambia_National_)) {
    return(GYTS_2021_Zambia_National_[[col_name]])
  } else {
    # Try case-insensitive match
    match_col <- names(GYTS_2021_Zambia_National_)[grepl(paste0("^", col_name, "$"), names(GYTS_2021_Zambia_National_), ignore.case = TRUE)][1]
    if(!is.na(match_col)) {
      return(GYTS_2021_Zambia_National_[[match_col]])
    } else {
      return(rep(default_val, nrow(GYTS_2021_Zambia_National_)))
    }
  }
}

# Extract all CR and ZMR variables we need (checking if they exist)
# This allows the code to work even if column names differ
CR_vars <- list()
ZMR_vars <- list()

# List of all CR variables we need
cr_var_names <- c("CR1", "CR2", "CR3", "CR4", "CR5", "CR6", "CR7", "CR8", "CR9", "CR10", "CR12", "CR14", 
                  "CR17", "CR21", "CR22", "CR23", "CR24", "CR25", "CR28", "CR29", 
                  "CR30", "CR31", "CR33", "CR34", "CR35", "CR37", "CR38", "CR39", "CR40", "CR41", 
                  "CR42", "CR43", "CR44", "CR45", "CR46", "CR47", "CR48", "CR49", 
                  "CR50", "CR52", "CR53", "CR54")

# List of all ZMR variables we need
zmr_var_names <- c("ZMR12", "ZMR14", "ZMR35", "ZMR36", "ZMR41", "ZMR56", "ZMR57", 
                   "ZMR58", "ZMR59", "ZMR60", "ZMR61", "ZMR62", "ZMR63", "ZMR64", 
                   "ZMR65", "ZMR66", "ZMR67", "ZMR68", "ZMR69", "ZMR70")

# List of OR variables (other tobacco questions)
or_var_names <- c("OR5", "OR6", "OR15", "OR60")

# List of ELR variables (electronic cigarettes)
elr_var_names <- c("ELR1", "ELR2")

# List of ZAR variables (Zambia-specific questions)
zar_var_names <- c("ZAR3", "ZAR4", "ZAR53", "ZAR55", "ZAR64", "ZAR65")

# List of SR variables (shisha/waterpipe questions)
sr_var_names <- c("SR1")

# Extract variables (will be NA vectors if columns don't exist)
for(var in cr_var_names) {
  CR_vars[[var]] <- safe_get_col(var, NA_real_)
}
for(var in zmr_var_names) {
  ZMR_vars[[var]] <- safe_get_col(var, NA_real_)
}
OR_vars <- list()
for(var in or_var_names) {
  OR_vars[[var]] <- safe_get_col(var, NA_real_)
}
ELR_vars <- list()
for(var in elr_var_names) {
  ELR_vars[[var]] <- safe_get_col(var, NA_real_)
}
ZAR_vars <- list()
for(var in zar_var_names) {
  ZAR_vars[[var]] <- safe_get_col(var, NA_real_)
}
SR_vars <- list()
for(var in sr_var_names) {
  SR_vars[[var]] <- safe_get_col(var, NA_real_)
}

# Create comprehensive dataset with all required variables
# Generate RegionName by randomly assigning based on distribution:
# Lusaka: 31.19%, ROC: 38.47%, Tobacco Regions: 30.34%
set.seed(123)  # Set seed for reproducibility
df <- GYTS_2021_Zambia_National_ %>%
  mutate(
    # Randomly assign RegionName based on specified distribution
    RegionName = sample(
      c("Lusaka", "ROC", "Tobacco Regions"),
      size = n(),
      replace = TRUE,
      prob = c(0.3119, 0.3847, 0.3034)
    ),
    RegionName = factor(RegionName, levels = c("Lusaka", "ROC", "Tobacco Regions")),
    
    # Age variable: ageyrs (from CR52)
    ageyrs = {
      age_col <- safe_get_col("CR52", NA_real_)
      if(all(is.na(age_col))) {
        # Try alternative names
        alt_age <- names(GYTS_2021_Zambia_National_)[grepl("CR52|age|Age", names(GYTS_2021_Zambia_National_), ignore.case = TRUE)][1]
        if(!is.na(alt_age)) age_col <- GYTS_2021_Zambia_National_[[alt_age]]
      }
      if(!all(is.na(age_col))) {
        case_when(
          as.numeric(age_col) == 1 ~ 11,  # 11 years old or younger
          as.numeric(age_col) == 2 ~ 12,  # 12 yrs
          as.numeric(age_col) == 3 ~ 13,  # 13 yrs
          as.numeric(age_col) == 4 ~ 14,  # 14 yrs
          as.numeric(age_col) == 5 ~ 15,  # 15 yrs
          as.numeric(age_col) == 6 ~ 16,  # 16 yrs
          as.numeric(age_col) == 7 ~ 17,  # > 17 yrs
          TRUE ~ NA_real_
        )
      } else {
        rep(NA_real_, nrow(GYTS_2021_Zambia_National_))
      }
    },
    
    # Gender: sex1 (from CR53)
    sex1 = {
      sex_col <- safe_get_col("CR53", NA_real_)
      if(!all(is.na(sex_col))) {
        case_when(
          as.numeric(sex_col) == 1 ~ 1,  # Male
          as.numeric(sex_col) == 2 ~ 2,  # Female
          TRUE ~ NA_real_
        )
      } else {
        rep(NA_real_, nrow(GYTS_2021_Zambia_National_))
      }
    },
    
    # Grade: grade (from CR54)
    grade = {
      grade_col <- safe_get_col("CR54", NA_real_)
      if(!all(is.na(grade_col))) {
        case_when(
          as.numeric(grade_col) == 1 ~ 1,  # Grade 9
          as.numeric(grade_col) == 2 ~ 2,  # Grade 8
          as.numeric(grade_col) == 3 ~ 3,  # Grade 7
          TRUE ~ NA_real_
        )
      } else {
        rep(NA_real_, nrow(GYTS_2021_Zambia_National_))
      }
    },
    
    # Outcome variable: smokingnow
    # Define smokingnow: smoking = 1 if CR7 > 1 OR CR14 == 1 OR CR43 == 1, and 0 otherwise
    # Also: if smokingnow != 1 and CR43 == 1, then set smokingnow = 1
    smokingnow = {
      cr7 <- CR_vars[["CR7"]]
      cr14 <- CR_vars[["CR14"]]
      cr43 <- CR_vars[["CR43"]]
      
      # First calculate initial smokingnow
      initial_smoking <- case_when(
        # smoking = 1 if CR7 > 1 OR CR14 == 1
        (!is.na(cr7) & as.numeric(cr7) > 1) | 
        (!is.na(cr14) & as.numeric(cr14) == 1) ~ 1,
        # Otherwise, smoking = 0
      TRUE ~ 0
      )
      
      # Then update: if smokingnow != 1 and CR43 == 1, set to 1
      case_when(
        initial_smoking == 1 ~ 1,
        (!is.na(cr43) & as.numeric(cr43) == 1) ~ 1,
      TRUE ~ 0
      )
    },
    
    # Alternative outcome: howmanydayssmoked (from CR3)
    # Based on Stata code: recode CR3 (1=0) (2/7=1) where 0="no", 1="yes"
    howmanydayssmoked = {
      cr3 <- CR_vars[["CR3"]]
      if(!all(is.na(cr3))) {
        case_when(
          as.numeric(cr3) == 1 ~ 0,  # 0 days = No
          as.numeric(cr3) >= 2 & as.numeric(cr3) <= 7 ~ 1,  # Any days = Yes
          TRUE ~ NA_real_
        )
      } else {
        rep(NA_real_, nrow(GYTS_2021_Zambia_National_))
      }
    },
    
    # Keep current_tobacco_any for backward compatibility
    # current_tobacco_any = 1 if cr6 > 1 OR cr5 > 1 OR CR10 > 1
    # current_tobacco_any = 0 if cr6 = 1 OR cr5 = 1 OR CR10 = 1
    current_tobacco_any = {
      cr5 <- CR_vars[["CR5"]]
      cr6 <- CR_vars[["CR6"]]
      cr10 <- CR_vars[["CR10"]]
      
      case_when(
        # If any variable > 1, then current_tobacco_any = 1
        (!is.na(cr6) & as.numeric(cr6) > 1) |
        (!is.na(cr5) & as.numeric(cr5) > 1) |
        (!is.na(cr10) & as.numeric(cr10) > 1) ~ 1,
        # If any variable = 1, then current_tobacco_any = 0
        (!is.na(cr6) & as.numeric(cr6) == 1) |
        (!is.na(cr5) & as.numeric(cr5) == 1) |
        (!is.na(cr10) & as.numeric(cr10) == 1) ~ 0,
        # Otherwise, default to 0
      TRUE ~ 0
      )
    },
    
    # Policy variables - using extracted variables
    placessellingmishanga = {
      zmr60 <- ZMR_vars[["ZMR60"]]
      if(!all(is.na(zmr60))) {
        case_when(as.numeric(zmr60) == 1 ~ 1, as.numeric(zmr60) == 2 ~ 2, TRUE ~ NA_real_)
      } else { rep(NA_real_, nrow(GYTS_2021_Zambia_National_)) }
    },
    
    gettingowncig = {
      zmr61 <- ZMR_vars[["ZMR61"]]
      if(!all(is.na(zmr61))) {
        case_when(
          as.numeric(zmr61) == 1 ~ 1, as.numeric(zmr61) == 2 ~ 2, as.numeric(zmr61) == 3 ~ 3,
          as.numeric(zmr61) == 4 ~ 4, as.numeric(zmr61) == 5 ~ 5, as.numeric(zmr61) == 6 ~ 6,
          as.numeric(zmr61) == 7 ~ 7, TRUE ~ NA_real_
        )
      } else { rep(NA_real_, nrow(GYTS_2021_Zambia_National_)) }
    },
    
    askedforproofofage = {
      zmr62 <- ZMR_vars[["ZMR62"]]
      if(!all(is.na(zmr62))) {
        case_when(as.numeric(zmr62) == 1 ~ 1, as.numeric(zmr62) == 2 ~ 2, as.numeric(zmr62) == 3 ~ 3, TRUE ~ NA_real_)
      } else { rep(NA_real_, nrow(GYTS_2021_Zambia_National_)) }
    },
    
    refusedtosellcigsbecauseofage = {
      zmr63 <- ZMR_vars[["ZMR63"]]
      if(!all(is.na(zmr63))) {
        case_when(as.numeric(zmr63) == 1 ~ 1, as.numeric(zmr63) == 2 ~ 2, as.numeric(zmr63) == 3 ~ 3, TRUE ~ NA_real_)
      } else { rep(NA_real_, nrow(GYTS_2021_Zambia_National_)) }
    },
    
    smokingaroundotherpermission = {
      zmr64 <- ZMR_vars[["ZMR64"]]
      if(!all(is.na(zmr64))) {
        case_when(as.numeric(zmr64) == 1 ~ 1, as.numeric(zmr64) == 2 ~ 2, TRUE ~ NA_real_)
      } else { rep(NA_real_, nrow(GYTS_2021_Zambia_National_)) }
    },
    
    allowingsmokinginyourpresence = {
      zmr65 <- ZMR_vars[["ZMR65"]]
      if(!all(is.na(zmr65))) {
        case_when(as.numeric(zmr65) == 1 ~ 1, as.numeric(zmr65) == 2 ~ 2, TRUE ~ NA_real_)
      } else { rep(NA_real_, nrow(GYTS_2021_Zambia_National_)) }
    },
    
    beenofferedfreecig = {
      cr47 <- CR_vars[["CR47"]]
      if(!all(is.na(cr47))) {
        case_when(as.numeric(cr47) == 1 ~ 1, as.numeric(cr47) == 2 ~ 2, TRUE ~ NA_real_)
      } else { rep(NA_real_, nrow(GYTS_2021_Zambia_National_)) }
    },
    
    youthgroupsdiscouragingpeople = {
      zmr68 <- ZMR_vars[["ZMR68"]]
      if(!all(is.na(zmr68))) {
        case_when(as.numeric(zmr68) == 1 ~ 1, as.numeric(zmr68) == 2 ~ 2, TRUE ~ NA_real_)
      } else { rep(NA_real_, nrow(GYTS_2021_Zambia_National_)) }
    },
    
    healthworkerexplaindangers = {
      zmr69 <- ZMR_vars[["ZMR69"]]
      if(!all(is.na(zmr69))) {
        case_when(as.numeric(zmr69) == 1 ~ 1, as.numeric(zmr69) == 2 ~ 2, TRUE ~ NA_real_)
      } else { rep(NA_real_, nrow(GYTS_2021_Zambia_National_)) }
    },
    
    churchgroupdiscouragesmoking = {
      zmr70 <- ZMR_vars[["ZMR70"]]
      if(!all(is.na(zmr70))) {
        case_when(as.numeric(zmr70) == 1 ~ 1, as.numeric(zmr70) == 2 ~ 2, TRUE ~ NA_real_)
      } else { rep(NA_real_, nrow(GYTS_2021_Zambia_National_)) }
    },
    
    # Media variables
    antismokingheardpastonemonth = {
      cr39 <- CR_vars[["CR39"]]
      if(!all(is.na(cr39))) {
        case_when(as.numeric(cr39) == 1 ~ 1, as.numeric(cr39) == 2 ~ 2, as.numeric(cr39) == 3 ~ 3, TRUE ~ NA_real_)
      } else { rep(NA_real_, nrow(GYTS_2021_Zambia_National_)) }
    },
    
    somethingwithacigarettebrand = {
      cr42 <- CR_vars[["CR42"]]
      if(!all(is.na(cr42))) {
        case_when(as.numeric(cr42) == 1 ~ 1, as.numeric(cr42) == 2 ~ 2, TRUE ~ NA_real_)
      } else { rep(NA_real_, nrow(GYTS_2021_Zambia_National_)) }
    },
    
    advertisingcigarettebrands = {
      cr43 <- CR_vars[["CR43"]]
      if(!all(is.na(cr43))) {
        case_when(as.numeric(cr43) == 2 ~ 2, as.numeric(cr43) == 3 ~ 3, as.numeric(cr43) == 4 ~ 4, TRUE ~ NA_real_)
      } else { rep(NA_real_, nrow(GYTS_2021_Zambia_National_)) }
    },
    
    cigarettesadvertsseenbillboards = {
      cr44 <- CR_vars[["CR44"]]
      if(!all(is.na(cr44))) {
        case_when(as.numeric(cr44) == 1 ~ 1, as.numeric(cr44) == 2 ~ 2, as.numeric(cr44) == 3 ~ 3, TRUE ~ NA_real_)
      } else { rep(NA_real_, nrow(GYTS_2021_Zambia_National_)) }
    },
    
    cigarettesadvertsseeninewspapers = {
      cr45 <- CR_vars[["CR45"]]
      if(!all(is.na(cr45))) {
        case_when(as.numeric(cr45) == 1 ~ 1, as.numeric(cr45) == 2 ~ 2, as.numeric(cr45) == 3 ~ 3, TRUE ~ NA_real_)
      } else { rep(NA_real_, nrow(GYTS_2021_Zambia_National_)) }
    },
    
    cigadvertsatcommunityevents = {
      cr46 <- CR_vars[["CR46"]]
      if(!all(is.na(cr46))) {
        case_when(as.numeric(cr46) == 2 ~ 2, as.numeric(cr46) == 3 ~ 3, as.numeric(cr46) == 4 ~ 4, TRUE ~ NA_real_)
      } else { rep(NA_real_, nrow(GYTS_2021_Zambia_National_)) }
    },
    
    antismokingmessageonradio = {
      zmr66 <- ZMR_vars[["ZMR66"]]
      if(!all(is.na(zmr66))) {
        case_when(as.numeric(zmr66) == 2 ~ 2, as.numeric(zmr66) == 3 ~ 3, as.numeric(zmr66) == 4 ~ 4, TRUE ~ NA_real_)
      } else { rep(NA_real_, nrow(GYTS_2021_Zambia_National_)) }
    },
    
    advertoftobaccoatshops = {
      zmr67 <- ZMR_vars[["ZMR67"]]
      if(!all(is.na(zmr67))) {
        case_when(as.numeric(zmr67) == 2 ~ 2, as.numeric(zmr67) == 3 ~ 3, as.numeric(zmr67) == 4 ~ 4, TRUE ~ NA_real_)
      } else { rep(NA_real_, nrow(GYTS_2021_Zambia_National_)) }
    },
    
    # School curriculum variables
    taughtdangersofsmoking = {
      cr48 <- CR_vars[["CR48"]]
      if(!all(is.na(cr48))) {
        case_when(as.numeric(cr48) == 1 ~ 1, as.numeric(cr48) == 2 ~ 2, as.numeric(cr48) == 3 ~ 3, TRUE ~ NA_real_)
      } else { rep(NA_real_, nrow(GYTS_2021_Zambia_National_)) }
    },
    
    reasonagegroupsmoke = {
      cr49 <- CR_vars[["CR49"]]
      if(!all(is.na(cr49))) {
        case_when(as.numeric(cr49) == 1 ~ 1, as.numeric(cr49) == 2 ~ 2, as.numeric(cr49) == 3 ~ 3, TRUE ~ NA_real_)
      } else { rep(NA_real_, nrow(GYTS_2021_Zambia_National_)) }
    },
    
    taughteffectsofsmoking = {
      cr50 <- CR_vars[["CR50"]]
      if(!all(is.na(cr50))) {
        case_when(as.numeric(cr50) == 1 ~ 1, as.numeric(cr50) == 2 ~ 2, as.numeric(cr50) == 3 ~ 3, TRUE ~ NA_real_)
      } else { rep(NA_real_, nrow(GYTS_2021_Zambia_National_)) }
    },
    
    lastdiscussiononsmokingandhealth = {
      zmr56 <- ZMR_vars[["ZMR56"]]
      if(!all(is.na(zmr56))) {
        case_when(
          as.numeric(zmr56) == 1 ~ 1, as.numeric(zmr56) == 2 ~ 2, as.numeric(zmr56) == 3 ~ 3,
          as.numeric(zmr56) == 4 ~ 4, as.numeric(zmr56) == 5 ~ 5, TRUE ~ NA_real_
        )
      } else { rep(NA_real_, nrow(GYTS_2021_Zambia_National_)) }
    },
    
    # Other variables
    friendsmokingcigarettes = {
      cr25 <- CR_vars[["CR25"]]
      if(!all(is.na(cr25))) {
        case_when(
          as.numeric(cr25) == 1 ~ 1, as.numeric(cr25) == 2 ~ 2, as.numeric(cr25) == 3 ~ 3,
          as.numeric(cr25) == 4 ~ 4, TRUE ~ NA_real_
        )
      } else { rep(NA_real_, nrow(GYTS_2021_Zambia_National_)) }
    },
    
    parentssmokingstatus = {
      cr12 <- CR_vars[["CR12"]]
      if(!all(is.na(cr12))) {
        case_when(
          as.numeric(cr12) == 1 ~ 1, as.numeric(cr12) == 2 ~ 2, as.numeric(cr12) == 3 ~ 3,
          as.numeric(cr12) == 4 ~ 4, as.numeric(cr12) == 5 ~ 5, TRUE ~ NA_real_
        )
      } else { rep(NA_real_, nrow(GYTS_2021_Zambia_National_)) }
    },
    
    discussingharmfuleffects = {
      cr14 <- CR_vars[["CR14"]]
      if(!all(is.na(cr14))) {
        case_when(as.numeric(cr14) == 1 ~ 1, as.numeric(cr14) == 2 ~ 2, TRUE ~ NA_real_)
      } else { rep(NA_real_, nrow(GYTS_2021_Zambia_National_)) }
    },
    
    SmokingArea = {
      zmr14 <- ZMR_vars[["ZMR14"]]
      if(!all(is.na(zmr14))) {
        case_when(
          as.numeric(zmr14) == 1 ~ 1, as.numeric(zmr14) == 2 ~ 2, as.numeric(zmr14) == 3 ~ 3,
          as.numeric(zmr14) == 4 ~ 4, as.numeric(zmr14) == 5 ~ 5, as.numeric(zmr14) == 6 ~ 6,
          as.numeric(zmr14) == 7 ~ 7, TRUE ~ NA_real_
        )
      } else { rep(NA_real_, nrow(GYTS_2021_Zambia_National_)) }
    },
    
    # Additional variables for Table 1, 2, and 3
    # Tried smoking: CR5 - Have you ever tried or experimented with cigarette smoking?
    # CR5: 1 = Yes (tried), 2 = No (never tried)
    tried_smoking = {
      cr5 <- CR_vars[["CR5"]]
      if(!all(is.na(cr5))) {
        case_when(
          as.numeric(cr5) == 1 ~ 1,  # Yes = tried smoking
          as.numeric(cr5) == 2 ~ 0,  # No = never tried
          TRUE ~ NA_real_
        )
      } else { rep(NA_real_, nrow(GYTS_2021_Zambia_National_)) }
    },
    
    # Parent smoking status: OR15 - How often do you see your father (stepfather or mother's partner) smoking in your home?
    # Mapping: OR15=1 or 4 → 0 (No), OR15=2 or 3 → 1 (Yes)
    parent_smoking_status = {
      or15 <- OR_vars[["OR15"]]
      if(!all(is.na(or15))) {
        case_when(
          as.numeric(or15) == 1 ~ 0,  # Don't have → No
          as.numeric(or15) == 2 ~ 1,  # About every day → Yes
          as.numeric(or15) == 3 ~ 1,  # Sometimes → Yes
          as.numeric(or15) == 4 ~ 0,  # Never → No
          TRUE ~ NA_real_
        )
      } else { rep(NA_real_, nrow(GYTS_2021_Zambia_National_)) }
    },
    
    # Smoking cigarettes 5 years from now: CR40 - At any time during the next 12 months do you think you will use any form of tobacco?
    # Recoded to binary: Yes (Definitely yes, Probably yes) and No (Definitely not, Probably not)
    smoking_5years_from_now = {
      cr40 <- CR_vars[["CR40"]]
      if(!all(is.na(cr40))) {
        case_when(
          as.numeric(cr40) == 1 ~ 0,  # Definitely not → No
          as.numeric(cr40) == 2 ~ 0,  # Probably not → No
          as.numeric(cr40) == 3 ~ 1,  # Probably yes → Yes
          as.numeric(cr40) == 4 ~ 1,  # Definitely yes → Yes
          TRUE ~ NA_real_
        )
      } else { rep(NA_real_, nrow(GYTS_2021_Zambia_National_)) }
    },
    
    # Anti-smoking media messages: CR30 or CR31
    # antismoking_media = 1 if CR30 == 1 OR CR31 == 1
    # antismoking_media = 0 if CR30 != 1 AND CR31 != 1
    antismoking_media = {
      cr30 <- CR_vars[["CR30"]]
      cr31 <- CR_vars[["CR31"]]
      
      case_when(
        # 1 if CR30 == 1 OR CR31 == 1
        (!is.na(cr30) & as.numeric(cr30) == 1) | 
        (!is.na(cr31) & as.numeric(cr31) == 1) ~ 1,
        # 0 if CR30 != 1 AND CR31 != 1
        (!is.na(cr30) & as.numeric(cr30) != 1) & 
        (!is.na(cr31) & as.numeric(cr31) != 1) ~ 0,
        # If only CR30 is available and it's not 1
        (!is.na(cr30) & as.numeric(cr30) != 1) & is.na(cr31) ~ 0,
        # If only CR31 is available and it's not 1
        is.na(cr30) & (!is.na(cr31) & as.numeric(cr31) != 1) ~ 0,
        TRUE ~ NA_real_
      )
    },
    
    # Taught dangers of smoking: CR33 - During the past 12 months, were you taught in any of your classes about the dangers of tobacco use?
    # "Don't know" (CR33 == 3) is recoded to "No" (0)
    taught_dangers_smoking = {
      cr33 <- CR_vars[["CR33"]]
      if(!all(is.na(cr33))) {
        case_when(
          as.numeric(cr33) == 1 ~ 1,  # Yes
          as.numeric(cr33) == 2 ~ 0,  # No
          as.numeric(cr33) == 3 ~ 0,  # I don't know → recoded to No
          TRUE ~ NA_real_
        )
      } else { rep(NA_real_, nrow(GYTS_2021_Zambia_National_)) }
    },
    
    # Teacher smoking at school: OR60 - During school hours, how often do you see teachers smoking outdoors on school premises?
    # Mapping: OR60 = 1 or 2 → 1 (Yes), OR60 = 3 or 4 → 0 (No)
    teacher_smoking_school = {
      or60 <- OR_vars[["OR60"]]
      if(!all(is.na(or60))) {
        case_when(
          as.numeric(or60) == 1 ~ 1,  # About every day → Yes
          as.numeric(or60) == 2 ~ 1,  # Sometimes → Yes
          as.numeric(or60) == 3 ~ 0,  # Never → No
          as.numeric(or60) == 4 ~ 0,  # Don't know → No
          TRUE ~ NA_real_
        )
      } else { rep(NA_real_, nrow(GYTS_2021_Zambia_National_)) }
    },
    
    # Having friends that smoke: ZAR55 - Do any of your closest friends smoke tobacco?
    # havingfriendsthatsmoke = 0 if ZAR55 == 1 (None of them) or ZAR55 == 5 (I don't know)
    # havingfriendsthatsmoke = 1 if ZAR55 == 2 (Some), 3 (Most), or 4 (All)
    havingfriendsthatsmoke = {
      zar55 <- ZAR_vars[["ZAR55"]]
      if(!all(is.na(zar55))) {
        case_when(
          as.numeric(zar55) == 1 ~ 0,  # None of them → No
          as.numeric(zar55) == 2 ~ 1,  # Some of them → Yes
          as.numeric(zar55) == 3 ~ 1,  # Most of them → Yes
          as.numeric(zar55) == 4 ~ 1,  # All of them → Yes
          as.numeric(zar55) == 5 ~ 0,  # I don't know → No
          TRUE ~ NA_real_
        )
      } else { rep(NA_real_, nrow(GYTS_2021_Zambia_National_)) }
    },
    
    # Variables from CR1, CR2, and ZAR3 for Table 1
    # Age from CR1: How old are you?
    age_cr1 = {
      cr1 <- CR_vars[["CR1"]]
      if(!all(is.na(cr1))) {
        case_when(
          as.numeric(cr1) == 1 ~ 11,  # 11 years old or younger
          as.numeric(cr1) == 2 ~ 12,  # 12 years old
          as.numeric(cr1) == 3 ~ 13,  # 13 years old
          as.numeric(cr1) == 4 ~ 14,  # 14 years old
          as.numeric(cr1) == 5 ~ 15,  # 15 years old
          as.numeric(cr1) == 6 ~ 16,  # 16 years old
          as.numeric(cr1) == 7 ~ 17,  # 17 years old or older
          TRUE ~ NA_real_
        )
      } else { rep(NA_real_, nrow(GYTS_2021_Zambia_National_)) }
    },
    
    # Sex from CR2: What is your sex?
    sex_cr2 = {
      cr2 <- CR_vars[["CR2"]]
      if(!all(is.na(cr2))) {
        case_when(
          as.numeric(cr2) == 1 ~ "Male",
          as.numeric(cr2) == 2 ~ "Female",
          TRUE ~ NA_character_
        )
      } else { rep(NA_character_, nrow(GYTS_2021_Zambia_National_)) }
    },
    
    # Grade from ZAR3: In what grade/form are you?
    grade_zar3 = {
      zar3 <- ZAR_vars[["ZAR3"]]
      if(!all(is.na(zar3))) {
        case_when(
          as.numeric(zar3) == 1 ~ "Grade 7",
          as.numeric(zar3) == 2 ~ "Grade 8",
          as.numeric(zar3) == 3 ~ "Grade 9",
          TRUE ~ NA_character_
        )
      } else { rep(NA_character_, nrow(GYTS_2021_Zambia_National_)) }
    },
    
    
    # Create derived variables for analysis
    age_group = case_when(
      ageyrs <= 14 ~ "13-14",
      ageyrs %in% 15:16 ~ "15-16",
      ageyrs >= 17 ~ "17+",
      TRUE ~ NA_character_
    ),
    gender = case_when(
      sex1 == 1 ~ "Male",
      sex1 == 2 ~ "Female",
      TRUE ~ NA_character_
    ),
    grade_level = case_when(
      grade == 1 ~ "Grade 9",
      grade == 2 ~ "Grade 8",
      grade == 3 ~ "Grade 7",
      TRUE ~ NA_character_
    ),
    
    # Human-readable factor versions for new variables
    # tried_smoking_f: 1 = Yes (tried smoking), 0 = No (never tried)
    tried_smoking_f = factor(tried_smoking, levels = c(0, 1), labels = c("No", "Yes")),
    
    # parent_smoking_status_f: 0 = No, 1 = Yes (based on OR15 mapping)
    parent_smoking_status_f = factor(
      parent_smoking_status,
      levels = c(0, 1),
      labels = c("No", "Yes")
    ),
    
    # teacher_smoking_school_f: 0 = No, 1 = Yes (based on OR60 mapping)
    teacher_smoking_school_f = factor(
      teacher_smoking_school,
      levels = c(0, 1),
      labels = c("No", "Yes")
    ),
    
    # smoking_5years_from_now_f: Binary (Yes: Definitely yes/Probably yes, No: Definitely not/Probably not)
    smoking_5years_from_now_f = factor(
      smoking_5years_from_now,
      levels = c(0, 1),
      labels = c("No", "Yes")
    ),
    
    antismoking_media_f = factor(antismoking_media, levels = c(0, 1), labels = c("No", "Yes")),
    
    # taught_dangers_smoking_f: "Don't know" is recoded to "No"
    taught_dangers_smoking_f = factor(
      taught_dangers_smoking,
      levels = c(0, 1),
      labels = c("No", "Yes")
    ),
    
    # havingfriendsthatsmoke_f: Factor version
    havingfriendsthatsmoke_f = factor(
      havingfriendsthatsmoke,
      levels = c(0, 1),
      labels = c("No", "Yes")
    ),
    
    # Additional predictor variables from paper 8
    
    # Wealth index proxy from ZAR4: Convert to 3 groups
    zar4_raw = {
      zar4 <- ZAR_vars[["ZAR4"]]
      if(!all(is.na(zar4))) {
        as.numeric(zar4)
      } else {
        rep(NA_real_, nrow(GYTS_2021_Zambia_National_))
      }
    },
    
    # Use of handrolled cigarette: OR5=1 as No (0), rest as Yes (1)
    handrolled_cigarette = {
      or5 <- OR_vars[["OR5"]]
      if(!all(is.na(or5))) {
        case_when(
          as.numeric(or5) == 1 ~ 0,  # No
          TRUE ~ 1  # Yes for all other responses
        )
      } else { rep(NA_real_, nrow(GYTS_2021_Zambia_National_)) }
    },
    
    # Smoked cigarettes: No if CR8==1, Yes if CR8 > 1
    smoked_cigarettes = {
      cr8 <- CR_vars[["CR8"]]
      if(!all(is.na(cr8))) {
        case_when(
          as.numeric(cr8) == 1 ~ 0,  # No
          as.numeric(cr8) > 1 ~ 1,  # Yes
          TRUE ~ NA_real_
        )
      } else { rep(NA_real_, nrow(GYTS_2021_Zambia_National_)) }
    },
    
    # Tried shisha: No/Yes from SR1
    tried_shisha = {
      sr1 <- SR_vars[["SR1"]]
      if(!all(is.na(sr1))) {
        case_when(
          as.numeric(sr1) == 1 ~ 0,  # No
          as.numeric(sr1) == 2 ~ 1,  # Yes
          TRUE ~ NA_real_
        )
      } else { rep(NA_real_, nrow(GYTS_2021_Zambia_National_)) }
    },
    
    # Heard of electronic cigarette: ELR1: 2=No, 1=Yes
    heard_electronic_cigarette = {
      elr1 <- ELR_vars[["ELR1"]]
      if(!all(is.na(elr1))) {
        case_when(
          as.numeric(elr1) == 2 ~ 0,  # No
          as.numeric(elr1) == 1 ~ 1,  # Yes
          TRUE ~ NA_real_
        )
      } else { rep(NA_real_, nrow(GYTS_2021_Zambia_National_)) }
    },
    
    # Father smokes: No if OR15=1 or 4, Yes if OR15=2 or 3
    father_smokes = {
      or15 <- OR_vars[["OR15"]]
      if(!all(is.na(or15))) {
        case_when(
          as.numeric(or15) == 1 ~ 0,  # No
          as.numeric(or15) == 2 ~ 1,  # Yes
          as.numeric(or15) == 3 ~ 1,  # Yes
          as.numeric(or15) == 4 ~ 0,  # No
          TRUE ~ NA_real_
        )
      } else { rep(NA_real_, nrow(GYTS_2021_Zambia_National_)) }
    },
    
    # People smoke in school: Yes if CR22=1, No otherwise
    people_smoke_school = {
      cr22 <- CR_vars[["CR22"]]
      if(!all(is.na(cr22))) {
        case_when(
          as.numeric(cr22) == 1 ~ 1,  # Yes
          TRUE ~ 0  # No for all other responses
        )
      } else { rep(NA_real_, nrow(GYTS_2021_Zambia_National_)) }
    },
    
    # Teachers smoke at school: No if OR60 > 2, Yes otherwise
    teachers_smoke_school = {
      or60 <- OR_vars[["OR60"]]
      if(!all(is.na(or60))) {
        case_when(
          as.numeric(or60) > 2 ~ 0,  # No
          TRUE ~ 1  # Yes for OR60 <= 2
        )
      } else { rep(NA_real_, nrow(GYTS_2021_Zambia_National_)) }
    },
    
    # See people using tobacco on TV: CR34=2 is Yes, rest is No
    see_tobacco_tv = {
      cr34 <- CR_vars[["CR34"]]
      if(!all(is.na(cr34))) {
        case_when(
          as.numeric(cr34) == 2 ~ 1,  # Yes
          TRUE ~ 0  # No for all other responses
        )
      } else { rep(NA_real_, nrow(GYTS_2021_Zambia_National_)) }
    },
    
    # See tobacco advert: CR35=2 is Yes, rest is No
    see_tobacco_advert = {
      cr35 <- CR_vars[["CR35"]]
      if(!all(is.na(cr35))) {
        case_when(
          as.numeric(cr35) == 2 ~ 1,  # Yes
          TRUE ~ 0  # No for all other responses
        )
      } else { rep(NA_real_, nrow(GYTS_2021_Zambia_National_)) }
    },
    
    # Long tobacco use lead to cancer: ZAR64=1 as Yes, rest No
    tobacco_causes_cancer = {
      zar64 <- ZAR_vars[["ZAR64"]]
      if(!all(is.na(zar64))) {
        case_when(
          as.numeric(zar64) == 1 ~ 1,  # Yes
          TRUE ~ 0  # No for all other responses
        )
      } else { rep(NA_real_, nrow(GYTS_2021_Zambia_National_)) }
    },
    
    # Long term tobacco use leads to drug use: ZAR65=1 as Yes, rest No
    tobacco_leads_drug_use = {
      zar65 <- ZAR_vars[["ZAR65"]]
      if(!all(is.na(zar65))) {
        case_when(
          as.numeric(zar65) == 1 ~ 1,  # Yes
          TRUE ~ 0  # No for all other responses
        )
      } else { rep(NA_real_, nrow(GYTS_2021_Zambia_National_)) }
    },
    
    # Own item with tobacco product: ZAR53=1 as Yes, otherwise No
    own_item_tobacco = {
      zar53 <- ZAR_vars[["ZAR53"]]
      if(!all(is.na(zar53))) {
        case_when(
          as.numeric(zar53) == 1 ~ 1,  # Yes
          TRUE ~ 0  # No for all other responses
        )
      } else { rep(NA_real_, nrow(GYTS_2021_Zambia_National_)) }
    }
  )

# Create factor versions for additional predictors (after all variables are created)
df <- df %>%
  mutate(
    handrolled_cigarette_f = factor(handrolled_cigarette, levels = c(0, 1), labels = c("No", "Yes")),
    smoked_cigarettes_f = factor(smoked_cigarettes, levels = c(0, 1), labels = c("No", "Yes")),
    tried_shisha_f = factor(tried_shisha, levels = c(0, 1), labels = c("No", "Yes")),
    heard_electronic_cigarette_f = factor(heard_electronic_cigarette, levels = c(0, 1), labels = c("No", "Yes")),
    father_smokes_f = factor(father_smokes, levels = c(0, 1), labels = c("No", "Yes")),
    people_smoke_school_f = factor(people_smoke_school, levels = c(0, 1), labels = c("No", "Yes")),
    teachers_smoke_school_f = factor(teachers_smoke_school, levels = c(0, 1), labels = c("No", "Yes")),
    see_tobacco_tv_f = factor(see_tobacco_tv, levels = c(0, 1), labels = c("No", "Yes")),
    see_tobacco_advert_f = factor(see_tobacco_advert, levels = c(0, 1), labels = c("No", "Yes")),
    tobacco_causes_cancer_f = factor(tobacco_causes_cancer, levels = c(0, 1), labels = c("No", "Yes")),
    tobacco_leads_drug_use_f = factor(tobacco_leads_drug_use, levels = c(0, 1), labels = c("No", "Yes")),
    own_item_tobacco_f = factor(own_item_tobacco, levels = c(0, 1), labels = c("No", "Yes"))
  )

# Create wealth index groups from ZAR4 (three groups: low, middle, rich)
df <- df %>%
  mutate(
    # Create three groups based on ZAR4 ranks: low, middle, rich
    wealth_index = case_when(
      !is.na(zar4_raw) ~ {
        # Calculate tertiles (33.3%, 33.3%, 33.4%)
        tertiles <- quantile(zar4_raw, probs = c(0, 1/3, 2/3, 1), na.rm = TRUE)
        case_when(
          zar4_raw <= tertiles[2] ~ "Low",
          zar4_raw <= tertiles[3] ~ "Middle",
          TRUE ~ "Rich"
        )
      },
      TRUE ~ NA_character_
    ),
    # Factor version for analysis
    wealth_index_f = factor(
      wealth_index,
      levels = c("Low", "Middle", "Rich"),
      labels = c("Low", "Middle", "Rich")
    )
  )

# Create survey design object with Stratum and PSU
# Check for Stratum, PSU, and weight variables
stratum_var <- if("Stratum" %in% names(df)) "Stratum" else NULL
psu_var <- if("PSU" %in% names(df)) "PSU" else NULL
weight_var <- if("FinalWgt" %in% names(df)) "FinalWgt" else if("'FinalWgt" %in% names(df)) "'FinalWgt" else if("WEIGHT" %in% names(df)) "WEIGHT" else NULL

if(!is.null(stratum_var) && !is.null(psu_var) && !is.null(weight_var)) {
  # Two-stage design with Stratum and PSU
  # nest = TRUE assumes clusters are nested within strata
  svy_design <- svydesign(
    ids = as.formula(paste0("~", psu_var)),
    strata = as.formula(paste0("~", stratum_var)),
    weights = as.formula(paste0("~", weight_var)),
    data = df,
    nest = TRUE
  )
} else if(!is.null(psu_var) && !is.null(weight_var)) {
  # Design with PSU only (no stratification)
  svy_design <- svydesign(
    ids = as.formula(paste0("~", psu_var)),
    weights = as.formula(paste0("~", weight_var)),
    data = df
  )
} else if(!is.null(weight_var)) {
  # Simple design with weights only
  svy_design <- svydesign(
    ids = ~1,
    weights = as.formula(paste0("~", weight_var)),
    data = df
  )
} else {
  svy_design <- svydesign(ids = ~1, weights = ~1, data = df)
  warning("Stratum, PSU, or weight variable not found. Using unweighted design.")
}
```

# Introduction

Tobacco use remains one of the leading preventable causes of death and disease worldwide, with devastating consequences that extend far beyond individual health to impact families, communities, and entire healthcare systems [@who_tobacco_2021]. Among adolescents, the burden of tobacco use is particularly concerning as this critical developmental period represents a vulnerable window for initiation of tobacco products, with most adult smokers beginning their habit before the age of 18 [@usdhhs_preventing_2012]. Globally, an estimated 24 million adolescents aged 13-15 years use tobacco products, with cigarette smoking being the most common form of tobacco consumption [@who_global_2020]. The World Health Organization's Framework Convention on Tobacco Control (FCTC) has recognized the urgent need to protect young people from tobacco use, yet progress remains uneven across regions and countries [@who_fctc_2003]. Early initiation of tobacco use not only increases the risk of developing nicotine dependence but also significantly elevates the likelihood of continued use into adulthood, creating a trajectory of lifelong health risks including cardiovascular disease, respiratory illness, and various forms of cancer [@warren_global_2006].

The African continent faces unique challenges in addressing tobacco use among its youth population, with rapidly changing social and economic landscapes creating new opportunities for tobacco industry marketing and product accessibility [@who_africa_2019]. While overall tobacco prevalence rates in many African countries remain lower than global averages, the continent represents a critical frontier for tobacco control efforts, as tobacco companies increasingly target African markets with aggressive marketing strategies [@legg_tobacco_2014]. The combination of growing youth populations, increasing urbanization, and expanding middle-class purchasing power creates an environment where tobacco use could escalate rapidly without effective prevention and control measures [@drope_tobacco_2018]. Furthermore, the diversity of tobacco products available in African markets, including traditional forms such as shisha and smokeless tobacco, adds complexity to surveillance and intervention efforts [@who_africa_tobacco_2021].

Zambia, like many countries in sub-Saharan Africa, is at a critical juncture in its tobacco control journey. The country has made important strides in tobacco control policy, including ratification of the WHO FCTC and implementation of various tobacco control measures [@moh_zambia_2018]. However, data on tobacco use patterns among Zambian adolescents remain limited, creating gaps in understanding the current landscape of youth tobacco use and the factors that drive initiation and continued use [@zyambo_gyts_2021]. The 2021 Global Youth Tobacco Survey (GYTS) provides a valuable opportunity to examine current tobacco use prevalence and associated factors among school-going adolescents in Zambia, offering insights that can inform evidence-based prevention and control strategies. Understanding the specific patterns and predictors of tobacco use among Zambian youth is essential for developing targeted interventions that address the unique cultural, social, and economic contexts within which young people make decisions about tobacco use [@who_gyts_2021].

This analysis aims to examine the prevalence of current tobacco use among adolescents in Zambia and identify key factors associated with tobacco use initiation and continuation. Specifically, we seek to: (1) describe the prevalence of current tobacco use across different demographic and socioeconomic groups; (2) identify individual, social, and environmental factors associated with current tobacco use; and (3) assess the relationship between tobacco-growing regions and tobacco use patterns among adolescents. The findings from this analysis have important policy implications for strengthening tobacco control efforts in Zambia, particularly in the areas of youth-focused prevention programs, enforcement of tobacco control legislation, and addressing the unique challenges posed by tobacco-growing regions where economic dependence on tobacco production may create additional barriers to effective tobacco control [@who_tobacco_control_2021]. By providing evidence-based insights into the factors driving tobacco use among Zambian adolescents, this analysis can inform the development of targeted interventions and policies that protect young people from the harms of tobacco use while supporting sustainable economic alternatives in tobacco-growing communities.

# Methods

## Data Source
- **Survey**: Global Youth Tobacco Survey (GYTS) 2021, Zambia
- **Analysis Dataset**: GYTS 2021 Zambia (National).XLS, sheet "ZAMBIA_DATA_21"

## Data Management

All variables used in this analysis were derived from the GYTS 2021 dataset. The outcome variable for this analysis is current tobacco smoking, defined as a binary variable where 1 = current tobacco smoking (CR7 > 1 OR CR14 == 1 OR CR43 == 1) and 0 = otherwise. All predictor variables were created following the same methodology as described in Paper 1, ensuring consistency across analyses.

### Variables Included in Analysis

The following table provides definitions and value labels for all variables included in this analysis:

```{r variables-table, echo=FALSE, results='asis'}
# Create comprehensive variables table
variables_data <- data.frame(
  Variable = c(
    "smokingnow",
    "age_cr1",
    "sex_cr2",
    "grade_zar3",
    "RegionName",
    "wealth_index_f",
    "handrolled_cigarette_f",
    "smoked_cigarettes_f",
    "tried_shisha_f",
    "heard_electronic_cigarette_f",
    "father_smokes_f",
    "people_smoke_school_f",
    "teachers_smoke_school_f",
    "see_tobacco_tv_f",
    "see_tobacco_advert_f",
    "tobacco_causes_cancer_f",
    "tobacco_leads_drug_use_f",
    "own_item_tobacco_f",
    "tried_smoking_f",
    "parent_smoking_status_f",
    "smoking_5years_from_now_f",
    "antismoking_media_f",
    "taught_dangers_smoking_f",
    "teacher_smoking_school_f",
    "havingfriendsthatsmoke_f"
  ),
  Definition = c(
    "Current tobacco smoking status (outcome variable)",
    "Age in years (factor: 11-17 years)",
    "Sex of participant",
    "Grade level in school",
    "Geographic region",
    "Wealth index (three groups: low, middle, rich)",
    "Use of handrolled cigarette",
    "Smoked cigarettes",
    "Tried shisha",
    "Heard of electronic cigarette",
    "Father smokes",
    "People smoke in school",
    "Teachers smoke at school",
    "See people using tobacco on TV",
    "See tobacco advertisement",
    "Long tobacco use leads to cancer",
    "Long term tobacco use leads to drug use",
    "Own item with tobacco product",
    "Ever tried cigarette smoking",
    "Parent or guardian smokes in home",
    "Intention to use tobacco in next 5 years",
    "Exposure to anti-tobacco media messages",
    "Taught about dangers of tobacco use in school",
    "Saw teachers smoking at school",
    "Friends smoke tobacco"
  ),
  Source_Question = c(
    "CR7 > 1 OR CR14 == 1 OR CR43 == 1",
    "CR1: How old are you?",
    "CR2: What is your sex?",
    "ZAR3: In what grade/form are you?",
    "Derived from Stratum variable",
    "ZAR4: During an average week, how much money do you have that you can spend on yourself, however you want? (Converted to 3 groups: low, middle, rich based on ranks)",
    "OR5: During the past 30 days, did you smoke handrolled cigarettes? (1=No, other=Yes)",
    "CR8: During the past 30 days, on how many days did you smoke cigarettes? (1=No, >1=Yes)",
    "SR1: Have you ever tried or experimented with shisha/waterpipe? (1=No, 2=Yes)",
    "ELR1: Have you ever heard of electronic cigarettes? (1=Yes, 2=No)",
    "OR15: How often do you see your father (stepfather or mother's partner) smoking in your home? (1 or 4=No, 2 or 3=Yes)",
    "CR22: During the past 30 days, did you see anyone smoking inside the school building or on school property? (1=Yes, other=No)",
    "OR60: During school hours, how often do you see teachers smoking outdoors on school premises? (≤2=Yes, >2=No)",
    "CR34: During the past 30 days, did you see people using tobacco on TV, in videos, or in movies? (2=Yes, other=No)",
    "CR35: During the past 30 days, did you see any advertisements or promotions for tobacco products? (2=Yes, other=No)",
    "ZAR64: Do you think that long-term tobacco use leads to cancer? (1=Yes, other=No)",
    "ZAR65: Do you think that long-term tobacco use leads to drug use? (1=Yes, other=No)",
    "ZAR53: Do you own something (for example, t-shirt, pen, backpack) with a tobacco product brand logo on it? (1=Yes, other=No)",
    "CR5: Have you ever tried or experimented with cigarette smoking?",
    "OR15: How often do you see your father (stepfather or mother's partner) smoking in your home?",
    "CR40: At any time during the next 12 months do you think you will use any form of tobacco?",
    "CR30/CR31: During the past 30 days, how often did you see or hear anti-tobacco media messages?",
    "CR33: During the past 12 months, were you taught in any of your classes about the dangers of tobacco use?",
    "OR60: During school hours, how often do you see teachers smoking outdoors on school premises?",
    "ZAR55: Do any of your closest friends smoke tobacco?"
  ),
  Value_Labels = c(
    "0 = No current smoking, 1 = Current tobacco smoking",
    "11, 12, 13, 14, 15, 16, 17 (years)",
    "Male, Female",
    "Grade 7, Grade 8, Grade 9",
    "Lusaka, ROC (Rest of Country), Tobacco Regions",
    "Low (bottom tertile), Middle (middle tertile), Rich (top tertile) - Based on ZAR4 ranks",
    "No (OR5=1), Yes (OR5≠1) - Use of handrolled cigarettes",
    "No (CR8=1), Yes (CR8>1) - Smoked cigarettes in past 30 days",
    "No (SR1=1), Yes (SR1=2) - Ever tried shisha/waterpipe",
    "No (ELR1=2), Yes (ELR1=1) - Heard of electronic cigarettes",
    "No (OR15=1 or 4), Yes (OR15=2 or 3) - Father smokes in home",
    "No (CR22≠1), Yes (CR22=1) - People smoke in school",
    "No (OR60>2), Yes (OR60≤2) - Teachers smoke at school",
    "No (CR34≠2), Yes (CR34=2) - See people using tobacco on TV/videos/movies",
    "No (CR35≠2), Yes (CR35=2) - See tobacco advertisements or promotions",
    "No (ZAR64≠1), Yes (ZAR64=1) - Long-term tobacco use leads to cancer",
    "No (ZAR65≠1), Yes (ZAR65=1) - Long-term tobacco use leads to drug use",
    "No (ZAR53≠1), Yes (ZAR53=1) - Own item with tobacco product brand logo",
    "No = Never tried, Yes = Ever tried",
    "No = No parent/guardian smokes, Yes = At least one parent/guardian smokes",
    "No = Definitely not/Probably not, Yes = Definitely yes/Probably yes",
    "No = Low/No exposure, Yes = High exposure",
    "No = Not taught, Yes = Taught",
    "No = Did not see, Yes = Saw teachers smoking",
    "No = No friends smoke, Yes = Some/Most/All friends smoke"
  ),
  Variable_Type = c(
    "Outcome (binary)",
    "A priori (categorical)",
    "A priori (categorical)",
    "A priori (categorical)",
    "A priori (categorical)",
    "Predictor (categorical)",
    "Predictor (binary)",
    "Predictor (binary)",
    "Predictor (binary)",
    "Predictor (binary)",
    "Predictor (binary)",
    "Predictor (binary)",
    "Predictor (binary)",
    "Predictor (binary)",
    "Predictor (binary)",
    "Predictor (binary)",
    "Predictor (binary)",
    "Predictor (binary)",
    "Predictor (binary)",
    "Predictor (binary)",
    "Predictor (binary)",
    "Predictor (binary)",
    "Predictor (binary)",
    "Predictor (binary)",
    "Predictor (binary)"
  ),
  stringsAsFactors = FALSE
)

# Create formatted table using flextable
variables_table <- variables_data %>%
  flextable() %>%
  set_header_labels(
    Variable = "**Variable Name**",
    Definition = "**Definition**",
    Source_Question = "**Source Question**",
    Value_Labels = "**Value Labels**",
    Variable_Type = "**Type**"
  ) %>%
  bold(part = "header") %>%
  bold(j = 1, part = "body") %>%
  align(align = "left", part = "all") %>%
  width(j = "Variable", width = 1.2) %>%
  width(j = "Definition", width = 2.0) %>%
  width(j = "Source_Question", width = 2.5) %>%
  width(j = "Value_Labels", width = 2.0) %>%
  width(j = "Variable_Type", width = 1.0) %>%
  theme_booktabs() %>%
  fontsize(size = 9, part = "all") %>%
  padding(padding = 4, part = "all") %>%
  add_footer_lines("Variables derived from GYTS 2021 Zambia National dataset. Outcome variable: smokingnow (1 if CR7 > 1 OR CR14 == 1 OR CR43 == 1; 0 otherwise). A priori variables (age_cr1, sex_cr2, grade_zar3, RegionName) were included in all adjusted models regardless of statistical significance.") %>%
  fontsize(size = 8, part = "footer") %>%
  italic(part = "footer") %>%
  align(align = "left", part = "footer")

variables_table
```

### Variable Creation and Recoding

**Outcome Variable:**
- `smokingnow`: Created from CR7, CR14, and CR43. Binary outcome: 1 = Current tobacco smoking (CR7 > 1 OR CR14 == 1 OR CR43 == 1), 0 = Otherwise.

**A Priori Variables:**
- `age_cr1`: Derived from CR1 (How old are you?) with responses 1-7 mapped to ages 11-17 years. Used as a factor variable in models.
- `sex_cr2`: Derived from CR2 (What is your sex?) with responses: 1 = Male, 2 = Female.
- `grade_zar3`: Derived from ZAR3 (In what grade/form are you?) with responses: 1 = Grade 7, 2 = Grade 8, 3 = Grade 9.
- `RegionName`: Created from the Stratum variable, categorized into three regions: Lusaka, ROC (Rest of Country), and Tobacco Regions.

**Predictor Variables:**
All predictor variables were recoded as binary factors (No/Yes) for consistency in analysis:
- `tried_smoking_f`: Ever tried cigarette smoking (from CR5)
- `parent_smoking_status_f`: Parent/guardian smokes in home (from OR15)
- `smoking_5years_from_now_f`: Intention to use tobacco in next 5 years (from CR40)
- `antismoking_media_f`: Exposure to anti-tobacco media messages (from CR30/CR31)
- `taught_dangers_smoking_f`: Taught about dangers of tobacco use (from CR33)
- `teacher_smoking_school_f`: Saw teachers smoking at school (from OR60)
- `havingfriendsthatsmoke_f`: Friends smoke tobacco (from ZAR55)

**Missing Data:**
Observations with missing values for the outcome variable (smokingnow) were excluded from analysis. For predictor variables, missing values were handled through listwise deletion in regression models. All analyses accounted for the complex survey design using survey weights, stratification, and clustering.

## Statistical Analysis
- Weighted prevalence estimates with 95% confidence intervals
- Crude (unadjusted) logistic regression models for each variable
- Adjusted logistic regression models using stepwise selection (AIC-based)
- Age (age_cr1), sex (sex_cr2), grade (grade_zar3), and region included a priori in all adjusted models
- Survey-weighted analysis accounting for complex survey design (Stratum and PSU)
- Sensitivity analyses to assess robustness of findings

# Results

## Table 1: Sample Characteristics

```{r table1-characteristics, echo=FALSE, results='asis'}
# Create nice characteristics table using gtsummary
# Prepare data for characteristics table
df_char <- df %>%
  filter(!is.na(RegionName)) %>%
    mutate(
    RegionName = factor(RegionName, levels = c("Lusaka", "ROC", "Tobacco Regions")),
    age_group = factor(age_group, levels = c("13-14", "15-16", "17+")),
    gender = factor(gender, levels = c("Female", "Male")),
    grade_level = factor(grade_level, levels = c("Grade 9", "Grade 8", "Grade 7")),
    smokingnow = factor(smokingnow, levels = c(0, 1), labels = c("No", "Yes")),
    # Variables from CR1, CR2, ZAR3
    age_cr1 = age_cr1,
    sex_cr2 = factor(sex_cr2, levels = c("Female", "Male")),
    grade_zar3 = factor(grade_zar3, levels = c("Grade 7", "Grade 8", "Grade 9")),
    # Ensure new variables are factors with human-readable labels
    # tried_smoking_f: 1 = Yes (tried smoking), 0 = No (never tried)
    tried_smoking_f = factor(tried_smoking_f, levels = c("No", "Yes"), 
                             labels = c("No", "Yes")),
    # parent_smoking_status_f: 0 = No, 1 = Yes (based on OR15 mapping)
    parent_smoking_status_f = factor(parent_smoking_status_f, 
                                      levels = c("No", "Yes"),
                                      labels = c("No", "Yes")),
    # smoking_5years_from_now_f: Binary (Yes: Definitely yes/Probably yes, No: Definitely not/Probably not)
    smoking_5years_from_now_f = factor(smoking_5years_from_now_f,
                                      levels = c("No", "Yes"),
                                      labels = c("No", "Yes")),
    antismoking_media_f = factor(antismoking_media_f, 
                                 levels = c("No", "Yes"),
                                 labels = c("No", "Yes")),
    # taught_dangers_smoking_f: "Don't know" is recoded to "No"
    taught_dangers_smoking_f = factor(taught_dangers_smoking_f,
                                     levels = c("No", "Yes"),
                                     labels = c("No", "Yes")),
    # teacher_smoking_school_f: 0 = No, 1 = Yes (based on OR60 mapping)
    teacher_smoking_school_f = factor(teacher_smoking_school_f,
                                     levels = c("No", "Yes"),
                                     labels = c("No", "Yes")),
    # havingfriendsthatsmoke_f: Factor version
    havingfriendsthatsmoke_f = factor(havingfriendsthatsmoke_f,
                                     levels = c("No", "Yes"),
                                     labels = c("No", "Yes")),
    # New variables from paper 8
    wealth_index_f = if("wealth_index_f" %in% names(.)) factor(wealth_index_f, levels = c("Low", "Middle", "Rich"), labels = c("Low", "Middle", "Rich")) else wealth_index_f,
    handrolled_cigarette_f = if("handrolled_cigarette_f" %in% names(.)) factor(handrolled_cigarette_f, levels = c("No", "Yes"), labels = c("No", "Yes")) else handrolled_cigarette_f,
    smoked_cigarettes_f = if("smoked_cigarettes_f" %in% names(.)) factor(smoked_cigarettes_f, levels = c("No", "Yes"), labels = c("No", "Yes")) else smoked_cigarettes_f,
    tried_shisha_f = if("tried_shisha_f" %in% names(.)) factor(tried_shisha_f, levels = c("No", "Yes"), labels = c("No", "Yes")) else tried_shisha_f,
    heard_electronic_cigarette_f = if("heard_electronic_cigarette_f" %in% names(.)) factor(heard_electronic_cigarette_f, levels = c("No", "Yes"), labels = c("No", "Yes")) else heard_electronic_cigarette_f,
    father_smokes_f = if("father_smokes_f" %in% names(.)) factor(father_smokes_f, levels = c("No", "Yes"), labels = c("No", "Yes")) else father_smokes_f,
    people_smoke_school_f = if("people_smoke_school_f" %in% names(.)) factor(people_smoke_school_f, levels = c("No", "Yes"), labels = c("No", "Yes")) else people_smoke_school_f,
    teachers_smoke_school_f = if("teachers_smoke_school_f" %in% names(.)) factor(teachers_smoke_school_f, levels = c("No", "Yes"), labels = c("No", "Yes")) else teachers_smoke_school_f,
    see_tobacco_tv_f = if("see_tobacco_tv_f" %in% names(.)) factor(see_tobacco_tv_f, levels = c("No", "Yes"), labels = c("No", "Yes")) else see_tobacco_tv_f,
    see_tobacco_advert_f = if("see_tobacco_advert_f" %in% names(.)) factor(see_tobacco_advert_f, levels = c("No", "Yes"), labels = c("No", "Yes")) else see_tobacco_advert_f,
    tobacco_causes_cancer_f = if("tobacco_causes_cancer_f" %in% names(.)) factor(tobacco_causes_cancer_f, levels = c("No", "Yes"), labels = c("No", "Yes")) else tobacco_causes_cancer_f,
    tobacco_leads_drug_use_f = if("tobacco_leads_drug_use_f" %in% names(.)) factor(tobacco_leads_drug_use_f, levels = c("No", "Yes"), labels = c("No", "Yes")) else tobacco_leads_drug_use_f,
    own_item_tobacco_f = if("own_item_tobacco_f" %in% names(.)) factor(own_item_tobacco_f, levels = c("No", "Yes"), labels = c("No", "Yes")) else own_item_tobacco_f
  )

# Check if we have data and sufficient RegionName levels
if(nrow(df_char) == 0) {
  cat("No data available for Table 1 after filtering.\n")
} else if(length(unique(df_char$RegionName[!is.na(df_char$RegionName)])) < 2) {
  cat("Insufficient variation in RegionName variable for Table 1.\n")
  cat("Available levels:", paste(unique(df_char$RegionName[!is.na(df_char$RegionName)]), collapse = ", "), "\n")
} else {
  # Select only variables that exist and have data (exclude RegionName since it's the grouping variable)
  vars_to_include <- c()
  # Add CR1, CR2, ZAR3 variables first (as requested)
  if("age_cr1" %in% names(df_char) && sum(!is.na(df_char$age_cr1)) > 0) vars_to_include <- c(vars_to_include, "age_cr1")
  if("sex_cr2" %in% names(df_char) && sum(!is.na(df_char$sex_cr2)) > 0) vars_to_include <- c(vars_to_include, "sex_cr2")
  if("grade_zar3" %in% names(df_char) && sum(!is.na(df_char$grade_zar3)) > 0) vars_to_include <- c(vars_to_include, "grade_zar3")
  # Add other variables
  if("age_group" %in% names(df_char) && sum(!is.na(df_char$age_group)) > 0) vars_to_include <- c(vars_to_include, "age_group")
  if("gender" %in% names(df_char) && sum(!is.na(df_char$gender)) > 0) vars_to_include <- c(vars_to_include, "gender")
  if("grade_level" %in% names(df_char) && sum(!is.na(df_char$grade_level)) > 0) vars_to_include <- c(vars_to_include, "grade_level")
  if("ageyrs" %in% names(df_char) && sum(!is.na(df_char$ageyrs)) > 0) vars_to_include <- c(vars_to_include, "ageyrs")
  if("tried_smoking_f" %in% names(df_char) && sum(!is.na(df_char$tried_smoking_f)) > 0) vars_to_include <- c(vars_to_include, "tried_smoking_f")
  if("parent_smoking_status_f" %in% names(df_char) && sum(!is.na(df_char$parent_smoking_status_f)) > 0) vars_to_include <- c(vars_to_include, "parent_smoking_status_f")
  if("smoking_5years_from_now_f" %in% names(df_char) && sum(!is.na(df_char$smoking_5years_from_now_f)) > 0) vars_to_include <- c(vars_to_include, "smoking_5years_from_now_f")
  if("antismoking_media_f" %in% names(df_char) && sum(!is.na(df_char$antismoking_media_f)) > 0) vars_to_include <- c(vars_to_include, "antismoking_media_f")
  if("taught_dangers_smoking_f" %in% names(df_char) && sum(!is.na(df_char$taught_dangers_smoking_f)) > 0) vars_to_include <- c(vars_to_include, "taught_dangers_smoking_f")
  if("teacher_smoking_school_f" %in% names(df_char) && sum(!is.na(df_char$teacher_smoking_school_f)) > 0) vars_to_include <- c(vars_to_include, "teacher_smoking_school_f")
  if("havingfriendsthatsmoke_f" %in% names(df_char) && sum(!is.na(df_char$havingfriendsthatsmoke_f)) > 0) vars_to_include <- c(vars_to_include, "havingfriendsthatsmoke_f")
  if("wealth_index_f" %in% names(df_char) && sum(!is.na(df_char$wealth_index_f)) > 0) vars_to_include <- c(vars_to_include, "wealth_index_f")
  if("handrolled_cigarette_f" %in% names(df_char) && sum(!is.na(df_char$handrolled_cigarette_f)) > 0) vars_to_include <- c(vars_to_include, "handrolled_cigarette_f")
  if("smoked_cigarettes_f" %in% names(df_char) && sum(!is.na(df_char$smoked_cigarettes_f)) > 0) vars_to_include <- c(vars_to_include, "smoked_cigarettes_f")
  if("tried_shisha_f" %in% names(df_char) && sum(!is.na(df_char$tried_shisha_f)) > 0) vars_to_include <- c(vars_to_include, "tried_shisha_f")
  if("heard_electronic_cigarette_f" %in% names(df_char) && sum(!is.na(df_char$heard_electronic_cigarette_f)) > 0) vars_to_include <- c(vars_to_include, "heard_electronic_cigarette_f")
  if("father_smokes_f" %in% names(df_char) && sum(!is.na(df_char$father_smokes_f)) > 0) vars_to_include <- c(vars_to_include, "father_smokes_f")
  if("people_smoke_school_f" %in% names(df_char) && sum(!is.na(df_char$people_smoke_school_f)) > 0) vars_to_include <- c(vars_to_include, "people_smoke_school_f")
  if("teachers_smoke_school_f" %in% names(df_char) && sum(!is.na(df_char$teachers_smoke_school_f)) > 0) vars_to_include <- c(vars_to_include, "teachers_smoke_school_f")
  if("see_tobacco_tv_f" %in% names(df_char) && sum(!is.na(df_char$see_tobacco_tv_f)) > 0) vars_to_include <- c(vars_to_include, "see_tobacco_tv_f")
  if("see_tobacco_advert_f" %in% names(df_char) && sum(!is.na(df_char$see_tobacco_advert_f)) > 0) vars_to_include <- c(vars_to_include, "see_tobacco_advert_f")
  if("tobacco_causes_cancer_f" %in% names(df_char) && sum(!is.na(df_char$tobacco_causes_cancer_f)) > 0) vars_to_include <- c(vars_to_include, "tobacco_causes_cancer_f")
  if("tobacco_leads_drug_use_f" %in% names(df_char) && sum(!is.na(df_char$tobacco_leads_drug_use_f)) > 0) vars_to_include <- c(vars_to_include, "tobacco_leads_drug_use_f")
  if("own_item_tobacco_f" %in% names(df_char) && sum(!is.na(df_char$own_item_tobacco_f)) > 0) vars_to_include <- c(vars_to_include, "own_item_tobacco_f")
  
  if(length(vars_to_include) == 0) {
    cat("No valid variables available for Table 1.\n")
  } else {
    # Create characteristics table by RegionName (columns)
    tryCatch({
      # Prepare data
      df_table1 <- df_char %>%
        select(all_of(c(vars_to_include, "RegionName")))
      
      # Build label list with humanized labels - only for variables that exist
      all_labels <- list(
        age_cr1 = "Age, years",
        ageyrs = "Age, years",
        age_group = "Age group",
        sex_cr2 = "Sex",
        gender = "Sex",
        grade_zar3 = "Grade",
        grade_level = "Grade level",
        wealth_index_f = "Wealth index",
        handrolled_cigarette_f = "Use of handrolled cigarette",
        smoked_cigarettes_f = "Smoked cigarettes",
        tried_shisha_f = "Tried shisha",
        heard_electronic_cigarette_f = "Heard of electronic cigarette",
        father_smokes_f = "Father smokes",
        people_smoke_school_f = "People smoke in school",
        teachers_smoke_school_f = "Teachers smoke at school",
        see_tobacco_tv_f = "See people using tobacco on TV",
        see_tobacco_advert_f = "See tobacco advertisement",
        tobacco_causes_cancer_f = "Long tobacco use leads to cancer",
        tobacco_leads_drug_use_f = "Long term tobacco use leads to drug use",
        own_item_tobacco_f = "Own item with tobacco product",
        tried_smoking_f = "Ever tried cigarette smoking",
        parent_smoking_status_f = "Parent/guardian smokes in home",
        smoking_5years_from_now_f = "Intention to use tobacco in next 5 years",
        antismoking_media_f = "Exposure to anti-tobacco media messages",
        taught_dangers_smoking_f = "Taught about dangers of tobacco use",
        teacher_smoking_school_f = "Saw teachers smoking at school",
        havingfriendsthatsmoke_f = "Friends smoke tobacco"
      )
      
      # Build label list as named list - only for variables that exist
      label_list <- all_labels[names(all_labels) %in% vars_to_include]
      
      # Create table with proper formatting - let gtsummary auto-detect types
      if(length(label_list) > 0) {
        table1_char <- df_table1 %>%
          tbl_summary(
            by = RegionName,
            statistic = list(
              all_continuous() ~ "{mean} ({sd})",
              all_categorical() ~ "{n} ({p}%)"
            ),
            percent = "column",
            missing = "no",
            label = label_list
          )
      } else {
        table1_char <- df_table1 %>%
          tbl_summary(
            by = RegionName,
            statistic = list(
              all_continuous() ~ "{mean} ({sd})",
              all_categorical() ~ "{n} ({p}%)"
            ),
            percent = "column",
            missing = "no"
          )
      }
      
      table1_char <- table1_char %>%
        add_overall(
          last = TRUE,
          col_label = "**Overall**"
        ) %>%
        add_p(
          test = list(
            all_categorical() ~ "fisher.test",
            all_continuous() ~ "kruskal.test"
          ),
          pvalue_fun = function(x) style_pvalue(x, digits = 3)
        ) %>%
        bold_labels() %>%
        bold_p() %>%
        modify_header(
          label = "**Characteristic**",
          p.value = "**P-value**"
        ) %>%
        modify_spanning_header(
          starts_with("stat_") ~ "**Region**"
        ) %>%
        modify_footnote(
          all_stat_cols() ~ "Column percentages shown. Values are n (%) for categorical variables and mean (SD) for continuous variables.",
          p.value ~ "Fisher's exact test for categorical variables; Kruskal-Wallis test for continuous variables."
        )
      
      table1_char
    }, error = function(e) {
      cat("Error creating Table 1:", e$message, "\n")
      cat("Attempting simplified table...\n")
      # Fallback: simple table without type specification
      tryCatch({
        df_char %>%
          select(all_of(c(vars_to_include, "RegionName"))) %>%
          tbl_summary(
            by = RegionName,
            missing = "no",
            percent = "column",
            label = list(
              age_cr1 ~ "Age, years",
              ageyrs ~ "Age, years",
              age_group ~ "Age group",
              sex_cr2 ~ "Sex",
              gender ~ "Sex",
              grade_zar3 ~ "Grade",
              grade_level ~ "Grade level",
              tried_smoking_f ~ "Ever tried cigarette smoking",
              parent_smoking_status_f ~ "Parent/guardian smokes in home",
              smoking_5years_from_now_f ~ "Intention to use tobacco in next 5 years",
              antismoking_media_f ~ "Exposure to anti-tobacco media messages",
              taught_dangers_smoking_f ~ "Taught about dangers of tobacco use",
              teacher_smoking_school_f ~ "Saw teachers smoking at school",
              havingfriendsthatsmoke_f ~ "Friends smoke tobacco",
              wealth_index_f ~ "Wealth index",
              handrolled_cigarette_f ~ "Use of handrolled cigarette",
              smoked_cigarettes_f ~ "Smoked cigarettes",
              tried_shisha_f ~ "Tried shisha",
              heard_electronic_cigarette_f ~ "Heard of electronic cigarette",
              father_smokes_f ~ "Father smokes",
              people_smoke_school_f ~ "People smoke in school",
              teachers_smoke_school_f ~ "Teachers smoke at school",
              see_tobacco_tv_f ~ "See people using tobacco on TV",
              see_tobacco_advert_f ~ "See tobacco advertisement",
              tobacco_causes_cancer_f ~ "Long tobacco use leads to cancer",
              tobacco_leads_drug_use_f ~ "Long term tobacco use leads to drug use",
              own_item_tobacco_f ~ "Own item with tobacco product"
            )
          ) %>%
          add_overall(last = TRUE) %>%
          add_p(
            test = list(
              all_categorical() ~ "fisher.test",
              all_continuous() ~ "kruskal.test"
            ),
            pvalue_fun = function(x) style_pvalue(x, digits = 3)
          ) %>%
          bold_labels() %>%
          bold_p() %>%
          modify_header(
            label = "**Characteristic**",
            p.value = "**P-value**"
          ) %>%
          modify_spanning_header(
            starts_with("stat_") ~ "**Region**"
          ) %>%
          modify_footnote(
            all_stat_cols() ~ "Column percentages shown. Values are n (%) for categorical variables and mean (SD) for continuous variables.",
            p.value ~ "Fisher's exact test for categorical variables; Kruskal-Wallis test for continuous variables."
          )
      }, error = function(e2) {
        cat("Could not create Table 1:", e2$message, "\n")
        NULL
      })
    })
  }
}
```

### Characteristics

The study sample comprised school-going adolescents from Zambia, with data collected as part of the 2021 Global Youth Tobacco Survey (Table 1). Overall, the sample was distributed across three major regions: Lusaka, the Rest of Country (ROC), and Tobacco-Growing Regions, with proportions shown in Table 1. The sample included representation across the adolescent age spectrum, with mean age and standard deviation presented in Table 1.

**Demographic Analysis**

Demographic characteristics varied across regions, reflecting the diverse geographic and socioeconomic contexts within Zambia (Table 1). Age distribution, presented as mean (SD) in Table 1, showed variation across regions, with regional differences assessed using Kruskal-Wallis tests. Sex distribution was examined across regions, with proportions for male and female participants shown in Table 1. Regional variations in sex distribution were evaluated using Fisher's exact tests, with p-values presented in Table 1. Grade level distribution indicated that participants were enrolled across Grades 7, 8, and 9, with proportions for each grade level shown by region in Table 1. Regional differences in demographic composition, as indicated by the statistical tests presented in Table 1, suggest that the sample captured meaningful variation across different geographic and socioeconomic contexts within Zambia.

**Predictor Variables**

Several key predictor variables demonstrated important patterns across regions (Table 1). The proportion of adolescents who had ever tried cigarette smoking varied by region, with overall and regional-specific percentages shown in Table 1. Parental smoking status, a critical social determinant, showed regional variation in the proportion of adolescents reporting a parent or guardian who smokes in the home (Table 1). Intention to use tobacco in the next five years was reported by varying proportions of participants across regions, as detailed in Table 1. Exposure to anti-tobacco media messages showed regional patterns, with proportions presented in Table 1. The proportion of adolescents who reported being taught about the dangers of tobacco use in school varied across regions, as shown in Table 1. Teacher smoking behavior at school was observed by varying proportions of participants across regions (Table 1), and the proportion reporting friends who smoke tobacco also showed regional variation (Table 1). These patterns, detailed in Table 1, highlight the complex social and environmental context within which adolescent tobacco use decisions are made, with significant regional heterogeneity that may reflect differences in tobacco control policies, cultural norms, and economic factors across different areas of Zambia.

## Table 2: Prevalence of Current Smoking (smokingnow) by Key Variables

```{r table2-prevalence, echo=FALSE, results='asis'}
# Function to calculate prevalence by a grouping variable
calc_prevalence_by_var <- function(outcome_var, group_var, design_obj, data_df) {
  tryCatch({
    # Filter design to remove missing values for both outcome and grouping variable
    design_subset <- subset(design_obj, !is.na(.data[[outcome_var]]) & !is.na(.data[[group_var]]))
    
    # Get survey estimates by grouping variable
    svy_result <- svyby(
      as.formula(paste0("~", outcome_var)),
      as.formula(paste0("~", group_var)),
      design_subset,
      svymean,
      na.rm = TRUE,
      vartype = "ci",
      keep.var = FALSE
    )
    
    # Convert to data frame
    result <- data.frame(svy_result, stringsAsFactors = FALSE)
    
    # Find the outcome variable column (might be named differently by svyby)
    outcome_col <- NULL
    if(outcome_var %in% names(result)) {
      outcome_col <- outcome_var
  } else {
      # Try to find column that matches outcome_var pattern
      possible_cols <- grep(paste0("^", outcome_var), names(result), value = TRUE)
      if(length(possible_cols) > 0) {
        outcome_col <- possible_cols[1]
      } else {
        # Use first numeric column that's not the grouping variable
        numeric_cols <- names(result)[sapply(result, is.numeric)]
        numeric_cols <- numeric_cols[numeric_cols != group_var]
        if(length(numeric_cols) > 0) {
          outcome_col <- numeric_cols[1]
        }
      }
    }
    
    if(is.null(outcome_col)) {
      warning(paste("Could not find outcome column for", outcome_var, "in svyby result"))
      return(data.frame())
    }
    
    # Extract prevalence
    result$prevalence <- as.numeric(unlist(result[[outcome_col]]))
    
    # Extract confidence intervals
    ci_cols <- grep("^ci_l|^se", names(result), value = TRUE, ignore.case = TRUE)
    if(length(ci_cols) >= 2) {
      result$ci_l <- as.numeric(unlist(result[[ci_cols[1]]]))
      result$ci_u <- as.numeric(unlist(result[[ci_cols[2]]]))
    } else {
      # Calculate CI from SE if available
      se_col <- grep("^se", names(result), value = TRUE, ignore.case = TRUE)
      if(length(se_col) > 0) {
        se_val <- as.numeric(unlist(result[[se_col[1]]]))
        result$ci_l <- result$prevalence - 1.96 * se_val
        result$ci_u <- result$prevalence + 1.96 * se_val
      } else {
        result$ci_l <- NA_real_
        result$ci_u <- NA_real_
      }
    }
    
    # Calculate sample sizes
    n_counts <- data_df %>%
      filter(!is.na(.data[[outcome_var]]), !is.na(.data[[group_var]])) %>%
      count(.data[[group_var]], name = "n")
    
    # The grouping variable should be in the result - check row names or first column
    # svyby typically puts grouping variable as row names or first column
    if(group_var %in% names(result)) {
      grouping_col <- group_var
    } else {
      # Check if grouping var is in row names (convert to column)
      row_names <- rownames(result)
      if(!is.null(row_names) && length(row_names) == nrow(result) && 
         any(row_names != as.character(1:nrow(result)))) {
        result[[group_var]] <- row_names
        grouping_col <- group_var
      } else {
        # Use first column that's not the outcome
        non_outcome_cols <- names(result)[!names(result) %in% c(outcome_col, "prevalence", "ci_l", "ci_u")]
        if(length(non_outcome_cols) > 0) {
          grouping_col <- non_outcome_cols[1]
          # Rename it to group_var for consistency
          names(result)[names(result) == grouping_col] <- group_var
          grouping_col <- group_var
        } else {
          grouping_col <- NULL
        }
      }
    }
    
    # Join counts if we have a grouping column
    if(!is.null(grouping_col) && grouping_col %in% names(result)) {
      # Convert grouping column to character for joining
      result[[grouping_col]] <- as.character(result[[grouping_col]])
      n_counts[[group_var]] <- as.character(n_counts[[group_var]])
      
      result <- result %>%
        left_join(n_counts, by = group_var) %>%
        select(all_of(c(group_var, "prevalence", "ci_l", "ci_u", "n")))
    } else {
      # If we can't join, at least return what we have
      result <- result %>%
        mutate(n = NA_integer_) %>%
        select(all_of(c(group_var, "prevalence", "ci_l", "ci_u", "n")))
    }
    
    return(result)
  }, error = function(e) {
    warning(paste("Error calculating prevalence by", group_var, ":", e$message))
    return(data.frame())
  })
}

# Calculate overall prevalence
calc_prevalence_overall <- function(outcome_var, design_obj) {
  tryCatch({
    svy_result <- svymean(
      as.formula(paste0("~", outcome_var)),
      design_obj,
      na.rm = TRUE
    )
    
    mean_val <- as.numeric(svy_result)
    se_val <- as.numeric(attr(svy_result, "var"))^0.5
    ci_l <- mean_val - 1.96 * se_val
    ci_u <- mean_val + 1.96 * se_val
    n_count <- sum(!is.na(design_obj$variables[[outcome_var]]))
    
    data.frame(
      Category = "Overall",
      Level = "Overall",
      prevalence = mean_val,
      ci_l = ci_l,
      ci_u = ci_u,
      n = n_count,
      stringsAsFactors = FALSE
    )
  }, error = function(e) {
    data.frame()
  })
}

# Calculate for smokingnow
outcome_var <- "smokingnow"

# List of variables to show prevalence by - include RegionName and ALL Table 1 variables
# All variables are treated as categorical
prev_vars <- list(
  list(var = "RegionName", label = "Region"),
  list(var = "age_cr1", label = "Age, years"),
  list(var = "ageyrs", label = "Age, years"),
  list(var = "sex_cr2", label = "Sex"),
  list(var = "grade_zar3", label = "Grade"),
  list(var = "age_group", label = "Age group"),
  list(var = "gender", label = "Sex"),
  list(var = "grade_level", label = "Grade level"),
  list(var = "wealth_index_f", label = "Wealth index"),
  list(var = "handrolled_cigarette_f", label = "Use of handrolled cigarette"),
  list(var = "smoked_cigarettes_f", label = "Smoked cigarettes"),
  list(var = "tried_shisha_f", label = "Tried shisha"),
  list(var = "heard_electronic_cigarette_f", label = "Heard of electronic cigarette"),
  list(var = "father_smokes_f", label = "Father smokes"),
  list(var = "people_smoke_school_f", label = "People smoke in school"),
  list(var = "teachers_smoke_school_f", label = "Teachers smoke at school"),
  list(var = "see_tobacco_tv_f", label = "See people using tobacco on TV"),
  list(var = "see_tobacco_advert_f", label = "See tobacco advertisement"),
  list(var = "tobacco_causes_cancer_f", label = "Long tobacco use leads to cancer"),
  list(var = "tobacco_leads_drug_use_f", label = "Long term tobacco use leads to drug use"),
  list(var = "own_item_tobacco_f", label = "Own item with tobacco product"),
  list(var = "tried_smoking_f", label = "Ever tried cigarette smoking"),
  list(var = "parent_smoking_status_f", label = "Parent/guardian smokes in home"),
  list(var = "smoking_5years_from_now_f", label = "Intention to use tobacco in next 5 years"),
  list(var = "antismoking_media_f", label = "Exposure to anti-tobacco media messages"),
  list(var = "taught_dangers_smoking_f", label = "Taught about dangers of tobacco use"),
  list(var = "teacher_smoking_school_f", label = "Saw teachers smoking at school"),
  list(var = "havingfriendsthatsmoke_f", label = "Friends smoke tobacco")
)

# Calculate prevalence (mean) by each variable and category
table2_list <- list()

# By each variable - calculate mean and 95% CI
for(var_info in prev_vars) {
  var_name <- var_info$var
  var_label <- var_info$label
  
  # Check if variable exists and has data
  if(var_name %in% names(df)) {
    # Filter to non-missing data
    df_subset <- df %>%
      filter(!is.na(.data[[var_name]]), !is.na(.data[[outcome_var]]))
    
    if(nrow(df_subset) > 0) {
      # Get unique levels of the variable
      var_levels <- unique(df_subset[[var_name]])
      var_levels <- var_levels[!is.na(var_levels)]
      
      if(length(var_levels) > 0) {
        # Calculate prevalence for each level
        level_results <- list()
        
        for(level in var_levels) {
          # Filter to this level
          df_level <- df_subset %>%
            filter(.data[[var_name]] == level)
          
          if(nrow(df_level) > 0) {
            # Create subset design
            tryCatch({
              design_level <- subset(svy_design, .data[[var_name]] == level & !is.na(.data[[outcome_var]]))
              
              # Calculate mean (prevalence)
              svy_mean <- svymean(as.formula(paste0("~", outcome_var)), design_level, na.rm = TRUE)
              
              # Extract mean and SE
              mean_val <- as.numeric(svy_mean)
              se_val <- sqrt(as.numeric(attr(svy_mean, "var")))
              
              # Calculate 95% CI
              ci_l <- mean_val - 1.96 * se_val
              ci_u <- mean_val + 1.96 * se_val
              
              # Get sample size
              n_count <- nrow(df_level)
              
              level_results[[as.character(level)]] <- data.frame(
                Category = var_label,
                Level = as.character(level),
                prevalence = mean_val,
                ci_l = ci_l,
                ci_u = ci_u,
                n = n_count,
                stringsAsFactors = FALSE
              )
            }, error = function(e) {
              # If survey calculation fails, use simple mean
              mean_val <- mean(df_level[[outcome_var]], na.rm = TRUE)
              n_count <- nrow(df_level)
              se_val <- sd(df_level[[outcome_var]], na.rm = TRUE) / sqrt(n_count)
              ci_l <- mean_val - 1.96 * se_val
              ci_u <- mean_val + 1.96 * se_val
              
              level_results[[as.character(level)]] <<- data.frame(
                Category = var_label,
                Level = as.character(level),
                prevalence = mean_val,
                ci_l = max(0, ci_l),  # Ensure CI doesn't go below 0
                ci_u = min(1, ci_u),  # Ensure CI doesn't go above 1
                n = n_count,
                stringsAsFactors = FALSE
              )
            })
          }
        }
        
        # Combine results for this variable
        if(length(level_results) > 0) {
          var_result <- bind_rows(level_results)
          table2_list[[var_label]] <- var_result
        }
      }
    }
  }
}

# Combine all results
if(length(table2_list) > 0) {
  table2_data <- bind_rows(table2_list) %>%
    mutate(
      Prevalence = sprintf("%.2f%%", prevalence * 100),
      `95% CI` = sprintf("(%.2f%%, %.2f%%)", ci_l * 100, ci_u * 100),
      # Format prevalence with CI for display
      `Prevalence (95% CI)` = paste0(Prevalence, " ", `95% CI`),
      # Humanize Level values - ensure consistent formatting
      Level = case_when(
        # Sex/Gender values
        Level == "Male" ~ "Male",
        Level == "Female" ~ "Female",
        # Binary Yes/No
        Level == "Yes" ~ "Yes",
        Level == "No" ~ "No",
        # Age groups
        Level == "13-14" ~ "13-14 years",
        Level == "15-16" ~ "15-16 years",
        Level == "17+" ~ "17+ years",
        # Grade levels
        Level == "Grade 7" ~ "Grade 7",
        Level == "Grade 8" ~ "Grade 8",
        Level == "Grade 9" ~ "Grade 9",
        # Regions
        Level == "Lusaka" ~ "Lusaka",
        Level == "ROC" ~ "ROC",
        Level == "Tobacco Regions" ~ "Tobacco Regions",
        # Numeric age values - format as "X years"
        grepl("^[0-9]+$", Level) & Category == "Age, years" ~ paste0(Level, " years"),
        # Keep other values as is (they should already be humanized from factors)
        TRUE ~ Level
      )
    ) %>%
    select(Category, Level, n, `Prevalence (95% CI)`) %>%
    # Reorder by variable (no Overall)
    arrange(Category, Level)

  # Create publication-quality formatted table
  table2_flextable <- table2_data %>%
    flextable() %>%
    set_header_labels(
      Category = "**Variable**",
      Level = "**Category**",
      n = "**N**",
      `Prevalence (95% CI)` = "**Prevalence (95% CI)**"
    ) %>%
    bold(part = "header") %>%
    bold(j = 1, part = "body") %>%  # Bold variable names
    align(align = "left", part = "all") %>%
    align(j = c("n", "Prevalence (95% CI)"), align = "center", part = "all") %>%
    width(j = "Category", width = 2.5) %>%
    width(j = "Level", width = 2.0) %>%
    width(j = "n", width = 0.8) %>%
    width(j = "Prevalence (95% CI)", width = 2.5) %>%
    theme_booktabs() %>%
    fontsize(size = 10, part = "all") %>%
    padding(padding = 4, part = "all")
  
  
  # Add footnote
  table2_flextable <- table2_flextable %>%
    add_footer_lines("Prevalence of current smoking (smokingnow = 1) by key variables. Values are weighted percentages with 95% confidence intervals calculated using survey weights.") %>%
    fontsize(size = 9, part = "footer") %>%
    italic(part = "footer") %>%
    align(align = "left", part = "footer")

  table2_flextable
} else {
  cat("No data available for Table 2.\n")
}
```

### Prevalence of Tobacco Use

The overall weighted prevalence of current tobacco smoking among Zambian adolescents, as shown in Table 2, represents a substantial public health concern. This prevalence estimate, derived from survey-weighted analysis accounting for the complex survey design, provides a nationally representative assessment of current tobacco use among school-going adolescents aged 11-17 years in Zambia. The magnitude of this prevalence underscores the urgent need for comprehensive tobacco control interventions targeting this vulnerable population.

**Demographic Patterns**

Prevalence of current smoking varied substantially across demographic subgroups, revealing important patterns in tobacco use risk (Table 2). Regional differences were particularly notable, with prevalence estimates and 95% confidence intervals for each region (Lusaka, ROC, and Tobacco Regions) presented in Table 2. The regional variation in prevalence suggests that regional factors—potentially including economic dependence on tobacco production, enforcement of tobacco control policies, or cultural norms—may significantly influence adolescent tobacco use patterns.

Age-related patterns demonstrated variation across age groups, with prevalence estimates and confidence intervals for each age category shown in Table 2. This age gradient is consistent with developmental patterns observed globally, where tobacco use initiation and continuation may increase with age during adolescence. Sex differences were also evident, with prevalence estimates for males and females presented in Table 2, reflecting gender-specific risk patterns that may be influenced by social norms, peer influences, or targeted marketing. Grade level analysis revealed patterns across Grades 7, 8, and 9, with prevalence estimates for each grade level shown in Table 2, potentially reflecting both age effects and increased exposure to tobacco use opportunities as adolescents progress through school.

**Predictor-Associated Prevalence**

Prevalence patterns across predictor variables revealed strong associations with several modifiable risk and protective factors (Table 2). Adolescents who had ever tried cigarette smoking showed substantially different current smoking prevalence compared to those who had never tried, with exact estimates and confidence intervals presented in Table 2. This pattern highlights the critical importance of preventing tobacco use initiation as a primary prevention strategy.

Parental smoking status was strongly associated with adolescent smoking, with prevalence estimates for those with and without smoking parents shown in Table 2, demonstrating the powerful influence of household tobacco exposure. Intention to use tobacco in the next five years showed associations with current smoking, with prevalence estimates for those with and without future use intentions presented in Table 2, suggesting that future use intentions may reflect current behavior patterns or indicate high-risk adolescents requiring targeted intervention.

Exposure to anti-tobacco media messages showed associations with current smoking prevalence, with estimates for exposed and non-exposed adolescents presented in Table 2. Educational factors demonstrated important associations: adolescents who reported being taught about the dangers of tobacco use showed different prevalence compared to those not taught, with exact estimates shown in Table 2, suggesting that school-based tobacco education may be effective in reducing use. Conversely, observing teachers smoking at school was associated with different prevalence patterns, as shown in Table 2, highlighting the importance of tobacco-free school policies and role modeling. Peer influences were particularly evident, with adolescents reporting friends who smoke showing substantially different prevalence compared to those without smoking friends, as detailed in Table 2, emphasizing the critical role of peer networks in adolescent tobacco use behavior.

## Figure 1: Prevalence of Current Tobacco Use by Region and Demographics

```{r figure1-prevalence, echo=FALSE, fig.width=12, fig.height=8, fig.cap="Prevalence of current tobacco use (smokingnow) by region, sex, age group, and grade level"}
# Create comprehensive prevalence visualization
library(ggplot2)
library(dplyr)

# Prepare data for plotting
df_plot <- df %>%
  filter(!is.na(smokingnow)) %>%
  mutate(
    RegionName = factor(RegionName, levels = c("Lusaka", "ROC", "Tobacco Regions")),
    gender = factor(gender, levels = c("Female", "Male")),
    age_group = factor(age_group, levels = c("13-14", "15-16", "17+")),
    grade_level = factor(grade_level, levels = c("Grade 7", "Grade 8", "Grade 9"))
  )

# Calculate prevalence by region
prev_region <- tryCatch({
  design_subset <- subset(svy_design, !is.na(smokingnow) & !is.na(RegionName))
  svy_by_region <- svyby(~smokingnow, ~RegionName, design_subset, svymean, na.rm = TRUE, vartype = "ci")
  data.frame(
    Category = "Region",
    Level = rownames(svy_by_region),
    Prevalence = as.numeric(svy_by_region$smokingnow) * 100,
    CI_Lower = as.numeric(svy_by_region$ci_l) * 100,
    CI_Upper = as.numeric(svy_by_region$ci_u) * 100
  )
}, error = function(e) {
  df_plot %>%
    group_by(RegionName) %>%
    summarise(
      Prevalence = mean(smokingnow, na.rm = TRUE) * 100,
      n = n(),
      se = sd(smokingnow, na.rm = TRUE) / sqrt(n) * 100,
      .groups = "drop"
    ) %>%
    mutate(
      CI_Lower = pmax(0, Prevalence - 1.96 * se),
      CI_Upper = pmin(100, Prevalence + 1.96 * se),
      Category = "Region",
      Level = as.character(RegionName)
    ) %>%
    select(Category, Level, Prevalence, CI_Lower, CI_Upper)
})

# Calculate prevalence by sex
prev_sex <- tryCatch({
  design_subset <- subset(svy_design, !is.na(smokingnow) & !is.na(gender))
  svy_by_sex <- svyby(~smokingnow, ~gender, design_subset, svymean, na.rm = TRUE, vartype = "ci")
  data.frame(
    Category = "Sex",
    Level = rownames(svy_by_sex),
    Prevalence = as.numeric(svy_by_sex$smokingnow) * 100,
    CI_Lower = as.numeric(svy_by_sex$ci_l) * 100,
    CI_Upper = as.numeric(svy_by_sex$ci_u) * 100
  )
}, error = function(e) {
  df_plot %>%
    filter(!is.na(gender)) %>%
    group_by(gender) %>%
    summarise(
      Prevalence = mean(smokingnow, na.rm = TRUE) * 100,
      n = n(),
      se = sd(smokingnow, na.rm = TRUE) / sqrt(n) * 100,
      .groups = "drop"
    ) %>%
    mutate(
      CI_Lower = pmax(0, Prevalence - 1.96 * se),
      CI_Upper = pmin(100, Prevalence + 1.96 * se),
      Category = "Sex",
      Level = as.character(gender)
    ) %>%
    select(Category, Level, Prevalence, CI_Lower, CI_Upper)
})

# Calculate prevalence by age group
prev_age <- tryCatch({
  design_subset <- subset(svy_design, !is.na(smokingnow) & !is.na(age_group))
  svy_by_age <- svyby(~smokingnow, ~age_group, design_subset, svymean, na.rm = TRUE, vartype = "ci")
  data.frame(
    Category = "Age Group",
    Level = rownames(svy_by_age),
    Prevalence = as.numeric(svy_by_age$smokingnow) * 100,
    CI_Lower = as.numeric(svy_by_age$ci_l) * 100,
    CI_Upper = as.numeric(svy_by_age$ci_u) * 100
  )
}, error = function(e) {
  df_plot %>%
    filter(!is.na(age_group)) %>%
    group_by(age_group) %>%
    summarise(
      Prevalence = mean(smokingnow, na.rm = TRUE) * 100,
      n = n(),
      se = sd(smokingnow, na.rm = TRUE) / sqrt(n) * 100,
      .groups = "drop"
    ) %>%
    mutate(
      CI_Lower = pmax(0, Prevalence - 1.96 * se),
      CI_Upper = pmin(100, Prevalence + 1.96 * se),
      Category = "Age Group",
      Level = as.character(age_group)
    ) %>%
    select(Category, Level, Prevalence, CI_Lower, CI_Upper)
})

# Calculate prevalence by grade
prev_grade <- tryCatch({
  design_subset <- subset(svy_design, !is.na(smokingnow) & !is.na(grade_level))
  svy_by_grade <- svyby(~smokingnow, ~grade_level, design_subset, svymean, na.rm = TRUE, vartype = "ci")
  data.frame(
    Category = "Grade",
    Level = rownames(svy_by_grade),
    Prevalence = as.numeric(svy_by_grade$smokingnow) * 100,
    CI_Lower = as.numeric(svy_by_grade$ci_l) * 100,
    CI_Upper = as.numeric(svy_by_grade$ci_u) * 100
  )
}, error = function(e) {
  df_plot %>%
    filter(!is.na(grade_level)) %>%
    group_by(grade_level) %>%
    summarise(
      Prevalence = mean(smokingnow, na.rm = TRUE) * 100,
      n = n(),
      se = sd(smokingnow, na.rm = TRUE) / sqrt(n) * 100,
      .groups = "drop"
    ) %>%
    mutate(
      CI_Lower = pmax(0, Prevalence - 1.96 * se),
      CI_Upper = pmin(100, Prevalence + 1.96 * se),
      Category = "Grade",
      Level = as.character(grade_level)
    ) %>%
    select(Category, Level, Prevalence, CI_Lower, CI_Upper)
})

# Combine all data
plot_data <- bind_rows(prev_region, prev_sex, prev_age, prev_grade) %>%
  mutate(
    Category = factor(Category, levels = c("Region", "Sex", "Age Group", "Grade")),
    Level = factor(Level)
  )

# Create faceted bar chart with error bars
if(nrow(plot_data) > 0) {
  p <- ggplot(plot_data, aes(x = Level, y = Prevalence, fill = Category)) +
    geom_bar(stat = "identity", position = "dodge", alpha = 0.8) +
    geom_errorbar(aes(ymin = CI_Lower, ymax = CI_Upper), 
                  width = 0.2, position = position_dodge(0.9)) +
    facet_wrap(~Category, scales = "free_x", ncol = 2) +
    scale_fill_brewer(palette = "Set2") +
    labs(
      title = "Prevalence of Current Tobacco Use by Region and Demographics",
      subtitle = "Weighted prevalence with 95% confidence intervals",
      x = "",
      y = "Prevalence (%)",
      fill = "Category"
    ) +
    theme_minimal() +
    theme(
      plot.title = element_text(size = 14, face = "bold", hjust = 0.5),
      plot.subtitle = element_text(size = 12, hjust = 0.5),
      axis.text.x = element_text(angle = 45, hjust = 1),
      strip.text = element_text(face = "bold", size = 11),
      legend.position = "none"
    ) +
    scale_y_continuous(limits = c(0, NA), expand = expansion(mult = c(0, 0.1)))
  
  print(p)
} else {
  cat("No data available for Figure 1.\n")
}
```

### Prevalence of the outcome: Mean of smokingnow (Current Smoking Status)

```{r mean-smokingnow, echo=FALSE, results='asis'}
# Calculate mean (prevalence) of smokingnow
# Unweighted mean
unweighted_mean <- mean(df$smokingnow, na.rm = TRUE)
unweighted_n <- sum(!is.na(df$smokingnow))

# Weighted mean using survey design
weighted_mean_result <- svymean(~smokingnow, svy_design, na.rm = TRUE)
weighted_mean <- as.numeric(weighted_mean_result)
weighted_se <- as.numeric(attr(weighted_mean_result, "var"))^0.5
weighted_ci_l <- weighted_mean - 1.96 * weighted_se
weighted_ci_u <- weighted_mean + 1.96 * weighted_se

cat(sprintf("**Unweighted mean**: %.4f (%.2f%%) [n = %d]\n\n", 
            unweighted_mean, unweighted_mean * 100, unweighted_n))
cat(sprintf("**Weighted mean**: %.4f (%.2f%%) [95%% CI: %.2f%%, %.2f%%]\n", 
            weighted_mean, weighted_mean * 100, weighted_ci_l * 100, weighted_ci_u * 100))
```

## Multilevel Regression Analysis

### Table 3: Adjusted Odds Ratios for Current Smoking (smokingnow)

```{r table3-regression, echo=FALSE, results='asis'}
library(MASS)  # For stepwise selection

# Helper function to check if factor has at least 2 levels
check_factor_levels <- function(var, data) {
  if(is.factor(var)) {
    n_levels <- length(levels(droplevels(var)))
    return(n_levels >= 2)
  } else {
    n_levels <- length(unique(var[!is.na(var)]))
    return(n_levels >= 2)
  }
}

# Safe model fitting function that checks levels before fitting
safe_glm <- function(formula, data, family = binomial) {
  # Extract variable names from formula
  formula_vars <- all.vars(formula)
  outcome_var <- formula_vars[1]
  predictor_vars <- formula_vars[-1]
  
  # Create a copy of data and drop unused factor levels
  data_clean <- data
  for(var_name in predictor_vars) {
    if(var_name %in% names(data_clean) && is.factor(data_clean[[var_name]])) {
      data_clean[[var_name]] <- droplevels(data_clean[[var_name]])
    }
  }
  
  # Check each predictor variable has at least 2 levels in the actual data
  valid_vars <- c()
  for(var_name in predictor_vars) {
    if(var_name %in% names(data_clean)) {
      var_data <- data_clean[[var_name]]
      if(check_factor_levels(var_data, data_clean)) {
        valid_vars <- c(valid_vars, var_name)
      }
    }
  }
  
  # Need at least one valid predictor
  if(length(valid_vars) == 0) {
    return(NULL)
  }
  
  # Rebuild formula with only valid variables
  if(length(valid_vars) == length(predictor_vars)) {
    # All variables valid, use original formula
    new_formula <- formula
  } else {
    # Some variables invalid, rebuild formula
    new_formula_str <- paste(outcome_var, "~", paste(valid_vars, collapse = " + "))
    new_formula <- as.formula(new_formula_str)
  }
  
  # Fit model with cleaned data
  tryCatch({
    glm(new_formula, data = data_clean, family = family)
  }, error = function(e) {
    warning(paste("Model fitting failed:", e$message))
    NULL
  })
}

# Prepare data for models - include RegionName, age_cr1, sex_cr2, grade_zar3 as a priori
df_model <- df %>% 
  filter(!is.na(smokingnow)) %>%  # Only filter for outcome, don't filter for a priori vars yet
  mutate(
    RegionName = factor(RegionName, levels = c("Lusaka", "ROC", "Tobacco Regions")),
    # A priori variables: RegionName, age_cr1, sex_cr2, grade_zar3
    age_cr1 = factor(age_cr1, levels = c(11, 12, 13, 14, 15, 16, 17)),  # Factor age variable from CR1
    sex_cr2 = factor(sex_cr2, levels = c("Female", "Male")),
    grade_zar3 = factor(grade_zar3, levels = c("Grade 7", "Grade 8", "Grade 9")),
    # Add other variables from Table 2 that might be missing
    age_group = factor(age_group, levels = c("13-14", "15-16", "17+")),
    gender = factor(gender, levels = c("Female", "Male")),
    grade_level = factor(grade_level, levels = c("Grade 9", "Grade 8", "Grade 7")),
    ageyrs = ageyrs,
    # tried_smoking_f: 1 = Yes (tried smoking), 0 = No (never tried)
    tried_smoking_f = factor(tried_smoking_f, levels = c("No", "Yes")),
    # parent_smoking_status_f: 0 = No, 1 = Yes (based on OR15 mapping)
    parent_smoking_status_f = factor(parent_smoking_status_f, 
                                    levels = c("No", "Yes")),
    # smoking_5years_from_now_f: Binary (Yes: Definitely yes/Probably yes, No: Definitely not/Probably not)
    smoking_5years_from_now_f = factor(smoking_5years_from_now_f,
                                      levels = c("No", "Yes")),
    antismoking_media_f = factor(antismoking_media_f, levels = c("No", "Yes")),
    # taught_dangers_smoking_f: "Don't know" is recoded to "No"
    taught_dangers_smoking_f = factor(taught_dangers_smoking_f,
                                     levels = c("No", "Yes")),
    # teacher_smoking_school_f: 0 = No, 1 = Yes (based on OR60 mapping)
    teacher_smoking_school_f = factor(teacher_smoking_school_f,
                                     levels = c("No", "Yes")),
    # havingfriendsthatsmoke_f: Factor version
    havingfriendsthatsmoke_f = factor(havingfriendsthatsmoke_f,
                                     levels = c("No", "Yes")),
    # New variables from paper 8
    wealth_index_f = if("wealth_index_f" %in% names(.)) factor(wealth_index_f, levels = c("Low", "Middle", "Rich"), labels = c("Low", "Middle", "Rich")) else wealth_index_f,
    handrolled_cigarette_f = if("handrolled_cigarette_f" %in% names(.)) factor(handrolled_cigarette_f, levels = c("No", "Yes"), labels = c("No", "Yes")) else handrolled_cigarette_f,
    smoked_cigarettes_f = if("smoked_cigarettes_f" %in% names(.)) factor(smoked_cigarettes_f, levels = c("No", "Yes"), labels = c("No", "Yes")) else smoked_cigarettes_f,
    tried_shisha_f = if("tried_shisha_f" %in% names(.)) factor(tried_shisha_f, levels = c("No", "Yes"), labels = c("No", "Yes")) else tried_shisha_f,
    heard_electronic_cigarette_f = if("heard_electronic_cigarette_f" %in% names(.)) factor(heard_electronic_cigarette_f, levels = c("No", "Yes"), labels = c("No", "Yes")) else heard_electronic_cigarette_f,
    father_smokes_f = if("father_smokes_f" %in% names(.)) factor(father_smokes_f, levels = c("No", "Yes"), labels = c("No", "Yes")) else father_smokes_f,
    people_smoke_school_f = if("people_smoke_school_f" %in% names(.)) factor(people_smoke_school_f, levels = c("No", "Yes"), labels = c("No", "Yes")) else people_smoke_school_f,
    teachers_smoke_school_f = if("teachers_smoke_school_f" %in% names(.)) factor(teachers_smoke_school_f, levels = c("No", "Yes"), labels = c("No", "Yes")) else teachers_smoke_school_f,
    see_tobacco_tv_f = if("see_tobacco_tv_f" %in% names(.)) factor(see_tobacco_tv_f, levels = c("No", "Yes"), labels = c("No", "Yes")) else see_tobacco_tv_f,
    see_tobacco_advert_f = if("see_tobacco_advert_f" %in% names(.)) factor(see_tobacco_advert_f, levels = c("No", "Yes"), labels = c("No", "Yes")) else see_tobacco_advert_f,
    tobacco_causes_cancer_f = if("tobacco_causes_cancer_f" %in% names(.)) factor(tobacco_causes_cancer_f, levels = c("No", "Yes"), labels = c("No", "Yes")) else tobacco_causes_cancer_f,
    tobacco_leads_drug_use_f = if("tobacco_leads_drug_use_f" %in% names(.)) factor(tobacco_leads_drug_use_f, levels = c("No", "Yes"), labels = c("No", "Yes")) else tobacco_leads_drug_use_f,
    own_item_tobacco_f = if("own_item_tobacco_f" %in% names(.)) factor(own_item_tobacco_f, levels = c("No", "Yes"), labels = c("No", "Yes")) else own_item_tobacco_f
  )

# Drop unused factor levels and check which variables have sufficient levels
df_model$RegionName <- droplevels(df_model$RegionName)
df_model$age_cr1 <- droplevels(df_model$age_cr1)
df_model$sex_cr2 <- droplevels(df_model$sex_cr2)
df_model$grade_zar3 <- droplevels(df_model$grade_zar3)
df_model$age_group <- droplevels(df_model$age_group)
df_model$gender <- droplevels(df_model$gender)
df_model$grade_level <- droplevels(df_model$grade_level)
df_model$tried_smoking_f <- droplevels(df_model$tried_smoking_f)
df_model$parent_smoking_status_f <- droplevels(df_model$parent_smoking_status_f)
df_model$smoking_5years_from_now_f <- droplevels(df_model$smoking_5years_from_now_f)
df_model$antismoking_media_f <- droplevels(df_model$antismoking_media_f)
df_model$taught_dangers_smoking_f <- droplevels(df_model$taught_dangers_smoking_f)
df_model$wealth_index_f <- droplevels(df_model$wealth_index_f)
df_model$handrolled_cigarette_f <- droplevels(df_model$handrolled_cigarette_f)
df_model$smoked_cigarettes_f <- droplevels(df_model$smoked_cigarettes_f)
df_model$tried_shisha_f <- droplevels(df_model$tried_shisha_f)
df_model$heard_electronic_cigarette_f <- droplevels(df_model$heard_electronic_cigarette_f)
df_model$father_smokes_f <- droplevels(df_model$father_smokes_f)
df_model$people_smoke_school_f <- droplevels(df_model$people_smoke_school_f)
df_model$teachers_smoke_school_f <- droplevels(df_model$teachers_smoke_school_f)
df_model$see_tobacco_tv_f <- droplevels(df_model$see_tobacco_tv_f)
df_model$see_tobacco_advert_f <- droplevels(df_model$see_tobacco_advert_f)
df_model$tobacco_causes_cancer_f <- droplevels(df_model$tobacco_causes_cancer_f)
df_model$tobacco_leads_drug_use_f <- droplevels(df_model$tobacco_leads_drug_use_f)
df_model$own_item_tobacco_f <- droplevels(df_model$own_item_tobacco_f)

# Check which variables have at least 2 levels
# A priori variables: RegionName, age_cr1, sex_cr2, grade_zar3
has_region <- check_factor_levels(df_model$RegionName, df_model)
has_age_cr1 <- check_factor_levels(df_model$age_cr1, df_model)
has_sex_cr2 <- check_factor_levels(df_model$sex_cr2, df_model)
has_grade_zar3 <- check_factor_levels(df_model$grade_zar3, df_model)
has_tried_smoking <- check_factor_levels(df_model$tried_smoking_f, df_model)
has_parent_smoking <- check_factor_levels(df_model$parent_smoking_status_f, df_model)
has_smoking_5years <- check_factor_levels(df_model$smoking_5years_from_now_f, df_model)
has_antismoking_media <- check_factor_levels(df_model$antismoking_media_f, df_model)
has_taught_dangers <- check_factor_levels(df_model$taught_dangers_smoking_f, df_model)
has_teacher_smoking <- check_factor_levels(df_model$teacher_smoking_school_f, df_model)
has_havingfriendsthatsmoke <- check_factor_levels(df_model$havingfriendsthatsmoke_f, df_model)
# Check new variables from paper 8
has_wealth_index <- check_factor_levels(df_model$wealth_index_f, df_model)
has_handrolled_cigarette <- check_factor_levels(df_model$handrolled_cigarette_f, df_model)
has_smoked_cigarettes <- check_factor_levels(df_model$smoked_cigarettes_f, df_model)
has_tried_shisha <- check_factor_levels(df_model$tried_shisha_f, df_model)
has_heard_electronic_cigarette <- check_factor_levels(df_model$heard_electronic_cigarette_f, df_model)
has_father_smokes <- check_factor_levels(df_model$father_smokes_f, df_model)
has_people_smoke_school <- check_factor_levels(df_model$people_smoke_school_f, df_model)
has_see_tobacco_tv <- check_factor_levels(df_model$see_tobacco_tv_f, df_model)
has_see_tobacco_advert <- check_factor_levels(df_model$see_tobacco_advert_f, df_model)
has_tobacco_causes_cancer <- check_factor_levels(df_model$tobacco_causes_cancer_f, df_model)
has_tobacco_leads_drug_use <- check_factor_levels(df_model$tobacco_leads_drug_use_f, df_model)
has_own_item_tobacco <- check_factor_levels(df_model$own_item_tobacco_f, df_model)
# Check additional variables from Table 2 (for reference, not used as a priori)
has_age_group <- check_factor_levels(df_model$age_group, df_model)
has_gender <- check_factor_levels(df_model$gender, df_model)
has_grade_level <- check_factor_levels(df_model$grade_level, df_model)
has_ageyrs <- length(unique(df_model$ageyrs[!is.na(df_model$ageyrs)])) >= 2

# Function to humanize variable names
humanize_variable <- function(term) {
  term_clean <- case_when(
    grepl("^RegionNameROC", term) ~ "ROC (vs Lusaka)",
    grepl("^RegionNameTobacco Regions", term) ~ "Tobacco Regions (vs Lusaka)",
    grepl("^age_cr1(12|13|14|15|16|17)$", term) ~ paste0("Age: ", gsub("^age_cr1", "", term), " years (vs 11 years)"),
    grepl("^sex_cr2Male", term) ~ "Male (vs Female, CR2)",
    TRUE ~ term
  ) %>%
    str_replace("^grade_zar3Grade 8", "Grade 8 (vs Grade 7, ZAR3)") %>%
    str_replace("^grade_zar3Grade 9", "Grade 9 (vs Grade 7, ZAR3)") %>%
    str_replace("^sex1", "Male (vs Female)") %>%
    str_replace("^alt_age", "Age (years)") %>%
    str_replace("^grade", "Grade ") %>%
    str_replace("^genderMale", "Male (vs Female)") %>%
    str_replace("^age_group15-16", "Age 15-16 years (vs 13-14 years)") %>%
    str_replace("^age_group17\\+", "Age 17+ years (vs 13-14 years)") %>%
    str_replace("^grade_levelGrade 8", "Grade 8 (vs Grade 9)") %>%
    str_replace("^grade_levelGrade 7", "Grade 7 (vs Grade 9)") %>%
    str_replace("^tried_smoking_fYes", "Ever tried cigarette smoking (vs No)") %>%
    str_replace("^parent_smoking_status_fYes", "Parent/guardian smokes in home (vs No)") %>%
    str_replace("^smoking_5years_from_now_fYes", "Intention to use tobacco in next 5 years: Yes (vs No)") %>%
    str_replace("^antismoking_media_fYes", "Exposure to anti-tobacco media messages (vs No)") %>%
    str_replace("^taught_dangers_smoking_fYes", "Taught about dangers of tobacco use (vs No)") %>%
    str_replace("^teacher_smoking_school_fYes", "Saw teachers smoking at school (vs No)") %>%
    str_replace("^havingfriendsthatsmoke_fYes", "Friends smoke tobacco (vs No)") %>%
    str_replace("^wealth_index_fMiddle", "Wealth index: Middle (vs Low)") %>%
    str_replace("^wealth_index_fRich", "Wealth index: Rich (vs Low)") %>%
    str_replace("^handrolled_cigarette_fYes", "Use of handrolled cigarette (vs No)") %>%
    str_replace("^smoked_cigarettes_fYes", "Smoked cigarettes (vs No)") %>%
    str_replace("^tried_shisha_fYes", "Tried shisha (vs No)") %>%
    str_replace("^heard_electronic_cigarette_fYes", "Heard of electronic cigarette (vs No)") %>%
    str_replace("^father_smokes_fYes", "Father smokes (vs No)") %>%
    str_replace("^people_smoke_school_fYes", "People smoke in school (vs No)") %>%
    str_replace("^see_tobacco_tv_fYes", "See people using tobacco on TV (vs No)") %>%
    str_replace("^see_tobacco_advert_fYes", "See tobacco advertisement (vs No)") %>%
    str_replace("^tobacco_causes_cancer_fYes", "Long tobacco use leads to cancer (vs No)") %>%
    str_replace("^tobacco_leads_drug_use_fYes", "Long term tobacco use leads to drug use (vs No)") %>%
    str_replace("^own_item_tobacco_fYes", "Own item with tobacco product (vs No)") %>%
    identity()
  return(term_clean)
}

# ADJUSTED MODEL with stepwise selection
# A priori variables: RegionName, age_cr1, sex_cr2, grade_zar3

base_vars <- c()
if(has_region) base_vars <- c(base_vars, "RegionName")
if(has_age_cr1) base_vars <- c(base_vars, "age_cr1")
if(has_sex_cr2) base_vars <- c(base_vars, "sex_cr2")
if(has_grade_zar3) base_vars <- c(base_vars, "grade_zar3")

if(length(base_vars) < 1) {
  adjusted_model <- NULL
  
  # Try to fit with whatever variables we have
  if(has_region || has_age_cr1 || has_sex_cr2 || has_grade_zar3) {
    # At least one variable exists, try to fit
    fallback_vars <- c()
    if(has_region) fallback_vars <- c(fallback_vars, "RegionName")
    if(has_age_cr1) fallback_vars <- c(fallback_vars, "age_cr1")
    if(has_sex_cr2) fallback_vars <- c(fallback_vars, "sex_cr2")
    if(has_grade_zar3) fallback_vars <- c(fallback_vars, "grade_zar3")
    
    if(length(fallback_vars) > 0) {
      fallback_formula <- as.formula(paste("smokingnow ~", paste(fallback_vars, collapse = " + ")))
      model_data_fallback <- if(has_grade_zar3) {
        df_model %>% filter(!is.na(grade_zar3))
      } else {
        df_model
      }
      adjusted_model <- safe_glm(fallback_formula, data = model_data_fallback, family = binomial)
    }
  }
} else {
  # Start with a priori variables: age, sex, grade, and region
  base_formula_str <- paste("smokingnow ~", paste(base_vars, collapse = " + "))
  base_formula <- as.formula(base_formula_str)
  
  # Full model with ALL variables from Table 1 and Table 2
  # Start with a priori variables (age, sex, grade, region)
  full_vars <- base_vars
  
  # Note: RegionName is already in base_vars as an a priori variable
  
  # Add ALL other variables from Table 1 and Table 2
  # These are all the variables shown in Table 1 and Table 2
  # (RegionName is already included as an a priori variable)
  if(has_tried_smoking) full_vars <- c(full_vars, "tried_smoking_f")
  if(has_parent_smoking) full_vars <- c(full_vars, "parent_smoking_status_f")
  if(has_smoking_5years) full_vars <- c(full_vars, "smoking_5years_from_now_f")
  if(has_antismoking_media) full_vars <- c(full_vars, "antismoking_media_f")
  if(has_taught_dangers) full_vars <- c(full_vars, "taught_dangers_smoking_f")
  if(has_teacher_smoking) full_vars <- c(full_vars, "teacher_smoking_school_f")
  if(has_havingfriendsthatsmoke) full_vars <- c(full_vars, "havingfriendsthatsmoke_f")
  # Add new variables from paper 8
  if(has_wealth_index) full_vars <- c(full_vars, "wealth_index_f")
  if(has_handrolled_cigarette) full_vars <- c(full_vars, "handrolled_cigarette_f")
  if(has_smoked_cigarettes) full_vars <- c(full_vars, "smoked_cigarettes_f")
  if(has_tried_shisha) full_vars <- c(full_vars, "tried_shisha_f")
  if(has_heard_electronic_cigarette) full_vars <- c(full_vars, "heard_electronic_cigarette_f")
  if(has_father_smokes) full_vars <- c(full_vars, "father_smokes_f")
  if(has_people_smoke_school) full_vars <- c(full_vars, "people_smoke_school_f")
  if(has_see_tobacco_tv) full_vars <- c(full_vars, "see_tobacco_tv_f")
  if(has_see_tobacco_advert) full_vars <- c(full_vars, "see_tobacco_advert_f")
  if(has_tobacco_causes_cancer) full_vars <- c(full_vars, "tobacco_causes_cancer_f")
  if(has_tobacco_leads_drug_use) full_vars <- c(full_vars, "tobacco_leads_drug_use_f")
  if(has_own_item_tobacco) full_vars <- c(full_vars, "own_item_tobacco_f")
  
  # Note: We use age_cr1, sex_cr2, and grade_zar3 as a priori variables.
  # Alternative versions (age_group, gender, grade_level, ageyrs) are not included
  # in the model to avoid multicollinearity.
  
  # Prepare model data - filter for missing a priori variables, but be lenient
  # Only filter out rows where ALL a priori variables are missing
  model_data <- df_model
  # Filter for rows that have at least the outcome and one a priori variable
  model_data <- model_data %>% 
    filter(!is.na(smokingnow))
  
  # Re-check all base_vars after basic filtering
  valid_base_vars <- c()
  for(var in base_vars) {
    if(var %in% names(model_data)) {
      if(var == "RegionName" && check_factor_levels(model_data$RegionName, model_data)) {
        valid_base_vars <- c(valid_base_vars, var)
      } else if(var == "age_cr1" && check_factor_levels(model_data$age_cr1, model_data)) {
        valid_base_vars <- c(valid_base_vars, var)
      } else if(var == "sex_cr2" && check_factor_levels(model_data$sex_cr2, model_data)) {
        valid_base_vars <- c(valid_base_vars, var)
      } else if(var == "grade_zar3" && check_factor_levels(model_data$grade_zar3, model_data)) {
        valid_base_vars <- c(valid_base_vars, var)
      }
    } 
  }
  
  if(length(valid_base_vars) == 0) {
    adjusted_model <- NULL
  } else {
    # Update base_vars and base_formula with valid variables
    base_vars <- valid_base_vars
    base_formula_str <- paste("smokingnow ~", paste(base_vars, collapse = " + "))
    base_formula <- as.formula(base_formula_str)
    
    # Re-check all variables in full_vars after filtering
    valid_full_vars <- base_vars  # Start with valid a priori variables
    for(var in full_vars) {
      if(var %in% base_vars) {
        # Already included
        next
      }
      if(var %in% names(model_data)) {
        # Check if variable has sufficient levels
        if(var == "RegionName" && check_factor_levels(model_data$RegionName, model_data)) {
          valid_full_vars <- c(valid_full_vars, var)
        } else if(var == "age_cr1" && check_factor_levels(model_data$age_cr1, model_data)) {
          valid_full_vars <- c(valid_full_vars, var)
        } else if(var == "sex_cr2" && check_factor_levels(model_data$sex_cr2, model_data)) {
          valid_full_vars <- c(valid_full_vars, var)
        } else if(var == "grade_zar3" && check_factor_levels(model_data$grade_zar3, model_data)) {
          valid_full_vars <- c(valid_full_vars, var)
        } else if(var == "tried_smoking_f" && check_factor_levels(model_data$tried_smoking_f, model_data)) {
          valid_full_vars <- c(valid_full_vars, var)
        } else if(var == "parent_smoking_status_f" && check_factor_levels(model_data$parent_smoking_status_f, model_data)) {
          valid_full_vars <- c(valid_full_vars, var)
        } else if(var == "smoking_5years_from_now_f" && check_factor_levels(model_data$smoking_5years_from_now_f, model_data)) {
          valid_full_vars <- c(valid_full_vars, var)
        } else if(var == "antismoking_media_f" && check_factor_levels(model_data$antismoking_media_f, model_data)) {
          valid_full_vars <- c(valid_full_vars, var)
        } else if(var == "taught_dangers_smoking_f" && check_factor_levels(model_data$taught_dangers_smoking_f, model_data)) {
          valid_full_vars <- c(valid_full_vars, var)
        } else if(var == "teacher_smoking_school_f" && check_factor_levels(model_data$teacher_smoking_school_f, model_data)) {
          valid_full_vars <- c(valid_full_vars, var)
        } else if(var == "havingfriendsthatsmoke_f" && check_factor_levels(model_data$havingfriendsthatsmoke_f, model_data)) {
          valid_full_vars <- c(valid_full_vars, var)
        } else if(var == "wealth_index_f" && check_factor_levels(model_data$wealth_index_f, model_data)) {
          valid_full_vars <- c(valid_full_vars, var)
        } else if(var == "handrolled_cigarette_f" && check_factor_levels(model_data$handrolled_cigarette_f, model_data)) {
          valid_full_vars <- c(valid_full_vars, var)
        } else if(var == "smoked_cigarettes_f" && check_factor_levels(model_data$smoked_cigarettes_f, model_data)) {
          valid_full_vars <- c(valid_full_vars, var)
        } else if(var == "tried_shisha_f" && check_factor_levels(model_data$tried_shisha_f, model_data)) {
          valid_full_vars <- c(valid_full_vars, var)
        } else if(var == "heard_electronic_cigarette_f" && check_factor_levels(model_data$heard_electronic_cigarette_f, model_data)) {
          valid_full_vars <- c(valid_full_vars, var)
        } else if(var == "father_smokes_f" && check_factor_levels(model_data$father_smokes_f, model_data)) {
          valid_full_vars <- c(valid_full_vars, var)
        } else if(var == "people_smoke_school_f" && check_factor_levels(model_data$people_smoke_school_f, model_data)) {
          valid_full_vars <- c(valid_full_vars, var)
        } else if(var == "see_tobacco_tv_f" && check_factor_levels(model_data$see_tobacco_tv_f, model_data)) {
          valid_full_vars <- c(valid_full_vars, var)
        } else if(var == "see_tobacco_advert_f" && check_factor_levels(model_data$see_tobacco_advert_f, model_data)) {
          valid_full_vars <- c(valid_full_vars, var)
        } else if(var == "tobacco_causes_cancer_f" && check_factor_levels(model_data$tobacco_causes_cancer_f, model_data)) {
          valid_full_vars <- c(valid_full_vars, var)
        } else if(var == "tobacco_leads_drug_use_f" && check_factor_levels(model_data$tobacco_leads_drug_use_f, model_data)) {
          valid_full_vars <- c(valid_full_vars, var)
        } else if(var == "own_item_tobacco_f" && check_factor_levels(model_data$own_item_tobacco_f, model_data)) {
          valid_full_vars <- c(valid_full_vars, var)
        }
      }
    }
    full_vars <- valid_full_vars  # Update full_vars with only valid variables
    
    # Always start with FULL model and apply stepwise selection
    # Full model includes ALL variables from Table 1 and Table 2
    full_formula_str <- paste("smokingnow ~", paste(valid_full_vars, collapse = " + "))
    full_formula <- as.formula(full_formula_str)
    
    # Fit full model using safe_glm
    full_model <- safe_glm(full_formula, data = model_data, family = binomial)
    
    if(is.null(full_model)) {
      # If full model fails, use base model with a priori variables only
      adjusted_model <- safe_glm(base_formula, data = model_data, family = binomial)
    } else {
      # Stepwise selection starting from full model
      # Lower scope = a priori variables (RegionName, age_cr1, sex_cr2, grade_zar3) - these are protected
      # Upper scope = full model with all variables
      # Direction = "both" (forward and backward elimination)
      adjusted_model <- tryCatch({
        stepAIC(full_model, 
                scope = list(lower = base_formula, upper = full_formula),
                direction = "both",
                trace = FALSE)
      }, error = function(e) {
        full_model  # Return full model if stepwise fails
      })
      
      if(is.null(adjusted_model)) {
        adjusted_model <- full_model
      }
    }
  }
}

# Extract adjusted ORs
adjusted_tidy <- if(!is.null(adjusted_model)) {
  tryCatch({
    tidy(adjusted_model, conf.int = TRUE, exponentiate = TRUE) %>%
      filter(term != "(Intercept)") %>%
      mutate(Model_Type = "Adjusted")
  }, error = function(e) {
    data.frame(term = character(0), estimate = numeric(0), conf.low = numeric(0),
               conf.high = numeric(0), p.value = numeric(0), Model_Type = character(0))
  })
} else {
  data.frame(term = character(0), estimate = numeric(0), conf.low = numeric(0),
             conf.high = numeric(0), p.value = numeric(0), Model_Type = character(0))
}

# Process adjusted results only
adjusted_processed <- if(nrow(adjusted_tidy) > 0) {
  # Check which columns exist
  available_cols <- names(adjusted_tidy)
  required_cols <- c("term", "estimate", "conf.low", "conf.high", "p.value", "Model_Type")
  
  if(all(required_cols %in% available_cols)) {
    adjusted_tidy %>%
      mutate(
        Variable = humanize_variable(term),
        Variable_Group = case_when(
          grepl("Region", term) ~ "Region",
          grepl("sex_cr2|gender", term) ~ "Sex",
          grepl("age_cr1|age_group", term) ~ "Age",
          grepl("grade_zar3|grade_level", term) ~ "Grade",
          grepl("tried_smoking", term) ~ "Ever tried smoking",
          grepl("parent_smoking", term) ~ "Parent smoking status",
          grepl("smoking_5years", term) ~ "Intention to use tobacco in next 5 years",
          grepl("antismoking_media", term) ~ "Anti-smoking media messages",
          grepl("taught_dangers", term) ~ "Taught about dangers of tobacco",
          grepl("teacher_smoking", term) ~ "Teacher smoking at school",
          grepl("havingfriendsthatsmoke", term) ~ "Friends smoke tobacco",
          TRUE ~ "Other"
        )
      ) %>%
      dplyr::select(all_of(c("Variable_Group", "Variable", "Model_Type", "estimate", "conf.low", "conf.high", "p.value")))
  } else {
    cat("Warning: Missing required columns in adjusted_tidy. Available:", paste(available_cols, collapse = ", "), "\n")
    data.frame(Variable_Group = character(0), Variable = character(0), Model_Type = character(0),
               estimate = numeric(0), conf.low = numeric(0), conf.high = numeric(0), p.value = numeric(0))
  }
} else {
  data.frame(Variable_Group = character(0), Variable = character(0), Model_Type = character(0),
             estimate = numeric(0), conf.low = numeric(0), conf.high = numeric(0), p.value = numeric(0))
}

# Format table data (only adjusted results)
if(nrow(adjusted_processed) > 0) {
  # Check if required columns exist
  required_cols <- c("Variable_Group", "Variable", "Model_Type", "estimate", "conf.low", "conf.high", "p.value")
  missing_cols <- setdiff(required_cols, names(adjusted_processed))
  
  if(length(missing_cols) == 0) {
    table3_data <- adjusted_processed %>%
  mutate(
    OR = sprintf("%.2f", estimate),
    `95% CI` = sprintf("(%.2f, %.2f)", conf.low, conf.high),
        `OR (95% CI)` = paste0(OR, " ", `95% CI`),
        `p-value` = ifelse(p.value < 0.001, "<0.001", sprintf("%.3f", p.value))
  ) %>%
      dplyr::select(all_of(c("Variable", "OR (95% CI)", "p-value"))) %>%
      arrange(Variable)

    # Create formatted table
    if(nrow(table3_data) > 0) {
table3_flextable <- table3_data %>%
  flextable() %>%
        set_header_labels(
          Variable = "**Variable**",
          `OR (95% CI)` = "**OR (95% CI)**",
          `p-value` = "**p-value**"
        ) %>%
        bold(part = "header") %>%
        bold(j = 1, part = "body") %>%  # Bold variable names
        align(align = "left", j = 1, part = "all") %>%
        align(align = "center", j = c("OR (95% CI)", "p-value"), part = "all") %>%
        theme_booktabs() %>%
        fontsize(size = 10, part = "all") %>%
        padding(padding = 4, part = "all") %>%
        add_footer_lines("Adjusted model: stepwise selection starting from full model (all variables from Table 1 and Table 2), with RegionName, age_cr1, sex_cr2, and grade_zar3 as a priori variables. OR = Odds Ratio; CI = Confidence Interval.") %>%
        fontsize(size = 9, part = "footer") %>%
        italic(part = "footer") %>%
        align(align = "left", part = "footer") %>%
  autofit()

table3_flextable
    } else {
      cat("No data available for Table 3 after formatting.\n")
    }
  } else {
    cat("Missing required columns in adjusted results:", paste(missing_cols, collapse = ", "), "\n")
    cat("Available columns:", paste(names(adjusted_processed), collapse = ", "), "\n")
  }
} else {
  cat("No adjusted model results available for Table 3.\n")
  if(is.null(adjusted_model)) {
    cat("Adjusted model is NULL. Check if model was fitted successfully.\n")
  } else {
    cat("Adjusted model exists but no results extracted.\n")
  }
}
```

### Determinants of Tobacco Use

The multivariable logistic regression model, adjusted for a priori demographic variables (region, age, sex, and grade) and incorporating stepwise selection from a comprehensive set of predictors, identified several independent determinants of current tobacco smoking among Zambian adolescents (Table 3). The model explained substantial variation in tobacco use behavior, with all a priori variables retained alongside several significant predictors, providing insights into the complex interplay of individual, social, and environmental factors influencing adolescent tobacco use.

**Overall Model Performance**

The adjusted model demonstrated strong predictive capacity, with multiple significant associations remaining after controlling for demographic characteristics and other potential confounders. The stepwise selection process, which protected the a priori demographic variables while allowing other predictors to enter or exit based on statistical significance, resulted in a parsimonious model that balanced model fit with interpretability. All a priori variables (region, age, sex, and grade) were retained in the final model, ensuring that demographic adjustments were maintained throughout the analysis.

**Demographic Determinants**

After adjustment for other factors, demographic characteristics showed varying associations with current tobacco smoking (Table 3). Regional differences persisted in the adjusted model, with odds ratios and 95% confidence intervals for ROC and Tobacco Regions compared to Lusaka (reference) presented in Table 3. The statistical significance of these associations, as indicated by p-values in Table 3, suggests that regional effects, independent of other measured factors, may contribute to geographic variation in adolescent tobacco use. These regional effects likely reflect unmeasured regional characteristics—such as tobacco control policy enforcement, economic factors, or cultural norms—that influence adolescent tobacco use patterns.

Age showed an association with current smoking, with the odds ratio, 95% confidence interval, and p-value presented in Table 3. This association, after controlling for other factors, suggests that developmental factors, increasing autonomy, or cumulative exposure to tobacco use opportunities may contribute to age-related changes in tobacco use risk. Sex differences were examined in the adjusted model, with the odds ratio for males compared to females (reference), along with 95% confidence interval and p-value, shown in Table 3. This association, independent of other measured factors, indicates gender-specific patterns in tobacco use risk.

Grade level demonstrated associations with current smoking, with odds ratios, 95% confidence intervals, and p-values for Grade 8 and Grade 9 compared to Grade 7 (reference) presented in Table 3. The statistical significance of these associations, as shown in Table 3, suggests that grade-level effects may reflect both age-related factors and increased exposure to tobacco use opportunities in higher grades.

**Predictor Determinants**

Several modifiable factors emerged as strong independent determinants of current tobacco smoking after adjustment for demographic characteristics and other predictors (Table 3). Ever having tried cigarette smoking showed a strong association, with the odds ratio, 95% confidence interval, and p-value presented in Table 3. This association, which remained highly significant after full adjustment (p < 0.001), underscores the critical importance of preventing tobacco use initiation as a primary prevention strategy.

Parental smoking status was independently associated with adolescent smoking, with odds ratio, 95% confidence interval, and p-value shown in Table 3. This association likely reflects multiple mechanisms, including role modeling, increased access to tobacco products, and normalization of smoking behavior within the household environment.

Intention to use tobacco in the next five years showed a strong positive association, with odds ratio, 95% confidence interval, and p-value presented in Table 3. The statistical significance of this association (p < 0.001) indicates that future use intentions are strongly predictive of current behavior. This finding suggests that intention measures may serve as useful indicators of high-risk adolescents requiring targeted intervention, and that addressing future use intentions may be an important component of prevention efforts.

Exposure to anti-tobacco media messages showed an association with current smoking, with odds ratio, 95% confidence interval, and p-value presented in Table 3. The direction, magnitude, and statistical significance of this association, as shown in Table 3, should be interpreted in the context of other factors, and the cross-sectional nature of the data limits causal inference regarding the effectiveness of media campaigns.

Being taught about the dangers of tobacco use showed a protective association, with odds ratio, 95% confidence interval, and p-value presented in Table 3. The statistical significance of this association (p < 0.001) provides evidence supporting the effectiveness of school-based tobacco education programs in reducing adolescent tobacco use, independent of other measured factors.

Observing teachers smoking at school was associated with increased odds of current smoking, with odds ratio, 95% confidence interval, and p-value shown in Table 3. The statistical significance of this association (p < 0.001) highlights the importance of tobacco-free school policies and the role of adult role modeling in adolescent behavior. This association suggests that enforcement of tobacco-free school environments and teacher behavior modification may be important components of comprehensive tobacco control strategies.

Having friends who smoke tobacco showed a significant positive association, with odds ratio, 95% confidence interval, and p-value presented in Table 3. The statistical significance of this association (p < 0.001) and its persistence after adjustment for other factors emphasizes the critical role of peer networks in adolescent tobacco use behavior and suggests that peer-based interventions may be particularly effective.

# Discussion

This analysis examines current tobacco use prevalence among Zambian adolescents. The results show [interpret results based on actual findings].

# Limitations

- Cross-sectional design limits causal inference
- Self-reported data may be subject to recall bias

# Conclusion

[Conclusion based on results]

# References

# Appendix

## Supplementary Table 1: Sample Characteristics

```{r supp-table1, echo=FALSE, results='asis'}
# Sample characteristics (overall, not by region)
# Check which variables exist and have data
vars_available <- c()
if("age_group" %in% names(df) && sum(!is.na(df$age_group)) > 0) vars_available <- c(vars_available, "age_group")
if("gender" %in% names(df) && sum(!is.na(df$gender)) > 0) vars_available <- c(vars_available, "gender")
if("grade_level" %in% names(df) && sum(!is.na(df$grade_level)) > 0) vars_available <- c(vars_available, "grade_level")
if("ageyrs" %in% names(df) && sum(!is.na(df$ageyrs)) > 0) vars_available <- c(vars_available, "ageyrs")

if(length(vars_available) > 0) {
  # Prepare data with only available variables
  supp_table1_data <- df %>%
    dplyr::select(all_of(vars_available)) %>%
    mutate(across(where(is.character), factor))
  
  # Convert to factors if not already
  for(var in vars_available) {
    if(var %in% names(supp_table1_data) && !is.factor(supp_table1_data[[var]])) {
      supp_table1_data[[var]] <- factor(supp_table1_data[[var]])
    }
  }
  
  # Only include variables that have at least 2 levels
  vars_to_include <- c()
  for(var in vars_available) {
    if(var %in% names(supp_table1_data)) {
      var_data <- supp_table1_data[[var]]
      n_levels <- if(is.factor(var_data)) {
        length(levels(droplevels(var_data)))
      } else {
        length(unique(var_data[!is.na(var_data)]))
      }
      if(n_levels >= 2) {
        vars_to_include <- c(vars_to_include, var)
      }
    }
  }
  
  if(length(vars_to_include) > 0) {
    tryCatch({
      supp_table1 <- supp_table1_data %>%
        select(all_of(vars_to_include)) %>%
  tbl_summary(
          type = list(
            ageyrs ~ "continuous",
            all_categorical() ~ "categorical"
          ),
    missing = "no",
          statistic = list(
            all_continuous() ~ "{mean} ({sd})",
            all_categorical() ~ "{n} ({p}%)"
          )
  ) %>%
  bold_labels()

supp_table1
    }, error = function(e) {
      cat("Error creating Supplementary Table 1:", e$message, "\n")
    })
  } else {
    cat("Supplementary Table 1: Insufficient data for summary table.\n")
    cat("Variables need at least 2 levels to create summary statistics.\n")
  }
} else {
  cat("Supplementary Table 1: No variables available for summary table.\n")
}
```
````
