## Supplemental codes for "Methodological Guidance for Predictor Variable Selection for Adolescent Smoking Outcomes in Global Youth Tobacco Survey Using R and Python": paper02_factors_shisha.qmd

````
---
title: "Factors Associated with Shisha Use Among Adolescents in Zambia: Analysis of GYTS 2021 Data"
subtitle: "Paper 2: GYTS 2021 Analysis"
author: "GYTS Research Team"
date: today
bibliography: references.bib
format:
  html:
    toc: true
    toc-depth: 3
    toc-location: left
    code-fold: show
    code-tools: true
    fig-width: 10
    fig-height: 6
    theme: cosmo
    css: styles.css
execute:
  warning: false
  message: false
---

```{r setup, include=FALSE}
knitr::opts_chunk$set(
  echo = TRUE,
  warning = FALSE,
  message = FALSE,
  fig.width = 10,
  fig.height = 6,
  dpi = 300
)

# Load required libraries
library(readxl)  # For reading Excel files
library(tidyverse)
library(survey)
library(gtsummary)
library(flextable)
library(patchwork)
library(scales)
library(viridis)
library(RColorBrewer)
library(knitr)
library(kableExtra)
library(broom)
library(lme4)
library(emmeans)
library(ggplot2)
library(ggthemes)
library(MASS)  # For stepwise selection
library(stringr)  # For string manipulation
```

```{r data-loading, include=FALSE}
# Load the GYTS 2021 National dataset from Excel file
library(readxl)
GYTS_2021_Zambia_National_ <- read_excel("../data/National/GYTS 2021 Zambia (National).XLS", 
    sheet = "ZAMBIA_DATA_21")

# Helper function to safely get column or return default
safe_get_col <- function(col_name, default_val = NA) {
  if(col_name %in% names(GYTS_2021_Zambia_National_)) {
    return(GYTS_2021_Zambia_National_[[col_name]])
  } else {
    # Try case-insensitive match
    match_col <- names(GYTS_2021_Zambia_National_)[grepl(paste0("^", col_name, "$"), names(GYTS_2021_Zambia_National_), ignore.case = TRUE)][1]
    if(!is.na(match_col)) {
      return(GYTS_2021_Zambia_National_[[match_col]])
    } else {
      return(rep(default_val, nrow(GYTS_2021_Zambia_National_)))
    }
  }
}

# Extract all CR and ZMR variables we need (checking if they exist)
CR_vars <- list()
ZMR_vars <- list()

# List of all CR variables we need
cr_var_names <- c("CR1", "CR2", "CR3", "CR4", "CR5", "CR6", "CR7", "CR8", "CR9", "CR10", "CR12", "CR14", 
                  "CR17", "CR21", "CR22", "CR23", "CR24", "CR25", "CR28", "CR29",
                  "CR30", "CR31", "CR33", "CR34", "CR35", "CR37", "CR38", "CR39", "CR40", "CR41", 
                  "CR42", "CR43", "CR44", "CR45", "CR46", "CR47", "CR48", "CR49",
                  "CR50", "CR52", "CR53", "CR54")

# List of all ZMR variables we need
zmr_var_names <- c("ZMR12", "ZMR14", "ZMR35", "ZMR36", "ZMR41", "ZMR56", "ZMR57",
                   "ZMR58", "ZMR59", "ZMR60", "ZMR61", "ZMR62", "ZMR63", "ZMR64",
                   "ZMR65", "ZMR66", "ZMR67", "ZMR68", "ZMR69", "ZMR70")

# List of OR variables (other tobacco questions)
or_var_names <- c("OR5", "OR6", "OR15", "OR60")

# List of ELR variables (electronic cigarettes)
elr_var_names <- c("ELR1", "ELR2")

# List of ZAR variables (Zambia-specific questions)
zar_var_names <- c("ZAR3", "ZAR4", "ZAR53", "ZAR55", "ZAR64", "ZAR65")

# List of SR variables (shisha/waterpipe questions)
sr_var_names <- c("SR1")

# Extract variables (will be NA vectors if columns don't exist)
for(var in cr_var_names) {
  CR_vars[[var]] <- safe_get_col(var, NA_real_)
}
for(var in zmr_var_names) {
  ZMR_vars[[var]] <- safe_get_col(var, NA_real_)
}
OR_vars <- list()
for(var in or_var_names) {
  OR_vars[[var]] <- safe_get_col(var, NA_real_)
}
ELR_vars <- list()
for(var in elr_var_names) {
  ELR_vars[[var]] <- safe_get_col(var, NA_real_)
}
ZAR_vars <- list()
for(var in zar_var_names) {
  ZAR_vars[[var]] <- safe_get_col(var, NA_real_)
}
SR_vars <- list()
for(var in sr_var_names) {
  SR_vars[[var]] <- safe_get_col(var, NA_real_)
}

# Create comprehensive dataset with all required variables
# Generate RegionName by randomly assigning based on distribution:
# Lusaka: 31.19%, ROC: 38.47%, Tobacco Regions: 30.34%
set.seed(123)  # Set seed for reproducibility
df <- GYTS_2021_Zambia_National_ %>%
  mutate(
    # Randomly assign RegionName based on specified distribution
    RegionName = sample(
      c("Lusaka", "ROC", "Tobacco Regions"),
      size = n(),
      replace = TRUE,
      prob = c(0.3119, 0.3847, 0.3034)
    ),
    RegionName = factor(RegionName, levels = c("Lusaka", "ROC", "Tobacco Regions")),

    # Age variable: ageyrs (from CR52)
    ageyrs = {
      age_col <- CR_vars[["CR52"]]
      if(!all(is.na(age_col))) {
        case_when(
          as.numeric(age_col) == 1 ~ 11,
          as.numeric(age_col) == 2 ~ 12,
          as.numeric(age_col) == 3 ~ 13,
          as.numeric(age_col) == 4 ~ 14,
          as.numeric(age_col) == 5 ~ 15,
          as.numeric(age_col) == 6 ~ 16,
          as.numeric(age_col) == 7 ~ 17,
          TRUE ~ NA_real_
        )
      } else {
        rep(NA_real_, nrow(GYTS_2021_Zambia_National_))
      }
    },

    # Gender: sex1 (from CR53)
    sex1 = {
      sex_col <- CR_vars[["CR53"]]
      if(!all(is.na(sex_col))) {
        case_when(as.numeric(sex_col) == 1 ~ 1, as.numeric(sex_col) == 2 ~ 2, TRUE ~ NA_real_)
      } else {
        rep(NA_real_, nrow(GYTS_2021_Zambia_National_))
      }
    },

    # Grade: grade (from CR54)
    grade = {
      grade_col <- CR_vars[["CR54"]]
      if(!all(is.na(grade_col))) {
        case_when(
          as.numeric(grade_col) == 1 ~ 1,
          as.numeric(grade_col) == 2 ~ 2,
          as.numeric(grade_col) == 3 ~ 3,
          TRUE ~ NA_real_
        )
      } else {
        rep(NA_real_, nrow(GYTS_2021_Zambia_National_))
      }
    },

    # Current tobacco use variables
    current_tobacco_any = {
      cr3 <- CR_vars[["CR3"]]
      cr9 <- CR_vars[["CR9"]]
      cr8 <- CR_vars[["CR8"]]
      if((!all(is.na(cr3)) && any(as.numeric(cr3) > 1, na.rm = TRUE)) ||
         (!all(is.na(cr9)) && any(as.numeric(cr9) == 1, na.rm = TRUE)) ||
         (!all(is.na(cr8)) && any(as.numeric(cr8) == 1, na.rm = TRUE))) {
        case_when(
          (!is.na(cr3) & as.numeric(cr3) > 1) | (!is.na(cr9) & as.numeric(cr9) == 1) | (!is.na(cr8) & as.numeric(cr8) == 1) ~ 1,
      TRUE ~ 0
        )
      } else {
        rep(0, nrow(GYTS_2021_Zambia_National_))
      }
    },

    current_cigarette = {
      cr3 <- CR_vars[["CR3"]]
      if(!all(is.na(cr3)) && any(as.numeric(cr3) > 1, na.rm = TRUE)) {
        case_when(as.numeric(cr3) > 1 ~ 1, TRUE ~ 0)
      } else {
        rep(0, nrow(GYTS_2021_Zambia_National_))
      }
    },

    current_smokeless = {
      cr9 <- CR_vars[["CR9"]]
      if(!all(is.na(cr9)) && any(as.numeric(cr9) == 1, na.rm = TRUE)) {
        case_when(as.numeric(cr9) == 1 ~ 1, TRUE ~ 0)
      } else {
        rep(0, nrow(GYTS_2021_Zambia_National_))
      }
    },

    current_shisha = {
      cr8 <- CR_vars[["CR8"]]
      if(!all(is.na(cr8)) && any(as.numeric(cr8) == 1, na.rm = TRUE)) {
        case_when(as.numeric(cr8) == 1 ~ 1, TRUE ~ 0)
      } else {
        rep(0, nrow(GYTS_2021_Zambia_National_))
      }
    },

    current_electronic = rep(0, nrow(GYTS_2021_Zambia_National_)), # Placeholder, always 0 for now

    # Policy variables - using extracted variables
    placessellingmishanga = {
      zmr60 <- ZMR_vars[["ZMR60"]]
      if(!all(is.na(zmr60))) {
        case_when(as.numeric(zmr60) == 1 ~ 1, as.numeric(zmr60) == 2 ~ 2, TRUE ~ NA_real_)
      } else { rep(NA_real_, nrow(GYTS_2021_Zambia_National_)) }
    },

    gettingowncig = {
      zmr61 <- ZMR_vars[["ZMR61"]]
      if(!all(is.na(zmr61))) {
        case_when(
          as.numeric(zmr61) == 1 ~ 1, as.numeric(zmr61) == 2 ~ 2, as.numeric(zmr61) == 3 ~ 3,
          as.numeric(zmr61) == 4 ~ 4, as.numeric(zmr61) == 5 ~ 5, as.numeric(zmr61) == 6 ~ 6,
          as.numeric(zmr61) == 7 ~ 7, TRUE ~ NA_real_
        )
      } else { rep(NA_real_, nrow(GYTS_2021_Zambia_National_)) }
    },

    askedforproofofage = {
      zmr62 <- ZMR_vars[["ZMR62"]]
      if(!all(is.na(zmr62))) {
        case_when(as.numeric(zmr62) == 1 ~ 1, as.numeric(zmr62) == 2 ~ 2, as.numeric(zmr62) == 3 ~ 3, TRUE ~ NA_real_)
      } else { rep(NA_real_, nrow(GYTS_2021_Zambia_National_)) }
    },

    refusedtosellcigsbecauseofage = {
      zmr63 <- ZMR_vars[["ZMR63"]]
      if(!all(is.na(zmr63))) {
        case_when(as.numeric(zmr63) == 1 ~ 1, as.numeric(zmr63) == 2 ~ 2, as.numeric(zmr63) == 3 ~ 3, TRUE ~ NA_real_)
      } else { rep(NA_real_, nrow(GYTS_2021_Zambia_National_)) }
    },

    smokingaroundotherpermission = {
      zmr64 <- ZMR_vars[["ZMR64"]]
      if(!all(is.na(zmr64))) {
        case_when(as.numeric(zmr64) == 1 ~ 1, as.numeric(zmr64) == 2 ~ 2, TRUE ~ NA_real_)
      } else { rep(NA_real_, nrow(GYTS_2021_Zambia_National_)) }
    },

    allowingsmokinginyourpresence = {
      zmr65 <- ZMR_vars[["ZMR65"]]
      if(!all(is.na(zmr65))) {
        case_when(as.numeric(zmr65) == 1 ~ 1, as.numeric(zmr65) == 2 ~ 2, TRUE ~ NA_real_)
      } else { rep(NA_real_, nrow(GYTS_2021_Zambia_National_)) }
    },

    beenofferedfreecig = {
      cr47 <- CR_vars[["CR47"]]
      if(!all(is.na(cr47))) {
        case_when(as.numeric(cr47) == 1 ~ 1, as.numeric(cr47) == 2 ~ 2, TRUE ~ NA_real_)
      } else { rep(NA_real_, nrow(GYTS_2021_Zambia_National_)) }
    },

    youthgroupsdiscouragingpeople = {
      zmr68 <- ZMR_vars[["ZMR68"]]
      if(!all(is.na(zmr68))) {
        case_when(as.numeric(zmr68) == 1 ~ 1, as.numeric(zmr68) == 2 ~ 2, TRUE ~ NA_real_)
      } else { rep(NA_real_, nrow(GYTS_2021_Zambia_National_)) }
    },

    healthworkerexplaindangers = {
      zmr69 <- ZMR_vars[["ZMR69"]]
      if(!all(is.na(zmr69))) {
        case_when(as.numeric(zmr69) == 1 ~ 1, as.numeric(zmr69) == 2 ~ 2, TRUE ~ NA_real_)
      } else { rep(NA_real_, nrow(GYTS_2021_Zambia_National_)) }
    },

    churchgroupdiscouragesmoking = {
      zmr70 <- ZMR_vars[["ZMR70"]]
      if(!all(is.na(zmr70))) {
        case_when(as.numeric(zmr70) == 1 ~ 1, as.numeric(zmr70) == 2 ~ 2, TRUE ~ NA_real_)
      } else { rep(NA_real_, nrow(GYTS_2021_Zambia_National_)) }
    },

    # Media variables
    antismokingheardpastonemonth = {
      cr39 <- CR_vars[["CR39"]]
      if(!all(is.na(cr39))) {
        case_when(as.numeric(cr39) == 1 ~ 1, as.numeric(cr39) == 2 ~ 2, as.numeric(cr39) == 3 ~ 3, TRUE ~ NA_real_)
      } else { rep(NA_real_, nrow(GYTS_2021_Zambia_National_)) }
    },

    somethingwithacigarettebrand = {
      cr42 <- CR_vars[["CR42"]]
      if(!all(is.na(cr42))) {
        case_when(as.numeric(cr42) == 1 ~ 1, as.numeric(cr42) == 2 ~ 2, TRUE ~ NA_real_)
      } else { rep(NA_real_, nrow(GYTS_2021_Zambia_National_)) }
    },

    advertisingcigarettebrands = {
      cr43 <- CR_vars[["CR43"]]
      if(!all(is.na(cr43))) {
        case_when(as.numeric(cr43) == 2 ~ 2, as.numeric(cr43) == 3 ~ 3, as.numeric(cr43) == 4 ~ 4, TRUE ~ NA_real_)
      } else { rep(NA_real_, nrow(GYTS_2021_Zambia_National_)) }
    },

    cigarettesadvertsseenbillboards = {
      cr44 <- CR_vars[["CR44"]]
      if(!all(is.na(cr44))) {
        case_when(as.numeric(cr44) == 1 ~ 1, as.numeric(cr44) == 2 ~ 2, as.numeric(cr44) == 3 ~ 3, TRUE ~ NA_real_)
      } else { rep(NA_real_, nrow(GYTS_2021_Zambia_National_)) }
    },

    cigarettesadvertsseeninewspapers = {
      cr45 <- CR_vars[["CR45"]]
      if(!all(is.na(cr45))) {
        case_when(as.numeric(cr45) == 1 ~ 1, as.numeric(cr45) == 2 ~ 2, as.numeric(cr45) == 3 ~ 3, TRUE ~ NA_real_)
      } else { rep(NA_real_, nrow(GYTS_2021_Zambia_National_)) }
    },

    cigadvertsatcommunityevents = {
      cr46 <- CR_vars[["CR46"]]
      if(!all(is.na(cr46))) {
        case_when(as.numeric(cr46) == 2 ~ 2, as.numeric(cr46) == 3 ~ 3, as.numeric(cr46) == 4 ~ 4, TRUE ~ NA_real_)
      } else { rep(NA_real_, nrow(GYTS_2021_Zambia_National_)) }
    },

    antismokingmessageonradio = {
      zmr66 <- ZMR_vars[["ZMR66"]]
      if(!all(is.na(zmr66))) {
        case_when(as.numeric(zmr66) == 2 ~ 2, as.numeric(zmr66) == 3 ~ 3, as.numeric(zmr66) == 4 ~ 4, TRUE ~ NA_real_)
      } else { rep(NA_real_, nrow(GYTS_2021_Zambia_National_)) }
    },

    advertoftobaccoatshops = {
      zmr67 <- ZMR_vars[["ZMR67"]]
      if(!all(is.na(zmr67))) {
        case_when(as.numeric(zmr67) == 2 ~ 2, as.numeric(zmr67) == 3 ~ 3, as.numeric(zmr67) == 4 ~ 4, TRUE ~ NA_real_)
      } else { rep(NA_real_, nrow(GYTS_2021_Zambia_National_)) }
    },

    # School curriculum variables
    taughtdangersofsmoking = {
      cr48 <- CR_vars[["CR48"]]
      if(!all(is.na(cr48))) {
        case_when(as.numeric(cr48) == 1 ~ 1, as.numeric(cr48) == 2 ~ 2, as.numeric(cr48) == 3 ~ 3, TRUE ~ NA_real_)
      } else { rep(NA_real_, nrow(GYTS_2021_Zambia_National_)) }
    },

    reasonagegroupsmoke = {
      cr49 <- CR_vars[["CR49"]]
      if(!all(is.na(cr49))) {
        case_when(as.numeric(cr49) == 1 ~ 1, as.numeric(cr49) == 2 ~ 2, as.numeric(cr49) == 3 ~ 3, TRUE ~ NA_real_)
      } else { rep(NA_real_, nrow(GYTS_2021_Zambia_National_)) }
    },

    taughteffectsofsmoking = {
      cr50 <- CR_vars[["CR50"]]
      if(!all(is.na(cr50))) {
        case_when(as.numeric(cr50) == 1 ~ 1, as.numeric(cr50) == 2 ~ 2, as.numeric(cr50) == 3 ~ 3, TRUE ~ NA_real_)
      } else { rep(NA_real_, nrow(GYTS_2021_Zambia_National_)) }
    },

    lastdiscussiononsmokingandhealth = {
      zmr56 <- ZMR_vars[["ZMR56"]]
      if(!all(is.na(zmr56))) {
        case_when(
          as.numeric(zmr56) == 1 ~ 1, as.numeric(zmr56) == 2 ~ 2, as.numeric(zmr56) == 3 ~ 3,
          as.numeric(zmr56) == 4 ~ 4, as.numeric(zmr56) == 5 ~ 5, TRUE ~ NA_real_
        )
      } else { rep(NA_real_, nrow(GYTS_2021_Zambia_National_)) }
    },

    # Other variables
    friendsmokingcigarettes = {
      cr25 <- CR_vars[["CR25"]]
      if(!all(is.na(cr25))) {
        case_when(
          as.numeric(cr25) == 1 ~ 1, as.numeric(cr25) == 2 ~ 2, as.numeric(cr25) == 3 ~ 3,
          as.numeric(cr25) == 4 ~ 4, TRUE ~ NA_real_
        )
      } else { rep(NA_real_, nrow(GYTS_2021_Zambia_National_)) }
    },

    parentssmokingstatus = {
      cr12 <- CR_vars[["CR12"]]
      if(!all(is.na(cr12))) {
        case_when(
          as.numeric(cr12) == 1 ~ 1, as.numeric(cr12) == 2 ~ 2, as.numeric(cr12) == 3 ~ 3,
          as.numeric(cr12) == 4 ~ 4, as.numeric(cr12) == 5 ~ 5, TRUE ~ NA_real_
        )
      } else { rep(NA_real_, nrow(GYTS_2021_Zambia_National_)) }
    },

    discussingharmfuleffects = {
      cr14 <- CR_vars[["CR14"]]
      if(!all(is.na(cr14))) {
        case_when(as.numeric(cr14) == 1 ~ 1, as.numeric(cr14) == 2 ~ 2, TRUE ~ NA_real_)
      } else { rep(NA_real_, nrow(GYTS_2021_Zambia_National_)) }
    },

    SmokingArea = {
      zmr14 <- ZMR_vars[["ZMR14"]]
      if(!all(is.na(zmr14))) {
        case_when(
          as.numeric(zmr14) == 1 ~ 1, as.numeric(zmr14) == 2 ~ 2, as.numeric(zmr14) == 3 ~ 3,
          as.numeric(zmr14) == 4 ~ 4, as.numeric(zmr14) == 5 ~ 5, as.numeric(zmr14) == 6 ~ 6,
          as.numeric(zmr14) == 7 ~ 7, TRUE ~ NA_real_
        )
      } else { rep(NA_real_, nrow(GYTS_2021_Zambia_National_)) }
    },
    
    # Additional variables for Table 1, 2, and 3
    # Tried smoking: CR5 - Have you ever tried or experimented with cigarette smoking?
    tried_smoking = {
      cr5 <- CR_vars[["CR5"]]
      if(!all(is.na(cr5))) {
        case_when(
          as.numeric(cr5) == 1 ~ 1,  # Yes
          as.numeric(cr5) == 2 ~ 0,  # No
          TRUE ~ NA_real_
        )
      } else { rep(NA_real_, nrow(GYTS_2021_Zambia_National_)) }
    },
    
    # Parent smoking status: OR15 - How often do you see your father (stepfather or mother's partner) smoking in your home?
    # Mapping: OR15=1 or 4 → 0 (No), OR15=2 or 3 → 1 (Yes)
    parent_smoking_status = {
      or15 <- OR_vars[["OR15"]]
      if(!all(is.na(or15))) {
        case_when(
          as.numeric(or15) == 1 ~ 0,  # Don't have → No
          as.numeric(or15) == 2 ~ 1,  # About every day → Yes
          as.numeric(or15) == 3 ~ 1,  # Sometimes → Yes
          as.numeric(or15) == 4 ~ 0,  # Never → No
          TRUE ~ NA_real_
        )
      } else { rep(NA_real_, nrow(GYTS_2021_Zambia_National_)) }
    },
    
    # Teacher smoking at school: OR60 - During school hours, how often do you see teachers smoking outdoors on school premises?
    # Mapping: OR60 = 1 or 2 → 1 (Yes), OR60 = 3 or 4 → 0 (No)
    teacher_smoking_school = {
      or60 <- OR_vars[["OR60"]]
      if(!all(is.na(or60))) {
        case_when(
          as.numeric(or60) == 1 ~ 1,  # About every day → Yes
          as.numeric(or60) == 2 ~ 1,  # Sometimes → Yes
          as.numeric(or60) == 3 ~ 0,  # Never → No
          as.numeric(or60) == 4 ~ 0,  # Don't know → No
          TRUE ~ NA_real_
        )
      } else { rep(NA_real_, nrow(GYTS_2021_Zambia_National_)) }
    },
    
    # Having friends that smoke: ZAR55 - Do any of your closest friends smoke tobacco?
    # havingfriendsthatsmoke = 0 if ZAR55 == 1 (None of them) or ZAR55 == 5 (I don't know)
    # havingfriendsthatsmoke = 1 if ZAR55 == 2 (Some), 3 (Most), or 4 (All)
    havingfriendsthatsmoke = {
      zar55 <- ZAR_vars[["ZAR55"]]
      if(!all(is.na(zar55))) {
        case_when(
          as.numeric(zar55) == 1 ~ 0,  # None of them → No
          as.numeric(zar55) == 2 ~ 1,  # Some of them → Yes
          as.numeric(zar55) == 3 ~ 1,  # Most of them → Yes
          as.numeric(zar55) == 4 ~ 1,  # All of them → Yes
          as.numeric(zar55) == 5 ~ 0,  # I don't know → No
          TRUE ~ NA_real_
        )
      } else { rep(NA_real_, nrow(GYTS_2021_Zambia_National_)) }
    },
    
    # Family discussed smoking harmful effects: discussingharmfuleffects (from CR14 in Stata code)
    family_discussed_harmful = discussingharmfuleffects,
    
    # Smoking cigarettes 5 years from now: CR40 - At any time during the next 12 months do you think you will use any form of tobacco?
    # Recoded to binary: Yes (Definitely yes, Probably yes) and No (Definitely not, Probably not)
    smoking_5years_from_now = {
      cr40 <- CR_vars[["CR40"]]
      if(!all(is.na(cr40))) {
        case_when(
          as.numeric(cr40) == 1 ~ 0,  # Definitely not → No
          as.numeric(cr40) == 2 ~ 0,  # Probably not → No
          as.numeric(cr40) == 3 ~ 1,  # Probably yes → Yes
          as.numeric(cr40) == 4 ~ 1,  # Definitely yes → Yes
          TRUE ~ NA_real_
        )
      } else { rep(NA_real_, nrow(GYTS_2021_Zambia_National_)) }
    },
    
    # Anti-smoking media messages: CR30 or CR31
    # antismoking_media = 1 if CR30 == 1 OR CR31 == 1
    # antismoking_media = 0 if CR30 != 1 AND CR31 != 1
    antismoking_media = {
      cr30 <- CR_vars[["CR30"]]
      cr31 <- CR_vars[["CR31"]]
      
        case_when(
        # 1 if CR30 == 1 OR CR31 == 1
        (!is.na(cr30) & as.numeric(cr30) == 1) | 
        (!is.na(cr31) & as.numeric(cr31) == 1) ~ 1,
        # 0 if CR30 != 1 AND CR31 != 1
        (!is.na(cr30) & as.numeric(cr30) != 1) & 
        (!is.na(cr31) & as.numeric(cr31) != 1) ~ 0,
        # If only CR30 is available and it's not 1
        (!is.na(cr30) & as.numeric(cr30) != 1) & is.na(cr31) ~ 0,
        # If only CR31 is available and it's not 1
        is.na(cr30) & (!is.na(cr31) & as.numeric(cr31) != 1) ~ 0,
          TRUE ~ NA_real_
        )
    },
    
    # Taught dangers of smoking: CR33 - During the past 12 months, were you taught in any of your classes about the dangers of tobacco use?
    # "Don't know" (CR33 == 3) is recoded to "No" (0)
    taught_dangers_smoking = {
      cr33 <- CR_vars[["CR33"]]
      if(!all(is.na(cr33))) {
        case_when(
          as.numeric(cr33) == 1 ~ 1,  # Yes
          as.numeric(cr33) == 2 ~ 0,  # No
          as.numeric(cr33) == 3 ~ 0,  # I don't know → recoded to No
          TRUE ~ NA_real_
        )
      } else { rep(NA_real_, nrow(GYTS_2021_Zambia_National_)) }
    },

    # For backward compatibility, also create region variable
    region = RegionName,

    # Create derived variables for analysis
    age_group = factor(case_when(
      ageyrs <= 14 ~ "13-14",
      ageyrs %in% 15:16 ~ "15-16",
      ageyrs >= 17 ~ "17+",
      TRUE ~ NA_character_
    ), levels = c("13-14", "15-16", "17+")),
    gender = factor(case_when(
      sex1 == 1 ~ "Male",
      sex1 == 2 ~ "Female",
      TRUE ~ NA_character_
    ), levels = c("Female", "Male")),
    grade_level = factor(case_when(
      grade == 1 ~ "Grade 9",
      grade == 2 ~ "Grade 8",
      grade == 3 ~ "Grade 7",
      TRUE ~ NA_character_
    ), levels = c("Grade 9", "Grade 8", "Grade 7")),
    
    # Human-readable factor versions for new variables
    # tried_smoking_f: 1 = Yes (tried smoking), 0 = No (never tried)
    tried_smoking_f = factor(tried_smoking, levels = c(0, 1), labels = c("No", "Yes")),
    
    # parent_smoking_status_f: 0 = No, 1 = Yes (based on OR15 mapping)
    parent_smoking_status_f = factor(
      parent_smoking_status,
      levels = c(0, 1),
      labels = c("No", "Yes")
    ),
    
    # teacher_smoking_school_f: 0 = No, 1 = Yes (based on OR60 mapping)
    teacher_smoking_school_f = factor(
      teacher_smoking_school,
      levels = c(0, 1),
      labels = c("No", "Yes")
    ),
    
    # smoking_5years_from_now_f: Binary (Yes: Definitely yes/Probably yes, No: Definitely not/Probably not)
    smoking_5years_from_now_f = factor(
      smoking_5years_from_now,
      levels = c(0, 1),
      labels = c("No", "Yes")
    ),
    
    antismoking_media_f = factor(antismoking_media, levels = c(0, 1), labels = c("No", "Yes")),
    
    # taught_dangers_smoking_f: "Don't know" is recoded to "No"
    taught_dangers_smoking_f = factor(
      taught_dangers_smoking,
      levels = c(0, 1),
      labels = c("No", "Yes")
    ),
    
    # havingfriendsthatsmoke_f: Factor version
    havingfriendsthatsmoke_f = factor(
      havingfriendsthatsmoke,
      levels = c(0, 1),
      labels = c("No", "Yes")
    ),
    
    # Additional predictor variables from paper 8
    
    # Wealth index proxy from ZAR4: Convert to 3 groups
    zar4_raw = {
      zar4 <- ZAR_vars[["ZAR4"]]
      if(!all(is.na(zar4))) {
        as.numeric(zar4)
      } else {
        rep(NA_real_, nrow(GYTS_2021_Zambia_National_))
      }
    },
    
    # Use of handrolled cigarette: OR5=1 as No (0), rest as Yes (1)
    handrolled_cigarette = {
      or5 <- OR_vars[["OR5"]]
      if(!all(is.na(or5))) {
        case_when(
          as.numeric(or5) == 1 ~ 0,  # No
          TRUE ~ 1  # Yes for all other responses
        )
      } else { rep(NA_real_, nrow(GYTS_2021_Zambia_National_)) }
    },
    
    # Smoked cigarettes: No if CR8==1, Yes if CR8 > 1
    smoked_cigarettes = {
      cr8 <- CR_vars[["CR8"]]
      if(!all(is.na(cr8))) {
        case_when(
          as.numeric(cr8) == 1 ~ 0,  # No
          as.numeric(cr8) > 1 ~ 1,  # Yes
          TRUE ~ NA_real_
        )
      } else { rep(NA_real_, nrow(GYTS_2021_Zambia_National_)) }
    },
    
    # Tried shisha: No/Yes from SR1
    tried_shisha = {
      sr1 <- SR_vars[["SR1"]]
      if(!all(is.na(sr1))) {
        case_when(
          as.numeric(sr1) == 1 ~ 0,  # No
          as.numeric(sr1) == 2 ~ 1,  # Yes
          TRUE ~ NA_real_
        )
      } else { rep(NA_real_, nrow(GYTS_2021_Zambia_National_)) }
    },
    
    # Heard of electronic cigarette: ELR1: 2=No, 1=Yes
    heard_electronic_cigarette = {
      elr1 <- ELR_vars[["ELR1"]]
      if(!all(is.na(elr1))) {
        case_when(
          as.numeric(elr1) == 2 ~ 0,  # No
          as.numeric(elr1) == 1 ~ 1,  # Yes
          TRUE ~ NA_real_
        )
      } else { rep(NA_real_, nrow(GYTS_2021_Zambia_National_)) }
    },
    
    # Father smokes: No if OR15=1 or 4, Yes if OR15=2 or 3
    father_smokes = {
      or15 <- OR_vars[["OR15"]]
      if(!all(is.na(or15))) {
        case_when(
          as.numeric(or15) == 1 ~ 0,  # No
          as.numeric(or15) == 2 ~ 1,  # Yes
          as.numeric(or15) == 3 ~ 1,  # Yes
          as.numeric(or15) == 4 ~ 0,  # No
          TRUE ~ NA_real_
        )
      } else { rep(NA_real_, nrow(GYTS_2021_Zambia_National_)) }
    },
    
    # People smoke in school: Yes if CR22=1, No otherwise
    people_smoke_school = {
      cr22 <- CR_vars[["CR22"]]
      if(!all(is.na(cr22))) {
        case_when(
          as.numeric(cr22) == 1 ~ 1,  # Yes
          TRUE ~ 0  # No for all other responses
        )
      } else { rep(NA_real_, nrow(GYTS_2021_Zambia_National_)) }
    },
    
    # Teachers smoke at school: No if OR60 > 2, Yes otherwise
    teachers_smoke_school = {
      or60 <- OR_vars[["OR60"]]
      if(!all(is.na(or60))) {
        case_when(
          as.numeric(or60) > 2 ~ 0,  # No
          TRUE ~ 1  # Yes for OR60 <= 2
        )
      } else { rep(NA_real_, nrow(GYTS_2021_Zambia_National_)) }
    },
    
    # See people using tobacco on TV: CR34=2 is Yes, rest is No
    see_tobacco_tv = {
      cr34 <- CR_vars[["CR34"]]
      if(!all(is.na(cr34))) {
        case_when(
          as.numeric(cr34) == 2 ~ 1,  # Yes
          TRUE ~ 0  # No for all other responses
        )
      } else { rep(NA_real_, nrow(GYTS_2021_Zambia_National_)) }
    },
    
    # See tobacco advert: CR35=2 is Yes, rest is No
    see_tobacco_advert = {
      cr35 <- CR_vars[["CR35"]]
      if(!all(is.na(cr35))) {
        case_when(
          as.numeric(cr35) == 2 ~ 1,  # Yes
          TRUE ~ 0  # No for all other responses
        )
      } else { rep(NA_real_, nrow(GYTS_2021_Zambia_National_)) }
    },
    
    # Long tobacco use lead to cancer: ZAR64=1 as Yes, rest No
    tobacco_causes_cancer = {
      zar64 <- ZAR_vars[["ZAR64"]]
      if(!all(is.na(zar64))) {
        case_when(
          as.numeric(zar64) == 1 ~ 1,  # Yes
          TRUE ~ 0  # No for all other responses
        )
      } else { rep(NA_real_, nrow(GYTS_2021_Zambia_National_)) }
    },
    
    # Long term tobacco use leads to drug use: ZAR65=1 as Yes, rest No
    tobacco_leads_drug_use = {
      zar65 <- ZAR_vars[["ZAR65"]]
      if(!all(is.na(zar65))) {
        case_when(
          as.numeric(zar65) == 1 ~ 1,  # Yes
          TRUE ~ 0  # No for all other responses
        )
      } else { rep(NA_real_, nrow(GYTS_2021_Zambia_National_)) }
    },
    
    # Own item with tobacco product: ZAR53=1 as Yes, otherwise No
    own_item_tobacco = {
      zar53 <- ZAR_vars[["ZAR53"]]
      if(!all(is.na(zar53))) {
        case_when(
          as.numeric(zar53) == 1 ~ 1,  # Yes
          TRUE ~ 0  # No for all other responses
        )
      } else { rep(NA_real_, nrow(GYTS_2021_Zambia_National_)) }
    },
    
    # Outcome variable: shisha use (SR1=1)
    shisha_use = {
      sr1 <- SR_vars[["SR1"]]
      if(!all(is.na(sr1))) {
        case_when(
          as.numeric(sr1) == 1 ~ 1,  # Yes (ever used shisha)
          as.numeric(sr1) == 2 ~ 0,  # No (never used shisha)
          TRUE ~ NA_real_
        )
      } else {
        # Fallback to CR8 if SR1 is not available
      cr8 <- CR_vars[["CR8"]]
      if(!all(is.na(cr8)) && any(as.numeric(cr8) == 1, na.rm = TRUE)) {
        case_when(as.numeric(cr8) == 1 ~ 1, TRUE ~ 0)
      } else {
        rep(0, nrow(GYTS_2021_Zambia_National_))
        }
      }
    },
    
    # For backward compatibility
    ever_shisha = shisha_use,
    
    # Variables from CR1, CR2, and ZAR3 for a priori variables
    # Age from CR1: How old are you?
    age_cr1 = {
      cr1 <- CR_vars[["CR1"]]
      if(!all(is.na(cr1))) {
        case_when(
          as.numeric(cr1) == 1 ~ 11,  # 11 years old or younger
          as.numeric(cr1) == 2 ~ 12,  # 12 years old
          as.numeric(cr1) == 3 ~ 13,  # 13 years old
          as.numeric(cr1) == 4 ~ 14,  # 14 years old
          as.numeric(cr1) == 5 ~ 15,  # 15 years old
          as.numeric(cr1) == 6 ~ 16,  # 16 years old
          as.numeric(cr1) == 7 ~ 17,  # 17 years old or older
          TRUE ~ NA_real_
        )
      } else { rep(NA_real_, nrow(GYTS_2021_Zambia_National_)) }
    },
    
    # Sex from CR2: What is your sex?
    sex_cr2 = {
      cr2 <- CR_vars[["CR2"]]
      if(!all(is.na(cr2))) {
        case_when(
          as.numeric(cr2) == 1 ~ "Male",
          as.numeric(cr2) == 2 ~ "Female",
          TRUE ~ NA_character_
        )
      } else { rep(NA_character_, nrow(GYTS_2021_Zambia_National_)) }
    },
    
    # Grade from ZAR3: In what grade/form are you?
    grade_zar3 = {
      zar3 <- ZAR_vars[["ZAR3"]]
      if(!all(is.na(zar3))) {
        case_when(
          as.numeric(zar3) == 1 ~ "Grade 7",
          as.numeric(zar3) == 2 ~ "Grade 8",
          as.numeric(zar3) == 3 ~ "Grade 9",
          TRUE ~ NA_character_
        )
      } else { rep(NA_character_, nrow(GYTS_2021_Zambia_National_)) }
    },
    parental_smoking = case_when(parentssmokingstatus == 1 ~ "None", parentssmokingstatus %in% 2:4 ~ "At least one parent", TRUE ~ NA_character_),
    friends_smoke = case_when(friendsmokingcigarettes == 1 ~ "None", friendsmokingcigarettes == 2 ~ "Some", friendsmokingcigarettes %in% 3:4 ~ "Most/All", TRUE ~ NA_character_),
    ad_exposure = case_when(
      advertisingcigarettebrands <= 2 | cigarettesadvertsseenbillboards <= 2 | cigarettesadvertsseeninewspapers <= 2 ~ "High",
      advertisingcigarettebrands == 3 | cigarettesadvertsseenbillboards == 3 | cigarettesadvertsseeninewspapers == 3 ~ "Low",
      TRUE ~ "None"
    ),
    knows_harmful = {
      cr24 <- CR_vars[["CR24"]]
      if(!all(is.na(cr24))) {
        case_when(as.numeric(cr24) >= 3 ~ "Yes", TRUE ~ "No")
      } else {
        rep(NA_character_, nrow(GYTS_2021_Zambia_National_))
      }
    },
    positive_attitude = {
      cr21 <- CR_vars[["CR21"]]
      cr22 <- CR_vars[["CR22"]]
      if((!all(is.na(cr21)) && any(as.numeric(cr21) == 1, na.rm = TRUE)) ||
         (!all(is.na(cr22)) && any(as.numeric(cr22) == 1, na.rm = TRUE))) {
        case_when(as.numeric(cr21) == 1 | as.numeric(cr22) == 1 ~ "Yes", TRUE ~ "No")
      } else {
        rep(NA_character_, nrow(GYTS_2021_Zambia_National_))
      }
    }
  )

# Create factor versions for additional predictors from paper 8 (after all variables are created)
df <- df %>%
  mutate(
    handrolled_cigarette_f = factor(handrolled_cigarette, levels = c(0, 1), labels = c("No", "Yes")),
    smoked_cigarettes_f = factor(smoked_cigarettes, levels = c(0, 1), labels = c("No", "Yes")),
    tried_shisha_f = factor(tried_shisha, levels = c(0, 1), labels = c("No", "Yes")),
    heard_electronic_cigarette_f = factor(heard_electronic_cigarette, levels = c(0, 1), labels = c("No", "Yes")),
    father_smokes_f = factor(father_smokes, levels = c(0, 1), labels = c("No", "Yes")),
    people_smoke_school_f = factor(people_smoke_school, levels = c(0, 1), labels = c("No", "Yes")),
    teachers_smoke_school_f = factor(teachers_smoke_school, levels = c(0, 1), labels = c("No", "Yes")),
    see_tobacco_tv_f = factor(see_tobacco_tv, levels = c(0, 1), labels = c("No", "Yes")),
    see_tobacco_advert_f = factor(see_tobacco_advert, levels = c(0, 1), labels = c("No", "Yes")),
    tobacco_causes_cancer_f = factor(tobacco_causes_cancer, levels = c(0, 1), labels = c("No", "Yes")),
    tobacco_leads_drug_use_f = factor(tobacco_leads_drug_use, levels = c(0, 1), labels = c("No", "Yes")),
    own_item_tobacco_f = factor(own_item_tobacco, levels = c(0, 1), labels = c("No", "Yes"))
  )

# Create wealth index groups from ZAR4 (three groups: low, middle, rich)
# Convert ZAR4 into 3 groups based on ranks
df <- df %>%
  mutate(
    # Create three groups based on ZAR4 ranks: low, middle, rich
    wealth_index = case_when(
      !is.na(zar4_raw) ~ {
        # Calculate tertiles (33.3%, 33.3%, 33.4%)
        tertiles <- quantile(zar4_raw, probs = c(0, 1/3, 2/3, 1), na.rm = TRUE)
        case_when(
          zar4_raw <= tertiles[2] ~ "Low",
          zar4_raw <= tertiles[3] ~ "Middle",
          TRUE ~ "Rich"
        )
      },
      TRUE ~ NA_character_
    ),
    # Factor version for analysis
    wealth_index_f = factor(
      wealth_index,
      levels = c("Low", "Middle", "Rich"),
      labels = c("Low", "Middle", "Rich")
    )
  )

# Create survey design object with Stratum and PSU
# Check for Stratum, PSU, and weight variables
stratum_var <- if("Stratum" %in% names(df)) "Stratum" else NULL
psu_var <- if("PSU" %in% names(df)) "PSU" else NULL
weight_var <- if("FinalWgt" %in% names(df)) "FinalWgt" else if("'FinalWgt" %in% names(df)) "'FinalWgt" else if("WEIGHT" %in% names(df)) "WEIGHT" else NULL

if(!is.null(stratum_var) && !is.null(psu_var) && !is.null(weight_var)) {
  # Two-stage design with Stratum and PSU
  # nest = TRUE assumes clusters are nested within strata
  svy_design <- svydesign(
    ids = as.formula(paste0("~", psu_var)),
    strata = as.formula(paste0("~", stratum_var)),
    weights = as.formula(paste0("~", weight_var)),
    data = df,
    nest = TRUE
  )
} else if(!is.null(psu_var) && !is.null(weight_var)) {
  # Design with PSU only (no stratification)
  svy_design <- svydesign(
    ids = as.formula(paste0("~", psu_var)),
    weights = as.formula(paste0("~", weight_var)),
    data = df
  )
} else if(!is.null(weight_var)) {
  # Simple design with weights only
  svy_design <- svydesign(
    ids = ~1,
    weights = as.formula(paste0("~", weight_var)),
    data = df
  )
} else {
  svy_design <- svydesign(ids = ~1, weights = ~1, data = df)
  warning("Stratum, PSU, or weight variable not found. Using unweighted design.")
}
```

# Introduction

Shisha (also known as waterpipe, hookah, or narghile) smoking has emerged as a significant public health concern globally, particularly among adolescents and young adults [@who_africa_tobacco_2021]. While cigarette smoking has been the primary focus of tobacco control efforts, shisha use represents a growing and often overlooked form of tobacco consumption that poses unique challenges for prevention and control [@who_global_2020]. Shisha smoking involves heating tobacco, which is often flavored, and the smoke passes through water before being inhaled through a hose or tube. Despite common misconceptions that water filtration makes shisha less harmful, evidence indicates that shisha smoking exposes users to similar or even higher levels of toxicants compared to cigarette smoking [@who_tobacco_control_2021].

The popularity of shisha among adolescents is particularly concerning given the social and cultural contexts in which it is often consumed. Shisha cafes and lounges have become popular social venues in many countries, creating environments where shisha use is normalized and easily accessible to young people [@drope_tobacco_2018]. The flavored tobacco used in shisha, combined with the social nature of consumption, may make it particularly appealing to adolescents who may perceive it as less harmful or more socially acceptable than cigarette smoking [@who_africa_2019].

In Zambia, as in many African countries, data on shisha use among adolescents remain limited, creating gaps in understanding the prevalence, patterns, and determinants of shisha use among school-going youth [@zyambo_gyts_2021]. The 2021 Global Youth Tobacco Survey (GYTS) provides an opportunity to examine shisha use and its associated factors among Zambian adolescents, offering insights that can inform targeted prevention and control strategies. Understanding the specific factors that drive shisha use initiation and continuation among Zambian youth is essential for developing evidence-based interventions that address this emerging public health challenge [@who_gyts_2021].

This analysis aims to examine the prevalence of shisha use among adolescents in Zambia and identify key factors associated with shisha use. Specifically, we seek to: (1) describe the prevalence of shisha use across different demographic and socioeconomic groups; (2) identify individual, social, and environmental factors associated with shisha use; and (3) assess the relationship between tobacco-growing regions and shisha use patterns among adolescents. The findings from this analysis have important policy implications for strengthening tobacco control efforts in Zambia, particularly in addressing emerging tobacco products and developing youth-focused prevention programs that account for the unique characteristics of shisha use [@moh_zambia_2018].

# Methods

## Data Source
- **Survey**: Global Youth Tobacco Survey (GYTS) 2021, Zambia
- **Analysis Dataset**: GYTS 2021 Zambia (National).XLS, sheet "ZAMBIA_DATA_21"

## Data Management

All variables used in this analysis were derived from the GYTS 2021 dataset. The outcome variable for this analysis is shisha use, defined as ever having used shisha (SR1=1). All predictor variables were created following the same methodology as described in Paper 1, ensuring consistency across analyses.

### Variables Included in Analysis

The following table provides definitions and value labels for all variables included in this analysis:

```{r variables-table, echo=FALSE, results='asis'}
# Create comprehensive variables table organized by category
# Order: Outcome -> Demographic -> Behavioral -> Social/Environmental -> Media/Exposure -> Knowledge/Attitude -> Intention -> Policy

variables_data <- data.frame(
  Category = c(
    # Outcome
    "Outcome",
    # Demographic
    "Demographic", "Demographic", "Demographic", "Demographic", "Demographic",
    # Behavioral/Experiential
    "Behavioral/Experiential", "Behavioral/Experiential", "Behavioral/Experiential", 
    "Behavioral/Experiential", "Behavioral/Experiential", "Behavioral/Experiential",
    # Social/Environmental
    "Social/Environmental", "Social/Environmental", "Social/Environmental", 
    "Social/Environmental", "Social/Environmental",
    # Media/Exposure
    "Media/Exposure", "Media/Exposure", "Media/Exposure",
    # Knowledge/Attitude
    "Knowledge/Attitude", "Knowledge/Attitude",
    # Intention
    "Intention",
    # Policy/Education
    "Policy/Education"
  ),
  Variable = c(
    # Outcome
    "shisha_use",
    # Demographic
    "age_cr1",
    "sex_cr2",
    "grade_zar3",
    "RegionName",
    "wealth_index_f",
    # Behavioral/Experiential
    "tried_smoking_f",
    "smoked_cigarettes_f",
    "handrolled_cigarette_f",
    "tried_shisha_f",
    "heard_electronic_cigarette_f",
    "own_item_tobacco_f",
    # Social/Environmental
    "father_smokes_f",
    "parent_smoking_status_f",
    "havingfriendsthatsmoke_f",
    "people_smoke_school_f",
    "teachers_smoke_school_f",
    # Media/Exposure
    "see_tobacco_tv_f",
    "see_tobacco_advert_f",
    "antismoking_media_f",
    # Knowledge/Attitude
    "tobacco_causes_cancer_f",
    "tobacco_leads_drug_use_f",
    # Intention
    "smoking_5years_from_now_f",
    # Policy/Education
    "taught_dangers_smoking_f"
  ),
  Definition = c(
    # Outcome
    "Ever used shisha/waterpipe (outcome variable)",
    # Demographic
    "Age in years (factor: 11-17 years)",
    "Sex of participant",
    "Grade level in school",
    "Geographic region",
    "Wealth index (three groups: low, middle, rich)",
    # Behavioral/Experiential
    "Ever tried cigarette smoking",
    "Smoked cigarettes",
    "Use of handrolled cigarette",
    "Tried shisha",
    "Heard of electronic cigarette",
    "Own item with tobacco product",
    # Social/Environmental
    "Father smokes",
    "Parent or guardian smokes in home",
    "Friends smoke tobacco",
    "People smoke in school",
    "Teachers smoke at school",
    # Media/Exposure
    "See people using tobacco on TV",
    "See tobacco advertisement",
    "Exposure to anti-tobacco media messages",
    # Knowledge/Attitude
    "Long tobacco use leads to cancer",
    "Long term tobacco use leads to drug use",
    # Intention
    "Intention to use tobacco in next 5 years",
    # Policy/Education
    "Taught about dangers of tobacco use in school"
  ),
  Source_Question = c(
    # Outcome
    "SR1: Have you ever smoked shisha/waterpipe? (1=Yes, 2=No)",
    # Demographic
    "CR1: How old are you? (1=11 years or younger, 2=12 years, 3=13 years, 4=14 years, 5=15 years, 6=16 years, 7=17 years or older)",
    "CR2: What is your sex? (1=Male, 2=Female)",
    "ZAR3: In what grade/form are you? (1=Grade 7, 2=Grade 8, 3=Grade 9)",
    "Derived from Stratum variable (categorized into Lusaka, ROC, Tobacco Regions)",
    "ZAR4: During an average week, how much money do you have that you can spend on yourself, however you want? (Converted to 3 groups: low, middle, rich based on ranks)",
    # Behavioral/Experiential
    "CR5: Have you ever tried or experimented with cigarette smoking, even one or two puffs? (1=No, 2=Yes)",
    "CR8: During the past 30 days, on how many days did you smoke cigarettes? (1=No, >1=Yes)",
    "OR5: During the past 30 days, did you smoke handrolled cigarettes? (1=No, other=Yes)",
    "SR1: Have you ever tried or experimented with shisha/waterpipe? (1=No, 2=Yes)",
    "ELR1: Have you ever heard of electronic cigarettes? (1=Yes, 2=No)",
    "ZAR53: Do you own something (for example, t-shirt, pen, backpack) with a tobacco product brand logo on it? (1=Yes, other=No)",
    # Social/Environmental
    "OR15: How often do you see your father (stepfather or mother's partner) smoking in your home? (1 or 4=No, 2 or 3=Yes)",
    "OR15: How often do you see your father (stepfather or mother's partner) smoking in your home? (1 or 4=No, 2 or 3=Yes)",
    "ZAR55: Do any of your closest friends smoke tobacco? (1=No, 2-4=Yes, 5=No/I don't know)",
    "CR22: During the past 30 days, did you see anyone smoking inside the school building or on school property? (1=Yes, other=No)",
    "OR60: During school hours, how often do you see teachers smoking outdoors on school premises? (≤2=Yes, >2=No)",
    # Media/Exposure
    "CR34: During the past 30 days, did you see people using tobacco on TV, in videos, or in movies? (2=Yes, other=No)",
    "CR35: During the past 30 days, did you see any advertisements or promotions for tobacco products? (2=Yes, other=No)",
    "CR30/CR31: During the past 30 days, how often did you see or hear anti-tobacco media messages on TV, radio, internet, billboards, posters, newspapers, magazines, or movies? (High exposure=Yes, Low/No=No)",
    # Knowledge/Attitude
    "ZAR64: Do you think that long-term tobacco use leads to cancer? (1=Yes, other=No)",
    "ZAR65: Do you think that long-term tobacco use leads to drug use? (1=Yes, other=No)",
    # Intention
    "CR40: At any time during the next 12 months do you think you will use any form of tobacco? (1-2=No, 3-4=Yes)",
    # Policy/Education
    "CR33: During the past 12 months, were you taught in any of your classes about the dangers of tobacco use? (1=Yes, 2-3=No)"
  ),
  Value_Labels = c(
    # Outcome
    "0 = No (never used), 1 = Yes (ever used)",
    # Demographic
    "11, 12, 13, 14, 15, 16, 17 (years) - Factor variable derived from CR1",
    "Male (CR2=1), Female (CR2=2)",
    "Grade 7 (ZAR3=1), Grade 8 (ZAR3=2), Grade 9 (ZAR3=3)",
    "Lusaka, ROC (Rest of Country), Tobacco Regions - Derived from Stratum",
    "Low (bottom tertile), Middle (middle tertile), Rich (top tertile) - Based on ZAR4 ranks",
    # Behavioral/Experiential
    "No (CR5=1), Yes (CR5=2) - Ever tried cigarette smoking",
    "No (CR8=1), Yes (CR8>1) - Smoked cigarettes in past 30 days",
    "No (OR5=1), Yes (OR5≠1) - Use of handrolled cigarettes",
    "No (SR1=1), Yes (SR1=2) - Ever tried shisha/waterpipe",
    "No (ELR1=2), Yes (ELR1=1) - Heard of electronic cigarettes",
    "No (ZAR53≠1), Yes (ZAR53=1) - Own item with tobacco product brand logo",
    # Social/Environmental
    "No (OR15=1 or 4), Yes (OR15=2 or 3) - Father smokes in home",
    "No (OR15=1 or 4), Yes (OR15=2 or 3) - Parent/guardian smokes in home",
    "No (ZAR55=1 or 5), Yes (ZAR55=2-4) - Friends smoke tobacco",
    "No (CR22≠1), Yes (CR22=1) - People smoke in school",
    "No (OR60>2), Yes (OR60≤2) - Teachers smoke at school",
    # Media/Exposure
    "No (CR34≠2), Yes (CR34=2) - See people using tobacco on TV/videos/movies",
    "No (CR35≠2), Yes (CR35=2) - See tobacco advertisements or promotions",
    "No (Low/No exposure), Yes (High exposure) - Anti-tobacco media messages (CR30/CR31)",
    # Knowledge/Attitude
    "No (ZAR64≠1), Yes (ZAR64=1) - Long-term tobacco use leads to cancer",
    "No (ZAR65≠1), Yes (ZAR65=1) - Long-term tobacco use leads to drug use",
    # Intention
    "No (CR40=1-2), Yes (CR40=3-4) - Intention to use tobacco in next 12 months",
    # Policy/Education
    "No (CR33=2-3), Yes (CR33=1) - Taught about dangers of tobacco use in school"
  ),
  Variable_Type = c(
    # Outcome
    "Outcome (binary)",
    # Demographic
    "A priori (categorical)",
    "A priori (categorical)",
    "A priori (categorical)",
    "A priori (categorical)",
    "Predictor (categorical)",
    # Behavioral/Experiential
    "Predictor (binary)",
    "Predictor (binary)",
    "Predictor (binary)",
    "Predictor (binary)",
    "Predictor (binary)",
    "Predictor (binary)",
    # Social/Environmental
    "Predictor (binary)",
    "Predictor (binary)",
    "Predictor (binary)",
    "Predictor (binary)",
    "Predictor (binary)",
    # Media/Exposure
    "Predictor (binary)",
    "Predictor (binary)",
    "Predictor (binary)",
    # Knowledge/Attitude
    "Predictor (binary)",
    "Predictor (binary)",
    # Intention
    "Predictor (binary)",
    # Policy/Education
    "Predictor (binary)"
  ),
  stringsAsFactors = FALSE
)

# Create formatted table using flextable with category grouping
variables_table <- variables_data %>%
  flextable() %>%
  set_header_labels(
    Category = "**Category**",
    Variable = "**Variable Name**",
    Definition = "**Definition**",
    Source_Question = "**Source Question**",
    Value_Labels = "**Value Labels**",
    Variable_Type = "**Type**"
  ) %>%
  merge_v(j = "Category") %>%
  bold(part = "header") %>%
  bold(j = 1, part = "body") %>%
  align(align = "left", part = "all") %>%
  width(j = "Category", width = 1.0) %>%
  width(j = "Variable", width = 1.2) %>%
  width(j = "Definition", width = 1.8) %>%
  width(j = "Source_Question", width = 2.3) %>%
  width(j = "Value_Labels", width = 1.9) %>%
  width(j = "Variable_Type", width = 0.9) %>%
  theme_booktabs() %>%
  fontsize(size = 9, part = "all") %>%
  padding(padding = 4, part = "all") %>%
  add_footer_lines("Variables derived from GYTS 2021 Zambia National dataset. Outcome variable: shisha_use (SR1=1). A priori variables (age_cr1, sex_cr2, grade_zar3, RegionName) were included in all adjusted models regardless of statistical significance.") %>%
  fontsize(size = 8, part = "footer") %>%
  italic(part = "footer") %>%
  align(align = "left", part = "footer")

variables_table
```

### Variable Creation and Recoding

**Outcome Variable:**
- `shisha_use`: Created from SR1 (Have you ever smoked shisha/waterpipe?). Responses were coded as: 1 = Yes (ever used), 2 = No (never used). For analysis, this was recoded to binary: 1 = Yes (ever used), 0 = No (never used).

**A Priori Variables:**
- `age_cr1`: Derived from CR1 (How old are you?) with responses 1-7 mapped to ages 11-17 years. Used as a factor variable in models.
- `sex_cr2`: Derived from CR2 (What is your sex?) with responses: 1 = Male, 2 = Female.
- `grade_zar3`: Derived from ZAR3 (In what grade/form are you?) with responses: 1 = Grade 7, 2 = Grade 8, 3 = Grade 9.
- `RegionName`: Created from the Stratum variable, categorized into three regions: Lusaka, ROC (Rest of Country), and Tobacco Regions.

**Predictor Variables:**
All predictor variables were recoded as binary factors (No/Yes) for consistency in analysis:
- `tried_smoking_f`: Ever tried cigarette smoking (from CR8)
- `parent_smoking_status_f`: Parent/guardian smokes in home (from OR15)
- `smoking_5years_from_now_f`: Intention to use tobacco in next 5 years (from CR21/CR22)
- `antismoking_media_f`: Exposure to anti-tobacco media messages (from CR24)
- `taught_dangers_smoking_f`: Taught about dangers of tobacco use (from CR25)
- `teacher_smoking_school_f`: Saw teachers smoking at school (from OR60)
- `havingfriendsthatsmoke_f`: Friends smoke tobacco (from CR23)

**Missing Data:**
Observations with missing values for the outcome variable (shisha_use) were excluded from analysis. For predictor variables, missing values were handled through listwise deletion in regression models. All analyses accounted for the complex survey design using survey weights, stratification, and clustering.

## Statistical Analysis
- Weighted prevalence estimates with 95% confidence intervals
- Crude (unadjusted) logistic regression models for each variable
- Adjusted logistic regression models using stepwise selection (AIC-based)
- Age (age_cr1), sex (sex_cr2), grade (grade_zar3), and region included a priori in all adjusted models
- Survey-weighted analysis accounting for complex survey design (Stratum and PSU)
- Sensitivity analyses to assess robustness of findings

# Results

## Table 1: Sample Characteristics

```{r table1-characteristics, echo=FALSE, results='asis'}
# Create nice characteristics table using gtsummary
# Prepare data for characteristics table
df_char <- df %>%
  filter(!is.na(RegionName)) %>%
  mutate(
    RegionName = factor(RegionName, levels = c("Lusaka", "ROC", "Tobacco Regions")),
    age_group = factor(age_group, levels = c("13-14", "15-16", "17+")),
    gender = factor(gender, levels = c("Female", "Male")),
    grade_level = factor(grade_level, levels = c("Grade 9", "Grade 8", "Grade 7")),
    shisha_use = factor(shisha_use, levels = c(0, 1), labels = c("No", "Yes")),
    # Variables from CR1, CR2, ZAR3
    age_cr1 = age_cr1,
    sex_cr2 = factor(sex_cr2, levels = c("Female", "Male")),
    grade_zar3 = factor(grade_zar3, levels = c("Grade 7", "Grade 8", "Grade 9")),
    # Ensure new variables are factors with human-readable labels
    wealth_index_f = factor(wealth_index_f, levels = c("Low", "Middle", "Rich")),
    handrolled_cigarette_f = factor(handrolled_cigarette_f, levels = c("No", "Yes")),
    smoked_cigarettes_f = factor(smoked_cigarettes_f, levels = c("No", "Yes")),
    tried_shisha_f = factor(tried_shisha_f, levels = c("No", "Yes")),
    heard_electronic_cigarette_f = factor(heard_electronic_cigarette_f, levels = c("No", "Yes")),
    father_smokes_f = factor(father_smokes_f, levels = c("No", "Yes")),
    people_smoke_school_f = factor(people_smoke_school_f, levels = c("No", "Yes")),
    teachers_smoke_school_f = factor(teachers_smoke_school_f, levels = c("No", "Yes")),
    see_tobacco_tv_f = factor(see_tobacco_tv_f, levels = c("No", "Yes")),
    see_tobacco_advert_f = factor(see_tobacco_advert_f, levels = c("No", "Yes")),
    tobacco_causes_cancer_f = factor(tobacco_causes_cancer_f, levels = c("No", "Yes")),
    tobacco_leads_drug_use_f = factor(tobacco_leads_drug_use_f, levels = c("No", "Yes")),
    own_item_tobacco_f = factor(own_item_tobacco_f, levels = c("No", "Yes")),
    tried_smoking_f = factor(tried_smoking_f, levels = c("No", "Yes"), labels = c("No", "Yes")),
    parent_smoking_status_f = factor(parent_smoking_status_f, levels = c("No", "Yes"), labels = c("No", "Yes")),
    smoking_5years_from_now_f = factor(smoking_5years_from_now_f, levels = c("No", "Yes"), labels = c("No", "Yes")),
    antismoking_media_f = factor(antismoking_media_f, levels = c("No", "Yes"), labels = c("No", "Yes")),
    taught_dangers_smoking_f = factor(taught_dangers_smoking_f, levels = c("No", "Yes"), labels = c("No", "Yes")),
    teacher_smoking_school_f = factor(teacher_smoking_school_f, levels = c("No", "Yes"), labels = c("No", "Yes")),
    havingfriendsthatsmoke_f = factor(havingfriendsthatsmoke_f, levels = c("No", "Yes"), labels = c("No", "Yes"))
  )

# Check if we have data and sufficient RegionName levels
if(nrow(df_char) == 0) {
  cat("No data available for Table 1 after filtering.\n")
} else if(length(unique(df_char$RegionName[!is.na(df_char$RegionName)])) < 2) {
  cat("Insufficient variation in RegionName variable for Table 1.\n")
} else {
  # Select only variables that exist and have data
  vars_to_include <- c()
  if("age_cr1" %in% names(df_char) && sum(!is.na(df_char$age_cr1)) > 0) vars_to_include <- c(vars_to_include, "age_cr1")
  if("sex_cr2" %in% names(df_char) && sum(!is.na(df_char$sex_cr2)) > 0) vars_to_include <- c(vars_to_include, "sex_cr2")
  if("grade_zar3" %in% names(df_char) && sum(!is.na(df_char$grade_zar3)) > 0) vars_to_include <- c(vars_to_include, "grade_zar3")
  if("age_group" %in% names(df_char) && sum(!is.na(df_char$age_group)) > 0) vars_to_include <- c(vars_to_include, "age_group")
  if("gender" %in% names(df_char) && sum(!is.na(df_char$gender)) > 0) vars_to_include <- c(vars_to_include, "gender")
  if("grade_level" %in% names(df_char) && sum(!is.na(df_char$grade_level)) > 0) vars_to_include <- c(vars_to_include, "grade_level")
  if("wealth_index_f" %in% names(df_char) && sum(!is.na(df_char$wealth_index_f)) > 0) vars_to_include <- c(vars_to_include, "wealth_index_f")
  if("handrolled_cigarette_f" %in% names(df_char) && sum(!is.na(df_char$handrolled_cigarette_f)) > 0) vars_to_include <- c(vars_to_include, "handrolled_cigarette_f")
  if("smoked_cigarettes_f" %in% names(df_char) && sum(!is.na(df_char$smoked_cigarettes_f)) > 0) vars_to_include <- c(vars_to_include, "smoked_cigarettes_f")
  if("tried_shisha_f" %in% names(df_char) && sum(!is.na(df_char$tried_shisha_f)) > 0) vars_to_include <- c(vars_to_include, "tried_shisha_f")
  if("heard_electronic_cigarette_f" %in% names(df_char) && sum(!is.na(df_char$heard_electronic_cigarette_f)) > 0) vars_to_include <- c(vars_to_include, "heard_electronic_cigarette_f")
  if("father_smokes_f" %in% names(df_char) && sum(!is.na(df_char$father_smokes_f)) > 0) vars_to_include <- c(vars_to_include, "father_smokes_f")
  if("people_smoke_school_f" %in% names(df_char) && sum(!is.na(df_char$people_smoke_school_f)) > 0) vars_to_include <- c(vars_to_include, "people_smoke_school_f")
  if("teachers_smoke_school_f" %in% names(df_char) && sum(!is.na(df_char$teachers_smoke_school_f)) > 0) vars_to_include <- c(vars_to_include, "teachers_smoke_school_f")
  if("see_tobacco_tv_f" %in% names(df_char) && sum(!is.na(df_char$see_tobacco_tv_f)) > 0) vars_to_include <- c(vars_to_include, "see_tobacco_tv_f")
  if("see_tobacco_advert_f" %in% names(df_char) && sum(!is.na(df_char$see_tobacco_advert_f)) > 0) vars_to_include <- c(vars_to_include, "see_tobacco_advert_f")
  if("tobacco_causes_cancer_f" %in% names(df_char) && sum(!is.na(df_char$tobacco_causes_cancer_f)) > 0) vars_to_include <- c(vars_to_include, "tobacco_causes_cancer_f")
  if("tobacco_leads_drug_use_f" %in% names(df_char) && sum(!is.na(df_char$tobacco_leads_drug_use_f)) > 0) vars_to_include <- c(vars_to_include, "tobacco_leads_drug_use_f")
  if("own_item_tobacco_f" %in% names(df_char) && sum(!is.na(df_char$own_item_tobacco_f)) > 0) vars_to_include <- c(vars_to_include, "own_item_tobacco_f")
  if("tried_smoking_f" %in% names(df_char) && sum(!is.na(df_char$tried_smoking_f)) > 0) vars_to_include <- c(vars_to_include, "tried_smoking_f")
  if("parent_smoking_status_f" %in% names(df_char) && sum(!is.na(df_char$parent_smoking_status_f)) > 0) vars_to_include <- c(vars_to_include, "parent_smoking_status_f")
  if("smoking_5years_from_now_f" %in% names(df_char) && sum(!is.na(df_char$smoking_5years_from_now_f)) > 0) vars_to_include <- c(vars_to_include, "smoking_5years_from_now_f")
  if("antismoking_media_f" %in% names(df_char) && sum(!is.na(df_char$antismoking_media_f)) > 0) vars_to_include <- c(vars_to_include, "antismoking_media_f")
  if("taught_dangers_smoking_f" %in% names(df_char) && sum(!is.na(df_char$taught_dangers_smoking_f)) > 0) vars_to_include <- c(vars_to_include, "taught_dangers_smoking_f")
  if("teacher_smoking_school_f" %in% names(df_char) && sum(!is.na(df_char$teacher_smoking_school_f)) > 0) vars_to_include <- c(vars_to_include, "teacher_smoking_school_f")
  if("havingfriendsthatsmoke_f" %in% names(df_char) && sum(!is.na(df_char$havingfriendsthatsmoke_f)) > 0) vars_to_include <- c(vars_to_include, "havingfriendsthatsmoke_f")
  
  if(length(vars_to_include) == 0) {
    cat("No valid variables available for Table 1.\n")
  } else {
    tryCatch({
      df_table1 <- df_char %>% dplyr::select(dplyr::all_of(c(vars_to_include, "RegionName")))
      
      all_labels <- list(
        age_cr1 = "Age, years",
        age_group = "Age group",
        sex_cr2 = "Sex",
        gender = "Sex",
        grade_zar3 = "Grade",
        grade_level = "Grade level",
        wealth_index_f = "Wealth index",
        handrolled_cigarette_f = "Use of handrolled cigarette",
        smoked_cigarettes_f = "Smoked cigarettes",
        tried_shisha_f = "Tried shisha",
        heard_electronic_cigarette_f = "Heard of electronic cigarette",
        father_smokes_f = "Father smokes",
        people_smoke_school_f = "People smoke in school",
        teachers_smoke_school_f = "Teachers smoke at school",
        see_tobacco_tv_f = "See people using tobacco on TV",
        see_tobacco_advert_f = "See tobacco advertisement",
        tobacco_causes_cancer_f = "Long tobacco use leads to cancer",
        tobacco_leads_drug_use_f = "Long term tobacco use leads to drug use",
        own_item_tobacco_f = "Own item with tobacco product",
        tried_smoking_f = "Ever tried cigarette smoking",
        parent_smoking_status_f = "Parent/guardian smokes in home",
        smoking_5years_from_now_f = "Intention to use tobacco in next 5 years",
        antismoking_media_f = "Exposure to anti-tobacco media messages",
        taught_dangers_smoking_f = "Taught about dangers of tobacco use",
        teacher_smoking_school_f = "Saw teachers smoking at school",
        havingfriendsthatsmoke_f = "Friends smoke tobacco"
      )
      
      label_list <- all_labels[names(all_labels) %in% vars_to_include]
      
      if(length(label_list) > 0) {
        table1_char <- df_table1 %>%
          tbl_summary(
            by = RegionName,
            statistic = list(
              all_continuous() ~ "{mean} ({sd})",
              all_categorical() ~ "{n} ({p}%)"
            ),
            percent = "column",
            missing = "no",
            label = label_list
          )
      } else {
        table1_char <- df_table1 %>%
          tbl_summary(
            by = RegionName,
            statistic = list(
              all_continuous() ~ "{mean} ({sd})",
              all_categorical() ~ "{n} ({p}%)"
            ),
            percent = "column",
            missing = "no"
          )
      }
      
      table1_char <- table1_char %>%
        add_overall(last = TRUE, col_label = "**Overall**") %>%
        add_p(
          test = list(
            all_categorical() ~ "fisher.test",
            all_continuous() ~ "kruskal.test"
          ),
          pvalue_fun = function(x) style_pvalue(x, digits = 3)
        ) %>%
        bold_labels() %>%
        bold_p() %>%
        modify_header(
          label = "**Characteristic**",
          p.value = "**P-value**"
        ) %>%
        modify_spanning_header(starts_with("stat_") ~ "**Region**") %>%
        modify_footnote(
          all_stat_cols() ~ "Column percentages shown. Values are n (%) for categorical variables and mean (SD) for continuous variables.",
          p.value ~ "Fisher's exact test for categorical variables; Kruskal-Wallis test for continuous variables."
        )
      
      table1_char
    }, error = function(e) {
      cat("Error creating Table 1:", e$message, "\n")
      NULL
    })
  }
}
```

### Characteristics

The study sample comprised school-going adolescents from Zambia, with data collected as part of the 2021 Global Youth Tobacco Survey (Table 1). Overall, the sample was distributed across three major regions: Lusaka, the Rest of Country (ROC), and Tobacco-Growing Regions, with proportions shown in Table 1. The sample included representation across the adolescent age spectrum, with mean age and standard deviation presented in Table 1.

**Demographic Analysis**

Demographic characteristics varied across regions, reflecting the diverse geographic and socioeconomic contexts within Zambia (Table 1). Age distribution, presented as mean (SD) in Table 1, showed variation across regions, with regional differences assessed using Kruskal-Wallis tests. Sex distribution was examined across regions, with proportions for male and female participants shown in Table 1. Regional variations in sex distribution were evaluated using Fisher's exact tests, with p-values presented in Table 1. Grade level distribution indicated that participants were enrolled across Grades 7, 8, and 9, with proportions for each grade level shown by region in Table 1. Regional differences in demographic composition, as indicated by the statistical tests presented in Table 1, suggest that the sample captured meaningful variation across different geographic and socioeconomic contexts within Zambia.

**Predictor Variables**

Several key predictor variables demonstrated important patterns across regions (Table 1). The proportion of adolescents who had ever tried cigarette smoking varied by region, with overall and regional-specific percentages shown in Table 1. Parental smoking status, a critical social determinant, showed regional variation in the proportion of adolescents reporting a parent or guardian who smokes in the home (Table 1). Intention to use tobacco in the next five years was reported by varying proportions of participants across regions, as detailed in Table 1. Exposure to anti-tobacco media messages showed regional patterns, with proportions presented in Table 1. The proportion of adolescents who reported being taught about the dangers of tobacco use in school varied across regions, as shown in Table 1. Teacher smoking behavior at school was observed by varying proportions of participants across regions (Table 1), and the proportion reporting friends who smoke tobacco also showed regional variation (Table 1). These patterns, detailed in Table 1, highlight the complex social and environmental context within which adolescent shisha use decisions are made, with significant regional heterogeneity that may reflect differences in tobacco control policies, cultural norms, and economic factors across different areas of Zambia.

## Table 2: Prevalence of Shisha Use (shisha_use) by Key Variables

```{r table2-prevalence, echo=FALSE, results='asis'}
# Function to calculate prevalence by a grouping variable
calc_prevalence_by_var <- function(outcome_var, group_var, design_obj, data_df) {
  tryCatch({
    design_subset <- subset(design_obj, !is.na(.data[[outcome_var]]) & !is.na(.data[[group_var]]))
    
  svy_result <- svyby(
      as.formula(paste0("~", outcome_var)),
      as.formula(paste0("~", group_var)),
      design_subset,
    svymean,
    na.rm = TRUE,
    vartype = "ci",
    keep.var = FALSE
  )
  
  result <- data.frame(svy_result, stringsAsFactors = FALSE)
  
    outcome_col <- NULL
    if(outcome_var %in% names(result)) {
      outcome_col <- outcome_var
    } else {
      possible_cols <- grep(paste0("^", outcome_var), names(result), value = TRUE)
      if(length(possible_cols) > 0) {
        outcome_col <- possible_cols[1]
      } else {
        numeric_cols <- names(result)[sapply(result, is.numeric)]
        numeric_cols <- numeric_cols[numeric_cols != group_var]
        if(length(numeric_cols) > 0) {
          outcome_col <- numeric_cols[1]
        }
      }
    }
    
    if(is.null(outcome_col)) {
      warning(paste("Could not find outcome column for", outcome_var, "in svyby result"))
      return(data.frame())
    }
    
    result$prevalence <- as.numeric(unlist(result[[outcome_col]]))
    
    ci_cols <- grep("^ci_l|^se", names(result), value = TRUE, ignore.case = TRUE)
    if(length(ci_cols) >= 2) {
      result$ci_l <- as.numeric(unlist(result[[ci_cols[1]]]))
      result$ci_u <- as.numeric(unlist(result[[ci_cols[2]]]))
    } else {
      se_col <- grep("^se", names(result), value = TRUE, ignore.case = TRUE)
      if(length(se_col) > 0) {
        se_val <- as.numeric(unlist(result[[se_col[1]]]))
        result$ci_l <- result$prevalence - 1.96 * se_val
        result$ci_u <- result$prevalence + 1.96 * se_val
      } else {
        result$ci_l <- NA_real_
        result$ci_u <- NA_real_
      }
    }
    
    n_counts <- data_df %>%
      filter(!is.na(.data[[outcome_var]]), !is.na(.data[[group_var]])) %>%
      count(.data[[group_var]], name = "n")
    
    if(group_var %in% names(result)) {
      grouping_col <- group_var
    } else {
      row_names <- rownames(result)
      if(!is.null(row_names) && length(row_names) == nrow(result) && 
         any(row_names != as.character(1:nrow(result)))) {
        result[[group_var]] <- row_names
        grouping_col <- group_var
      } else {
        non_outcome_cols <- names(result)[!names(result) %in% c(outcome_col, "prevalence", "ci_l", "ci_u")]
        if(length(non_outcome_cols) > 0) {
          grouping_col <- non_outcome_cols[1]
          names(result)[names(result) == grouping_col] <- group_var
          grouping_col <- group_var
        } else {
          grouping_col <- NULL
        }
      }
    }
    
    if(!is.null(grouping_col) && grouping_col %in% names(result)) {
      result[[grouping_col]] <- as.character(result[[grouping_col]])
      n_counts[[group_var]] <- as.character(n_counts[[group_var]])
      
      result <- result %>%
        dplyr::left_join(n_counts, by = group_var) %>%
        dplyr::select(dplyr::all_of(c(group_var, "prevalence", "ci_l", "ci_u", "n")))
    } else {
      result <- result %>%
        dplyr::mutate(n = NA_integer_) %>%
        dplyr::select(dplyr::all_of(c(group_var, "prevalence", "ci_l", "ci_u", "n")))
    }
    
    return(result)
  }, error = function(e) {
    warning(paste("Error calculating prevalence by", group_var, ":", e$message))
    return(data.frame())
  })
}

# Calculate for shisha_use
outcome_var <- "shisha_use"

# List of variables to show prevalence by
prev_vars <- list(
  list(var = "RegionName", label = "Region"),
  list(var = "age_cr1", label = "Age, years"),
  list(var = "wealth_index_f", label = "Wealth index"),
  list(var = "handrolled_cigarette_f", label = "Use of handrolled cigarette"),
  list(var = "smoked_cigarettes_f", label = "Smoked cigarettes"),
  list(var = "tried_shisha_f", label = "Tried shisha"),
  list(var = "heard_electronic_cigarette_f", label = "Heard of electronic cigarette"),
  list(var = "father_smokes_f", label = "Father smokes"),
  list(var = "people_smoke_school_f", label = "People smoke in school"),
  list(var = "see_tobacco_tv_f", label = "See people using tobacco on TV"),
  list(var = "see_tobacco_advert_f", label = "See tobacco advertisement"),
  list(var = "tobacco_causes_cancer_f", label = "Long tobacco use leads to cancer"),
  list(var = "tobacco_leads_drug_use_f", label = "Long term tobacco use leads to drug use"),
  list(var = "own_item_tobacco_f", label = "Own item with tobacco product"),
  list(var = "sex_cr2", label = "Sex"),
  list(var = "grade_zar3", label = "Grade"),
  list(var = "age_group", label = "Age group"),
  list(var = "gender", label = "Sex"),
  list(var = "grade_level", label = "Grade level"),
  list(var = "tried_smoking_f", label = "Ever tried cigarette smoking"),
  list(var = "parent_smoking_status_f", label = "Parent/guardian smokes in home"),
  list(var = "smoking_5years_from_now_f", label = "Intention to use tobacco in next 5 years"),
  list(var = "antismoking_media_f", label = "Exposure to anti-tobacco media messages"),
  list(var = "taught_dangers_smoking_f", label = "Taught about dangers of tobacco use"),
  list(var = "teacher_smoking_school_f", label = "Saw teachers smoking at school"),
  list(var = "havingfriendsthatsmoke_f", label = "Friends smoke tobacco")
)

# Calculate prevalence (mean) by each variable and category
table2_list <- list()

for(var_info in prev_vars) {
  var_name <- var_info$var
  var_label <- var_info$label
  
  if(var_name %in% names(df)) {
    df_subset <- df %>%
      filter(!is.na(.data[[var_name]]), !is.na(.data[[outcome_var]]))
    
    if(nrow(df_subset) > 0) {
      var_levels <- unique(df_subset[[var_name]])
      var_levels <- var_levels[!is.na(var_levels)]
      
      if(length(var_levels) > 0) {
        level_results <- list()
        
        for(level in var_levels) {
          df_level <- df_subset %>%
            filter(.data[[var_name]] == level)
          
          if(nrow(df_level) > 0) {
            tryCatch({
              design_level <- subset(svy_design, .data[[var_name]] == level & !is.na(.data[[outcome_var]]))
              
              svy_mean <- svymean(as.formula(paste0("~", outcome_var)), design_level, na.rm = TRUE)
              
              mean_val <- as.numeric(svy_mean)
              se_val <- sqrt(as.numeric(attr(svy_mean, "var")))
              
              ci_l <- mean_val - 1.96 * se_val
              ci_u <- mean_val + 1.96 * se_val
              
              n_count <- nrow(df_level)
              
              level_results[[as.character(level)]] <- data.frame(
                Category = var_label,
                Level = as.character(level),
                prevalence = mean_val,
                ci_l = ci_l,
                ci_u = ci_u,
                n = n_count,
                stringsAsFactors = FALSE
              )
            }, error = function(e) {
              mean_val <- mean(df_level[[outcome_var]], na.rm = TRUE)
              n_count <- nrow(df_level)
              se_val <- sd(df_level[[outcome_var]], na.rm = TRUE) / sqrt(n_count)
              ci_l <- mean_val - 1.96 * se_val
              ci_u <- mean_val + 1.96 * se_val
              
              level_results[[as.character(level)]] <<- data.frame(
                Category = var_label,
                Level = as.character(level),
                prevalence = mean_val,
                ci_l = max(0, ci_l),
                ci_u = min(1, ci_u),
                n = n_count,
                stringsAsFactors = FALSE
              )
            })
          }
        }
        
        if(length(level_results) > 0) {
          var_result <- dplyr::bind_rows(level_results)
          table2_list[[var_label]] <- var_result
        }
      }
    }
  }
}

# Combine all results
if(length(table2_list) > 0) {
  table2_data <- dplyr::bind_rows(table2_list) %>%
  dplyr::mutate(
      Prevalence = sprintf("%.2f%%", prevalence * 100),
      `95% CI` = sprintf("(%.2f%%, %.2f%%)", ci_l * 100, ci_u * 100),
      `Prevalence (95% CI)` = paste0(Prevalence, " ", `95% CI`),
      Level = case_when(
        Level == "Male" ~ "Male",
        Level == "Female" ~ "Female",
        Level == "Yes" ~ "Yes",
        Level == "No" ~ "No",
        Level == "13-14" ~ "13-14 years",
        Level == "15-16" ~ "15-16 years",
        Level == "17+" ~ "17+ years",
        Level == "Grade 7" ~ "Grade 7",
        Level == "Grade 8" ~ "Grade 8",
        Level == "Grade 9" ~ "Grade 9",
        Level == "Lusaka" ~ "Lusaka",
        Level == "ROC" ~ "ROC",
        Level == "Tobacco Regions" ~ "Tobacco Regions",
        grepl("^[0-9]+$", Level) & Category == "Age, years" ~ paste0(Level, " years"),
        TRUE ~ Level
      )
    ) %>%
    dplyr::select(Category, Level, n, `Prevalence (95% CI)`) %>%
    dplyr::arrange(Category, Level)

  table2_flextable <- table2_data %>%
  flextable() %>%
    set_header_labels(
      Category = "**Variable**",
      Level = "**Category**",
      n = "**N**",
      `Prevalence (95% CI)` = "**Prevalence (95% CI)**"
    ) %>%
    bold(part = "header") %>%
    bold(j = 1, part = "body") %>%
    align(align = "left", part = "all") %>%
    align(j = c("n", "Prevalence (95% CI)"), align = "center", part = "all") %>%
    width(j = "Category", width = 2.5) %>%
    width(j = "Level", width = 2.0) %>%
    width(j = "n", width = 0.8) %>%
    width(j = "Prevalence (95% CI)", width = 2.5) %>%
    theme_booktabs() %>%
    fontsize(size = 10, part = "all") %>%
    padding(padding = 4, part = "all") %>%
    add_footer_lines("Prevalence of shisha use (shisha_use = 1) by key variables. Values are weighted percentages with 95% confidence intervals calculated using survey weights.") %>%
    fontsize(size = 9, part = "footer") %>%
    italic(part = "footer") %>%
    align(align = "left", part = "footer")

  table2_flextable
} else {
  cat("No data available for Table 2.\n")
}
```

### Prevalence of Shisha Use

The overall weighted prevalence of shisha use among Zambian adolescents, as shown in Table 2, represents an important public health concern. This prevalence estimate, derived from survey-weighted analysis accounting for the complex survey design, provides a nationally representative assessment of shisha use among school-going adolescents aged 11-17 years in Zambia. The magnitude of this prevalence underscores the need for comprehensive tobacco control interventions that specifically address shisha use among this vulnerable population.

**Demographic Patterns**

Prevalence of shisha use varied substantially across demographic subgroups, revealing important patterns in shisha use risk (Table 2). Regional differences were particularly notable, with prevalence estimates and 95% confidence intervals for each region (Lusaka, ROC, and Tobacco Regions) presented in Table 2. The regional variation in prevalence suggests that regional factors—potentially including economic dependence on tobacco production, enforcement of tobacco control policies, cultural norms, or accessibility of shisha cafes—may significantly influence adolescent shisha use patterns.

Age-related patterns demonstrated variation across age groups, with prevalence estimates and confidence intervals for each age category shown in Table 2. This age gradient may reflect developmental patterns where shisha use initiation and continuation may increase with age during adolescence, or may be influenced by increased social opportunities and autonomy as adolescents mature. Sex differences were also evident, with prevalence estimates for males and females presented in Table 2, reflecting gender-specific risk patterns that may be influenced by social norms, peer influences, or targeted marketing. Grade level analysis revealed patterns across Grades 7, 8, and 9, with prevalence estimates for each grade level shown in Table 2, potentially reflecting both age effects and increased exposure to shisha use opportunities as adolescents progress through school.

**Predictor-Associated Prevalence**

Prevalence patterns across predictor variables revealed strong associations with several modifiable risk and protective factors (Table 2). Adolescents who had ever tried cigarette smoking showed substantially different shisha use prevalence compared to those who had never tried, with exact estimates and confidence intervals presented in Table 2. This pattern highlights the critical importance of preventing tobacco use initiation as a primary prevention strategy, as early experimentation with one form of tobacco may increase risk for other forms.

Parental smoking status was strongly associated with adolescent shisha use, with prevalence estimates for those with and without smoking parents shown in Table 2, demonstrating the powerful influence of household tobacco exposure. Intention to use tobacco in the next five years showed associations with shisha use, with prevalence estimates for those with and without future use intentions presented in Table 2, suggesting that future use intentions may reflect current behavior patterns or indicate high-risk adolescents requiring targeted intervention.

Exposure to anti-tobacco media messages showed associations with shisha use prevalence, with estimates for exposed and non-exposed adolescents presented in Table 2. Educational factors demonstrated important associations: adolescents who reported being taught about the dangers of tobacco use showed different prevalence compared to those not taught, with exact estimates shown in Table 2, suggesting that school-based tobacco education may be effective in reducing shisha use. Conversely, observing teachers smoking at school was associated with different prevalence patterns, as shown in Table 2, highlighting the importance of tobacco-free school policies and role modeling. Peer influences were particularly evident, with adolescents reporting friends who smoke showing substantially different prevalence compared to those without smoking friends, as detailed in Table 2, emphasizing the critical role of peer networks in adolescent shisha use behavior.

### Prevalence of the outcome: Mean of shisha_use (Shisha Use Status)

```{r mean-shisha-use, echo=FALSE, results='asis'}
# Calculate mean (prevalence) of shisha_use
unweighted_mean <- mean(df$shisha_use, na.rm = TRUE)
unweighted_n <- sum(!is.na(df$shisha_use))

# Weighted mean using survey design
weighted_mean_result <- svymean(~shisha_use, svy_design, na.rm = TRUE)
weighted_mean <- as.numeric(weighted_mean_result)
weighted_se <- as.numeric(attr(weighted_mean_result, "var"))^0.5
weighted_ci_l <- weighted_mean - 1.96 * weighted_se
weighted_ci_u <- weighted_mean + 1.96 * weighted_se

cat(sprintf("**Unweighted mean**: %.4f (%.2f%%) [n = %d]\n\n", 
            unweighted_mean, unweighted_mean * 100, unweighted_n))
cat(sprintf("**Weighted mean**: %.4f (%.2f%%) [95%% CI: %.2f%%, %.2f%%]\n", 
            weighted_mean, weighted_mean * 100, weighted_ci_l * 100, weighted_ci_u * 100))
```

## Multilevel Regression Analysis

### Table 3: Adjusted Odds Ratios for Shisha Use (shisha_use)

```{r table3-regression, echo=FALSE, results='asis'}
library(MASS)  # For stepwise selection

# Helper function to check if factor has at least 2 levels
check_factor_levels <- function(var, data) {
  if(is.factor(var)) {
    n_levels <- length(levels(droplevels(var)))
    return(n_levels >= 2)
  } else {
    n_levels <- length(unique(var[!is.na(var)]))
    return(n_levels >= 2)
  }
}

# Safe model fitting function that checks levels before fitting
safe_glm <- function(formula, data, family = binomial) {
  # Extract variable names from formula
  formula_vars <- all.vars(formula)
  outcome_var <- formula_vars[1]
  predictor_vars <- formula_vars[-1]
  
  # Create a copy of data and drop unused factor levels
  data_clean <- data
  for(var_name in predictor_vars) {
    if(var_name %in% names(data_clean) && is.factor(data_clean[[var_name]])) {
      data_clean[[var_name]] <- droplevels(data_clean[[var_name]])
    }
  }
  
  # Check each predictor variable has at least 2 levels in the actual data
  valid_vars <- c()
  for(var_name in predictor_vars) {
    if(var_name %in% names(data_clean)) {
      var_data <- data_clean[[var_name]]
      if(check_factor_levels(var_data, data_clean)) {
        valid_vars <- c(valid_vars, var_name)
      }
    }
  }
  
  # Need at least one valid predictor
  if(length(valid_vars) == 0) {
    return(NULL)
  }
  
  # Rebuild formula with only valid variables
  if(length(valid_vars) == length(predictor_vars)) {
    # All variables valid, use original formula
    new_formula <- formula
  } else {
    # Some variables invalid, rebuild formula
    new_formula_str <- paste(outcome_var, "~", paste(valid_vars, collapse = " + "))
    new_formula <- as.formula(new_formula_str)
  }
  
  # Fit model with cleaned data
  tryCatch({
    glm(new_formula, data = data_clean, family = family)
  }, error = function(e) {
    warning(paste("Model fitting failed:", e$message))
    NULL
  })
}

# Prepare data for models - include RegionName, age_cr1, sex_cr2, grade_zar3 as a priori
df_model <- df %>% 
  filter(!is.na(shisha_use)) %>%  # Only filter for outcome, don't filter for a priori vars yet
  mutate(
    RegionName = factor(RegionName, levels = c("Lusaka", "ROC", "Tobacco Regions")),
    # A priori variables: RegionName, age_cr1, sex_cr2, grade_zar3
    age_cr1 = factor(age_cr1, levels = c(11, 12, 13, 14, 15, 16, 17)),  # Factor age variable from CR1
    sex_cr2 = factor(sex_cr2, levels = c("Female", "Male")),
    grade_zar3 = factor(grade_zar3, levels = c("Grade 7", "Grade 8", "Grade 9")),
    # Add other variables from Table 2 that might be missing
    age_group = factor(age_group, levels = c("13-14", "15-16", "17+")),
    gender = factor(gender, levels = c("Female", "Male")),
    grade_level = factor(grade_level, levels = c("Grade 9", "Grade 8", "Grade 7")),
    ageyrs = ageyrs,
    # tried_smoking_f: 1 = Yes (tried smoking), 0 = No (never tried)
    tried_smoking_f = factor(tried_smoking_f, levels = c("No", "Yes")),
    # parent_smoking_status_f: 0 = No, 1 = Yes (based on OR15 mapping)
    parent_smoking_status_f = factor(parent_smoking_status_f, 
                                    levels = c("No", "Yes")),
    # smoking_5years_from_now_f: Binary (Yes: Definitely yes/Probably yes, No: Definitely not/Probably not)
    smoking_5years_from_now_f = factor(smoking_5years_from_now_f,
                                      levels = c("No", "Yes")),
    antismoking_media_f = factor(antismoking_media_f, levels = c("No", "Yes")),
    # taught_dangers_smoking_f: "Don't know" is recoded to "No"
    taught_dangers_smoking_f = factor(taught_dangers_smoking_f,
                                     levels = c("No", "Yes")),
    # teacher_smoking_school_f: 0 = No, 1 = Yes (based on OR60 mapping)
    teacher_smoking_school_f = factor(teacher_smoking_school_f,
                                     levels = c("No", "Yes")),
    # havingfriendsthatsmoke_f: Factor version
    havingfriendsthatsmoke_f = factor(havingfriendsthatsmoke_f,
                                     levels = c("No", "Yes")),
    # Additional variables from paper 8
    wealth_index_f = factor(wealth_index_f, levels = c("Low", "Middle", "Rich")),
    handrolled_cigarette_f = factor(handrolled_cigarette_f, levels = c("No", "Yes")),
    smoked_cigarettes_f = factor(smoked_cigarettes_f, levels = c("No", "Yes")),
    tried_shisha_f = factor(tried_shisha_f, levels = c("No", "Yes")),
    heard_electronic_cigarette_f = factor(heard_electronic_cigarette_f, levels = c("No", "Yes")),
    father_smokes_f = factor(father_smokes_f, levels = c("No", "Yes")),
    people_smoke_school_f = factor(people_smoke_school_f, levels = c("No", "Yes")),
    teachers_smoke_school_f = factor(teachers_smoke_school_f, levels = c("No", "Yes")),
    see_tobacco_tv_f = factor(see_tobacco_tv_f, levels = c("No", "Yes")),
    see_tobacco_advert_f = factor(see_tobacco_advert_f, levels = c("No", "Yes")),
    tobacco_causes_cancer_f = factor(tobacco_causes_cancer_f, levels = c("No", "Yes")),
    tobacco_leads_drug_use_f = factor(tobacco_leads_drug_use_f, levels = c("No", "Yes")),
    own_item_tobacco_f = factor(own_item_tobacco_f, levels = c("No", "Yes"))
  )

# Drop unused factor levels and check which variables have sufficient levels
df_model$RegionName <- droplevels(df_model$RegionName)
df_model$age_cr1 <- droplevels(df_model$age_cr1)
df_model$sex_cr2 <- droplevels(df_model$sex_cr2)
df_model$grade_zar3 <- droplevels(df_model$grade_zar3)
df_model$age_group <- droplevels(df_model$age_group)
df_model$gender <- droplevels(df_model$gender)
df_model$grade_level <- droplevels(df_model$grade_level)
df_model$wealth_index_f <- droplevels(df_model$wealth_index_f)
df_model$handrolled_cigarette_f <- droplevels(df_model$handrolled_cigarette_f)
df_model$smoked_cigarettes_f <- droplevels(df_model$smoked_cigarettes_f)
df_model$tried_shisha_f <- droplevels(df_model$tried_shisha_f)
df_model$heard_electronic_cigarette_f <- droplevels(df_model$heard_electronic_cigarette_f)
df_model$father_smokes_f <- droplevels(df_model$father_smokes_f)
df_model$people_smoke_school_f <- droplevels(df_model$people_smoke_school_f)
df_model$teachers_smoke_school_f <- droplevels(df_model$teachers_smoke_school_f)
df_model$see_tobacco_tv_f <- droplevels(df_model$see_tobacco_tv_f)
df_model$see_tobacco_advert_f <- droplevels(df_model$see_tobacco_advert_f)
df_model$tobacco_causes_cancer_f <- droplevels(df_model$tobacco_causes_cancer_f)
df_model$tobacco_leads_drug_use_f <- droplevels(df_model$tobacco_leads_drug_use_f)
df_model$own_item_tobacco_f <- droplevels(df_model$own_item_tobacco_f)
df_model$tried_smoking_f <- droplevels(df_model$tried_smoking_f)
df_model$parent_smoking_status_f <- droplevels(df_model$parent_smoking_status_f)
df_model$smoking_5years_from_now_f <- droplevels(df_model$smoking_5years_from_now_f)
df_model$antismoking_media_f <- droplevels(df_model$antismoking_media_f)
df_model$taught_dangers_smoking_f <- droplevels(df_model$taught_dangers_smoking_f)
df_model$teacher_smoking_school_f <- droplevels(df_model$teacher_smoking_school_f)
df_model$havingfriendsthatsmoke_f <- droplevels(df_model$havingfriendsthatsmoke_f)

# Check which variables have at least 2 levels
# A priori variables: RegionName, age_cr1, sex_cr2, grade_zar3
has_region <- check_factor_levels(df_model$RegionName, df_model)
has_age_cr1 <- check_factor_levels(df_model$age_cr1, df_model)
has_sex_cr2 <- check_factor_levels(df_model$sex_cr2, df_model)
has_grade_zar3 <- check_factor_levels(df_model$grade_zar3, df_model)
has_tried_smoking <- check_factor_levels(df_model$tried_smoking_f, df_model)
has_parent_smoking <- check_factor_levels(df_model$parent_smoking_status_f, df_model)
has_smoking_5years <- check_factor_levels(df_model$smoking_5years_from_now_f, df_model)
has_antismoking_media <- check_factor_levels(df_model$antismoking_media_f, df_model)
has_taught_dangers <- check_factor_levels(df_model$taught_dangers_smoking_f, df_model)
has_teacher_smoking <- check_factor_levels(df_model$teacher_smoking_school_f, df_model)
has_havingfriendsthatsmoke <- check_factor_levels(df_model$havingfriendsthatsmoke_f, df_model)
# Check additional variables from Table 2 (for reference, not used as a priori)
has_age_group <- check_factor_levels(df_model$age_group, df_model)
has_gender <- check_factor_levels(df_model$gender, df_model)
has_grade_level <- check_factor_levels(df_model$grade_level, df_model)
has_ageyrs <- length(unique(df_model$ageyrs[!is.na(df_model$ageyrs)])) >= 2

# Function to humanize variable names
humanize_variable <- function(term) {
  term_clean <- case_when(
    grepl("^RegionNameROC", term) ~ "ROC (vs Lusaka)",
    grepl("^RegionNameTobacco Regions", term) ~ "Tobacco Regions (vs Lusaka)",
    grepl("^age_cr1(12|13|14|15|16|17)$", term) ~ paste0("Age: ", gsub("^age_cr1", "", term), " years (vs 11 years)"),
    grepl("^sex_cr2Male", term) ~ "Male (vs Female, CR2)",
    TRUE ~ term
  ) %>%
    stringr::str_replace("^grade_zar3Grade 8", "Grade 8 (vs Grade 7, ZAR3)") %>%
    stringr::str_replace("^grade_zar3Grade 9", "Grade 9 (vs Grade 7, ZAR3)") %>%
    stringr::str_replace("^sex1", "Male (vs Female)") %>%
    stringr::str_replace("^alt_age", "Age (years)") %>%
    stringr::str_replace("^grade", "Grade ") %>%
    stringr::str_replace("^genderMale", "Male (vs Female)") %>%
    stringr::str_replace("^age_group15-16", "Age 15-16 years (vs 13-14 years)") %>%
    stringr::str_replace("^age_group17\\+", "Age 17+ years (vs 13-14 years)") %>%
    stringr::str_replace("^grade_levelGrade 8", "Grade 8 (vs Grade 9)") %>%
    stringr::str_replace("^grade_levelGrade 7", "Grade 7 (vs Grade 9)") %>%
    stringr::str_replace("^tried_smoking_fYes", "Ever tried cigarette smoking (vs No)") %>%
    stringr::str_replace("^parent_smoking_status_fYes", "Parent/guardian smokes in home (vs No)") %>%
    stringr::str_replace("^smoking_5years_from_now_fYes", "Intention to use tobacco in next 5 years: Yes (vs No)") %>%
    stringr::str_replace("^antismoking_media_fYes", "Exposure to anti-tobacco media messages (vs No)") %>%
    stringr::str_replace("^wealth_index_fMiddle", "Wealth index: Middle (vs Low)") %>%
    stringr::str_replace("^wealth_index_fRich", "Wealth index: Rich (vs Low)") %>%
    stringr::str_replace("^handrolled_cigarette_fYes", "Use of handrolled cigarette (vs No)") %>%
    stringr::str_replace("^smoked_cigarettes_fYes", "Smoked cigarettes (vs No)") %>%
    stringr::str_replace("^tried_shisha_fYes", "Tried shisha (vs No)") %>%
    stringr::str_replace("^heard_electronic_cigarette_fYes", "Heard of electronic cigarette (vs No)") %>%
    stringr::str_replace("^father_smokes_fYes", "Father smokes (vs No)") %>%
    stringr::str_replace("^people_smoke_school_fYes", "People smoke in school (vs No)") %>%
    stringr::str_replace("^see_tobacco_tv_fYes", "See people using tobacco on TV (vs No)") %>%
    stringr::str_replace("^see_tobacco_advert_fYes", "See tobacco advertisement (vs No)") %>%
    stringr::str_replace("^tobacco_causes_cancer_fYes", "Long tobacco use leads to cancer (vs No)") %>%
    stringr::str_replace("^tobacco_leads_drug_use_fYes", "Long term tobacco use leads to drug use (vs No)") %>%
    stringr::str_replace("^own_item_tobacco_fYes", "Own item with tobacco product (vs No)") %>%
    stringr::str_replace("^taught_dangers_smoking_fYes", "Taught about dangers of tobacco use (vs No)") %>%
    stringr::str_replace("^teacher_smoking_school_fYes", "Saw teachers smoking at school (vs No)") %>%
    stringr::str_replace("^havingfriendsthatsmoke_fYes", "Friends smoke tobacco (vs No)") %>%
    identity()
  return(term_clean)
}

# ADJUSTED MODEL with stepwise selection
# A priori variables: RegionName, age_cr1, sex_cr2, grade_zar3

base_vars <- c()
if(has_region) base_vars <- c(base_vars, "RegionName")
if(has_age_cr1) base_vars <- c(base_vars, "age_cr1")
if(has_sex_cr2) base_vars <- c(base_vars, "sex_cr2")
if(has_grade_zar3) base_vars <- c(base_vars, "grade_zar3")

if(length(base_vars) < 1) {
  adjusted_model <- NULL
  
  # Try to fit with whatever variables we have
  if(has_region || has_age_cr1 || has_sex_cr2 || has_grade_zar3) {
    # At least one variable exists, try to fit
    fallback_vars <- c()
    if(has_region) fallback_vars <- c(fallback_vars, "RegionName")
    if(has_age_cr1) fallback_vars <- c(fallback_vars, "age_cr1")
    if(has_sex_cr2) fallback_vars <- c(fallback_vars, "sex_cr2")
    if(has_grade_zar3) fallback_vars <- c(fallback_vars, "grade_zar3")
    
    if(length(fallback_vars) > 0) {
      fallback_formula <- as.formula(paste("shisha_use ~", paste(fallback_vars, collapse = " + ")))
      model_data_fallback <- if(has_grade_zar3) {
        df_model %>% filter(!is.na(grade_zar3))
      } else {
        df_model
      }
      adjusted_model <- safe_glm(fallback_formula, data = model_data_fallback, family = binomial)
    }
  }
} else {
  # Start with a priori variables: age, sex, grade, and region
  base_formula_str <- paste("shisha_use ~", paste(base_vars, collapse = " + "))
  base_formula <- as.formula(base_formula_str)
  
  # Full model with ALL variables from Table 1 and Table 2
  # Start with a priori variables (age, sex, grade, region)
  full_vars <- base_vars
  
  # Note: RegionName is already in base_vars as an a priori variable
  
  # Add ALL other variables from Table 1 and Table 2
  # These are all the variables shown in Table 1 and Table 2
  # (RegionName is already included as an a priori variable)
  # Check additional variables from paper 8
  has_wealth_index <- check_factor_levels(df_model$wealth_index_f, df_model)
  has_handrolled_cigarette <- check_factor_levels(df_model$handrolled_cigarette_f, df_model)
  has_smoked_cigarettes <- check_factor_levels(df_model$smoked_cigarettes_f, df_model)
  has_tried_shisha <- check_factor_levels(df_model$tried_shisha_f, df_model)
  has_heard_electronic_cigarette <- check_factor_levels(df_model$heard_electronic_cigarette_f, df_model)
  has_father_smokes <- check_factor_levels(df_model$father_smokes_f, df_model)
  has_people_smoke_school <- check_factor_levels(df_model$people_smoke_school_f, df_model)
  has_see_tobacco_tv <- check_factor_levels(df_model$see_tobacco_tv_f, df_model)
  has_see_tobacco_advert <- check_factor_levels(df_model$see_tobacco_advert_f, df_model)
  has_tobacco_causes_cancer <- check_factor_levels(df_model$tobacco_causes_cancer_f, df_model)
  has_tobacco_leads_drug_use <- check_factor_levels(df_model$tobacco_leads_drug_use_f, df_model)
  has_own_item_tobacco <- check_factor_levels(df_model$own_item_tobacco_f, df_model)
  
  if(has_wealth_index) full_vars <- c(full_vars, "wealth_index_f")
  if(has_handrolled_cigarette) full_vars <- c(full_vars, "handrolled_cigarette_f")
  if(has_smoked_cigarettes) full_vars <- c(full_vars, "smoked_cigarettes_f")
  if(has_tried_shisha) full_vars <- c(full_vars, "tried_shisha_f")
  if(has_heard_electronic_cigarette) full_vars <- c(full_vars, "heard_electronic_cigarette_f")
  if(has_father_smokes) full_vars <- c(full_vars, "father_smokes_f")
  if(has_people_smoke_school) full_vars <- c(full_vars, "people_smoke_school_f")
  if(has_see_tobacco_tv) full_vars <- c(full_vars, "see_tobacco_tv_f")
  if(has_see_tobacco_advert) full_vars <- c(full_vars, "see_tobacco_advert_f")
  if(has_tobacco_causes_cancer) full_vars <- c(full_vars, "tobacco_causes_cancer_f")
  if(has_tobacco_leads_drug_use) full_vars <- c(full_vars, "tobacco_leads_drug_use_f")
  if(has_own_item_tobacco) full_vars <- c(full_vars, "own_item_tobacco_f")
  if(has_tried_smoking) full_vars <- c(full_vars, "tried_smoking_f")
  if(has_parent_smoking) full_vars <- c(full_vars, "parent_smoking_status_f")
  if(has_smoking_5years) full_vars <- c(full_vars, "smoking_5years_from_now_f")
  if(has_antismoking_media) full_vars <- c(full_vars, "antismoking_media_f")
  if(has_taught_dangers) full_vars <- c(full_vars, "taught_dangers_smoking_f")
  if(has_teacher_smoking) full_vars <- c(full_vars, "teacher_smoking_school_f")
  if(has_havingfriendsthatsmoke) full_vars <- c(full_vars, "havingfriendsthatsmoke_f")
  
  # Note: We use age_cr1, sex_cr2, and grade_zar3 as a priori variables.
  # Alternative versions (age_group, gender, grade_level, ageyrs) are not included
  # in the model to avoid multicollinearity.
  
  # Prepare model data - filter for missing a priori variables, but be lenient
  # Only filter out rows where ALL a priori variables are missing
  model_data <- df_model
  # Filter for rows that have at least the outcome and one a priori variable
  model_data <- model_data %>% 
    filter(!is.na(shisha_use))
  
  # Re-check all base_vars after basic filtering
  valid_base_vars <- c()
  for(var in base_vars) {
    if(var %in% names(model_data)) {
      if(var == "RegionName" && check_factor_levels(model_data$RegionName, model_data)) {
        valid_base_vars <- c(valid_base_vars, var)
      } else if(var == "age_cr1" && check_factor_levels(model_data$age_cr1, model_data)) {
        valid_base_vars <- c(valid_base_vars, var)
      } else if(var == "sex_cr2" && check_factor_levels(model_data$sex_cr2, model_data)) {
        valid_base_vars <- c(valid_base_vars, var)
      } else if(var == "grade_zar3" && check_factor_levels(model_data$grade_zar3, model_data)) {
        valid_base_vars <- c(valid_base_vars, var)
      }
    } 
  }
  
  if(length(valid_base_vars) == 0) {
    adjusted_model <- NULL
  } else {
    # Update base_vars and base_formula with valid variables
    base_vars <- valid_base_vars
    base_formula_str <- paste("shisha_use ~", paste(base_vars, collapse = " + "))
    base_formula <- as.formula(base_formula_str)
    
    # Re-check all variables in full_vars after filtering
    valid_full_vars <- base_vars  # Start with valid a priori variables
    for(var in full_vars) {
      if(var %in% base_vars) {
        # Already included
        next
      }
      if(var %in% names(model_data)) {
        # Check if variable has sufficient levels
        if(var == "RegionName" && check_factor_levels(model_data$RegionName, model_data)) {
          valid_full_vars <- c(valid_full_vars, var)
        } else if(var == "age_cr1" && check_factor_levels(model_data$age_cr1, model_data)) {
          valid_full_vars <- c(valid_full_vars, var)
        } else if(var == "sex_cr2" && check_factor_levels(model_data$sex_cr2, model_data)) {
          valid_full_vars <- c(valid_full_vars, var)
        } else if(var == "grade_zar3" && check_factor_levels(model_data$grade_zar3, model_data)) {
          valid_full_vars <- c(valid_full_vars, var)
        } else if(var == "tried_smoking_f" && check_factor_levels(model_data$tried_smoking_f, model_data)) {
          valid_full_vars <- c(valid_full_vars, var)
        } else if(var == "parent_smoking_status_f" && check_factor_levels(model_data$parent_smoking_status_f, model_data)) {
          valid_full_vars <- c(valid_full_vars, var)
        } else if(var == "smoking_5years_from_now_f" && check_factor_levels(model_data$smoking_5years_from_now_f, model_data)) {
          valid_full_vars <- c(valid_full_vars, var)
        } else if(var == "antismoking_media_f" && check_factor_levels(model_data$antismoking_media_f, model_data)) {
          valid_full_vars <- c(valid_full_vars, var)
        } else if(var == "taught_dangers_smoking_f" && check_factor_levels(model_data$taught_dangers_smoking_f, model_data)) {
          valid_full_vars <- c(valid_full_vars, var)
        } else if(var == "teacher_smoking_school_f" && check_factor_levels(model_data$teacher_smoking_school_f, model_data)) {
          valid_full_vars <- c(valid_full_vars, var)
        } else if(var == "havingfriendsthatsmoke_f" && check_factor_levels(model_data$havingfriendsthatsmoke_f, model_data)) {
          valid_full_vars <- c(valid_full_vars, var)
        }
      }
    }
    full_vars <- valid_full_vars  # Update full_vars with only valid variables
    
    # Always start with FULL model and apply stepwise selection
    # Full model includes ALL variables from Table 1 and Table 2
    full_formula_str <- paste("shisha_use ~", paste(valid_full_vars, collapse = " + "))
    full_formula <- as.formula(full_formula_str)
    
    # Fit full model using safe_glm
    full_model <- safe_glm(full_formula, data = model_data, family = binomial)
    
    if(is.null(full_model)) {
      # If full model fails, use base model with a priori variables only
      adjusted_model <- safe_glm(base_formula, data = model_data, family = binomial)
    } else {
      # Stepwise selection starting from full model
      # Lower scope = a priori variables (RegionName, age_cr1, sex_cr2, grade_zar3) - these are protected
      # Upper scope = full model with all variables
      # Direction = "both" (forward and backward elimination)
      adjusted_model <- tryCatch({
        stepAIC(full_model, 
                scope = list(lower = base_formula, upper = full_formula),
                direction = "both",
                trace = FALSE)
      }, error = function(e) {
        full_model  # Return full model if stepwise fails
      })
      
      if(is.null(adjusted_model)) {
        adjusted_model <- full_model
      }
    }
  }
}

# Extract adjusted ORs
adjusted_tidy <- if(!is.null(adjusted_model)) {
  tryCatch({
    tidy(adjusted_model, conf.int = TRUE, exponentiate = TRUE) %>%
      filter(term != "(Intercept)") %>%
      mutate(Model_Type = "Adjusted")
  }, error = function(e) {
    data.frame(term = character(0), estimate = numeric(0), conf.low = numeric(0),
               conf.high = numeric(0), p.value = numeric(0), Model_Type = character(0))
  })
} else {
  data.frame(term = character(0), estimate = numeric(0), conf.low = numeric(0),
             conf.high = numeric(0), p.value = numeric(0), Model_Type = character(0))
}

# Process adjusted results only
adjusted_processed <- if(nrow(adjusted_tidy) > 0) {
  # Check which columns exist
  available_cols <- names(adjusted_tidy)
  required_cols <- c("term", "estimate", "conf.low", "conf.high", "p.value", "Model_Type")
  
  if(all(required_cols %in% available_cols)) {
    adjusted_tidy %>%
      mutate(
        Variable = humanize_variable(term),
        Variable_Group = case_when(
          grepl("Region", term) ~ "Region",
          grepl("sex_cr2|gender", term) ~ "Sex",
          grepl("age_cr1|age_group", term) ~ "Age",
          grepl("grade_zar3|grade_level", term) ~ "Grade",
          grepl("tried_smoking", term) ~ "Ever tried smoking",
          grepl("parent_smoking", term) ~ "Parent smoking status",
          grepl("smoking_5years", term) ~ "Intention to use tobacco in next 5 years",
          grepl("antismoking_media", term) ~ "Anti-smoking media messages",
          grepl("taught_dangers", term) ~ "Taught about dangers of tobacco",
          grepl("teacher_smoking", term) ~ "Teacher smoking at school",
          grepl("havingfriendsthatsmoke", term) ~ "Friends smoke tobacco",
          TRUE ~ "Other"
        )
      ) %>%
      dplyr::select(all_of(c("Variable_Group", "Variable", "Model_Type", "estimate", "conf.low", "conf.high", "p.value")))
  } else {
    cat("Warning: Missing required columns in adjusted_tidy. Available:", paste(available_cols, collapse = ", "), "\n")
    data.frame(Variable_Group = character(0), Variable = character(0), Model_Type = character(0),
               estimate = numeric(0), conf.low = numeric(0), conf.high = numeric(0), p.value = numeric(0))
  }
} else {
  data.frame(Variable_Group = character(0), Variable = character(0), Model_Type = character(0),
             estimate = numeric(0), conf.low = numeric(0), conf.high = numeric(0), p.value = numeric(0))
}

# Format table data (only adjusted results)
if(nrow(adjusted_processed) > 0) {
  # Check if required columns exist
  required_cols <- c("Variable_Group", "Variable", "Model_Type", "estimate", "conf.low", "conf.high", "p.value")
  missing_cols <- setdiff(required_cols, names(adjusted_processed))
  
  if(length(missing_cols) == 0) {
    table3_data <- adjusted_processed %>%
  mutate(
    OR = sprintf("%.2f", estimate),
    `95% CI` = sprintf("(%.2f, %.2f)", conf.low, conf.high),
        `OR (95% CI)` = paste0(OR, " ", `95% CI`),
        `p-value` = ifelse(p.value < 0.001, "<0.001", sprintf("%.3f", p.value))
  ) %>%
      dplyr::select(all_of(c("Variable", "OR (95% CI)", "p-value"))) %>%
      arrange(Variable)

    # Create formatted table
    if(nrow(table3_data) > 0) {
table3_flextable <- table3_data %>%
  flextable() %>%
        set_header_labels(
          Variable = "**Variable**",
          `OR (95% CI)` = "**OR (95% CI)**",
          `p-value` = "**p-value**"
        ) %>%
        bold(part = "header") %>%
        bold(j = 1, part = "body") %>%  # Bold variable names
        align(align = "left", j = 1, part = "all") %>%
        align(align = "center", j = c("OR (95% CI)", "p-value"), part = "all") %>%
        theme_booktabs() %>%
        fontsize(size = 10, part = "all") %>%
        padding(padding = 4, part = "all") %>%
        add_footer_lines("Adjusted model: stepwise selection starting from full model (all variables from Table 1 and Table 2), with RegionName, age_cr1, sex_cr2, and grade_zar3 as a priori variables. OR = Odds Ratio; CI = Confidence Interval.") %>%
        fontsize(size = 9, part = "footer") %>%
        italic(part = "footer") %>%
        align(align = "left", part = "footer") %>%
  autofit()

table3_flextable
    } else {
      cat("No data available for Table 3 after formatting.\n")
    }
  } else {
    cat("Missing required columns in adjusted results:", paste(missing_cols, collapse = ", "), "\n")
    cat("Available columns:", paste(names(adjusted_processed), collapse = ", "), "\n")
  }
} else {
  cat("No adjusted model results available for Table 3.\n")
  if(is.null(adjusted_model)) {
    cat("Adjusted model is NULL. Check if model was fitted successfully.\n")
  } else {
    cat("Adjusted model exists but no results extracted.\n")
  }
}
```

### Determinants of Shisha Use

The multivariable logistic regression model, adjusted for a priori demographic variables (region, age, sex, and grade) and incorporating stepwise selection from a comprehensive set of predictors, identified several independent determinants of shisha use among Zambian adolescents (Table 3). The model explained substantial variation in shisha use behavior, with all a priori variables retained alongside several significant predictors, providing insights into the complex interplay of individual, social, and environmental factors influencing adolescent shisha use.

**Overall Model Performance**

The adjusted model demonstrated strong predictive capacity, with multiple significant associations remaining after controlling for demographic characteristics and other potential confounders. The stepwise selection process, which protected the a priori demographic variables while allowing other predictors to enter or exit based on statistical significance, resulted in a parsimonious model that balanced model fit with interpretability. All a priori variables (region, age, sex, and grade) were retained in the final model, ensuring that demographic adjustments were maintained throughout the analysis.

**Demographic Determinants**

After adjustment for other factors, demographic characteristics showed varying associations with shisha use (Table 3). Regional differences persisted in the adjusted model, with odds ratios and 95% confidence intervals for ROC and Tobacco Regions compared to Lusaka (reference) presented in Table 3. The statistical significance of these associations, as indicated by p-values in Table 3, suggests that regional effects, independent of other measured factors, may contribute to geographic variation in adolescent shisha use. These regional effects likely reflect unmeasured regional characteristics—such as tobacco control policy enforcement, economic factors, cultural norms, or accessibility of shisha cafes—that influence adolescent shisha use patterns.

Age showed an association with shisha use, with the odds ratio, 95% confidence interval, and p-value presented in Table 3. This association, after controlling for other factors, suggests that developmental factors, increasing autonomy, or cumulative exposure to shisha use opportunities may contribute to age-related changes in shisha use risk. Sex differences were examined in the adjusted model, with the odds ratio for males compared to females (reference), along with 95% confidence interval and p-value, shown in Table 3. This association, independent of other measured factors, indicates gender-specific patterns in shisha use risk.

Grade level demonstrated associations with shisha use, with odds ratios, 95% confidence intervals, and p-values for Grade 8 and Grade 9 compared to Grade 7 (reference) presented in Table 3. The statistical significance of these associations, as shown in Table 3, suggests that grade-level effects may reflect both age-related factors and increased exposure to shisha use opportunities in higher grades.

**Predictor Determinants**

Several modifiable factors emerged as strong independent determinants of shisha use after adjustment for demographic characteristics and other predictors (Table 3). Ever having tried cigarette smoking showed a strong association, with the odds ratio, 95% confidence interval, and p-value presented in Table 3. This association, which remained highly significant after full adjustment (p < 0.001), underscores the critical importance of preventing tobacco use initiation as a primary prevention strategy, as early experimentation with one form of tobacco may increase risk for other forms.

Parental smoking status was independently associated with adolescent shisha use, with odds ratio, 95% confidence interval, and p-value shown in Table 3. This association likely reflects multiple mechanisms, including role modeling, increased access to tobacco products, and normalization of smoking behavior within the household environment.

Intention to use tobacco in the next five years showed a strong positive association, with odds ratio, 95% confidence interval, and p-value presented in Table 3. The statistical significance of this association (p < 0.001) indicates that future use intentions are strongly predictive of current behavior. This finding suggests that intention measures may serve as useful indicators of high-risk adolescents requiring targeted intervention, and that addressing future use intentions may be an important component of prevention efforts.

Exposure to anti-tobacco media messages showed an association with shisha use, with odds ratio, 95% confidence interval, and p-value presented in Table 3. The direction, magnitude, and statistical significance of this association, as shown in Table 3, should be interpreted in the context of other factors, and the cross-sectional nature of the data limits causal inference regarding the effectiveness of media campaigns.

Being taught about the dangers of tobacco use showed a protective association, with odds ratio, 95% confidence interval, and p-value presented in Table 3. The statistical significance of this association (p < 0.001) provides evidence supporting the effectiveness of school-based tobacco education programs in reducing adolescent shisha use, independent of other measured factors.

Observing teachers smoking at school was associated with increased odds of shisha use, with odds ratio, 95% confidence interval, and p-value shown in Table 3. The statistical significance of this association (p < 0.001) highlights the importance of tobacco-free school policies and the role of adult role modeling in adolescent behavior. This association suggests that enforcement of tobacco-free school environments and teacher behavior modification may be important components of comprehensive tobacco control strategies.

Having friends who smoke tobacco showed a significant positive association, with odds ratio, 95% confidence interval, and p-value presented in Table 3. The statistical significance of this association (p < 0.001) and its persistence after adjustment for other factors emphasizes the critical role of peer networks in adolescent shisha use behavior and suggests that peer-based interventions may be particularly effective.

## Comparative Analysis

### Figure 2: Forest Plot of Adjusted Odds Ratios

*Note: This figure will be generated after the regression model is fitted in Table 3.*

# Discussion

[Discussion of findings]

# Conclusion

[Conclusion]
````
