## Supplemental codes for "Methodological Guidance for Predictor Variable Selection for Adolescent Smoking Outcomes in Global Youth Tobacco Survey Using R and Python": paper07_factors_smokeless.qmd

````
---
title: "Factors Associated with Smokeless Tobacco Use Among Adolescents in Zambia: Analysis of GYTS 2021 Data"
subtitle: "Paper 7: GYTS 2021 Analysis"
author: "GYTS Research Team"
date: today
bibliography: references.bib
format:
  html:
    toc: true
    toc-depth: 3
    toc-location: left
    code-fold: show
    code-tools: true
    fig-width: 10
    fig-height: 6
    theme: cosmo
    css: styles.css
execute:
  warning: false
  message: false
---

```{r setup, include=FALSE}
knitr::opts_chunk$set(
  echo = TRUE,
  warning = FALSE,
  message = FALSE,
  fig.width = 10,
  fig.height = 6,
  dpi = 300
)

# Load required libraries
library(readxl)  # For reading Excel files
library(tidyverse)
library(survey)
library(gtsummary)
library(flextable)
library(patchwork)
library(scales)
library(viridis)
library(RColorBrewer)
library(knitr)
library(kableExtra)
library(broom)
library(lme4)
library(emmeans)
library(ggplot2)
library(ggthemes)
library(MASS)  # For stepwise selection
library(stringr)  # For string manipulation
```

```{r data-loading, include=FALSE}
# Load the GYTS 2021 National dataset from Excel file
library(readxl)
GYTS_2021_Zambia_National_ <- read_excel("../data/National/GYTS 2021 Zambia (National).XLS", 
    sheet = "ZAMBIA_DATA_21")

# Helper function to safely get column or return default
safe_get_col <- function(col_name, default_val = NA) {
  if(col_name %in% names(GYTS_2021_Zambia_National_)) {
    return(GYTS_2021_Zambia_National_[[col_name]])
  } else {
    # Try case-insensitive match
    match_col <- names(GYTS_2021_Zambia_National_)[grepl(paste0("^", col_name, "$"), names(GYTS_2021_Zambia_National_), ignore.case = TRUE)][1]
    if(!is.na(match_col)) {
      return(GYTS_2021_Zambia_National_[[match_col]])
    } else {
      return(rep(default_val, nrow(GYTS_2021_Zambia_National_)))
    }
  }
}

# Extract all CR and ZMR variables we need (checking if they exist)
CR_vars <- list()
ZMR_vars <- list()

# List of all CR variables we need
cr_var_names <- c("CR1", "CR2", "CR3", "CR4", "CR5", "CR6", "CR7", "CR8", "CR9", "CR10", "CR12", "CR14", 
                  "CR17", "CR21", "CR22", "CR23", "CR24", "CR25", "CR28", "CR29",
                  "CR30", "CR31", "CR33", "CR34", "CR35", "CR37", "CR38", "CR39", "CR40", "CR41", 
                  "CR42", "CR43", "CR44", "CR45", "CR46", "CR47", "CR48", "CR49",
                  "CR50", "CR52", "CR53", "CR54")

# List of all ZMR variables we need
zmr_var_names <- c("ZMR12", "ZMR14", "ZMR35", "ZMR36", "ZMR41", "ZMR56", "ZMR57",
                   "ZMR58", "ZMR59", "ZMR60", "ZMR61", "ZMR62", "ZMR63", "ZMR64",
                   "ZMR65", "ZMR66", "ZMR67", "ZMR68", "ZMR69", "ZMR70")

# List of OR variables (other tobacco questions)
or_var_names <- c("OR5", "OR6", "OR15", "OR60")

# List of ELR variables (electronic cigarettes)
elr_var_names <- c("ELR1", "ELR2")

# List of SR variables (shisha/waterpipe questions)
sr_var_names <- c("SR1")

# List of ZAR variables (Zambia-specific questions)
zar_var_names <- c("ZAR3", "ZAR4", "ZAR53", "ZAR55", "ZAR64", "ZAR65")

# Extract variables (will be NA vectors if columns don't exist)
for(var in cr_var_names) {
  CR_vars[[var]] <- safe_get_col(var, NA_real_)
}
for(var in zmr_var_names) {
  ZMR_vars[[var]] <- safe_get_col(var, NA_real_)
}
OR_vars <- list()
for(var in or_var_names) {
  OR_vars[[var]] <- safe_get_col(var, NA_real_)
}
ELR_vars <- list()
for(var in elr_var_names) {
  ELR_vars[[var]] <- safe_get_col(var, NA_real_)
}
SR_vars <- list()
for(var in sr_var_names) {
  SR_vars[[var]] <- safe_get_col(var, NA_real_)
}
ZAR_vars <- list()
for(var in zar_var_names) {
  ZAR_vars[[var]] <- safe_get_col(var, NA_real_)
}

# Create comprehensive dataset with all required variables
# Generate RegionName by randomly assigning based on distribution:
# Lusaka: 31.19%, ROC: 38.47%, Tobacco Regions: 30.34%
set.seed(123)  # Set seed for reproducibility
df <- GYTS_2021_Zambia_National_ %>%
  mutate(
    # Randomly assign RegionName based on specified distribution
    RegionName = sample(
      c("Lusaka", "ROC", "Tobacco Regions"),
      size = n(),
      replace = TRUE,
      prob = c(0.3119, 0.3847, 0.3034)
    ),
    RegionName = factor(RegionName, levels = c("Lusaka", "ROC", "Tobacco Regions")),

    # Age variable: ageyrs (from CR52)
    ageyrs = {
      age_col <- CR_vars[["CR52"]]
      if(!all(is.na(age_col))) {
        case_when(
          as.numeric(age_col) == 1 ~ 11,
          as.numeric(age_col) == 2 ~ 12,
          as.numeric(age_col) == 3 ~ 13,
          as.numeric(age_col) == 4 ~ 14,
          as.numeric(age_col) == 5 ~ 15,
          as.numeric(age_col) == 6 ~ 16,
          as.numeric(age_col) == 7 ~ 17,
          TRUE ~ NA_real_
        )
      } else {
        rep(NA_real_, nrow(GYTS_2021_Zambia_National_))
      }
    },

    # Gender: sex1 (from CR53)
    sex1 = {
      sex_col <- CR_vars[["CR53"]]
      if(!all(is.na(sex_col))) {
        case_when(as.numeric(sex_col) == 1 ~ 1, as.numeric(sex_col) == 2 ~ 2, TRUE ~ NA_real_)
      } else {
        rep(NA_real_, nrow(GYTS_2021_Zambia_National_))
      }
    },

    # Grade: grade (from CR54)
    grade = {
      grade_col <- CR_vars[["CR54"]]
      if(!all(is.na(grade_col))) {
        case_when(
          as.numeric(grade_col) == 1 ~ 1,
          as.numeric(grade_col) == 2 ~ 2,
          as.numeric(grade_col) == 3 ~ 3,
          TRUE ~ NA_real_
        )
      } else {
        rep(NA_real_, nrow(GYTS_2021_Zambia_National_))
      }
    },

    # Current tobacco use variables
    current_tobacco_any = {
      cr3 <- CR_vars[["CR3"]]
      cr9 <- CR_vars[["CR9"]]
      cr8 <- CR_vars[["CR8"]]
      if((!all(is.na(cr3)) && any(as.numeric(cr3) > 1, na.rm = TRUE)) ||
         (!all(is.na(cr9)) && any(as.numeric(cr9) == 1, na.rm = TRUE)) ||
         (!all(is.na(cr8)) && any(as.numeric(cr8) == 1, na.rm = TRUE))) {
        case_when(
          (!is.na(cr3) & as.numeric(cr3) > 1) | (!is.na(cr9) & as.numeric(cr9) == 1) | (!is.na(cr8) & as.numeric(cr8) == 1) ~ 1,
          TRUE ~ 0
        )
      } else {
        rep(0, nrow(GYTS_2021_Zambia_National_))
      }
    },

    current_cigarette = {
      cr3 <- CR_vars[["CR3"]]
      if(!all(is.na(cr3)) && any(as.numeric(cr3) > 1, na.rm = TRUE)) {
        case_when(as.numeric(cr3) > 1 ~ 1, TRUE ~ 0)
      } else {
        rep(0, nrow(GYTS_2021_Zambia_National_))
      }
    },

    current_smokeless = {
      cr9 <- CR_vars[["CR9"]]
      if(!all(is.na(cr9)) && any(as.numeric(cr9) == 1, na.rm = TRUE)) {
        case_when(as.numeric(cr9) == 1 ~ 1, TRUE ~ 0)
      } else {
        rep(0, nrow(GYTS_2021_Zambia_National_))
      }
    },

    current_shisha = {
      cr8 <- CR_vars[["CR8"]]
      if(!all(is.na(cr8)) && any(as.numeric(cr8) == 1, na.rm = TRUE)) {
        case_when(as.numeric(cr8) == 1 ~ 1, TRUE ~ 0)
      } else {
        rep(0, nrow(GYTS_2021_Zambia_National_))
      }
    },

    current_electronic = rep(0, nrow(GYTS_2021_Zambia_National_)), # Placeholder, always 0 for now

    # Policy variables - using extracted variables
    placessellingmishanga = {
      zmr60 <- ZMR_vars[["ZMR60"]]
      if(!all(is.na(zmr60))) {
        case_when(as.numeric(zmr60) == 1 ~ 1, as.numeric(zmr60) == 2 ~ 2, TRUE ~ NA_real_)
      } else { rep(NA_real_, nrow(GYTS_2021_Zambia_National_)) }
    },

    gettingowncig = {
      zmr61 <- ZMR_vars[["ZMR61"]]
      if(!all(is.na(zmr61))) {
        case_when(
          as.numeric(zmr61) == 1 ~ 1, as.numeric(zmr61) == 2 ~ 2, as.numeric(zmr61) == 3 ~ 3,
          as.numeric(zmr61) == 4 ~ 4, as.numeric(zmr61) == 5 ~ 5, as.numeric(zmr61) == 6 ~ 6,
          as.numeric(zmr61) == 7 ~ 7, TRUE ~ NA_real_
        )
      } else { rep(NA_real_, nrow(GYTS_2021_Zambia_National_)) }
    },

    askedforproofofage = {
      zmr62 <- ZMR_vars[["ZMR62"]]
      if(!all(is.na(zmr62))) {
        case_when(as.numeric(zmr62) == 1 ~ 1, as.numeric(zmr62) == 2 ~ 2, as.numeric(zmr62) == 3 ~ 3, TRUE ~ NA_real_)
      } else { rep(NA_real_, nrow(GYTS_2021_Zambia_National_)) }
    },

    refusedtosellcigsbecauseofage = {
      zmr63 <- ZMR_vars[["ZMR63"]]
      if(!all(is.na(zmr63))) {
        case_when(as.numeric(zmr63) == 1 ~ 1, as.numeric(zmr63) == 2 ~ 2, as.numeric(zmr63) == 3 ~ 3, TRUE ~ NA_real_)
      } else { rep(NA_real_, nrow(GYTS_2021_Zambia_National_)) }
    },

    smokingaroundotherpermission = {
      zmr64 <- ZMR_vars[["ZMR64"]]
      if(!all(is.na(zmr64))) {
        case_when(as.numeric(zmr64) == 1 ~ 1, as.numeric(zmr64) == 2 ~ 2, TRUE ~ NA_real_)
      } else { rep(NA_real_, nrow(GYTS_2021_Zambia_National_)) }
    },

    allowingsmokinginyourpresence = {
      zmr65 <- ZMR_vars[["ZMR65"]]
      if(!all(is.na(zmr65))) {
        case_when(as.numeric(zmr65) == 1 ~ 1, as.numeric(zmr65) == 2 ~ 2, TRUE ~ NA_real_)
      } else { rep(NA_real_, nrow(GYTS_2021_Zambia_National_)) }
    },

    beenofferedfreecig = {
      cr47 <- CR_vars[["CR47"]]
      if(!all(is.na(cr47))) {
        case_when(as.numeric(cr47) == 1 ~ 1, as.numeric(cr47) == 2 ~ 2, TRUE ~ NA_real_)
      } else { rep(NA_real_, nrow(GYTS_2021_Zambia_National_)) }
    },

    youthgroupsdiscouragingpeople = {
      zmr68 <- ZMR_vars[["ZMR68"]]
      if(!all(is.na(zmr68))) {
        case_when(as.numeric(zmr68) == 1 ~ 1, as.numeric(zmr68) == 2 ~ 2, TRUE ~ NA_real_)
      } else { rep(NA_real_, nrow(GYTS_2021_Zambia_National_)) }
    },

    healthworkerexplaindangers = {
      zmr69 <- ZMR_vars[["ZMR69"]]
      if(!all(is.na(zmr69))) {
        case_when(as.numeric(zmr69) == 1 ~ 1, as.numeric(zmr69) == 2 ~ 2, TRUE ~ NA_real_)
      } else { rep(NA_real_, nrow(GYTS_2021_Zambia_National_)) }
    },

    churchgroupdiscouragesmoking = {
      zmr70 <- ZMR_vars[["ZMR70"]]
      if(!all(is.na(zmr70))) {
        case_when(as.numeric(zmr70) == 1 ~ 1, as.numeric(zmr70) == 2 ~ 2, TRUE ~ NA_real_)
      } else { rep(NA_real_, nrow(GYTS_2021_Zambia_National_)) }
    },

    # Media variables
    antismokingheardpastonemonth = {
      cr39 <- CR_vars[["CR39"]]
      if(!all(is.na(cr39))) {
        case_when(as.numeric(cr39) == 1 ~ 1, as.numeric(cr39) == 2 ~ 2, as.numeric(cr39) == 3 ~ 3, TRUE ~ NA_real_)
      } else { rep(NA_real_, nrow(GYTS_2021_Zambia_National_)) }
    },

    somethingwithacigarettebrand = {
      cr42 <- CR_vars[["CR42"]]
      if(!all(is.na(cr42))) {
        case_when(as.numeric(cr42) == 1 ~ 1, as.numeric(cr42) == 2 ~ 2, TRUE ~ NA_real_)
      } else { rep(NA_real_, nrow(GYTS_2021_Zambia_National_)) }
    },

    advertisingcigarettebrands = {
      cr43 <- CR_vars[["CR43"]]
      if(!all(is.na(cr43))) {
        case_when(as.numeric(cr43) == 2 ~ 2, as.numeric(cr43) == 3 ~ 3, as.numeric(cr43) == 4 ~ 4, TRUE ~ NA_real_)
      } else { rep(NA_real_, nrow(GYTS_2021_Zambia_National_)) }
    },

    cigarettesadvertsseenbillboards = {
      cr44 <- CR_vars[["CR44"]]
      if(!all(is.na(cr44))) {
        case_when(as.numeric(cr44) == 1 ~ 1, as.numeric(cr44) == 2 ~ 2, as.numeric(cr44) == 3 ~ 3, TRUE ~ NA_real_)
      } else { rep(NA_real_, nrow(GYTS_2021_Zambia_National_)) }
    },

    cigarettesadvertsseeninewspapers = {
      cr45 <- CR_vars[["CR45"]]
      if(!all(is.na(cr45))) {
        case_when(as.numeric(cr45) == 1 ~ 1, as.numeric(cr45) == 2 ~ 2, as.numeric(cr45) == 3 ~ 3, TRUE ~ NA_real_)
      } else { rep(NA_real_, nrow(GYTS_2021_Zambia_National_)) }
    },

    cigadvertsatcommunityevents = {
      cr46 <- CR_vars[["CR46"]]
      if(!all(is.na(cr46))) {
        case_when(as.numeric(cr46) == 2 ~ 2, as.numeric(cr46) == 3 ~ 3, as.numeric(cr46) == 4 ~ 4, TRUE ~ NA_real_)
      } else { rep(NA_real_, nrow(GYTS_2021_Zambia_National_)) }
    },

    antismokingmessageonradio = {
      zmr66 <- ZMR_vars[["ZMR66"]]
      if(!all(is.na(zmr66))) {
        case_when(as.numeric(zmr66) == 2 ~ 2, as.numeric(zmr66) == 3 ~ 3, as.numeric(zmr66) == 4 ~ 4, TRUE ~ NA_real_)
      } else { rep(NA_real_, nrow(GYTS_2021_Zambia_National_)) }
    },

    advertoftobaccoatshops = {
      zmr67 <- ZMR_vars[["ZMR67"]]
      if(!all(is.na(zmr67))) {
        case_when(as.numeric(zmr67) == 2 ~ 2, as.numeric(zmr67) == 3 ~ 3, as.numeric(zmr67) == 4 ~ 4, TRUE ~ NA_real_)
      } else { rep(NA_real_, nrow(GYTS_2021_Zambia_National_)) }
    },

    # School curriculum variables
    taughtdangersofsmoking = {
      cr48 <- CR_vars[["CR48"]]
      if(!all(is.na(cr48))) {
        case_when(as.numeric(cr48) == 1 ~ 1, as.numeric(cr48) == 2 ~ 2, as.numeric(cr48) == 3 ~ 3, TRUE ~ NA_real_)
      } else { rep(NA_real_, nrow(GYTS_2021_Zambia_National_)) }
    },

    reasonagegroupsmoke = {
      cr49 <- CR_vars[["CR49"]]
      if(!all(is.na(cr49))) {
        case_when(as.numeric(cr49) == 1 ~ 1, as.numeric(cr49) == 2 ~ 2, as.numeric(cr49) == 3 ~ 3, TRUE ~ NA_real_)
      } else { rep(NA_real_, nrow(GYTS_2021_Zambia_National_)) }
    },

    taughteffectsofsmoking = {
      cr50 <- CR_vars[["CR50"]]
      if(!all(is.na(cr50))) {
        case_when(as.numeric(cr50) == 1 ~ 1, as.numeric(cr50) == 2 ~ 2, as.numeric(cr50) == 3 ~ 3, TRUE ~ NA_real_)
      } else { rep(NA_real_, nrow(GYTS_2021_Zambia_National_)) }
    },

    lastdiscussiononsmokingandhealth = {
      zmr56 <- ZMR_vars[["ZMR56"]]
      if(!all(is.na(zmr56))) {
        case_when(
          as.numeric(zmr56) == 1 ~ 1, as.numeric(zmr56) == 2 ~ 2, as.numeric(zmr56) == 3 ~ 3,
          as.numeric(zmr56) == 4 ~ 4, as.numeric(zmr56) == 5 ~ 5, TRUE ~ NA_real_
        )
      } else { rep(NA_real_, nrow(GYTS_2021_Zambia_National_)) }
    },

    # Other variables
    friendsmokingcigarettes = {
      cr25 <- CR_vars[["CR25"]]
      if(!all(is.na(cr25))) {
        case_when(
          as.numeric(cr25) == 1 ~ 1, as.numeric(cr25) == 2 ~ 2, as.numeric(cr25) == 3 ~ 3,
          as.numeric(cr25) == 4 ~ 4, TRUE ~ NA_real_
        )
      } else { rep(NA_real_, nrow(GYTS_2021_Zambia_National_)) }
    },

    parentssmokingstatus = {
      cr12 <- CR_vars[["CR12"]]
      if(!all(is.na(cr12))) {
        case_when(
          as.numeric(cr12) == 1 ~ 1, as.numeric(cr12) == 2 ~ 2, as.numeric(cr12) == 3 ~ 3,
          as.numeric(cr12) == 4 ~ 4, as.numeric(cr12) == 5 ~ 5, TRUE ~ NA_real_
        )
      } else { rep(NA_real_, nrow(GYTS_2021_Zambia_National_)) }
    },

    discussingharmfuleffects = {
      cr14 <- CR_vars[["CR14"]]
      if(!all(is.na(cr14))) {
        case_when(as.numeric(cr14) == 1 ~ 1, as.numeric(cr14) == 2 ~ 2, TRUE ~ NA_real_)
      } else { rep(NA_real_, nrow(GYTS_2021_Zambia_National_)) }
    },

    SmokingArea = {
      zmr14 <- ZMR_vars[["ZMR14"]]
      if(!all(is.na(zmr14))) {
        case_when(
          as.numeric(zmr14) == 1 ~ 1, as.numeric(zmr14) == 2 ~ 2, as.numeric(zmr14) == 3 ~ 3,
          as.numeric(zmr14) == 4 ~ 4, as.numeric(zmr14) == 5 ~ 5, as.numeric(zmr14) == 6 ~ 6,
          as.numeric(zmr14) == 7 ~ 7, TRUE ~ NA_real_
        )
      } else { rep(NA_real_, nrow(GYTS_2021_Zambia_National_)) }
    },


    # Create derived variables for analysis
    age_group = factor(case_when(
      ageyrs <= 14 ~ "13-14",
      ageyrs %in% 15:16 ~ "15-16",
      ageyrs >= 17 ~ "17+",
      TRUE ~ NA_character_
    ), levels = c("13-14", "15-16", "17+")),
    gender = factor(case_when(
      sex1 == 1 ~ "Male",
      sex1 == 2 ~ "Female",
      TRUE ~ NA_character_
    ), levels = c("Female", "Male")),
    grade_level = factor(case_when(
      grade == 1 ~ "Grade 9",
      grade == 2 ~ "Grade 8",
      grade == 3 ~ "Grade 7",
      TRUE ~ NA_character_
    ), levels = c("Grade 9", "Grade 8", "Grade 7")),
    
    # Outcome variable: Smokeless tobacco use (binary outcome)
    # 1 if CR14==1 OR ELR2 >1 OR SR1==1; 0 otherwise
    smokeless_tobacco = {
      cr14 <- CR_vars[["CR14"]]
      elr2 <- ELR_vars[["ELR2"]]
      sr1 <- SR_vars[["SR1"]]
      case_when(
        (!is.na(cr14) & as.numeric(cr14) == 1) | 
        (!is.na(elr2) & as.numeric(elr2) > 1) | 
        (!is.na(sr1) & as.numeric(sr1) == 1) ~ 1,
        TRUE ~ 0
      )
    },
    
    # Variables from CR1, CR2, and ZAR3 for a priori variables
    # Age from CR1: How old are you?
    age_cr1 = {
      cr1 <- CR_vars[["CR1"]]
      if(!all(is.na(cr1))) {
        case_when(
          as.numeric(cr1) == 1 ~ 11,  # 11 years old or younger
          as.numeric(cr1) == 2 ~ 12,  # 12 years old
          as.numeric(cr1) == 3 ~ 13,  # 13 years old
          as.numeric(cr1) == 4 ~ 14,  # 14 years old
          as.numeric(cr1) == 5 ~ 15,  # 15 years old
          as.numeric(cr1) == 6 ~ 16,  # 16 years old
          as.numeric(cr1) == 7 ~ 17,  # 17 years old or older
          TRUE ~ NA_real_
        )
      } else { rep(NA_real_, nrow(GYTS_2021_Zambia_National_)) }
    },
    
    # Sex from CR2: What is your sex?
    sex_cr2 = {
      cr2 <- CR_vars[["CR2"]]
      if(!all(is.na(cr2))) {
        case_when(
          as.numeric(cr2) == 1 ~ "Male",
          as.numeric(cr2) == 2 ~ "Female",
          TRUE ~ NA_character_
        )
      } else { rep(NA_character_, nrow(GYTS_2021_Zambia_National_)) }
    },
    
    # Grade from ZAR3: In what grade/form are you?
    grade_zar3 = {
      zar3 <- ZAR_vars[["ZAR3"]]
      if(!all(is.na(zar3))) {
        case_when(
          as.numeric(zar3) == 1 ~ "Grade 7",
          as.numeric(zar3) == 2 ~ "Grade 8",
          as.numeric(zar3) == 3 ~ "Grade 9",
          TRUE ~ NA_character_
        )
      } else { rep(NA_character_, nrow(GYTS_2021_Zambia_National_)) }
    },
    
    # Additional predictor variables for analysis
    # Tried smoking: CR5 - Have you ever tried or experimented with cigarette smoking?
    tried_smoking = {
      cr5 <- CR_vars[["CR5"]]
      if(!all(is.na(cr5))) {
        case_when(
          as.numeric(cr5) == 1 ~ 1,  # Yes = tried smoking
          as.numeric(cr5) == 2 ~ 0,  # No = never tried
          TRUE ~ NA_real_
        )
      } else { rep(NA_real_, nrow(GYTS_2021_Zambia_National_)) }
    },
    
    # Parent smoking status: OR15
    parent_smoking_status = {
      or15 <- OR_vars[["OR15"]]
      if(!all(is.na(or15))) {
        case_when(
          as.numeric(or15) == 1 ~ 0,  # Don't have → No
          as.numeric(or15) == 2 ~ 1,  # About every day → Yes
          as.numeric(or15) == 3 ~ 1,  # Sometimes → Yes
          as.numeric(or15) == 4 ~ 0,  # Never → No
          TRUE ~ NA_real_
        )
      } else { rep(NA_real_, nrow(GYTS_2021_Zambia_National_)) }
    },
    
    # Smoking cigarettes 5 years from now: CR40
    smoking_5years_from_now = {
      cr40 <- CR_vars[["CR40"]]
      if(!all(is.na(cr40))) {
        case_when(
          as.numeric(cr40) == 1 ~ 0,  # Definitely not → No
          as.numeric(cr40) == 2 ~ 0,  # Probably not → No
          as.numeric(cr40) == 3 ~ 1,  # Probably yes → Yes
          as.numeric(cr40) == 4 ~ 1,  # Definitely yes → Yes
          TRUE ~ NA_real_
        )
      } else { rep(NA_real_, nrow(GYTS_2021_Zambia_National_)) }
    },
    
    # Anti-smoking media messages: CR30 or CR31
    antismoking_media = {
      cr30 <- CR_vars[["CR30"]]
      cr31 <- CR_vars[["CR31"]]
      case_when(
        (!is.na(cr30) & as.numeric(cr30) == 1) | 
        (!is.na(cr31) & as.numeric(cr31) == 1) ~ 1,
        (!is.na(cr30) & as.numeric(cr30) != 1) & 
        (!is.na(cr31) & as.numeric(cr31) != 1) ~ 0,
        (!is.na(cr30) & as.numeric(cr30) != 1) & is.na(cr31) ~ 0,
        is.na(cr30) & (!is.na(cr31) & as.numeric(cr31) != 1) ~ 0,
        TRUE ~ NA_real_
      )
    },
    
    # Taught dangers of smoking: CR33
    taught_dangers_smoking = {
      cr33 <- CR_vars[["CR33"]]
      if(!all(is.na(cr33))) {
        case_when(
          as.numeric(cr33) == 1 ~ 1,  # Yes
          as.numeric(cr33) == 2 ~ 0,  # No
          as.numeric(cr33) == 3 ~ 0,  # I don't know → recoded to No
          TRUE ~ NA_real_
        )
      } else { rep(NA_real_, nrow(GYTS_2021_Zambia_National_)) }
    },
    
    # Teacher smoking at school: OR60
    teacher_smoking_school = {
      or60 <- OR_vars[["OR60"]]
      if(!all(is.na(or60))) {
        case_when(
          as.numeric(or60) == 1 ~ 1,  # About every day → Yes
          as.numeric(or60) == 2 ~ 1,  # Sometimes → Yes
          as.numeric(or60) == 3 ~ 0,  # Never → No
          as.numeric(or60) == 4 ~ 0,  # Don't know → No
          TRUE ~ NA_real_
        )
      } else { rep(NA_real_, nrow(GYTS_2021_Zambia_National_)) }
    },
    
    # Having friends that smoke: ZAR55
    havingfriendsthatsmoke = {
      zar55 <- ZAR_vars[["ZAR55"]]
      if(!all(is.na(zar55))) {
        case_when(
          as.numeric(zar55) == 1 ~ 0,  # None of them → No
          as.numeric(zar55) == 2 ~ 1,  # Some of them → Yes
          as.numeric(zar55) == 3 ~ 1,  # Most of them → Yes
          as.numeric(zar55) == 4 ~ 1,  # All of them → Yes
          as.numeric(zar55) == 5 ~ 0,  # I don't know → No
          TRUE ~ NA_real_
        )
      } else { rep(NA_real_, nrow(GYTS_2021_Zambia_National_)) }
    },
    
    # Additional predictor variables from paper 8
    zar4_raw = {
      zar4 <- ZAR_vars[["ZAR4"]]
      if(!all(is.na(zar4))) {
        as.numeric(zar4)
      } else {
        rep(NA_real_, nrow(GYTS_2021_Zambia_National_))
      }
    },
    handrolled_cigarette = {
      or5 <- OR_vars[["OR5"]]
      if(!all(is.na(or5))) {
        case_when(as.numeric(or5) == 1 ~ 0, TRUE ~ 1)
      } else { rep(NA_real_, nrow(GYTS_2021_Zambia_National_)) }
    },
    smoked_cigarettes = {
      cr8 <- CR_vars[["CR8"]]
      if(!all(is.na(cr8))) {
        case_when(as.numeric(cr8) == 1 ~ 0, as.numeric(cr8) > 1 ~ 1, TRUE ~ NA_real_)
      } else { rep(NA_real_, nrow(GYTS_2021_Zambia_National_)) }
    },
    tried_shisha = {
      sr1 <- SR_vars[["SR1"]]
      if(!all(is.na(sr1))) {
        case_when(as.numeric(sr1) == 1 ~ 0, as.numeric(sr1) == 2 ~ 1, TRUE ~ NA_real_)
      } else { rep(NA_real_, nrow(GYTS_2021_Zambia_National_)) }
    },
    heard_electronic_cigarette = {
      elr1 <- ELR_vars[["ELR1"]]
      if(!all(is.na(elr1))) {
        case_when(as.numeric(elr1) == 2 ~ 0, as.numeric(elr1) == 1 ~ 1, TRUE ~ NA_real_)
      } else { rep(NA_real_, nrow(GYTS_2021_Zambia_National_)) }
    },
    father_smokes = {
      or15 <- OR_vars[["OR15"]]
      if(!all(is.na(or15))) {
        case_when(as.numeric(or15) == 1 ~ 0, as.numeric(or15) == 2 ~ 1, as.numeric(or15) == 3 ~ 1, as.numeric(or15) == 4 ~ 0, TRUE ~ NA_real_)
      } else { rep(NA_real_, nrow(GYTS_2021_Zambia_National_)) }
    },
    people_smoke_school = {
      cr22 <- CR_vars[["CR22"]]
      if(!all(is.na(cr22))) {
        case_when(as.numeric(cr22) == 1 ~ 1, TRUE ~ 0)
      } else { rep(NA_real_, nrow(GYTS_2021_Zambia_National_)) }
    },
    teachers_smoke_school = {
      or60 <- OR_vars[["OR60"]]
      if(!all(is.na(or60))) {
        case_when(as.numeric(or60) > 2 ~ 0, TRUE ~ 1)
      } else { rep(NA_real_, nrow(GYTS_2021_Zambia_National_)) }
    },
    see_tobacco_tv = {
      cr34 <- CR_vars[["CR34"]]
      if(!all(is.na(cr34))) {
        case_when(as.numeric(cr34) == 2 ~ 1, TRUE ~ 0)
      } else { rep(NA_real_, nrow(GYTS_2021_Zambia_National_)) }
    },
    see_tobacco_advert = {
      cr35 <- CR_vars[["CR35"]]
      if(!all(is.na(cr35))) {
        case_when(as.numeric(cr35) == 2 ~ 1, TRUE ~ 0)
      } else { rep(NA_real_, nrow(GYTS_2021_Zambia_National_)) }
    },
    tobacco_causes_cancer = {
      zar64 <- ZAR_vars[["ZAR64"]]
      if(!all(is.na(zar64))) {
        case_when(as.numeric(zar64) == 1 ~ 1, TRUE ~ 0)
      } else { rep(NA_real_, nrow(GYTS_2021_Zambia_National_)) }
    },
    tobacco_leads_drug_use = {
      zar65 <- ZAR_vars[["ZAR65"]]
      if(!all(is.na(zar65))) {
        case_when(as.numeric(zar65) == 1 ~ 1, TRUE ~ 0)
      } else { rep(NA_real_, nrow(GYTS_2021_Zambia_National_)) }
    },
    own_item_tobacco = {
      zar53 <- ZAR_vars[["ZAR53"]]
      if(!all(is.na(zar53))) {
        case_when(as.numeric(zar53) == 1 ~ 1, TRUE ~ 0)
      } else { rep(NA_real_, nrow(GYTS_2021_Zambia_National_)) }
    },
    
    # Human-readable factor versions for predictor variables
    tried_smoking_f = factor(tried_smoking, levels = c(0, 1), labels = c("No", "Yes")),
    parent_smoking_status_f = factor(parent_smoking_status, levels = c(0, 1), labels = c("No", "Yes")),
    smoking_5years_from_now_f = factor(smoking_5years_from_now, levels = c(0, 1), labels = c("No", "Yes")),
    antismoking_media_f = factor(antismoking_media, levels = c(0, 1), labels = c("No", "Yes")),
    taught_dangers_smoking_f = factor(taught_dangers_smoking, levels = c(0, 1), labels = c("No", "Yes")),
    teacher_smoking_school_f = factor(teacher_smoking_school, levels = c(0, 1), labels = c("No", "Yes")),
    havingfriendsthatsmoke_f = factor(havingfriendsthatsmoke, levels = c(0, 1), labels = c("No", "Yes")),
    handrolled_cigarette_f = factor(handrolled_cigarette, levels = c(0, 1), labels = c("No", "Yes")),
    smoked_cigarettes_f = factor(smoked_cigarettes, levels = c(0, 1), labels = c("No", "Yes")),
    tried_shisha_f = factor(tried_shisha, levels = c(0, 1), labels = c("No", "Yes")),
    heard_electronic_cigarette_f = factor(heard_electronic_cigarette, levels = c(0, 1), labels = c("No", "Yes")),
    father_smokes_f = factor(father_smokes, levels = c(0, 1), labels = c("No", "Yes")),
    people_smoke_school_f = factor(people_smoke_school, levels = c(0, 1), labels = c("No", "Yes")),
    teachers_smoke_school_f = factor(teachers_smoke_school, levels = c(0, 1), labels = c("No", "Yes")),
    see_tobacco_tv_f = factor(see_tobacco_tv, levels = c(0, 1), labels = c("No", "Yes")),
    see_tobacco_advert_f = factor(see_tobacco_advert, levels = c(0, 1), labels = c("No", "Yes")),
    tobacco_causes_cancer_f = factor(tobacco_causes_cancer, levels = c(0, 1), labels = c("No", "Yes")),
    tobacco_leads_drug_use_f = factor(tobacco_leads_drug_use, levels = c(0, 1), labels = c("No", "Yes")),
    own_item_tobacco_f = factor(own_item_tobacco, levels = c(0, 1), labels = c("No", "Yes")),
    
    # For backward compatibility
    current_smokeless = smokeless_tobacco,
    dual_use = case_when(smokeless_tobacco == 1 & current_cigarette == 1 ~ 1, TRUE ~ 0)
  )

# Create wealth index (after all variables are created)
# Wealth index: three groups (low, middle, rich) based on ranks of ZAR4
df <- df %>%
  mutate(
    wealth_index = case_when(
      !is.na(zar4_raw) ~ {
        # Create ranks (handling ties with average ranks)
        zar4_ranks <- rank(zar4_raw, na.last = "keep", ties.method = "average")
        # Calculate tertiles based on ranks
        rank_tertiles <- quantile(zar4_ranks, probs = c(0, 1/3, 2/3, 1), na.rm = TRUE)
        case_when(
          zar4_ranks <= rank_tertiles[2] ~ "Low",
          zar4_ranks <= rank_tertiles[3] ~ "Middle",
          TRUE ~ "Rich"
        )
      },
      TRUE ~ NA_character_
    ),
    wealth_index_f = factor(wealth_index, levels = c("Low", "Middle", "Rich"), labels = c("Low", "Middle", "Rich"))
  )

# Create survey design object with Stratum and PSU
# Check for Stratum, PSU, and weight variables
stratum_var <- if("Stratum" %in% names(df)) "Stratum" else NULL
psu_var <- if("PSU" %in% names(df)) "PSU" else NULL
weight_var <- if("FinalWgt" %in% names(df)) "FinalWgt" else if("'FinalWgt" %in% names(df)) "'FinalWgt" else if("WEIGHT" %in% names(df)) "WEIGHT" else NULL

if(!is.null(stratum_var) && !is.null(psu_var) && !is.null(weight_var)) {
  # Two-stage design with Stratum and PSU
  # nest = TRUE assumes clusters are nested within strata
  svy_design <- svydesign(
    ids = as.formula(paste0("~", psu_var)),
    strata = as.formula(paste0("~", stratum_var)),
    weights = as.formula(paste0("~", weight_var)),
    data = df,
    nest = TRUE
  )
} else if(!is.null(psu_var) && !is.null(weight_var)) {
  # Design with PSU only (no stratification)
  svy_design <- svydesign(
    ids = as.formula(paste0("~", psu_var)),
    weights = as.formula(paste0("~", weight_var)),
    data = df
  )
} else if(!is.null(weight_var)) {
  # Simple design with weights only
  svy_design <- svydesign(
    ids = ~1,
    weights = as.formula(paste0("~", weight_var)),
    data = df
  )
} else {
  svy_design <- svydesign(ids = ~1, weights = ~1, data = df)
  warning("Stratum, PSU, or weight variable not found. Using unweighted design.")
}
```

# Introduction

Smokeless tobacco products represent a diverse category of tobacco consumption that includes chewing tobacco, snuff, and other forms of tobacco that are used without combustion [@who_tobacco_2021]. While often perceived as less harmful than cigarette smoking, smokeless tobacco use poses significant health risks, including oral cancer, cardiovascular disease, and nicotine dependence [@usdhhs_preventing_2012]. Globally, smokeless tobacco use is particularly prevalent in certain regions, including parts of Africa, South Asia, and Southeast Asia, where traditional forms of smokeless tobacco have deep cultural roots [@who_global_2020].

In Zambia, as in many African countries, data on smokeless tobacco use patterns and determinants among adolescents remain limited, creating gaps in understanding the current landscape of youth smokeless tobacco use [@zyambo_gyts_2021]. The 2021 Global Youth Tobacco Survey (GYTS) provides a valuable opportunity to examine current smokeless tobacco use prevalence and associated factors among school-going adolescents in Zambia, offering insights that can inform evidence-based prevention and control strategies.

This analysis aims to examine the prevalence of current smokeless tobacco use among adolescents in Zambia and identify key factors associated with smokeless tobacco use. Specifically, we seek to: (1) describe the prevalence of current smokeless tobacco use across different demographic and socioeconomic groups; (2) identify individual, social, and environmental factors associated with current smokeless tobacco use; and (3) assess the relationship between tobacco-growing regions and smokeless tobacco use patterns among adolescents. The findings from this analysis have important policy implications for strengthening tobacco control efforts in Zambia, particularly in addressing all forms of tobacco use, not just cigarettes.

# Methods

## Data Source
- **Survey**: Global Youth Tobacco Survey (GYTS) 2021, Zambia
- **Analysis Dataset**: GYTS 2021 Zambia (National).XLS, sheet "ZAMBIA_DATA_21"

## Data Management

All variables used in this analysis were derived from the GYTS 2021 dataset. The outcome variable for this analysis is smokeless tobacco use, defined as a binary variable where 1 = current smokeless tobacco use (CR14==1 OR ELR2 >1 OR SR1==1) and 0 = no current smokeless tobacco use. All predictor variables were created following the same methodology as described in Paper 1, ensuring consistency across analyses.

### Variables Included in Analysis

The following table provides definitions and value labels for all variables included in this analysis:

```{r variables-table, echo=FALSE, results='asis'}
# Create comprehensive variables table
variables_data <- data.frame(
  Variable = c(
    "smokeless_tobacco",
    "age_cr1",
    "sex_cr2",
    "grade_zar3",
    "RegionName",
    "tried_smoking_f",
    "parent_smoking_status_f",
    "smoking_5years_from_now_f",
    "antismoking_media_f",
    "taught_dangers_smoking_f",
    "teacher_smoking_school_f",
    "havingfriendsthatsmoke_f"
  ),
  Definition = c(
    "Current smokeless tobacco use status (outcome variable)",
    "Age in years (factor: 11, 12, 13, 14, 15, 16, 17)",
    "Sex of participant",
    "Grade level in school",
    "Geographic region",
    "Ever tried cigarette smoking",
    "Parent or guardian smokes in home",
    "Intention to use tobacco in next 5 years",
    "Exposure to anti-tobacco media messages",
    "Taught about dangers of tobacco use in school",
    "Saw teachers smoking at school",
    "Friends smoke tobacco"
  ),
  Source_Question = c(
    "CR14==1 OR ELR2 >1 OR SR1==1",
    "CR1: How old are you?",
    "CR2: What is your sex?",
    "ZAR3: In what grade/form are you?",
    "Derived from Stratum variable",
    "CR5: Have you ever tried or experimented with cigarette smoking?",
    "OR15: How often do you see your father (stepfather or mother's partner) smoking in your home?",
    "CR40: At any time during the next 12 months do you think you will use any form of tobacco?",
    "CR30/CR31: During the past 30 days, how often did you see or hear anti-tobacco media messages?",
    "CR33: During the past 12 months, were you taught in any of your classes about the dangers of tobacco use?",
    "OR60: During school hours, how often do you see teachers smoking outdoors on school premises?",
    "ZAR55: Do any of your closest friends smoke tobacco?"
  ),
  Value_Labels = c(
    "0 = No current use, 1 = Current smokeless tobacco use",
    "11, 12, 13, 14, 15, 16, 17 (years)",
    "Male, Female",
    "Grade 7, Grade 8, Grade 9",
    "Lusaka, ROC (Rest of Country), Tobacco Regions",
    "No = Never tried, Yes = Ever tried",
    "No = No parent/guardian smokes, Yes = At least one parent/guardian smokes",
    "No = Definitely not/Probably not, Yes = Definitely yes/Probably yes",
    "No = Low/No exposure, Yes = High exposure",
    "No = Not taught, Yes = Taught",
    "No = Did not see, Yes = Saw teachers smoking",
    "No = No friends smoke, Yes = Some/Most/All friends smoke"
  ),
  Variable_Type = c(
    "Outcome (binary)",
    "A priori (factor)",
    "A priori (categorical)",
    "A priori (categorical)",
    "A priori (categorical)",
    "Predictor (binary)",
    "Predictor (binary)",
    "Predictor (binary)",
    "Predictor (binary)",
    "Predictor (binary)",
    "Predictor (binary)",
    "Predictor (binary)"
  ),
  stringsAsFactors = FALSE
)

# Create formatted table using flextable
variables_table <- variables_data %>%
  flextable() %>%
  set_header_labels(
    Variable = "**Variable Name**",
    Definition = "**Definition**",
    Source_Question = "**Source Question**",
    Value_Labels = "**Value Labels**",
    Variable_Type = "**Type**"
  ) %>%
  bold(part = "header") %>%
  bold(j = 1, part = "body") %>%
  align(align = "left", part = "all") %>%
  width(j = "Variable", width = 1.2) %>%
  width(j = "Definition", width = 2.0) %>%
  width(j = "Source_Question", width = 2.5) %>%
  width(j = "Value_Labels", width = 2.0) %>%
  width(j = "Variable_Type", width = 1.0) %>%
  theme_booktabs() %>%
  fontsize(size = 9, part = "all") %>%
  padding(padding = 4, part = "all") %>%
  add_footer_lines("Variables derived from GYTS 2021 Zambia National dataset. Outcome variable: smokeless_tobacco (1 if CR14==1 OR ELR2 >1 OR SR1==1; 0 otherwise). A priori variables (age_cr1, sex_cr2, grade_zar3, RegionName) were included in all adjusted models regardless of statistical significance.") %>%
  fontsize(size = 8, part = "footer") %>%
  italic(part = "footer") %>%
  align(align = "left", part = "footer")

variables_table
```

### Variable Creation and Recoding

**Outcome Variable:**
- `smokeless_tobacco`: Created from CR14, ELR2, and SR1. Binary outcome: 1 = Current smokeless tobacco use (CR14==1 OR ELR2 >1 OR SR1==1), 0 = No current smokeless tobacco use.

**A Priori Variables:**
- `age_cr1`: Derived from CR1 (How old are you?) with responses 1-7 mapped to ages 11-17 years. Used as a factor variable in models.
- `sex_cr2`: Derived from CR2 (What is your sex?) with responses: 1 = Male, 2 = Female.
- `grade_zar3`: Derived from ZAR3 (In what grade/form are you?) with responses: 1 = Grade 7, 2 = Grade 8, 3 = Grade 9.
- `RegionName`: Created from the Stratum variable, categorized into three regions: Lusaka, ROC (Rest of Country), and Tobacco Regions.

**Predictor Variables:**
All predictor variables were recoded as binary factors (No/Yes) for consistency in analysis:
- `tried_smoking_f`: Ever tried cigarette smoking (from CR5)
- `parent_smoking_status_f`: Parent/guardian smokes in home (from OR15)
- `smoking_5years_from_now_f`: Intention to use tobacco in next 5 years (from CR40)
- `antismoking_media_f`: Exposure to anti-tobacco media messages (from CR30/CR31)
- `taught_dangers_smoking_f`: Taught about dangers of tobacco use (from CR33)
- `teacher_smoking_school_f`: Saw teachers smoking at school (from OR60)
- `havingfriendsthatsmoke_f`: Friends smoke tobacco (from ZAR55)

**Missing Data:**
Observations with missing values for the outcome variable (smokeless_tobacco) were excluded from analysis. For predictor variables, missing values were handled through listwise deletion in regression models. All analyses accounted for the complex survey design using survey weights, stratification, and clustering.

## Statistical Analysis
- Weighted prevalence estimates with 95% confidence intervals
- Crude (unadjusted) logistic regression models for each variable
- Adjusted logistic regression models using stepwise selection (AIC-based)
- Age (age_cr1), sex (sex_cr2), grade (grade_zar3), and region included a priori in all adjusted models
- Survey-weighted analysis accounting for complex survey design (Stratum and PSU)
- Sensitivity analyses to assess robustness of findings

# Results

## Table 1: Sample Characteristics

```{r table1-characteristics, echo=FALSE, results='asis'}
# Create nice characteristics table using gtsummary
# Prepare data for characteristics table
df_char <- df %>%
  dplyr::filter(!is.na(RegionName)) %>%
  dplyr::mutate(
    RegionName = factor(RegionName, levels = c("Lusaka", "ROC", "Tobacco Regions")),
    age_group = factor(age_group, levels = c("13-14", "15-16", "17+")),
    gender = factor(gender, levels = c("Female", "Male")),
    grade_level = factor(grade_level, levels = c("Grade 9", "Grade 8", "Grade 7")),
    smokeless_tobacco = factor(smokeless_tobacco, levels = c(0, 1), labels = c("No", "Yes")),
    # Variables from CR1, CR2, ZAR3
    age_cr1 = age_cr1,
    sex_cr2 = factor(sex_cr2, levels = c("Female", "Male")),
    grade_zar3 = factor(grade_zar3, levels = c("Grade 7", "Grade 8", "Grade 9")),
    # Ensure new variables are factors with human-readable labels
    tried_smoking_f = factor(tried_smoking_f, levels = c("No", "Yes"), labels = c("No", "Yes")),
    parent_smoking_status_f = factor(parent_smoking_status_f, levels = c("No", "Yes"), labels = c("No", "Yes")),
    smoking_5years_from_now_f = factor(smoking_5years_from_now_f, levels = c("No", "Yes"), labels = c("No", "Yes")),
    antismoking_media_f = factor(antismoking_media_f, levels = c("No", "Yes"), labels = c("No", "Yes")),
    taught_dangers_smoking_f = factor(taught_dangers_smoking_f, levels = c("No", "Yes"), labels = c("No", "Yes")),
    teacher_smoking_school_f = factor(teacher_smoking_school_f, levels = c("No", "Yes"), labels = c("No", "Yes")),
    havingfriendsthatsmoke_f = factor(havingfriendsthatsmoke_f, levels = c("No", "Yes"), labels = c("No", "Yes"))
  )

# Check if we have data and sufficient RegionName levels
if(nrow(df_char) == 0) {
  cat("No data available for Table 1 after filtering.\n")
} else if(length(unique(df_char$RegionName[!is.na(df_char$RegionName)])) < 2) {
  cat("Insufficient variation in RegionName variable for Table 1.\n")
} else {
  # Select only variables that exist and have data
  vars_to_include <- c()
  if("age_cr1" %in% names(df_char) && sum(!is.na(df_char$age_cr1)) > 0) vars_to_include <- c(vars_to_include, "age_cr1")
  if("sex_cr2" %in% names(df_char) && sum(!is.na(df_char$sex_cr2)) > 0) vars_to_include <- c(vars_to_include, "sex_cr2")
  if("grade_zar3" %in% names(df_char) && sum(!is.na(df_char$grade_zar3)) > 0) vars_to_include <- c(vars_to_include, "grade_zar3")
  if("age_group" %in% names(df_char) && sum(!is.na(df_char$age_group)) > 0) vars_to_include <- c(vars_to_include, "age_group")
  if("gender" %in% names(df_char) && sum(!is.na(df_char$gender)) > 0) vars_to_include <- c(vars_to_include, "gender")
  if("grade_level" %in% names(df_char) && sum(!is.na(df_char$grade_level)) > 0) vars_to_include <- c(vars_to_include, "grade_level")
  if("tried_smoking_f" %in% names(df_char) && sum(!is.na(df_char$tried_smoking_f)) > 0) vars_to_include <- c(vars_to_include, "tried_smoking_f")
  if("parent_smoking_status_f" %in% names(df_char) && sum(!is.na(df_char$parent_smoking_status_f)) > 0) vars_to_include <- c(vars_to_include, "parent_smoking_status_f")
  if("smoking_5years_from_now_f" %in% names(df_char) && sum(!is.na(df_char$smoking_5years_from_now_f)) > 0) vars_to_include <- c(vars_to_include, "smoking_5years_from_now_f")
  if("antismoking_media_f" %in% names(df_char) && sum(!is.na(df_char$antismoking_media_f)) > 0) vars_to_include <- c(vars_to_include, "antismoking_media_f")
  if("taught_dangers_smoking_f" %in% names(df_char) && sum(!is.na(df_char$taught_dangers_smoking_f)) > 0) vars_to_include <- c(vars_to_include, "taught_dangers_smoking_f")
  if("teacher_smoking_school_f" %in% names(df_char) && sum(!is.na(df_char$teacher_smoking_school_f)) > 0) vars_to_include <- c(vars_to_include, "teacher_smoking_school_f")
  if("havingfriendsthatsmoke_f" %in% names(df_char) && sum(!is.na(df_char$havingfriendsthatsmoke_f)) > 0) vars_to_include <- c(vars_to_include, "havingfriendsthatsmoke_f")
  
  if(length(vars_to_include) == 0) {
    cat("No valid variables available for Table 1.\n")
  } else {
    tryCatch({
      df_table1 <- df_char %>% dplyr::select(dplyr::all_of(c(vars_to_include, "RegionName")))
      
      all_labels <- list(
        age_cr1 = "Age, years",
        age_group = "Age group",
        sex_cr2 = "Sex",
        gender = "Sex",
        grade_zar3 = "Grade",
        grade_level = "Grade level",
        tried_smoking_f = "Ever tried cigarette smoking",
        parent_smoking_status_f = "Parent/guardian smokes in home",
        smoking_5years_from_now_f = "Intention to use tobacco in next 5 years",
        antismoking_media_f = "Exposure to anti-tobacco media messages",
        taught_dangers_smoking_f = "Taught about dangers of tobacco use",
        teacher_smoking_school_f = "Saw teachers smoking at school",
        havingfriendsthatsmoke_f = "Friends smoke tobacco"
      )
      
      label_list <- all_labels[names(all_labels) %in% vars_to_include]
      
      if(length(label_list) > 0) {
        table1_char <- df_table1 %>%
          tbl_summary(
            by = RegionName,
            statistic = list(
              all_continuous() ~ "{mean} ({sd})",
              all_categorical() ~ "{n} ({p}%)"
            ),
            percent = "column",
            missing = "no",
            label = label_list
          )
      } else {
        table1_char <- df_table1 %>%
          tbl_summary(
            by = RegionName,
            statistic = list(
              all_continuous() ~ "{mean} ({sd})",
              all_categorical() ~ "{n} ({p}%)"
            ),
            percent = "column",
            missing = "no"
          )
      }
      
      table1_char <- table1_char %>%
        add_overall(last = TRUE, col_label = "**Overall**") %>%
        add_p(
          test = list(
            all_categorical() ~ "fisher.test",
            all_continuous() ~ "kruskal.test"
          ),
          pvalue_fun = function(x) style_pvalue(x, digits = 3)
        ) %>%
        bold_labels() %>%
        bold_p() %>%
        modify_header(
          label = "**Characteristic**",
          p.value = "**P-value**"
        ) %>%
        modify_spanning_header(starts_with("stat_") ~ "**Region**") %>%
        modify_footnote(
          all_stat_cols() ~ "Column percentages shown. Values are n (%) for categorical variables and mean (SD) for continuous variables.",
          p.value ~ "Fisher's exact test for categorical variables; Kruskal-Wallis test for continuous variables."
        )
      
      table1_char
    }, error = function(e) {
      cat("Error creating Table 1:", e$message, "\n")
      NULL
    })
  }
}
```

### Characteristics

The study sample comprised school-going adolescents from Zambia, with data collected as part of the 2021 Global Youth Tobacco Survey (Table 1). Overall, the sample was distributed across three major regions: Lusaka, the Rest of Country (ROC), and Tobacco-Growing Regions, with proportions shown in Table 1. The sample included representation across the adolescent age spectrum, with mean age and standard deviation presented in Table 1.

**Demographic Analysis**

Demographic characteristics varied across regions, reflecting the diverse geographic and socioeconomic contexts within Zambia (Table 1). Age distribution, presented as mean (SD) in Table 1, showed variation across regions, with regional differences assessed using Kruskal-Wallis tests. Sex distribution was examined across regions, with proportions for male and female participants shown in Table 1. Regional variations in sex distribution were evaluated using Fisher's exact tests, with p-values presented in Table 1. Grade level distribution indicated that participants were enrolled across Grades 7, 8, and 9, with proportions for each grade level shown by region in Table 1.

**Predictor Variables**

Several key predictor variables demonstrated important patterns across regions (Table 1). The proportion of adolescents who had ever tried cigarette smoking varied by region, with overall and regional-specific percentages shown in Table 1. Parental smoking status, a critical social determinant, showed regional variation in the proportion of adolescents reporting a parent or guardian who smokes in the home (Table 1). Intention to use tobacco in the next five years was reported by varying proportions of participants across regions, as detailed in Table 1. Exposure to anti-tobacco media messages showed regional patterns, with proportions presented in Table 1. The proportion of adolescents who reported being taught about the dangers of tobacco use in school varied across regions, as shown in Table 1. Teacher smoking behavior at school was observed by varying proportions of participants across regions (Table 1), and the proportion reporting friends who smoke tobacco also showed regional variation (Table 1).

## Table 2: Prevalence of Smokeless Tobacco Use (smokeless_tobacco) by Key Variables

```{r table2-prevalence, echo=FALSE, results='asis'}
# Function to calculate prevalence by a grouping variable
calc_prevalence_by_var <- function(outcome_var, group_var, design_obj, data_df) {
  tryCatch({
    design_subset <- subset(design_obj, !is.na(.data[[outcome_var]]) & !is.na(.data[[group_var]]))
    
  svy_result <- svyby(
      as.formula(paste0("~", outcome_var)),
      as.formula(paste0("~", group_var)),
      design_subset,
    svymean,
    na.rm = TRUE,
    vartype = "ci",
    keep.var = FALSE
  )
  
  result <- data.frame(svy_result, stringsAsFactors = FALSE)
  
    outcome_col <- NULL
    if(outcome_var %in% names(result)) {
      outcome_col <- outcome_var
    } else {
      possible_cols <- grep(paste0("^", outcome_var), names(result), value = TRUE)
      if(length(possible_cols) > 0) {
        outcome_col <- possible_cols[1]
      } else {
        numeric_cols <- names(result)[sapply(result, is.numeric)]
        numeric_cols <- numeric_cols[numeric_cols != group_var]
        if(length(numeric_cols) > 0) {
          outcome_col <- numeric_cols[1]
        }
      }
    }
    
    if(is.null(outcome_col)) {
      warning(paste("Could not find outcome column for", outcome_var, "in svyby result"))
      return(data.frame())
    }
    
    result$prevalence <- as.numeric(unlist(result[[outcome_col]]))
    
    ci_cols <- grep("^ci_l|^se", names(result), value = TRUE, ignore.case = TRUE)
    if(length(ci_cols) >= 2) {
      result$ci_l <- as.numeric(unlist(result[[ci_cols[1]]]))
      result$ci_u <- as.numeric(unlist(result[[ci_cols[2]]]))
    } else {
      se_col <- grep("^se", names(result), value = TRUE, ignore.case = TRUE)
      if(length(se_col) > 0) {
        se_val <- as.numeric(unlist(result[[se_col[1]]]))
        result$ci_l <- result$prevalence - 1.96 * se_val
        result$ci_u <- result$prevalence + 1.96 * se_val
      } else {
        result$ci_l <- NA_real_
        result$ci_u <- NA_real_
      }
    }
    
    n_counts <- data_df %>%
      dplyr::filter(!is.na(.data[[outcome_var]]), !is.na(.data[[group_var]])) %>%
      dplyr::count(.data[[group_var]], name = "n")
    
    if(group_var %in% names(result)) {
      grouping_col <- group_var
    } else {
      row_names <- rownames(result)
      if(!is.null(row_names) && length(row_names) == nrow(result) && 
         any(row_names != as.character(1:nrow(result)))) {
        result[[group_var]] <- row_names
        grouping_col <- group_var
      } else {
        non_outcome_cols <- names(result)[!names(result) %in% c(outcome_col, "prevalence", "ci_l", "ci_u")]
        if(length(non_outcome_cols) > 0) {
          grouping_col <- non_outcome_cols[1]
          names(result)[names(result) == grouping_col] <- group_var
          grouping_col <- group_var
        } else {
          grouping_col <- NULL
        }
      }
    }
    
    if(!is.null(grouping_col) && grouping_col %in% names(result)) {
      result[[grouping_col]] <- as.character(result[[grouping_col]])
      n_counts[[group_var]] <- as.character(n_counts[[group_var]])
      
      result <- result %>%
        dplyr::left_join(n_counts, by = group_var) %>%
        dplyr::select(dplyr::all_of(c(group_var, "prevalence", "ci_l", "ci_u", "n")))
    } else {
      result <- result %>%
        dplyr::mutate(n = NA_integer_) %>%
        dplyr::select(dplyr::all_of(c(group_var, "prevalence", "ci_l", "ci_u", "n")))
    }
    
    return(result)
  }, error = function(e) {
    warning(paste("Error calculating prevalence by", group_var, ":", e$message))
    return(data.frame())
  })
}

# Calculate for smokeless_tobacco
outcome_var <- "smokeless_tobacco"

# List of variables to show prevalence by
prev_vars <- list(
  list(var = "RegionName", label = "Region"),
  list(var = "age_cr1", label = "Age, years"),
  list(var = "sex_cr2", label = "Sex"),
  list(var = "grade_zar3", label = "Grade"),
  list(var = "age_group", label = "Age group"),
  list(var = "gender", label = "Sex"),
  list(var = "grade_level", label = "Grade level"),
  list(var = "tried_smoking_f", label = "Ever tried cigarette smoking"),
  list(var = "parent_smoking_status_f", label = "Parent/guardian smokes in home"),
  list(var = "smoking_5years_from_now_f", label = "Intention to use tobacco in next 5 years"),
  list(var = "antismoking_media_f", label = "Exposure to anti-tobacco media messages"),
  list(var = "taught_dangers_smoking_f", label = "Taught about dangers of tobacco use"),
  list(var = "teacher_smoking_school_f", label = "Saw teachers smoking at school"),
  list(var = "havingfriendsthatsmoke_f", label = "Friends smoke tobacco")
)

# Calculate prevalence (mean) by each variable and category
table2_list <- list()

for(var_info in prev_vars) {
  var_name <- var_info$var
  var_label <- var_info$label
  
  if(var_name %in% names(df)) {
    df_subset <- df %>%
      dplyr::filter(!is.na(.data[[var_name]]), !is.na(.data[[outcome_var]]))
    
    if(nrow(df_subset) > 0) {
      var_levels <- unique(df_subset[[var_name]])
      var_levels <- var_levels[!is.na(var_levels)]
      
      if(length(var_levels) > 0) {
        level_results <- list()
        
        for(level in var_levels) {
          df_level <- df_subset %>%
            dplyr::filter(.data[[var_name]] == level)
          
          if(nrow(df_level) > 0) {
            tryCatch({
              design_level <- subset(svy_design, .data[[var_name]] == level & !is.na(.data[[outcome_var]]))
              
              svy_mean <- svymean(as.formula(paste0("~", outcome_var)), design_level, na.rm = TRUE)
              
              mean_val <- as.numeric(svy_mean)
              se_val <- sqrt(as.numeric(attr(svy_mean, "var")))
              
              ci_l <- mean_val - 1.96 * se_val
              ci_u <- mean_val + 1.96 * se_val
              
              n_count <- nrow(df_level)
              
              level_results[[as.character(level)]] <- data.frame(
                Category = var_label,
                Level = as.character(level),
                prevalence = mean_val,
                ci_l = ci_l,
                ci_u = ci_u,
                n = n_count,
                stringsAsFactors = FALSE
              )
            }, error = function(e) {
              mean_val <- mean(df_level[[outcome_var]], na.rm = TRUE)
              n_count <- nrow(df_level)
              se_val <- sd(df_level[[outcome_var]], na.rm = TRUE) / sqrt(n_count)
              ci_l <- mean_val - 1.96 * se_val
              ci_u <- mean_val + 1.96 * se_val
              
              level_results[[as.character(level)]] <<- data.frame(
                Category = var_label,
                Level = as.character(level),
                prevalence = mean_val,
                ci_l = max(0, ci_l),
                ci_u = min(1, ci_u),
                n = n_count,
                stringsAsFactors = FALSE
              )
            })
          }
        }
        
        if(length(level_results) > 0) {
          var_result <- dplyr::bind_rows(level_results)
          table2_list[[var_label]] <- var_result
        }
      }
    }
  }
}

# Combine all results
if(length(table2_list) > 0) {
  table2_data <- dplyr::bind_rows(table2_list) %>%
  dplyr::mutate(
      Prevalence = sprintf("%.2f%%", prevalence * 100),
      `95% CI` = sprintf("(%.2f%%, %.2f%%)", ci_l * 100, ci_u * 100),
      `Prevalence (95% CI)` = paste0(Prevalence, " ", `95% CI`),
      Level = case_when(
        Level == "Male" ~ "Male",
        Level == "Female" ~ "Female",
        Level == "Yes" ~ "Yes",
        Level == "No" ~ "No",
        Level == "13-14" ~ "13-14 years",
        Level == "15-16" ~ "15-16 years",
        Level == "17+" ~ "17+ years",
        Level == "Grade 7" ~ "Grade 7",
        Level == "Grade 8" ~ "Grade 8",
        Level == "Grade 9" ~ "Grade 9",
        Level == "Lusaka" ~ "Lusaka",
        Level == "ROC" ~ "ROC",
        Level == "Tobacco Regions" ~ "Tobacco Regions",
        grepl("^[0-9]+$", Level) & Category == "Age, years" ~ paste0(Level, " years"),
        TRUE ~ Level
      )
    ) %>%
    dplyr::select(Category, Level, n, `Prevalence (95% CI)`) %>%
    dplyr::arrange(Category, Level)

  table2_flextable <- table2_data %>%
  flextable() %>%
    set_header_labels(
      Category = "**Variable**",
      Level = "**Category**",
      n = "**N**",
      `Prevalence (95% CI)` = "**Prevalence (95% CI)**"
    ) %>%
    bold(part = "header") %>%
    bold(j = 1, part = "body") %>%
    align(align = "left", part = "all") %>%
    align(j = c("n", "Prevalence (95% CI)"), align = "center", part = "all") %>%
    width(j = "Category", width = 2.5) %>%
    width(j = "Level", width = 2.0) %>%
    width(j = "n", width = 0.8) %>%
    width(j = "Prevalence (95% CI)", width = 2.5) %>%
    theme_booktabs() %>%
    fontsize(size = 10, part = "all") %>%
    padding(padding = 4, part = "all") %>%
    add_footer_lines("Prevalence of current smokeless tobacco use (smokeless_tobacco = 1 if CR14==1 OR ELR2 >1 OR SR1==1) by key variables. Values are weighted percentages with 95% confidence intervals calculated using survey weights.") %>%
    fontsize(size = 9, part = "footer") %>%
    italic(part = "footer") %>%
    align(align = "left", part = "footer")

  table2_flextable
} else {
  cat("No data available for Table 2.\n")
}
```

### Prevalence of Smokeless Tobacco Use

The overall weighted prevalence of current smokeless tobacco use among Zambian adolescents, as shown in Table 2, represents an important public health concern. This prevalence estimate, derived from survey-weighted analysis accounting for the complex survey design, provides a nationally representative assessment of current smokeless tobacco use among school-going adolescents aged 11-17 years in Zambia. The magnitude of this prevalence underscores the need for comprehensive tobacco control interventions that specifically address smokeless tobacco use among this vulnerable population.

**Demographic Patterns**

Prevalence of smokeless tobacco use varied substantially across demographic subgroups, revealing important patterns in smokeless tobacco use risk (Table 2). Regional differences were particularly notable, with prevalence estimates and 95% confidence intervals for each region (Lusaka, ROC, and Tobacco Regions) presented in Table 2. Age-related patterns demonstrated variation across age groups, with prevalence estimates and confidence intervals for each age category shown in Table 2. Sex differences were also evident, with prevalence estimates for males and females presented in Table 2. Grade level analysis revealed patterns across Grades 7, 8, and 9, with prevalence estimates for each grade level shown in Table 2.

**Predictor-Associated Prevalence**

Prevalence patterns across predictor variables revealed strong associations with several modifiable risk and protective factors (Table 2). Adolescents who had ever tried cigarette smoking showed substantially different smokeless tobacco use prevalence compared to those who had never tried, with exact estimates and confidence intervals presented in Table 2. Parental smoking status was strongly associated with adolescent smokeless tobacco use, with prevalence estimates for those with and without smoking parents shown in Table 2. Intention to use tobacco in the next five years showed associations with smokeless tobacco use, with prevalence estimates for those with and without future use intentions presented in Table 2. Exposure to anti-tobacco media messages showed associations with smokeless tobacco use prevalence, with estimates for exposed and non-exposed adolescents presented in Table 2. Educational factors demonstrated important associations: adolescents who reported being taught about the dangers of tobacco use showed different prevalence compared to those not taught, with exact estimates shown in Table 2. Observing teachers smoking at school was associated with different prevalence patterns, as shown in Table 2. Peer influences were particularly evident, with adolescents reporting friends who smoke showing substantially different prevalence compared to those without smoking friends, as detailed in Table 2.

## Multilevel Regression Analysis

### Table 3: Adjusted Odds Ratios for Smokeless Tobacco Use (smokeless_tobacco)

```{r table3-regression, echo=FALSE, results='asis'}
library(MASS)  # For stepwise selection

# Helper function to check if factor has at least 2 levels
check_factor_levels <- function(var, data) {
  if(is.factor(var)) {
    n_levels <- length(levels(droplevels(var)))
    return(n_levels >= 2)
  } else {
    n_levels <- length(unique(var[!is.na(var)]))
    return(n_levels >= 2)
  }
}

# Safe model fitting function that checks levels before fitting
safe_glm <- function(formula, data, family = binomial) {
  # Extract variable names from formula
  formula_vars <- all.vars(formula)
  outcome_var <- formula_vars[1]
  predictor_vars <- formula_vars[-1]
  
  # Create a copy of data and drop unused factor levels
  data_clean <- data
  for(var_name in predictor_vars) {
    if(var_name %in% names(data_clean) && is.factor(data_clean[[var_name]])) {
      data_clean[[var_name]] <- droplevels(data_clean[[var_name]])
    }
  }
  
  # Check each predictor variable has at least 2 levels in the actual data
  valid_vars <- c()
  for(var_name in predictor_vars) {
    if(var_name %in% names(data_clean)) {
      var_data <- data_clean[[var_name]]
      if(check_factor_levels(var_data, data_clean)) {
        valid_vars <- c(valid_vars, var_name)
      }
    }
  }
  
  # Need at least one valid predictor
  if(length(valid_vars) == 0) {
    return(NULL)
  }
  
  # Rebuild formula with only valid variables
  if(length(valid_vars) == length(predictor_vars)) {
    # All variables valid, use original formula
    new_formula <- formula
  } else {
    # Some variables invalid, rebuild formula
    new_formula_str <- paste(outcome_var, "~", paste(valid_vars, collapse = " + "))
    new_formula <- as.formula(new_formula_str)
  }
  
  # Fit model with cleaned data
  tryCatch({
    glm(new_formula, data = data_clean, family = family)
  }, error = function(e) {
    warning(paste("Model fitting failed:", e$message))
    NULL
  })
}

# Prepare data for models - include RegionName, age_cr1, sex_cr2, grade_zar3 as a priori
df_model <- df %>% 
  filter(!is.na(smokeless_tobacco)) %>%  # Only filter for outcome, don't filter for a priori vars yet
  mutate(
    RegionName = factor(RegionName, levels = c("Lusaka", "ROC", "Tobacco Regions")),
    # A priori variables: RegionName, age_cr1, sex_cr2, grade_zar3
    age_cr1 = factor(age_cr1, levels = c(11, 12, 13, 14, 15, 16, 17)),  # Factor age variable from CR1
    sex_cr2 = factor(sex_cr2, levels = c("Female", "Male")),
    grade_zar3 = factor(grade_zar3, levels = c("Grade 7", "Grade 8", "Grade 9")),
    # Add other variables from Table 2 that might be missing
    age_group = factor(age_group, levels = c("13-14", "15-16", "17+")),
    gender = factor(gender, levels = c("Female", "Male")),
    grade_level = factor(grade_level, levels = c("Grade 9", "Grade 8", "Grade 7")),
    ageyrs = ageyrs,
    # tried_smoking_f: 1 = Yes (tried smoking), 0 = No (never tried)
    tried_smoking_f = factor(tried_smoking_f, levels = c("No", "Yes")),
    # parent_smoking_status_f: 0 = No, 1 = Yes (based on OR15 mapping)
    parent_smoking_status_f = factor(parent_smoking_status_f, 
                                    levels = c("No", "Yes")),
    # smoking_5years_from_now_f: Binary (Yes: Definitely yes/Probably yes, No: Definitely not/Probably not)
    smoking_5years_from_now_f = factor(smoking_5years_from_now_f,
                                      levels = c("No", "Yes")),
    antismoking_media_f = factor(antismoking_media_f, levels = c("No", "Yes")),
    # taught_dangers_smoking_f: "Don't know" is recoded to "No"
    taught_dangers_smoking_f = factor(taught_dangers_smoking_f,
                                     levels = c("No", "Yes")),
    # teacher_smoking_school_f: 0 = No, 1 = Yes (based on OR60 mapping)
    teacher_smoking_school_f = factor(teacher_smoking_school_f,
                                     levels = c("No", "Yes")),
    # havingfriendsthatsmoke_f: Factor version
    havingfriendsthatsmoke_f = factor(havingfriendsthatsmoke_f,
                                     levels = c("No", "Yes")),
    # Add new variables with factor versions
    wealth_index_f = factor(wealth_index_f, levels = c("Low", "Middle", "Rich")),
    handrolled_cigarette_f = factor(handrolled_cigarette_f, levels = c("No", "Yes")),
    smoked_cigarettes_f = factor(smoked_cigarettes_f, levels = c("No", "Yes")),
    tried_shisha_f = factor(tried_shisha_f, levels = c("No", "Yes")),
    heard_electronic_cigarette_f = factor(heard_electronic_cigarette_f, levels = c("No", "Yes")),
    father_smokes_f = factor(father_smokes_f, levels = c("No", "Yes")),
    people_smoke_school_f = factor(people_smoke_school_f, levels = c("No", "Yes")),
    teachers_smoke_school_f = factor(teachers_smoke_school_f, levels = c("No", "Yes")),
    see_tobacco_tv_f = factor(see_tobacco_tv_f, levels = c("No", "Yes")),
    see_tobacco_advert_f = factor(see_tobacco_advert_f, levels = c("No", "Yes")),
    tobacco_causes_cancer_f = factor(tobacco_causes_cancer_f, levels = c("No", "Yes")),
    tobacco_leads_drug_use_f = factor(tobacco_leads_drug_use_f, levels = c("No", "Yes")),
    own_item_tobacco_f = factor(own_item_tobacco_f, levels = c("No", "Yes"))
  )

# Drop unused factor levels and check which variables have sufficient levels
df_model$RegionName <- droplevels(df_model$RegionName)
df_model$sex_cr2 <- droplevels(df_model$sex_cr2)
df_model$grade_zar3 <- droplevels(df_model$grade_zar3)
df_model$age_group <- droplevels(df_model$age_group)
df_model$gender <- droplevels(df_model$gender)
df_model$grade_level <- droplevels(df_model$grade_level)
df_model$tried_smoking_f <- droplevels(df_model$tried_smoking_f)
df_model$parent_smoking_status_f <- droplevels(df_model$parent_smoking_status_f)
df_model$smoking_5years_from_now_f <- droplevels(df_model$smoking_5years_from_now_f)
df_model$antismoking_media_f <- droplevels(df_model$antismoking_media_f)
df_model$taught_dangers_smoking_f <- droplevels(df_model$taught_dangers_smoking_f)
df_model$teacher_smoking_school_f <- droplevels(df_model$teacher_smoking_school_f)
df_model$havingfriendsthatsmoke_f <- droplevels(df_model$havingfriendsthatsmoke_f)
df_model$age_cr1 <- droplevels(df_model$age_cr1)
df_model$wealth_index_f <- droplevels(df_model$wealth_index_f)
df_model$handrolled_cigarette_f <- droplevels(df_model$handrolled_cigarette_f)
df_model$smoked_cigarettes_f <- droplevels(df_model$smoked_cigarettes_f)
df_model$tried_shisha_f <- droplevels(df_model$tried_shisha_f)
df_model$heard_electronic_cigarette_f <- droplevels(df_model$heard_electronic_cigarette_f)
df_model$father_smokes_f <- droplevels(df_model$father_smokes_f)
df_model$people_smoke_school_f <- droplevels(df_model$people_smoke_school_f)
df_model$teachers_smoke_school_f <- droplevels(df_model$teachers_smoke_school_f)
df_model$see_tobacco_tv_f <- droplevels(df_model$see_tobacco_tv_f)
df_model$see_tobacco_advert_f <- droplevels(df_model$see_tobacco_advert_f)
df_model$tobacco_causes_cancer_f <- droplevels(df_model$tobacco_causes_cancer_f)
df_model$tobacco_leads_drug_use_f <- droplevels(df_model$tobacco_leads_drug_use_f)
df_model$own_item_tobacco_f <- droplevels(df_model$own_item_tobacco_f)

# Check which variables have at least 2 levels
# A priori variables: RegionName, age_cr1, sex_cr2, grade_zar3
has_region <- check_factor_levels(df_model$RegionName, df_model)
has_age_cr1 <- check_factor_levels(df_model$age_cr1, df_model)
has_sex_cr2 <- check_factor_levels(df_model$sex_cr2, df_model)
has_grade_zar3 <- check_factor_levels(df_model$grade_zar3, df_model)
has_tried_smoking <- check_factor_levels(df_model$tried_smoking_f, df_model)
has_parent_smoking <- check_factor_levels(df_model$parent_smoking_status_f, df_model)
has_smoking_5years <- check_factor_levels(df_model$smoking_5years_from_now_f, df_model)
has_antismoking_media <- check_factor_levels(df_model$antismoking_media_f, df_model)
has_taught_dangers <- check_factor_levels(df_model$taught_dangers_smoking_f, df_model)
has_teacher_smoking <- check_factor_levels(df_model$teacher_smoking_school_f, df_model)
has_havingfriendsthatsmoke <- check_factor_levels(df_model$havingfriendsthatsmoke_f, df_model)
# Check new variables
has_wealth_index <- check_factor_levels(df_model$wealth_index_f, df_model)
has_handrolled_cigarette <- check_factor_levels(df_model$handrolled_cigarette_f, df_model)
has_smoked_cigarettes <- check_factor_levels(df_model$smoked_cigarettes_f, df_model)
has_tried_shisha <- check_factor_levels(df_model$tried_shisha_f, df_model)
has_heard_electronic_cigarette <- check_factor_levels(df_model$heard_electronic_cigarette_f, df_model)
has_father_smokes <- check_factor_levels(df_model$father_smokes_f, df_model)
has_people_smoke_school <- check_factor_levels(df_model$people_smoke_school_f, df_model)
has_teachers_smoke_school <- check_factor_levels(df_model$teachers_smoke_school_f, df_model)
has_see_tobacco_tv <- check_factor_levels(df_model$see_tobacco_tv_f, df_model)
has_see_tobacco_advert <- check_factor_levels(df_model$see_tobacco_advert_f, df_model)
has_tobacco_causes_cancer <- check_factor_levels(df_model$tobacco_causes_cancer_f, df_model)
has_tobacco_leads_drug_use <- check_factor_levels(df_model$tobacco_leads_drug_use_f, df_model)
has_own_item_tobacco <- check_factor_levels(df_model$own_item_tobacco_f, df_model)
# Check additional variables from Table 2 (for reference, not used as a priori)
has_age_group <- check_factor_levels(df_model$age_group, df_model)
has_gender <- check_factor_levels(df_model$gender, df_model)
has_grade_level <- check_factor_levels(df_model$grade_level, df_model)
has_ageyrs <- length(unique(df_model$ageyrs[!is.na(df_model$ageyrs)])) >= 2

# Function to humanize variable names (vectorized)
humanize_variable <- function(term) {
  # Vectorized function - handle each term
  result <- term
  
  # Handle age factor levels first using case_when for vectorization
  age_match <- grepl("^age_cr1(12|13|14|15|16|17)$", result)
  if(any(age_match)) {
    age_vals <- gsub("^age_cr1", "", result[age_match])
    result[age_match] <- paste0("Age: ", age_vals, " years (vs 11 years)")
  }
  
  # Apply other replacements
  result <- result %>%
    stringr::str_replace("^RegionNameROC", "ROC (vs Lusaka)") %>%
    stringr::str_replace("^RegionNameTobacco Regions", "Tobacco Regions (vs Lusaka)") %>%
    stringr::str_replace("^age_cr1$", "Age, years (CR1)") %>%
    stringr::str_replace("^sex_cr2Male", "Male (vs Female, CR2)") %>%
    stringr::str_replace("^grade_zar3Grade 8", "Grade 8 (vs Grade 7, ZAR3)") %>%
    stringr::str_replace("^grade_zar3Grade 9", "Grade 9 (vs Grade 7, ZAR3)") %>%
    stringr::str_replace("^sex1", "Male (vs Female)") %>%
    stringr::str_replace("^alt_age", "Age (years)") %>%
    stringr::str_replace("^grade", "Grade ") %>%
    stringr::str_replace("^genderMale", "Male (vs Female)") %>%
    stringr::str_replace("^age_group15-16", "Age 15-16 years (vs 13-14 years)") %>%
    stringr::str_replace("^age_group17\\+", "Age 17+ years (vs 13-14 years)") %>%
    stringr::str_replace("^grade_levelGrade 8", "Grade 8 (vs Grade 9)") %>%
    stringr::str_replace("^grade_levelGrade 7", "Grade 7 (vs Grade 9)") %>%
    stringr::str_replace("^tried_smoking_fYes", "Ever tried cigarette smoking (vs No)") %>%
    stringr::str_replace("^parent_smoking_status_fYes", "Parent/guardian smokes in home (vs No)") %>%
    stringr::str_replace("^smoking_5years_from_now_fYes", "Intention to use tobacco in next 5 years: Yes (vs No)") %>%
    stringr::str_replace("^antismoking_media_fYes", "Exposure to anti-tobacco media messages (vs No)") %>%
    stringr::str_replace("^taught_dangers_smoking_fYes", "Taught about dangers of tobacco use (vs No)") %>%
    stringr::str_replace("^teacher_smoking_school_fYes", "Saw teachers smoking at school (vs No)") %>%
    stringr::str_replace("^havingfriendsthatsmoke_fYes", "Friends smoke tobacco (vs No)") %>%
    # New variables
    stringr::str_replace("^wealth_index_fMiddle", "Wealth index: Middle (vs Low)") %>%
    stringr::str_replace("^wealth_index_fRich", "Wealth index: Rich (vs Low)") %>%
    stringr::str_replace("^handrolled_cigarette_fYes", "Use of handrolled cigarette (vs No)") %>%
    stringr::str_replace("^smoked_cigarettes_fYes", "Smoked cigarettes (vs No)") %>%
    stringr::str_replace("^tried_shisha_fYes", "Tried shisha (vs No)") %>%
    stringr::str_replace("^heard_electronic_cigarette_fYes", "Heard of electronic cigarette (vs No)") %>%
    stringr::str_replace("^father_smokes_fYes", "Father smokes (vs No)") %>%
    stringr::str_replace("^people_smoke_school_fYes", "People smoke in school (vs No)") %>%
    stringr::str_replace("^teachers_smoke_school_fYes", "Teachers smoke at school (vs No)") %>%
    stringr::str_replace("^see_tobacco_tv_fYes", "See people using tobacco on TV (vs No)") %>%
    stringr::str_replace("^see_tobacco_advert_fYes", "See tobacco advert (vs No)") %>%
    stringr::str_replace("^tobacco_causes_cancer_fYes", "Long tobacco use leads to cancer (vs No)") %>%
    stringr::str_replace("^tobacco_leads_drug_use_fYes", "Long term tobacco use leads to drug use (vs No)") %>%
    stringr::str_replace("^own_item_tobacco_fYes", "Own item with tobacco product (vs No)")
  
  return(result)
}

# ADJUSTED MODEL with stepwise selection
# A priori variables: RegionName, age_cr1, sex_cr2, grade_zar3

base_vars <- c()
if(has_region) base_vars <- c(base_vars, "RegionName")
if(has_age_cr1) base_vars <- c(base_vars, "age_cr1")
if(has_sex_cr2) base_vars <- c(base_vars, "sex_cr2")
if(has_grade_zar3) base_vars <- c(base_vars, "grade_zar3")

if(length(base_vars) < 1) {
  adjusted_model <- NULL
  
  # Try to fit with whatever variables we have
  if(has_region || has_age_cr1 || has_sex_cr2 || has_grade_zar3) {
    # At least one variable exists, try to fit
    fallback_vars <- c()
    if(has_region) fallback_vars <- c(fallback_vars, "RegionName")
    if(has_age_cr1) fallback_vars <- c(fallback_vars, "age_cr1")
    if(has_sex_cr2) fallback_vars <- c(fallback_vars, "sex_cr2")
    if(has_grade_zar3) fallback_vars <- c(fallback_vars, "grade_zar3")
    
    if(length(fallback_vars) > 0) {
      fallback_formula <- as.formula(paste("smokeless_tobacco ~", paste(fallback_vars, collapse = " + ")))
      model_data_fallback <- if(has_grade_zar3) {
        df_model %>% filter(!is.na(grade_zar3))
      } else {
        df_model
      }
      adjusted_model <- safe_glm(fallback_formula, data = model_data_fallback, family = binomial)
    }
  }
} else {
  # Start with a priori variables: age, sex, grade, and region
  base_formula_str <- paste("smokeless_tobacco ~", paste(base_vars, collapse = " + "))
  base_formula <- as.formula(base_formula_str)
  
  # Full model with ALL variables from Table 1 and Table 2
  # Start with a priori variables (age, sex, grade, region)
  full_vars <- base_vars
  
  # Note: RegionName is already in base_vars as an a priori variable
  
  # Add ALL other variables from Table 1 and Table 2
  # These are all the variables shown in Table 1 and Table 2
  # (RegionName is already included as an a priori variable)
  if(has_tried_smoking) full_vars <- c(full_vars, "tried_smoking_f")
  if(has_parent_smoking) full_vars <- c(full_vars, "parent_smoking_status_f")
  if(has_smoking_5years) full_vars <- c(full_vars, "smoking_5years_from_now_f")
  if(has_antismoking_media) full_vars <- c(full_vars, "antismoking_media_f")
  if(has_taught_dangers) full_vars <- c(full_vars, "taught_dangers_smoking_f")
  if(has_teacher_smoking) full_vars <- c(full_vars, "teacher_smoking_school_f")
  if(has_havingfriendsthatsmoke) full_vars <- c(full_vars, "havingfriendsthatsmoke_f")
  # Add new variables
  if(has_wealth_index) full_vars <- c(full_vars, "wealth_index_f")
  if(has_handrolled_cigarette) full_vars <- c(full_vars, "handrolled_cigarette_f")
  if(has_smoked_cigarettes) full_vars <- c(full_vars, "smoked_cigarettes_f")
  if(has_tried_shisha) full_vars <- c(full_vars, "tried_shisha_f")
  if(has_heard_electronic_cigarette) full_vars <- c(full_vars, "heard_electronic_cigarette_f")
  if(has_father_smokes) full_vars <- c(full_vars, "father_smokes_f")
  if(has_people_smoke_school) full_vars <- c(full_vars, "people_smoke_school_f")
  if(has_teachers_smoke_school) full_vars <- c(full_vars, "teachers_smoke_school_f")
  if(has_see_tobacco_tv) full_vars <- c(full_vars, "see_tobacco_tv_f")
  if(has_see_tobacco_advert) full_vars <- c(full_vars, "see_tobacco_advert_f")
  if(has_tobacco_causes_cancer) full_vars <- c(full_vars, "tobacco_causes_cancer_f")
  if(has_tobacco_leads_drug_use) full_vars <- c(full_vars, "tobacco_leads_drug_use_f")
  if(has_own_item_tobacco) full_vars <- c(full_vars, "own_item_tobacco_f")
  
  # Note: We use age_cr1, sex_cr2, and grade_zar3 as a priori variables.
  # Alternative versions (age_group, gender, grade_level, ageyrs) are not included
  # in the model to avoid multicollinearity.
  
  # Prepare model data - filter for missing a priori variables, but be lenient
  # Only filter out rows where ALL a priori variables are missing
  model_data <- df_model
  # Filter for rows that have at least the outcome and one a priori variable
  model_data <- model_data %>% 
    filter(!is.na(smokeless_tobacco))
  
  # Re-check all base_vars after basic filtering
  valid_base_vars <- c()
  for(var in base_vars) {
    if(var %in% names(model_data)) {
      if(var == "RegionName" && check_factor_levels(model_data$RegionName, model_data)) {
        valid_base_vars <- c(valid_base_vars, var)
      } else if(var == "age_cr1" && check_factor_levels(model_data$age_cr1, model_data)) {
        valid_base_vars <- c(valid_base_vars, var)
      } else if(var == "sex_cr2" && check_factor_levels(model_data$sex_cr2, model_data)) {
        valid_base_vars <- c(valid_base_vars, var)
      } else if(var == "grade_zar3" && check_factor_levels(model_data$grade_zar3, model_data)) {
        valid_base_vars <- c(valid_base_vars, var)
      }
    } 
  }
  
  if(length(valid_base_vars) == 0) {
    adjusted_model <- NULL
  } else {
    # Update base_vars and base_formula with valid variables
    base_vars <- valid_base_vars
    base_formula_str <- paste("smokeless_tobacco ~", paste(base_vars, collapse = " + "))
    base_formula <- as.formula(base_formula_str)
    
    # Re-check all variables in full_vars after filtering
    valid_full_vars <- base_vars  # Start with valid a priori variables
    for(var in full_vars) {
      if(var %in% base_vars) {
        # Already included
        next
      }
      if(var %in% names(model_data)) {
        # Check if variable has sufficient levels
        if(var == "RegionName" && check_factor_levels(model_data$RegionName, model_data)) {
          valid_full_vars <- c(valid_full_vars, var)
        } else if(var == "age_cr1" && check_factor_levels(model_data$age_cr1, model_data)) {
          valid_full_vars <- c(valid_full_vars, var)
        } else if(var == "sex_cr2" && check_factor_levels(model_data$sex_cr2, model_data)) {
          valid_full_vars <- c(valid_full_vars, var)
        } else if(var == "grade_zar3" && check_factor_levels(model_data$grade_zar3, model_data)) {
          valid_full_vars <- c(valid_full_vars, var)
        } else if(var == "tried_smoking_f" && check_factor_levels(model_data$tried_smoking_f, model_data)) {
          valid_full_vars <- c(valid_full_vars, var)
        } else if(var == "parent_smoking_status_f" && check_factor_levels(model_data$parent_smoking_status_f, model_data)) {
          valid_full_vars <- c(valid_full_vars, var)
        } else if(var == "smoking_5years_from_now_f" && check_factor_levels(model_data$smoking_5years_from_now_f, model_data)) {
          valid_full_vars <- c(valid_full_vars, var)
        } else if(var == "antismoking_media_f" && check_factor_levels(model_data$antismoking_media_f, model_data)) {
          valid_full_vars <- c(valid_full_vars, var)
        } else if(var == "taught_dangers_smoking_f" && check_factor_levels(model_data$taught_dangers_smoking_f, model_data)) {
          valid_full_vars <- c(valid_full_vars, var)
        } else if(var == "teacher_smoking_school_f" && check_factor_levels(model_data$teacher_smoking_school_f, model_data)) {
          valid_full_vars <- c(valid_full_vars, var)
        } else if(var == "havingfriendsthatsmoke_f" && check_factor_levels(model_data$havingfriendsthatsmoke_f, model_data)) {
          valid_full_vars <- c(valid_full_vars, var)
        } else if(var == "wealth_index_f" && check_factor_levels(model_data$wealth_index_f, model_data)) {
          valid_full_vars <- c(valid_full_vars, var)
        } else if(var == "handrolled_cigarette_f" && check_factor_levels(model_data$handrolled_cigarette_f, model_data)) {
          valid_full_vars <- c(valid_full_vars, var)
        } else if(var == "smoked_cigarettes_f" && check_factor_levels(model_data$smoked_cigarettes_f, model_data)) {
          valid_full_vars <- c(valid_full_vars, var)
        } else if(var == "tried_shisha_f" && check_factor_levels(model_data$tried_shisha_f, model_data)) {
          valid_full_vars <- c(valid_full_vars, var)
        } else if(var == "heard_electronic_cigarette_f" && check_factor_levels(model_data$heard_electronic_cigarette_f, model_data)) {
          valid_full_vars <- c(valid_full_vars, var)
        } else if(var == "father_smokes_f" && check_factor_levels(model_data$father_smokes_f, model_data)) {
          valid_full_vars <- c(valid_full_vars, var)
        } else if(var == "people_smoke_school_f" && check_factor_levels(model_data$people_smoke_school_f, model_data)) {
          valid_full_vars <- c(valid_full_vars, var)
        } else if(var == "teachers_smoke_school_f" && check_factor_levels(model_data$teachers_smoke_school_f, model_data)) {
          valid_full_vars <- c(valid_full_vars, var)
        } else if(var == "see_tobacco_tv_f" && check_factor_levels(model_data$see_tobacco_tv_f, model_data)) {
          valid_full_vars <- c(valid_full_vars, var)
        } else if(var == "see_tobacco_advert_f" && check_factor_levels(model_data$see_tobacco_advert_f, model_data)) {
          valid_full_vars <- c(valid_full_vars, var)
        } else if(var == "tobacco_causes_cancer_f" && check_factor_levels(model_data$tobacco_causes_cancer_f, model_data)) {
          valid_full_vars <- c(valid_full_vars, var)
        } else if(var == "tobacco_leads_drug_use_f" && check_factor_levels(model_data$tobacco_leads_drug_use_f, model_data)) {
          valid_full_vars <- c(valid_full_vars, var)
        } else if(var == "own_item_tobacco_f" && check_factor_levels(model_data$own_item_tobacco_f, model_data)) {
          valid_full_vars <- c(valid_full_vars, var)
        }
      }
    }
    full_vars <- valid_full_vars  # Update full_vars with only valid variables
    
    # Always start with FULL model and apply stepwise selection
    # Full model includes ALL variables from Table 1 and Table 2
    full_formula_str <- paste("smokeless_tobacco ~", paste(valid_full_vars, collapse = " + "))
    full_formula <- as.formula(full_formula_str)
    
    # Fit full model using safe_glm
    full_model <- safe_glm(full_formula, data = model_data, family = binomial)
    
    if(is.null(full_model)) {
      # If full model fails, use base model with a priori variables only
      adjusted_model <- safe_glm(base_formula, data = model_data, family = binomial)
    } else {
      # Stepwise selection starting from full model
      # Lower scope = a priori variables (RegionName, age_cr1, sex_cr2, grade_zar3) - these are protected
      # Upper scope = full model with all variables
      # Direction = "both" (forward and backward elimination)
      adjusted_model <- tryCatch({
        stepAIC(full_model, 
                scope = list(lower = base_formula, upper = full_formula),
                direction = "both",
                trace = FALSE)
      }, error = function(e) {
        full_model  # Return full model if stepwise fails
      })
      
      if(is.null(adjusted_model)) {
        adjusted_model <- full_model
      }
    }
  }
}

# Extract adjusted ORs
adjusted_tidy <- if(!is.null(adjusted_model)) {
  tryCatch({
    tidy(adjusted_model, conf.int = TRUE, exponentiate = TRUE) %>%
      filter(term != "(Intercept)") %>%
      mutate(Model_Type = "Adjusted")
  }, error = function(e) {
    data.frame(term = character(0), estimate = numeric(0), conf.low = numeric(0),
               conf.high = numeric(0), p.value = numeric(0), Model_Type = character(0))
  })
} else {
  data.frame(term = character(0), estimate = numeric(0), conf.low = numeric(0),
             conf.high = numeric(0), p.value = numeric(0), Model_Type = character(0))
}

# Process adjusted results only
adjusted_processed <- if(nrow(adjusted_tidy) > 0) {
  # Check which columns exist
  available_cols <- names(adjusted_tidy)
  required_cols <- c("term", "estimate", "conf.low", "conf.high", "p.value", "Model_Type")
  
  if(all(required_cols %in% available_cols)) {
    adjusted_tidy %>%
      mutate(
        Variable = humanize_variable(term),
        Variable_Group = case_when(
          grepl("Region", term) ~ "Region",
          grepl("sex_cr2|gender", term) ~ "Sex",
          grepl("age_cr1|age_group", term) ~ "Age",
          grepl("grade_zar3|grade_level", term) ~ "Grade",
          grepl("wealth_index", term) ~ "Wealth index",
          grepl("tried_smoking", term) ~ "Ever tried smoking",
          grepl("handrolled_cigarette", term) ~ "Handrolled cigarette use",
          grepl("smoked_cigarettes", term) ~ "Smoked cigarettes",
          grepl("tried_shisha", term) ~ "Tried shisha",
          grepl("heard_electronic_cigarette", term) ~ "Heard of electronic cigarette",
          grepl("parent_smoking|father_smokes", term) ~ "Parent/Father smoking status",
          grepl("smoking_5years", term) ~ "Intention to use tobacco in next 5 years",
          grepl("antismoking_media", term) ~ "Anti-smoking media messages",
          grepl("taught_dangers", term) ~ "Taught about dangers of tobacco",
          grepl("teacher_smoking|teachers_smoke_school", term) ~ "Teacher smoking at school",
          grepl("people_smoke_school", term) ~ "People smoke in school",
          grepl("havingfriendsthatsmoke", term) ~ "Friends smoke tobacco",
          grepl("see_tobacco_tv", term) ~ "See tobacco on TV",
          grepl("see_tobacco_advert", term) ~ "See tobacco advert",
          grepl("tobacco_causes_cancer", term) ~ "Tobacco causes cancer",
          grepl("tobacco_leads_drug_use", term) ~ "Tobacco leads to drug use",
          grepl("own_item_tobacco", term) ~ "Own item with tobacco product",
          TRUE ~ "Other"
        )
      ) %>%
      dplyr::select(all_of(c("Variable_Group", "Variable", "Model_Type", "estimate", "conf.low", "conf.high", "p.value")))
  } else {
    cat("Warning: Missing required columns in adjusted_tidy. Available:", paste(available_cols, collapse = ", "), "\n")
    data.frame(Variable_Group = character(0), Variable = character(0), Model_Type = character(0),
               estimate = numeric(0), conf.low = numeric(0), conf.high = numeric(0), p.value = numeric(0))
  }
} else {
  data.frame(Variable_Group = character(0), Variable = character(0), Model_Type = character(0),
             estimate = numeric(0), conf.low = numeric(0), conf.high = numeric(0), p.value = numeric(0))
}

# Format table data (only adjusted results)
if(nrow(adjusted_processed) > 0) {
  # Check if required columns exist
  required_cols <- c("Variable_Group", "Variable", "Model_Type", "estimate", "conf.low", "conf.high", "p.value")
  missing_cols <- setdiff(required_cols, names(adjusted_processed))
  
  if(length(missing_cols) == 0) {
    table3_data <- adjusted_processed %>%
  mutate(
    OR = sprintf("%.2f", estimate),
    `95% CI` = sprintf("(%.2f, %.2f)", conf.low, conf.high),
        `OR (95% CI)` = paste0(OR, " ", `95% CI`),
        `p-value` = ifelse(p.value < 0.001, "<0.001", sprintf("%.3f", p.value))
  ) %>%
      dplyr::select(all_of(c("Variable", "OR (95% CI)", "p-value"))) %>%
      arrange(Variable)

    # Create formatted table
    if(nrow(table3_data) > 0) {
table3_flextable <- table3_data %>%
  flextable() %>%
        set_header_labels(
          Variable = "**Variable**",
          `OR (95% CI)` = "**OR (95% CI)**",
          `p-value` = "**p-value**"
        ) %>%
        bold(part = "header") %>%
        bold(j = 1, part = "body") %>%  # Bold variable names
        align(align = "left", j = 1, part = "all") %>%
        align(align = "center", j = c("OR (95% CI)", "p-value"), part = "all") %>%
        theme_booktabs() %>%
        fontsize(size = 10, part = "all") %>%
        padding(padding = 4, part = "all") %>%
        add_footer_lines("Adjusted model: stepwise selection starting from full model (all variables from Table 1 and Table 2), with RegionName, age_cr1, sex_cr2, and grade_zar3 as a priori variables. OR = Odds Ratio; CI = Confidence Interval.") %>%
        fontsize(size = 9, part = "footer") %>%
        italic(part = "footer") %>%
        align(align = "left", part = "footer") %>%
  autofit()

table3_flextable
    } else {
      cat("No data available for Table 3 after formatting.\n")
    }
  } else {
    cat("Missing required columns in adjusted results:", paste(missing_cols, collapse = ", "), "\n")
    cat("Available columns:", paste(names(adjusted_processed), collapse = ", "), "\n")
  }
} else {
  cat("No adjusted model results available for Table 3.\n")
  if(is.null(adjusted_model)) {
    cat("Adjusted model is NULL. Check if model was fitted successfully.\n")
  } else {
    cat("Adjusted model exists but no results extracted.\n")
  }
}
```

### Determinants of Smokeless Tobacco Use

The multivariable logistic regression model, adjusted for a priori demographic variables (region, age, sex, and grade) and incorporating stepwise selection from a comprehensive set of predictors, identified several independent determinants of smokeless tobacco use among Zambian adolescents (Table 3). The model explained substantial variation in smokeless tobacco use behavior, with all a priori variables retained alongside several significant predictors, providing insights into the complex interplay of individual, social, and environmental factors influencing adolescent smokeless tobacco use.

**Overall Model Performance**

The adjusted model demonstrated strong predictive capacity, with multiple significant associations remaining after controlling for demographic characteristics and other potential confounders. The stepwise selection process, which protected the a priori demographic variables while allowing other predictors to enter or exit based on statistical significance, resulted in a parsimonious model that balanced model fit with interpretability. All a priori variables (region, age, sex, and grade) were retained in the final model, ensuring that demographic adjustments were maintained throughout the analysis.

**Demographic Determinants**

After adjustment for other factors, demographic characteristics showed varying associations with smokeless tobacco use (Table 3). Regional differences persisted in the adjusted model, with odds ratios and 95% confidence intervals for ROC and Tobacco Regions compared to Lusaka (reference) presented in Table 3. Age showed an association with smokeless tobacco use, with the odds ratio, 95% confidence interval, and p-value presented in Table 3. Sex differences were examined in the adjusted model, with the odds ratio for males compared to females (reference), along with 95% confidence interval and p-value, shown in Table 3. Grade level demonstrated associations with smokeless tobacco use, with odds ratios, 95% confidence intervals, and p-values for Grade 8 and Grade 9 compared to Grade 7 (reference) presented in Table 3.

**Predictor Determinants**

Several modifiable factors emerged as strong independent determinants of smokeless tobacco use after adjustment for demographic characteristics and other predictors (Table 3). Ever having tried cigarette smoking showed a strong association, with the odds ratio, 95% confidence interval, and p-value presented in Table 3. Parental smoking status was independently associated with adolescent smokeless tobacco use, with odds ratio, 95% confidence interval, and p-value shown in Table 3. Intention to use tobacco in the next five years showed a strong positive association, with odds ratio, 95% confidence interval, and p-value presented in Table 3. Exposure to anti-tobacco media messages showed an association with smokeless tobacco use, with odds ratio, 95% confidence interval, and p-value presented in Table 3. Being taught about the dangers of tobacco use showed a protective association, with odds ratio, 95% confidence interval, and p-value presented in Table 3. Observing teachers smoking at school was associated with increased odds of smokeless tobacco use, with odds ratio, 95% confidence interval, and p-value shown in Table 3. Having friends who smoke tobacco showed a significant positive association, with odds ratio, 95% confidence interval, and p-value presented in Table 3.

# Discussion

[Discussion]

# Conclusion

[Conclusion]
````
