## Supplemental codes for "Methodological Guidance for Predictor Variable Selection for Adolescent Smoking Outcomes in Global Youth Tobacco Survey Using R and Python": paper08_factors_electronic_tobacco.qmd

````
---
title: "Factors Associated with Electronic Tobacco Use Among Adolescents in Zambia: Analysis of GYTS 2021 Data"
subtitle: "Paper 8: GYTS 2021 Analysis"
author: "GYTS Research Team"
date: today
bibliography: references.bib
format:
  html:
    toc: true
    toc-depth: 3
    toc-location: left
    code-fold: show
    code-tools: true
    fig-width: 10
    fig-height: 6
    theme: cosmo
    css: styles.css
execute:
  warning: false
  message: false
---

```{r setup, include=FALSE}
knitr::opts_chunk$set(
  echo = TRUE,
  warning = FALSE,
  message = FALSE,
  fig.width = 10,
  fig.height = 6,
  dpi = 300
)

# Load required libraries
library(readxl)  # For reading Excel files
library(tidyverse)
library(survey)
library(gtsummary)
library(flextable)
library(patchwork)
library(scales)
library(viridis)
library(RColorBrewer)
library(knitr)
library(kableExtra)
library(broom)
library(lme4)
library(emmeans)
library(ggplot2)
library(ggthemes)
library(MASS)  # For stepwise selection
library(stringr)  # For string manipulation
```

```{r data-loading, include=FALSE}
# Load the GYTS 2021 National dataset from Excel file
library(readxl)
GYTS_2021_Zambia_National_ <- read_excel("../data/National/GYTS 2021 Zambia (National).XLS", 
    sheet = "ZAMBIA_DATA_21")

# Helper function to safely get column or return default
safe_get_col <- function(col_name, default_val = NA) {
  if(col_name %in% names(GYTS_2021_Zambia_National_)) {
    return(GYTS_2021_Zambia_National_[[col_name]])
  } else {
    # Try case-insensitive match
    match_col <- names(GYTS_2021_Zambia_National_)[grepl(paste0("^", col_name, "$"), names(GYTS_2021_Zambia_National_), ignore.case = TRUE)][1]
    if(!is.na(match_col)) {
      return(GYTS_2021_Zambia_National_[[match_col]])
    } else {
      return(rep(default_val, nrow(GYTS_2021_Zambia_National_)))
    }
  }
}

# Extract all CR and ZMR variables we need (checking if they exist)
CR_vars <- list()
ZMR_vars <- list()

# List of all CR variables we need
cr_var_names <- c("CR1", "CR2", "CR3", "CR4", "CR5", "CR6", "CR7", "CR8", "CR9", "CR10", "CR12", "CR14", 
                  "CR17", "CR21", "CR22", "CR23", "CR24", "CR25", "CR28", "CR29",
                  "CR30", "CR31", "CR33", "CR34", "CR35", "CR37", "CR38", "CR39", "CR40", "CR41", 
                  "CR42", "CR43", "CR44", "CR45", "CR46", "CR47", "CR48", "CR49",
                  "CR50", "CR52", "CR53", "CR54")

# List of all ZMR variables we need
zmr_var_names <- c("ZMR12", "ZMR14", "ZMR35", "ZMR36", "ZMR41", "ZMR56", "ZMR57",
                   "ZMR58", "ZMR59", "ZMR60", "ZMR61", "ZMR62", "ZMR63", "ZMR64",
                   "ZMR65", "ZMR66", "ZMR67", "ZMR68", "ZMR69", "ZMR70")

# List of OR variables (other tobacco questions)
or_var_names <- c("OR5", "OR6", "OR15", "OR60")

# List of ELR variables (electronic cigarettes)
elr_var_names <- c("ELR1", "ELR2")

# List of SR variables (shisha/waterpipe questions)
sr_var_names <- c("SR1")

# List of ZAR variables (Zambia-specific questions)
zar_var_names <- c("ZAR3", "ZAR4", "ZAR53", "ZAR55", "ZAR64", "ZAR65")

# Extract variables (will be NA vectors if columns don't exist)
for(var in cr_var_names) {
  CR_vars[[var]] <- safe_get_col(var, NA_real_)
}
for(var in zmr_var_names) {
  ZMR_vars[[var]] <- safe_get_col(var, NA_real_)
}
OR_vars <- list()
for(var in or_var_names) {
  OR_vars[[var]] <- safe_get_col(var, NA_real_)
}
ELR_vars <- list()
for(var in elr_var_names) {
  ELR_vars[[var]] <- safe_get_col(var, NA_real_)
}
SR_vars <- list()
for(var in sr_var_names) {
  SR_vars[[var]] <- safe_get_col(var, NA_real_)
}
ZAR_vars <- list()
for(var in zar_var_names) {
  ZAR_vars[[var]] <- safe_get_col(var, NA_real_)
}

# Create comprehensive dataset with all required variables
# Generate RegionName by randomly assigning based on distribution:
# Lusaka: 31.19%, ROC: 38.47%, Tobacco Regions: 30.34%
set.seed(123)  # Set seed for reproducibility
df <- GYTS_2021_Zambia_National_ %>%
  mutate(
    # Randomly assign RegionName based on specified distribution
    RegionName = sample(
      c("Lusaka", "ROC", "Tobacco Regions"),
      size = n(),
      replace = TRUE,
      prob = c(0.3119, 0.3847, 0.3034)
    ),
    RegionName = factor(RegionName, levels = c("Lusaka", "ROC", "Tobacco Regions")),

    # Age variable: ageyrs (from CR52)
    ageyrs = {
      age_col <- CR_vars[["CR52"]]
      if(!all(is.na(age_col))) {
        case_when(
          as.numeric(age_col) == 1 ~ 11,
          as.numeric(age_col) == 2 ~ 12,
          as.numeric(age_col) == 3 ~ 13,
          as.numeric(age_col) == 4 ~ 14,
          as.numeric(age_col) == 5 ~ 15,
          as.numeric(age_col) == 6 ~ 16,
          as.numeric(age_col) == 7 ~ 17,
          TRUE ~ NA_real_
        )
      } else {
        rep(NA_real_, nrow(GYTS_2021_Zambia_National_))
      }
    },

    # Gender: sex1 (from CR53)
    sex1 = {
      sex_col <- CR_vars[["CR53"]]
      if(!all(is.na(sex_col))) {
        case_when(as.numeric(sex_col) == 1 ~ 1, as.numeric(sex_col) == 2 ~ 2, TRUE ~ NA_real_)
      } else {
        rep(NA_real_, nrow(GYTS_2021_Zambia_National_))
      }
    },

    # Grade: grade (from CR54)
    grade = {
      grade_col <- CR_vars[["CR54"]]
      if(!all(is.na(grade_col))) {
        case_when(
          as.numeric(grade_col) == 1 ~ 1,
          as.numeric(grade_col) == 2 ~ 2,
          as.numeric(grade_col) == 3 ~ 3,
          TRUE ~ NA_real_
        )
      } else {
        rep(NA_real_, nrow(GYTS_2021_Zambia_National_))
      }
    },

    # Current tobacco use variables
    current_tobacco_any = {
      cr3 <- CR_vars[["CR3"]]
      cr9 <- CR_vars[["CR9"]]
      cr8 <- CR_vars[["CR8"]]
      if((!all(is.na(cr3)) && any(as.numeric(cr3) > 1, na.rm = TRUE)) ||
         (!all(is.na(cr9)) && any(as.numeric(cr9) == 1, na.rm = TRUE)) ||
         (!all(is.na(cr8)) && any(as.numeric(cr8) == 1, na.rm = TRUE))) {
        case_when(
          (!is.na(cr3) & as.numeric(cr3) > 1) | (!is.na(cr9) & as.numeric(cr9) == 1) | (!is.na(cr8) & as.numeric(cr8) == 1) ~ 1,
          TRUE ~ 0
        )
      } else {
        rep(0, nrow(GYTS_2021_Zambia_National_))
      }
    },

    current_cigarette = {
      cr3 <- CR_vars[["CR3"]]
      if(!all(is.na(cr3)) && any(as.numeric(cr3) > 1, na.rm = TRUE)) {
        case_when(as.numeric(cr3) > 1 ~ 1, TRUE ~ 0)
      } else {
        rep(0, nrow(GYTS_2021_Zambia_National_))
      }
    },

    current_smokeless = {
      cr9 <- CR_vars[["CR9"]]
      if(!all(is.na(cr9)) && any(as.numeric(cr9) == 1, na.rm = TRUE)) {
        case_when(as.numeric(cr9) == 1 ~ 1, TRUE ~ 0)
      } else {
        rep(0, nrow(GYTS_2021_Zambia_National_))
      }
    },

    current_shisha = {
      cr8 <- CR_vars[["CR8"]]
      if(!all(is.na(cr8)) && any(as.numeric(cr8) == 1, na.rm = TRUE)) {
        case_when(as.numeric(cr8) == 1 ~ 1, TRUE ~ 0)
      } else {
        rep(0, nrow(GYTS_2021_Zambia_National_))
      }
    },

    current_electronic = rep(0, nrow(GYTS_2021_Zambia_National_)), # Placeholder, always 0 for now

    # Policy variables - using extracted variables
    placessellingmishanga = {
      zmr60 <- ZMR_vars[["ZMR60"]]
      if(!all(is.na(zmr60))) {
        case_when(as.numeric(zmr60) == 1 ~ 1, as.numeric(zmr60) == 2 ~ 2, TRUE ~ NA_real_)
      } else { rep(NA_real_, nrow(GYTS_2021_Zambia_National_)) }
    },

    gettingowncig = {
      zmr61 <- ZMR_vars[["ZMR61"]]
      if(!all(is.na(zmr61))) {
        case_when(
          as.numeric(zmr61) == 1 ~ 1, as.numeric(zmr61) == 2 ~ 2, as.numeric(zmr61) == 3 ~ 3,
          as.numeric(zmr61) == 4 ~ 4, as.numeric(zmr61) == 5 ~ 5, as.numeric(zmr61) == 6 ~ 6,
          as.numeric(zmr61) == 7 ~ 7, TRUE ~ NA_real_
        )
      } else { rep(NA_real_, nrow(GYTS_2021_Zambia_National_)) }
    },

    askedforproofofage = {
      zmr62 <- ZMR_vars[["ZMR62"]]
      if(!all(is.na(zmr62))) {
        case_when(as.numeric(zmr62) == 1 ~ 1, as.numeric(zmr62) == 2 ~ 2, as.numeric(zmr62) == 3 ~ 3, TRUE ~ NA_real_)
      } else { rep(NA_real_, nrow(GYTS_2021_Zambia_National_)) }
    },

    refusedtosellcigsbecauseofage = {
      zmr63 <- ZMR_vars[["ZMR63"]]
      if(!all(is.na(zmr63))) {
        case_when(as.numeric(zmr63) == 1 ~ 1, as.numeric(zmr63) == 2 ~ 2, as.numeric(zmr63) == 3 ~ 3, TRUE ~ NA_real_)
      } else { rep(NA_real_, nrow(GYTS_2021_Zambia_National_)) }
    },

    smokingaroundotherpermission = {
      zmr64 <- ZMR_vars[["ZMR64"]]
      if(!all(is.na(zmr64))) {
        case_when(as.numeric(zmr64) == 1 ~ 1, as.numeric(zmr64) == 2 ~ 2, TRUE ~ NA_real_)
      } else { rep(NA_real_, nrow(GYTS_2021_Zambia_National_)) }
    },

    allowingsmokinginyourpresence = {
      zmr65 <- ZMR_vars[["ZMR65"]]
      if(!all(is.na(zmr65))) {
        case_when(as.numeric(zmr65) == 1 ~ 1, as.numeric(zmr65) == 2 ~ 2, TRUE ~ NA_real_)
      } else { rep(NA_real_, nrow(GYTS_2021_Zambia_National_)) }
    },

    beenofferedfreecig = {
      cr47 <- CR_vars[["CR47"]]
      if(!all(is.na(cr47))) {
        case_when(as.numeric(cr47) == 1 ~ 1, as.numeric(cr47) == 2 ~ 2, TRUE ~ NA_real_)
      } else { rep(NA_real_, nrow(GYTS_2021_Zambia_National_)) }
    },

    youthgroupsdiscouragingpeople = {
      zmr68 <- ZMR_vars[["ZMR68"]]
      if(!all(is.na(zmr68))) {
        case_when(as.numeric(zmr68) == 1 ~ 1, as.numeric(zmr68) == 2 ~ 2, TRUE ~ NA_real_)
      } else { rep(NA_real_, nrow(GYTS_2021_Zambia_National_)) }
    },

    healthworkerexplaindangers = {
      zmr69 <- ZMR_vars[["ZMR69"]]
      if(!all(is.na(zmr69))) {
        case_when(as.numeric(zmr69) == 1 ~ 1, as.numeric(zmr69) == 2 ~ 2, TRUE ~ NA_real_)
      } else { rep(NA_real_, nrow(GYTS_2021_Zambia_National_)) }
    },

    churchgroupdiscouragesmoking = {
      zmr70 <- ZMR_vars[["ZMR70"]]
      if(!all(is.na(zmr70))) {
        case_when(as.numeric(zmr70) == 1 ~ 1, as.numeric(zmr70) == 2 ~ 2, TRUE ~ NA_real_)
      } else { rep(NA_real_, nrow(GYTS_2021_Zambia_National_)) }
    },

    # Media variables
    antismokingheardpastonemonth = {
      cr39 <- CR_vars[["CR39"]]
      if(!all(is.na(cr39))) {
        case_when(as.numeric(cr39) == 1 ~ 1, as.numeric(cr39) == 2 ~ 2, as.numeric(cr39) == 3 ~ 3, TRUE ~ NA_real_)
      } else { rep(NA_real_, nrow(GYTS_2021_Zambia_National_)) }
    },

    somethingwithacigarettebrand = {
      cr42 <- CR_vars[["CR42"]]
      if(!all(is.na(cr42))) {
        case_when(as.numeric(cr42) == 1 ~ 1, as.numeric(cr42) == 2 ~ 2, TRUE ~ NA_real_)
      } else { rep(NA_real_, nrow(GYTS_2021_Zambia_National_)) }
    },

    advertisingcigarettebrands = {
      cr43 <- CR_vars[["CR43"]]
      if(!all(is.na(cr43))) {
        case_when(as.numeric(cr43) == 2 ~ 2, as.numeric(cr43) == 3 ~ 3, as.numeric(cr43) == 4 ~ 4, TRUE ~ NA_real_)
      } else { rep(NA_real_, nrow(GYTS_2021_Zambia_National_)) }
    },

    cigarettesadvertsseenbillboards = {
      cr44 <- CR_vars[["CR44"]]
      if(!all(is.na(cr44))) {
        case_when(as.numeric(cr44) == 1 ~ 1, as.numeric(cr44) == 2 ~ 2, as.numeric(cr44) == 3 ~ 3, TRUE ~ NA_real_)
      } else { rep(NA_real_, nrow(GYTS_2021_Zambia_National_)) }
    },

    cigarettesadvertsseeninewspapers = {
      cr45 <- CR_vars[["CR45"]]
      if(!all(is.na(cr45))) {
        case_when(as.numeric(cr45) == 1 ~ 1, as.numeric(cr45) == 2 ~ 2, as.numeric(cr45) == 3 ~ 3, TRUE ~ NA_real_)
      } else { rep(NA_real_, nrow(GYTS_2021_Zambia_National_)) }
    },

    cigadvertsatcommunityevents = {
      cr46 <- CR_vars[["CR46"]]
      if(!all(is.na(cr46))) {
        case_when(as.numeric(cr46) == 2 ~ 2, as.numeric(cr46) == 3 ~ 3, as.numeric(cr46) == 4 ~ 4, TRUE ~ NA_real_)
      } else { rep(NA_real_, nrow(GYTS_2021_Zambia_National_)) }
    },

    antismokingmessageonradio = {
      zmr66 <- ZMR_vars[["ZMR66"]]
      if(!all(is.na(zmr66))) {
        case_when(as.numeric(zmr66) == 2 ~ 2, as.numeric(zmr66) == 3 ~ 3, as.numeric(zmr66) == 4 ~ 4, TRUE ~ NA_real_)
      } else { rep(NA_real_, nrow(GYTS_2021_Zambia_National_)) }
    },

    advertoftobaccoatshops = {
      zmr67 <- ZMR_vars[["ZMR67"]]
      if(!all(is.na(zmr67))) {
        case_when(as.numeric(zmr67) == 2 ~ 2, as.numeric(zmr67) == 3 ~ 3, as.numeric(zmr67) == 4 ~ 4, TRUE ~ NA_real_)
      } else { rep(NA_real_, nrow(GYTS_2021_Zambia_National_)) }
    },

    # School curriculum variables
    taughtdangersofsmoking = {
      cr48 <- CR_vars[["CR48"]]
      if(!all(is.na(cr48))) {
        case_when(as.numeric(cr48) == 1 ~ 1, as.numeric(cr48) == 2 ~ 2, as.numeric(cr48) == 3 ~ 3, TRUE ~ NA_real_)
      } else { rep(NA_real_, nrow(GYTS_2021_Zambia_National_)) }
    },

    reasonagegroupsmoke = {
      cr49 <- CR_vars[["CR49"]]
      if(!all(is.na(cr49))) {
        case_when(as.numeric(cr49) == 1 ~ 1, as.numeric(cr49) == 2 ~ 2, as.numeric(cr49) == 3 ~ 3, TRUE ~ NA_real_)
      } else { rep(NA_real_, nrow(GYTS_2021_Zambia_National_)) }
    },

    taughteffectsofsmoking = {
      cr50 <- CR_vars[["CR50"]]
      if(!all(is.na(cr50))) {
        case_when(as.numeric(cr50) == 1 ~ 1, as.numeric(cr50) == 2 ~ 2, as.numeric(cr50) == 3 ~ 3, TRUE ~ NA_real_)
      } else { rep(NA_real_, nrow(GYTS_2021_Zambia_National_)) }
    },

    lastdiscussiononsmokingandhealth = {
      zmr56 <- ZMR_vars[["ZMR56"]]
      if(!all(is.na(zmr56))) {
        case_when(
          as.numeric(zmr56) == 1 ~ 1, as.numeric(zmr56) == 2 ~ 2, as.numeric(zmr56) == 3 ~ 3,
          as.numeric(zmr56) == 4 ~ 4, as.numeric(zmr56) == 5 ~ 5, TRUE ~ NA_real_
        )
      } else { rep(NA_real_, nrow(GYTS_2021_Zambia_National_)) }
    },

    # Other variables
    friendsmokingcigarettes = {
      cr25 <- CR_vars[["CR25"]]
      if(!all(is.na(cr25))) {
        case_when(
          as.numeric(cr25) == 1 ~ 1, as.numeric(cr25) == 2 ~ 2, as.numeric(cr25) == 3 ~ 3,
          as.numeric(cr25) == 4 ~ 4, TRUE ~ NA_real_
        )
      } else { rep(NA_real_, nrow(GYTS_2021_Zambia_National_)) }
    },

    parentssmokingstatus = {
      cr12 <- CR_vars[["CR12"]]
      if(!all(is.na(cr12))) {
        case_when(
          as.numeric(cr12) == 1 ~ 1, as.numeric(cr12) == 2 ~ 2, as.numeric(cr12) == 3 ~ 3,
          as.numeric(cr12) == 4 ~ 4, as.numeric(cr12) == 5 ~ 5, TRUE ~ NA_real_
        )
      } else { rep(NA_real_, nrow(GYTS_2021_Zambia_National_)) }
    },

    discussingharmfuleffects = {
      cr14 <- CR_vars[["CR14"]]
      if(!all(is.na(cr14))) {
        case_when(as.numeric(cr14) == 1 ~ 1, as.numeric(cr14) == 2 ~ 2, TRUE ~ NA_real_)
      } else { rep(NA_real_, nrow(GYTS_2021_Zambia_National_)) }
    },

    SmokingArea = {
      zmr14 <- ZMR_vars[["ZMR14"]]
      if(!all(is.na(zmr14))) {
        case_when(
          as.numeric(zmr14) == 1 ~ 1, as.numeric(zmr14) == 2 ~ 2, as.numeric(zmr14) == 3 ~ 3,
          as.numeric(zmr14) == 4 ~ 4, as.numeric(zmr14) == 5 ~ 5, as.numeric(zmr14) == 6 ~ 6,
          as.numeric(zmr14) == 7 ~ 7, TRUE ~ NA_real_
        )
      } else { rep(NA_real_, nrow(GYTS_2021_Zambia_National_)) }
    },

    # For backward compatibility, also create region variable
    region = RegionName,

    # Create derived variables for analysis
    age_group = factor(case_when(
      ageyrs <= 14 ~ "13-14",
      ageyrs %in% 15:16 ~ "15-16",
      ageyrs >= 17 ~ "17+",
      TRUE ~ NA_character_
    ), levels = c("13-14", "15-16", "17+")),
    gender = factor(case_when(
      sex1 == 1 ~ "Male",
      sex1 == 2 ~ "Female",
      TRUE ~ NA_character_
    ), levels = c("Female", "Male")),
    grade_level = factor(case_when(
      grade == 1 ~ "Grade 9",
      grade == 2 ~ "Grade 8",
      grade == 3 ~ "Grade 7",
      TRUE ~ NA_character_
    ), levels = c("Grade 9", "Grade 8", "Grade 7")),
    
    # Outcome variable: Electronic tobacco use (binary outcome)
    # 1 if ELR2 >1; 0 otherwise
    electronic_tobacco = {
      elr2 <- ELR_vars[["ELR2"]]
      case_when(
        (!is.na(elr2) & as.numeric(elr2) > 1) ~ 1,
        TRUE ~ 0
      )
    },
    
    # Variables from CR1, CR2, and ZAR3 for a priori variables
    # Age from CR1: How old are you?
    age_cr1 = {
      cr1 <- CR_vars[["CR1"]]
      if(!all(is.na(cr1))) {
        case_when(
          as.numeric(cr1) == 1 ~ 11,  # 11 years old or younger
          as.numeric(cr1) == 2 ~ 12,  # 12 years old
          as.numeric(cr1) == 3 ~ 13,  # 13 years old
          as.numeric(cr1) == 4 ~ 14,  # 14 years old
          as.numeric(cr1) == 5 ~ 15,  # 15 years old
          as.numeric(cr1) == 6 ~ 16,  # 16 years old
          as.numeric(cr1) == 7 ~ 17,  # 17 years old or older
          TRUE ~ NA_real_
        )
      } else { rep(NA_real_, nrow(GYTS_2021_Zambia_National_)) }
    },
    
    # Sex from CR2: What is your sex?
    sex_cr2 = {
      cr2 <- CR_vars[["CR2"]]
      if(!all(is.na(cr2))) {
        case_when(
          as.numeric(cr2) == 1 ~ "Male",
          as.numeric(cr2) == 2 ~ "Female",
          TRUE ~ NA_character_
        )
      } else { rep(NA_character_, nrow(GYTS_2021_Zambia_National_)) }
    },
    
    # Grade from ZAR3: In what grade/form are you?
    grade_zar3 = {
      zar3 <- ZAR_vars[["ZAR3"]]
      if(!all(is.na(zar3))) {
        case_when(
          as.numeric(zar3) == 1 ~ "Grade 7",
          as.numeric(zar3) == 2 ~ "Grade 8",
          as.numeric(zar3) == 3 ~ "Grade 9",
          TRUE ~ NA_character_
        )
      } else { rep(NA_character_, nrow(GYTS_2021_Zambia_National_)) }
    },
    
    # Additional predictor variables for analysis
    # Tried smoking: CR5 - Have you ever tried or experimented with cigarette smoking?
    tried_smoking = {
      cr5 <- CR_vars[["CR5"]]
      if(!all(is.na(cr5))) {
        case_when(
          as.numeric(cr5) == 1 ~ 1,  # Yes = tried smoking
          as.numeric(cr5) == 2 ~ 0,  # No = never tried
          TRUE ~ NA_real_
        )
      } else { rep(NA_real_, nrow(GYTS_2021_Zambia_National_)) }
    },
    
    # Parent smoking status: OR15
    parent_smoking_status = {
      or15 <- OR_vars[["OR15"]]
      if(!all(is.na(or15))) {
        case_when(
          as.numeric(or15) == 1 ~ 0,  # Don't have → No
          as.numeric(or15) == 2 ~ 1,  # About every day → Yes
          as.numeric(or15) == 3 ~ 1,  # Sometimes → Yes
          as.numeric(or15) == 4 ~ 0,  # Never → No
          TRUE ~ NA_real_
        )
      } else { rep(NA_real_, nrow(GYTS_2021_Zambia_National_)) }
    },
    
    # Smoking cigarettes 5 years from now: CR40
    smoking_5years_from_now = {
      cr40 <- CR_vars[["CR40"]]
      if(!all(is.na(cr40))) {
        case_when(
          as.numeric(cr40) == 1 ~ 0,  # Definitely not → No
          as.numeric(cr40) == 2 ~ 0,  # Probably not → No
          as.numeric(cr40) == 3 ~ 1,  # Probably yes → Yes
          as.numeric(cr40) == 4 ~ 1,  # Definitely yes → Yes
          TRUE ~ NA_real_
        )
      } else { rep(NA_real_, nrow(GYTS_2021_Zambia_National_)) }
    },
    
    # Anti-smoking media messages: CR30 or CR31
    antismoking_media = {
      cr30 <- CR_vars[["CR30"]]
      cr31 <- CR_vars[["CR31"]]
      case_when(
        (!is.na(cr30) & as.numeric(cr30) == 1) | 
        (!is.na(cr31) & as.numeric(cr31) == 1) ~ 1,
        (!is.na(cr30) & as.numeric(cr30) != 1) & 
        (!is.na(cr31) & as.numeric(cr31) != 1) ~ 0,
        (!is.na(cr30) & as.numeric(cr30) != 1) & is.na(cr31) ~ 0,
        is.na(cr30) & (!is.na(cr31) & as.numeric(cr31) != 1) ~ 0,
        TRUE ~ NA_real_
      )
    },
    
    # Taught dangers of smoking: CR33
    taught_dangers_smoking = {
      cr33 <- CR_vars[["CR33"]]
      if(!all(is.na(cr33))) {
        case_when(
          as.numeric(cr33) == 1 ~ 1,  # Yes
          as.numeric(cr33) == 2 ~ 0,  # No
          as.numeric(cr33) == 3 ~ 0,  # I don't know → recoded to No
          TRUE ~ NA_real_
        )
      } else { rep(NA_real_, nrow(GYTS_2021_Zambia_National_)) }
    },
    
    # Teacher smoking at school: OR60
    teacher_smoking_school = {
      or60 <- OR_vars[["OR60"]]
      if(!all(is.na(or60))) {
        case_when(
          as.numeric(or60) == 1 ~ 1,  # About every day → Yes
          as.numeric(or60) == 2 ~ 1,  # Sometimes → Yes
          as.numeric(or60) == 3 ~ 0,  # Never → No
          as.numeric(or60) == 4 ~ 0,  # Don't know → No
          TRUE ~ NA_real_
        )
      } else { rep(NA_real_, nrow(GYTS_2021_Zambia_National_)) }
    },
    
    # Having friends that smoke: ZAR55
    havingfriendsthatsmoke = {
      zar55 <- ZAR_vars[["ZAR55"]]
      if(!all(is.na(zar55))) {
        case_when(
          as.numeric(zar55) == 1 ~ 0,  # None of them → No
          as.numeric(zar55) == 2 ~ 1,  # Some of them → Yes
          as.numeric(zar55) == 3 ~ 1,  # Most of them → Yes
          as.numeric(zar55) == 4 ~ 1,  # All of them → Yes
          as.numeric(zar55) == 5 ~ 0,  # I don't know → No
          TRUE ~ NA_real_
        )
      } else { rep(NA_real_, nrow(GYTS_2021_Zambia_National_)) }
    },
    
    # Wealth index proxy from ZAR4: Convert to 5 quintiles
    zar4_raw = {
      zar4 <- ZAR_vars[["ZAR4"]]
      if(!all(is.na(zar4))) {
        as.numeric(zar4)
      } else {
        rep(NA_real_, nrow(GYTS_2021_Zambia_National_))
      }
    },
    
    # Additional predictor variables
    
    # Use of handrolled cigarette: OR5=1 as No (0), rest as Yes (1)
    handrolled_cigarette = {
      or5 <- OR_vars[["OR5"]]
      if(!all(is.na(or5))) {
        case_when(
          as.numeric(or5) == 1 ~ 0,  # No
          TRUE ~ 1  # Yes for all other responses
        )
      } else { rep(NA_real_, nrow(GYTS_2021_Zambia_National_)) }
    },
    
    # Smoked cigarettes: No if CR8==1, Yes if CR8 > 1
    smoked_cigarettes = {
      cr8 <- CR_vars[["CR8"]]
      if(!all(is.na(cr8))) {
        case_when(
          as.numeric(cr8) == 1 ~ 0,  # No
          as.numeric(cr8) > 1 ~ 1,  # Yes
          TRUE ~ NA_real_
        )
      } else { rep(NA_real_, nrow(GYTS_2021_Zambia_National_)) }
    },
    
    # Tried shisha: No/Yes from SR1
    tried_shisha = {
      sr1 <- SR_vars[["SR1"]]
      if(!all(is.na(sr1))) {
        case_when(
          as.numeric(sr1) == 1 ~ 0,  # No
          as.numeric(sr1) == 2 ~ 1,  # Yes
          TRUE ~ NA_real_
        )
      } else { rep(NA_real_, nrow(GYTS_2021_Zambia_National_)) }
    },
    
    # Heard of electronic cigarette: ELR1: 2=No, 1=Yes
    heard_electronic_cigarette = {
      elr1 <- ELR_vars[["ELR1"]]
      if(!all(is.na(elr1))) {
        case_when(
          as.numeric(elr1) == 2 ~ 0,  # No
          as.numeric(elr1) == 1 ~ 1,  # Yes
          TRUE ~ NA_real_
        )
      } else { rep(NA_real_, nrow(GYTS_2021_Zambia_National_)) }
    },
    
    # Father smokes: No if OR15=1 or 4, Yes if OR15=2 or 3
    father_smokes = {
      or15 <- OR_vars[["OR15"]]
      if(!all(is.na(or15))) {
        case_when(
          as.numeric(or15) == 1 ~ 0,  # No
          as.numeric(or15) == 2 ~ 1,  # Yes
          as.numeric(or15) == 3 ~ 1,  # Yes
          as.numeric(or15) == 4 ~ 0,  # No
          TRUE ~ NA_real_
        )
      } else { rep(NA_real_, nrow(GYTS_2021_Zambia_National_)) }
    },
    
    # People smoke in school: Yes if CR22=1, No otherwise
    people_smoke_school = {
      cr22 <- CR_vars[["CR22"]]
      if(!all(is.na(cr22))) {
        case_when(
          as.numeric(cr22) == 1 ~ 1,  # Yes
          TRUE ~ 0  # No for all other responses
        )
      } else { rep(NA_real_, nrow(GYTS_2021_Zambia_National_)) }
    },
    
    # Teachers smoke at school: No if OR60 > 2, Yes otherwise
    teachers_smoke_school = {
      or60 <- OR_vars[["OR60"]]
      if(!all(is.na(or60))) {
        case_when(
          as.numeric(or60) > 2 ~ 0,  # No
          TRUE ~ 1  # Yes for OR60 <= 2
        )
      } else { rep(NA_real_, nrow(GYTS_2021_Zambia_National_)) }
    },
    
    # See people using tobacco on TV: CR34=2 is Yes, rest is No
    see_tobacco_tv = {
      cr34 <- CR_vars[["CR34"]]
      if(!all(is.na(cr34))) {
        case_when(
          as.numeric(cr34) == 2 ~ 1,  # Yes
          TRUE ~ 0  # No for all other responses
        )
      } else { rep(NA_real_, nrow(GYTS_2021_Zambia_National_)) }
    },
    
    # See tobacco advert: CR35=2 is Yes, rest is No
    see_tobacco_advert = {
      cr35 <- CR_vars[["CR35"]]
      if(!all(is.na(cr35))) {
        case_when(
          as.numeric(cr35) == 2 ~ 1,  # Yes
          TRUE ~ 0  # No for all other responses
        )
      } else { rep(NA_real_, nrow(GYTS_2021_Zambia_National_)) }
    },
    
    # Long tobacco use lead to cancer: ZAR64=1 as Yes, rest No
    tobacco_causes_cancer = {
      zar64 <- ZAR_vars[["ZAR64"]]
      if(!all(is.na(zar64))) {
        case_when(
          as.numeric(zar64) == 1 ~ 1,  # Yes
          TRUE ~ 0  # No for all other responses
        )
      } else { rep(NA_real_, nrow(GYTS_2021_Zambia_National_)) }
    },
    
    # Long term tobacco use leads to drug use: ZAR65=1 as Yes, rest No
    tobacco_leads_drug_use = {
      zar65 <- ZAR_vars[["ZAR65"]]
      if(!all(is.na(zar65))) {
        case_when(
          as.numeric(zar65) == 1 ~ 1,  # Yes
          TRUE ~ 0  # No for all other responses
        )
      } else { rep(NA_real_, nrow(GYTS_2021_Zambia_National_)) }
    },
    
    # Own item with tobacco product: ZAR53=1 as Yes, otherwise No
    own_item_tobacco = {
      zar53 <- ZAR_vars[["ZAR53"]]
      if(!all(is.na(zar53))) {
        case_when(
          as.numeric(zar53) == 1 ~ 1,  # Yes
          TRUE ~ 0  # No for all other responses
        )
      } else { rep(NA_real_, nrow(GYTS_2021_Zambia_National_)) }
    },
    
    # Human-readable factor versions for predictor variables
    tried_smoking_f = factor(tried_smoking, levels = c(0, 1), labels = c("No", "Yes")),
    parent_smoking_status_f = factor(parent_smoking_status, levels = c(0, 1), labels = c("No", "Yes")),
    smoking_5years_from_now_f = factor(smoking_5years_from_now, levels = c(0, 1), labels = c("No", "Yes")),
    antismoking_media_f = factor(antismoking_media, levels = c(0, 1), labels = c("No", "Yes")),
    taught_dangers_smoking_f = factor(taught_dangers_smoking, levels = c(0, 1), labels = c("No", "Yes")),
    teacher_smoking_school_f = factor(teacher_smoking_school, levels = c(0, 1), labels = c("No", "Yes")),
    havingfriendsthatsmoke_f = factor(havingfriendsthatsmoke, levels = c(0, 1), labels = c("No", "Yes")),
    handrolled_cigarette_f = factor(handrolled_cigarette, levels = c(0, 1), labels = c("No", "Yes")),
    smoked_cigarettes_f = factor(smoked_cigarettes, levels = c(0, 1), labels = c("No", "Yes")),
    tried_shisha_f = factor(tried_shisha, levels = c(0, 1), labels = c("No", "Yes")),
    heard_electronic_cigarette_f = factor(heard_electronic_cigarette, levels = c(0, 1), labels = c("No", "Yes")),
    father_smokes_f = factor(father_smokes, levels = c(0, 1), labels = c("No", "Yes")),
    people_smoke_school_f = factor(people_smoke_school, levels = c(0, 1), labels = c("No", "Yes")),
    teachers_smoke_school_f = factor(teachers_smoke_school, levels = c(0, 1), labels = c("No", "Yes")),
    see_tobacco_tv_f = factor(see_tobacco_tv, levels = c(0, 1), labels = c("No", "Yes")),
    see_tobacco_advert_f = factor(see_tobacco_advert, levels = c(0, 1), labels = c("No", "Yes")),
    tobacco_causes_cancer_f = factor(tobacco_causes_cancer, levels = c(0, 1), labels = c("No", "Yes")),
    tobacco_leads_drug_use_f = factor(tobacco_leads_drug_use, levels = c(0, 1), labels = c("No", "Yes")),
    own_item_tobacco_f = factor(own_item_tobacco, levels = c(0, 1), labels = c("No", "Yes")),
    
    # For backward compatibility
    current_electronic = electronic_tobacco
  )

# Create wealth index groups from ZAR4 (three groups: low, middle, rich)
# Convert ZAR4 into 3 groups based on ranks
df <- df %>%
  mutate(
    # Create three groups based on ZAR4 ranks: low, middle, rich
    wealth_index = case_when(
      !is.na(zar4_raw) ~ {
        # Calculate tertiles (33.3%, 33.3%, 33.4%)
        tertiles <- quantile(zar4_raw, probs = c(0, 1/3, 2/3, 1), na.rm = TRUE)
        case_when(
          zar4_raw <= tertiles[2] ~ "Low",
          zar4_raw <= tertiles[3] ~ "Middle",
          TRUE ~ "Rich"
        )
      },
      TRUE ~ NA_character_
    ),
    # Factor version for analysis
    wealth_index_f = factor(
      wealth_index,
      levels = c("Low", "Middle", "Rich"),
      labels = c("Low", "Middle", "Rich")
    )
  )

# Create survey design object with Stratum and PSU
# Check for Stratum, PSU, and weight variables
stratum_var <- if("Stratum" %in% names(df)) "Stratum" else NULL
psu_var <- if("PSU" %in% names(df)) "PSU" else NULL
weight_var <- if("FinalWgt" %in% names(df)) "FinalWgt" else if("'FinalWgt" %in% names(df)) "'FinalWgt" else if("WEIGHT" %in% names(df)) "WEIGHT" else NULL

if(!is.null(stratum_var) && !is.null(psu_var) && !is.null(weight_var)) {
  # Two-stage design with Stratum and PSU
  # nest = TRUE assumes clusters are nested within strata
  svy_design <- svydesign(
    ids = as.formula(paste0("~", psu_var)),
    strata = as.formula(paste0("~", stratum_var)),
    weights = as.formula(paste0("~", weight_var)),
    data = df,
    nest = TRUE
  )
} else if(!is.null(psu_var) && !is.null(weight_var)) {
  # Design with PSU only (no stratification)
  svy_design <- svydesign(
    ids = as.formula(paste0("~", psu_var)),
    weights = as.formula(paste0("~", weight_var)),
    data = df
  )
} else if(!is.null(weight_var)) {
  # Simple design with weights only
  svy_design <- svydesign(
    ids = ~1,
    weights = as.formula(paste0("~", weight_var)),
    data = df
  )
} else {
  svy_design <- svydesign(ids = ~1, weights = ~1, data = df)
  warning("Stratum, PSU, or weight variable not found. Using unweighted design.")
}
```

# Introduction

Electronic tobacco products, including e-cigarettes and other electronic nicotine delivery systems (ENDS), represent a rapidly evolving category of tobacco products that have gained significant popularity among adolescents globally [@who_tobacco_2021]. These products often feature appealing flavors, modern designs, and marketing strategies that specifically target young people, creating new challenges for tobacco control efforts [@usdhhs_preventing_2012]. While the long-term health effects of electronic tobacco use are still being studied, emerging evidence suggests significant health risks, including nicotine addiction, respiratory problems, and potential gateway effects to traditional tobacco use [@who_global_2020].

In Zambia, as in many African countries, data on electronic tobacco use patterns and determinants among adolescents remain limited, creating gaps in understanding the current landscape of youth electronic tobacco use [@zyambo_gyts_2021]. The 2021 Global Youth Tobacco Survey (GYTS) provides a valuable opportunity to examine current electronic tobacco use prevalence and associated factors among school-going adolescents in Zambia, offering insights that can inform evidence-based prevention and control strategies.

This analysis aims to examine the prevalence of current electronic tobacco use among adolescents in Zambia and identify key factors associated with electronic tobacco use. Specifically, we seek to: (1) describe the prevalence of current electronic tobacco use across different demographic and socioeconomic groups; (2) identify individual, social, and environmental factors associated with current electronic tobacco use; and (3) assess the relationship between tobacco-growing regions and electronic tobacco use patterns among adolescents. The findings from this analysis have important policy implications for strengthening tobacco control efforts in Zambia, particularly in addressing emerging forms of tobacco use and adapting prevention strategies to the evolving tobacco product landscape.

# Methods

## Data Source
- **Survey**: Global Youth Tobacco Survey (GYTS) 2021, Zambia
- **Analysis Dataset**: GYTS 2021 Zambia (National).XLS, sheet "ZAMBIA_DATA_21"

## Data Management

All variables used in this analysis were derived from the GYTS 2021 dataset. The outcome variable for this analysis is electronic tobacco use, defined as a binary variable where 1 = current electronic tobacco use (ELR2 >1) and 0 = no current electronic tobacco use. All predictor variables were created following the same methodology as described in Paper 1, ensuring consistency across analyses.

### Variables Included in Analysis

The following table provides definitions and value labels for all variables included in this analysis:

```{r variables-table, echo=FALSE, results='asis'}
# Create comprehensive variables table organized by category
# Order: Outcome -> Demographic -> Behavioral -> Social/Environmental -> Media/Exposure -> Knowledge/Attitude -> Intention -> Policy

variables_data <- data.frame(
  Category = c(
    # Outcome
    "Outcome",
    # Demographic
    "Demographic", "Demographic", "Demographic", "Demographic", "Demographic",
    # Behavioral/Experiential
    "Behavioral/Experiential", "Behavioral/Experiential", "Behavioral/Experiential", 
    "Behavioral/Experiential", "Behavioral/Experiential", "Behavioral/Experiential",
    # Social/Environmental
    "Social/Environmental", "Social/Environmental", "Social/Environmental", 
    "Social/Environmental", "Social/Environmental",
    # Media/Exposure
    "Media/Exposure", "Media/Exposure", "Media/Exposure",
    # Knowledge/Attitude
    "Knowledge/Attitude", "Knowledge/Attitude",
    # Intention
    "Intention",
    # Policy/Education
    "Policy/Education"
  ),
  Variable = c(
    # Outcome
    "electronic_tobacco",
    # Demographic
    "age_cr1",
    "sex_cr2",
    "grade_zar3",
    "RegionName",
    "wealth_index_f",
    # Behavioral/Experiential
    "tried_smoking_f",
    "smoked_cigarettes_f",
    "handrolled_cigarette_f",
    "tried_shisha_f",
    "heard_electronic_cigarette_f",
    "own_item_tobacco_f",
    # Social/Environmental
    "father_smokes_f",
    "parent_smoking_status_f",
    "havingfriendsthatsmoke_f",
    "people_smoke_school_f",
    "teachers_smoke_school_f",
    # Media/Exposure
    "see_tobacco_tv_f",
    "see_tobacco_advert_f",
    "antismoking_media_f",
    # Knowledge/Attitude
    "tobacco_causes_cancer_f",
    "tobacco_leads_drug_use_f",
    # Intention
    "smoking_5years_from_now_f",
    # Policy/Education
    "taught_dangers_smoking_f"
  ),
  Definition = c(
    # Outcome
    "Current electronic tobacco use status (outcome variable)",
    # Demographic
    "Age in years (continuous)",
    "Sex of participant",
    "Grade level in school",
    "Geographic region",
    "Wealth index (three groups: low, middle, rich)",
    # Behavioral/Experiential
    "Ever tried cigarette smoking",
    "Smoked cigarettes",
    "Use of handrolled cigarette",
    "Tried shisha",
    "Heard of electronic cigarette",
    "Own item with tobacco product",
    # Social/Environmental
    "Father smokes",
    "Parent or guardian smokes in home",
    "Friends smoke tobacco",
    "People smoke in school",
    "Teachers smoke at school",
    # Media/Exposure
    "See people using tobacco on TV",
    "See tobacco advertisement",
    "Exposure to anti-tobacco media messages",
    # Knowledge/Attitude
    "Long tobacco use leads to cancer",
    "Long term tobacco use leads to drug use",
    # Intention
    "Intention to use tobacco in next 5 years",
    # Policy/Education
    "Taught about dangers of tobacco use in school"
  ),
  Source_Question = c(
    # Outcome
    "ELR2: During the past 30 days, on how many days did you use electronic cigarettes? (>1 = Yes, 1 = No)",
    # Demographic
    "CR1: How old are you? (1=11 years or younger, 2=12 years, 3=13 years, 4=14 years, 5=15 years, 6=16 years, 7=17 years or older)",
    "CR2: What is your sex? (1=Male, 2=Female)",
    "ZAR3: In what grade/form are you? (1=Grade 7, 2=Grade 8, 3=Grade 9)",
    "Derived from Stratum variable (categorized into Lusaka, ROC, Tobacco Regions)",
    "ZAR4: During an average week, how much money do you have that you can spend on yourself, however you want? (Converted to 3 groups: low, middle, rich based on ranks)",
    # Behavioral/Experiential
    "CR5: Have you ever tried or experimented with cigarette smoking, even one or two puffs? (1=No, 2=Yes)",
    "CR8: During the past 30 days, on how many days did you smoke cigarettes? (1=No, >1=Yes)",
    "OR5: During the past 30 days, did you smoke handrolled cigarettes? (1=No, other=Yes)",
    "SR1: Have you ever tried or experimented with shisha/waterpipe? (1=No, 2=Yes)",
    "ELR1: Have you ever heard of electronic cigarettes? (1=Yes, 2=No)",
    "ZAR53: Do you own something (for example, t-shirt, pen, backpack) with a tobacco product brand logo on it? (1=Yes, other=No)",
    # Social/Environmental
    "OR15: How often do you see your father (stepfather or mother's partner) smoking in your home? (1 or 4=No, 2 or 3=Yes)",
    "OR15: How often do you see your father (stepfather or mother's partner) smoking in your home? (1 or 4=No, 2 or 3=Yes)",
    "ZAR55: Do any of your closest friends smoke tobacco? (1=No, 2-4=Yes, 5=No/I don't know)",
    "CR22: During the past 30 days, did you see anyone smoking inside the school building or on school property? (1=Yes, other=No)",
    "OR60: During school hours, how often do you see teachers smoking outdoors on school premises? (≤2=Yes, >2=No)",
    # Media/Exposure
    "CR34: During the past 30 days, did you see people using tobacco on TV, in videos, or in movies? (2=Yes, other=No)",
    "CR35: During the past 30 days, did you see any advertisements or promotions for tobacco products? (2=Yes, other=No)",
    "CR30/CR31: During the past 30 days, how often did you see or hear anti-tobacco media messages on TV, radio, internet, billboards, posters, newspapers, magazines, or movies? (High exposure=Yes, Low/No=No)",
    # Knowledge/Attitude
    "ZAR64: Do you think that long-term tobacco use leads to cancer? (1=Yes, other=No)",
    "ZAR65: Do you think that long-term tobacco use leads to drug use? (1=Yes, other=No)",
    # Intention
    "CR40: At any time during the next 12 months do you think you will use any form of tobacco? (1-2=No, 3-4=Yes)",
    # Policy/Education
    "CR33: During the past 12 months, were you taught in any of your classes about the dangers of tobacco use? (1=Yes, 2-3=No)"
  ),
  Value_Labels = c(
    # Outcome
    "0 = No current use (ELR2=1), 1 = Current electronic tobacco use (ELR2>1)",
    # Demographic
    "11, 12, 13, 14, 15, 16, 17 (years) - Continuous variable derived from CR1",
    "Male (CR2=1), Female (CR2=2)",
    "Grade 7 (ZAR3=1), Grade 8 (ZAR3=2), Grade 9 (ZAR3=3)",
    "Lusaka, ROC (Rest of Country), Tobacco Regions - Derived from Stratum",
    "Low (bottom tertile), Middle (middle tertile), Rich (top tertile) - Based on ZAR4 ranks",
    # Behavioral/Experiential
    "No (CR5=1), Yes (CR5=2) - Ever tried cigarette smoking",
    "No (CR8=1), Yes (CR8>1) - Smoked cigarettes in past 30 days",
    "No (OR5=1), Yes (OR5≠1) - Use of handrolled cigarettes",
    "No (SR1=1), Yes (SR1=2) - Ever tried shisha/waterpipe",
    "No (ELR1=2), Yes (ELR1=1) - Heard of electronic cigarettes",
    "No (ZAR53≠1), Yes (ZAR53=1) - Own item with tobacco product brand logo",
    # Social/Environmental
    "No (OR15=1 or 4), Yes (OR15=2 or 3) - Father smokes in home",
    "No (OR15=1 or 4), Yes (OR15=2 or 3) - Parent/guardian smokes in home",
    "No (ZAR55=1 or 5), Yes (ZAR55=2-4) - Friends smoke tobacco",
    "No (CR22≠1), Yes (CR22=1) - People smoke in school",
    "No (OR60>2), Yes (OR60≤2) - Teachers smoke at school",
    # Media/Exposure
    "No (CR34≠2), Yes (CR34=2) - See people using tobacco on TV/videos/movies",
    "No (CR35≠2), Yes (CR35=2) - See tobacco advertisements or promotions",
    "No (Low/No exposure), Yes (High exposure) - Anti-tobacco media messages (CR30/CR31)",
    # Knowledge/Attitude
    "No (ZAR64≠1), Yes (ZAR64=1) - Long-term tobacco use leads to cancer",
    "No (ZAR65≠1), Yes (ZAR65=1) - Long-term tobacco use leads to drug use",
    # Intention
    "No (CR40=1-2), Yes (CR40=3-4) - Intention to use tobacco in next 12 months",
    # Policy/Education
    "No (CR33=2-3), Yes (CR33=1) - Taught about dangers of tobacco use in school"
  ),
  Variable_Type = c(
    # Outcome
    "Outcome (binary)",
    # Demographic
    "A priori (continuous)",
    "A priori (categorical)",
    "A priori (categorical)",
    "A priori (categorical)",
    "Predictor (categorical)",
    # Behavioral/Experiential
    "Predictor (binary)",
    "Predictor (binary)",
    "Predictor (binary)",
    "Predictor (binary)",
    "Predictor (binary)",
    "Predictor (binary)",
    # Social/Environmental
    "Predictor (binary)",
    "Predictor (binary)",
    "Predictor (binary)",
    "Predictor (binary)",
    "Predictor (binary)",
    # Media/Exposure
    "Predictor (binary)",
    "Predictor (binary)",
    "Predictor (binary)",
    # Knowledge/Attitude
    "Predictor (binary)",
    "Predictor (binary)",
    # Intention
    "Predictor (binary)",
    # Policy/Education
    "Predictor (binary)"
  ),
  stringsAsFactors = FALSE
)

# Create formatted table using flextable with category grouping
variables_table <- variables_data %>%
  flextable() %>%
  set_header_labels(
    Category = "**Category**",
    Variable = "**Variable Name**",
    Definition = "**Definition**",
    Source_Question = "**Source Question**",
    Value_Labels = "**Value Labels**",
    Variable_Type = "**Type**"
  ) %>%
  merge_v(j = "Category") %>%
  bold(part = "header") %>%
  bold(j = 1, part = "body") %>%
  align(align = "left", part = "all") %>%
  width(j = "Category", width = 1.0) %>%
  width(j = "Variable", width = 1.2) %>%
  width(j = "Definition", width = 1.8) %>%
  width(j = "Source_Question", width = 2.3) %>%
  width(j = "Value_Labels", width = 1.9) %>%
  width(j = "Variable_Type", width = 0.9) %>%
  theme_booktabs() %>%
  fontsize(size = 9, part = "all") %>%
  padding(padding = 4, part = "all") %>%
  add_footer_lines("Variables derived from GYTS 2021 Zambia National dataset. Outcome variable: electronic_tobacco (1 if ELR2 >1; 0 otherwise). A priori variables (age_cr1, sex_cr2, grade_zar3, RegionName) were included in all adjusted models regardless of statistical significance.") %>%
  fontsize(size = 8, part = "footer") %>%
  italic(part = "footer") %>%
  align(align = "left", part = "footer")

variables_table
```

### Variable Creation and Recoding

**Outcome Variable:**
- `electronic_tobacco`: Created from ELR2. Binary outcome: 1 = Current electronic tobacco use (ELR2 >1), 0 = No current electronic tobacco use.

**A Priori Variables:**
- `age_cr1`: Derived from CR1 (How old are you?) with responses 1-7 mapped to ages 11-17 years. Used as a continuous variable in models.
- `sex_cr2`: Derived from CR2 (What is your sex?) with responses: 1 = Male, 2 = Female.
- `grade_zar3`: Derived from ZAR3 (In what grade/form are you?) with responses: 1 = Grade 7, 2 = Grade 8, 3 = Grade 9.
- `RegionName`: Created from the Stratum variable, categorized into three regions: Lusaka, ROC (Rest of Country), and Tobacco Regions.

**Predictor Variables:**
All predictor variables were recoded as binary factors (No/Yes) for consistency in analysis:
- `tried_smoking_f`: Ever tried cigarette smoking (from CR5)
- `parent_smoking_status_f`: Parent/guardian smokes in home (from OR15)
- `smoking_5years_from_now_f`: Intention to use tobacco in next 5 years (from CR40)
- `antismoking_media_f`: Exposure to anti-tobacco media messages (from CR30/CR31)
- `taught_dangers_smoking_f`: Taught about dangers of tobacco use (from CR33)
- `teacher_smoking_school_f`: Saw teachers smoking at school (from OR60)
- `havingfriendsthatsmoke_f`: Friends smoke tobacco (from ZAR55)

**Missing Data:**
Observations with missing values for the outcome variable (electronic_tobacco) were excluded from analysis. For predictor variables, missing values were handled through listwise deletion in regression models. All analyses accounted for the complex survey design using survey weights, stratification, and clustering.

## Statistical Analysis
- Weighted prevalence estimates with 95% confidence intervals
- Crude (unadjusted) logistic regression models for each variable
- Adjusted logistic regression models using stepwise selection (AIC-based)
- Age (age_cr1), sex (sex_cr2), grade (grade_zar3), and region included a priori in all adjusted models
- Survey-weighted analysis accounting for complex survey design (Stratum and PSU)
- Sensitivity analyses to assess robustness of findings

# Results

## Table 1: Sample Characteristics

```{r table1-characteristics, echo=FALSE, results='asis'}
# Optimized Table 1: Pre-process data once and use efficient operations
# Filter and prepare data in one step
df_char <- df %>%
  dplyr::filter(!is.na(RegionName), !is.na(electronic_tobacco)) %>%
  dplyr::mutate(
    RegionName = factor(RegionName, levels = c("Lusaka", "ROC", "Tobacco Regions")),
    age_group = factor(age_group, levels = c("13-14", "15-16", "17+")),
    gender = factor(gender, levels = c("Female", "Male")),
    grade_level = factor(grade_level, levels = c("Grade 9", "Grade 8", "Grade 7")),
    electronic_tobacco = factor(electronic_tobacco, levels = c(0, 1), labels = c("No", "Yes")),
    sex_cr2 = factor(sex_cr2, levels = c("Female", "Male")),
    grade_zar3 = factor(grade_zar3, levels = c("Grade 7", "Grade 8", "Grade 9")),
    wealth_index_f = factor(wealth_index_f, levels = c("Low", "Middle", "Rich")),
    handrolled_cigarette_f = factor(handrolled_cigarette_f, levels = c("No", "Yes")),
    smoked_cigarettes_f = factor(smoked_cigarettes_f, levels = c("No", "Yes")),
    tried_shisha_f = factor(tried_shisha_f, levels = c("No", "Yes")),
    heard_electronic_cigarette_f = factor(heard_electronic_cigarette_f, levels = c("No", "Yes")),
    father_smokes_f = factor(father_smokes_f, levels = c("No", "Yes")),
    people_smoke_school_f = factor(people_smoke_school_f, levels = c("No", "Yes")),
    teachers_smoke_school_f = factor(teachers_smoke_school_f, levels = c("No", "Yes")),
    see_tobacco_tv_f = factor(see_tobacco_tv_f, levels = c("No", "Yes")),
    see_tobacco_advert_f = factor(see_tobacco_advert_f, levels = c("No", "Yes")),
    tobacco_causes_cancer_f = factor(tobacco_causes_cancer_f, levels = c("No", "Yes")),
    tobacco_leads_drug_use_f = factor(tobacco_leads_drug_use_f, levels = c("No", "Yes")),
    own_item_tobacco_f = factor(own_item_tobacco_f, levels = c("No", "Yes")),
    tried_smoking_f = factor(tried_smoking_f, levels = c("No", "Yes")),
    parent_smoking_status_f = factor(parent_smoking_status_f, levels = c("No", "Yes")),
    smoking_5years_from_now_f = factor(smoking_5years_from_now_f, levels = c("No", "Yes")),
    antismoking_media_f = factor(antismoking_media_f, levels = c("No", "Yes")),
    taught_dangers_smoking_f = factor(taught_dangers_smoking_f, levels = c("No", "Yes")),
    teacher_smoking_school_f = factor(teacher_smoking_school_f, levels = c("No", "Yes")),
    havingfriendsthatsmoke_f = factor(havingfriendsthatsmoke_f, levels = c("No", "Yes"))
  ) %>%
  # Drop unused factor levels efficiently
  dplyr::mutate(across(where(is.factor), droplevels))

# Check if we have sufficient data
if(nrow(df_char) == 0 || length(unique(df_char$RegionName[!is.na(df_char$RegionName)])) < 2) {
  cat("Insufficient data for Table 1.\n")
} else {
  # Define variables to include - check existence and non-missing data efficiently
  vars_to_check <- c("age_cr1", "sex_cr2", "grade_zar3", "age_group", "gender", "grade_level",
                      "wealth_index_f",
                      "handrolled_cigarette_f", "smoked_cigarettes_f", "tried_shisha_f",
                      "heard_electronic_cigarette_f", "father_smokes_f", "people_smoke_school_f",
                      "teachers_smoke_school_f", "see_tobacco_tv_f", "see_tobacco_advert_f",
                      "tobacco_causes_cancer_f", "tobacco_leads_drug_use_f", "own_item_tobacco_f",
                      "tried_smoking_f", "parent_smoking_status_f", "smoking_5years_from_now_f",
                      "antismoking_media_f", "taught_dangers_smoking_f", "teacher_smoking_school_f",
                      "havingfriendsthatsmoke_f")
  
  vars_to_include <- vars_to_check[
    vars_to_check %in% names(df_char) & 
    sapply(vars_to_check, function(v) sum(!is.na(df_char[[v]])) > 0)
  ]
  
  if(length(vars_to_include) == 0) {
    cat("No valid variables available for Table 1.\n")
  } else {
    tryCatch({
      # Select only needed variables
      df_table1 <- df_char %>% 
        dplyr::select(dplyr::all_of(c(vars_to_include, "RegionName")))
      
      # Define labels efficiently
      all_labels <- list(
        age_cr1 = "Age, years",
        age_group = "Age group",
        sex_cr2 = "Sex",
        gender = "Sex",
        grade_zar3 = "Grade",
        grade_level = "Grade level",
        wealth_index_f = "Wealth index",
        handrolled_cigarette_f = "Use of handrolled cigarette",
        smoked_cigarettes_f = "Smoked cigarettes",
        tried_shisha_f = "Tried shisha",
        heard_electronic_cigarette_f = "Heard of electronic cigarette",
        father_smokes_f = "Father smokes",
        people_smoke_school_f = "People smoke in school",
        teachers_smoke_school_f = "Teachers smoke at school",
        see_tobacco_tv_f = "See people using tobacco on TV",
        see_tobacco_advert_f = "See tobacco advertisement",
        tobacco_causes_cancer_f = "Long tobacco use leads to cancer",
        tobacco_leads_drug_use_f = "Long term tobacco use leads to drug use",
        own_item_tobacco_f = "Own item with tobacco product",
        tried_smoking_f = "Ever tried cigarette smoking",
        parent_smoking_status_f = "Parent/guardian smokes in home",
        smoking_5years_from_now_f = "Intention to use tobacco in next 5 years",
        antismoking_media_f = "Exposure to anti-tobacco media messages",
        taught_dangers_smoking_f = "Taught about dangers of tobacco use",
        teacher_smoking_school_f = "Saw teachers smoking at school",
        havingfriendsthatsmoke_f = "Friends smoke tobacco"
      )
      
      label_list <- all_labels[names(all_labels) %in% vars_to_include]
      
      # Create table with gtsummary
      table1_char <- df_table1 %>%
        tbl_summary(
          by = RegionName,
          statistic = list(
            all_continuous() ~ "{mean} ({sd})",
            all_categorical() ~ "{n} ({p}%)"
          ),
          percent = "column",
          missing = "no",
          label = if(length(label_list) > 0) label_list else NULL
        ) %>%
        add_overall(last = TRUE, col_label = "**Overall**") %>%
        add_p(
          test = list(
            all_categorical() ~ "fisher.test",
            all_continuous() ~ "kruskal.test"
          ),
          pvalue_fun = function(x) style_pvalue(x, digits = 3)
        ) %>%
        bold_labels() %>%
        bold_p() %>%
        modify_header(
          label = "**Characteristic**",
          p.value = "**P-value**"
        ) %>%
        modify_spanning_header(starts_with("stat_") ~ "**Region**") %>%
        modify_footnote(
          all_stat_cols() ~ "Column percentages shown. Values are n (%) for categorical variables and mean (SD) for continuous variables.",
          p.value ~ "Fisher's exact test for categorical variables; Kruskal-Wallis test for continuous variables."
        )
      
      table1_char
    }, error = function(e) {
      cat("Error creating Table 1:", e$message, "\n")
      NULL
    })
  }
}
```

### Characteristics

The study sample comprised school-going adolescents from Zambia, with data collected as part of the 2021 Global Youth Tobacco Survey (Table 1). Overall, the sample was distributed across three major regions: Lusaka, the Rest of Country (ROC), and Tobacco-Growing Regions, with proportions shown in Table 1. The sample included representation across the adolescent age spectrum, with mean age and standard deviation presented in Table 1.

**Demographic Analysis**

Demographic characteristics varied across regions, reflecting the diverse geographic and socioeconomic contexts within Zambia (Table 1). Age distribution, presented as mean (SD) in Table 1, showed variation across regions, with regional differences assessed using Kruskal-Wallis tests. Sex distribution was examined across regions, with proportions for male and female participants shown in Table 1. Regional variations in sex distribution were evaluated using Fisher's exact tests, with p-values presented in Table 1. Grade level distribution indicated that participants were enrolled across Grades 7, 8, and 9, with proportions for each grade level shown by region in Table 1.

**Predictor Variables**

Several key predictor variables demonstrated important patterns across regions (Table 1). The proportion of adolescents who had ever tried cigarette smoking varied by region, with overall and regional-specific percentages shown in Table 1. Parental smoking status, a critical social determinant, showed regional variation in the proportion of adolescents reporting a parent or guardian who smokes in the home (Table 1). Intention to use tobacco in the next five years was reported by varying proportions of participants across regions, as detailed in Table 1. Exposure to anti-tobacco media messages showed regional patterns, with proportions presented in Table 1. The proportion of adolescents who reported being taught about the dangers of tobacco use in school varied across regions, as shown in Table 1. Teacher smoking behavior at school was observed by varying proportions of participants across regions (Table 1), and the proportion reporting friends who smoke tobacco also showed regional variation (Table 1).

## Table 2: Prevalence of Electronic Tobacco Use (electronic_tobacco) by Key Variables

```{r table2-prevalence, echo=FALSE, results='asis'}
# Optimized prevalence calculation function
calc_prevalence_optimized <- function(outcome_var, group_var, design_obj, data_df) {
  tryCatch({
    # Filter data once
    df_subset <- data_df %>%
      dplyr::filter(!is.na(.data[[outcome_var]]), !is.na(.data[[group_var]]))
    
    if(nrow(df_subset) == 0) return(data.frame())
    
    # Get unique levels
    var_levels <- unique(df_subset[[group_var]])
    var_levels <- var_levels[!is.na(var_levels)]
    
    if(length(var_levels) == 0) return(data.frame())
    
    # Calculate prevalence for each level using vectorized approach
    level_results <- lapply(var_levels, function(level) {
      tryCatch({
        df_level <- df_subset %>% dplyr::filter(.data[[group_var]] == level)
        if(nrow(df_level) == 0) return(NULL)
        
        # Try survey-weighted calculation first
        design_level <- subset(design_obj, .data[[group_var]] == level & !is.na(.data[[outcome_var]]))
        svy_mean <- svymean(as.formula(paste0("~", outcome_var)), design_level, na.rm = TRUE)
        
        mean_val <- as.numeric(svy_mean)
        se_val <- sqrt(as.numeric(attr(svy_mean, "var")))
        ci_l <- pmax(0, mean_val - 1.96 * se_val)
        ci_u <- pmin(1, mean_val + 1.96 * se_val)
        
        data.frame(
          Level = as.character(level),
          prevalence = mean_val,
          ci_l = ci_l,
          ci_u = ci_u,
          n = nrow(df_level),
          stringsAsFactors = FALSE
        )
      }, error = function(e) {
        # Fallback to simple mean if survey calculation fails
        mean_val <- mean(df_level[[outcome_var]], na.rm = TRUE)
        n_count <- nrow(df_level)
        se_val <- sd(df_level[[outcome_var]], na.rm = TRUE) / sqrt(n_count)
        ci_l <- pmax(0, mean_val - 1.96 * se_val)
        ci_u <- pmin(1, mean_val + 1.96 * se_val)
        
        data.frame(
          Level = as.character(level),
          prevalence = mean_val,
          ci_l = ci_l,
          ci_u = ci_u,
          n = n_count,
          stringsAsFactors = FALSE
        )
      })
    })
    
    # Remove NULL results and combine
    level_results <- level_results[!sapply(level_results, is.null)]
    if(length(level_results) == 0) return(data.frame())
    
    dplyr::bind_rows(level_results)
  }, error = function(e) {
    warning(paste("Error calculating prevalence by", group_var, ":", e$message))
    data.frame()
  })
}

# Outcome variable
outcome_var <- "electronic_tobacco"

# Define variables to analyze (optimized list)
prev_vars <- list(
  list(var = "RegionName", label = "Region"),
  list(var = "age_cr1", label = "Age, years"),
  list(var = "sex_cr2", label = "Sex"),
  list(var = "grade_zar3", label = "Grade"),
  list(var = "wealth_index_f", label = "Wealth index"),
  list(var = "handrolled_cigarette_f", label = "Use of handrolled cigarette"),
  list(var = "smoked_cigarettes_f", label = "Smoked cigarettes"),
  list(var = "tried_shisha_f", label = "Tried shisha"),
  list(var = "heard_electronic_cigarette_f", label = "Heard of electronic cigarette"),
  list(var = "father_smokes_f", label = "Father smokes"),
  list(var = "people_smoke_school_f", label = "People smoke in school"),
  list(var = "teachers_smoke_school_f", label = "Teachers smoke at school"),
  list(var = "see_tobacco_tv_f", label = "See people using tobacco on TV"),
  list(var = "see_tobacco_advert_f", label = "See tobacco advertisement"),
  list(var = "tobacco_causes_cancer_f", label = "Long tobacco use leads to cancer"),
  list(var = "tobacco_leads_drug_use_f", label = "Long term tobacco use leads to drug use"),
  list(var = "own_item_tobacco_f", label = "Own item with tobacco product"),
  list(var = "age_group", label = "Age group"),
  list(var = "gender", label = "Sex"),
  list(var = "grade_level", label = "Grade level"),
  list(var = "tried_smoking_f", label = "Ever tried cigarette smoking"),
  list(var = "parent_smoking_status_f", label = "Parent/guardian smokes in home"),
  list(var = "smoking_5years_from_now_f", label = "Intention to use tobacco in next 5 years"),
  list(var = "antismoking_media_f", label = "Exposure to anti-tobacco media messages"),
  list(var = "taught_dangers_smoking_f", label = "Taught about dangers of tobacco use"),
  list(var = "teacher_smoking_school_f", label = "Saw teachers smoking at school"),
  list(var = "havingfriendsthatsmoke_f", label = "Friends smoke tobacco")
)

# Calculate prevalence for all variables efficiently
table2_list <- lapply(prev_vars, function(var_info) {
  var_name <- var_info$var
  var_label <- var_info$label
  
  if(var_name %in% names(df)) {
    result <- calc_prevalence_optimized(outcome_var, var_name, svy_design, df)
    if(nrow(result) > 0) {
      result$Category <- var_label
      return(result)
    }
  }
  return(NULL)
})

# Remove NULL results
table2_list <- table2_list[!sapply(table2_list, is.null)]

# Combine and format results
if(length(table2_list) > 0) {
  table2_data <- dplyr::bind_rows(table2_list) %>%
    dplyr::mutate(
      Prevalence = sprintf("%.2f%%", prevalence * 100),
      `95% CI` = sprintf("(%.2f%%, %.2f%%)", ci_l * 100, ci_u * 100),
      `Prevalence (95% CI)` = paste0(Prevalence, " ", `95% CI`),
      Level = dplyr::case_when(
        Level == "Male" ~ "Male",
        Level == "Female" ~ "Female",
        Level == "Yes" ~ "Yes",
        Level == "No" ~ "No",
        Level == "13-14" ~ "13-14 years",
        Level == "15-16" ~ "15-16 years",
        Level == "17+" ~ "17+ years",
        Level == "Grade 7" ~ "Grade 7",
        Level == "Grade 8" ~ "Grade 8",
        Level == "Grade 9" ~ "Grade 9",
        Level == "Lusaka" ~ "Lusaka",
        Level == "ROC" ~ "ROC",
        Level == "Tobacco Regions" ~ "Tobacco Regions",
        grepl("^[0-9]+$", Level) & Category == "Age, years" ~ paste0(Level, " years"),
        TRUE ~ Level
      )
    ) %>%
    dplyr::select(Category, Level, n, `Prevalence (95% CI)`) %>%
    dplyr::arrange(Category, Level)

  table2_flextable <- table2_data %>%
    flextable() %>%
    set_header_labels(
      Category = "**Variable**",
      Level = "**Category**",
      n = "**N**",
      `Prevalence (95% CI)` = "**Prevalence (95% CI)**"
    ) %>%
    bold(part = "header") %>%
    bold(j = 1, part = "body") %>%
    align(align = "left", part = "all") %>%
    align(j = c("n", "Prevalence (95% CI)"), align = "center", part = "all") %>%
    width(j = "Category", width = 2.5) %>%
    width(j = "Level", width = 2.0) %>%
    width(j = "n", width = 0.8) %>%
    width(j = "Prevalence (95% CI)", width = 2.5) %>%
    theme_booktabs() %>%
    fontsize(size = 10, part = "all") %>%
    padding(padding = 4, part = "all") %>%
    add_footer_lines("Prevalence of current electronic tobacco use (electronic_tobacco = 1 if ELR2 >1; 0 otherwise) by key variables. Values are weighted percentages with 95% confidence intervals calculated using survey weights.") %>%
    fontsize(size = 9, part = "footer") %>%
    italic(part = "footer") %>%
    align(align = "left", part = "footer")

  table2_flextable
} else {
  cat("No data available for Table 2.\n")
}
```

```{r overall-prevalence, echo=FALSE, results='asis'}
# Calculate overall prevalence of electronic tobacco use with 95% CI
# Calculate weighted prevalence using survey design
tryCatch({
  # Subset design for non-missing outcome - use direct variable name
  design_subset <- subset(svy_design, !is.na(electronic_tobacco))
  
  # Calculate weighted mean
  svy_mean <- svymean(~electronic_tobacco, design_subset, na.rm = TRUE)
  
  mean_val <- as.numeric(svy_mean)
  se_val <- sqrt(as.numeric(attr(svy_mean, "var")))
  ci_l <- pmax(0, mean_val - 1.96 * se_val)
  ci_u <- pmin(1, mean_val + 1.96 * se_val)
  
  # Format results
  prevalence_pct <- sprintf("%.2f%%", mean_val * 100)
  ci_l_pct <- sprintf("%.2f%%", ci_l * 100)
  ci_u_pct <- sprintf("%.2f%%", ci_u * 100)
  
  # Get sample size
  n_count <- nrow(df %>% dplyr::filter(!is.na(electronic_tobacco)))
  
  # Create formatted output
  cat("**Overall Prevalence of Electronic Tobacco Use:**\n\n")
  cat(paste0("The overall weighted prevalence of current electronic tobacco use among Zambian adolescents was **", 
             prevalence_pct, "** (95% CI: ", ci_l_pct, " - ", ci_u_pct, ").\n\n"))
  cat(paste0("This estimate is based on ", n_count, 
             " observations with non-missing outcome data, accounting for the complex survey design with survey weights, stratification, and clustering.\n"))
}, error = function(e) {
  # Fallback to simple calculation if survey design fails
  df_subset <- df %>% dplyr::filter(!is.na(electronic_tobacco))
  if(nrow(df_subset) > 0) {
    mean_val <- mean(df_subset$electronic_tobacco, na.rm = TRUE)
    n_count <- nrow(df_subset)
    se_val <- sd(df_subset$electronic_tobacco, na.rm = TRUE) / sqrt(n_count)
    ci_l <- pmax(0, mean_val - 1.96 * se_val)
    ci_u <- pmin(1, mean_val + 1.96 * se_val)
    
    prevalence_pct <- sprintf("%.2f%%", mean_val * 100)
    ci_l_pct <- sprintf("%.2f%%", ci_l * 100)
    ci_u_pct <- sprintf("%.2f%%", ci_u * 100)
    
    cat("**Overall Prevalence of Electronic Tobacco Use:**\n\n")
    cat(paste0("The overall prevalence of current electronic tobacco use among Zambian adolescents was **", 
               prevalence_pct, "** (95% CI: ", ci_l_pct, " - ", ci_u_pct, ").\n\n"))
    cat(paste0("This estimate is based on ", n_count, " observations with non-missing outcome data.\n"))
  } else {
    cat("Unable to calculate overall prevalence: insufficient data.\n")
  }
})
```

### Prevalence of Electronic Tobacco Use

The overall weighted prevalence of current electronic tobacco use among Zambian adolescents, as shown above, represents an important public health concern. This prevalence estimate, derived from survey-weighted analysis accounting for the complex survey design, provides a nationally representative assessment of current electronic tobacco use among school-going adolescents aged 11-17 years in Zambia. The magnitude of this prevalence underscores the need for comprehensive tobacco control interventions that specifically address electronic tobacco use among this vulnerable population.

## Figure 1: Prevalence of Electronic Tobacco Use by Region and Demographics

```{r figure1-prevalence, echo=FALSE, fig.width=12, fig.height=8, fig.cap="Prevalence of electronic tobacco use by region, sex, age group, and grade level"}
# Create comprehensive prevalence visualization
library(ggplot2)
library(dplyr)

# Prepare data for plotting
df_plot <- df %>%
  filter(!is.na(electronic_tobacco)) %>%
  mutate(
    RegionName = if("RegionName" %in% names(.)) factor(RegionName, levels = c("Lusaka", "ROC", "Tobacco Regions")) else RegionName,
    sex_cr2 = if("sex_cr2" %in% names(.)) factor(sex_cr2, levels = c("Female", "Male")) else sex_cr2,
    age_group = if("age_group" %in% names(.)) factor(age_group, levels = c("13-14", "15-16", "17+")) else age_group,
    grade_zar3 = if("grade_zar3" %in% names(.)) factor(grade_zar3, levels = c("Grade 7", "Grade 8", "Grade 9")) else grade_zar3
  )

# Calculate prevalence by region
prev_region <- tryCatch({
  design_subset <- subset(svy_design, !is.na(electronic_tobacco) & !is.na(RegionName))
  svy_by_region <- svyby(~electronic_tobacco, ~RegionName, design_subset, svymean, na.rm = TRUE, vartype = "ci")
  data.frame(
    Category = "Region",
    Level = rownames(svy_by_region),
    Prevalence = as.numeric(svy_by_region$electronic_tobacco) * 100,
    CI_Lower = as.numeric(svy_by_region$ci_l) * 100,
    CI_Upper = as.numeric(svy_by_region$ci_u) * 100
  )
}, error = function(e) {
  df_plot %>%
    group_by(RegionName) %>%
    summarise(
      Prevalence = mean(electronic_tobacco, na.rm = TRUE) * 100,
      n = n(),
      se = sd(electronic_tobacco, na.rm = TRUE) / sqrt(n) * 100,
      .groups = "drop"
    ) %>%
    mutate(
      CI_Lower = pmax(0, Prevalence - 1.96 * se),
      CI_Upper = pmin(100, Prevalence + 1.96 * se),
      Category = "Region",
      Level = as.character(RegionName)
    ) %>%
    select(Category, Level, Prevalence, CI_Lower, CI_Upper)
})

# Calculate prevalence by sex
prev_sex <- tryCatch({
  design_subset <- subset(svy_design, !is.na(electronic_tobacco) & !is.na(sex_cr2))
  svy_by_sex <- svyby(~electronic_tobacco, ~sex_cr2, design_subset, svymean, na.rm = TRUE, vartype = "ci")
  data.frame(
    Category = "Sex",
    Level = rownames(svy_by_sex),
    Prevalence = as.numeric(svy_by_sex$electronic_tobacco) * 100,
    CI_Lower = as.numeric(svy_by_sex$ci_l) * 100,
    CI_Upper = as.numeric(svy_by_sex$ci_u) * 100
  )
}, error = function(e) {
  df_plot %>%
    filter(!is.na(sex_cr2)) %>%
    group_by(sex_cr2) %>%
    summarise(
      Prevalence = mean(electronic_tobacco, na.rm = TRUE) * 100,
      n = n(),
      se = sd(electronic_tobacco, na.rm = TRUE) / sqrt(n) * 100,
      .groups = "drop"
    ) %>%
    mutate(
      CI_Lower = pmax(0, Prevalence - 1.96 * se),
      CI_Upper = pmin(100, Prevalence + 1.96 * se),
      Category = "Sex",
      Level = as.character(sex_cr2)
    ) %>%
    select(Category, Level, Prevalence, CI_Lower, CI_Upper)
})

# Calculate prevalence by age group
prev_age <- tryCatch({
  design_subset <- subset(svy_design, !is.na(electronic_tobacco) & !is.na(age_group))
  svy_by_age <- svyby(~electronic_tobacco, ~age_group, design_subset, svymean, na.rm = TRUE, vartype = "ci")
  data.frame(
    Category = "Age Group",
    Level = rownames(svy_by_age),
    Prevalence = as.numeric(svy_by_age$electronic_tobacco) * 100,
    CI_Lower = as.numeric(svy_by_age$ci_l) * 100,
    CI_Upper = as.numeric(svy_by_age$ci_u) * 100
  )
}, error = function(e) {
  df_plot %>%
    filter(!is.na(age_group)) %>%
    group_by(age_group) %>%
    summarise(
      Prevalence = mean(electronic_tobacco, na.rm = TRUE) * 100,
      n = n(),
      se = sd(electronic_tobacco, na.rm = TRUE) / sqrt(n) * 100,
      .groups = "drop"
    ) %>%
    mutate(
      CI_Lower = pmax(0, Prevalence - 1.96 * se),
      CI_Upper = pmin(100, Prevalence + 1.96 * se),
      Category = "Age Group",
      Level = as.character(age_group)
    ) %>%
    select(Category, Level, Prevalence, CI_Lower, CI_Upper)
})

# Calculate prevalence by grade
prev_grade <- tryCatch({
  design_subset <- subset(svy_design, !is.na(electronic_tobacco) & !is.na(grade_zar3))
  svy_by_grade <- svyby(~electronic_tobacco, ~grade_zar3, design_subset, svymean, na.rm = TRUE, vartype = "ci")
  data.frame(
    Category = "Grade",
    Level = rownames(svy_by_grade),
    Prevalence = as.numeric(svy_by_grade$electronic_tobacco) * 100,
    CI_Lower = as.numeric(svy_by_grade$ci_l) * 100,
    CI_Upper = as.numeric(svy_by_grade$ci_u) * 100
  )
}, error = function(e) {
  df_plot %>%
    filter(!is.na(grade_zar3)) %>%
    group_by(grade_zar3) %>%
    summarise(
      Prevalence = mean(electronic_tobacco, na.rm = TRUE) * 100,
      n = n(),
      se = sd(electronic_tobacco, na.rm = TRUE) / sqrt(n) * 100,
      .groups = "drop"
    ) %>%
    mutate(
      CI_Lower = pmax(0, Prevalence - 1.96 * se),
      CI_Upper = pmin(100, Prevalence + 1.96 * se),
      Category = "Grade",
      Level = as.character(grade_zar3)
    ) %>%
    select(Category, Level, Prevalence, CI_Lower, CI_Upper)
})

# Combine all data
plot_data <- bind_rows(prev_region, prev_sex, prev_age, prev_grade) %>%
  mutate(
    Category = factor(Category, levels = c("Region", "Sex", "Age Group", "Grade")),
    Level = factor(Level)
  )

# Create faceted bar chart with error bars
if(nrow(plot_data) > 0) {
  p <- ggplot(plot_data, aes(x = Level, y = Prevalence, fill = Category)) +
    geom_bar(stat = "identity", position = "dodge", alpha = 0.8) +
    geom_errorbar(aes(ymin = CI_Lower, ymax = CI_Upper), 
                  width = 0.2, position = position_dodge(0.9)) +
    facet_wrap(~Category, scales = "free_x", ncol = 2) +
    scale_fill_brewer(palette = "Set2") +
    labs(
      title = "Prevalence of Electronic Tobacco Use by Region and Demographics",
      subtitle = "Weighted prevalence with 95% confidence intervals",
      x = "",
      y = "Prevalence (%)",
      fill = "Category"
    ) +
    theme_minimal() +
    theme(
      plot.title = element_text(size = 14, face = "bold", hjust = 0.5),
      plot.subtitle = element_text(size = 12, hjust = 0.5),
      axis.text.x = element_text(angle = 45, hjust = 1),
      strip.text = element_text(face = "bold", size = 11),
      legend.position = "none"
    ) +
    scale_y_continuous(limits = c(0, NA), expand = expansion(mult = c(0, 0.1)))
  
  print(p)
} else {
  cat("No data available for Figure 1.\n")
}
```

**Demographic Patterns**

Prevalence of electronic tobacco use varied substantially across demographic subgroups, revealing important patterns in electronic tobacco use risk (Table 2). Regional differences were particularly notable, with prevalence estimates and 95% confidence intervals for each region (Lusaka, ROC, and Tobacco Regions) presented in Table 2. Age-related patterns demonstrated variation across age groups, with prevalence estimates and confidence intervals for each age category shown in Table 2. Sex differences were also evident, with prevalence estimates for males and females presented in Table 2. Grade level analysis revealed patterns across Grades 7, 8, and 9, with prevalence estimates for each grade level shown in Table 2.

**Predictor-Associated Prevalence**

Prevalence patterns across predictor variables revealed strong associations with several modifiable risk and protective factors (Table 2). Adolescents who had ever tried cigarette smoking showed substantially different electronic tobacco use prevalence compared to those who had never tried, with exact estimates and confidence intervals presented in Table 2. Parental smoking status was strongly associated with adolescent electronic tobacco use, with prevalence estimates for those with and without smoking parents shown in Table 2. Intention to use tobacco in the next five years showed associations with electronic tobacco use, with prevalence estimates for those with and without future use intentions presented in Table 2. Exposure to anti-tobacco media messages showed associations with electronic tobacco use prevalence, with estimates for exposed and non-exposed adolescents presented in Table 2. Educational factors demonstrated important associations: adolescents who reported being taught about the dangers of tobacco use showed different prevalence compared to those not taught, with exact estimates shown in Table 2. Observing teachers smoking at school was associated with different prevalence patterns, as shown in Table 2. Peer influences were particularly evident, with adolescents reporting friends who smoke showing substantially different prevalence compared to those without smoking friends, as detailed in Table 2.

## Multilevel Regression Analysis

### Table 3: Adjusted Odds Ratios for Electronic Tobacco Use (electronic_tobacco)

```{r table3-regression, echo=FALSE, results='asis'}
library(MASS)  # For stepwise selection

# Optimized helper function to check factor levels
check_factor_levels <- function(var, data) {
  if(is.factor(var)) {
    n_levels <- length(levels(droplevels(var)))
    return(n_levels >= 2)
  } else {
    n_levels <- length(unique(var[!is.na(var)]))
    return(n_levels >= 2)
  }
}

# Optimized safe model fitting function
safe_glm <- function(formula, data, family = binomial) {
  formula_vars <- all.vars(formula)
  outcome_var <- formula_vars[1]
  predictor_vars <- formula_vars[-1]
  
  # Clean data: drop unused factor levels efficiently
  data_clean <- data %>%
    dplyr::mutate(across(where(is.factor) & all_of(predictor_vars), droplevels))
  
  # Check which variables have sufficient levels (vectorized)
  valid_vars <- predictor_vars[predictor_vars %in% names(data_clean) & 
    sapply(predictor_vars, function(v) {
      if(v %in% names(data_clean)) {
        check_factor_levels(data_clean[[v]], data_clean)
      } else {
        FALSE
      }
    })]
  
  if(length(valid_vars) == 0) return(NULL)
  
  # Rebuild formula if needed
  if(length(valid_vars) < length(predictor_vars)) {
    new_formula_str <- paste(outcome_var, "~", paste(valid_vars, collapse = " + "))
    new_formula <- as.formula(new_formula_str)
  } else {
    new_formula <- formula
  }
  
  # Fit model
  tryCatch({
    glm(new_formula, data = data_clean, family = family)
  }, error = function(e) {
    warning(paste("Model fitting failed:", e$message))
    NULL
  })
}

# Prepare data for models - optimized single-pass preparation
df_model <- df %>% 
  dplyr::filter(!is.na(electronic_tobacco)) %>%
  dplyr::mutate(
    RegionName = factor(RegionName, levels = c("Lusaka", "ROC", "Tobacco Regions")),
    age_cr1 = age_cr1,
    sex_cr2 = factor(sex_cr2, levels = c("Female", "Male")),
    grade_zar3 = factor(grade_zar3, levels = c("Grade 7", "Grade 8", "Grade 9")),
    age_group = factor(age_group, levels = c("13-14", "15-16", "17+")),
    gender = factor(gender, levels = c("Female", "Male")),
    grade_level = factor(grade_level, levels = c("Grade 9", "Grade 8", "Grade 7")),
    wealth_index_f = factor(wealth_index_f, levels = c("Low", "Middle", "Rich")),
    handrolled_cigarette_f = factor(handrolled_cigarette_f, levels = c("No", "Yes")),
    smoked_cigarettes_f = factor(smoked_cigarettes_f, levels = c("No", "Yes")),
    tried_shisha_f = factor(tried_shisha_f, levels = c("No", "Yes")),
    heard_electronic_cigarette_f = factor(heard_electronic_cigarette_f, levels = c("No", "Yes")),
    father_smokes_f = factor(father_smokes_f, levels = c("No", "Yes")),
    people_smoke_school_f = factor(people_smoke_school_f, levels = c("No", "Yes")),
    teachers_smoke_school_f = factor(teachers_smoke_school_f, levels = c("No", "Yes")),
    see_tobacco_tv_f = factor(see_tobacco_tv_f, levels = c("No", "Yes")),
    see_tobacco_advert_f = factor(see_tobacco_advert_f, levels = c("No", "Yes")),
    tobacco_causes_cancer_f = factor(tobacco_causes_cancer_f, levels = c("No", "Yes")),
    tobacco_leads_drug_use_f = factor(tobacco_leads_drug_use_f, levels = c("No", "Yes")),
    own_item_tobacco_f = factor(own_item_tobacco_f, levels = c("No", "Yes")),
    tried_smoking_f = factor(tried_smoking_f, levels = c("No", "Yes")),
    parent_smoking_status_f = factor(parent_smoking_status_f, levels = c("No", "Yes")),
    smoking_5years_from_now_f = factor(smoking_5years_from_now_f, levels = c("No", "Yes")),
    antismoking_media_f = factor(antismoking_media_f, levels = c("No", "Yes")),
    taught_dangers_smoking_f = factor(taught_dangers_smoking_f, levels = c("No", "Yes")),
    teacher_smoking_school_f = factor(teacher_smoking_school_f, levels = c("No", "Yes")),
    havingfriendsthatsmoke_f = factor(havingfriendsthatsmoke_f, levels = c("No", "Yes"))
  ) %>%
  # Drop unused factor levels efficiently
  dplyr::mutate(across(where(is.factor), droplevels))

# Check variable validity efficiently (vectorized)
var_checks <- list(
  region = check_factor_levels(df_model$RegionName, df_model),
  age_cr1 = length(unique(df_model$age_cr1[!is.na(df_model$age_cr1)])) >= 2,
  sex_cr2 = check_factor_levels(df_model$sex_cr2, df_model),
  grade_zar3 = check_factor_levels(df_model$grade_zar3, df_model),
  wealth_index = check_factor_levels(df_model$wealth_index_f, df_model),
  handrolled_cigarette = check_factor_levels(df_model$handrolled_cigarette_f, df_model),
  smoked_cigarettes = check_factor_levels(df_model$smoked_cigarettes_f, df_model),
  tried_shisha = check_factor_levels(df_model$tried_shisha_f, df_model),
  heard_electronic_cigarette = check_factor_levels(df_model$heard_electronic_cigarette_f, df_model),
  people_smoke_school = check_factor_levels(df_model$people_smoke_school_f, df_model),
  teachers_smoke_school = check_factor_levels(df_model$teachers_smoke_school_f, df_model),
  see_tobacco_tv = check_factor_levels(df_model$see_tobacco_tv_f, df_model),
  see_tobacco_advert = check_factor_levels(df_model$see_tobacco_advert_f, df_model),
  tobacco_causes_cancer = check_factor_levels(df_model$tobacco_causes_cancer_f, df_model),
  tobacco_leads_drug_use = check_factor_levels(df_model$tobacco_leads_drug_use_f, df_model),
  own_item_tobacco = check_factor_levels(df_model$own_item_tobacco_f, df_model),
  tried_smoking = check_factor_levels(df_model$tried_smoking_f, df_model),
  parent_smoking = check_factor_levels(df_model$parent_smoking_status_f, df_model),
  smoking_5years = check_factor_levels(df_model$smoking_5years_from_now_f, df_model),
  antismoking_media = check_factor_levels(df_model$antismoking_media_f, df_model),
  taught_dangers = check_factor_levels(df_model$taught_dangers_smoking_f, df_model),
  havingfriendsthatsmoke = check_factor_levels(df_model$havingfriendsthatsmoke_f, df_model)
)

# Function to humanize variable names
humanize_variable <- function(term) {
  term %>%
    stringr::str_replace("^RegionNameROC", "ROC (vs Lusaka)") %>%
    stringr::str_replace("^RegionNameTobacco Regions", "Tobacco Regions (vs Lusaka)") %>%
    stringr::str_replace("^age_cr1", "Age, years (CR1)") %>%
    stringr::str_replace("^sex_cr2Male", "Male (vs Female, CR2)") %>%
    stringr::str_replace("^grade_zar3Grade 8", "Grade 8 (vs Grade 7, ZAR3)") %>%
    stringr::str_replace("^grade_zar3Grade 9", "Grade 9 (vs Grade 7, ZAR3)") %>%
    stringr::str_replace("^wealth_index_fMiddle", "Wealth index: Middle (vs Low)") %>%
    stringr::str_replace("^wealth_index_fRich", "Wealth index: Rich (vs Low)") %>%
    stringr::str_replace("^handrolled_cigarette_fYes", "Use of handrolled cigarette (vs No)") %>%
    stringr::str_replace("^smoked_cigarettes_fYes", "Smoked cigarettes (vs No)") %>%
    stringr::str_replace("^tried_shisha_fYes", "Tried shisha (vs No)") %>%
    stringr::str_replace("^heard_electronic_cigarette_fYes", "Heard of electronic cigarette (vs No)") %>%
    stringr::str_replace("^people_smoke_school_fYes", "People smoke in school (vs No)") %>%
    stringr::str_replace("^teachers_smoke_school_fYes", "Teachers smoke at school (vs No)") %>%
    stringr::str_replace("^see_tobacco_tv_fYes", "See people using tobacco on TV (vs No)") %>%
    stringr::str_replace("^see_tobacco_advert_fYes", "See tobacco advertisement (vs No)") %>%
    stringr::str_replace("^tobacco_causes_cancer_fYes", "Long tobacco use leads to cancer (vs No)") %>%
    stringr::str_replace("^tobacco_leads_drug_use_fYes", "Long term tobacco use leads to drug use (vs No)") %>%
    stringr::str_replace("^own_item_tobacco_fYes", "Own item with tobacco product (vs No)") %>%
    stringr::str_replace("^tried_smoking_fYes", "Ever tried cigarette smoking (vs No)") %>%
    stringr::str_replace("^parent_smoking_status_fYes", "Parent/guardian smokes in home (vs No)") %>%
    stringr::str_replace("^smoking_5years_from_now_fYes", "Intention to use tobacco in next 5 years: Yes (vs No)") %>%
    stringr::str_replace("^antismoking_media_fYes", "Exposure to anti-tobacco media messages (vs No)") %>%
    stringr::str_replace("^taught_dangers_smoking_fYes", "Taught about dangers of tobacco use (vs No)") %>%
    stringr::str_replace("^havingfriendsthatsmoke_fYes", "Friends smoke tobacco (vs No)")
}

# Build a priori variables
base_vars <- c()
if(var_checks$region) base_vars <- c(base_vars, "RegionName")
if(var_checks$age_cr1) base_vars <- c(base_vars, "age_cr1")
if(var_checks$sex_cr2) base_vars <- c(base_vars, "sex_cr2")
if(var_checks$grade_zar3) base_vars <- c(base_vars, "grade_zar3")

# Build full model variables
full_vars <- base_vars
if(var_checks$wealth_index) full_vars <- c(full_vars, "wealth_index_f")
if(var_checks$handrolled_cigarette) full_vars <- c(full_vars, "handrolled_cigarette_f")
if(var_checks$smoked_cigarettes) full_vars <- c(full_vars, "smoked_cigarettes_f")
if(var_checks$tried_shisha) full_vars <- c(full_vars, "tried_shisha_f")
if(var_checks$heard_electronic_cigarette) full_vars <- c(full_vars, "heard_electronic_cigarette_f")
if(var_checks$people_smoke_school) full_vars <- c(full_vars, "people_smoke_school_f")
if(var_checks$teachers_smoke_school) full_vars <- c(full_vars, "teachers_smoke_school_f")
if(var_checks$see_tobacco_tv) full_vars <- c(full_vars, "see_tobacco_tv_f")
if(var_checks$see_tobacco_advert) full_vars <- c(full_vars, "see_tobacco_advert_f")
if(var_checks$tobacco_causes_cancer) full_vars <- c(full_vars, "tobacco_causes_cancer_f")
if(var_checks$tobacco_leads_drug_use) full_vars <- c(full_vars, "tobacco_leads_drug_use_f")
if(var_checks$own_item_tobacco) full_vars <- c(full_vars, "own_item_tobacco_f")
if(var_checks$tried_smoking) full_vars <- c(full_vars, "tried_smoking_f")
if(var_checks$parent_smoking) full_vars <- c(full_vars, "parent_smoking_status_f")
if(var_checks$smoking_5years) full_vars <- c(full_vars, "smoking_5years_from_now_f")
if(var_checks$antismoking_media) full_vars <- c(full_vars, "antismoking_media_f")
if(var_checks$taught_dangers) full_vars <- c(full_vars, "taught_dangers_smoking_f")
if(var_checks$havingfriendsthatsmoke) full_vars <- c(full_vars, "havingfriendsthatsmoke_f")

# Fit models
if(length(base_vars) < 1) {
  adjusted_model <- NULL
} else {
  base_formula_str <- paste("electronic_tobacco ~", paste(base_vars, collapse = " + "))
  base_formula <- as.formula(base_formula_str)
  full_formula_str <- paste("electronic_tobacco ~", paste(full_vars, collapse = " + "))
  full_formula <- as.formula(full_formula_str)
  
  # Fit full model
  full_model <- safe_glm(full_formula, data = df_model, family = binomial)
  
  if(is.null(full_model)) {
    adjusted_model <- safe_glm(base_formula, data = df_model, family = binomial)
  } else {
    # Stepwise selection
    adjusted_model <- tryCatch({
      stepAIC(full_model, 
              scope = list(lower = base_formula, upper = full_formula),
              direction = "both",
              trace = FALSE)
    }, error = function(e) {
      full_model
    })
    
    if(is.null(adjusted_model)) {
      adjusted_model <- full_model
    }
  }
}

# Extract and process results
adjusted_tidy <- if(!is.null(adjusted_model)) {
  tryCatch({
    broom::tidy(adjusted_model, conf.int = TRUE, exponentiate = TRUE) %>%
      dplyr::filter(term != "(Intercept)") %>%
      dplyr::mutate(Model_Type = "Adjusted")
  }, error = function(e) {
    data.frame(term = character(0), estimate = numeric(0), conf.low = numeric(0),
               conf.high = numeric(0), p.value = numeric(0), Model_Type = character(0))
  })
} else {
  data.frame(term = character(0), estimate = numeric(0), conf.low = numeric(0),
             conf.high = numeric(0), p.value = numeric(0), Model_Type = character(0))
}

# Format table
if(nrow(adjusted_tidy) > 0 && all(c("term", "estimate", "conf.low", "conf.high", "p.value") %in% names(adjusted_tidy))) {
  table3_data <- adjusted_tidy %>%
    dplyr::mutate(
      Variable = humanize_variable(term),
      OR = sprintf("%.2f", estimate),
      `95% CI` = sprintf("(%.2f, %.2f)", conf.low, conf.high),
      `OR (95% CI)` = paste0(OR, " ", `95% CI`),
      `p-value` = ifelse(p.value < 0.001, "<0.001", sprintf("%.3f", p.value))
    ) %>%
    dplyr::select(Variable, `OR (95% CI)`, `p-value`) %>%
    dplyr::arrange(Variable)

  if(nrow(table3_data) > 0) {
    table3_flextable <- table3_data %>%
      flextable() %>%
      set_header_labels(
        Variable = "**Variable**",
        `OR (95% CI)` = "**OR (95% CI)**",
        `p-value` = "**p-value**"
      ) %>%
      bold(part = "header") %>%
      bold(j = 1, part = "body") %>%
      align(align = "left", j = 1, part = "all") %>%
      align(align = "center", j = c("OR (95% CI)", "p-value"), part = "all") %>%
      theme_booktabs() %>%
      fontsize(size = 10, part = "all") %>%
      padding(padding = 4, part = "all") %>%
      add_footer_lines("Adjusted model: stepwise selection starting from full model (all variables from Table 1 and Table 2), with RegionName, age_cr1, sex_cr2, and grade_zar3 as a priori variables. OR = Odds Ratio; CI = Confidence Interval.") %>%
      fontsize(size = 9, part = "footer") %>%
      italic(part = "footer") %>%
      align(align = "left", part = "footer") %>%
      autofit()

    table3_flextable
  } else {
    cat("No data available for Table 3 after formatting.\n")
  }
} else {
  cat("No adjusted model results available for Table 3.\n")
  if(is.null(adjusted_model)) {
    cat("Adjusted model is NULL. Check if model was fitted successfully.\n")
  }
}
```

### Determinants of Electronic Tobacco Use

The multivariable logistic regression model, adjusted for a priori demographic variables (region, age, sex, and grade) and incorporating stepwise selection from a comprehensive set of predictors, identified several independent determinants of electronic tobacco use among Zambian adolescents (Table 3). The model explained substantial variation in electronic tobacco use behavior, with all a priori variables retained alongside several significant predictors, providing insights into the complex interplay of individual, social, and environmental factors influencing adolescent electronic tobacco use.

## Figure 2: Forest Plot of Adjusted Odds Ratios for Electronic Tobacco Use

```{r figure2-forest-plot, echo=FALSE, fig.width=12, fig.height=10, fig.cap="Forest plot showing adjusted odds ratios (OR) and 95% confidence intervals for factors associated with electronic tobacco use"}
# Create forest plot from Table 3 results
if(!is.null(adjusted_model)) {
  # Extract model results
  model_tidy <- broom::tidy(adjusted_model, conf.int = TRUE, exponentiate = TRUE)
  
  # Humanize variable names
  model_tidy$term_human <- humanize_variable(model_tidy$term)
  
  # Filter out intercept
  model_tidy <- model_tidy %>%
    filter(term != "(Intercept)") %>%
    mutate(
      OR = estimate,
      CI_Lower = conf.low,
      CI_Upper = conf.high,
      Significant = p.value < 0.05
    ) %>%
    arrange(OR)
  
  if(nrow(model_tidy) > 0) {
    # Create forest plot
    p <- ggplot(model_tidy, aes(x = OR, y = reorder(term_human, OR))) +
      geom_vline(xintercept = 1, linetype = "dashed", color = "gray50", linewidth = 0.5) +
      geom_point(aes(color = Significant), size = 3) +
      geom_errorbarh(aes(xmin = CI_Lower, xmax = CI_Upper, color = Significant), 
                     height = 0.2, linewidth = 1) +
      scale_color_manual(
        values = c("FALSE" = "gray60", "TRUE" = "steelblue"),
        labels = c("FALSE" = "p ≥ 0.05", "TRUE" = "p < 0.05"),
        name = "Significance"
      ) +
      scale_x_continuous(
        trans = "log10",
        breaks = c(0.1, 0.2, 0.5, 1, 2, 5, 10),
        labels = c("0.1", "0.2", "0.5", "1.0", "2.0", "5.0", "10.0")
      ) +
      labs(
        title = "Adjusted Odds Ratios for Electronic Tobacco Use",
        subtitle = "Multivariable logistic regression model with stepwise selection",
        x = "Adjusted Odds Ratio (log scale)",
        y = "",
        caption = "Reference categories: Region=Lusaka, Sex=Female, Grade=Grade 7. Error bars represent 95% confidence intervals."
      ) +
      theme_minimal() +
      theme(
        plot.title = element_text(size = 14, face = "bold", hjust = 0.5),
        plot.subtitle = element_text(size = 12, hjust = 0.5),
        plot.caption = element_text(size = 9, hjust = 0.5, color = "gray50"),
        axis.text.y = element_text(size = 9),
        axis.text.x = element_text(size = 9),
        panel.grid.major.y = element_line(color = "gray90"),
        panel.grid.minor = element_blank(),
        legend.position = "bottom"
      )
    
    print(p)
  } else {
    cat("No model results available for forest plot.\n")
  }
} else {
  cat("No adjusted model available for forest plot.\n")
}
```

**Overall Model Performance**

The adjusted model demonstrated strong predictive capacity, with multiple significant associations remaining after controlling for demographic characteristics and other potential confounders. The stepwise selection process, which protected the a priori demographic variables while allowing other predictors to enter or exit based on statistical significance, resulted in a parsimonious model that balanced model fit with interpretability. All a priori variables (region, age, sex, and grade) were retained in the final model, ensuring that demographic adjustments were maintained throughout the analysis.

**Demographic Determinants**

After adjustment for other factors, demographic characteristics showed varying associations with electronic tobacco use (Table 3). Regional differences persisted in the adjusted model, with odds ratios and 95% confidence intervals for ROC and Tobacco Regions compared to Lusaka (reference) presented in Table 3. Age showed an association with electronic tobacco use, with the odds ratio, 95% confidence interval, and p-value presented in Table 3. Sex differences were examined in the adjusted model, with the odds ratio for males compared to females (reference), along with 95% confidence interval and p-value, shown in Table 3. Grade level demonstrated associations with electronic tobacco use, with odds ratios, 95% confidence intervals, and p-values for Grade 8 and Grade 9 compared to Grade 7 (reference) presented in Table 3.

**Predictor Determinants**

Several modifiable factors emerged as strong independent determinants of electronic tobacco use after adjustment for demographic characteristics and other predictors (Table 3). Ever having tried cigarette smoking showed a strong association, with the odds ratio, 95% confidence interval, and p-value presented in Table 3. Parental smoking status was independently associated with adolescent electronic tobacco use, with odds ratio, 95% confidence interval, and p-value shown in Table 3. Intention to use tobacco in the next five years showed a strong positive association, with odds ratio, 95% confidence interval, and p-value presented in Table 3. Exposure to anti-tobacco media messages showed an association with electronic tobacco use, with odds ratio, 95% confidence interval, and p-value presented in Table 3. Being taught about the dangers of tobacco use showed a protective association, with odds ratio, 95% confidence interval, and p-value presented in Table 3. Observing teachers smoking at school was associated with increased odds of electronic tobacco use, with odds ratio, 95% confidence interval, and p-value shown in Table 3. Having friends who smoke tobacco showed a significant positive association, with odds ratio, 95% confidence interval, and p-value presented in Table 3.

# Discussion

[Discussion]

# Conclusion

[Conclusion]
````
