## Supplemental codes for "Methodological Guidance for Predictor Variable Selection for Adolescent Smoking Outcomes in Global Youth Tobacco Survey Using R and Python": PUBLICATION_QUALITY_ENHANCEMENTS.md

# Publication Quality Assessment & Enhancement Recommendations

## Current Status Summary

### ✅ **STRENGTHS - What's Already Publication-Quality:**

1. **Tables:**
   - ✅ Using `flextable()` package (excellent for publication)
   - ✅ `theme_zebra()` for professional striped appearance
   - ✅ `autofit()` for proper column sizing
   - ✅ Proper numeric formatting with sprintf()
   - ✅ Confidence intervals included where relevant

2. **Figures:**
   - ✅ High-resolution output (300 DPI)
   - ✅ Consistent color scheme across all papers
   - ✅ Proper titles and axis labels
   - ✅ Error bars where appropriate (in main figures)
   - ✅ `theme_minimal()` for clean appearance

3. **Overall Structure:**
   - ✅ Professional YAML headers
   - ✅ Clear figure and table numbering
   - ✅ Proper statistical formatting (ORs, CIs, p-values)

### ⚠️ **AREAS FOR ENHANCEMENT:**

## 1. Table Enhancements Needed

### Issues Found:
- Some tables lack descriptive captions
- Number formatting could be more consistent
- Some tables missing proper headers/spanners
- Missing table footnotes for statistical tests

### Recommendations:
1. **Add descriptive captions** using `set_caption()` or in markdown
2. **Standardize number formatting** (consistent decimal places)
3. **Add table footnotes** explaining abbreviations, statistical tests
4. **Improve header formatting** with merged headers where appropriate
5. **Add borders** using `border_outer()` and `border_inner()` for PDF output

### Example Enhancement:
```r
table1_flextable <- table1_data %>%
  flextable() %>%
  set_caption(caption = "Table 1: Prevalence of Tobacco Use by Product Type and Region, Zambia GYTS 2021") %>%
  set_header_labels(...) %>%
  add_header_row(...) %>%
  theme_zebra() %>%
  border_outer(border = fp_border(color = "black", width = 1)) %>%
  border_inner_h(border = fp_border(color = "gray", width = 0.5)) %>%
  autofit() %>%
  footnote(i = 1, j = 1, 
           value = as_paragraph("CI = confidence interval. Prevalence estimates are weighted."),
           ref_symbols = "a")
```

## 2. Figure Enhancements Needed

### Issues Found:
- Some figures missing error bars/confidence intervals
- Font sizes not standardized across all figures
- Some figures could benefit from better legends
- Missing subtitles explaining what the figure shows
- Some figures don't have proper axis breaks/scales

### Recommendations:
1. **Add error bars** to all bar charts showing estimates
2. **Standardize font sizes**:
   - Title: 14pt bold
   - Subtitle: 11pt
   - Axis labels: 11pt
   - Legend: 10pt
3. **Improve legends**: Clear, descriptive labels
4. **Add figure notes** explaining sample sizes, statistical tests
5. **Use consistent color palettes** (already done - good!)
6. **Add reference lines** where appropriate (e.g., null value at 1.0 in forest plots)

### Example Enhancement:
```r
fig1 <- fig1_data %>%
  ggplot(aes(x = outcome_label, y = prevalence, fill = region)) +
  geom_col(position = position_dodge(width = 0.8), width = 0.7) +
  geom_errorbar(
    aes(ymin = ci_lower, ymax = ci_upper),
    position = position_dodge(width = 0.8),
    width = 0.2,
    color = "black",
    linewidth = 0.5  # Use linewidth instead of size for newer ggplot2
  ) +
  scale_fill_manual(values = region_colors, labels = ...) +
  labs(
    title = "Figure 1: Tobacco Use Prevalence by Region and Product Type",
    subtitle = "Weighted prevalence (%) with 95% confidence intervals. Sample sizes: Lusaka n=XXX, Tobacco Growing n=XXX, Other Zambia n=XXX.",
    x = "Tobacco Product Type",
    y = "Prevalence (%)",
    fill = "Region",
    caption = "Note: Error bars represent 95% confidence intervals. Estimates are weighted for survey design."
  ) +
  theme_minimal(base_size = 11) +
  theme(
    plot.title = element_text(size = 14, face = "bold", hjust = 0),
    plot.subtitle = element_text(size = 10, hjust = 0),
    plot.caption = element_text(size = 9, hjust = 0, face = "italic"),
    axis.title = element_text(size = 11),
    axis.text = element_text(size = 10),
    legend.title = element_text(size = 11, face = "bold"),
    legend.text = element_text(size = 10),
    panel.grid.minor = element_blank(),
    panel.grid.major.x = element_blank()
  )
```

## 3. Specific Files Needing Attention

### Papers with Minimal Tables/Figures:
- **Paper 5 (Reasons)**: Mostly placeholders - needs actual figures when variables identified
- **Paper 8 (Electronic tobacco)**: Placeholder content
- **Paper 14 (FCTC)**: Needs more detailed tables
- **Paper 15 (Availability)**: Missing price visualization

### Papers with Good Quality but Could Be Enhanced:
- **Paper 1**: Already very good, but could add table footnotes
- **Paper 2**: Good structure, could improve figure subtitles
- **Paper 6**: Could add variable importance plots
- **Paper 11**: Good, could add confidence intervals to all estimates

## 4. Overall Recommendations

### Immediate Actions:
1. ✅ **Add error bars to all bar charts** showing prevalence estimates
2. ✅ **Add figure captions/notes** explaining sample sizes and methods
3. ✅ **Add table footnotes** for abbreviations and statistical notes
4. ✅ **Standardize font sizes** across all figures
5. ✅ **Add borders to tables** for PDF output

### Optional Enhancements:
1. **Color-blind friendly palettes**: Consider using `viridis` or `RColorBrewer` palettes
2. **Faceted plots**: Where appropriate, use facets for better comparisons
3. **Annotation**: Add significance indicators (e.g., asterisks for p-values)
4. **Consistent styling**: Create a custom theme function to ensure consistency

### Code to Add to Each File's Setup:

```r
# Publication-quality theme
pub_theme <- function(base_size = 11) {
  theme_minimal(base_size = base_size) +
  theme(
    plot.title = element_text(size = 14, face = "bold", hjust = 0, margin = margin(b = 10)),
    plot.subtitle = element_text(size = 10, hjust = 0, margin = margin(b = 8)),
    plot.caption = element_text(size = 9, hjust = 0, face = "italic", margin = margin(t = 8)),
    axis.title = element_text(size = 11, face = "bold"),
    axis.text = element_text(size = 10),
    legend.title = element_text(size = 11, face = "bold"),
    legend.text = element_text(size = 10),
    legend.position = "bottom",
    panel.grid.minor = element_blank(),
    panel.grid.major.x = element_blank(),
    strip.text = element_text(size = 10, face = "bold")
  )
}

# Use: fig1 + pub_theme()
```

## 5. Publication Standards Checklist

For each table:
- [ ] Descriptive caption
- [ ] Clear column headers
- [ ] Proper number formatting (consistent decimals)
- [ ] Sample sizes included
- [ ] Confidence intervals where appropriate
- [ ] Footnotes for abbreviations
- [ ] Footnotes for statistical tests

For each figure:
- [ ] Clear, descriptive title
- [ ] Subtitle with sample size information
- [ ] Proper axis labels
- [ ] Error bars/confidence intervals
- [ ] Legend with clear labels
- [ ] Figure caption/note
- [ ] High resolution (300 DPI)
- [ ] Appropriate size for publication

## Conclusion

The files have a **solid foundation** for publication-quality output. With the suggested enhancements:
1. Adding error bars consistently
2. Improving table formatting with borders and footnotes
3. Standardizing font sizes and themes
4. Adding descriptive captions and notes

They will meet **top-tier journal publication standards**.
