## Supplemental codes for "Methodological Guidance for Predictor Variable Selection for Adolescent Smoking Outcomes in Global Youth Tobacco Survey Using R and Python": references.bib

@article{who_tobacco_2021,
  title = {Tobacco},
  author = {{World Health Organization}},
  year = {2021},
  journal = {World Health Organization Fact Sheets},
  url = {https://www.who.int/news-room/fact-sheets/detail/tobacco},
  note = {Accessed: 2021}
}

@report{usdhhs_preventing_2012,
  title = {Preventing Tobacco Use Among Youth and Young Adults: A Report of the Surgeon General},
  author = {{U.S. Department of Health and Human Services}},
  year = {2012},
  institution = {U.S. Department of Health and Human Services, Centers for Disease Control and Prevention, National Center for Chronic Disease Prevention and Health Promotion, Office on Smoking and Health},
  address = {Atlanta, GA}
}

@report{who_global_2020,
  title = {WHO Global Report on Trends in Prevalence of Tobacco Use 2000-2025},
  author = {{World Health Organization}},
  year = {2020},
  institution = {World Health Organization},
  address = {Geneva, Switzerland}
}

@misc{who_fctc_2003,
  title = {WHO Framework Convention on Tobacco Control},
  author = {{World Health Organization}},
  year = {2003},
  institution = {World Health Organization},
  address = {Geneva, Switzerland},
  url = {https://www.who.int/fctc/en/}
}

@article{warren_global_2006,
  title = {Global Tobacco Surveillance System (GTSS) Collaborative Group. Patterns of global tobacco use in young people and implications for future chronic disease burden in adults},
  author = {Warren, Charles W. and Jones, Nathan R. and Eriksen, Michael P. and Asma, Samira},
  year = {2006},
  journal = {The Lancet},
  volume = {367},
  number = {9512},
  pages = {749--753},
  doi = {10.1016/S0140-6736(06)68192-0}
}

@report{who_africa_2019,
  title = {Tobacco Control in the WHO African Region: 2019 Status Report},
  author = {{World Health Organization Regional Office for Africa}},
  year = {2019},
  institution = {World Health Organization},
  address = {Brazzaville, Republic of Congo}
}

@article{legg_tobacco_2014,
  title = {Tobacco industry tactics for resisting public policy on health},
  author = {Legg, Trevor and Hatchard, Jenny and Gilmore, Anna B.},
  year = {2014},
  journal = {Bulletin of the World Health Organization},
  volume = {92},
  number = {12},
  pages = {937--945},
  doi = {10.2471/BLT.13.128637}
}

@report{drope_tobacco_2018,
  title = {The Tobacco Atlas},
  author = {Drope, Jeffrey and Schluger, Neil W. and Cahn, Zachary and Drope, Jacqui and Hamill, Sarah and Islami, Farhad and Liber, Alex and Nargis, Nigar and Stoklosa, Michal},
  year = {2018},
  edition = {6th},
  publisher = {American Cancer Society and Vital Strategies},
  address = {Atlanta, GA}
}

@report{who_africa_tobacco_2021,
  title = {Tobacco Control in the WHO African Region: Progress and Challenges},
  author = {{World Health Organization Regional Office for Africa}},
  year = {2021},
  institution = {World Health Organization},
  address = {Brazzaville, Republic of Congo}
}

@report{moh_zambia_2018,
  title = {Zambia National Tobacco Control Policy},
  author = {{Ministry of Health, Zambia}},
  year = {2018},
  institution = {Ministry of Health},
  address = {Lusaka, Zambia}
}

@article{zyambo_gyts_2021,
  title = {Global Youth Tobacco Survey (GYTS) 2021: Zambia National Report},
  author = {Zyambo, M. and {Ministry of Health, Zambia} and {World Health Organization}},
  year = {2021},
  journal = {Ministry of Health, Zambia},
  note = {Unpublished report}
}

@report{who_gyts_2021,
  title = {Global Youth Tobacco Survey (GYTS): Data Summary and Statistics},
  author = {{World Health Organization}},
  year = {2021},
  institution = {World Health Organization},
  address = {Geneva, Switzerland}
}

@report{who_tobacco_control_2021,
  title = {WHO Report on the Global Tobacco Epidemic, 2021: Addressing New and Emerging Products},
  author = {{World Health Organization}},
  year = {2021},
  institution = {World Health Organization},
  address = {Geneva, Switzerland}
}

@article{gyts_collaborative_2002,
  title = {Tobacco use among youth: a cross country comparison},
  author = {{Global Youth Tobacco Survey Collaborative Group}},
  year = {2002},
  journal = {Tobacco Control},
  volume = {11},
  number = {3},
  pages = {252--270},
  doi = {10.1136/tc.11.3.252}
}
