## Supplemental codes for "Methodological Guidance for Predictor Variable Selection for Adolescent Smoking Outcomes in Global Youth Tobacco Survey Using R and Python": RENDERING_NOTE.md

# Rendering Instructions and Notes

## ✅ Data Paths Fixed

All files now use the correct relative path: `"../data/National/GYTS 2021 Zambia (National).XLS"` when rendered from the `draft_papers` directory.

## ⚠️ Before Rendering - Required Steps:

### 1. Verify Variable Names
The code assumes certain variable names exist in your dataset. You need to verify and update:
- **Region/Province variables**: Currently uses `Region` and `Province` - update to actual column names
- **Weight variable**: Currently checks for `WEIGHT` - update if different
- **School ID**: Currently uses `SCHOOL_ID` - update if different  
- **Questionnaire variables**: CR1, CR3, CR52, CR53, CR54, etc. - verify these match your dataset

### 2. Install Required R Packages
Make sure all required packages are installed:
```r
install.packages(c("readxl", "tidyverse", "survey", "gtsummary", "flextable", 
                   "patchwork", "scales", "viridis", "RColorBrewer", 
                   "knitr", "kableExtra", "broom", "lme4", "emmeans", 
                   "ggplot2", "ggthemes", "stringr", "purrr"))
```

### 3. Rendering Options

**Option A: Render from project root:**
```powershell
quarto render draft_papers/paper01_tobacco_growing_association.qmd
```

**Option B: Render from draft_papers directory:**
```powershell
cd draft_papers
quarto render paper01_tobacco_growing_association.qmd
```

**Option C: Render all papers:**
```powershell
cd draft_papers
quarto render *.qmd
```

## Current Status

- ✅ All data paths corrected
- ✅ Missing packages removed (forestplot, sjPlot)
- ✅ Humanization functions added to key files
- ⚠️ Variable names need verification against actual dataset
- ⚠️ First render will likely show variable name errors that need fixing

## Expected Errors on First Render

1. **Variable not found errors**: Region, Province, CR variables - need to verify actual column names
2. **Missing data**: If Excel file path is incorrect or sheet name doesn't match
3. **Model errors**: If SCHOOL_ID doesn't exist - models will need adjustment

## After Variable Names Are Verified

Once you update the variable names to match your actual dataset, the papers should render successfully with beautiful tables and figures ready for publication!
