## Supplemental codes for "Methodological Guidance for Predictor Variable Selection for Adolescent Smoking Outcomes in Global Youth Tobacco Survey Using R and Python": S1_Safe_extraction.R

```
#R: Safe Variable Extraction Function

# Safe variable extraction with fallback for missing columns
safe_get_col <- function(dataset, col_name, default_val = NA) {
  if(col_name %in% names(dataset)) {
    return(dataset[[col_name]])
  } else {
    # Case-insensitive match for variable naming variations
    match_col <- names(dataset)[grepl(
      paste0("^", col_name, "$"), 
      names(dataset), 
      ignore.case = TRUE
    )][1]
    if(!is.na(match_col)) {
      return(dataset[[match_col]])
    } else {
      return(rep(default_val, nrow(dataset)))
    }
  }
}

# Example: Age harmonization from CR1
age_cr1 <- case_when(
  as.numeric(CR_vars[["CR1"]]) == 1 ~ 11,  # 11 years or younger
  as.numeric(CR_vars[["CR1"]]) == 2 ~ 12,  # 12 years old
  as.numeric(CR_vars[["CR1"]]) == 3 ~ 13,  # 13 years old
  as.numeric(CR_vars[["CR1"]]) == 4 ~ 14,  # 14 years old
  as.numeric(CR_vars[["CR1"]]) == 5 ~ 15,  # 15 years old
  as.numeric(CR_vars[["CR1"]]) == 6 ~ 16,  # 16 years old
  as.numeric(CR_vars[["CR1"]]) == 7 ~ 17,  # 17 years old or older
  TRUE ~ NA_real_
)
```
