## Supplemental codes for "Methodological Guidance for Predictor Variable Selection for Adolescent Smoking Outcomes in Global Youth Tobacco Survey Using R and Python": S2a_Complex_survey_weighted_two_stage_design.R

```
#R Implementation (survey package):
#  R: Complex Survey Design Specification
library(survey)
# Two-stage design with nesting
svy_design <- svydesign(
  ids = ~PSU,                    # Cluster (school)
  strata = ~Stratum,             # Stratification variable
  weights = ~FinalWgt,           # Survey weights
  data = df,
  nest = TRUE                    # PSUs nested within strata
)
# Key features:
# - nest = TRUE ensures proper handling of PSU identifiers reused across strata
# - svydesign automatically handles finite population corrections
# - Supports both stratified and unstratified designs
```
