## Supplemental codes for "Methodological Guidance for Predictor Variable Selection for Adolescent Smoking Outcomes in Global Youth Tobacco Survey Using R and Python": S3a_Stepwise_selection.R

```
#R: Constrained Stepwise Selection

library(MASS)

# Define variable groups based on theoretical framework
apriori_vars <- c("RegionName", "age_cr1", "sex_cr2", "grade_zar3")

individual_vars <- c("tried_smoking_f", "smoked_cigarettes_f", "tried_shisha_f", 
                     "smoking_5years_from_now_f", "wealth_index_f")

social_vars <- c("parent_smoking_status_f", "havingfriendsthatsmoke_f", 
                 "people_smoke_school_f", "teachers_smoke_school_f")

media_vars <- c("tobacco_awareness_f", "see_tobacco_tv_f", 
                "see_tobacco_advert_f", "antismoking_media_f", "own_item_tobacco_f")

policy_vars <- c("taught_dangers_smoking_f", "ban_closed_smoking_f", 
                 "ban_public_smoking_f")

# Combine all candidate predictors
candidate_vars <- c(individual_vars, social_vars, media_vars, policy_vars)

# Build formulas: lower = a priori only, upper = full model
base_formula <- as.formula(
  paste("smoking_behaviour ~", paste(apriori_vars, collapse = " + "))
)

full_formula <- as.formula(
  paste("smoking_behaviour ~", 
        paste(c(apriori_vars, candidate_vars), collapse = " + "))
)

# Survey-weighted logistic regression
full_model <- svyglm(full_formula, 
                     design = svy_design, 
                     family = quasibinomial(link = "logit"))

# Stepwise selection with constraints
# scope = list(lower = base_formula, upper = full_formula) protects a priori vars
final_model <- stepAIC(
  full_model, 
  scope = list(lower = base_formula, upper = full_formula),
  direction = "both",  # Forward and backward selection
  trace = FALSE
)

# Extract results with odds ratios and confidence intervals
library(broom)
results <- tidy(final_model, exponentiate = TRUE, conf.int = TRUE)
```
