## Supplemental codes for "Methodological Guidance for Predictor Variable Selection for Adolescent Smoking Outcomes in Global Youth Tobacco Survey Using R and Python": update_papers_9_20.py

````
#!/usr/bin/env python3
"""
Script to update papers 9-20 with robust data-loading section.
This script reads each paper, finds the data-loading section, and replaces it
with the robust template while preserving paper-specific variables.
"""

import re
import os

# Read the robust template (from paper07)
template_file = "paper07_factors_smokeless.qmd"
with open(template_file, 'r', encoding='utf-8') as f:
    template_content = f.read()

# Extract the data-loading section from template
template_match = re.search(r'```\{r data-loading[^`]+```', template_content, re.DOTALL)
if not template_match:
    print("ERROR: Could not find data-loading section in template")
    exit(1)

robust_data_loading = template_match.group(0)

# Paper-specific variables for each paper (from grep results)
paper_specific_vars = {
    9: """    # Paper-specific variables
    age_initiation = {
      cr2 <- CR_vars[["CR2"]]
      if(!all(is.na(cr2))) {
        case_when(
          as.numeric(cr2) == 2 ~ "≤7", as.numeric(cr2) == 3 ~ "8-9", as.numeric(cr2) == 4 ~ "10-11",
          as.numeric(cr2) == 5 ~ "12-13", as.numeric(cr2) == 6 ~ "14-15", as.numeric(cr2) == 7 ~ "≥16",
          TRUE ~ "Never"
        )
      } else {
        rep("Never", nrow(df_raw))
      }
    },
    early_initiation = {
      cr2 <- CR_vars[["CR2"]]
      if(!all(is.na(cr2))) {
        case_when(as.numeric(cr2) <= 4 ~ 1, as.numeric(cr2) > 4 ~ 0, TRUE ~ NA_real_)
      } else {
        rep(NA_real_, nrow(df_raw))
      }
    },
    never_smoked = {
      cr1 <- CR_vars[["CR1"]]
      if(!all(is.na(cr1)) && any(as.numeric(cr1) == 2, na.rm = TRUE)) {
        case_when(as.numeric(cr1) == 2 ~ 1, TRUE ~ 0)
      } else {
        rep(0, nrow(df_raw))
      }
    },
    susceptibility = {
      cr13 <- CR_vars[["CR13"]]
      cr15 <- CR_vars[["CR15"]]
      if((!all(is.na(cr13)) && any(as.numeric(cr13) >= 3, na.rm = TRUE)) ||
         (!all(is.na(cr15)) && any(as.numeric(cr15) >= 3, na.rm = TRUE))) {
        case_when(as.numeric(cr13) >= 3 | as.numeric(cr15) >= 3 ~ 1, TRUE ~ 0)
      } else {
        rep(0, nrow(df_raw))
      }
    }""",
    10: """    # Paper-specific variables
    any_tobacco = {
      cr3 <- CR_vars[["CR3"]]
      cr9 <- CR_vars[["CR9"]]
      cr8 <- CR_vars[["CR8"]]
      if((!all(is.na(cr3)) && any(as.numeric(cr3) > 1, na.rm = TRUE)) ||
         (!all(is.na(cr9)) && any(as.numeric(cr9) == 1, na.rm = TRUE)) ||
         (!all(is.na(cr8)) && any(as.numeric(cr8) == 1, na.rm = TRUE))) {
        case_when(
          as.numeric(cr3) > 1 | as.numeric(cr9) == 1 | as.numeric(cr8) == 1 ~ 1,
          TRUE ~ 0
        )
      } else {
        rep(0, nrow(df_raw))
      }
    }""",
    11: """    # Paper-specific variables
    shs_home = {
      zmr35 <- ZMR_vars[["ZMR35"]]
      if(!all(is.na(zmr35)) && any(as.numeric(zmr35) > 1, na.rm = TRUE)) {
        case_when(as.numeric(zmr35) > 1 ~ 1, TRUE ~ 0)
      } else {
        rep(0, nrow(df_raw))
      }
    },
    shs_public = {
      zmr36 <- ZMR_vars[["ZMR36"]]
      if(!all(is.na(zmr36)) && any(as.numeric(zmr36) > 1, na.rm = TRUE)) {
        case_when(as.numeric(zmr36) > 1 ~ 1, TRUE ~ 0)
      } else {
        rep(0, nrow(df_raw))
      }
    },
    shs_any = case_when(shs_home == 1 | shs_public == 1 ~ 1, TRUE ~ 0)""",
    12: """    # Paper-specific variables
    ever_smoked = {
      cr1 <- CR_vars[["CR1"]]
      if(!all(is.na(cr1)) && any(as.numeric(cr1) == 1, na.rm = TRUE)) {
        case_when(as.numeric(cr1) == 1 ~ 1, TRUE ~ 0)
      } else {
        rep(0, nrow(df_raw))
      }
    },
    current_smoker = {
      cr3 <- CR_vars[["CR3"]]
      if(!all(is.na(cr3)) && any(as.numeric(cr3) > 1, na.rm = TRUE)) {
        case_when(as.numeric(cr3) > 1 ~ 1, TRUE ~ 0)
      } else {
        rep(0, nrow(df_raw))
      }
    },
    former_smoker = case_when(ever_smoked == 1 & current_smoker == 0 ~ 1, TRUE ~ 0),
    quit_attempt = {
      cr34 <- CR_vars[["CR34"]]
      if(!all(is.na(cr34))) {
        case_when(as.numeric(cr34) == 3 ~ 1, as.numeric(cr34) == 4 ~ 0, TRUE ~ NA_real_)
      } else {
        rep(NA_real_, nrow(df_raw))
      }
    },
    want_to_stop = {
      cr33 <- CR_vars[["CR33"]]
      if(!all(is.na(cr33))) {
        case_when(as.numeric(cr33) == 3 ~ 1, as.numeric(cr33) == 4 ~ 0, TRUE ~ NA_real_)
      } else {
        rep(NA_real_, nrow(df_raw))
      }
    },
    received_help = {
      cr38 <- CR_vars[["CR38"]]
      if(!all(is.na(cr38))) {
        case_when(as.numeric(cr38) >= 2 ~ 1, as.numeric(cr38) == 6 ~ 0, TRUE ~ NA_real_)
      } else {
        rep(NA_real_, nrow(df_raw))
      }
    }""",
    13: """    # Paper-specific variables
    intention_next12m = {
      cr15 <- CR_vars[["CR15"]]
      if(!all(is.na(cr15)) && any(as.numeric(cr15) >= 3, na.rm = TRUE)) {
        case_when(as.numeric(cr15) >= 3 ~ 1, TRUE ~ 0)
      } else {
        rep(0, nrow(df_raw))
      }
    },
    intention_5yr = {
      cr16 <- CR_vars[["CR16"]]
      if(!all(is.na(cr16)) && any(as.numeric(cr16) >= 3, na.rm = TRUE)) {
        case_when(as.numeric(cr16) >= 3 ~ 1, TRUE ~ 0)
      } else {
        rep(0, nrow(df_raw))
      }
    },
    attitude_positive = {
      cr21 <- CR_vars[["CR21"]]
      cr22 <- CR_vars[["CR22"]]
      if((!all(is.na(cr21)) && any(as.numeric(cr21) == 1, na.rm = TRUE)) ||
         (!all(is.na(cr22)) && any(as.numeric(cr22) == 1, na.rm = TRUE))) {
        case_when(as.numeric(cr21) == 1 | as.numeric(cr22) == 1 ~ 1, TRUE ~ 0)
      } else {
        rep(0, nrow(df_raw))
      }
    },
    friends_smoke = case_when(friendsmokingcigarettes >= 2 ~ 1, TRUE ~ 0),
    difficult_quit = {
      cr17 <- CR_vars[["CR17"]]
      if(!all(is.na(cr17)) && any(as.numeric(cr17) >= 3, na.rm = TRUE)) {
        case_when(as.numeric(cr17) >= 3 ~ 1, TRUE ~ 0)
      } else {
        rep(0, nrow(df_raw))
      }
    }""",
    14: """    # Paper-specific variables
    # FCTC indicators will be calculated in analysis section""",
    15: """    # Paper-specific variables
    easy_access = {
      cr7 <- CR_vars[["CR7"]]
      if(!all(is.na(cr7)) && any(as.numeric(cr7) == 3, na.rm = TRUE)) {
        case_when(as.numeric(cr7) == 3 ~ "Easy", TRUE ~ "Not easy")
      } else {
        rep("Not easy", nrow(df_raw))
      }
    },
    refused_sale = {
      cr7 <- CR_vars[["CR7"]]
      if(!all(is.na(cr7)) && any(as.numeric(cr7) == 2, na.rm = TRUE)) {
        case_when(as.numeric(cr7) == 2 ~ "Yes", TRUE ~ "No")
      } else {
        rep("No", nrow(df_raw))
      }
    },
    knows_mishanga = case_when(placessellingmishanga == 1 ~ "Yes", TRUE ~ "No"),
    price_paid = NA""",
    16: """    # Paper-specific variables
    taught_dangers = case_when(taughtdangersofsmoking == 1 ~ 1, TRUE ~ 0),
    discussed_reasons = case_when(reasonagegroupsmoke == 1 ~ 1, TRUE ~ 0),
    taught_effects = case_when(taughteffectsofsmoking == 1 ~ 1, TRUE ~ 0),
    policy_score = taught_dangers + discussed_reasons + taught_effects,
    policy_category = case_when(
      policy_score == 0 ~ "None", policy_score == 1 ~ "Low",
      policy_score == 2 ~ "Moderate", policy_score == 3 ~ "High"
    ),
    current_smoking = {
      cr3 <- CR_vars[["CR3"]]
      if(!all(is.na(cr3)) && any(as.numeric(cr3) > 1, na.rm = TRUE)) {
        case_when(as.numeric(cr3) > 1 ~ 1, TRUE ~ 0)
      } else {
        rep(0, nrow(df_raw))
      }
    }""",
    17: """    # Paper-specific variables
    knows_harmful = {
      cr24 <- CR_vars[["CR24"]]
      if(!all(is.na(cr24)) && any(as.numeric(cr24) >= 3, na.rm = TRUE)) {
        case_when(as.numeric(cr24) >= 3 ~ 1, TRUE ~ 0)
      } else {
        rep(0, nrow(df_raw))
      }
    },
    knows_passive_harmful = {
      cr29 <- CR_vars[["CR29"]]
      if(!all(is.na(cr29)) && any(as.numeric(cr29) >= 3, na.rm = TRUE)) {
        case_when(as.numeric(cr29) >= 3 ~ 1, TRUE ~ 0)
      } else {
        rep(0, nrow(df_raw))
      }
    },
    knows_safe_shortterm = {
      cr28 <- CR_vars[["CR28"]]
      if(!all(is.na(cr28)) && any(as.numeric(cr28) <= 2, na.rm = TRUE)) {
        case_when(as.numeric(cr28) <= 2 ~ 1, TRUE ~ 0)
      } else {
        rep(0, nrow(df_raw))
      }
    },
    boys_attractive = {
      cr21 <- CR_vars[["CR21"]]
      if(!all(is.na(cr21))) {
        case_when(as.numeric(cr21) == 1 ~ "More", as.numeric(cr21) == 2 ~ "Less", TRUE ~ "No diff")
      } else {
        rep("No diff", nrow(df_raw))
      }
    },
    girls_attractive = {
      cr22 <- CR_vars[["CR22"]]
      if(!all(is.na(cr22))) {
        case_when(as.numeric(cr22) == 1 ~ "More", as.numeric(cr22) == 2 ~ "Less", TRUE ~ "No diff")
      } else {
        rep("No diff", nrow(df_raw))
      }
    },
    weight_control = {
      cr23 <- CR_vars[["CR23"]]
      if(!all(is.na(cr23)) && any(as.numeric(cr23) == 2, na.rm = TRUE)) {
        case_when(as.numeric(cr23) == 2 ~ "Lose", TRUE ~ "Gain/No diff")
      } else {
        rep("Gain/No diff", nrow(df_raw))
      }
    },
    difficult_quit = {
      cr17 <- CR_vars[["CR17"]]
      if(!all(is.na(cr17)) && any(as.numeric(cr17) >= 3, na.rm = TRUE)) {
        case_when(as.numeric(cr17) >= 3 ~ 1, TRUE ~ 0)
      } else {
        rep(0, nrow(df_raw))
      }
    },
    current_smoking = {
      cr3 <- CR_vars[["CR3"]]
      if(!all(is.na(cr3)) && any(as.numeric(cr3) > 1, na.rm = TRUE)) {
        case_when(as.numeric(cr3) > 1 ~ 1, TRUE ~ 0)
      } else {
        rep(0, nrow(df_raw))
      }
    }""",
    18: """    # Paper-specific variables
    tv_ads = case_when(advertisingcigarettebrands <= 2 ~ "High", advertisingcigarettebrands == 3 ~ "Low", TRUE ~ "None"),
    billboard_ads = case_when(cigarettesadvertsseenbillboards <= 2 ~ "High", cigarettesadvertsseenbillboards == 3 ~ "Low", TRUE ~ "None"),
    print_ads = case_when(cigarettesadvertsseeninewspapers <= 2 ~ "High", cigarettesadvertsseeninewspapers == 3 ~ "Low", TRUE ~ "None"),
    pos_ads = case_when(advertoftobaccoatshops <= 2 ~ "High", advertoftobaccoatshops == 3 ~ "Low", TRUE ~ "None"),
    has_brand_item = case_when(somethingwithacigarettebrand == 1 ~ "Yes", TRUE ~ "No"),
    free_cig = case_when(beenofferedfreecig == 2 ~ "Yes", TRUE ~ "No"),
    ad_exposure_count = case_when(tv_ads == "High" ~ 1, TRUE ~ 0) + case_when(billboard_ads == "High" ~ 1, TRUE ~ 0) + case_when(print_ads == "High" ~ 1, TRUE ~ 0) + case_when(pos_ads == "High" ~ 1, TRUE ~ 0),
    current_smoking = {
      cr3 <- CR_vars[["CR3"]]
      if(!all(is.na(cr3)) && any(as.numeric(cr3) > 1, na.rm = TRUE)) {
        case_when(as.numeric(cr3) > 1 ~ 1, TRUE ~ 0)
      } else {
        rep(0, nrow(df_raw))
      }
    }""",
    19: """    # Paper-specific variables
    anti_smoking_media = case_when(antismokingheardpastonemonth == 1 ~ "A lot", antismokingheardpastonemonth == 2 ~ "A few", TRUE ~ "None"),
    anti_smoking_events = {
      cr40 <- CR_vars[["CR40"]]
      if(!all(is.na(cr40))) {
        case_when(as.numeric(cr40) == 2 ~ "A lot", as.numeric(cr40) == 3 ~ "Sometimes", TRUE ~ "Never")
      } else {
        rep("Never", nrow(df_raw))
      }
    },
    anti_smoking_radio = case_when(antismokingmessageonradio == 2 ~ "Many", antismokingmessageonradio == 3 ~ "Few", TRUE ~ "None"),
    youth_groups = case_when(youthgroupsdiscouragingpeople == 2 ~ "Yes", TRUE ~ "No"),
    health_worker = case_when(healthworkerexplaindangers == 2 ~ "Yes", TRUE ~ "No"),
    church_group = case_when(churchgroupdiscouragesmoking == 2 ~ "Yes", TRUE ~ "No"),
    current_smoking = {
      cr3 <- CR_vars[["CR3"]]
      if(!all(is.na(cr3)) && any(as.numeric(cr3) > 1, na.rm = TRUE)) {
        case_when(as.numeric(cr3) > 1 ~ 1, TRUE ~ 0)
      } else {
        rep(0, nrow(df_raw))
      }
    }""",
    20: """    # Paper-specific variables
    current_smoker = {
      cr3 <- CR_vars[["CR3"]]
      if(!all(is.na(cr3)) && any(as.numeric(cr3) > 1, na.rm = TRUE)) {
        case_when(as.numeric(cr3) > 1 ~ 1, TRUE ~ 0)
      } else {
        rep(0, nrow(df_raw))
      }
    },
    days_smoked = {
      cr3 <- CR_vars[["CR3"]]
      if(!all(is.na(cr3))) {
        case_when(
          as.numeric(cr3) == 7 ~ "Daily", as.numeric(cr3) >= 5 ~ "Frequent",
          as.numeric(cr3) >= 3 ~ "Occasional", TRUE ~ "None"
        )
      } else {
        rep("None", nrow(df_raw))
      }
    },
    cigs_per_day = {
      cr4 <- CR_vars[["CR4"]]
      if(!all(is.na(cr4))) {
        case_when(
          as.numeric(cr4) >= 7 ~ "20+", as.numeric(cr4) >= 5 ~ "6-20",
          as.numeric(cr4) >= 3 ~ "1-5", TRUE ~ "None"
        )
      } else {
        rep("None", nrow(df_raw))
      }
    }"""
}

# Update each paper
for paper_num in range(9, 21):
    paper_file = f"paper{paper_num:02d}_*.qmd"
    # Find the actual file name
    import glob
    files = glob.glob(paper_file)
    if not files:
        print(f"WARNING: Could not find paper {paper_num}")
        continue
    
    paper_file = files[0]
    print(f"Updating {paper_file}...")
    
    # Read the paper
    with open(paper_file, 'r', encoding='utf-8') as f:
        content = f.read()
    
    # Find the data-loading section
    pattern = r'```\{r data-loading[^`]+```'
    match = re.search(pattern, content, re.DOTALL)
    if not match:
        print(f"  WARNING: Could not find data-loading section in {paper_file}")
        continue
    
    # Replace the paper-specific variables section in the template
    # Find where to insert paper-specific variables (before the closing of mutate)
    template_with_vars = robust_data_loading.replace(
        '    # Paper-specific variables\n    dual_use = case_when(current_smokeless == 1 & current_cigarette == 1 ~ 1, TRUE ~ 0)',
        paper_specific_vars[paper_num]
    )
    
    # Replace in content
    new_content = content[:match.start()] + template_with_vars + content[match.end():]
    
    # Write back
    with open(paper_file, 'w', encoding='utf-8') as f:
        f.write(new_content)
    
    print(f"  ✓ Updated {paper_file}")

print("\nDone! All papers 9-20 have been updated.")
````
