## Supplemental codes for "Methodological Guidance for Predictor Variable Selection for Adolescent Smoking Outcomes in Global Youth Tobacco Survey Using R and Python": UPDATE_SUMMARY.md

# Summary of Predictor Variable Updates

## Papers Already Updated:
- ✅ Paper 1: tobacco_growing_association
- ✅ Paper 2: factors_shisha  
- ✅ Paper 6: factors_cigarette_smoking
- ✅ Paper 7: factors_smokeless
- ✅ Paper 8: factors_electronic_tobacco (reference)
- ✅ Paper 12: cessation_quitting (previously updated)
- ✅ Paper 13: intention_smoke (previously updated)
- ✅ Paper 18: advertisement_promotion (previously updated)
- ✅ Paper 19: antitobacco_messages (previously updated)

## Papers Still Needing Updates:
- ⏳ Paper 3: trends_lusaka
- ⏳ Paper 4: teaching_harmful_effects
- ⏳ Paper 5: reasons_smoke
- ⏳ Paper 9: initiating_smoking
- ⏳ Paper 10: factors_tobacco_general
- ⏳ Paper 11: secondhand_smoke
- ⏳ Paper 14: fctc_linkage
- ⏳ Paper 15: availability_affordability
- ⏳ Paper 16: school_policy
- ⏳ Paper 17: attitudes_beliefs
- ⏳ Paper 20: addiction_tobacco

## Variables to Add to Each Paper:

### 1. Update Variable Extraction Lists:
```r
# Add to elr_var_names:
elr_var_names <- c("ELR1", "ELR2")  # Add ELR1 if not present

# Add to zar_var_names:
zar_var_names <- c("ZAR3", "ZAR4", "ZAR53", "ZAR55", "ZAR64", "ZAR65")  # Add ZAR53, ZAR64, ZAR65

# Add SR variables if not present:
sr_var_names <- c("SR1")
```

### 2. Add Variable Extraction:
```r
ELR_vars <- list()
for(var in elr_var_names) {
  ELR_vars[[var]] <- safe_get_col(var, NA_real_)
}
SR_vars <- list()
for(var in sr_var_names) {
  SR_vars[[var]] <- safe_get_col(var, NA_real_)
}
```

### 3. Add New Predictor Variables (before closing mutate() or before factor versions):
```r
# Additional predictor variables from paper 8
zar4_raw = {
  zar4 <- ZAR_vars[["ZAR4"]]
  if(!all(is.na(zar4))) {
    as.numeric(zar4)
  } else {
    rep(NA_real_, nrow(GYTS_2021_Zambia_National_))
  }
},
handrolled_cigarette = {
  or5 <- OR_vars[["OR5"]]
  if(!all(is.na(or5))) {
    case_when(as.numeric(or5) == 1 ~ 0, TRUE ~ 1)
  } else { rep(NA_real_, nrow(GYTS_2021_Zambia_National_)) }
},
smoked_cigarettes = {
  cr8 <- CR_vars[["CR8"]]
  if(!all(is.na(cr8))) {
    case_when(as.numeric(cr8) == 1 ~ 0, as.numeric(cr8) > 1 ~ 1, TRUE ~ NA_real_)
  } else { rep(NA_real_, nrow(GYTS_2021_Zambia_National_)) }
},
tried_shisha = {
  sr1 <- SR_vars[["SR1"]]
  if(!all(is.na(sr1))) {
    case_when(as.numeric(sr1) == 1 ~ 0, as.numeric(sr1) == 2 ~ 1, TRUE ~ NA_real_)
  } else { rep(NA_real_, nrow(GYTS_2021_Zambia_National_)) }
},
heard_electronic_cigarette = {
  elr1 <- ELR_vars[["ELR1"]]
  if(!all(is.na(elr1))) {
    case_when(as.numeric(elr1) == 2 ~ 0, as.numeric(elr1) == 1 ~ 1, TRUE ~ NA_real_)
  } else { rep(NA_real_, nrow(GYTS_2021_Zambia_National_)) }
},
father_smokes = {
  or15 <- OR_vars[["OR15"]]
  if(!all(is.na(or15))) {
    case_when(as.numeric(or15) == 1 ~ 0, as.numeric(or15) == 2 ~ 1, as.numeric(or15) == 3 ~ 1, as.numeric(or15) == 4 ~ 0, TRUE ~ NA_real_)
  } else { rep(NA_real_, nrow(GYTS_2021_Zambia_National_)) }
},
people_smoke_school = {
  cr22 <- CR_vars[["CR22"]]
  if(!all(is.na(cr22))) {
    case_when(as.numeric(cr22) == 1 ~ 1, TRUE ~ 0)
  } else { rep(NA_real_, nrow(GYTS_2021_Zambia_National_)) }
},
teachers_smoke_school = {
  or60 <- OR_vars[["OR60"]]
  if(!all(is.na(or60))) {
    case_when(as.numeric(or60) > 2 ~ 0, TRUE ~ 1)
  } else { rep(NA_real_, nrow(GYTS_2021_Zambia_National_)) }
},
see_tobacco_tv = {
  cr34 <- CR_vars[["CR34"]]
  if(!all(is.na(cr34))) {
    case_when(as.numeric(cr34) == 2 ~ 1, TRUE ~ 0)
  } else { rep(NA_real_, nrow(GYTS_2021_Zambia_National_)) }
},
see_tobacco_advert = {
  cr35 <- CR_vars[["CR35"]]
  if(!all(is.na(cr35))) {
    case_when(as.numeric(cr35) == 2 ~ 1, TRUE ~ 0)
  } else { rep(NA_real_, nrow(GYTS_2021_Zambia_National_)) }
},
tobacco_causes_cancer = {
  zar64 <- ZAR_vars[["ZAR64"]]
  if(!all(is.na(zar64))) {
    case_when(as.numeric(zar64) == 1 ~ 1, TRUE ~ 0)
  } else { rep(NA_real_, nrow(GYTS_2021_Zambia_National_)) }
},
tobacco_leads_drug_use = {
  zar65 <- ZAR_vars[["ZAR65"]]
  if(!all(is.na(zar65))) {
    case_when(as.numeric(zar65) == 1 ~ 1, TRUE ~ 0)
  } else { rep(NA_real_, nrow(GYTS_2021_Zambia_National_)) }
},
own_item_tobacco = {
  zar53 <- ZAR_vars[["ZAR53"]]
  if(!all(is.na(zar53))) {
    case_when(as.numeric(zar53) == 1 ~ 1, TRUE ~ 0)
  } else { rep(NA_real_, nrow(GYTS_2021_Zambia_National_)) }
}
```

### 4. Add Factor Versions (after all variables are created, before survey design):
```r
# Create factor versions for additional predictors
df <- df %>%
  mutate(
    handrolled_cigarette_f = factor(handrolled_cigarette, levels = c(0, 1), labels = c("No", "Yes")),
    smoked_cigarettes_f = factor(smoked_cigarettes, levels = c(0, 1), labels = c("No", "Yes")),
    tried_shisha_f = factor(tried_shisha, levels = c(0, 1), labels = c("No", "Yes")),
    heard_electronic_cigarette_f = factor(heard_electronic_cigarette, levels = c(0, 1), labels = c("No", "Yes")),
    father_smokes_f = factor(father_smokes, levels = c(0, 1), labels = c("No", "Yes")),
    people_smoke_school_f = factor(people_smoke_school, levels = c(0, 1), labels = c("No", "Yes")),
    teachers_smoke_school_f = factor(teachers_smoke_school, levels = c(0, 1), labels = c("No", "Yes")),
    see_tobacco_tv_f = factor(see_tobacco_tv, levels = c(0, 1), labels = c("No", "Yes")),
    see_tobacco_advert_f = factor(see_tobacco_advert, levels = c(0, 1), labels = c("No", "Yes")),
    tobacco_causes_cancer_f = factor(tobacco_causes_cancer, levels = c(0, 1), labels = c("No", "Yes")),
    tobacco_leads_drug_use_f = factor(tobacco_leads_drug_use, levels = c(0, 1), labels = c("No", "Yes")),
    own_item_tobacco_f = factor(own_item_tobacco, levels = c(0, 1), labels = c("No", "Yes"))
  )

# Create wealth index groups from ZAR4 (three groups: low, middle, rich)
df <- df %>%
  mutate(
    wealth_index = case_when(
      !is.na(zar4_raw) ~ {
        tertiles <- quantile(zar4_raw, probs = c(0, 1/3, 2/3, 1), na.rm = TRUE)
        case_when(
          zar4_raw <= tertiles[2] ~ "Low",
          zar4_raw <= tertiles[3] ~ "Middle",
          TRUE ~ "Rich"
        )
      },
      TRUE ~ NA_character_
    ),
    wealth_index_f = factor(wealth_index, levels = c("Low", "Middle", "Rich"), labels = c("Low", "Middle", "Rich"))
  )
```

### 5. Update Tables:
- Update variables table to include new variables
- Update Table 1 (characteristics) to include new variables
- Update Table 2 (prevalence) to include new variables  
- Update Table 3 (regression) to include new variables in model
- Update humanize_variable function to include new variable labels
